# Creation of a tool for the Identification of Neurodevelopmental Disabilities to Improve Global Outcomes (INDIGO) in children aged 0-3 years in Malawi, Pakistan and Uganda – feasibility of implementation and diagnostic accuracy

**DOI:** 10.1101/2025.11.13.25340081

**Authors:** M. Gladstone, G. McCray, H Babikako, M Rasheed, E Stockdale, A Kakooza Mwesige, P Chand, E. Mbale, L Maliwichi, P. Lynch, K Bromley, A Abubakar, H Kitt, K.A Donald, M Van den Heuvel, H.M Nabwera, G. A. Lancaster

## Abstract

**Background:** Over 50 million children under five have disabilities, the majority residing in resource-limited settings. Many children with neurodevelopmental disabilities (NDDs) remain undetected due to lack of culturally appropriate tools for identification. This study aimed to develop and evaluate the feasibility of the Identification of Neurodevelopmental Disabilities to Improve Global Outcomes (INDIGO) tool to identify NDDs in children aged 0 −3 years.

**Methods:** A systematic approach was used to construct the INDIGO tool. First, 114 items of the Malawi Developmental Assessment Tool (MDAT) were mapped onto the International Classification of Functioning, Disability and Health for Children and Youth (ICF-CY). Gaps were identified leading to review of 2,158 items from existing tools. After consensus, 59 additional items (28 existing, 31 newly created) were incorporated, ensuring comprehensive coverage of ICF-CY domains. The preliminary INDIGO tool (203 items) was pilot tested in Uganda, Malawi and Pakistan with a gold standard assessment protocol developed to validate diagnostic accuracy. Feasibility outcomes including recruitment, diagnostic accuracy, cultural applicability and practical implementation were assessed. Items with over 80% diagnostic accuracy were retained for the final INDIGO prototype.

**Results:** A total of 425 children (151 Malawi, 145 Pakistan, 129 Uganda) were assessed using both INDIGO and gold standard evaluation. The feasibility study highlighted challenges in recruiting younger children with behavioural or socioemotional conditions and difficulties assessing sensory impairment due to resource constraints. Despite challenges, 99 high performing items were selected for the final INDIGO prototype emphasizing diagnostic accuracy, feasibility and cultural neutrality.

**Conclusions:** The INDIGO tool is a novel, rigorously developed instrument designed for early detection of NDDs in children 0-3 years in LMICs. By integrating a broad range of developmental domains, INDIGO addressed key limitations of existing tools. Future large-scale validation and implementation studies are needed to assess its effectiveness in routine child health surveillance programmes.

**Key Messages:** *What is already known on this topic:*

- Most existing developmental screening tools used globally are not culturally adapted for LMICs, require specialized training, and fail to comprehensively assess structural, functional, and environmental factors important for detecting NDDs in early childhood.

*What this study adds:*

- This study presents the development and feasibility testing of the INDIGO tool, a novel screening instrument for identifying moderate to severe NDDs in children under 3 years in resource-limited settings, incorporating items to assess structural anomalies, sensory impairments, and participation, offering a more holistic and contextually relevant approach to NDD detection.
- The study provides strong initial evidence of diagnostic accuracy across multiple LMIC settings, with high feasibility and acceptability among caregivers and health workers.

*How this study might affect research, practice or policy:*

- Future studies could enable scaled up implementation of tools such as INDIGO integrated into national child health systems for early NDD detection, enhancing referral and intervention in LMICs.

## Introduction

Globally, as medical care improves and becomes increasingly available, more children are surviving birth and the first few years of life. However, over 50 million children under 5 years have disabilities and the vast majority reside in low- and middle-income countries (LMICs)^1^. Young children are disproportionately affected by Neurodevelopmental Disabilities (NDD) in low resource settings due to environmental risks such as central nervous system infections occuring antenatally or postnatally, in-utero exposure to toxins, and traumatic incidents at or around the time of birth. Some of the most important and widespread NDD include; cerebral palsy, autism spectrum disorder, epilepsy and global developmental delay. The documented prevalence of these conditions has increased dramatically in recent years as more children survive the high-risk neonatal period, especially infections, birth asphyxia and prematurity ^2,3^. Many children with NDD remain undetected and hence untreated in resource-limited settings^4^. Reasons for this may include; community stigma, discrimination, limited training of professionals in identification of NDD at this young age, and limited access to health-care services for assessment or care once identified^5,6^. This is particularly due to the fact that the focus is often on saving lives with limited resources for the long term follow up of at-risk infants^7,8^.

Most currently available screening tools for NDD are proprietary, expensive, include items that might not be appropriate for children living in non-Western settings and are based on expected developmental milestone attainment for children living in high-resource Western settings^9–11^. Some tools created for global use to monitor child development require resource-intensive training^12,13^ and are designed to be conducted in primary care clinics by individuals with some expertise^14^. Most existing tools do not have specific items for identifying, sensory and hearing loss, congenital abnormalities or epilepsy. Children, therefore, have to rely on *ad hoc* assessments such as hearing screening^15^, vision screening^16^ and neonatal health checks^17^ when available.

One culturally appropriate developmental screening tool which has been designed for use in low resource settings, may have the potential to be adapted for identification of NDDs in these settings^18,19^. The Malawi Developmental Assessment Tool (MDAT)^20^, created and maintained by four of the authors, is used in many low-resource countries ^21–24^. The MDAT takes around 30 minutes to administer and is focused on child development. It does not directly measure other areas important for flagging developmental disabilities (e.g. hearing, vision, seizures, behaviour, hydrocephalus, spinal abnormality, etc.), or address relevant environmental circumstances, or the impact of the NDD on participation as embodied in the ICF-CY International Classification of Functioning and Disability for Children and Youth (ICF-CY) framework^25^.

There is a pressing need to create and validate a tool which is simple to administer, culturally relevant, freely available, and which can identify young children with neurodevelopmental disabilities in low-resource settings. In this study, we aimed to develop, pilot and field test a new tool, adapted from the MDAT and items on the ICF-CY, for the “Identification of Neurodevelopmental Disabilities to Improve Global Outcomes” (INDIGO), in young children (0-3 years) with a range of moderate to severe neurodevelopmental disabilities in preparation for a future large-scale validation study.

Our specific objectives were to:

1. Create a preliminary version of a new INDIGO tool to measure all aspects of functioning ascribed by the ICF-CY and develop a gold-standard measurement protocol of NDD for assessment of the INDIGO (Phase I).
2. Pilot and field test the feasibility of implementing the preliminary INDIGO tool and gold standard protocol in three low-resource countries, to determine the diagnostic accuracy of each item and create a final INDIGO Prototype I tool (Phase II).

## Methods

The study was conducted between March 2018 and December 2019. The study team comprised 14 researchers in psychology, paediatric neurodevelopment, paediatric neurology, general paediatrics, public health, education and biostatistics, from four countries; Uganda, Malawi, Pakistan and the UK as well as a wider team of 5 experts from India, Kenya, South Africa and Canada who provided advice on the protocol and item selection. These collaborations were formed through the Networking grant which also allowed for meetings held in the UK and Uganda as well as data collection activities in the three countries; Malawi, Pakistan and Uganda.

The study team held an initial meeting in the UK where we discussed the study protocol including; creation of the Preliminary INDIGO tool (Phase I) and the processes required for field testing and assessing diagnostic accuracy to create the Prototype INDIGO Version 1 tool (Phase II). This included making decisions on sampling processes, inclusion criteria and creation of a clinical gold (reference) standard assessment battery.

### PHASE 1: Creation of Preliminary INDIGO tool and a Gold Standard Assessment to test diagnostic accuracy

#### Creation of Preliminary INDIGO tool

During the initial networking meeting, a decision was made to utilise a recent systematic review conducted by one of the members of the network. This review which had pinpointed 18 tools used in low-and middle-income settings with the strongest evidence for screening young children with disabilities^26^. These 18 tools (see Supplementary File 1) contained relevant items and concepts which the study team could review to ensure comprehensive coverage of all areas of the ICF-CY in a single new tool.

**Coding process:** Following this meeting, we initiated a structured coding process starting with the MDAT and then identifying any gaps in its coverage of the ICF-CY framework. Initially, three expert assessors () all trained in neurodevelopmental paediatrics, coded all 144 MDAT items (36 Gross motor, 36 Fine Motor, 36 Language, 36 Social) onto the ICF-CY framework for specific age ranges (0-3m, 4-7m, 8-12m, 13-24m, 25m+) (Supplementary Figure 1). To facilitate this process, we used a coding spreadsheet that categorized items under 30 ICF-CY sub-categories distributed across four major domains (Body structure, Body Function, Activities and Participation, and Environmental Factors). Each MDAT item or concept was recorded in the spreadsheet and rated by the assessors.

We then identified ICF-CY sub-categories that were not covered by MDAT items across the designated age ranges. To address these gaps, six expert assessors () coded relevant items from the 18 tools identified in the systematic review^26^ onto the same ICF-CY spreadsheet in the same way as done with the MDAT items. This spreadsheet now includes both MDAT items and those from the additional 18 tools (Supplementary Table 1). The assessors, a diverse group of six paediatricians, one public health doctor, and one inclusive education specialist from Malawi, Uganda, Pakistan, Canada and the UK, selected items or concepts that addressed these gaps. The selected items included aspects such as child behaviour, sleep, feeding, body structure and environmental factors, ensuring that each met the following criteria: i) feasibility, ii) cross-cultural comparability, and iii) age relevance (0-3 years). As multiple items often covered the same concept, we conducted a consensus meeting to determine the most suitable items for ensuring that the INDIGO tool provided good coverage of the ICF-CY across all age groups.

Supplementary Figure 2 illustrates the domains of the ICF-CY (Body Structure, Body functions, Activities and Participation, Environmental Factors) in rows with subcategories in columns, along with the number of items identified from the different developmental screening tools in each ICF-CY domain. In total 2,302 items were coded onto the ICF-CY framework comprising 144 MDAT items and 2158 items from the 18 additional assessment tools. A high concentration of items was observed in the domains for Body Functions and Activities and Participation whereas fewer or no items were identified in the subcategories for Body Structure and Environmental Factors.

To address these gaps, we then identified 59 additional items. Twenty-eight items were selected from the 18 existing tools, and 31 entirely new items were developed and agreed by consensus particularly in the ICF-CY domains of Structure, Participation and Environmental factors. The final preliminary INDIGO tool comprised 203 items; 144 core MDAT items^20^ - 72 motor (fine and gross motor), 36 language and 36 socio-emotional items, and 59 additional items (Supplementary Table 2) to ensure full representation across all ICF-CY domains. This included 9 new items within the Body Structure domain of the ICF-CY (e.g., *“Does the child’s head seem abnormally small, large or unusually shaped?”, “Does the child have any unusual marks or sores or an unusual shape of their spine?”*). It also included 37 Functioning and Activity items including 28 taken from the 18 tools and 9 newly created ones (e.g., “Does your baby have trouble sucking from a bottle or breast?”, “Does the child play by pretending objects are something?”, “Do you have concerns about your child’s hearing?”). Finally, the tool included 13 new Participation and Environmental items addressing aspects such as social inclusion and access to play (e.g., *“Does the environment your child live in restrict your child’s potential to play outside the house?”* and *“Do you have any difficulties meeting up with your friends and family at social events because of your child?”).* These 59 additional items are detailed in Supplementary Table 2, where items sourced from the 18 existing tools are shaded and newly created items are left unshaded.

### Creation of Gold Standard

Prior to our first network meeting, each team gathered evidence on assessment tools which were well validated and had evidence of sensitivity and specificity for identifying children in the first five years of life through direct observation, which would identify moderate or severe NDDs according to our definition of NDD (global developmental delay, cerebral palsy, autism spectrum condition, epilepsy, hearing loss or vision impairment) for children. During the meeting, each NDD condition was discussed and consensus from the overall group was reached depending on the following factors; whether the tool had or could potentially be used in resource limited settings (cost), ease of use (cultural acceptability), training required to use it and time taken to conduct assessment. We created an assessment protocol after extensive consultation with team experts agreeing that the gold standard measure should include; i) detailed paediatric history, ii) a physical and neurological examination looking for any signs of neuromuscular, tonal changes, dysmorphic features and congenital anomalies, iii) Hammersmith Infant Neurological Exam^27^ (if under 24 months), iv) the Gross Motor Function Classification System Scoring^28^ (if greater than 24 months) and v) the RITA-T as an observational autism assessment (if over 24 months)^29^ (Figure 1). This battery of diagnostic tools served as an adjunct to clinical expertise providing quantifiable and objective data that enhanced diagnostic accuracy and reduced uncertainty by the senior clinical experts in their diagnosis and ensured that all teams were consistently using the same tools to make decisions on final diagnoses. The final diagnoses included; Autism Spectrum Disorder, Cerebral palsy, Spina Bifida, Hydrocephalus, Epilepsy, Global Developmental Delay, Hearing loss, vision impairment or other conditions diagnosed. The examining clinicians also assessed whether the child had a disability using a functional transdiagnostic approach using the functional domains of motor, cognition, hearing, vision, communication, emotion, behaviour or epilepsy.

**Figure 1:**
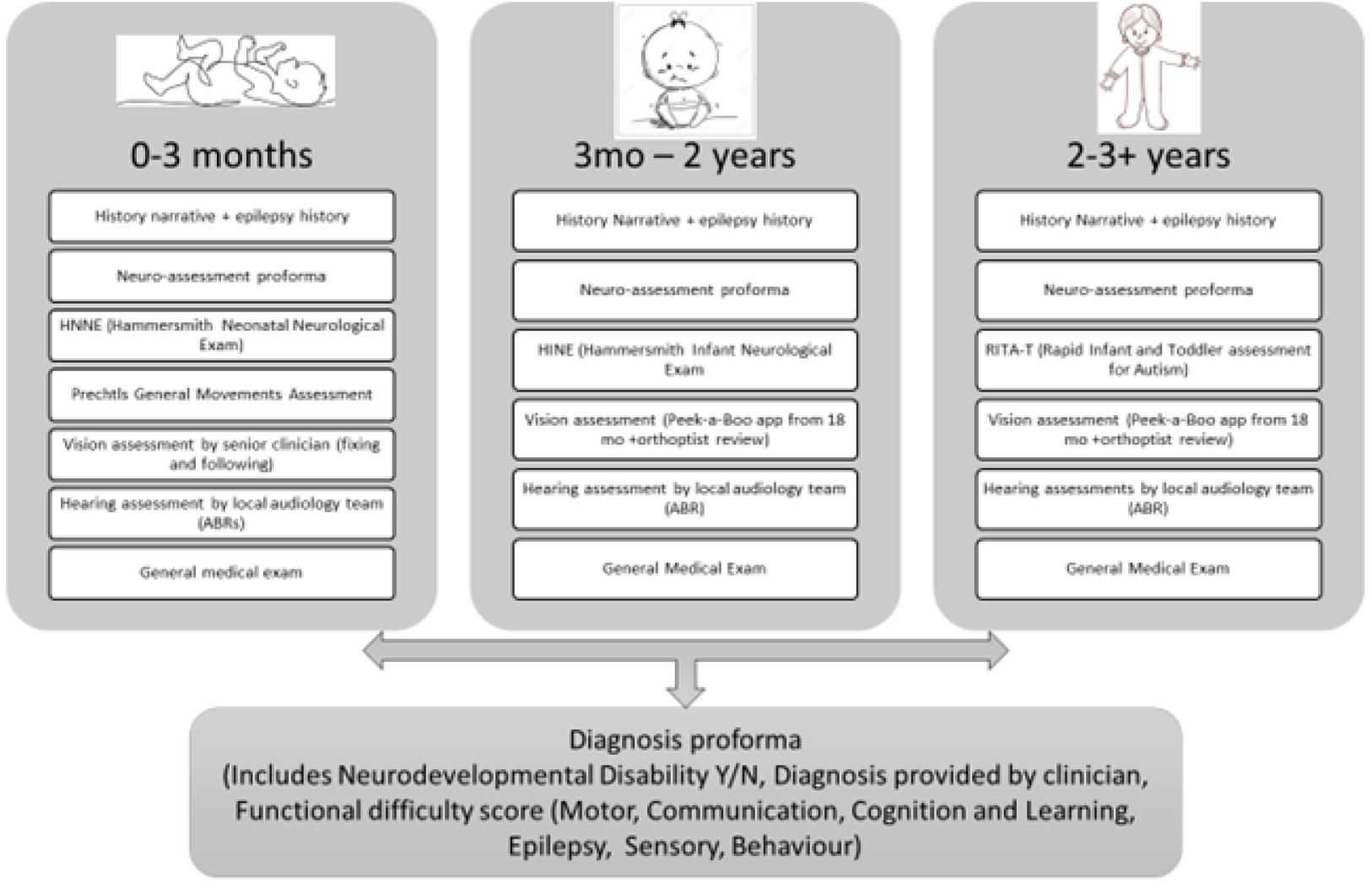
**Gold-standard Assessment for NDD including all tools chosen at consensus**

### Phase II. Implementation of Preliminary INDIGO tool alongside Gold (reference) Standard

In this section we describe the process for field testing and implementing the preliminary tool testing feasibility of implementation and gold standard assessment in Malawi, Pakistan and Uganda

We aimed to recruit 480 children overall (160 in total from each of the three countries). Each country sample comprised of 3 groups - 40 children recruited from a general community sample (Group A), 80 children coming from at-risk groups (Group B) i.e. children who were ‘at risk’ of having an NDD given environmental factors, and 80 children with a *known disability* (Group C) (Supplementary Table 3). These children were recruited using quota sampling across four age bands (0-3months, 4-12months, 13-24months, 25-36months). The sample size was based on pragmatic decisions in consultation with the country team leads, following CONSORT extension guidelines to pilot and feasibility studies^30,31^, and considering the personnel, time and funding available for data collection.

We included all parents/carers and their children from 0-3 years in the above three groups, where the parent was able to provide written informed consent, was resident within the catchment area of the clinical study site and who would continue to be resident for 12 months. Children who were unwell on the day of assessment were invited back for another visit or excluded if they did not return. When recruiting for Group A, any child who had chronic health needs e.g. those with a known NDD, HIV, malnutrition or recurrent infections, these were placed in Groups B or C.

### Recruitment

All potential participants received information, both verbally and in writing, in Chichewa (Malawi), Urdu (Pakistan), Luganda (Uganda) or English on the purpose and procedure of the study. Written consent was gained before commencing any data collection.

***Blantyre, Malawi***. Children and caregivers were recruited at Queen Elizabeth Central Hospital (QECH) in Blantyre. Children at risk (Group B) were recruited from wards and clinics (general outpatients, malnutrition unit, neonatal unit) and children with *known disabilities (Group C)* from neurology, physiotherapy, ophthalmology and audiology clinics. The *community sample (Group A)* was recruited through an existing community cohort of the CHAIN study where a list of participants was available for re-contacting^32^. Recruitment for at risk and known disability children occurred by research assistants doing a daily review of patients in wards and outpatient clinics and checking against inclusion criteria and checking with nurses in charge of respective areas to inquire when patients were to be discharged from the wards. Child assessments were done at QECH, Blantyre and for CHAIN study patients, in specific research clinics^33^.

***Karachi, Pakistan***. Children and caregivers were recruited from outpatient clinics at Aga Khan University Hospital (AKUH), Karachi including those attending neurology and developmental clinics. For the *community sample*, existing children were again identified as part of the CHAIN study and re-consented after information provision for this study^32^. Research assistants recruited patients in groups B (at risk) and C (known disability) by reviewing medical records of outpatients and checking against inclusion criteria. Assessments of groups B and C were done at AKUH whereas assessments of Group A (the *community sample)* were done at home.

***Kampala, Uganda***. Children were recruited in health units within Kampala district as well as at Mulago National Referral Hospital (MNRH). Study participants in the *at risk* and *known-disability* samples were recruited from paediatric inpatient wards, paediatric outpatient clinics (Physiotherapy, Occupational therapy, Audiology, Neurology Clinic and General Paediatric Clinics) and the in-patient paediatric nutrition unit. The *community sample* was recruited from typically developing children at immunization clinics in urban Kampala or from the CHAIN^33^ study cohort, serving the Kampala informal settlement (slum) community. Each day, research assistants would consult nurses in charge of respective inpatient wards and inquire which patients were to be discharged the following day. In addition, research assistants would visit outpatient clinics each day to screen for eligible children.

### Measures

Research assistants collected data on i) INDIGO ii) anthropometrics, iii) socioeconomic status and iv) home situation. They were blinded to the gold standard assessment (Figure 1) and vice-versa which was conducted by a senior clinician within two weeks of consent and INDIGO assessment. Data was entered directly into tablets using Open Data Kit (ODK)^34^ .

**INDIGO tool:** The INDIGO tool was created via methodology described in Phase I (Figure 2). It contained 203 items comprising of core MDAT items^20^ - 72 motor (fine and gross motor), 36 language and 36 socio-emotional items and the 59 additional items (Supplementary Table 2) to ensure that all domains of ICF-CY in structure, function, activities and participation, and environment were present.

**Figure 2:**
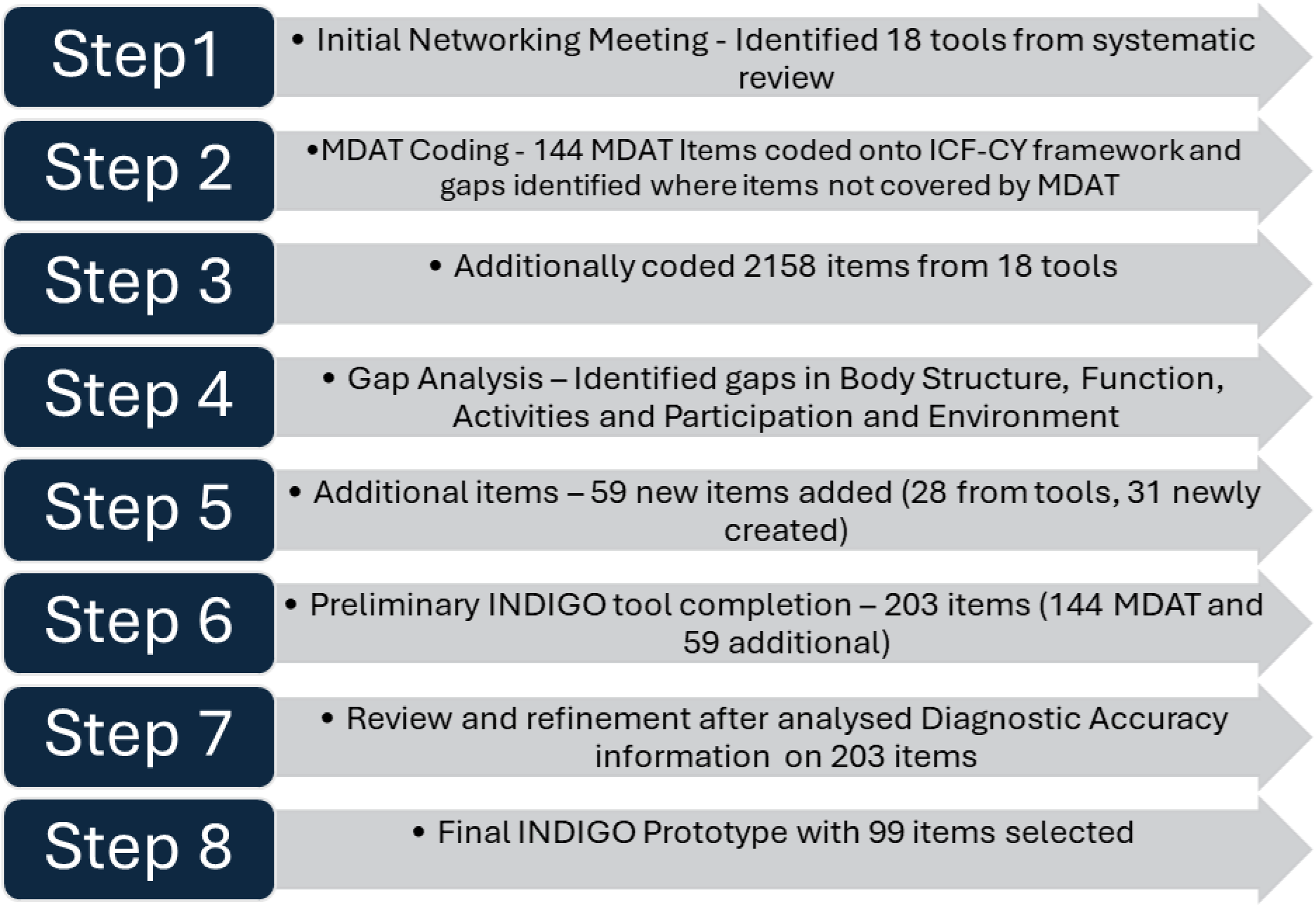
**Flow diagram showing process of creating INDIGO prototype**

**Figures 3a - 3f:**
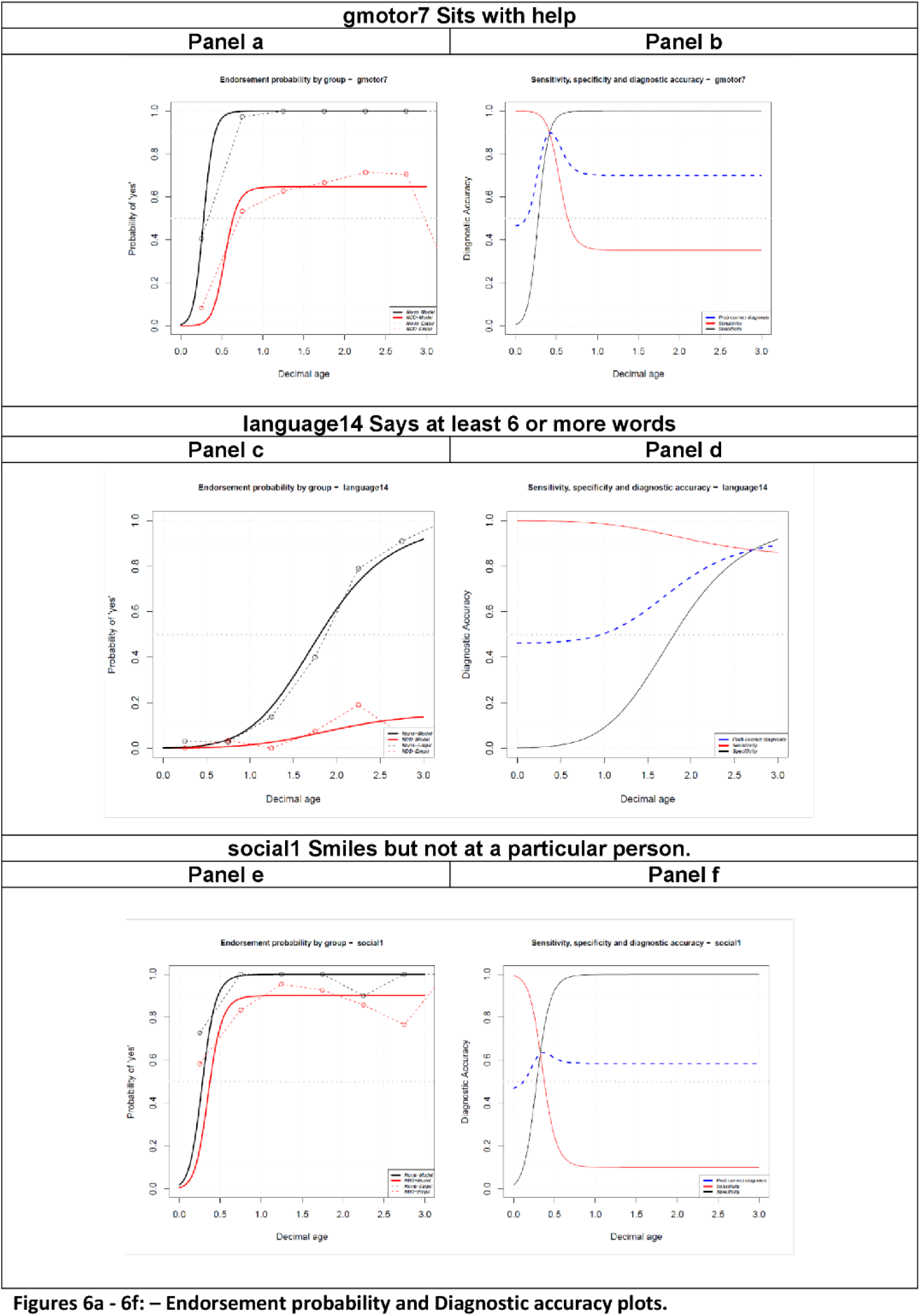
**Endorsement probability and Diagnostic accuracy plots.**

**Anthropometry:** Anthropometric measurements of every child in all sites (weight, height, MUAC and head circumference) were collected following standard WHO protocols^12^. These measures were used to create Length/Height-for-Age Z-score (LAZ/HAZ), Weight-for-Age Z-score (WAZ) and a Weight-for-Length/Height Z-score (WLZ/WHZ) where, in line with WHO recommendations, scores below -2 Z are concerning.

**Socioeconomic status and home stimulation**: Socioeconomic status (SES) variables included a wealth index and maternal education, which were assessed using Demographic & Health Survey (DHS) items standardized for each country^13^. Higher scores indicate a higher SES with a high number of assets. Scores have been converted and are reported on a to a z-score scale.

**Home situation:** Information on the stimulation and learning opportunities offered by the child’s home environment was collected with the Family Care Indicators (FCI)^14^. Higher scores indicate the child had a better number of opportunities for learning and stimulation in the home. Scores have been converted and are reported on a to a z-score scale.

**Gold Standard Assessment:** Full Gold Standard clinical assessments (Figure 1) were conducted in 1) Uganda at Mulago National Referral hospital, by study neurologist () and the country principal investigator (), 2) Malawi by study neurodevelopmental paediatrician () and 3) Pakistan by study neurologist () and psychologist (). Training was conducted across all sites to ensure consistency of measurement.

### Feasibility of implementation

The feasibility outcomes that we were interested in evaluating were: recruitment and consent rates of families, sampling processes across age bins and risk strata, training, fidelity and assessment of quality in using the Gold standard tools, acceptability of the assessments, adherence to returning for the Gold Standard after having completed the Preliminary version of the INDIGO tool, completion of all parts of the study^31^.

### Data analysis

The demographic and contextual data were summarized using mean and standard deviation (SD) for continuous data and count (percentage) for categorical data. The age of the child was computed as a decimal age. Missing item responses were not imputed, only complete case data was used.

### Assessing the diagnostic accuracy of the INDIGO at the item-level

Children with any diagnosis of disability (as confirmed through our Gold Standard assessment) were categorized into the ‘NDD’ group while those with no disabilities were categorized as ‘normally developing’ to obtain a ‘true’ diagnosis. The directly observed test data collected by the preliminary INDIGO tool was then modelled using a modified logistic regression to calculate (i) the probability of passing the item correctly (i.e., ‘yes’) by decimal age for the NDD and typically developing groups separately, and (ii) the probability of a correct diagnosis by decimal age for the sensitivity, specificity and diagnostic accuracy of each item in the tool. Empirical probabilities were also calculated and two graphs were created from the analysis for each item. The graphs allowed us to select the optimal items for the final INDIGO tool.

The logistic regression was modified in several ways to account for certain child characteristics. Firstly, since the diagnostic accuracy of a child development item is dependent on the age at which that item is asked, we needed to design a methodology to create *age dependent* diagnostic accuracy curves. Secondly, the logistic regression was constrained such that the probability of passing the item correctly was constrained to be close to zero at age zero, reflecting the fact that we know *a priori*, that a child who is only hours old cannot complete the item. Thirdly, the logistic curve in the NDD group was constrained so the endorsement curve would “plateau” rather than be assumed to be 1, to allow for those children who may never be able to successfully complete the item due to their disability. Bayes theorem was then used to convert the probability of passing the item correctly for both groups into a probability of making a correct diagnosis (i.e., the proportion of both true-positive and false negatives) denoted by the blue dotted line in the figures. It is important to note here that the diagnostic accuracy results presented are related to the prevalence of NDD in the sample. Thus, diagnostic accuracy should be interpreted in relative terms, i.e., the best items can be flagged, but their absolute levels of diagnostic accuracy may change dependent on the prevalence of NDD in the target population. More detail on this process is provided in Supplementary File 1.

### Selection of optimal items for final INDIGO Prototype I tool

We reviewed data on all 203 INDIGO items (144 MDAT and 59 additional items tested) and placed them back in the ICF-CY framework by age range and domain. All items with over 80% diagnostic accuracy in the age range of the children were included in a version which was then reviewed. Where multiple items were available for the same age, domain and child characteristic, we chose those items that were most feasible (needing the least materials to test a child), culturally neutral and easy to train on. A final list of 99 items that were included in the prototype INDIGO tool which was then tested is shown in Supplementary Table 4.

## Results: (Phase II)

We screened 831 participants (281 in Malawi, 350 in Pakistan and 200 in Uganda) and 479 (58%) who fulfilled the inclusion criteria were recruited into the study. Of those eligible, 404 (84%) were fully assessed with both INDIGO and the Gold-standard assessments. Those who were not fully assessed were those who had not returned for the second Gold-standard assessment (Supplementary Figure 3).

### Feasibility of implementation

***Recruitment and consent rates***: The lessons learned regarding the best ways to recruit across age groups in the community, to find those at-risk and those with NDDs are listed in Table 1. In terms of recruitment and consent of families (Figure 5), we had to approach almost twice as many families (831) as then consented and enrolled (479) to take part. The country teams found it much easier to recruit those with NDDs from a hospital environment due to the easy accessibility for the teams.

**Table 1.** Feasibility issues we identified within the study and potential solutions for future studies.

| Feasibility issue | Future solution |
| --- | --- |
| **Recruitment and consent rates** |  |
| Children with autism spectrum disorder and behavioural conditions were hard to identify<br>Many families place less value on a diagnosis where options for treatment are limited, and therefore some didn't consent as they felt there was little in the way of support and treatment for their children for these conditions.<br>While participants were recruited from the wards in the hospital and caregivers were willing to participate, the variability of the sample was restricted to those able to come to hospital and subject to bias | Conducting community engagement and liaison from earliest stages to ensure the community knows what you are looking for, may help to prevent parents from wanting to have their children assessed and identified.<br>Using a population-based sample and liaising with the key community stakeholders early in the study process to obtain the necessary support and encourage community engagement and family participation from varied populations.<br>Awareness talks in the hospitals/communities prior to study start to inform them of what is being studied and how they can be involved |
| **Sampling processes across age bins and strata** |  |
| Children with autism spectrum condition and behavioural conditions were hard to identify particularly at a young age.<br>Children with milder developmental and behavioural difficulties were hard to identify particularly at the youngest ages<br>Many families do not raise concerns and are therefore not easily identified in at-risk samples | Conducting a much larger population-based study would enable children across the spectrum to be identified OR using a larger "at risk" sample such as all of those discharged from a neonatal unit over 1-2 years of age<br>Using a larger population-based sample would also mean that children were not "singled out"<br>Employing multiple sample sources to identify "at risk" children, such as those discharged from neonatal intensive units, those attending premature clinics, CP clinics, genetics clinics, etc. |
| **Training** |  |
| Some tools were complicated and required knowledge of child development and working with children<br>Training for use of tools particularly in Gold Standard was extensive<br>Some retraining was required for staff who started in middle of study | In Pakistan, the team specifically recruited research assistants with experience in screening and managing children with developmental issues recognizing the importance of their expertise.<br>Consider hiring staff who can be retained for whole study |
| **Fidelity and assessment of quality using the Gold Standard tools** |  |
| ASD: The RITA-T was not a diagnostic tool and not useful for children < 2<br>Vision: The Peek-a-Boo app did not work well for children who had motor and cognitive difficulties and was not helpful for children less than 2 years. A tablet-based tool was also unfamiliar to many children and therefore difficult to use<br>Hearing: We did not have good portable systems for measuring ABR in the community and therefore could only use the systems employed at the central audiology departments in the hospital<br>We did not have a good tool for assessing behavioural difficulties in young children<br>Having a well trained neurodevelopmental paediatrician in each site gave us the ability to confirm clinically diagnoses however one was not always enough | Using a tool for use in resource limited settings that has better validity e.g. INCLEN ASD tool and the use of consensus diagnosis through video consultation across countries<br>Use of a definitive diagnosis through an orthoptist specially trained for all cases.<br>In future to use more portable systems that are available e.g. using mobile systems as conducted in India.<br>Further adaptation of tools to assess behaviour at the very youngest ages e.g. BITSEA<br>The need for a well-trained neurodevelopmental paediatrician in all sites where a study is taking place is vital. Training and supervising an additional medical doctor in the specific assessments to be carried out to minimize deferred appointments is essential. |
| **Acceptability of assessments** |  |
| Caregivers did not always want to come back for several appointments as they were busy.<br>Caregivers in general were satisfied with the assessments; however some children threw tantrums during the assessments and were difficult to placate | Assessments were integrated into the hospital visits in Pakistan and therefore not an issue<br>Provision of minimal breaks in between the different assessments, with some breaks spiced up with a snack, playing with a toy, etc |
| **Adherence to returning for Gold Standard** |  |
| A second assessment was needed for the Gold Standard which meant more time for the family<br>Due to changing priorities and circumstances, some caregivers did not keep the appointments | The team in Pakistan provided pick up and drop services and the reputation of AKUH played a key role in encouraging families to return.<br>Scheduling the Gold Standard assessments on the same day as the initial assessments or giving a shorter return date.<br>Having more than one well trained neurodevelopmental paediatrician at the site |
| **Completion of all assessments** |  |
| Most assessments were completed except for a few people who failed to come back for the gold standard. | Consider same day assessments |

***Sampling processes across age bins and strata***: We had some issues with sampling the youngest children particularly those with NDDs and those living in the community in Malawi, although we did manage to recruit enough to the at-risk group. In Pakistan, the level of recruitment in the at-risk group was low due to difficulties recruiting in the hospital, however levels of recruitment of those living in the community and those with NDDs were achieved. In Uganda recruitment was generally good with the number at risk only a little short of the target.

***Training***: Training for all Gold Standard tools took time and required a good level of knowledge to start with. Some needed to re-train and all staff needed to be reliably doing the same thing across sites. Training centrally at our networking meetings e.g. on RITA-T prior to starting enabled us to be conducting assessments in a similar way across sites.

***Fidelity and assessment of quality using the Gold Standard tools:*** We found that some tools did not perform well enough and would need replacing in a future study. Involvement of professionals was also important and was particularly the case with vision and hearing assessment where hiring an audiologist and an orthoptist at each site would have made things much easier in terms of following protocols and being confident with the results. Tools that require less expert input to provide a result are better but difficult to achieve.

***Acceptability of assessments*:** The country assessors reported that families were happy with the actual assessments however occasionally children got tired and difficult to manage during assessments.

***Adherence to returning for the Gold Standard*:** In Malawi 23/174 did not return for the Gold Standard assessments and in Pakistan 43/145 did not return with families often being less incentivized to return. Uganda had much better levels of completion with only 9/160 not completing.

***Completion of all assessments*:** In terms of completion rates of all assessments, we found that most children completed all assessments, however it was sometimes difficult to complete vision and hearing assessments.

Table 2 shows the numbers recruited by country and group that completed both the gold standard and INDIGO assessments and the demographics of the country samples. Children in all samples except the at-risk sample in Pakistan (N =5) were of average birthweight over 2.5 kg. There were slightly more males than females identified in the at risk and NDD samples. We had a good representation of mothers across different educational levels at all sites. Most mothers in all samples came from urban settings with more at risk or NDD children coming from rural settings in Malawi and Uganda. Missing data was very low and is shown in the last column of Table 2.

**Table 2.** Demographics of final sample who had INDIGO and Gold Standard Assessment.

|  | Malawi |  |  | Pakistan |  |  | Uganda |  |  | Total Mean(SD) | Missing (%) |
| --- | --- | --- | --- | --- | --- | --- | --- | --- | --- | --- | --- |
|  | Community<br>N=13 | At Risk<br>N=119 | NDD<br>N=19 | Community<br>N=47 | At Risk<br>N=5 | NDD<br>N=50 | Community<br>N=51 | At Risk<br>N=62 | NDD<br>N=38 |  |  |
| Total numbers of children assessed | N=151 |  |  | N=102 |  |  | N=151 |  |  | N=404 |  |
| Mean age | 1.25(0.79) | 0.92(0.96) | 1.57(0.75) | 1.25(0.89) | 1.26(0.93) | 1.49(0.86) | 1.16(0.80) | 1.37(0.84) | 1.56(0.76) |  | 13(3%) |
| Male (N) | 8 | 66 | 13 | 22 | 2 | 21 | 24 | 29 | 24 | 209 | 0(0%) |
| Female (N) | 5 | 53 | 6 | 25 | 3 | 29 | 27 | 33 | 14 | 195 | 0(0%) |
| Birthweight (kg) | 2.97(0.50) | 2.53(0.87) | 2.68(0.55) | 2.74(0.65) | 2.37(0.55) | 2.67(0.63) | 3.28(0.56) | 2.54(0.85) | 3.00(0.91) | 2.27(0.80) | 23(6%) |
| Maternal Education |  |  |  |  |  |  |  |  |  |  |  |
| None (N) | 0 | 4 | 0 | 9 | 0 | 3 | 4 | 7 | 6 | 33 | 11(3%) |
| Primary (N) | 3 | 41 | 7 | 2 | 3 | 3 | 11 | 22 | 11 | 103 |  |
| Started high school (N) | 4 | 33 | 4 | 12 | 0 | 6 | 16 | 15 | 7 | 97 |  |
| Completed High School (N) | 4 | 22 | 6 | 11 | 2 | 6 | 9 | 7 | 3 | 70 |  |
| Education beyond MSCE (N) | 1 | 10 | 1 | 12 | 0 | 32 | 3 | 5 | 5 | 69 |  |
| Completed MSCE/O'level (N) | 1 | 2 | 0 | 1 | 0 | 0 | 5 | 5 | 3 | 17 |  |
| Completed A'Levels (N) | 0 | 0 | 0 | 0 | 0 | 0 | 2 | 0 | 2 | 4 |  |
| Location of participants |  |  |  |  |  |  |  |  |  |  |  |
| Remote (N) | 0 | 0 | 0 | 2 | 0 | 1 | 1 | 0 | 0 | 4 | 9(2%) |
| Rural (N) | 1 | 36 | 4 | 0 | 0 | 2 | 9 | 15 | 10 | 77 |  |
| Urban (N) | 5 | 42 | 5 | 34 | 3 | 40 | 29 | 33 | 17 | 208 |  |
| Inner City (N) | 7 | 35 | 9 | 11 | 2 | 7 | 11 | 14 | 10 | 106 |  |
| Mean SES (DHS) (Z-score) | -0.12 (0.87) | -0.54 (0.85) | -0.61 (0.75) | 0.98 (0.70) | 0.48(0.20) | 1.16(0.60) | -0.06(0.54) | -0.29(0.59) | -0.29(0.60) | 0.00(1.00) | 20 (5%) |
| Mean FCI (Z-score) | 0.44(0.70) | 0.33(0.83) | -0.35(0.64) | 0.24(1.26) | -0.26(1.29) | -0.01(1.12) | -0.03(0.98) | -0.38(0.96) | -0.61(0.60) | 0.00(0.94) | 7(2%) |
| Mean Height for Age (Z-score) | -0.79 (1.75) | -1.24 (2.17) | 1.92 (2.24) | -0.38 (1.95) | 0.17 (0.91) | -1.26 (1.87) | -1.15 (2.20) | -2.31 (2.38) | -2.46 (1.96) | -1.39 (2.19) | 71(18%) |
| Weight for Age (Z-score) | -0.35 (1.30) | -1.06 (2.22) | -2.00 (2.04) | -0.92 (1.63) | -1.03 (0.75) | -1.85 (1.92) | -0.76 (1.36) | -2.35 (2.12) | -2.49 (1.85) | -1.46(2.03) | 26(6%) |
| Head circumference for age Z-score | 1.02 (1.55) | 0.17 (2.23) | -0.67 (1.83) | -0.99 (1.29) | -0.97 (1.03) | -1.70 (1.58) | -0.15 (1.50) | -0.84 (1.73) | -1.39 (2.21) | -0.52(1.98) | 55(14%) |

Footnote: Socioeconomic status measured using the Demographic Health Survey tools for each country and provided with a Z score, Home stimulation measured using Family Care Indicators using a Z score, Height for age, Weight for age and Head circumference for age Z score were all created using Anthrostat software.

### Diagnostic accuracy of the INDIGO items

The proportion of children that correctly passed an item, in both the NDD and normally developing groups, was modelled by decimal age. Graphs were drawn up for each item to aid in the item selection process. Examples of three different items are given in Figures 6a - 6f. In these graphs, an item which tends to be answered positively by the typically developing group and negatively by the NDD group will be more useful in discriminating between the two groups. For example, in Figure 6a we can see a big difference between the two groups in terms of the probability of a correct response at around 6 months. If we look at the corresponding diagnostic accuracy plot, Figure 6b, we can see that the probability of a correct diagnosis for that item is highest at around this age. This is indicative of the age at which the item is performing best. Contrast this with Figures 6c and 6d, where the item does not discriminate (i.e., chance [∼50%] level of correct diagnosis) until around 24 months where the gap between the normally developing and NDD children, in terms of developing language, becomes apparent. Figure 6e and 6f show an example of an item which is only marginally less likely to be answered correctly by the typically developing group than the NDD group. This leads to an item that offers poor diagnostic utility right across the age range.

The graphs for all the items are shown in Supplementary Figure 1 and more detailed methodology information in Supplementary File 1.

### Creation of the INDIGO prototype Version I

Through our testing of diagnostic accuracy, we identified items from the preliminary INDIGO tool which had excellent diagnostic accuracy for identifying children who had a definitive neurodevelopmental disorder (Supplementary Table 5 shows diagnostic accuracy per age group for all items in INDIGO). Using a criterion of having a diagnostic accuracy of greater than 80% for the age group, we chose items which had as high diagnostic accuracy for the age as possible (e.g. if 90% we chose that one), had good feasibility and used few items, were as culturally neutral as possible and covered as many areas of the ICF-CY as possible. As no items in MDAT looked at structure of the child’s body – we included ten new items created through consensus into the final tool (highlighted in bold and italics in Supplementary Table 2). Similarly, there were no existing items that focused on sensory functioning or epilepsy – so one item from our INDIGO analysis was included in the final tool. Two or three items were chosen from the MDAT within each age group for Gross Motor and Fine Motor Function however, for cognition and language, there were very few items with >80% diagnostic accuracy in the first 12 to 18 months of life. Between 18 months and 3 years of life, more items had diagnostic accuracy. We included items which were not in MDAT which covered areas of ICF-CY which we had identified on other tools. These included six extra “play” or “socioemotional” items (e.g. “does your child like to be around other children”, “does your child pretend objects are something else” or “does your child engage in pretend play” which provided diagnostic power (Supplementary Table 4).

The final prototype tool is given in Supplementary Table 4. The tool is divided into 10 age groups for ease of use with children in the community with more age groups at the lower ages when children’s development changes more. The prototype then has items within the ICF-CY categories of structure (ten items), function (sensory and mental functions) and activities (Mobility – gross and fine motor, Cognition, Communication (producing and receiving language), Major Life Areas (play) and Self-care and interpersonal skills. We had lots of items in mobility and communication but none for cognition below 18 months as this is hard to measure behaviourally in children at this age.

## Discussion

In this study we have created the INDIGO tool, using a novel approach for identifying moderate to severe NDDs in children aged 0-3 years in resource-limited settings. The INDIGO tool addresses significant gaps in existing developmental screening tools by incorporating items that assess body structure (congenital abnormalities, head size, spina bifida), functioning (vision, hearing, epilepsy, development) and activities and participation (developmental items). Unlike tools primarily designed for high-resource settings, INDIGO is culturally adapted, feasible and tailored for use in LMICs, potentially bridging critical gaps in early identification and intervention for NDDs.

Through a detailed and rigorous coding process, we have identified missing components in existing tools such as the Malawi Developmental Assessment Tool (MDAT). Items addressing structural anomalies (e.g. head size, spine abnormalities), sensory impairments or loss (vision and hearing, general health, feeding and sleeping) and activities and participation (play, socioemotional engagement) were added to ensure comprehensive coverage of the ICF-CY framework. These factors can significantly impact a child’s abilities, including the availability of family, community, and school support, financial resources, family dynamics, mental health, and access to necessary equipment or adaptations. A holistic approach will enhance a tool’s relevance in diverse LMIC settings where children face unique developmental challenges heavily influenced by socioenvironmental factors.

A key challenge for the study was establishing a gold-standard assessment in three LMIC countries for validating the INDIGO tool. Diagnostic consistency was ensured through a multidisciplinary approach involving highly trained neurodevelopmental paediatricians (ES, HK, AKM, MR, PC) alongside the use of standardized tools feasible for use in young ages for single assessments. This included the Hammersmith Infant Neurological Exam and a standard vision^35^ and hearing examination done through referral to professional services. Limitations were noted with some tools – particularly the RITA-T with issues regarding cultural relevance (the item with the “magic trick”) as well as assessing vision and hearing in very young children. The Peekaboo app^36^ (measures grating visual acuity) for young children was piloted however it was difficult to use in young children with motor and severe cognitive difficulties as it required the child to move their head to look towards the grating or to point at the appropriate grating. We relied on formal tone audiometry for hearing assessment and vision assessment through formalized local services optometry/paediatric ophthalmologists, audiologists and orthoptists. These challenges underscore the need for more portable, culturally validated diagnostic tools particularly for behavioural and sensory impairments. Since this study commenced, some newer tools have been developed and validated, such as the INCLEN tools which now have a version for ASD diagnostic assessment for young children and which could be helpful although have not been validated outside an Indian setting as yet^37^. In addition, new hearing screening processes recommended in community settings by teams in India, may be helpful for future studies^38^.

We demonstrated through an original and rigorous study, that many of the MDAT items retained excellent diagnostic accuracy or utility, whilst some additional items addressing socioemotional or play behaviours showed promise (e.g. “does your child like to be around other children”, “does your child pretend objects are something else” or “does your child engage in pretend play”) with good discriminative validity for moderate to severe NDD. However, subtle and age independent items like those assessing some socio-emotional areas, sleep or feeding did not perform as well and may require larger sample sizes to validate their diagnostic utility fully.

Despite achieving an 84% recruitment rate (404/480 children), the study faced difficulties in identifying enough very young children with mild behavioural, socioemotional and social communication difficulties. Future studies should consider larger population-based cohorts or expanded at-risk samples, such as neonates discharged from intensive care units, to improve the detection of diverse developmental profiles to fully understand the diagnostic accuracy of items.

Our findings also emphasize the importance of community engagement to address parental reluctance stemming from limited treatment options. Future efforts should focus on sensitizing communities about the value of early identification and support for children with NDDs before a study begins. Our recent related qualitative study made it clear that there was often a lack of skills among health care providers to support the initiative for early identification of children with NDD and to refer to appropriate services and that often these are not integrated^39^.

This study and our related published qualitative study provide vital evidence to inform the development of scalable, culturally relevant tools for NDD identification across these three LMICs. This study demonstrates that we can conduct diagnostic accuracy studies for NDDs across LMICs particularly through creating larger networks of expertise to generate context-relevant data for optimal NDD assessment of children <3 years. However, further work is needed to refine the INDIGO prototype tool, including integrating newer validated measures and portable community-based screening methods for sensory impairments. Additionally, future studies should have greater focus on implementation and process evaluation of training and utilization of a tool such as this in national child health surveillance systems.

## Conclusions

This study contributes new evidence to the field by taking a careful methodological approach to develop a set of items which have been tested to create a prototype version of a short tool to identify children <3 years with NDDs in community settings across two sub-Saharan African and one South Asian resource-limited countries. We demonstrate clear processes for conducting such studies and how to gain information on the diagnostic accuracy of items that may identify children with NDDs using a detailed protocol for gold-standard assessment which can be refined depending on the expertise available.

## Supporting information

Supplementary Figure 4 All graphs

## Data Availability

All data produced in the present study are available upon reasonable request to the authors

## Acknowledgements

We would like to thank all the parents, guardians and professionals who shared their valuable time and insights with us.

## PPI Statement

Direct patient or public involvement in the design and conduct of this study was limited however, this work was explicitly informed by a complementary qualitative study (Lynch et al., 2023) led by members of the same research team, including the senior author (M. Gladstone). That study explored the perspectives of caregivers and frontline health workers in Malawi, Uganda, and Pakistan regarding early identification of neurodevelopmental disabilities (NDDs), stigma, feasibility of assessment, and barriers to care. Findings from that study will directly influenced further implementation of the INDIGO tool. Results from both studies have been disseminated back to stakeholders in Malawi and Pakistan.

## Ethical Approval

The study was approved by the Research and Ethics Committee at the Liverpool School of Tropical Medicine, UK (18-001), College of Medicine, University of Malawi (P.06/18/2424) Makerere University, Uganda (2018-147) and the Aga Khan University, Pakistan (2018-0271-377).

## Supplementary Files

**Supplementary Table 1.** Names of tools reviewed for items which might have concepts relating to the ICF-CY which were not in MDAT (for coding)

|  |  |
| --- | --- |
| 23 Question Screen | Kakooza-Mwesige A, Ssebyala K, Karamagi C, et al. Adaptation of the "ten questions" to screen for autism and other neurodevelopmental disorders in Uganda. *Autism* 2014; **18**(4): 447-57. |
| Ages and Stages | Squires J, Bricker D. Ages & Stages Questionnaires®Third Edition (ASQ®-3) A Parent-Completed Child Monitoring System. Baltimore: Paul H. Brookes Publishing Co., Inc; 2009 |
| Ages and Stages Socio-emotional questionnaire | Squires J, Bricker D, Twombly E. Ages & Stages Questionnaires®: Social-Emotional, (ASQ®:SE-2) Second Edition A Parent-Completed Child Monitoring System for Social-Emotional Behaviours. Baltimore,; Paul H. Brookes Publishing Co., Inc.; 2015 |
| Child Behaviour Checklist | Achenbach TM, Rescorla LA. Manual for the ASEBA School-Age Forms & Profiles. Burlington, VT: University of Vermont, Research Center for Children, Youth, & Families; 2001. |
| CDC (Centre for Disease Control) Milestones | Centres for Disease Control. Child Development, Toddlers (1-2 years old) 2017. https://www.cdc.gov/ncbddd/childdevelopment/positiveparenting/toddlers.html (accessed 24th March, 2024). |
| CREDI | McCoy DC, Sudfeld CR, Bellinger DC, et al. Development and validation of an early childhood development scale for use in low-resourced settings. *Popul Health Metr* 2017; **15**(1): 3. |
| DMC (Developmental Milestones Checklist) | Prado EL, Abubakar AA, Abbeduto S, Jimenez EY, Some JW, Ouedraogo JB. Extending the Developmental Milestones Checklist for use in a different context in Sub-Saharan Africa. *Acta Paediatr* 2014; **103**(4): 447-54. |
| DSQ (Developmental Screening Questionnaire) | Khan NZ, Muslima H, Begum D, et al. Validation of rapid neurodevelopmental assessment instrument for under-two-year-old children in Bangladesh. *Paediatrics* 2010; **125**(4): e755-62. |
| IYCD (Infant and Young Child Developmental Indicators) | Gladstone M, Lancaster G, McCray G, et al. Validation of the Infant and Young Child Development (IYCD) Indicators in Three Countries: Brazil, Malawi and Pakistan. *Int J Environ Res Public Health* 2021; **18**(11). |
| Kilifi Developmental Inventory | Abubakar A, Holding P, van Baar A, Newton CR, van de Vijver FJ. Monitoring psychomotor development in a resource-limited setting: an evaluation of the Kilifi Developmental Inventory. *Ann Trop Paediatr* 2008; **28**(3): 217-26 |
| NDST (Neurodevelopmental Screening Tool) | Silberberg D, Arora N, Bhutani V, et al. Neuro-Developmental Disorders in India - From Epidemiology to Public Policy (I10-1.006). *Neurology* 2014; **82**(10 Supplement): I10-1.006. |
| RBSK | Kar A, Radhakrishnan B, Girase T, Ujagare D, Patil A. Community-Based Screening and Early Intervention for Birth Defects and Developmental Disabilities: Lessons from the RBSK Programme in India. *Disability, CBR & Inclusive Development* 2020; **31**(1): 30. |
| RITA-T | Lemay JF, Amin P, Langenberger S, McLeod S. Experience with the Rapid Interactive Test for Autism in Toddlers in an Autism Spectrum Disorder Diagnostic Clinic. *J Dev Behav Pediatr* 2020; **41**(2): 95-103. |
| RNDA (Rapid Neurodevelopmental Assessment) | Khan NZ, Muslima H, Shilpi AB, et al. Validation of rapid neurodevelopmental assessment for 2- to 5-year-old children in Bangladesh. *Paediatrics* 2013; **131**(2): e486-94. |
| SDQ (Strengths and Difficulties Questionnaire) | Goodman R. The Strengths and Difficulties Questionnaire: A Research Note. *Journal of Child Psychology and Psychiatry* 1997; **38**(5): 581-6. |
| SWYC (Survey of Well-being of Young Children) | Perrin E, Sheldrick C, Visco Z, Mattern K. The Survey of Well-being of Young Children (SWYC) User's Manual Version 1.01. 2016. https://www.tuftschildrenshospital.org/-/media/Brochures/Floating-Hospital/SWYC/SWYC-Manual-v101-Web-Format-33016.ashx?la=en&hash=E0C2802F003ED312E9D5268374C540A112151FB3 |
| Ten Question Questionnaire | Zaman SS, Khan NZ, Islam S, et al. Validity of the 'Ten Questions' for screening serious childhood disability: results from urban Bangladesh. *Int J Epidemiol* 1990; **19**(3): 613-20 |
| Washington Questions | WG Child Disability Workgroup. Module on Child Functioning and Disability. Washington Centre for Disease Control 2014. |

**Supplementary Table 2.**
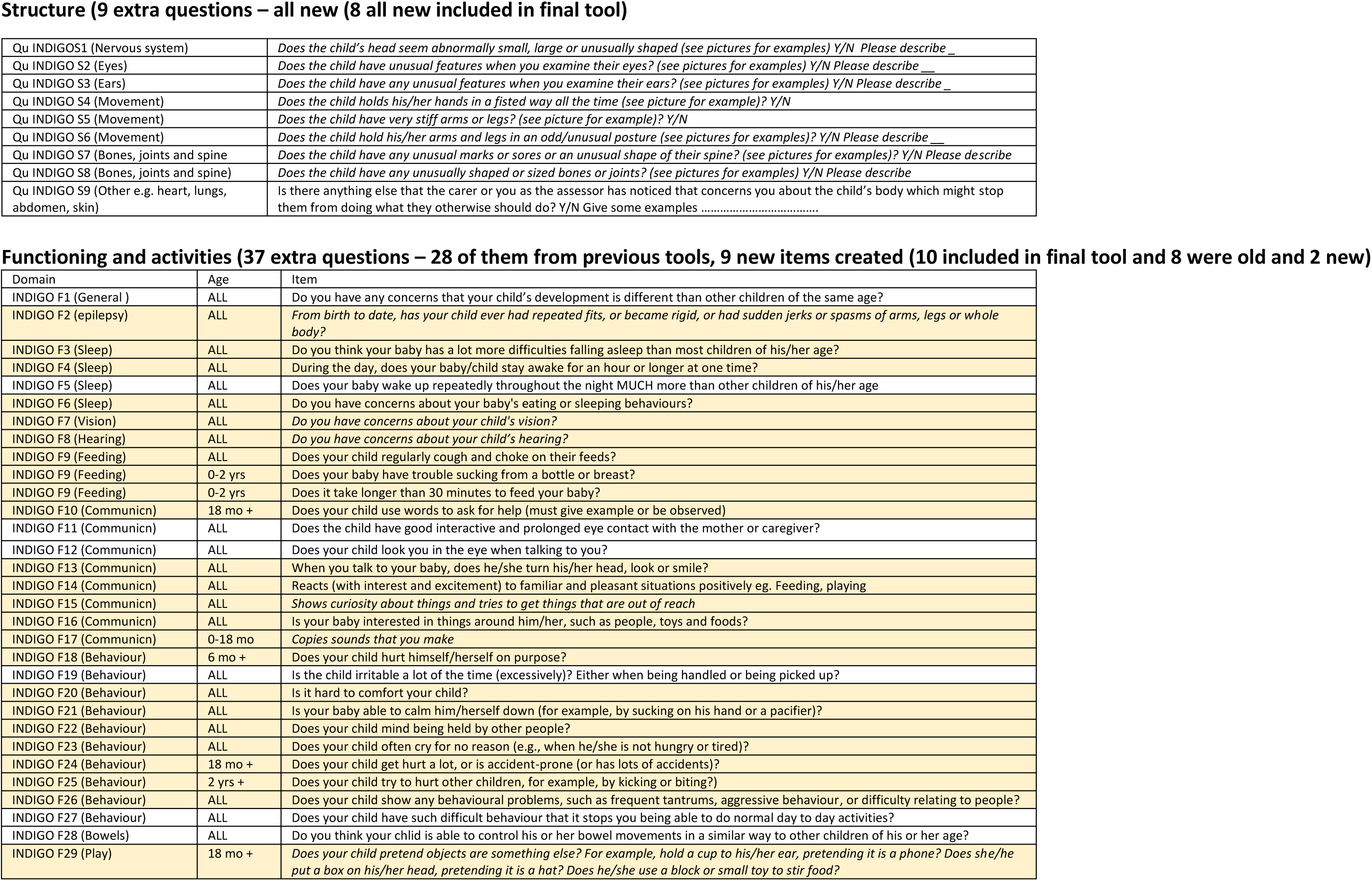
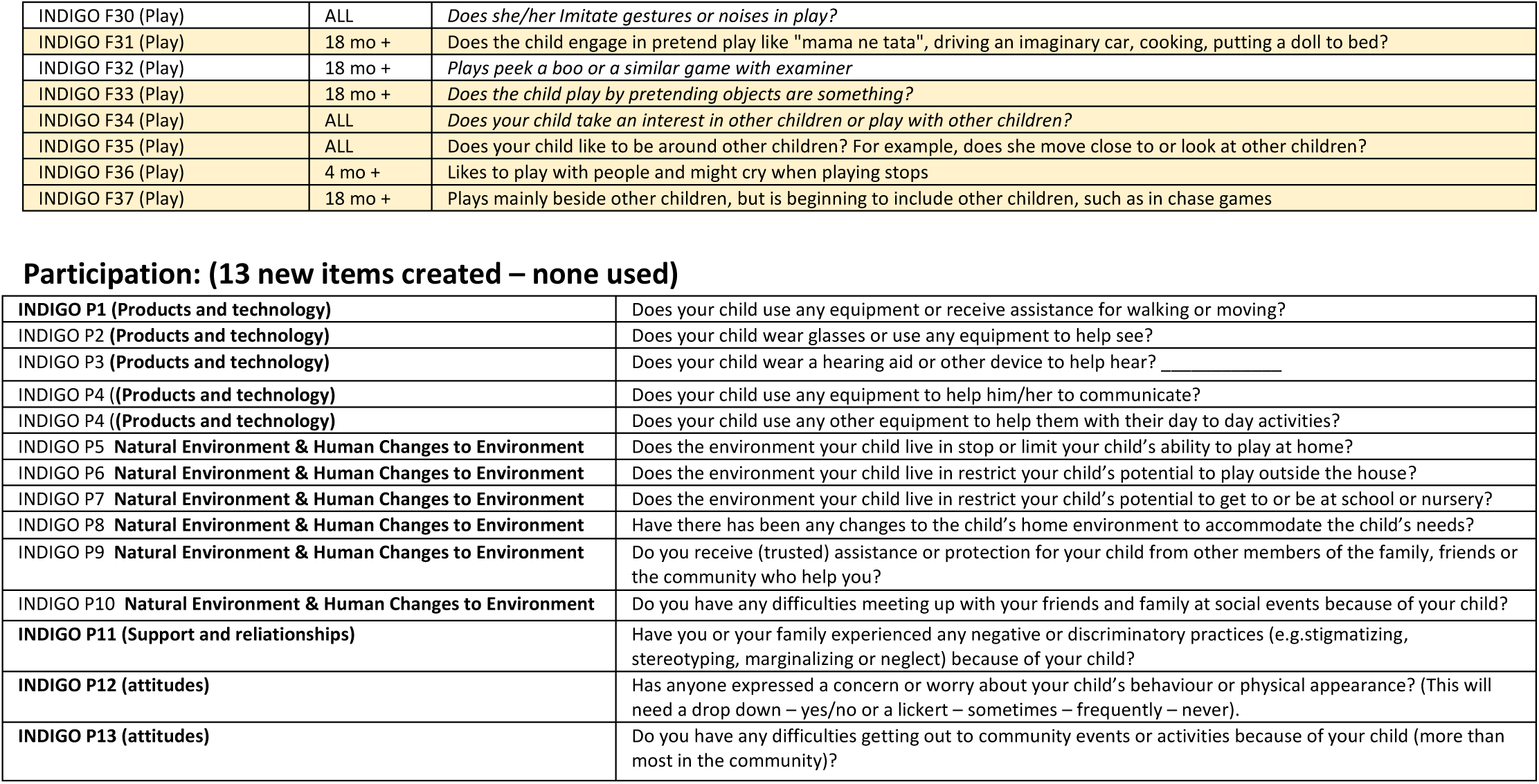
Additional items included in INDIGO tool for testing.

Structure (9 extra questions – all new (8 all new included in final tool))
|  |  |
| --- | --- |
| Qu INDIGO S1 (Nervous system) | *Does the child's head seem abnormally small, large or unusually shaped (see pictures for examples) Y/N Please describe _* |
| Qu INDIGO S2 (Eyes) | *Does the child have unusual features when you examine their eyes? (see pictures for examples) Y/N Please describe _* |
| Qu INDIGO S3 (Ears) | *Does the child have any unusual features when you examine their ears? (see pictures for examples) Y/N Please describe _* |
| Qu INDIGO S4 (Movement) | *Does the child holds his/her hands in a fisted way all the time (see picture for example)? Y/N* |
| Qu INDIGO S5 (Movement) | *Does the child have very stiff arms or legs? (see picture for example)? Y/N* |
| Qu INDIGO S6 (Movement) | *Does the child hold his/her arms and legs in an odd/unusual posture (see pictures for examples)? Y/N Please describe _* |
| Qu INDIGO S7 (Bones, joints and spine) | *Does the child have any unusual marks or sores or an unusual shape of their spine? (see pictures for examples)? Y/N Please describe* |
| Qu INDIGO S8 (Bones, joints and spine) | *Does the child have any unusually shaped or sized bones or joints? (see pictures for examples) Y/N Please describe* |
| Qu INDIGO S9 (Other e.g. heart, lungs, abdomen, skin) | Is there anything else that the carer or you as the assessor has noticed that concerns you about the child's body which might stop them from doing what they otherwise should do? Y/N Give some examples ..... |

Functioning and activities (37 extra questions – 28 of them from previous tools, 9 new items created (10 included in final tool and 8 were old and 2 new))
| Domain | Age | Item |
| --- | --- | --- |
| INDIGO F1 (General ) | ALL | Do you have any concerns that your child's development is different than other children of the same age? |
| INDIGO F2 (epilepsy) | ALL | *From birth to date, has your child ever had repeated fits, or became rigid, or had sudden jerks or spasms of arms, legs or whole body?* |
| INDIGO F3 (Sleep) | ALL | Do you think your baby has a lot more difficulties falling asleep than most children of his/her age? |
| INDIGO F4 (Sleep) | ALL | During the day, does your baby/child stay awake for an hour or longer at one time? |
| INDIGO F5 (Sleep) | ALL | Does your baby wake up repeatedly throughout the night MUCH more than other children of his/her age |
| INDIGO F6 (Sleep) | ALL | Do you have concerns about your baby's eating or sleeping behaviours? |
| INDIGO F7 (Vision) | ALL | *Do you have concerns about your child's vision?* |
| INDIGO F8 (Hearing) | ALL | *Do you have concerns about your child's hearing?* |
| INDIGO F9 (Feeding) | ALL | Does your child regularly cough and choke on their feeds? |
| INDIGO F9 (Feeding) | 0-2 yrs | Does your baby have trouble sucking from a bottle or breast? |
| INDIGO F9 (Feeding) | 0-2 yrs | Does it take longer than 30 minutes to feed your baby? |
| INDIGO F10 (Communicn) | 18 mo + | Does your child use words to ask for help (must give example or be observed) |
| INDIGO F11 (Communicn) | ALL | Does the child have good interactive and prolonged eye contact with the mother or caregiver? |
| INDIGO F12 (Communicn) | ALL | Does your child look you in the eye when talking to you? |
| INDIGO F13 (Communicn) | ALL | When you talk to your baby, does he/she turn his/her head, look or smile? |
| INDIGO F14 (Communicn) | ALL | Reacts (with interest and excitement) to familiar and pleasant situations positively eg. Feeding, playing |
| INDIGO F15 (Communicn) | ALL | *Shows curiosity about things and tries to get things that are out of reach* |
| INDIGO F16 (Communicn) | ALL | Is your baby interested in things around him/her, such as people, toys and foods? |
| INDIGO F17 (Communicn) | 0-18 mo | *Copies sounds that you make* |
| INDIGO F18 (Behaviour) | 6 mo + | Does your child hurt himself/herself on purpose? |
| INDIGO F19 (Behaviour) | ALL | Is the child irritable a lot of the time (excessively)? Either when being handled or being picked up? |
| INDIGO F20 (Behaviour) | ALL | Is it hard to comfort your child? |
| INDIGO F21 (Behaviour) | ALL | Is your baby able to calm him/herself down (for example, by sucking on his hand or a pacifier)? |
| INDIGO F22 (Behaviour) | ALL | Does your child mind being held by other people? |
| INDIGO F23 (Behaviour) | ALL | Does your child often cry for no reason (e.g., when he/she is not hungry or tired)? |
| INDIGO F24 (Behaviour) | 18 mo + | Does your child get hurt a lot, or is accident-prone (or has lots of accidents)? |
| INDIGO F25 (Behaviour) | 2 yrs + | Does your child try to hurt other children, for example, by kicking or biting?) |
| INDIGO F26 (Behaviour) | ALL | Does your child show any behavioural problems, such as frequent tantrums, aggressive behaviour, or difficulty relating to people? |
| INDIGO F27 (Behaviour) | ALL | Does your child have such difficult behaviour that it stops you being able to do normal day to day activities? |
| INDIGO F28 (Bowels) | ALL | Do you think your child is able to control his or her bowel movements in a similar way to other children of his or her age? |
| INDIGO F29 (Play) | 18 mo + | *Does your child pretend objects are something else? For example, hold a cup to his/her ear, pretending it is a phone? Does she/he put a box on his/her head, pretending it is a hat? Does he/she use a block or small toy to stir food?* |
| INDIGO F30 (Play) | ALL | *Does she/her Imitate gestures or noises in play?* |
| INDIGO F31 (Play) | 18 mo + | Does the child engage in pretend play like "mama ne tata", driving an imaginary car, cooking, putting a doll to bed? |
| INDIGO F32 (Play) | 18 mo + | *Plays peek a boo or a similar game with examiner* |
| INDIGO F33 (Play) | 18 mo + | *Does the child play by pretending objects are something?* |
| INDIGO F34 (Play) | ALL | *Does your child take an interest in other children or play with other children?* |
| INDIGO F35 (Play) | ALL | Does your child like to be around other children? For example, does she move close to or look at other children? |
| INDIGO F36 (Play) | 4 mo + | Likes to play with people and might cry when playing stops |
| INDIGO F37 (Play) | 18 mo + | Plays mainly beside other children, but is beginning to include other children, such as in chase games |

|  |  |
| --- | --- |
| **INDIGO P1 (Products and technology)** | Does your child use any equipment or receive assistance for walking or moving? |
| **INDIGO P2 (Products and technology)** | Does your child wear glasses or use any equipment to help see? |
| **INDIGO P3 (Products and technology)** | Does your child wear a hearing aid or other device to help hear? _____ |
| **INDIGO P4 ((Products and technology)** | Does your child use any equipment to help him/her to communicate? |
| **INDIGO P4 ((Products and technology)** | Does your child use any other equipment to help them with their day to day activities? |
| **INDIGO P5 Natural Environment & Human Changes to Environment** | Does the environment your child live in stop or limit your child's ability to play at home? |
| **INDIGO P6 Natural Environment & Human Changes to Environment** | Does the environment your child live in restrict your child's potential to play outside the house? |
| **INDIGO P7 Natural Environment & Human Changes to Environment** | Does the environment your child live in restrict your child's potential to get to or be at school or nursery? |
| **INDIGO P8 Natural Environment & Human Changes to Environment** | Have there has been any changes to the child's home environment to accommodate the child's needs? |
| **INDIGO P9 Natural Environment & Human Changes to Environment** | Do you receive (trusted) assistance or protection for your child from other members of the family, friends or the community who help you? |
| **INDIGO P10 Natural Environment & Human Changes to Environment** | Do you have any difficulties meeting up with your friends and family at social events because of your child? |
| **INDIGO P11 (Support and relationships)** | Have you or your family experienced any negative or discriminatory practices (e.g.stigmatizing, stereotyping, marginalizing or neglect) because of your child? |
| **INDIGO P12 (attitudes)** | Has anyone expressed a concern or worry about your child's behaviour or physical appearance? (This will need a drop down – yes/no or a lickert – sometimes – frequently – never). |
| **INDIGO P13 (attitudes)** | Do you have any difficulties getting out to community events or activities because of your child (more than most in the community)? |

**Supplementary Table 3.** Sampling characteristics of 3 groups for INDIGO study.

| **Group** | **Recruitment Criteria** |
| --- | --- |
| **A) Community sample** | -Children who have no risk factors or diagnoses in group c and no exclusion factors |
| **B) At risk** | -Signs of congenital deformities or syndromic features<br>-Episode of hypoxic ischaemic encephalopathy at birth<br>-Any child born as low birthweight (<1800g) or premature (<37 weeks)<br>-Concerns about vision by carer<br>-Concerns about hearing by the carer<br>-Signs of likely in-utero infection (congenital syphilis, congenital CMV)<br>-Abnormal looking spine<br>-Large head (macrocephaly)<br>-More than 2 seizures not associated with a fever<br>-Malnutrition (moderate or severe)<br>-Documented recent episode of head trauma |
| **C) Known disability** | -Cerebral palsy<br>-Severe neuromuscular conditions (including arthrogryposis), spina bifida and hydrocephalus.<br>-Severe global developmental delay (->2SD in all areas of a developmental assessment)<br>-Hearing loss (deafness) (moderate – profound) (41 dB – 81 dB+)<br>-Visual impairment (moderate to severe) (VA <3 – 6/60 in better eye when best corrected)<br>-Diagnosed epilepsy according to the revised ILAE definition<sup>40</sup><br>-ASD (DSM V) requiring substantial support as in DSM V definition<br>-Severe behavioural problems (pervasive and in all situations) |

**Supplementary Table 4.** Final items chosen for INDIGO Prototype Version 1.

|  |  |  |  |  |  |  |  |  |  |  |  |
| --- | --- | --- | --- | --- | --- | --- | --- | --- | --- | --- | --- |
| STRUCTURE |  | Do you think the child's head looks extremely small or extremely big (if possible measure it) |  |  |  |  |  |  |  |  |  |
|  |  | Do you think the child's eyes or ears look abnormal? |  |  |  |  |  |  |  |  |  |
|  |  | Does the child have any abnormal features on their spine or back? |  |  |  |  |  |  |  |  |  |
|  |  | Do you have any concerns that the child has a problem with their palate or lip? |  |  |  |  |  |  |  |  |  |
|  |  | Does the child hold their legs in a stiff position? |  |  |  |  |  |  |  |  |  |
|  |  | Does the child have any very abnormal joints or limbs? |  |  |  |  |  |  |  |  |  |
|  |  | Anthropometry |  |  |  |  |  |  |  |  |  |
| FUNCTION | Sensory functions | Do you think the child has any difficulty hearing? |  |  |  |  |  |  |  |  |  |
|  |  | Do you think the child has any difficulty seeing? |  |  |  |  |  |  |  |  |  |
|  | Mental functions | From birth to now has the child ever had repeated fits or repeated episodes of becoming rigid? |  |  |  |  |  |  |  |  |  |
|  |  | >6 weeks - <3 mo | >3-<6 mo | >6-<9 mo | >9-<12 mo | >12-<15 mo | >15 - <18 mo | <18 -<21 mo | >21 - <24 mo | >24-<30 mo | >30-<36 mo |
| ACTIVITIES | MOBILITY (Gross motor) | GM1 Lifts chin | GM3 Holds head upright | GM6 Bears weight on legs | GM8 Rolls over | GM11 Crawls (in any way) | GM13 Stands holding on | GM15 Walks with help | GM17 Stoops over | GM20 Kicks a ball (in any way) | GM23 Throws a ball overhand) |
|  |  | GM2 Lift head up 90 degrees | GM4 Pulls to sit - no head lag | GM7 Sits with help | GM10 Sits by self well. | GM12 Pulls self to stand | GM14 Walks using both hands | GM16 Walks -falls at times | GM18 Walks well | GM21 Runs well, | GM24 Kneels and can get up |
|  |  | S4 Frolics alone | GM5 Lifts head, shoulders & chest |  |  |  |  |  | GM19 Runs basic | GM22 Climb on to furniture |  |
|  | MOBILITY (Fine motor) (and sensory function) | FM4 Reaches | FM6 Grasps. | FM9 Transfers objects | FM11 Strikes objects | FM12 Finds object hidden | FM14 Puts blocks in and out of cup | FM15 Copies pushing a car | FM17 Dump blocks out of the jar | FM18 Scribbles in straight lines. | FM23 Tower of 4 |
|  |  | FM1 Follows face to mid line | FM7 Pick a larger object from ground |  |  | FM13 Can use a neat pincer grasp |  | FM16 Blocks in and out of a jar | S10 Drinks from a cup by self |  | FM24 Tower of 6 |
|  |  |  | FM2 Fixes and follows right to left |  |  |  |  | FM20 Tower of 2 |  |  | FM28 Threads 6 beads |
|  | COGNITION (Learning and Applying knowledge) |  |  |  |  |  |  | L10 Follows one stage commands | Shows curiosity get things out of reach | L15 2 stage commands | FM22 Understands "give me one" |
|  |  |  |  |  |  |  |  |  |  |  | L27 3 stage commands |
|  | COMMUNICATION (Producing) |  | L3Laughs and chuckles | L5 Makes single syllable sounds | S8 Stretches to be picked up. | L9 Shakes head or something for "No" | S11 Indicate by pointing | L11 Unclear talk or jabber | L12 2 words other than mama dada | L19 Names at least 5 things. | L23 Names at least 10 things. |
|  |  |  |  |  | L7 Says two syllable babble |  |  | Interactive eye contact | L13 Says two words together | L14 Says at least 6 or more words | L17 Speaks clearly in sentences |
|  |  |  |  |  |  |  |  |  | Copies sounds that you make | S16 Indicates need for a poo or a pee. | Use words to ask for help |
|  |  |  |  |  |  |  |  |  |  | S18 Able to greet |  |
|  |  |  |  |  |  |  |  |  |  | *Imitate gestures or noises in play* |  |
|  |  |  |  |  |  |  |  |  |  | S19 Understands to share things |  |
|  | Communication (Receiving) - | L1 Startles to sounds | L4 Turns to voice – | L6 Responds to name |  | L8 Understands when told “no” | L10 Follows one stage commands |  |  | L15 Follows two stage commands | L21 Knows actions of objects. |
|  |  |  |  |  |  |  |  |  |  | L18 Points to more than 1 body part. | L22 Identify up to 10 things. |
|  | Major Life Areas (Play) | S3 Frolics in response |  |  |  |  |  |  | *Plays mainly beside other children* | *Pretend objects are something else?* | S22 Pretend cooking or similar |
|  |  | S2 Smiles in response |  |  |  |  |  |  |  | *Take an interest or play with children* |  |
|  |  |  |  |  |  |  |  |  |  | S17 Wants to join in singing games. |  |
|  | Self Care and Interpersonal |  | S5 Recognises caregivers | S7 Helps hold a cup. |  | S12 Can eat by picking up morsels |  | S13 Puts hands out to be washed | S15 Hold a spoon by self spills some | *Engage in pretend play* | S21 Eats off spoon without spilling. |
|  |  |  |  |  |  | S9 Hold spoon not get to mouth well |  |  |  | *Plays peek a boo or a similar game* | S23 Can make and eat own morsels |
|  |  |  |  |  |  |  |  |  |  | S14 Pretending objects are something else? | S25 Wants to go visit friend’s house. |
| Total number of items in Activities | 99 | 8 | 9 | 6 | 5 | 8 | 5 | 9 | 11 | 20 | 18 |

**Supplementary Table 5.** Number of items in each age-group meeting minimal diagnostic accuracy levels.

| **MDAT Items (N = 144)** |  |  |  |  |  |  |
| --- | --- | --- | --- | --- | --- | --- |
|  | **0-6 Months** | **7-12 Months** | **13-18 Months** | **19-24 Months** | **25-30 Months** | **31-36 Months** |
| **90%** | 5 | 1 | 0 | 0 | 1 | 4 |
| **80%** | 23 | 22 | 9 | 29 | 46 | 65 |
| **70%** | 30 | 45 | 50 | 65 | 89 | 102 |
| **Additional items (n = 62)** |  |  |  |  |  |  |
|  | **0-6 Months** | **7-12 Months** | **13-18 Months** | **19-24 Months** | **25-30 Months** | **31-36 Months** |
| **90%** | 0 | 0 | 0 | 0 | 0 | 0 |
| **80%** | 0 | 0 | 0 | 3 | 11 | 12 |
| **70%** | 0 | 0 | 7 | 14 | 16 | 17 |

**Supplementary Figure 1.**
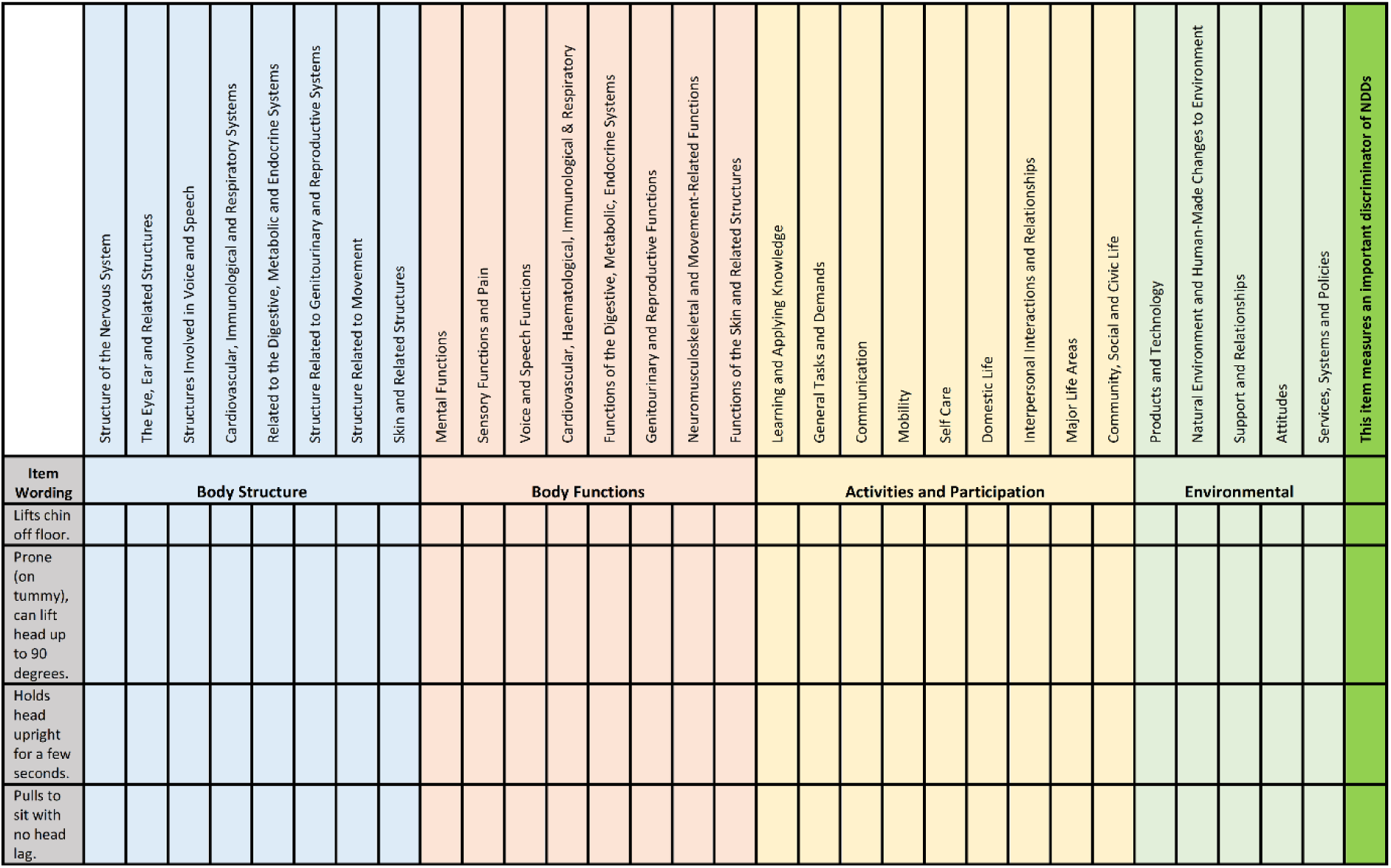
**A portion of the framework for coding items from the MDAT and 18 developmental screening tools to the ICF-CY framework**

**Supplementary Figure 2.**
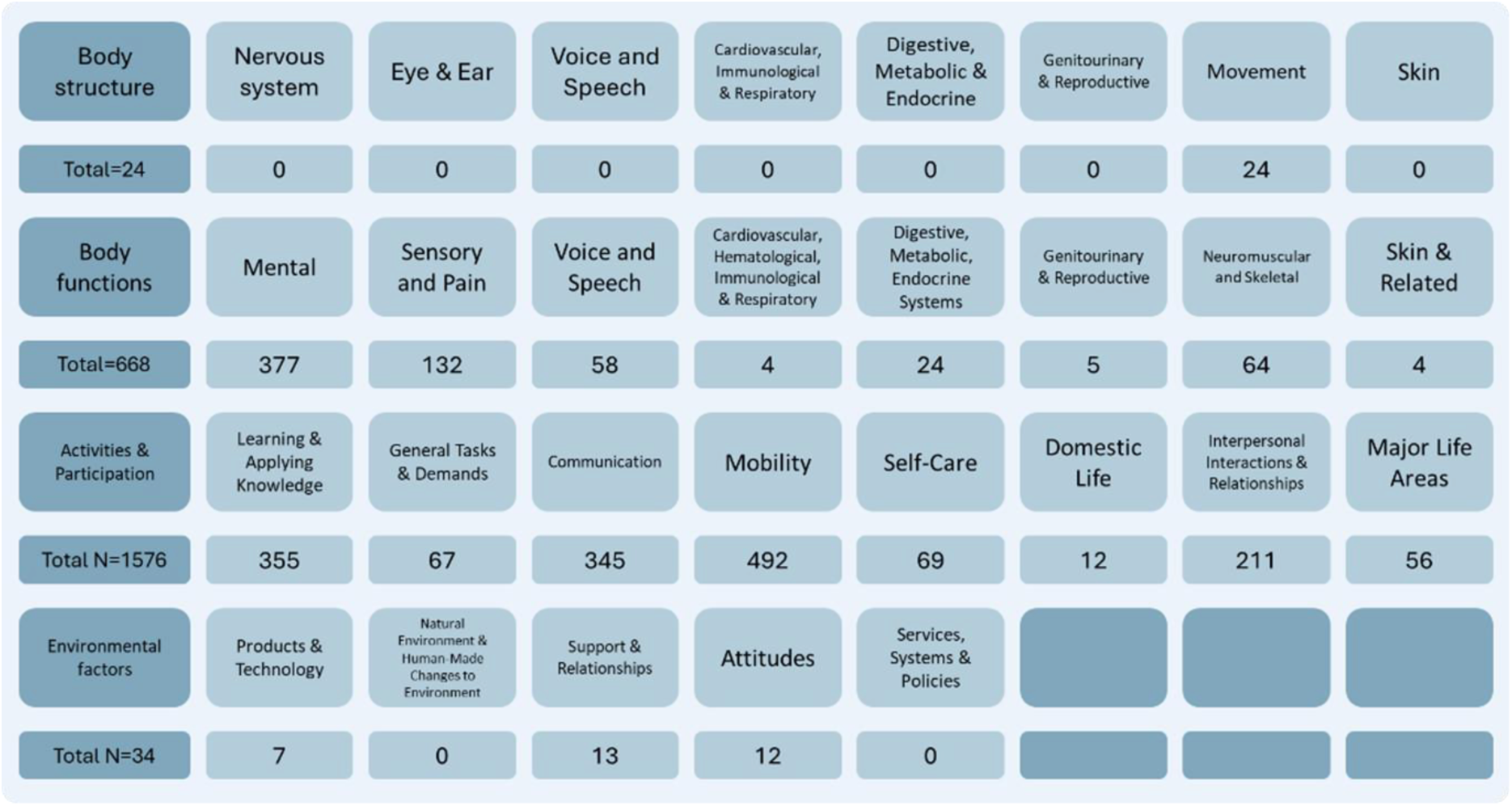
**Summary of items coded onto ICF-CY from MDAT and 18 developmental screening tools**

**Supplementary Figure 3.**
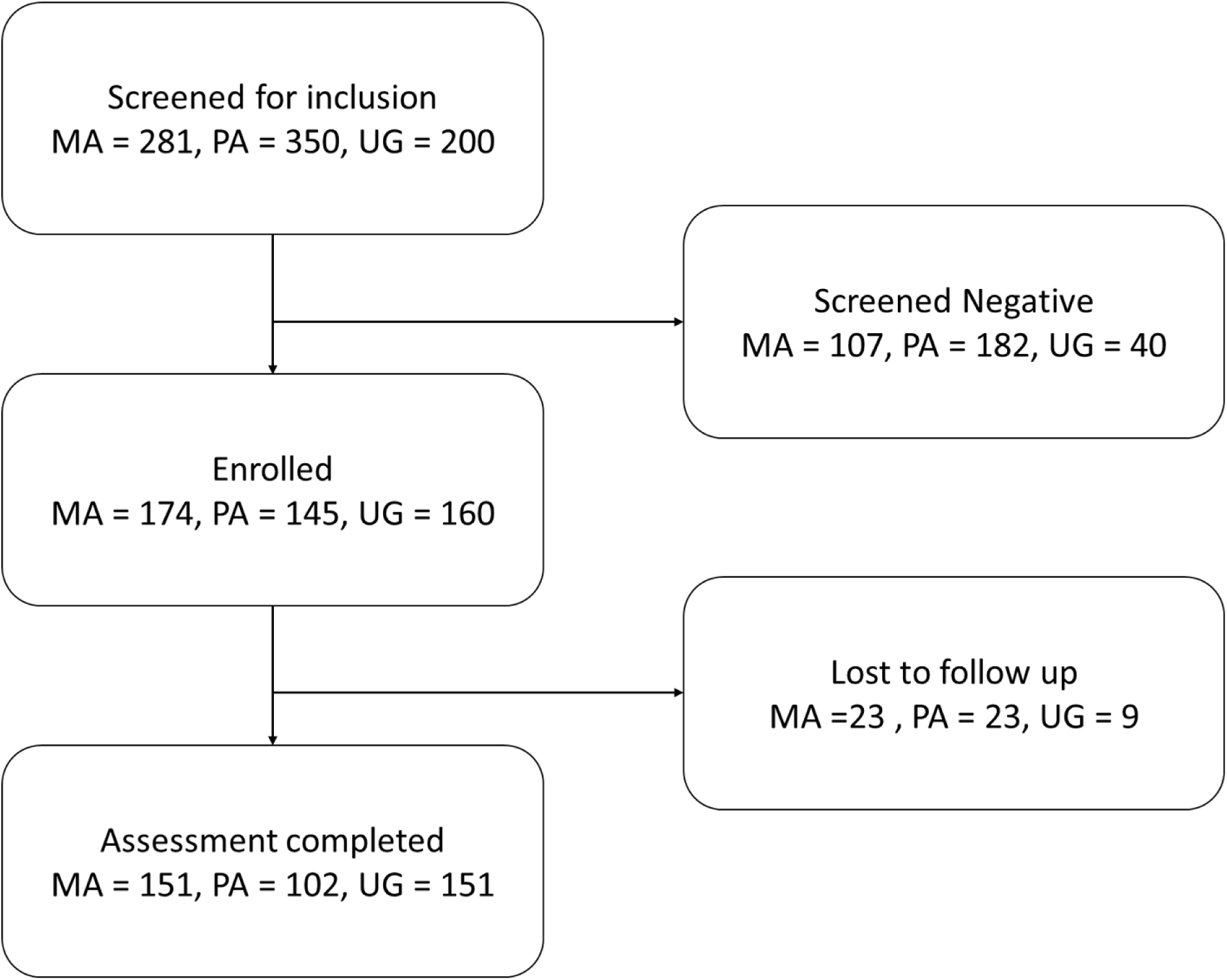
**Consort diagram for item selection analysis**

**Supplementary** Figure 4 **– Graphs of all items for diagnostic accuracy (uploaded separately)**

**Item performance**

The six panels of Figure 3 show the endorsement probability and diagnostic accuracy plots for three example items from the INDIGO. In the endorsement probability plots, (panels a, c,e) the proportion of children who responded “yes” to an item is given at each age, the dotted line is an empirical probability plot, and the solid red and black lines represent the NDD and Typically developing group, respectively. Note the age is expressed as a decimal age where 0.25 is 3 months, 0.5 is 6 months, 1 is 12 months, etc.

Panel a of Figure 3 lets us observe a number of features of the relationship between the item, age and the endorsement rate which hint at the diagnostic utility of the item across the age range. Firstly, we can see that at a few days old, neither group is able to *sit with help*. If we are told that at this age a child **could not** *sit with help*, we have virtually no information about whether they are likely to be NDD, as neither NDD children nor typically developing children **could** *sit with help* at this age. If we were told that the child could *sit with help* at this age we would assume the data was incorrect as this is entirely unfeasible. Secondly, at around 6 months old (decimal age 0.5) there is around a 95% chance of a typically developing child *sitting with help* and only about a 33% chance of an NDD child being able to *sit with help*. If we are told at this age that a child **could not** sit with help, we would consider them more likely to be in the NDD group. Thirdly we can see that at the age of one year, the probabilities of being able to *sit with help* level off and almost all of the typically developing children can do it with approximately 60% of the NDD children also doing it. If we are told at this age that a child **could not** *sit with help*, we would conclude they were almost certainly in the NDD group as all typically developing children at this age are able to do this. If we were told that the child **could** *sit with help*, we would conclude that there is about a 66% chance of them being in the typically developing group (in this sample with its specific prevalence) and a 33% chance of them being in the NDD group, as by this age, about half of the NDD group are able to sit with help. Though difficult to comprehend at first reading, from this example, we can see how an item can become more or less useful for diagnosis depending on the age at which it is asked.

In Figure 3 Panel b we have the diagnostic accuracy, sensitivity, and specificity of a single item for detecting whether a child does or does not belong to the NDD group across the age range. The diagnostic accuracy is the measure of the proportion of the time that we will make the correct diagnosis into, typically developing Vs. NDD, if we assume that a “Yes” answer to an item means that a child belongs to the typically developing group and a “No” means they belong to the NDD group. At age 0, we can see that the measure of diagnostic accuracy is around 50%. This tells us that the response to the question at this age gives us no information about the group membership of the child, obviously, this is because neither group can do the item at this age. At the age of 6 months, we see the diagnostic accuracy raise to almost 90%. Typically developing children are likely to be able to successfully complete this item at this age, while the children with NDD are not. This item is particularly good for diagnosing group membership at around this age this age. At the age of 1 year and onwards, we can see that the diagnostic accuracy levels off at around 0.75. This is because, even though we are almost certain that a response of “No” means a child has NDD, it has become more likely that a response of “Yes” could come from a member of the NDD group, so the overall level of diagnostic accuracy comes down. Note that the specificity, across the age range is equal to the probability of endorsement in the Normal group in panel a, and the sensitivity is equal to one minus the endorsement rate for the NDD group.

Examining panels c and d we can see that the item, *says 6 or more words*, does not become a useful diagnostic marker until around the age of 2-3 years old. Panels e and f, in Figure 3, show an item that is never a useful diagnostic marker, *Smiles but not at a particular person.* In this item, the probability of an NDD child being able to do the item is very close to that of a typically developing child meaning a response to this item tells us little about which group, typically developing Vs. NDD, a child likely belongs to.

The numbers of items, in this sample, meeting various levels of diagnostic accuracy in various age ranges are given in Supplementary Table 5. For example, we can see that for the MDAT, there are 23 items in the 0-6 month of range that at some point in that age range, have a diagnostic accuracy of greater than 80% and of these, five items have a diagnostic accuracy of greater than 90%, in the same age range. We can see from this table that I) There tend to be more items suitable at the higher age ranges, ii) there are few items suitable for 13–18-month-olds, iii) while there are many items that cross the 80% diagnostic accuracy threshold, few cross the 90% level, and iii) the additional added items have high levels of diagnostic accuracy in on this sample, only a few for the older children.

The full code used to create the graphs can be found in Figure S1 for those with further interest.

**Figure S1 - Code To create the graphs**

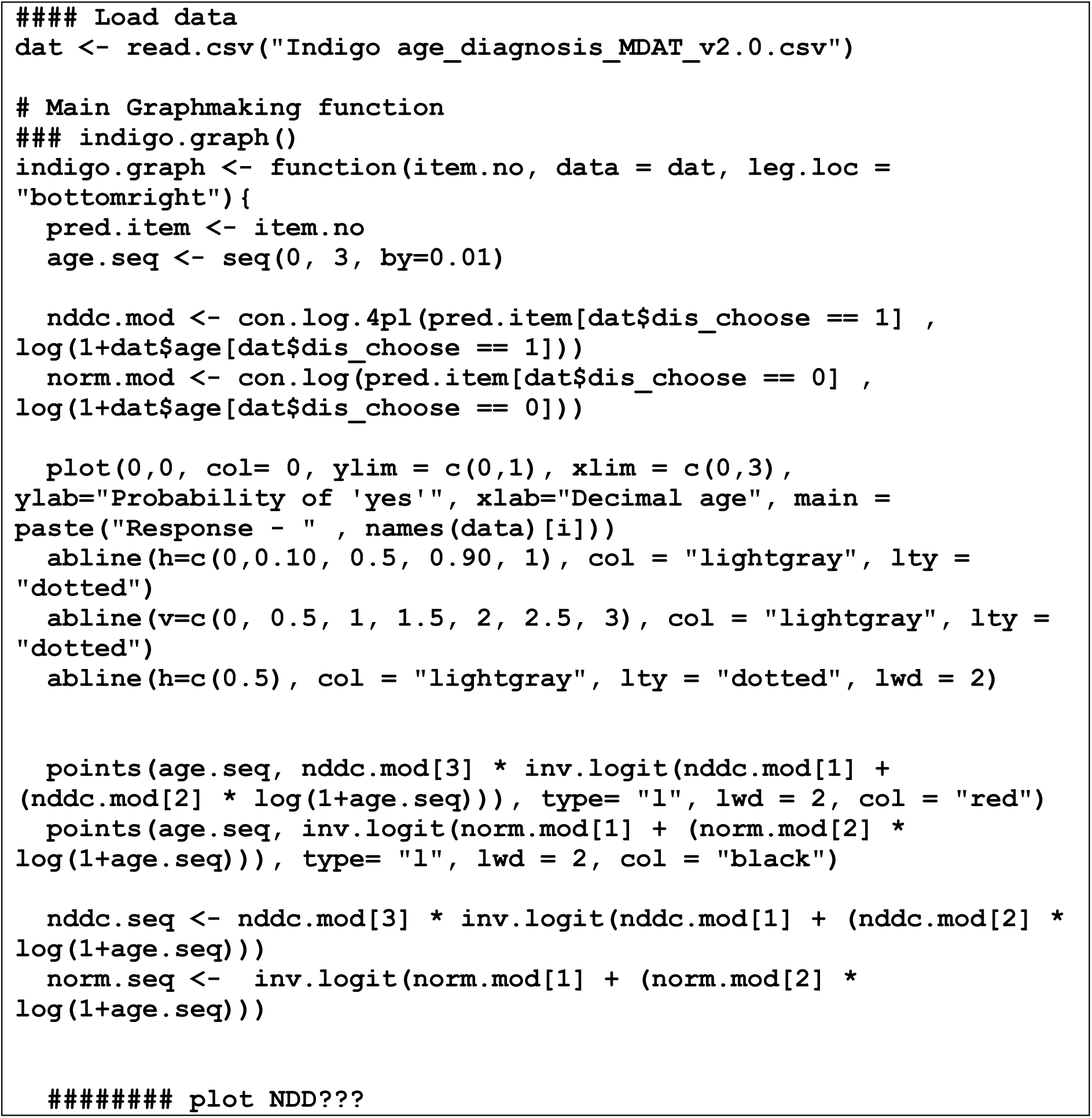
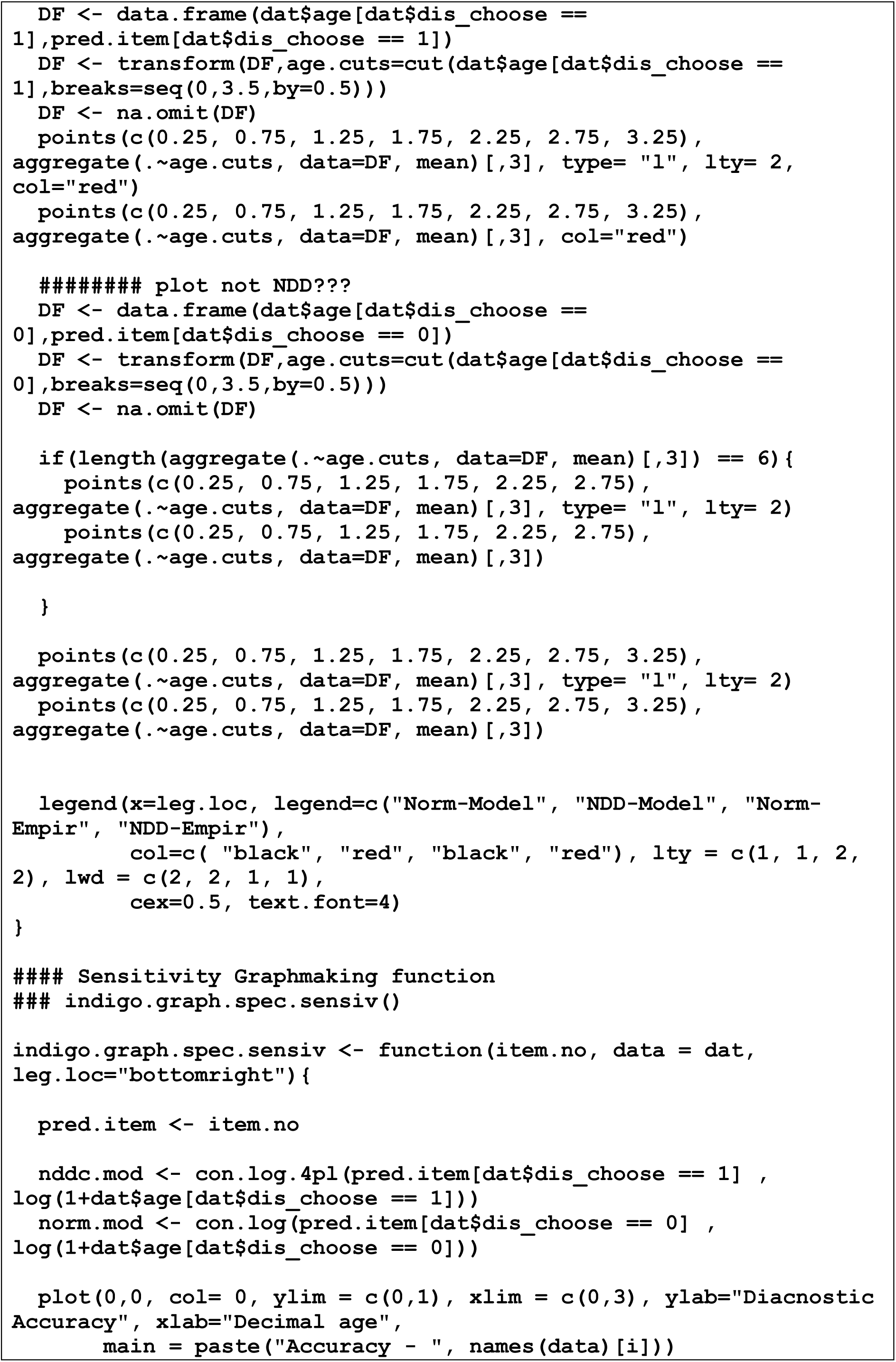
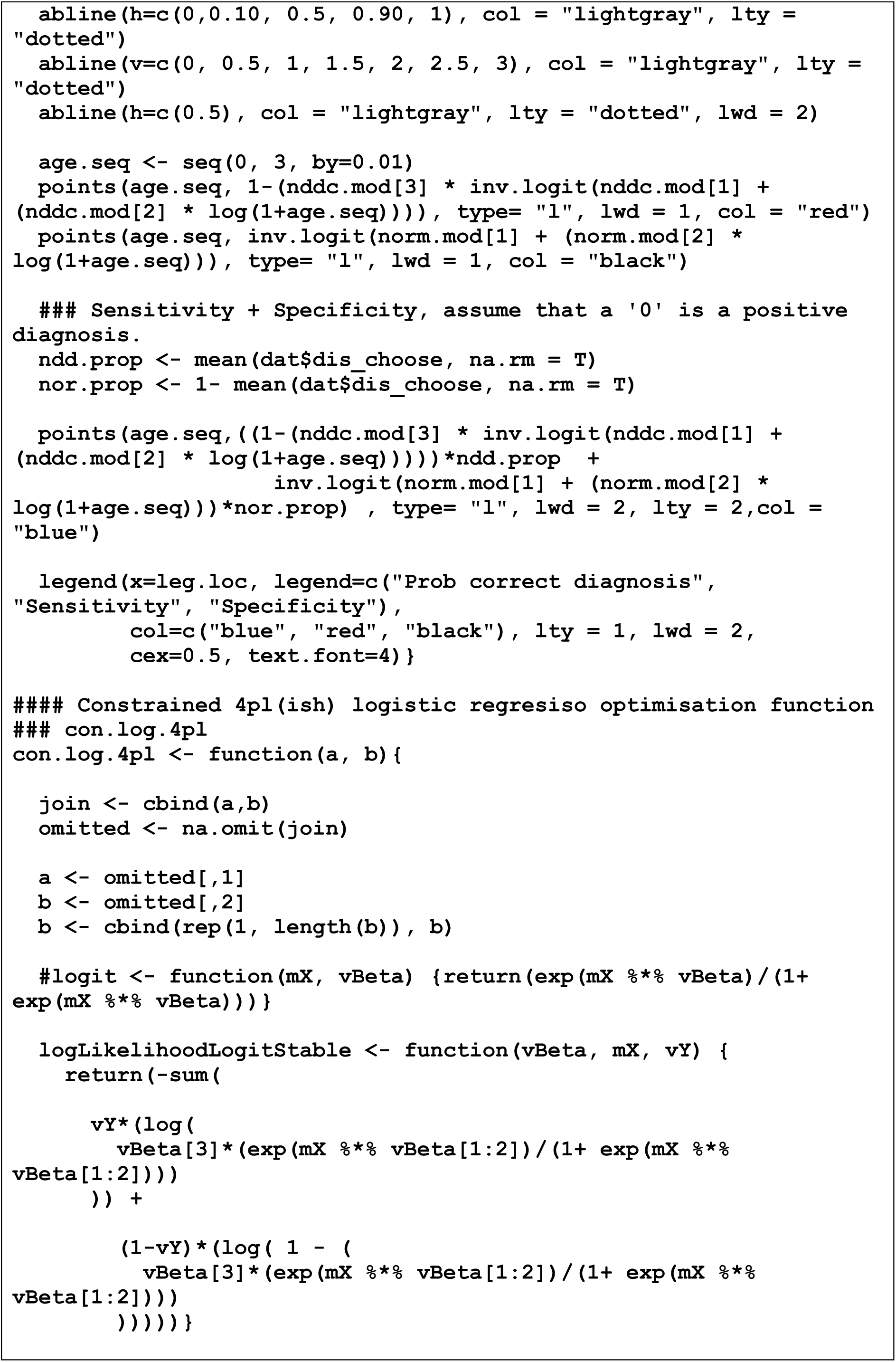
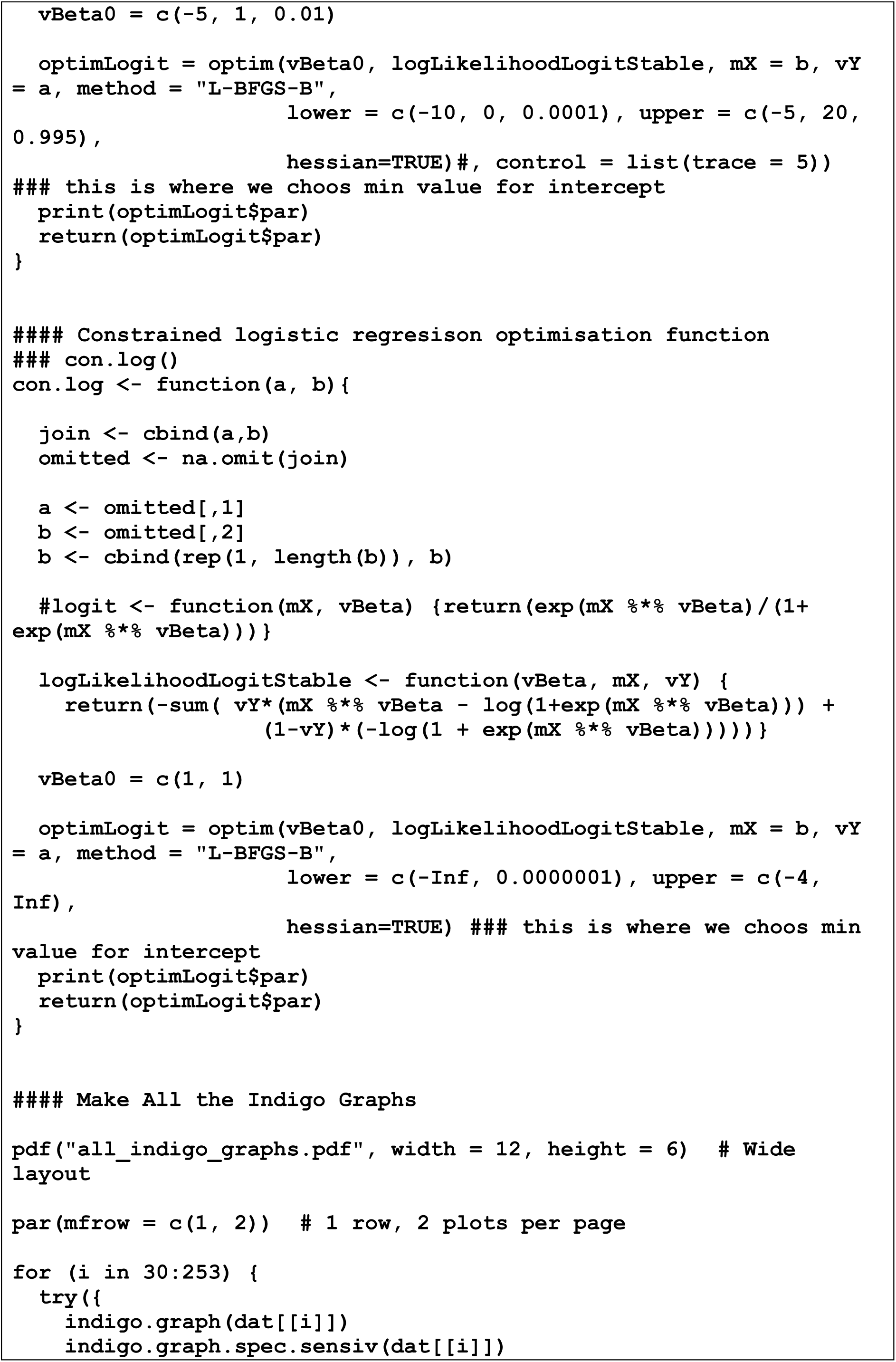
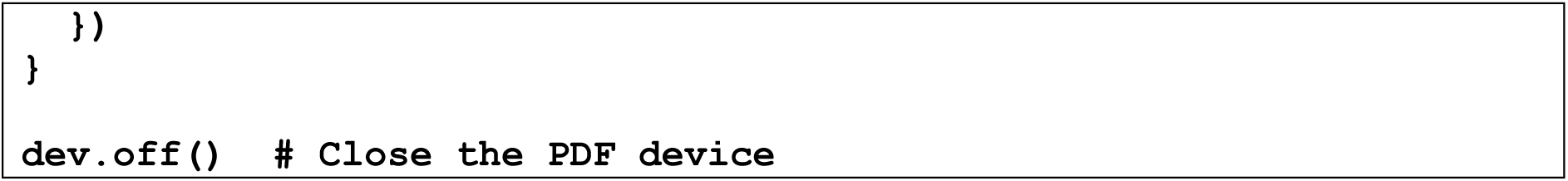

**Supplementary File 1 – Technical details of the item response model**

