## Supplementary Figure 4 All graphs for "Creation of a tool for the Identification of Neurodevelopmental Disabilities to Improve Global Outcomes (INDIGO) in children aged 0-3 years in Malawi, Pakistan and Uganda – feasibility of implementation and diagnostic accuracy"

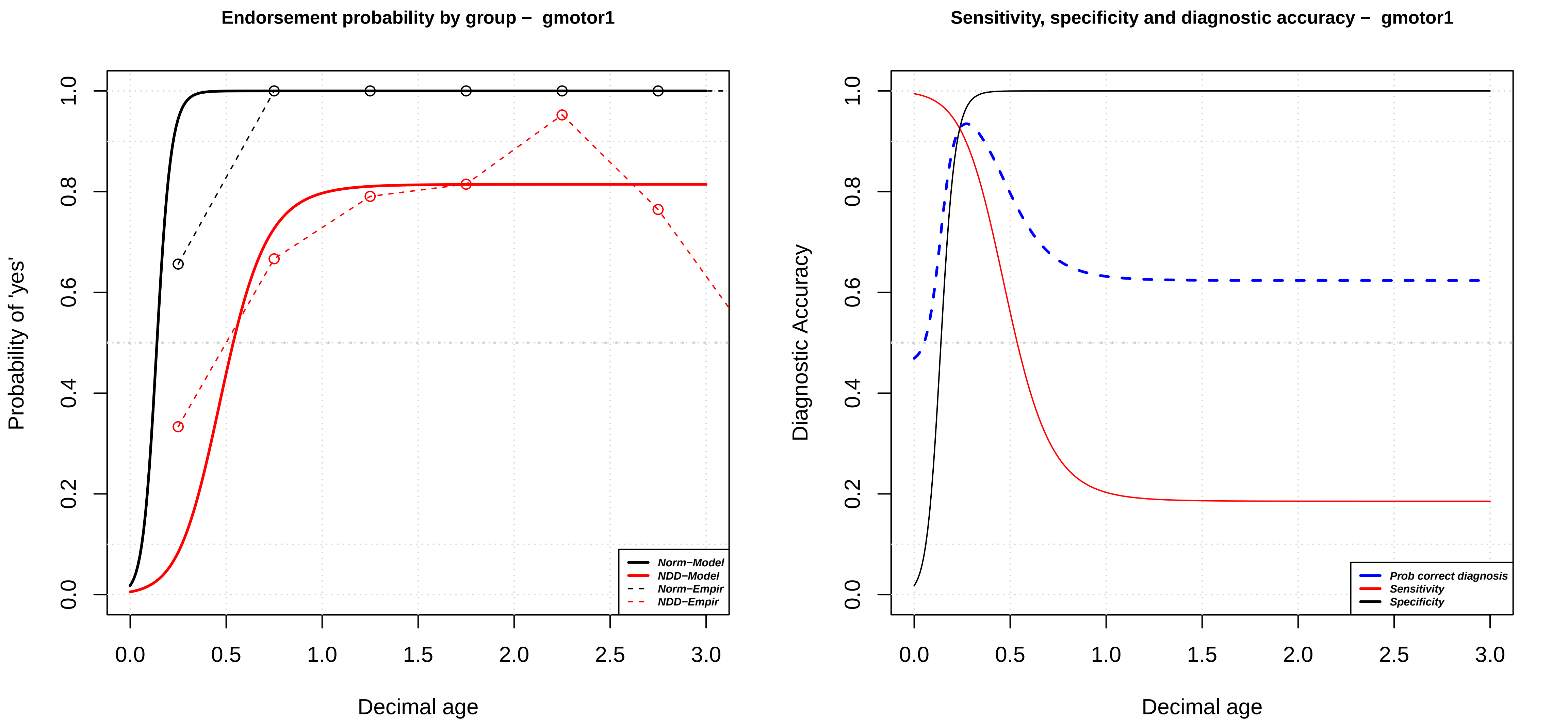

Endorsement probability by group − gmotor1
Probability of 'yes'
Decimal age
Norm−Model
NDD−Model
Norm−Empir
NDD−Empir
Sensitivity, specificity and diagnostic accuracy − gmotor1
Diagnostic Accuracy
Decimal age
Prob correct diagnosis
Sensitivity
Specificity

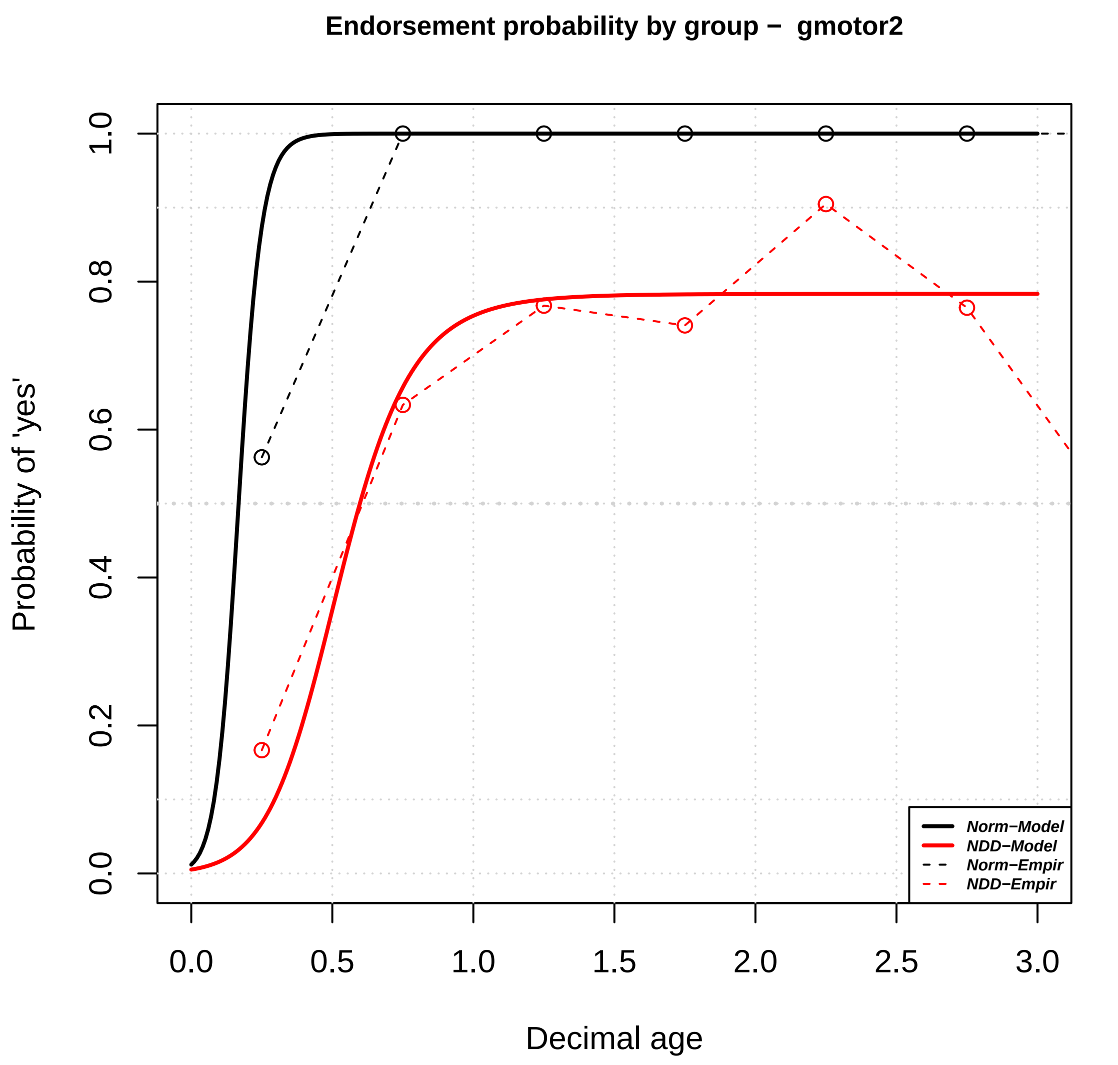
Endorsement probability by group − gmotor2
Probability of 'yes'
Decimal age
0.0
0.2
0.4
0.6
0.8
1.0
0.0
0.5
1.0
1.5
2.0
2.5
3.0
Norm−Model
NDD−Model
Norm−Empir
NDD−Empir

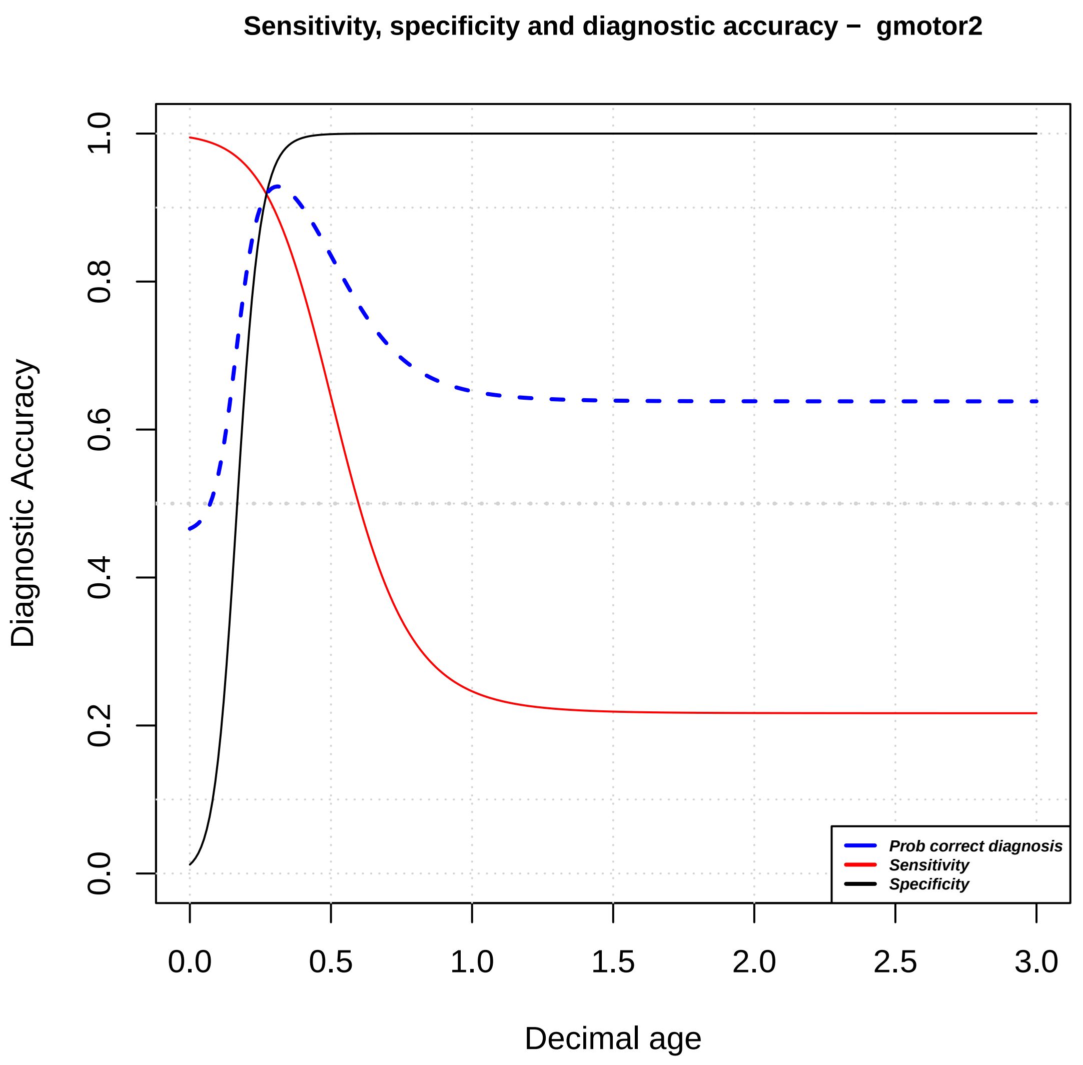
Sensitivity, specificity and diagnostic accuracy − gmotor2
Diagnostic Accuracy
Decimal age
0.0
0.2
0.4
0.6
0.8
1.0
0.0
0.5
1.0
1.5
2.0
2.5
3.0
Prob correct diagnosis
Sensitivity
Specificity

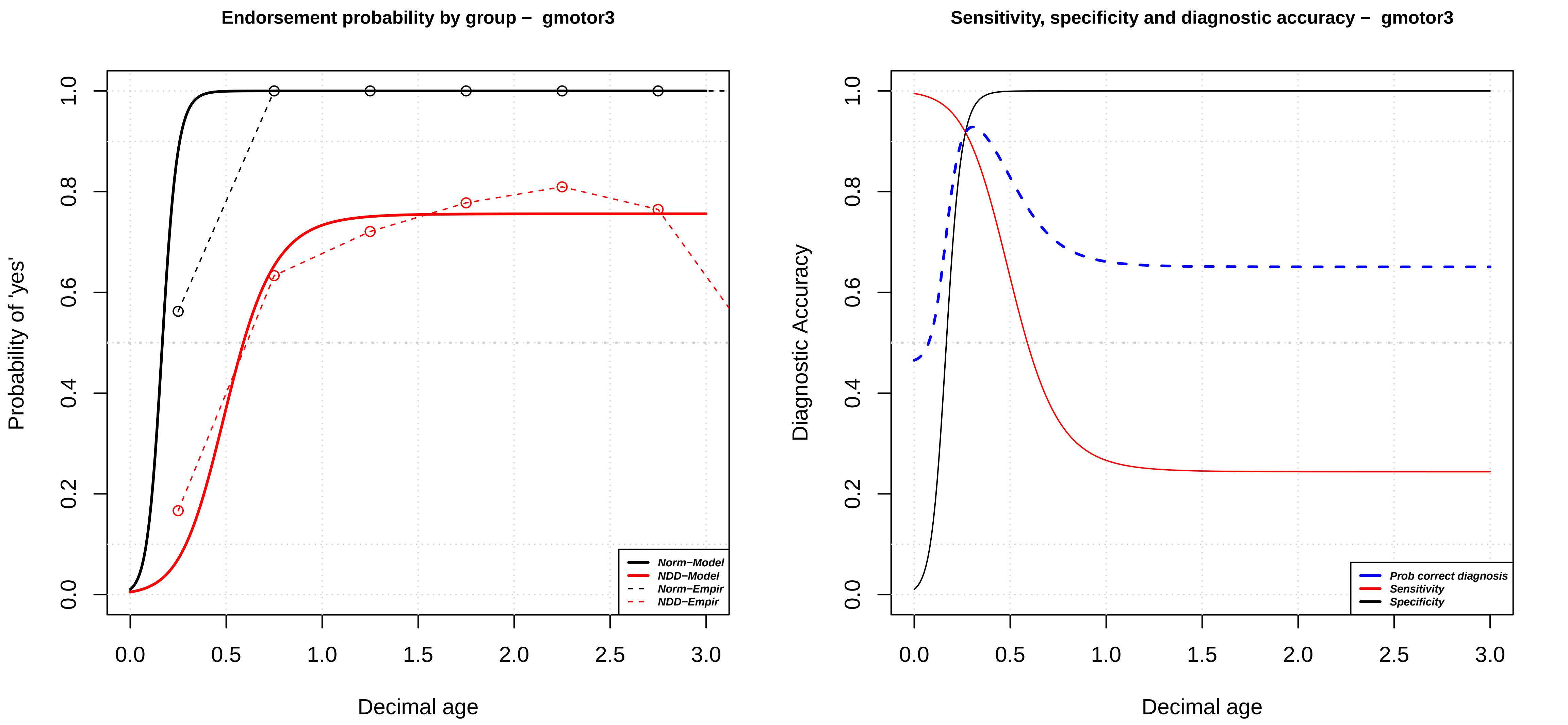

Endorsement probability by group − gmotor3
Probability of 'yes'
Decimal age
0.0
0.2
0.4
0.6
0.8
1.0
0.5
1.5
2.0
2.5
3.0
Norm−Model
NDD−Model
Norm−Empir
NDD−Empir
Sensitivity, specificity and diagnostic accuracy − gmotor3
Diagnostic Accuracy
Decimal age
Prob correct diagnosis
Sensitivity
Specificity

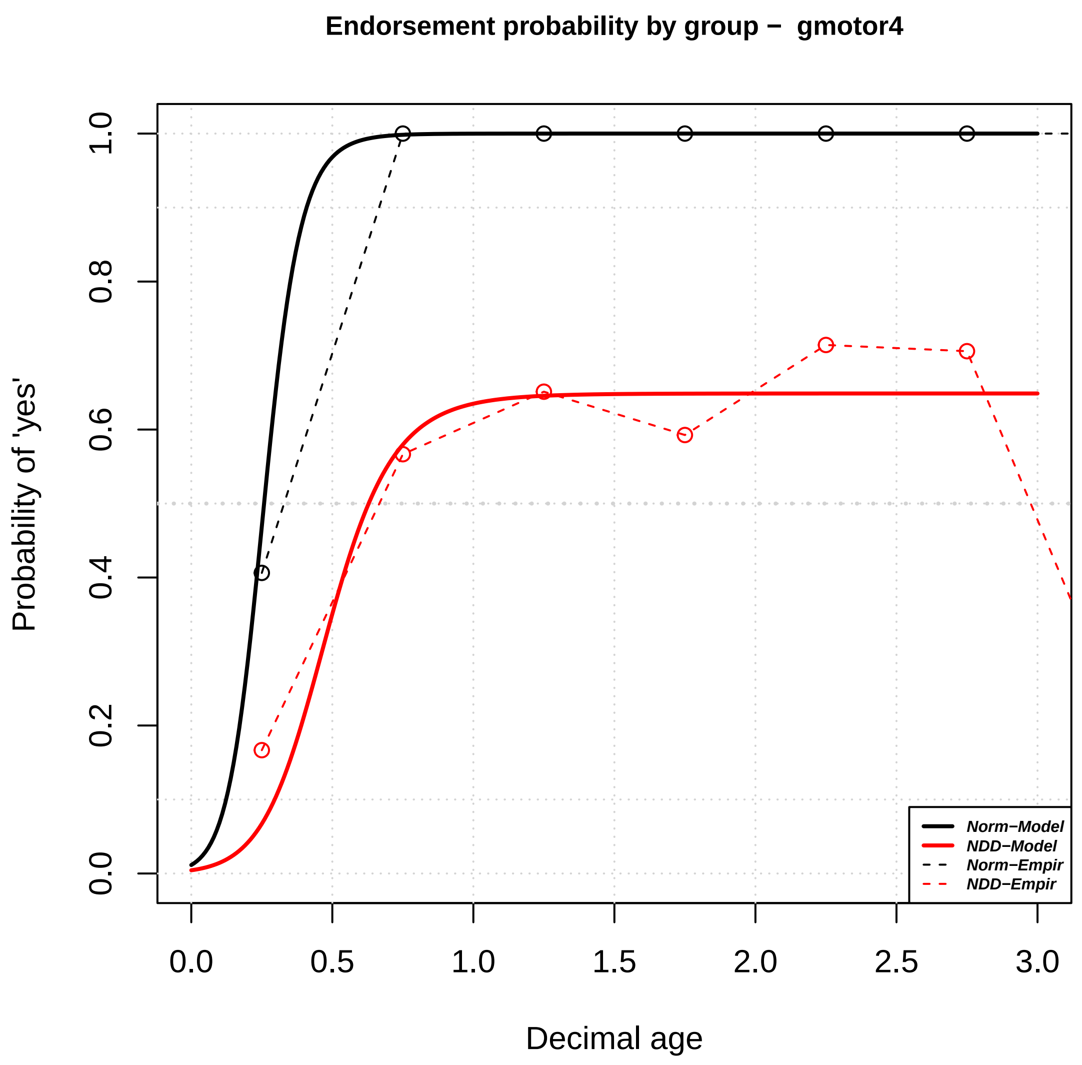

Endorsement probability by group – gmotor4
Probability of 'yes'
Decimal age
0.0
0.2
0.4
0.6
0.8
1.0
0.0
0.5
1.0
1.5
2.0
2.5
3.0
Norm−Model
NDD−Model
Norm−Empir
NDD−Empir


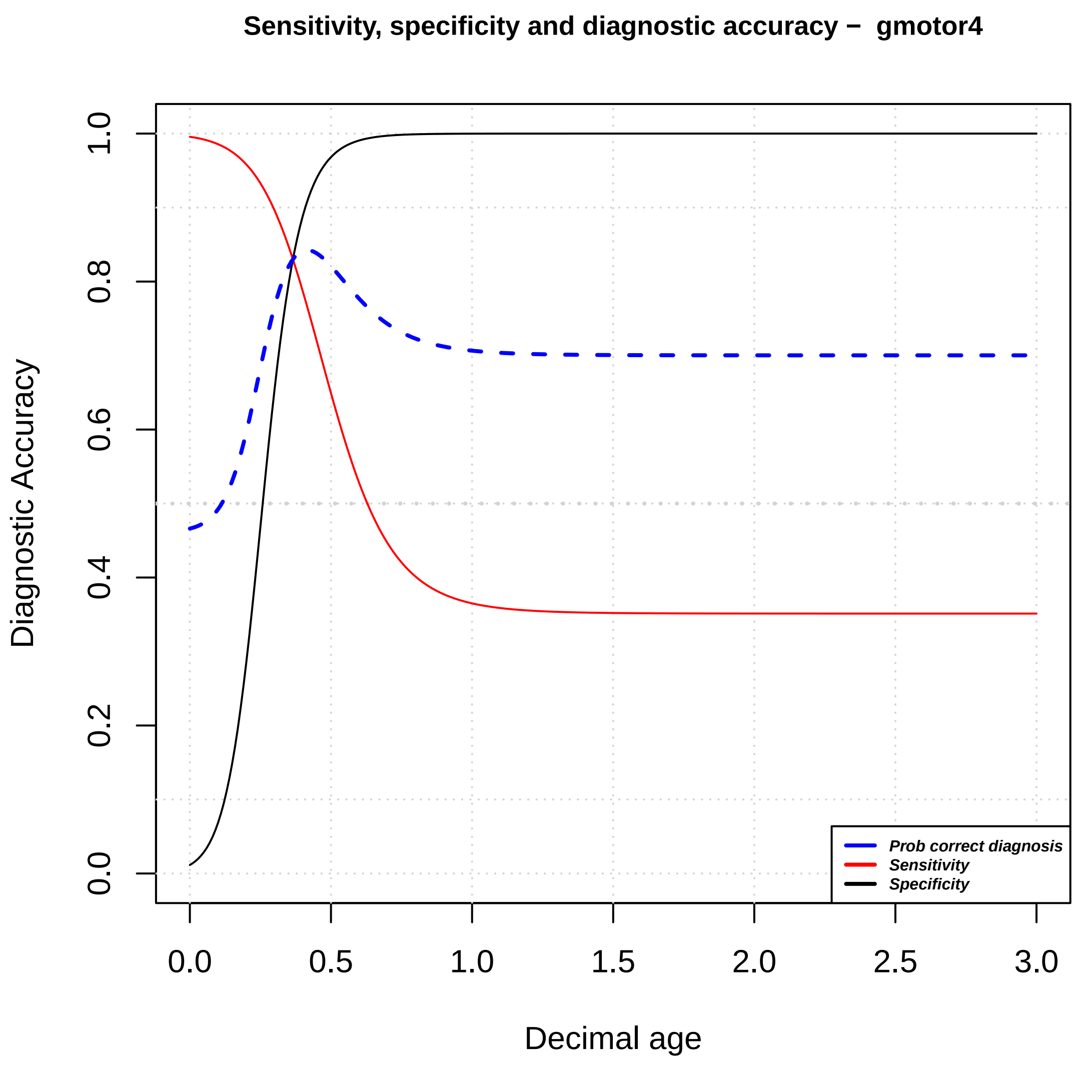

Sensitivity, specificity and diagnostic accuracy – gmotor4
Diagnostic Accuracy
Decimal age
0.0
0.2
0.4
0.6
0.8
1.0
0.0
0.5
1.0
1.5
2.0
2.5
3.0
Prob correct diagnosis
Sensitivity
Specificity

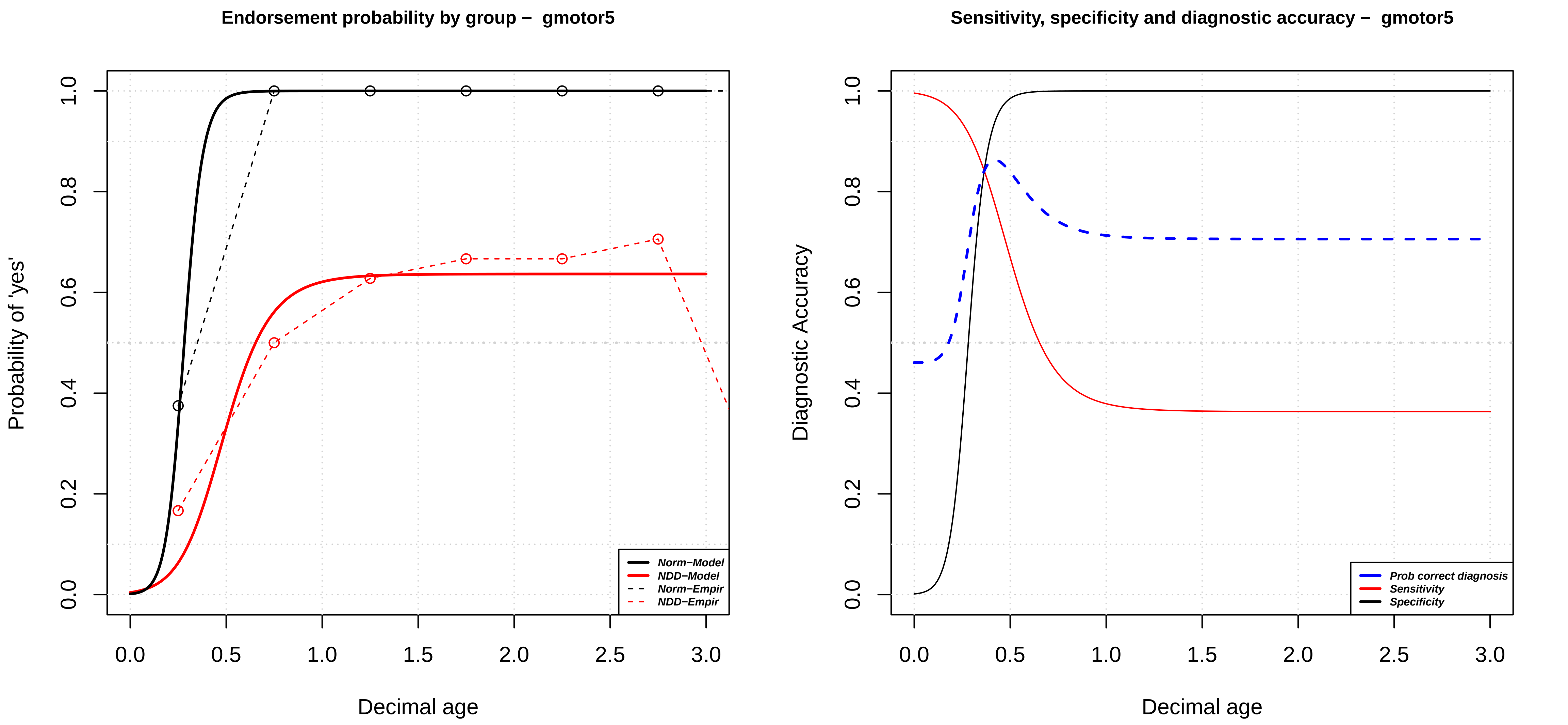

Endorsement probability by group – gmotor5
Probability of 'yes'
Decimal age
0.0
0.2
0.4
0.6
0.8
1.0
0.0
0.5
1.0
1.5
2.0
2.5
3.0
Norm–Model
NDD–Model
Norm–Empir
NDD–Empir
Sensitivity, specificity and diagnostic accuracy – gmotor5
Diagnostic Accuracy
Decimal age
0.0
0.2
0.4
0.6
0.8
1.0
0.0
0.5
1.0
1.5
2.0
2.5
3.0
Prob correct diagnosis
Sensitivity
Specificity

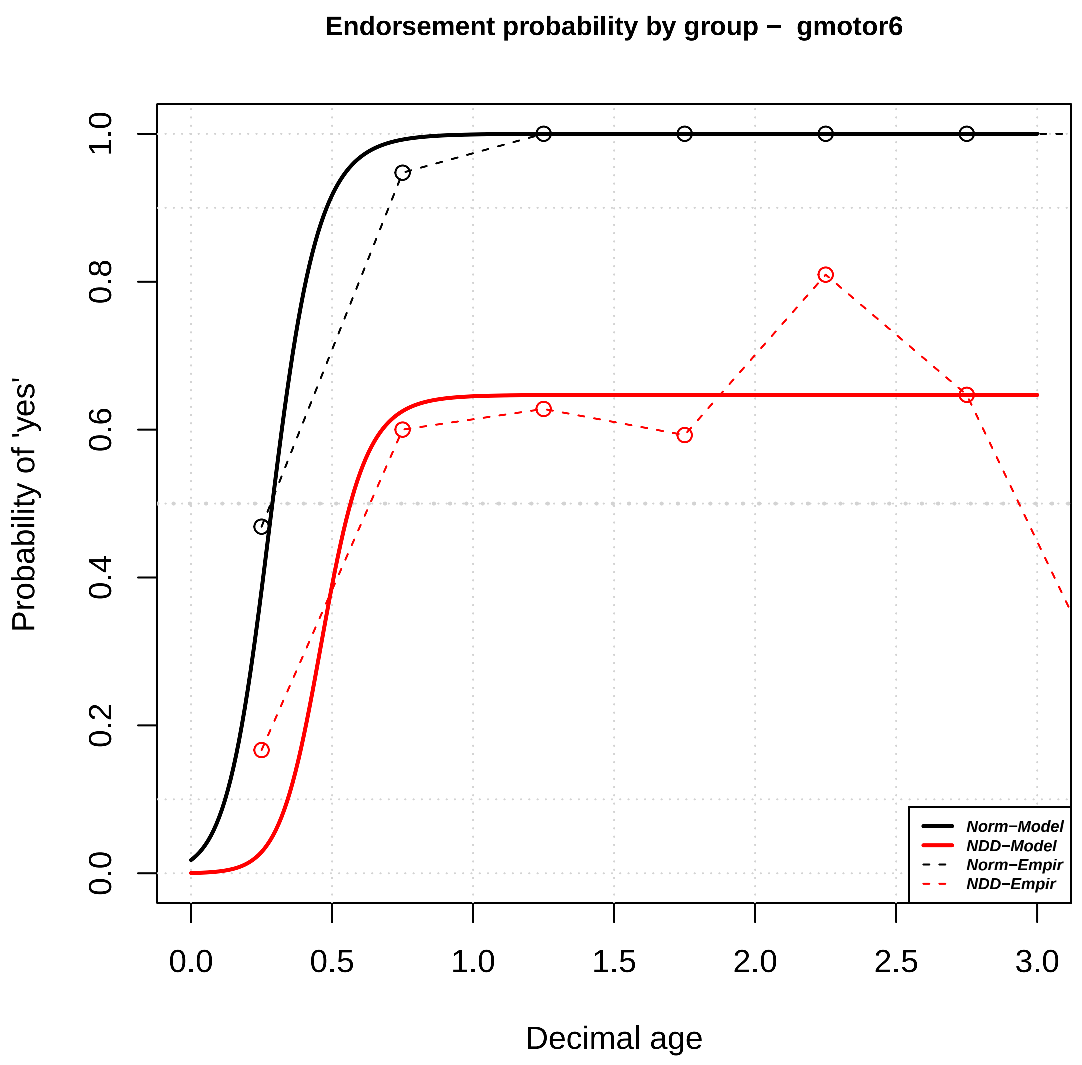
Endorsement probability by group – gmotor6
Probability of 'yes'
Decimal age
0.0
0.2
0.4
0.6
0.8
1.0
0.0
0.5
1.0
1.5
2.0
2.5
3.0
Norm–Model
NDD–Model
Norm–Empir
NDD–Empir

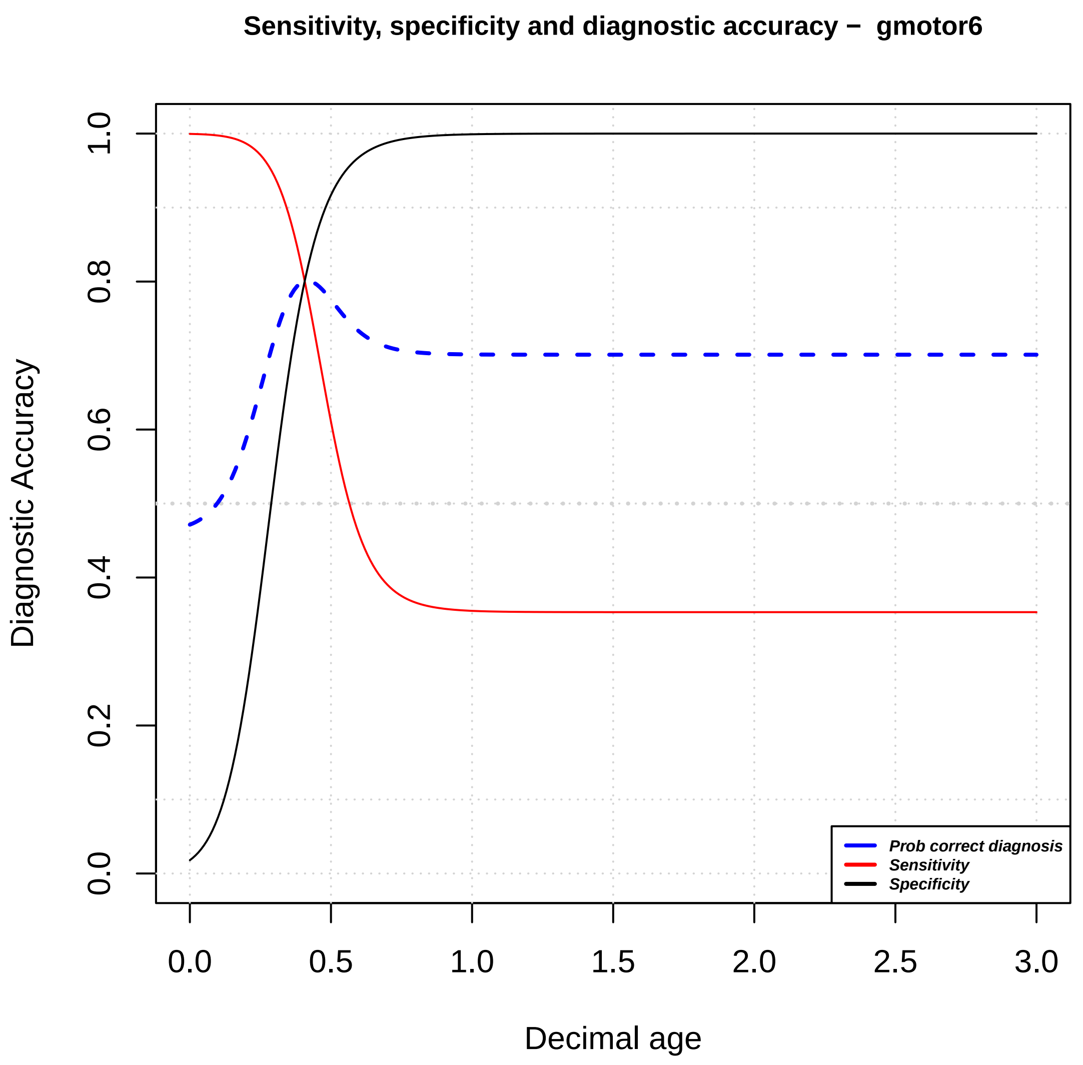
Sensitivity, specificity and diagnostic accuracy – gmotor6
Diagnostic Accuracy
Decimal age
0.0
0.2
0.4
0.6
0.8
1.0
0.0
0.5
1.0
1.5
2.0
2.5
3.0
Prob correct diagnosis
Sensitivity
Specificity

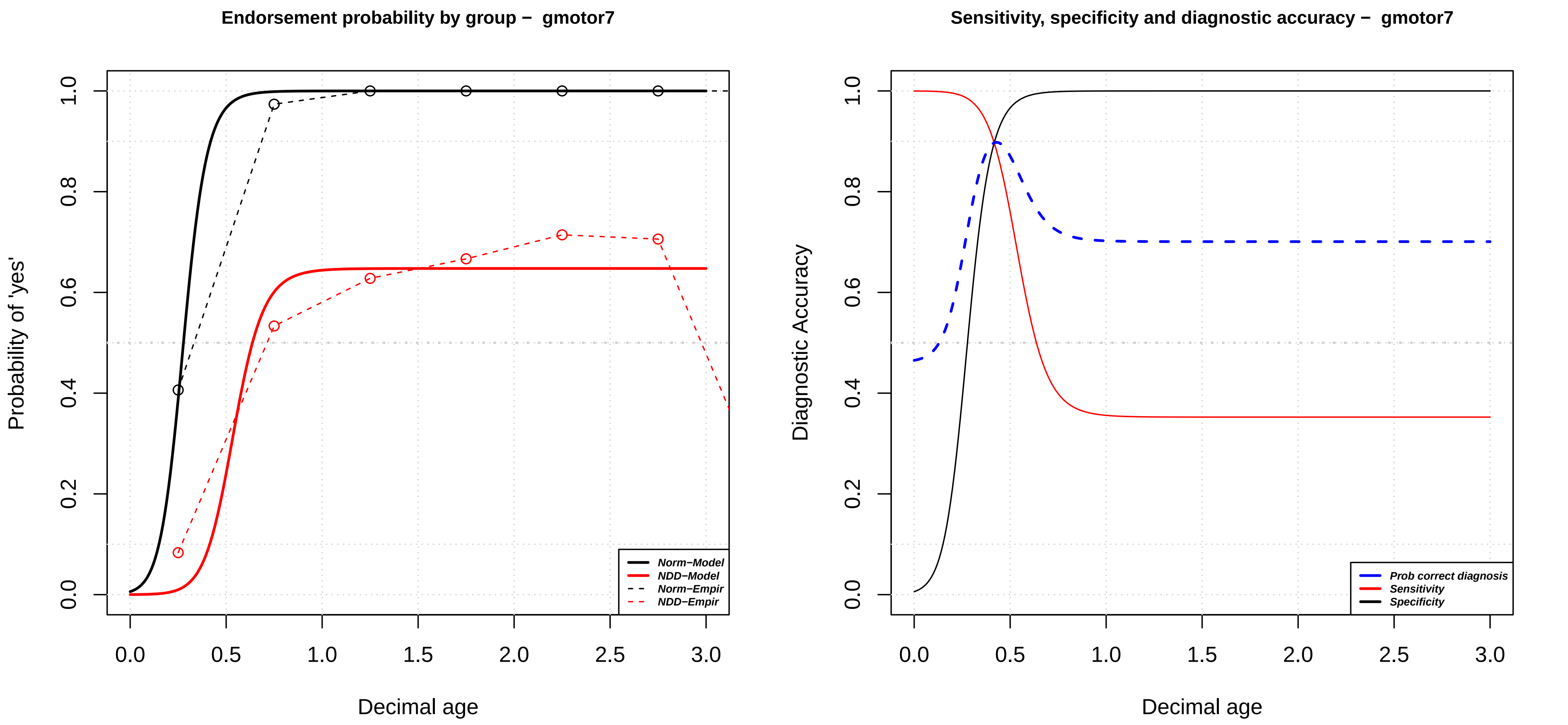

Endorsement probability by group − gmotor7
Probability of 'yes'
Decimal age
0.0
0.2
0.4
0.6
0.8
1.0
0.0
0.5
1.0
1.5
2.0
2.5
3.0
Norm−Model
NDD−Model
Norm−Empir
NDD−Empir
Sensitivity, specificity and diagnostic accuracy − gmotor7
Diagnostic Accuracy
Decimal age
Prob correct diagnosis
Sensitivity
Specificity

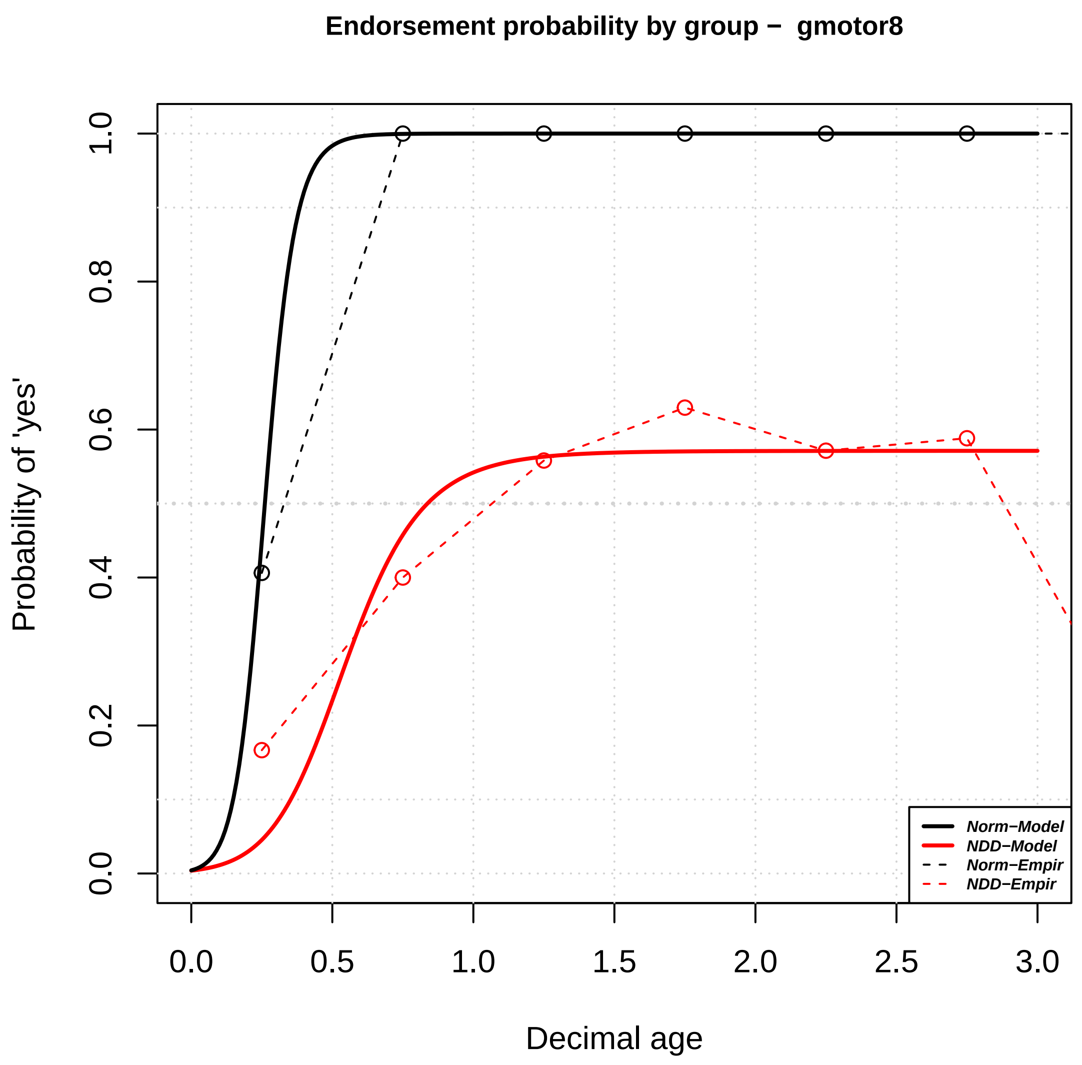
Endorsement probability by group − gmotor8
Probability of 'yes'
Decimal age
0.0
0.2
0.4
0.6
0.8
1.0
0.0
0.5
1.0
1.5
2.0
2.5
3.0
Norm−Model
NDD−Model
Norm−Empir
NDD−Empir

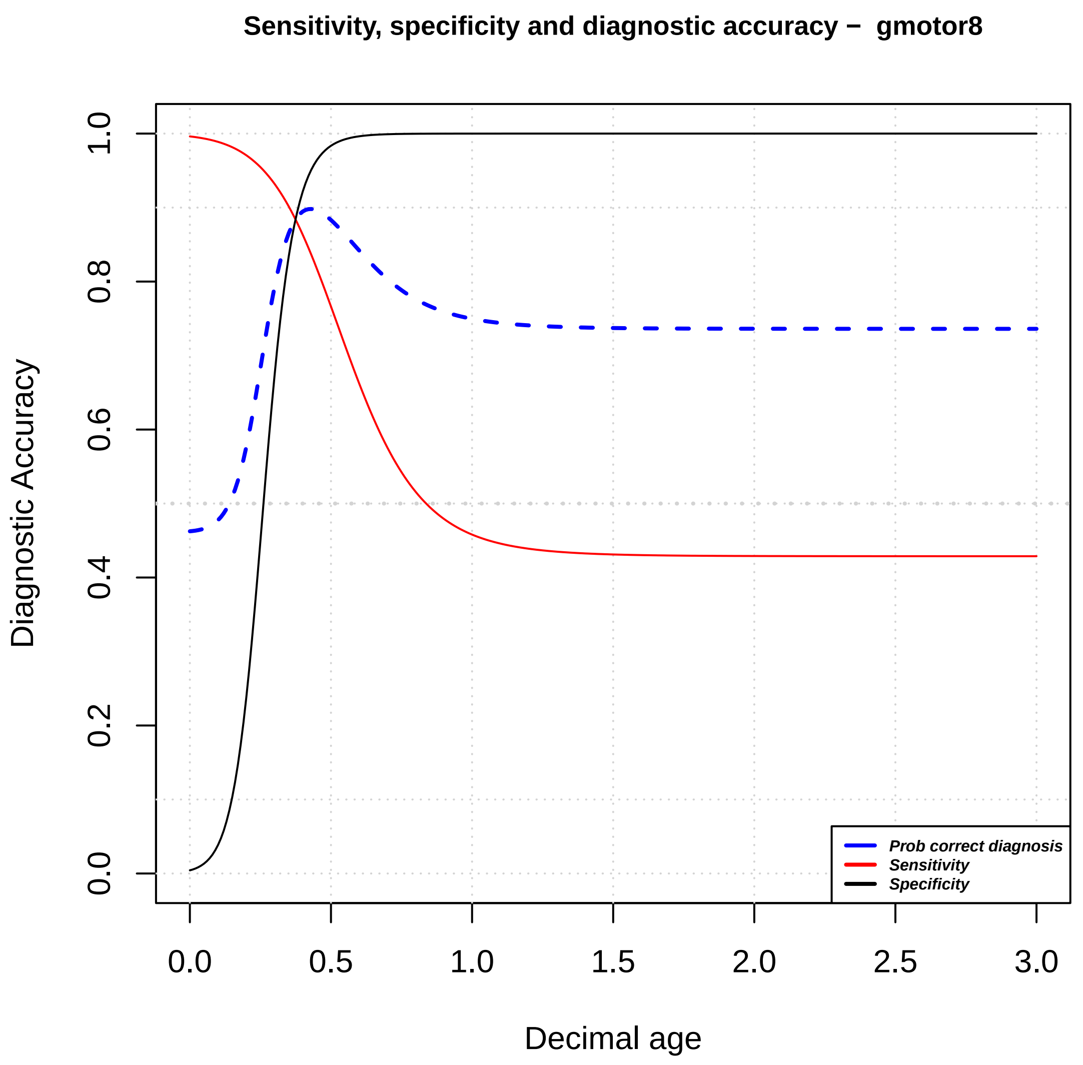
Sensitivity, specificity and diagnostic accuracy − gmotor8
Diagnostic Accuracy
Decimal age
0.0
0.2
0.4
0.6
0.8
1.0
0.0
0.5
1.0
1.5
2.0
2.5
3.0
Prob correct diagnosis
Sensitivity
Specificity

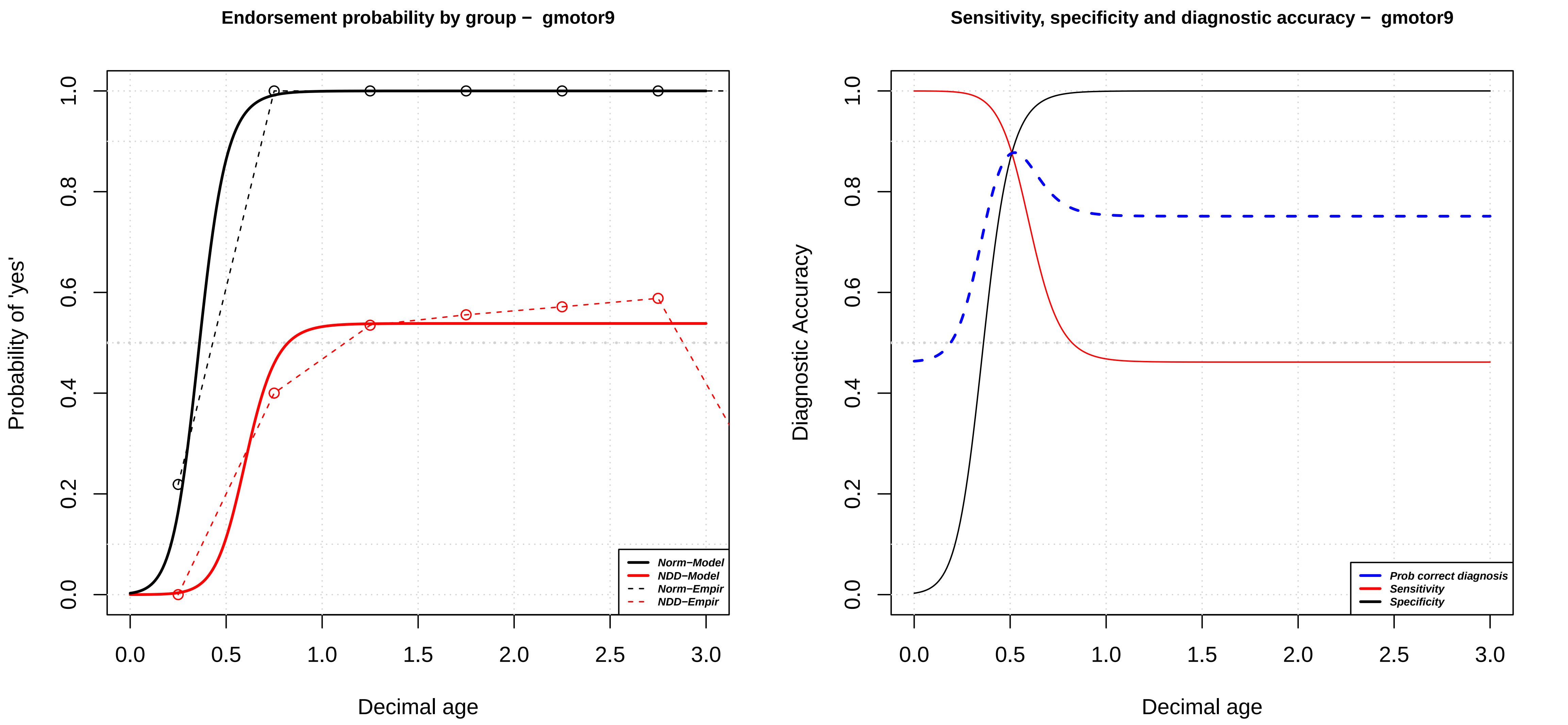

Endorsement probability by group – gmotor9
Probability of 'yes'
Decimal age
0.0
0.2
0.4
0.6
0.8
1.0
0.0
0.5
1.0
1.5
2.0
2.5
3.0
Norm−Model
NDD−Model
Norm−Empir
NDD−Empir
Sensitivity, specificity and diagnostic accuracy – gmotor9
Diagnostic Accuracy
Decimal age
Prob correct diagnosis
Sensitivity
Specificity

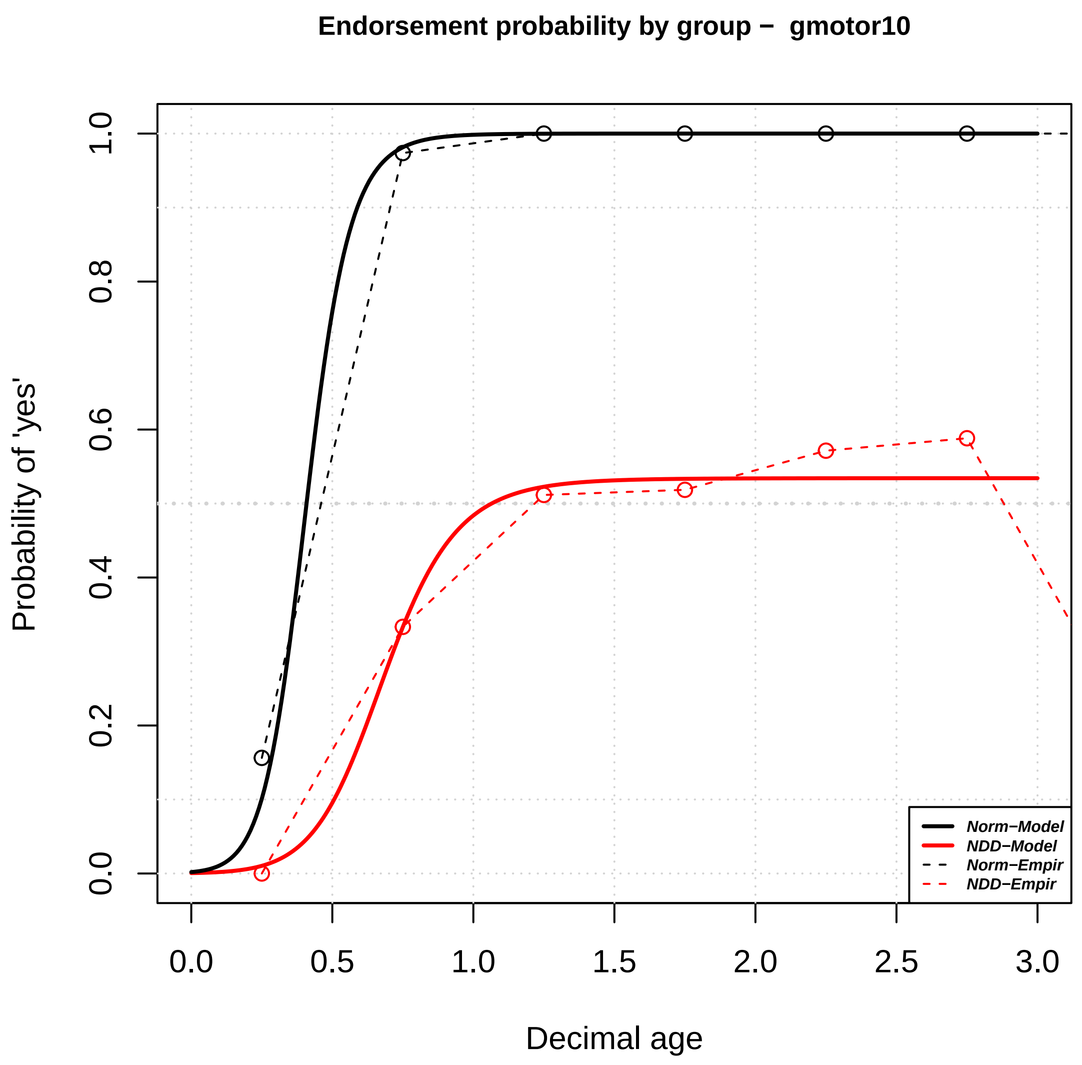

Endorsement probability by group – gmotor10
Probability of 'yes'
Decimal age
0.0
0.2
0.4
0.6
0.8
1.0
0.0
0.5
1.0
1.5
2.0
2.5
3.0
Norm−Model
NDD−Model
Norm−Empir
NDD−Empir


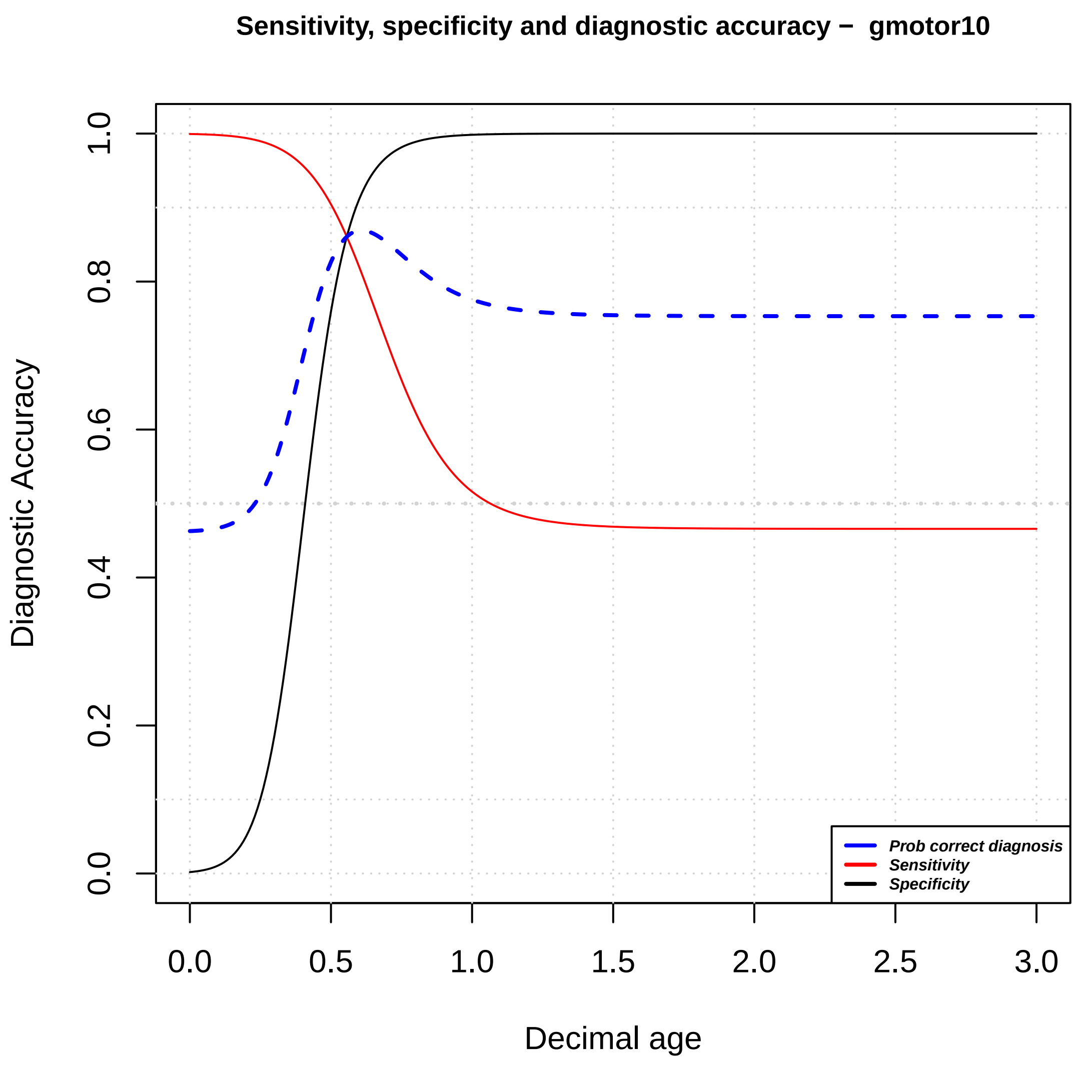

Sensitivity, specificity and diagnostic accuracy – gmotor10
Diagnostic Accuracy
Decimal age
0.0
0.2
0.4
0.6
0.8
1.0
0.0
0.5
1.0
1.5
2.0
2.5
3.0
Prob correct diagnosis
Sensitivity
Specificity

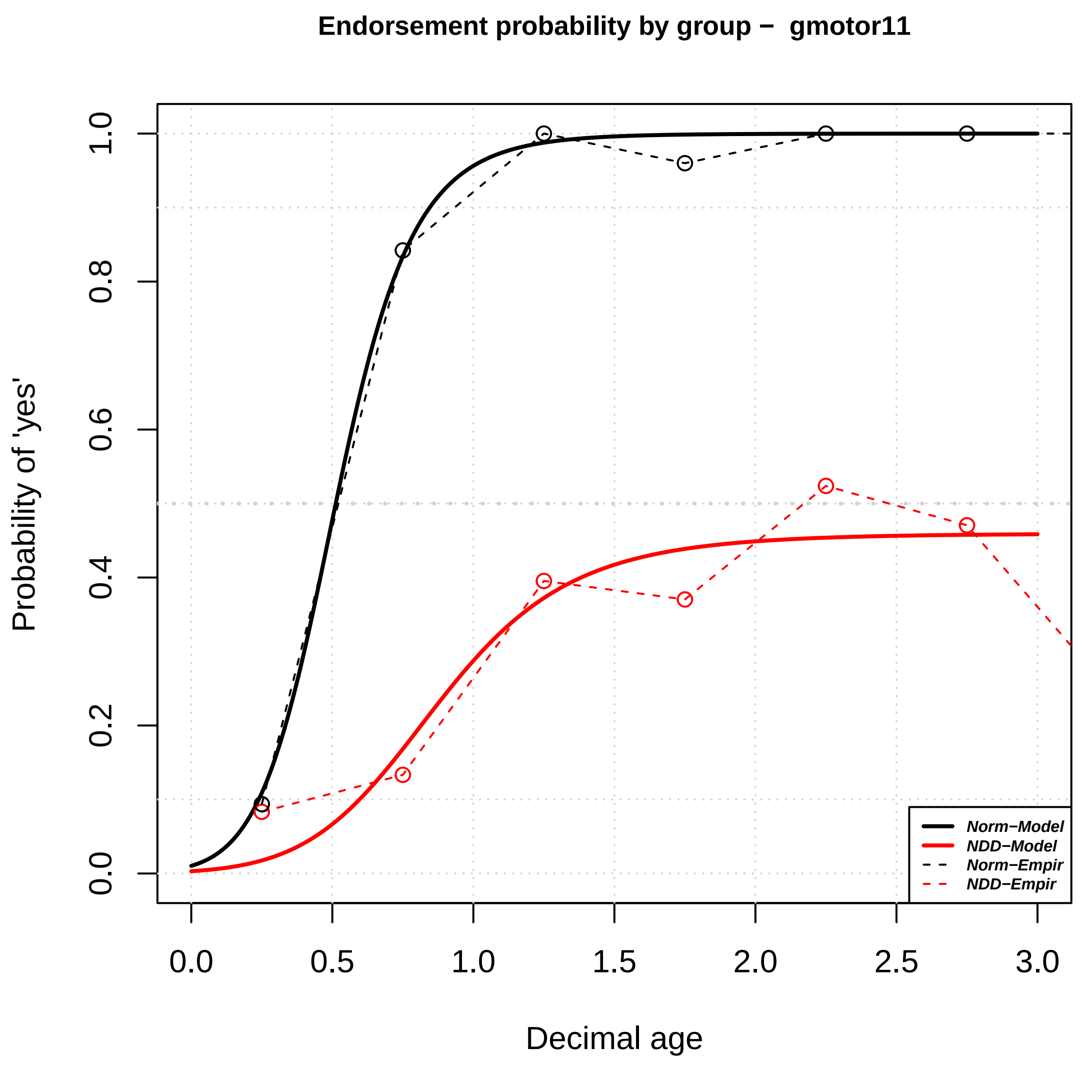

Endorsement probability by group – gmotor11
Probability of 'yes'
Decimal age
0.0
0.2
0.4
0.6
0.8
1.0
0.0
0.5
1.0
1.5
2.0
2.5
3.0
Norm−Model
NDD−Model
Norm−Empir
NDD−Empir


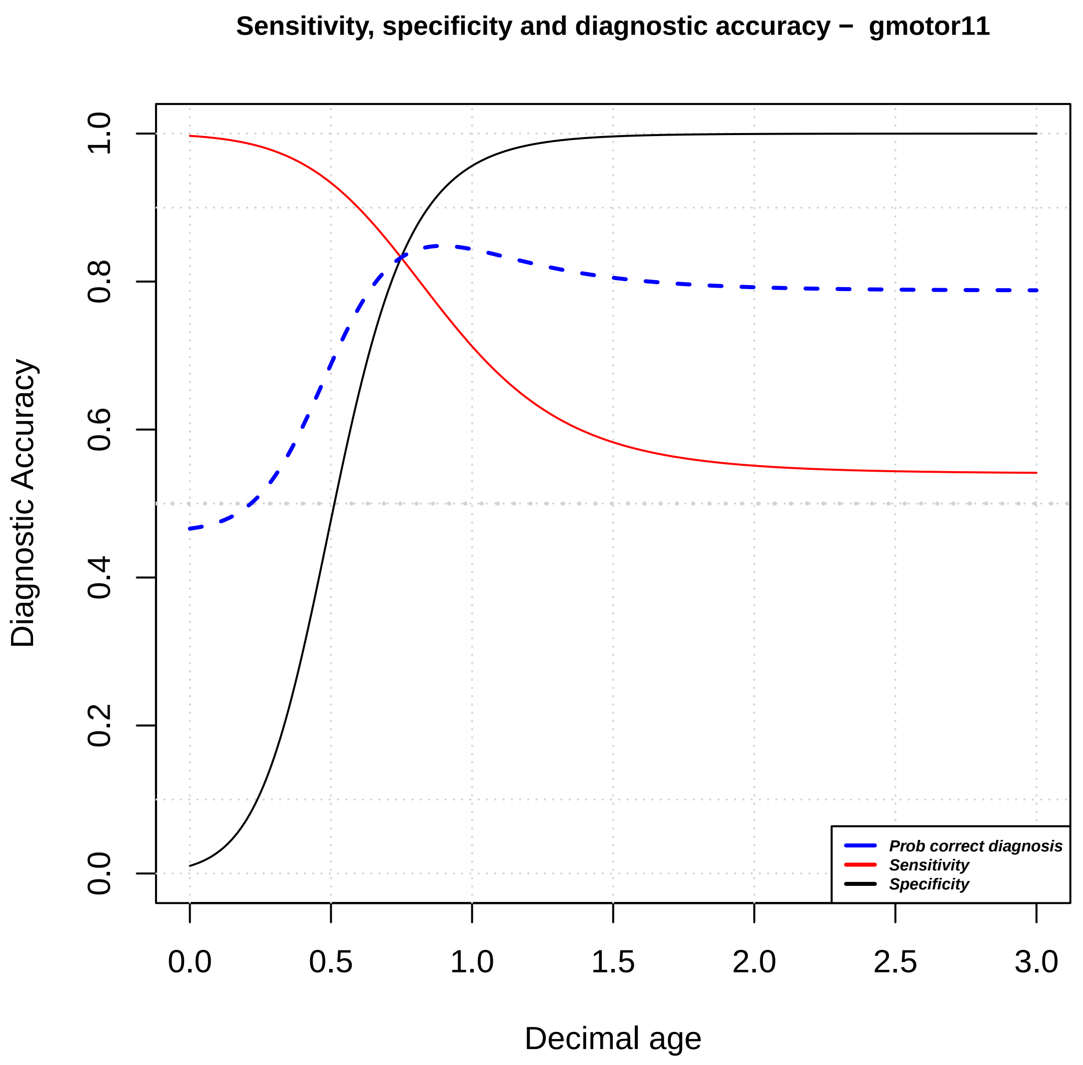

Sensitivity, specificity and diagnostic accuracy – gmotor11
Diagnostic Accuracy
Decimal age
0.0
0.2
0.4
0.6
0.8
1.0
0.0
0.5
1.0
1.5
2.0
2.5
3.0
Prob correct diagnosis
Sensitivity
Specificity

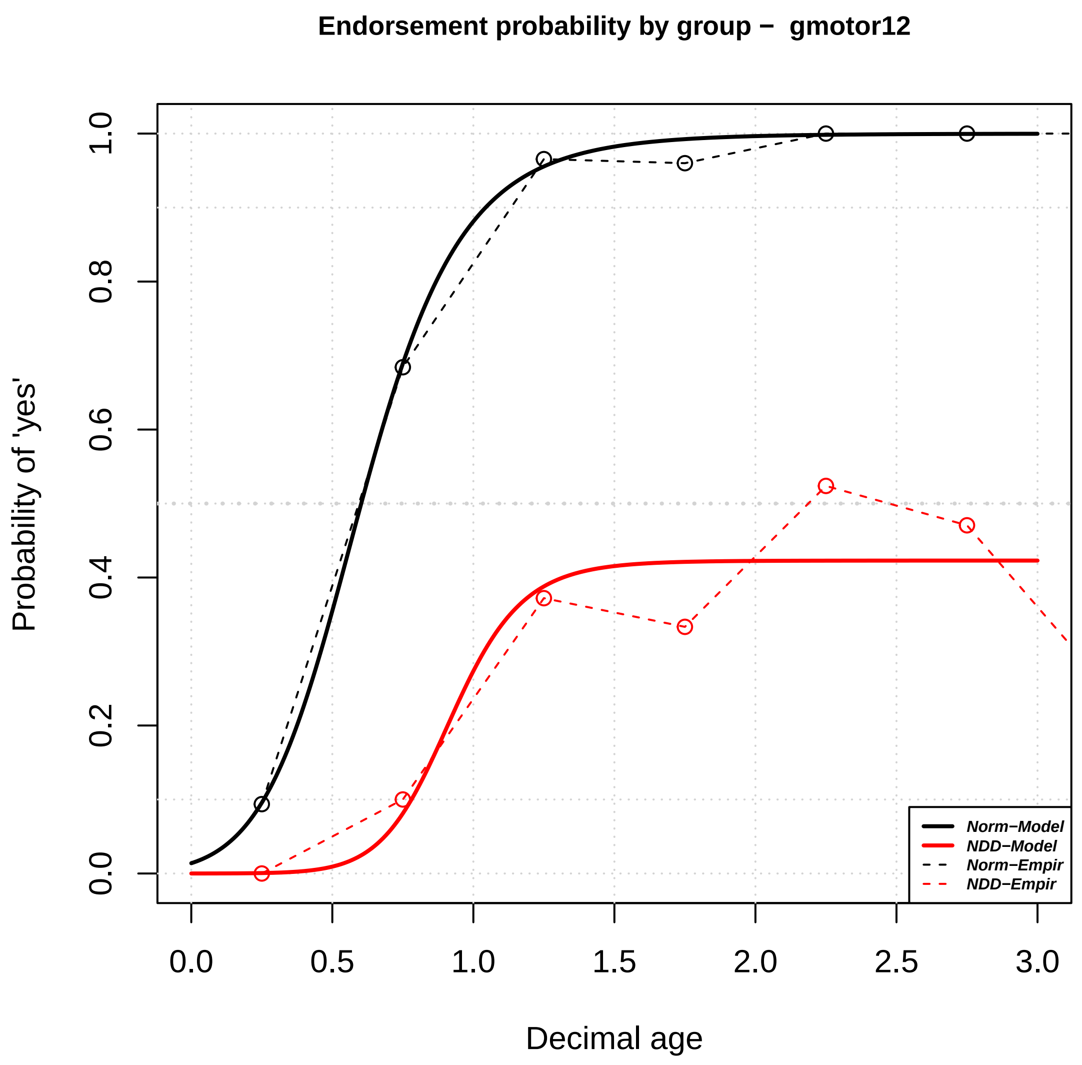

Endorsement probability by group – gmotor12
Probability of 'yes'
Decimal age
0.0
0.2
0.4
0.6
0.8
1.0
0.0
0.5
1.0
1.5
2.0
2.5
3.0
Norm−Model
NDD−Model
Norm−Empir
NDD−Empir


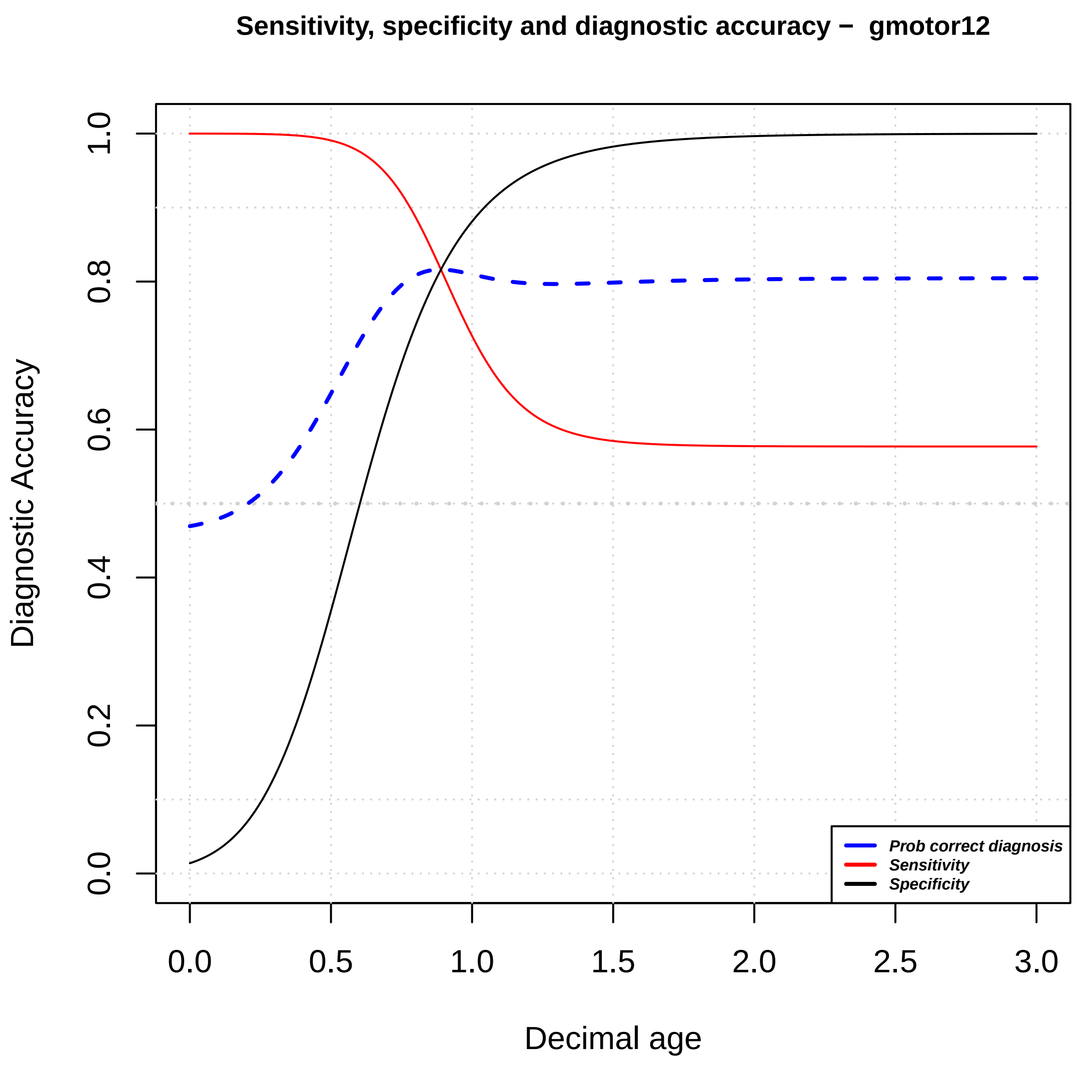

Sensitivity, specificity and diagnostic accuracy – gmotor12
Diagnostic Accuracy
Decimal age
0.0
0.2
0.4
0.6
0.8
1.0
0.0
0.5
1.0
1.5
2.0
2.5
3.0
Prob correct diagnosis
Sensitivity
Specificity

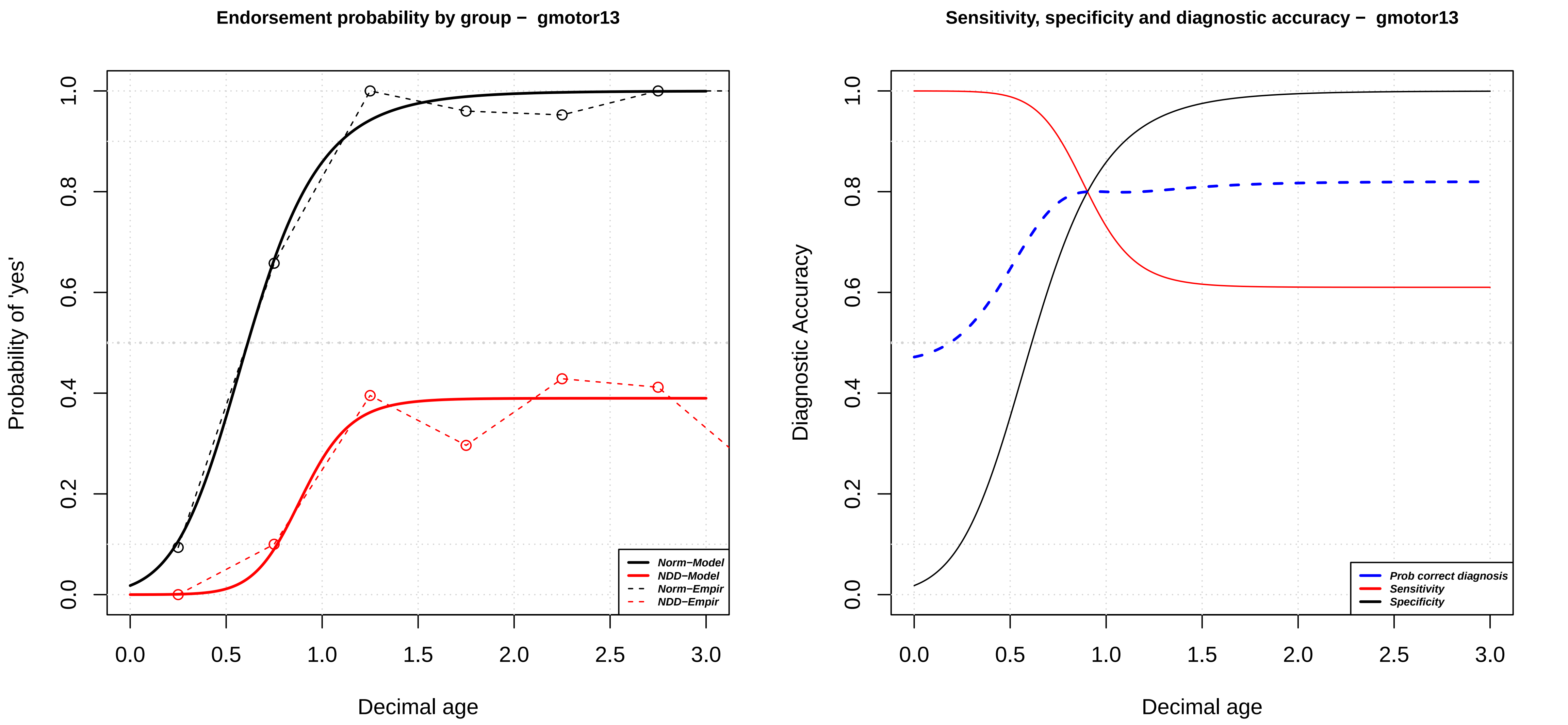
Endorsement probability by group – gmotor13
Probability of 'yes'
Decimal age
Norm–Model
NDD–Model
Norm–Empir
NDD–Empir
Sensitivity, specificity and diagnostic accuracy – gmotor13
Diagnostic Accuracy
Decimal age
Prob correct diagnosis
Sensitivity
Specificity

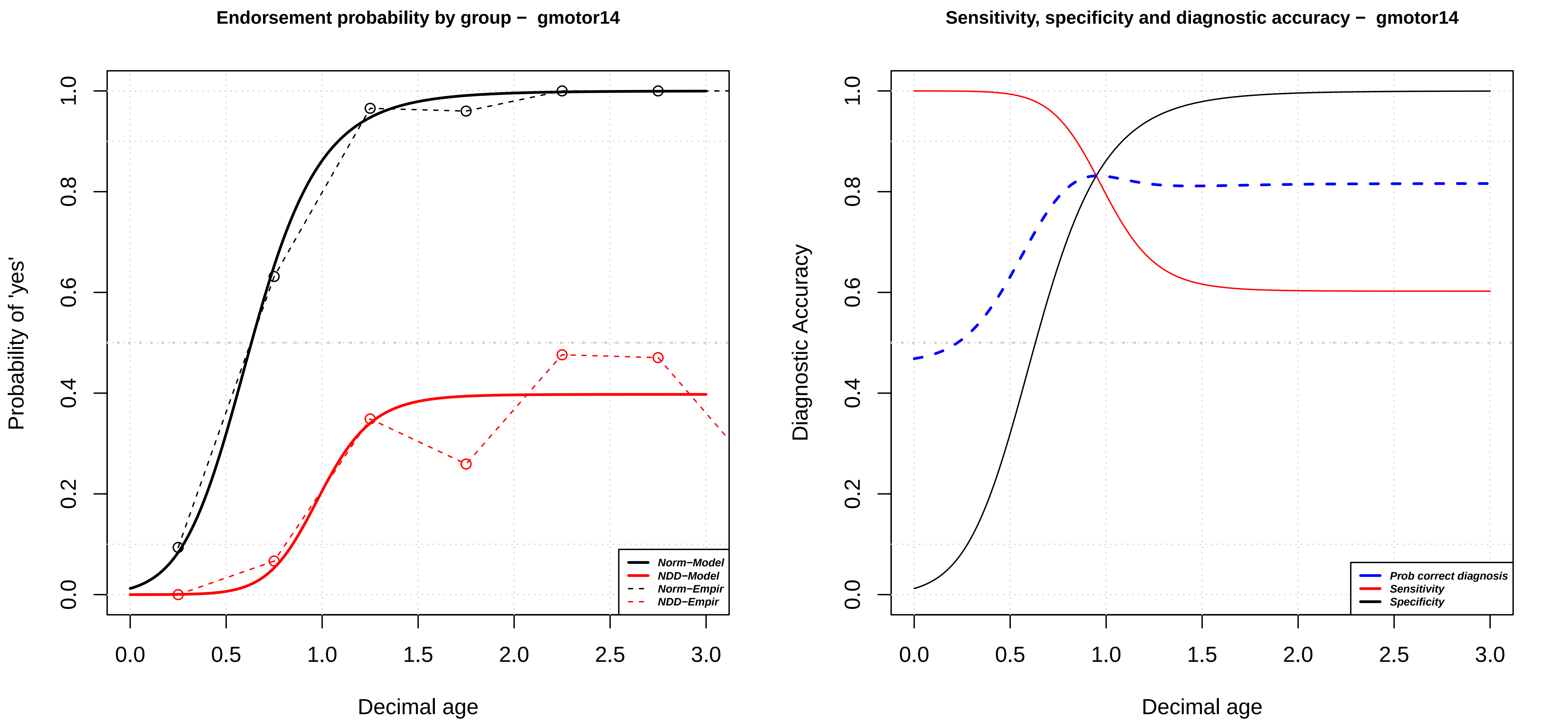
Endorsement probability by group − gmotor14
Probability of 'yes'
Decimal age
Norm−Model
NDD−Model
Norm−Empir
NDD−Empir
0.0
0.2
0.4
0.6
0.8
1.0
0.0
0.5
1.0
1.5
2.0
2.5
3.0
Sensitivity, specificity and diagnostic accuracy − gmotor14
Diagnostic Accuracy
Decimal age
Prob correct diagnosis
Sensitivity
Specificity

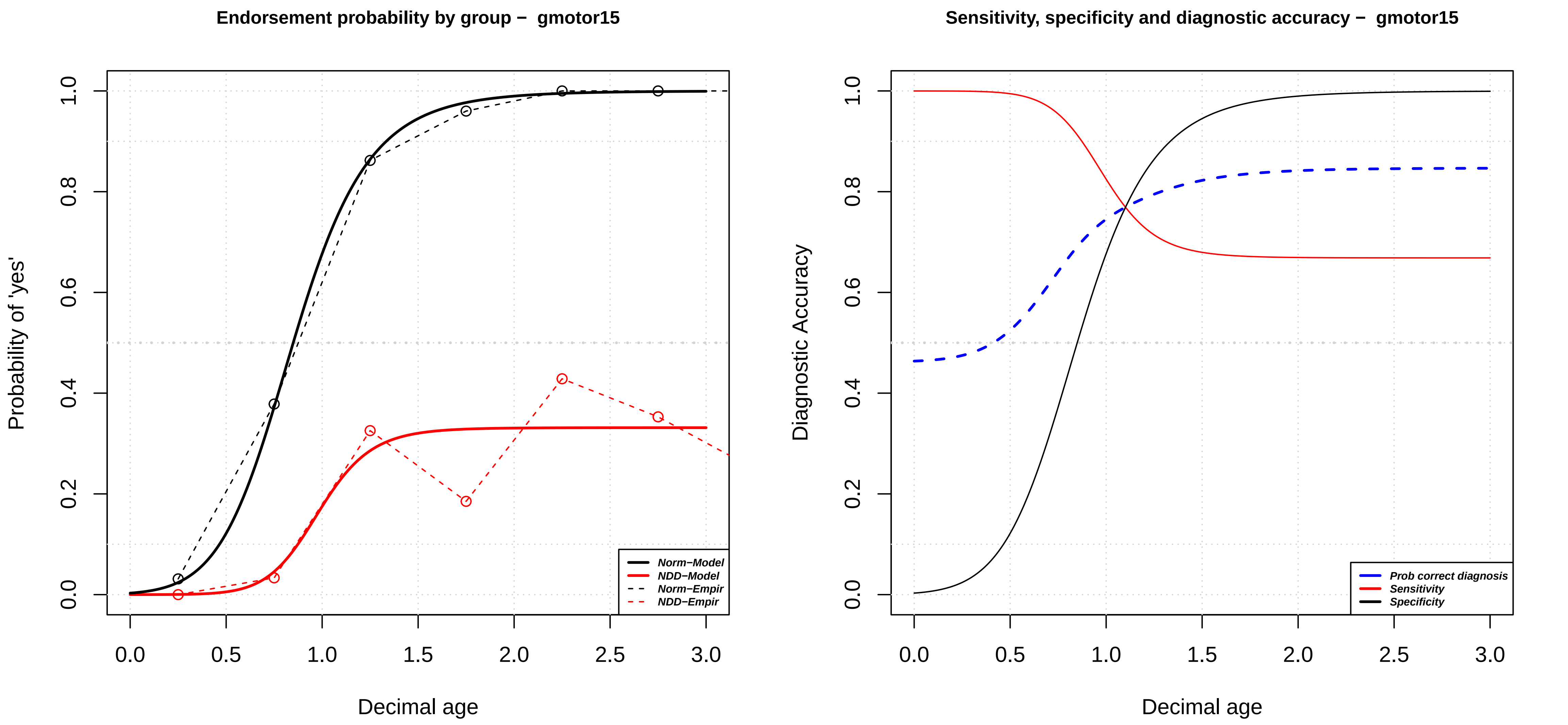
Endorsement probability by group – gmotor15
Probability of 'yes'
Decimal age
Norm−Model
NDD−Model
Norm−Empir
NDD−Empir
Sensitivity, specificity and diagnostic accuracy – gmotor15
Diagnostic Accuracy
Decimal age
Prob correct diagnosis
Sensitivity
Specificity

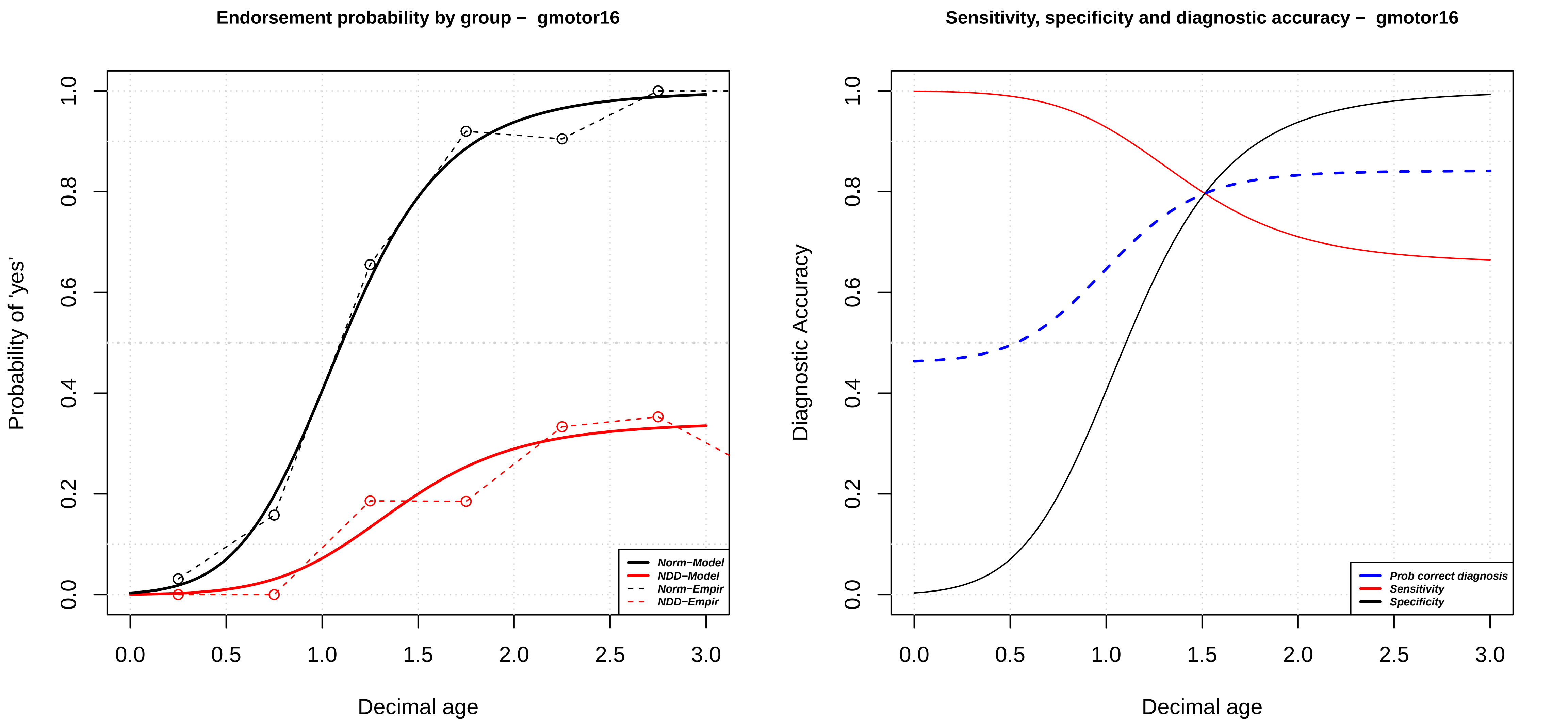

Endorsement probability by group – gmotor16
Probability of 'yes'
Decimal age
Norm−Model
NDD−Model
Norm−Empir
NDD−Empir
0.0
0.2
0.4
0.6
0.8
1.0
0.0
0.5
1.0
1.5
2.0
2.5
3.0
Sensitivity, specificity and diagnostic accuracy – gmotor16
Diagnostic Accuracy
Decimal age
Prob correct diagnosis
Sensitivity
Specificity

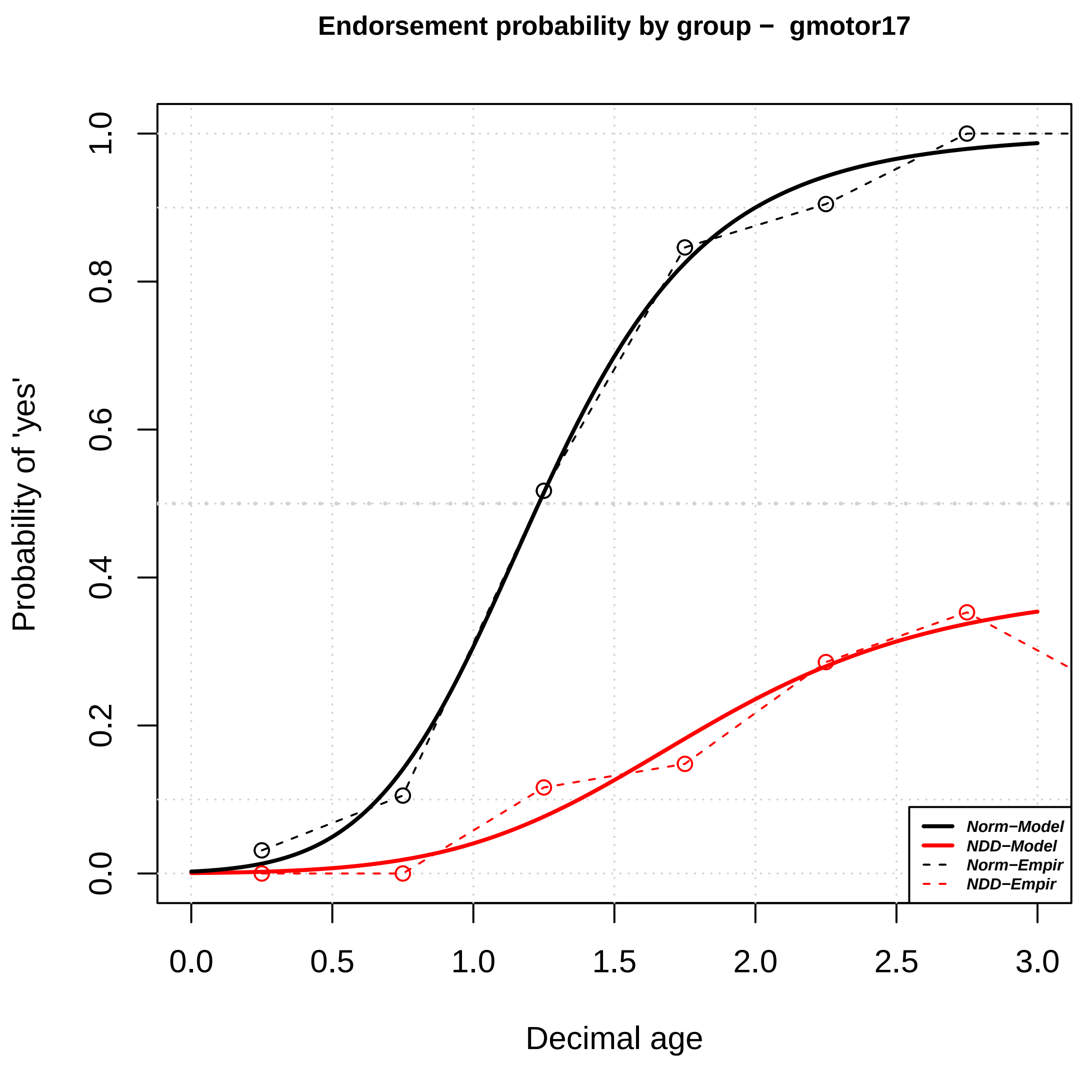

Endorsement probability by group − gmotor17
Probability of 'yes'
Decimal age
0.0
0.2
0.4
0.6
0.8
1.0
0.0
0.5
1.0
1.5
2.0
2.5
3.0
Norm−Model
NDD−Model
Norm−Empir
NDD−Empir


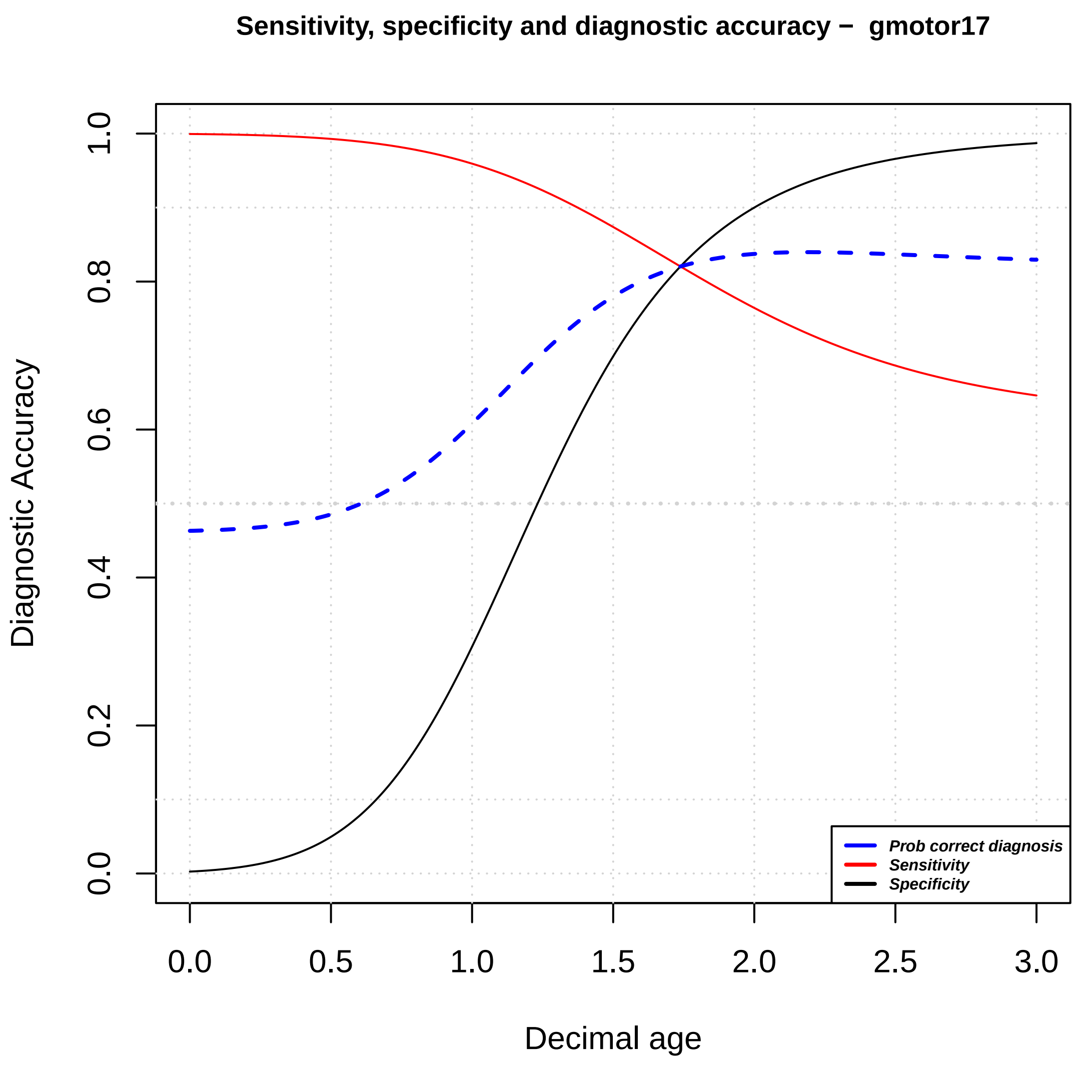

Sensitivity, specificity and diagnostic accuracy − gmotor17
Diagnostic Accuracy
Decimal age
0.0
0.2
0.4
0.6
0.8
1.0
0.0
0.5
1.0
1.5
2.0
2.5
3.0
Prob correct diagnosis
Sensitivity
Specificity

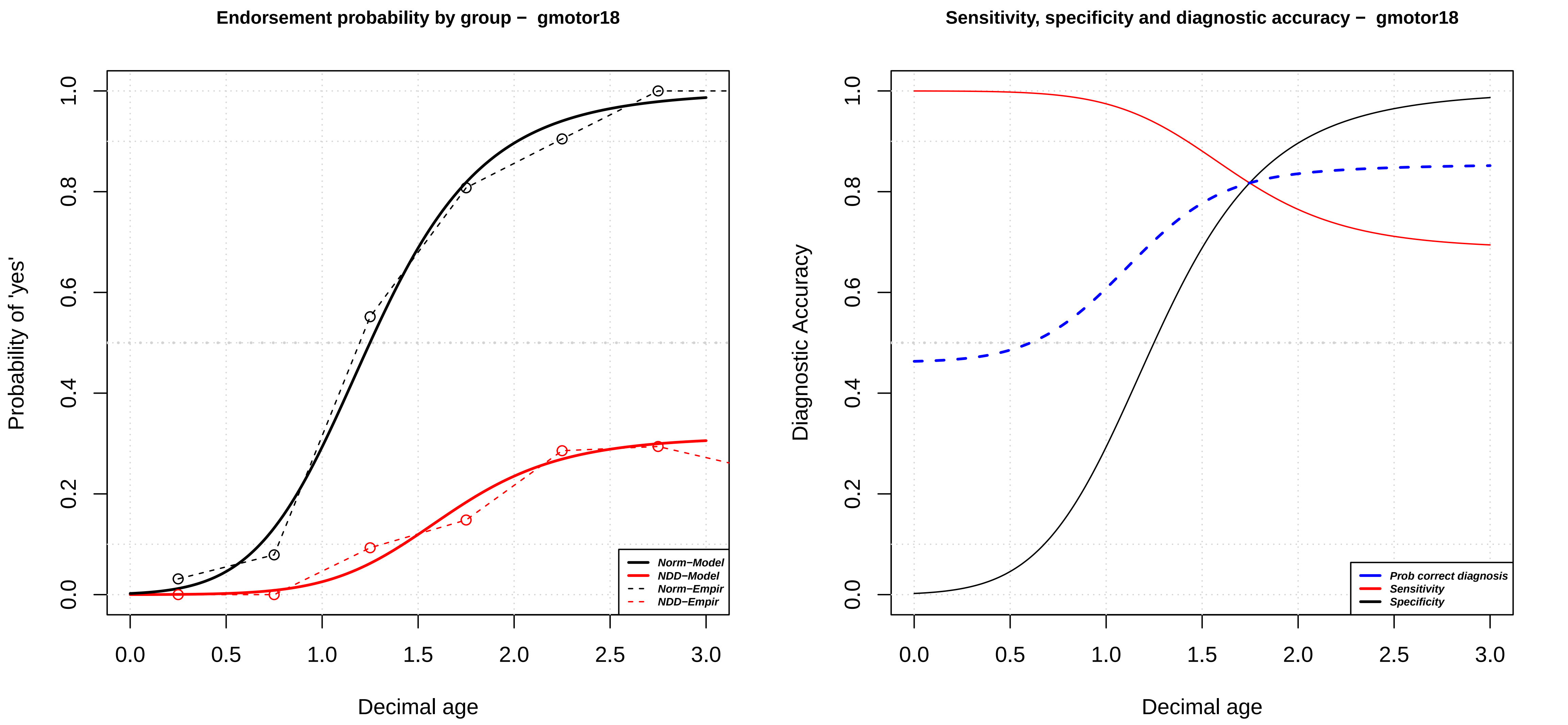

Endorsement probability by group − gmotor18
Probability of 'yes'
Decimal age
0.0
0.2
0.4
0.6
0.8
1.0
0.0
0.5
1.0
1.5
2.0
2.5
3.0
Norm−Model
NDD−Model
Norm−Empir
NDD−Empir
Sensitivity, specificity and diagnostic accuracy − gmotor18
Diagnostic Accuracy
Decimal age
Prob correct diagnosis
Sensitivity
Specificity

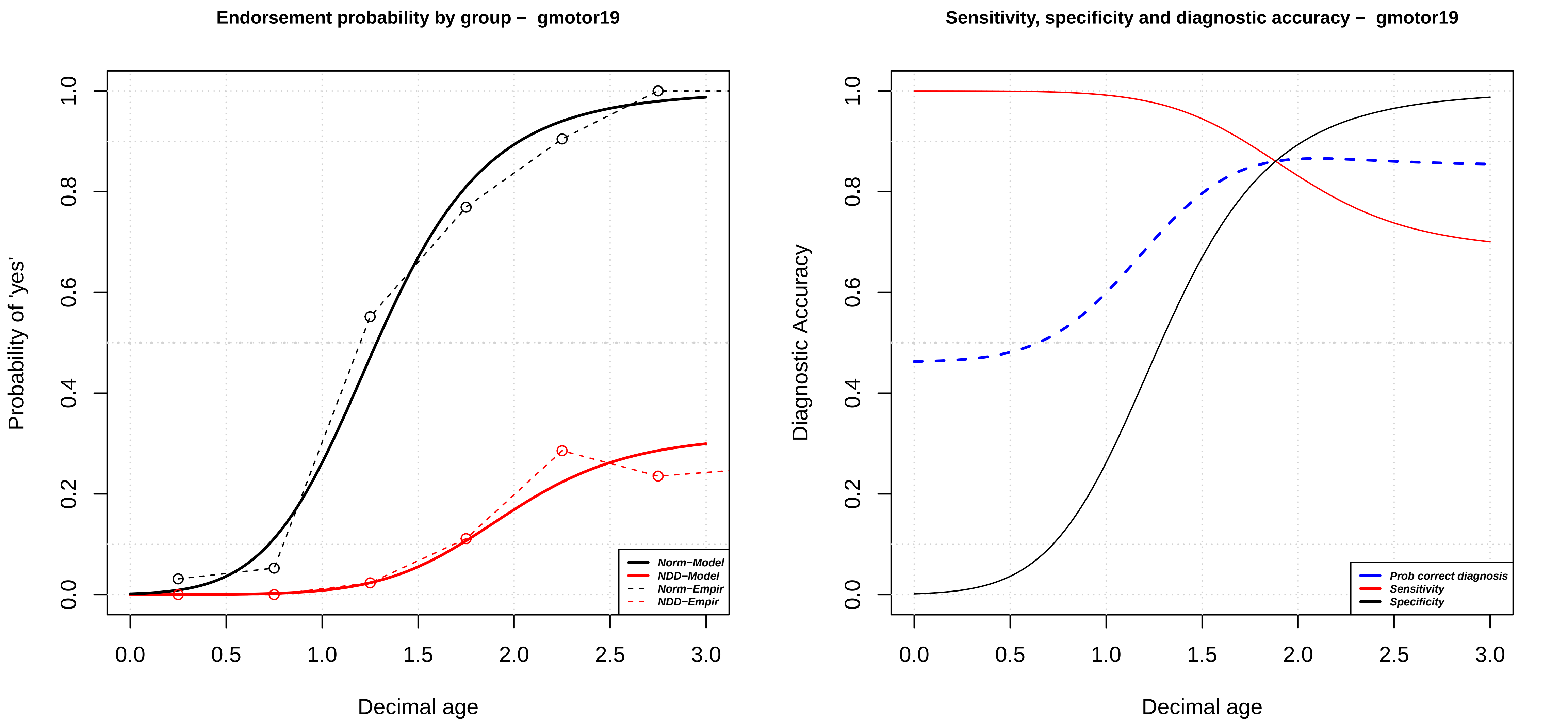

Endorsement probability by group – gmotor19
Probability of 'yes'
Decimal age
Norm−Model
NDD−Model
Norm−Empir
NDD−Empir
Sensitivity, specificity and diagnostic accuracy – gmotor19
Diagnostic Accuracy
Decimal age
Prob correct diagnosis
Sensitivity
Specificity

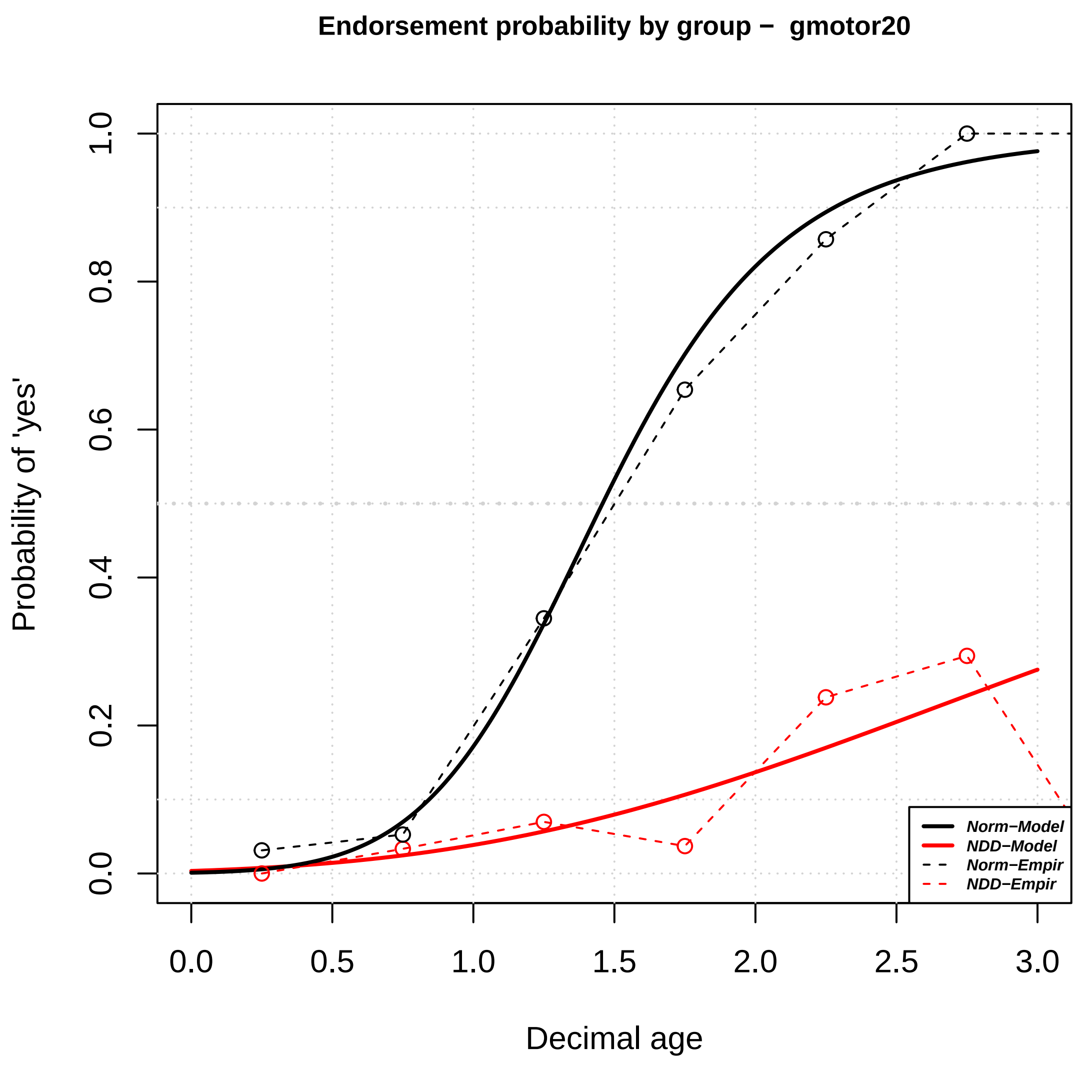
Endorsement probability by group − gmotor20
Probability of 'yes'
Decimal age
0.0
0.2
0.4
0.6
0.8
1.0
0.0
0.5
1.0
1.5
2.0
2.5
3.0
Norm−Model
NDD−Model
Norm−Empir
NDD−Empir

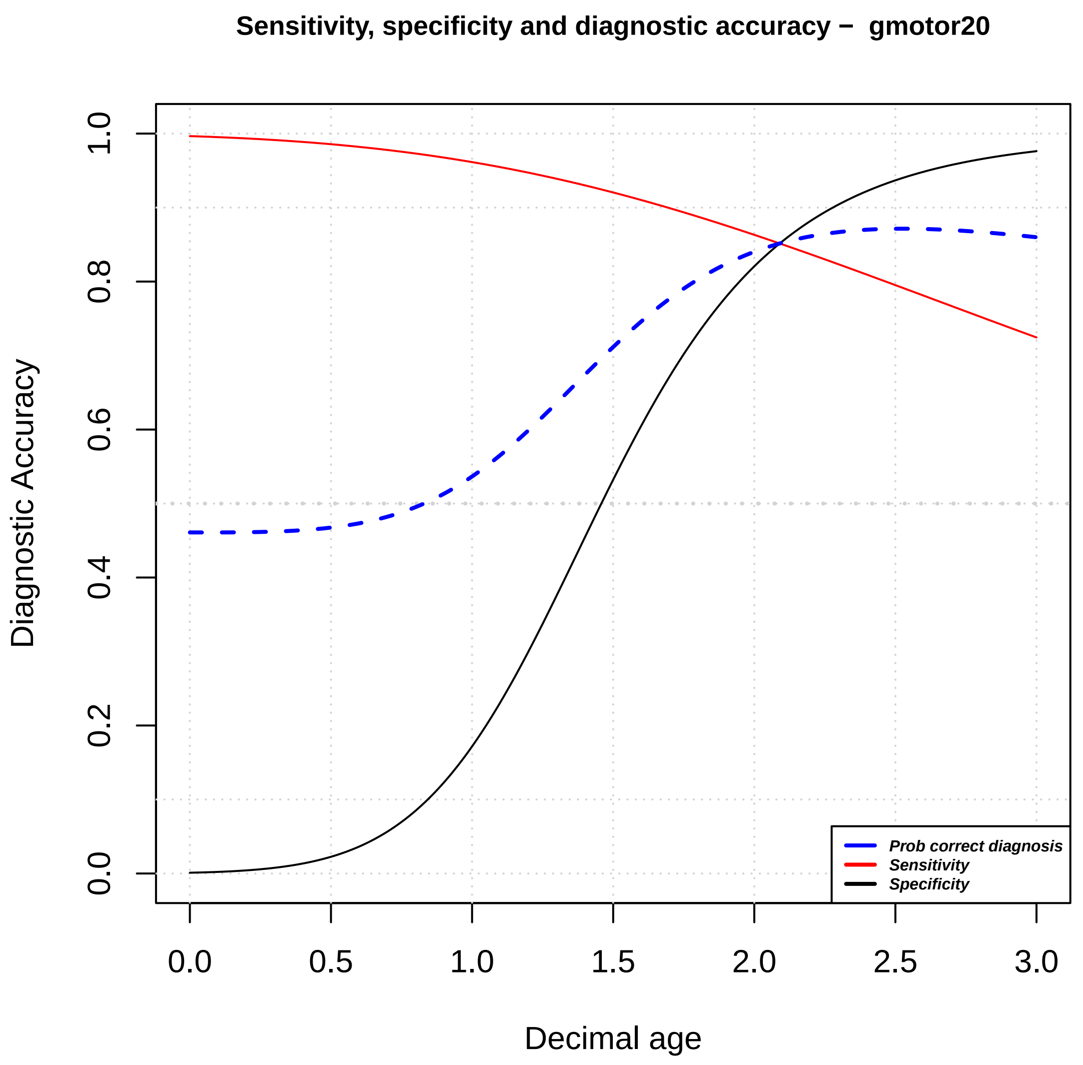
Sensitivity, specificity and diagnostic accuracy − gmotor20
Diagnostic Accuracy
Decimal age
0.0
0.2
0.4
0.6
0.8
1.0
0.0
0.5
1.0
1.5
2.0
2.5
3.0
Prob correct diagnosis
Sensitivity
Specificity

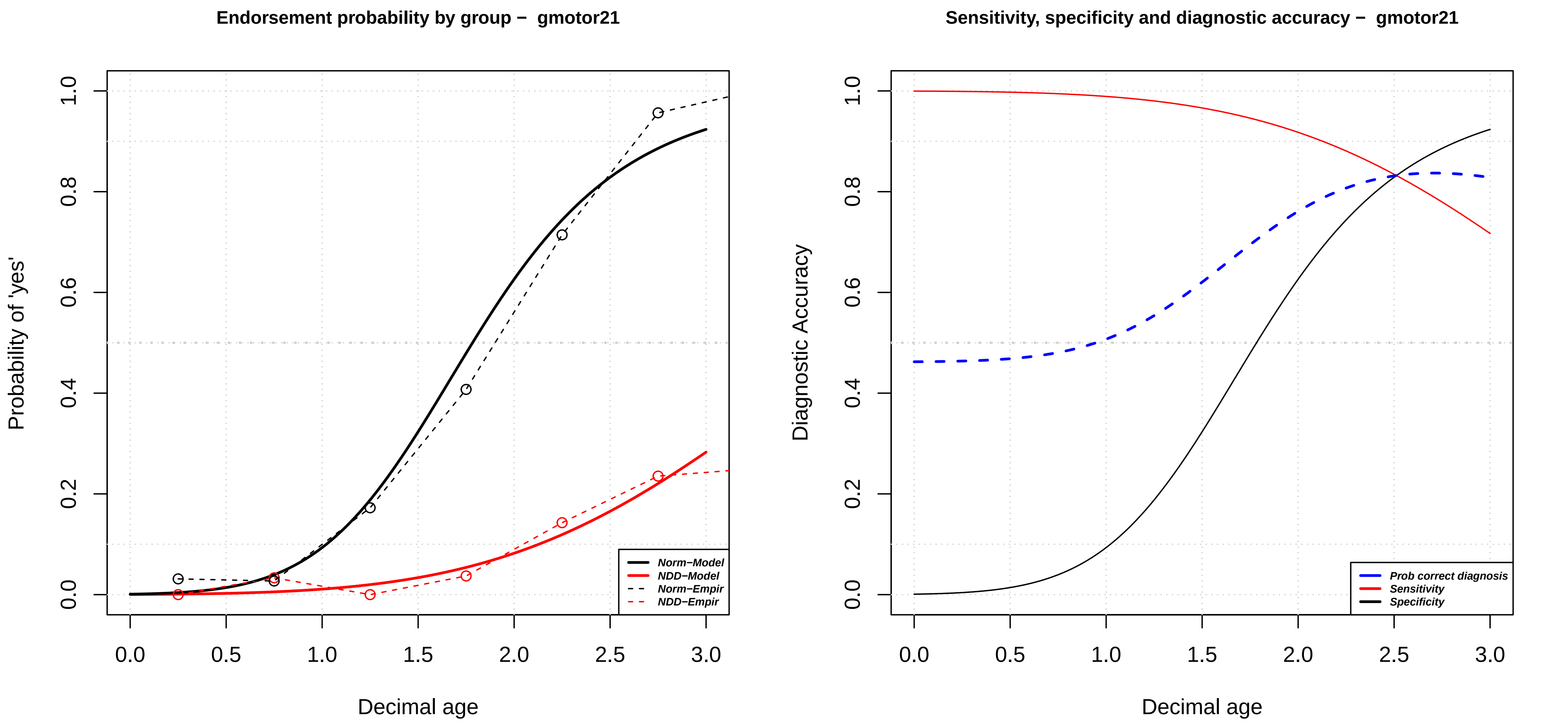

Endorsement probability by group – gmotor21
Probability of 'yes'
Decimal age
Norm−Model
NDD−Model
Norm−Empir
NDD−Empir
0.0
0.2
0.4
0.6
0.8
1.0
0.0
0.5
1.0
1.5
2.0
2.5
3.0
Sensitivity, specificity and diagnostic accuracy – gmotor21
Diagnostic Accuracy
Decimal age
Prob correct diagnosis
Sensitivity
Specificity

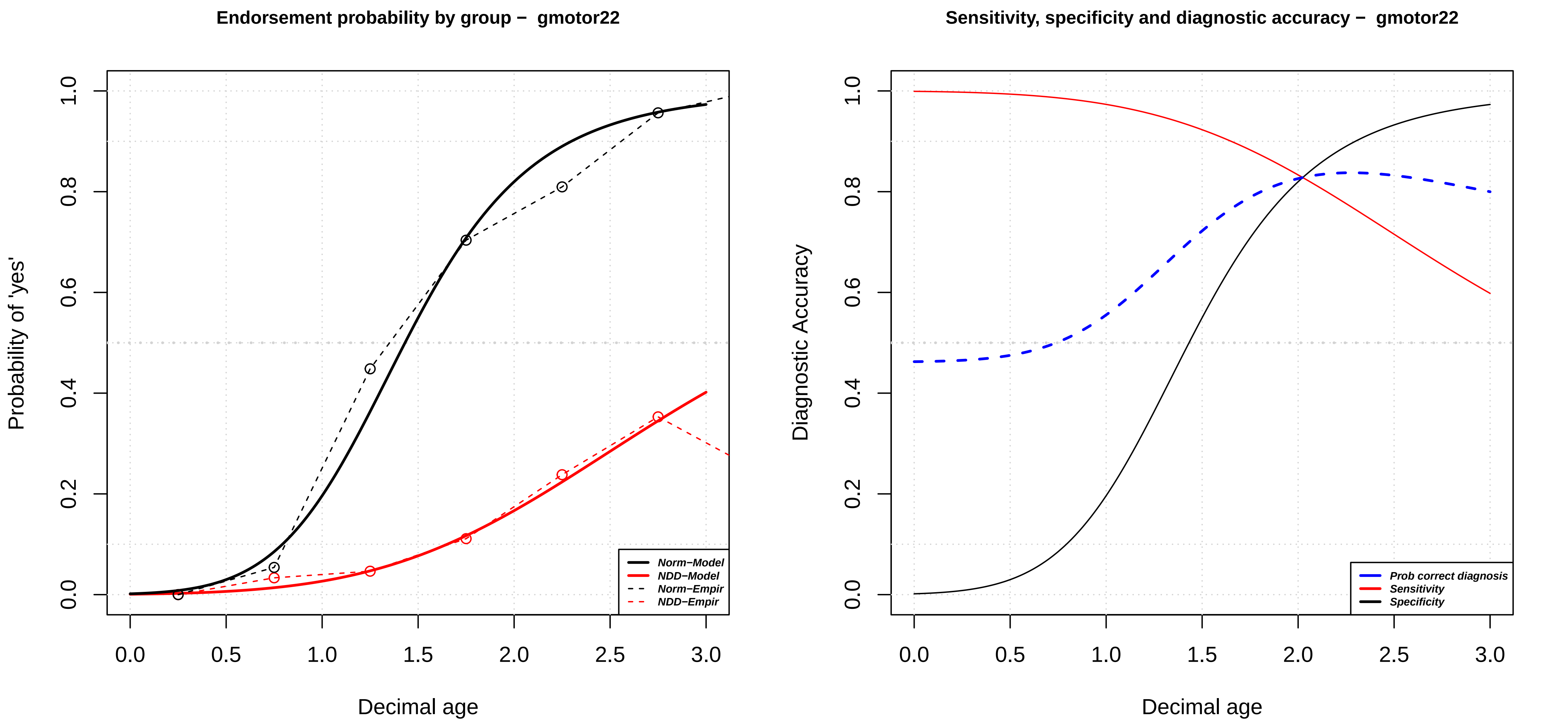

Endorsement probability by group − gmotor22
Probability of 'yes'
Decimal age
Norm−Model
NDD−Model
Norm−Empir
NDD−Empir
Sensitivity, specificity and diagnostic accuracy − gmotor22
Diagnostic Accuracy
Decimal age
Prob correct diagnosis
Sensitivity
Specificity

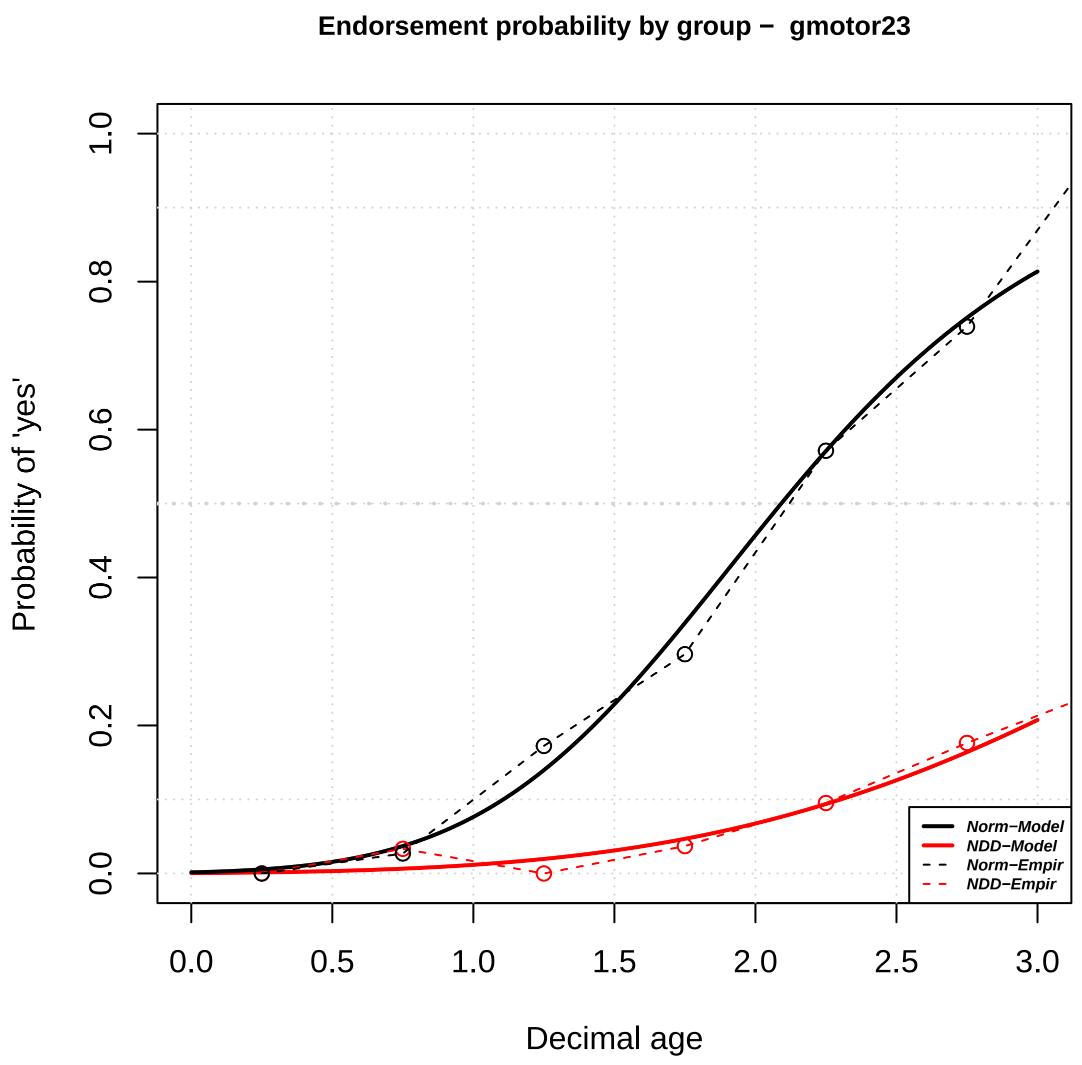
Endorsement probability by group − gmotor23
Probability of 'yes'
Decimal age
0.0
0.2
0.4
0.6
0.8
1.0
0.0
0.5
1.0
1.5
2.0
2.5
3.0
Norm−Model
NDD−Model
Norm−Empir
NDD−Empir

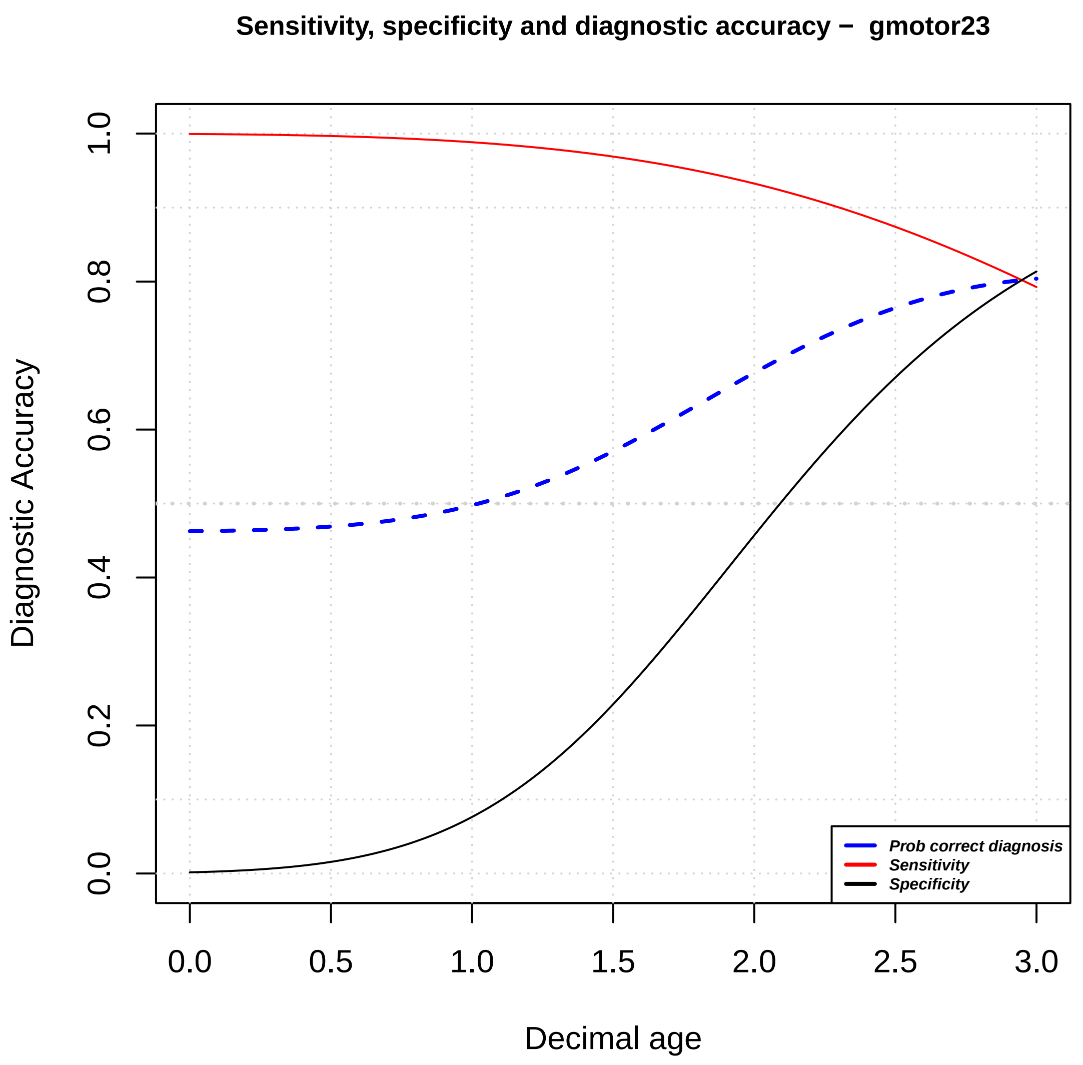
Sensitivity, specificity and diagnostic accuracy − gmotor23
Diagnostic Accuracy
Decimal age
0.0
0.2
0.4
0.6
0.8
1.0
0.0
0.5
1.0
1.5
2.0
2.5
3.0
Prob correct diagnosis
Sensitivity
Specificity

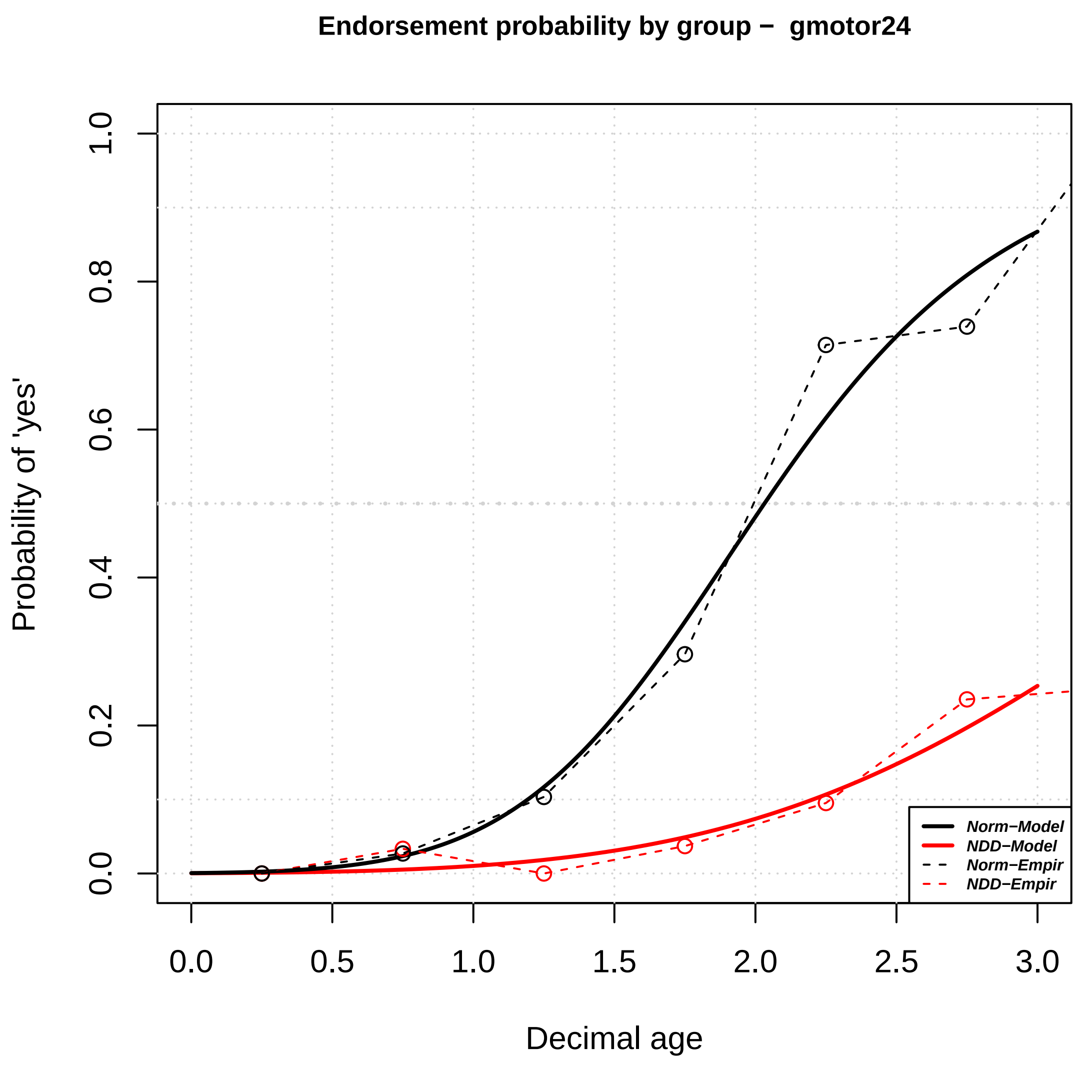

Endorsement probability by group – gmotor24
Probability of 'yes'
Decimal age
0.0
0.2
0.4
0.6
0.8
1.0
0.0
0.5
1.0
1.5
2.0
2.5
3.0
Norm–Model
NDD–Model
Norm–Empir
NDD–Empir


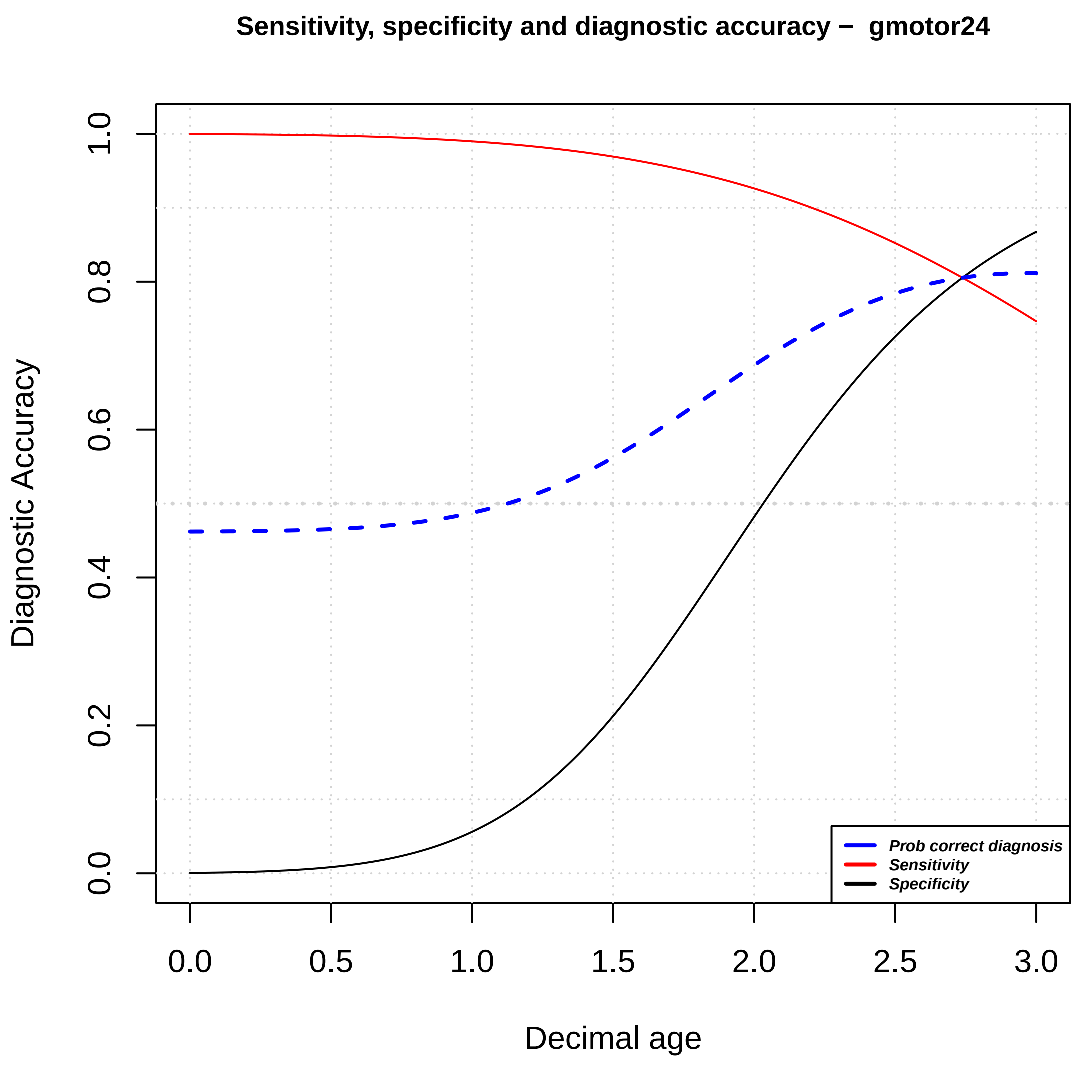

Sensitivity, specificity and diagnostic accuracy – gmotor24
Diagnostic Accuracy
Decimal age
0.0
0.2
0.4
0.6
0.8
1.0
0.0
0.5
1.0
1.5
2.0
2.5
3.0
Prob correct diagnosis
Sensitivity
Specificity

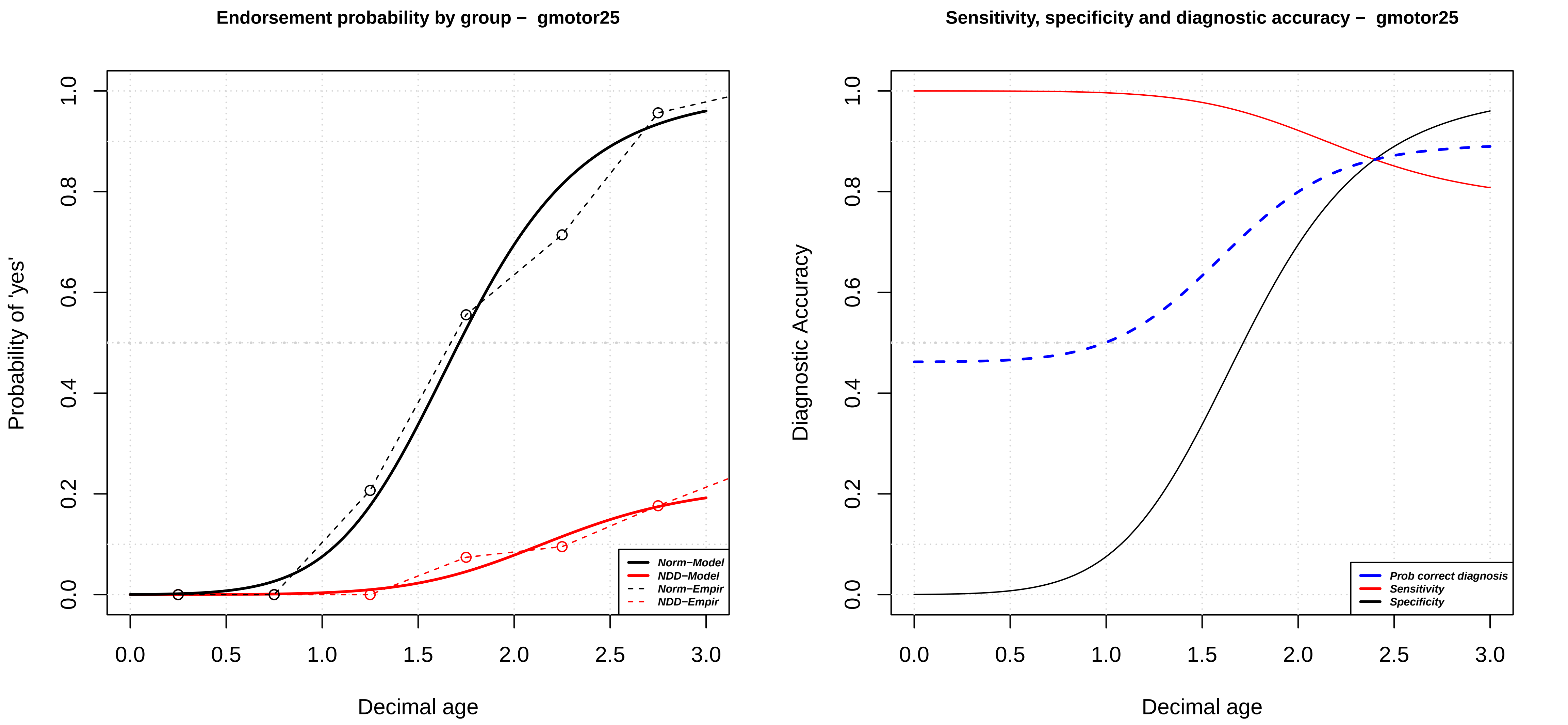
Endorsement probability by group – gmotor25
Probability of 'yes'
Decimal age
Norm−Model
NDD−Model
Norm−Empir
NDD−Empir
Sensitivity, specificity and diagnostic accuracy – gmotor25
Diagnostic Accuracy
Decimal age
Prob correct diagnosis
Sensitivity
Specificity

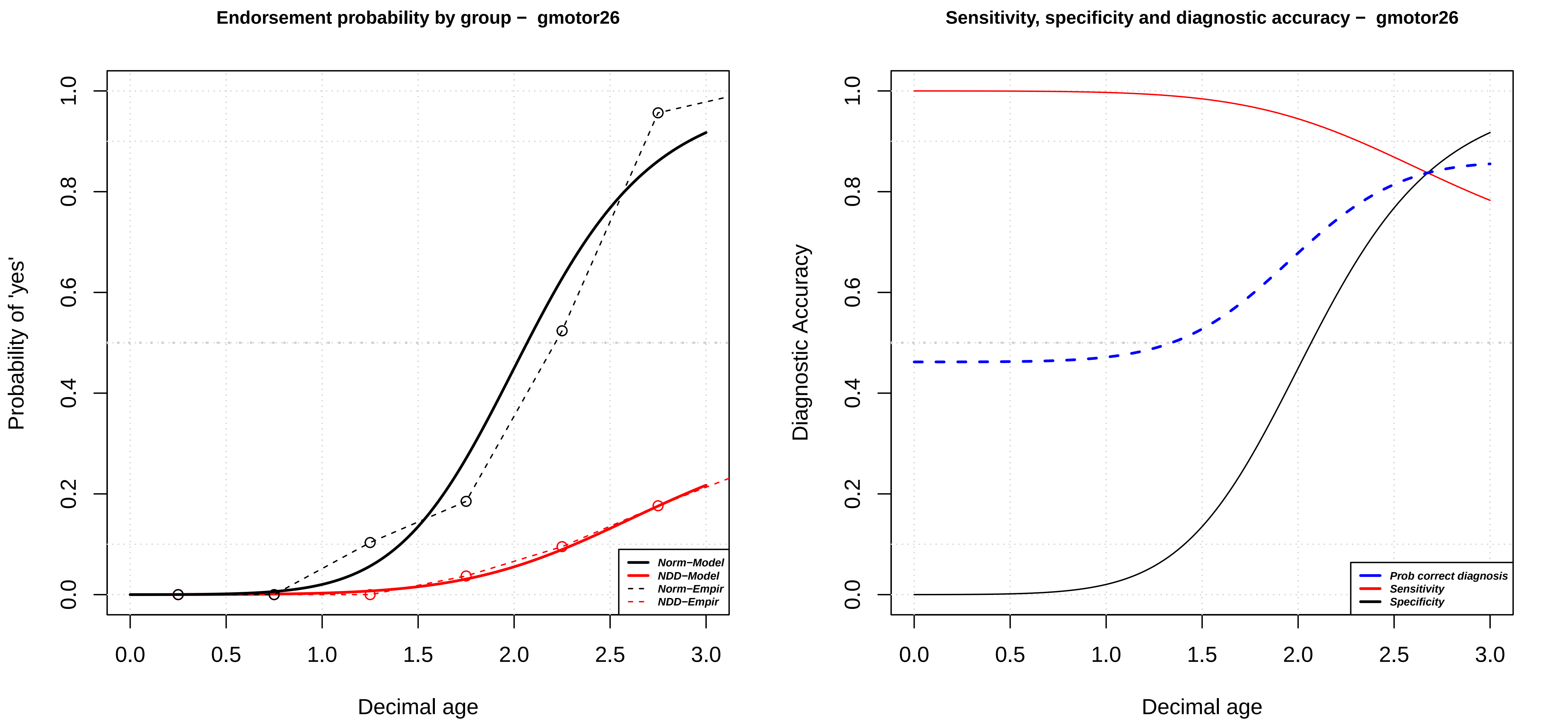
Endorsement probability by group − gmotor26
Probability of 'yes'
Decimal age
0.0
0.2
0.4
0.6
0.8
1.0
0.0
0.5
1.0
1.5
2.0
2.5
3.0
Norm−Model
NDD−Model
Norm−Empir
NDD−Empir
Sensitivity, specificity and diagnostic accuracy − gmotor26
Diagnostic Accuracy
Decimal age
Prob correct diagnosis
Sensitivity
Specificity

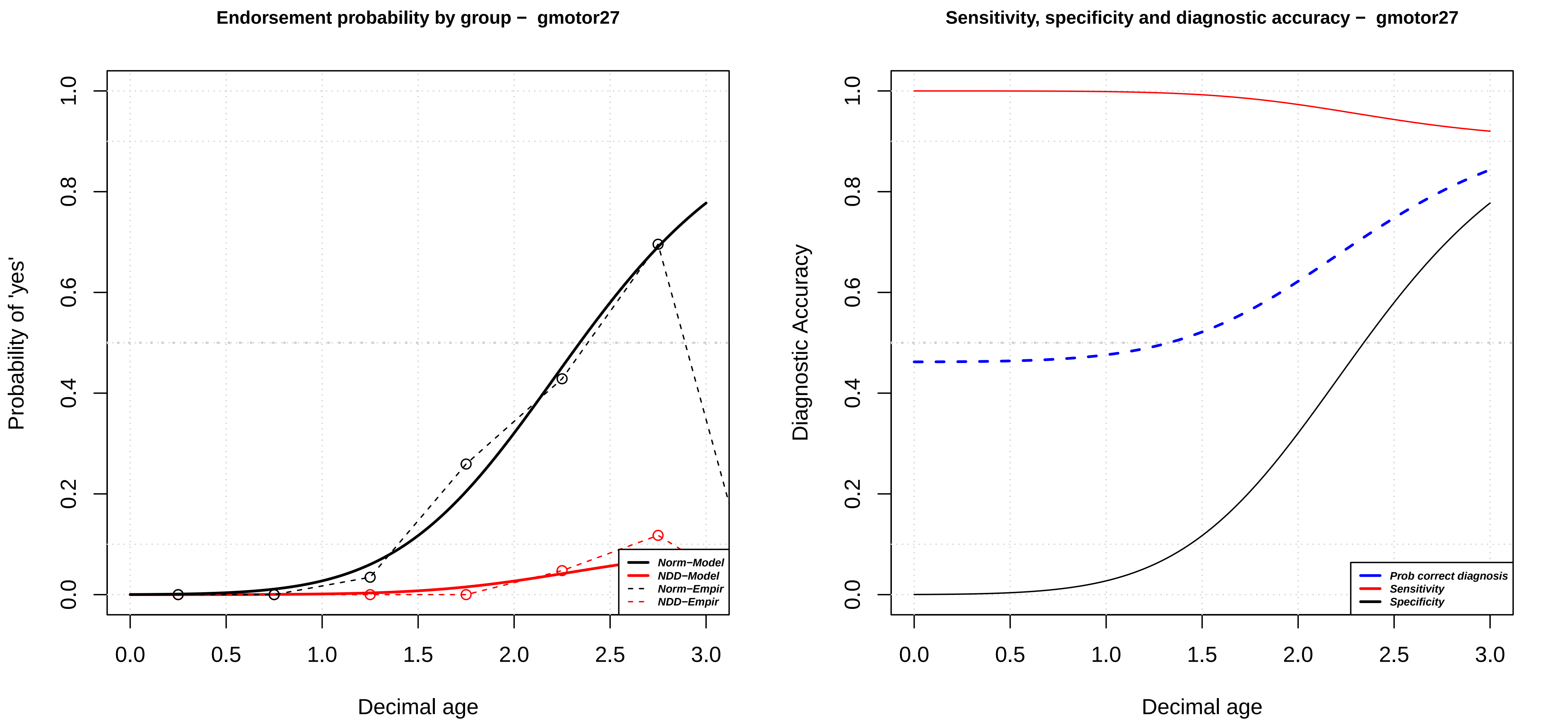

Endorsement probability by group – gmotor27
Probability of 'yes'
Decimal age
0.0
0.2
0.4
0.6
0.8
1.0
0.0
0.5
1.0
1.5
2.0
2.5
3.0
Norm−Model
NDD−Model
Norm−Empir
NDD−Empir
Sensitivity, specificity and diagnostic accuracy – gmotor27
Diagnostic Accuracy
Decimal age
0.0
0.2
0.4
0.6
0.8
1.0
0.0
0.5
1.0
1.5
2.0
2.5
3.0
Prob correct diagnosis
Sensitivity
Specificity

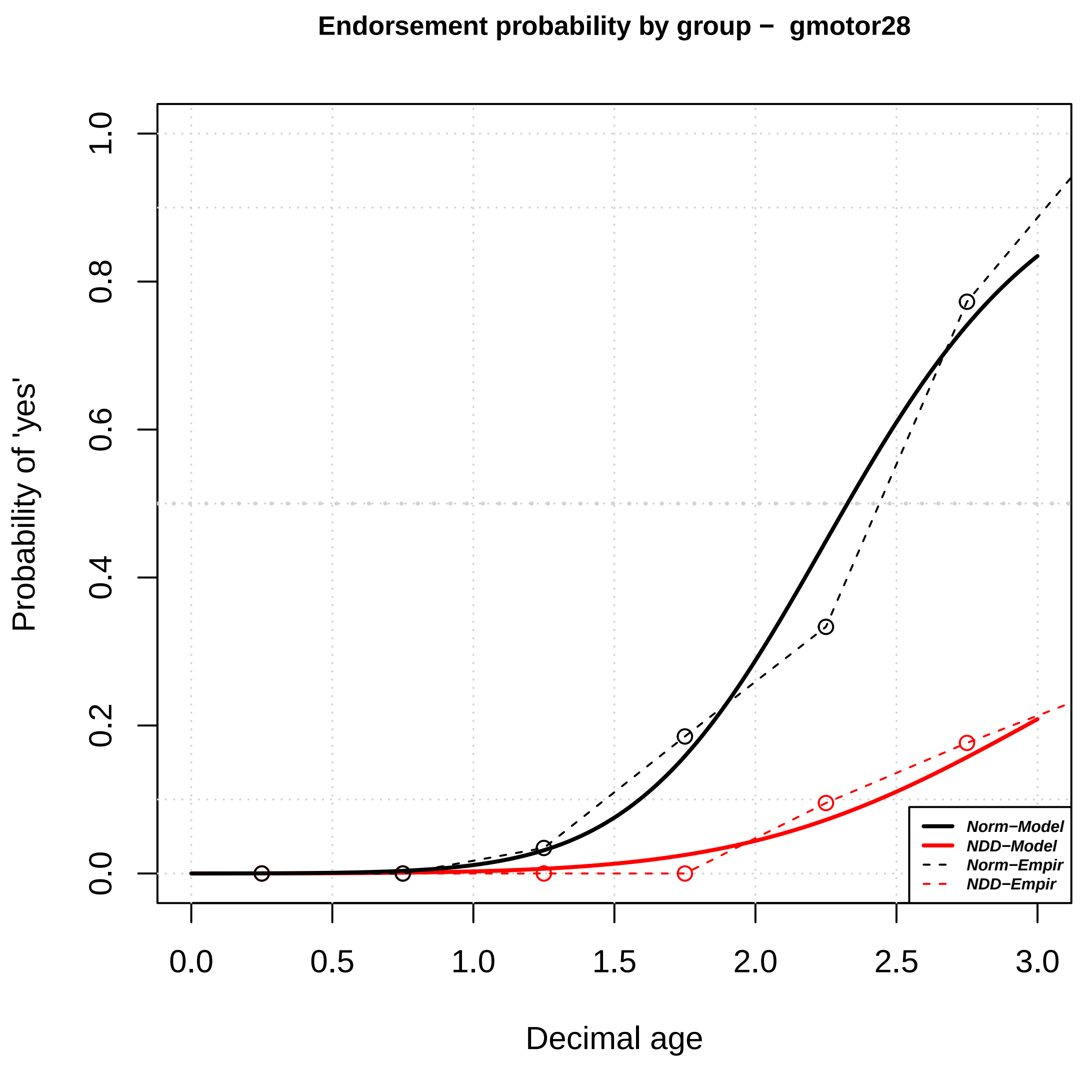
Endorsement probability by group − gmotor28
Probability of 'yes'
Decimal age
0.0 0.2 0.4 0.6 0.8 1.0
0.0 0.5 1.0 1.5 2.0 2.5 3.0
Norm−Model
NDD−Model
Norm−Empir
NDD−Empir

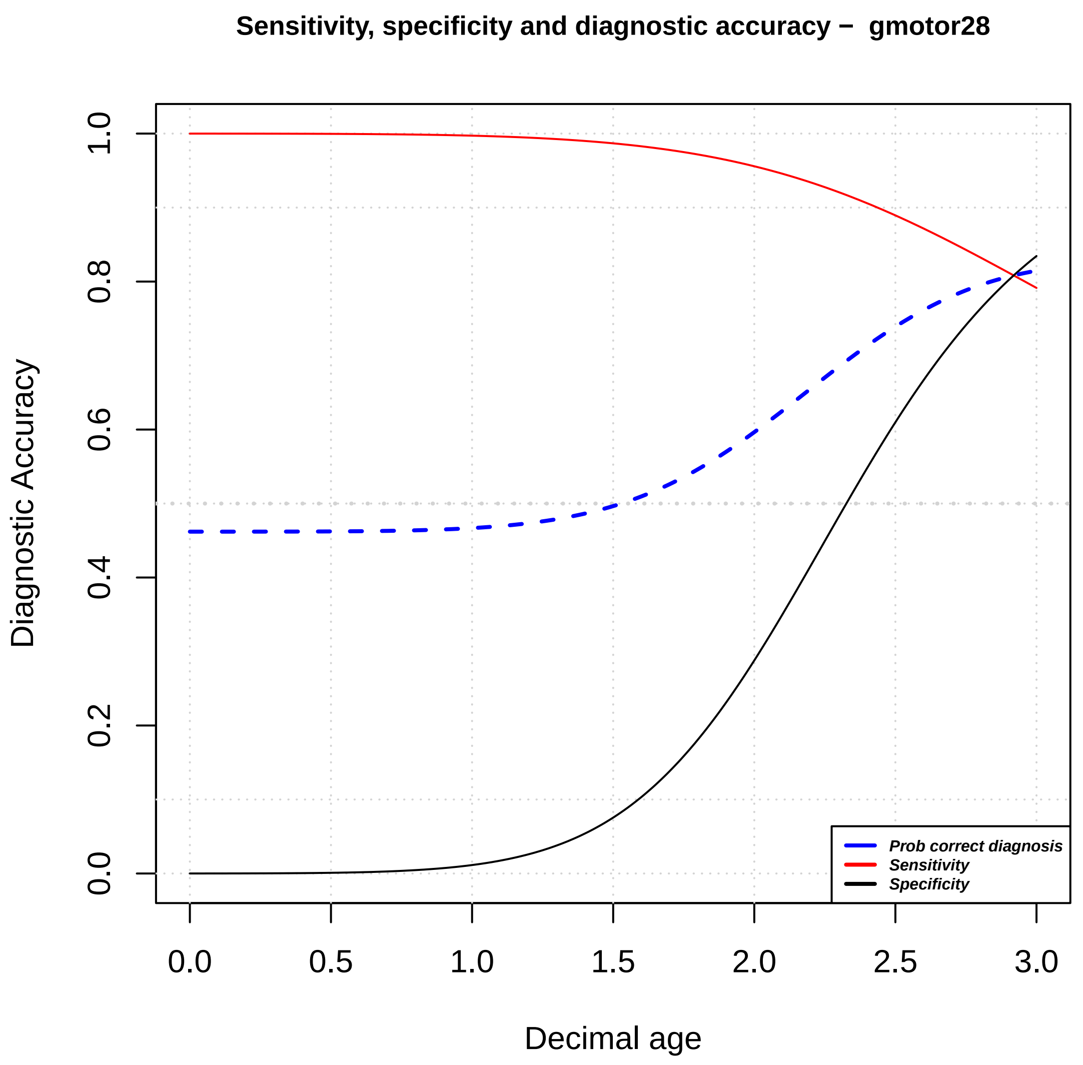
Sensitivity, specificity and diagnostic accuracy − gmotor28
Diagnostic Accuracy
Decimal age
0.0 0.2 0.4 0.6 0.8 1.0
0.0 0.5 1.0 1.5 2.0 2.5 3.0
Prob correct diagnosis
Sensitivity
Specificity

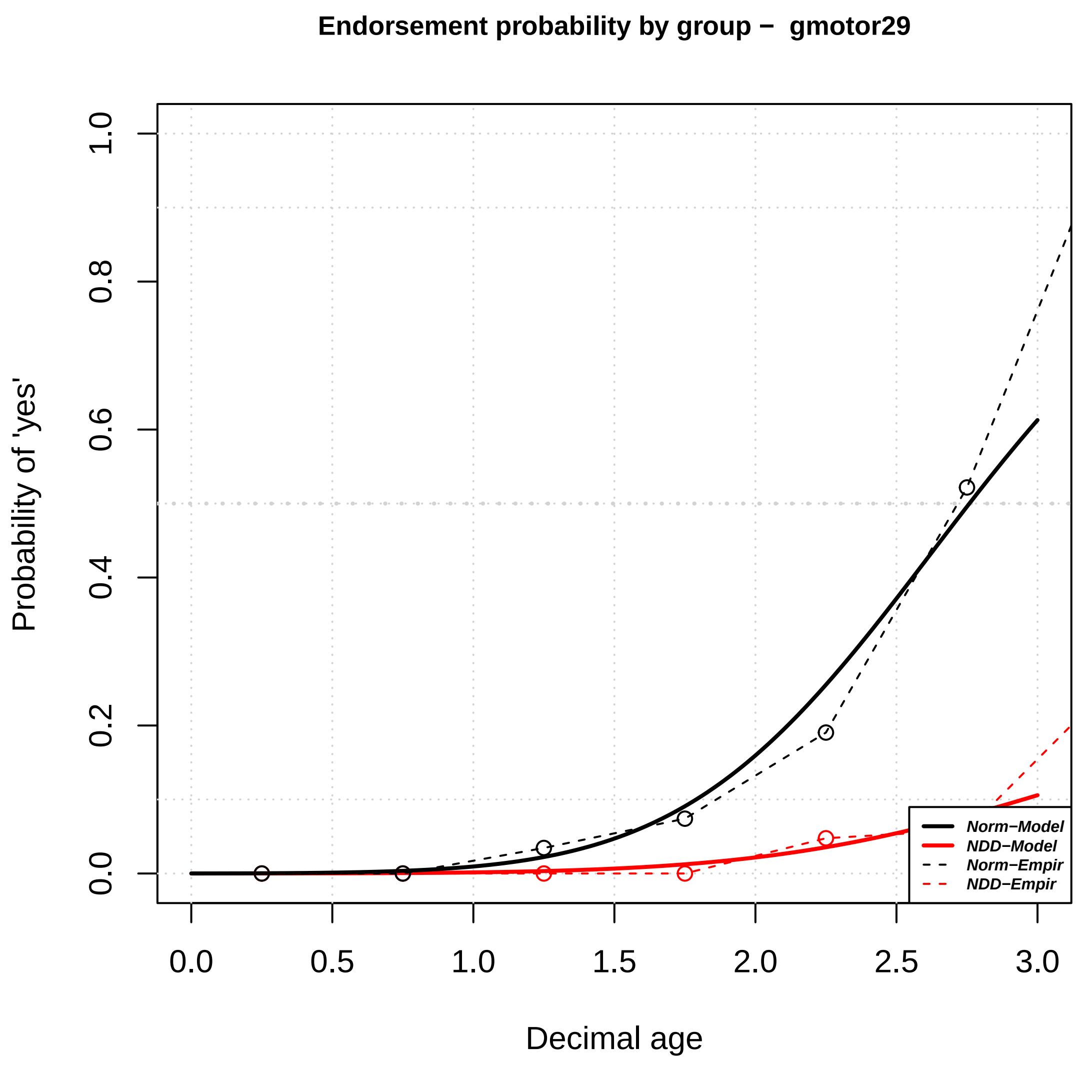

Endorsement probability by group – gmotor29
Probability of 'yes'
Decimal age
0.0
0.2
0.4
0.6
0.8
1.0
0.0
0.5
1.0
1.5
2.0
2.5
3.0
Norm–Model
NDD–Model
Norm–Empir
NDD–Empir


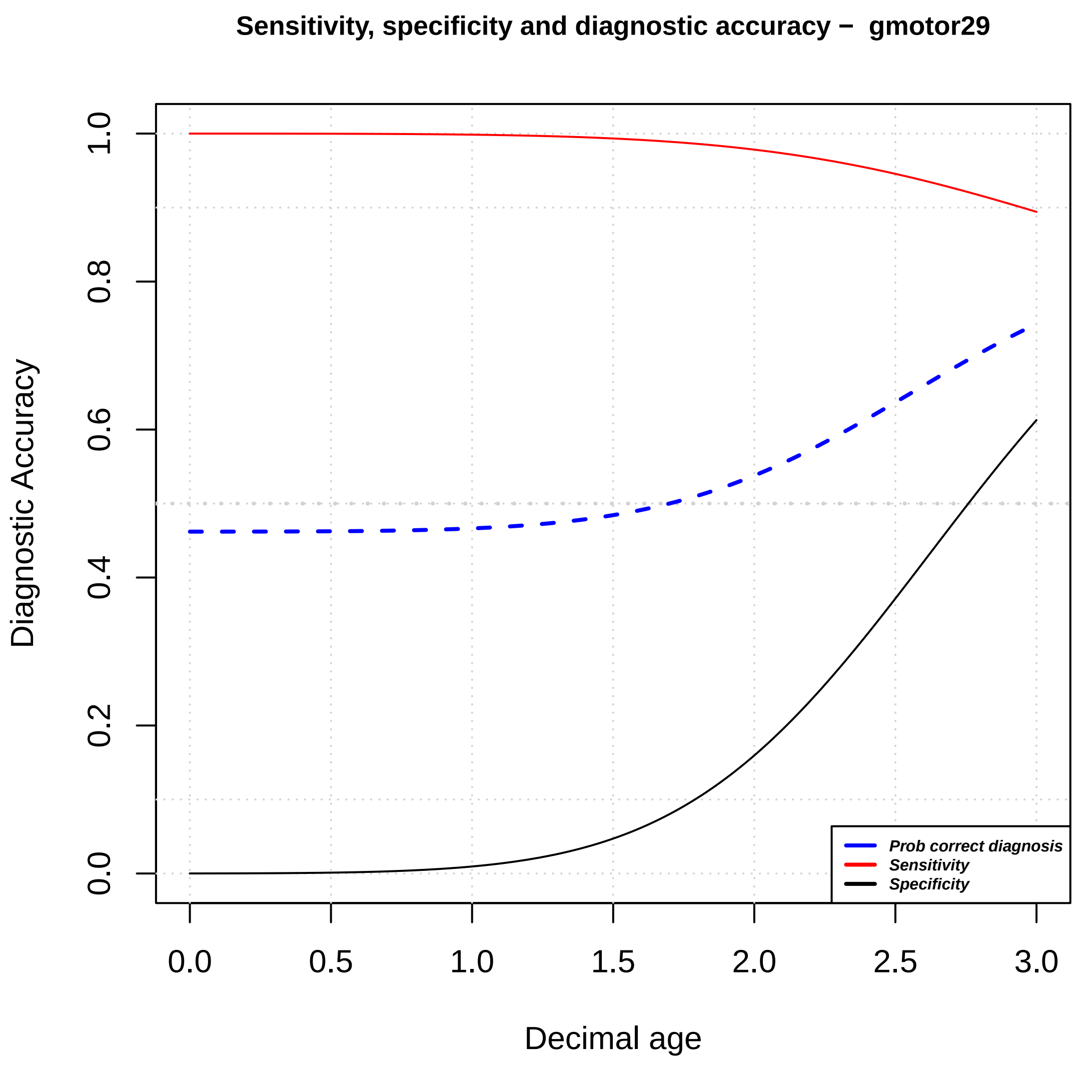

Sensitivity, specificity and diagnostic accuracy – gmotor29
Diagnostic Accuracy
Decimal age
0.0
0.2
0.4
0.6
0.8
1.0
0.0
0.5
1.0
1.5
2.0
2.5
3.0
Prob correct diagnosis
Sensitivity
Specificity

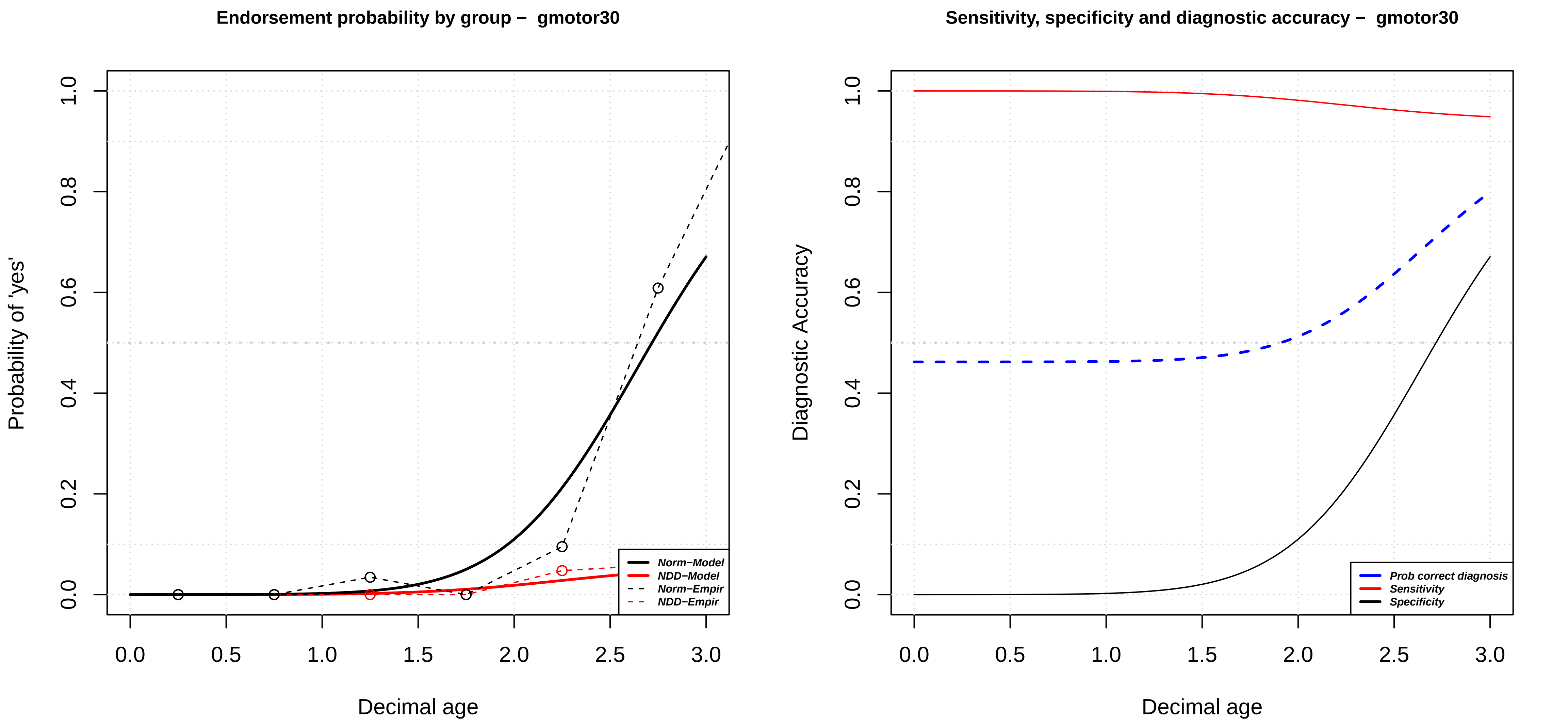
Endorsement probability by group – gmotor30
Probability of 'yes'
Decimal age
0.0
0.2
0.4
0.6
0.8
1.0
0.0
0.5
1.0
1.5
2.0
2.5
3.0
Norm–Model
NDD–Model
Norm–Empir
NDD–Empir
Sensitivity, specificity and diagnostic accuracy – gmotor30
Diagnostic Accuracy
Decimal age
Prob correct diagnosis
Sensitivity
Specificity

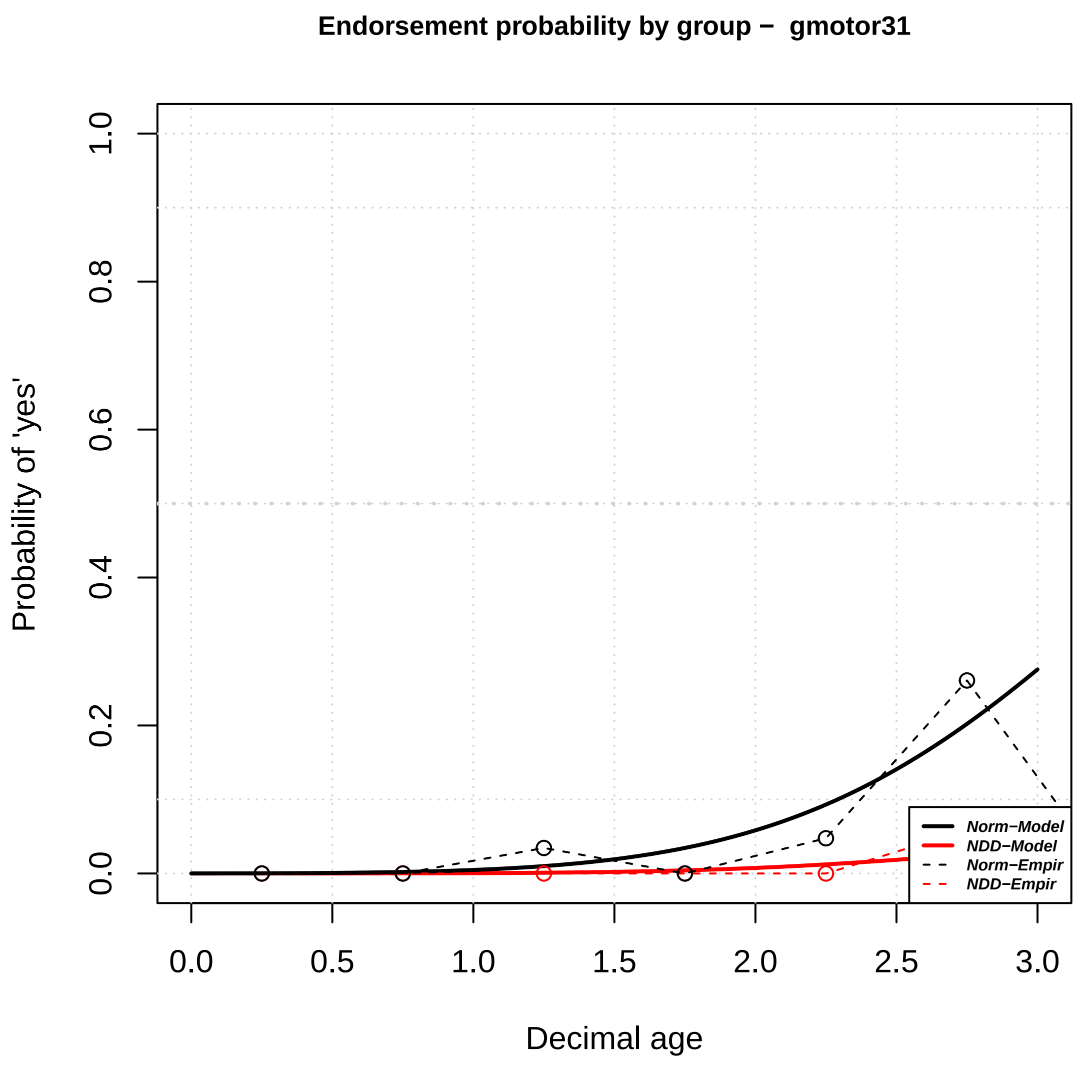
Endorsement probability by group − gmotor31
Probability of 'yes'
Decimal age
0.0
0.2
0.4
0.6
0.8
1.0
0.0
0.5
1.0
1.5
2.0
2.5
3.0
Norm−Model
NDD−Model
Norm−Empir
NDD−Empir

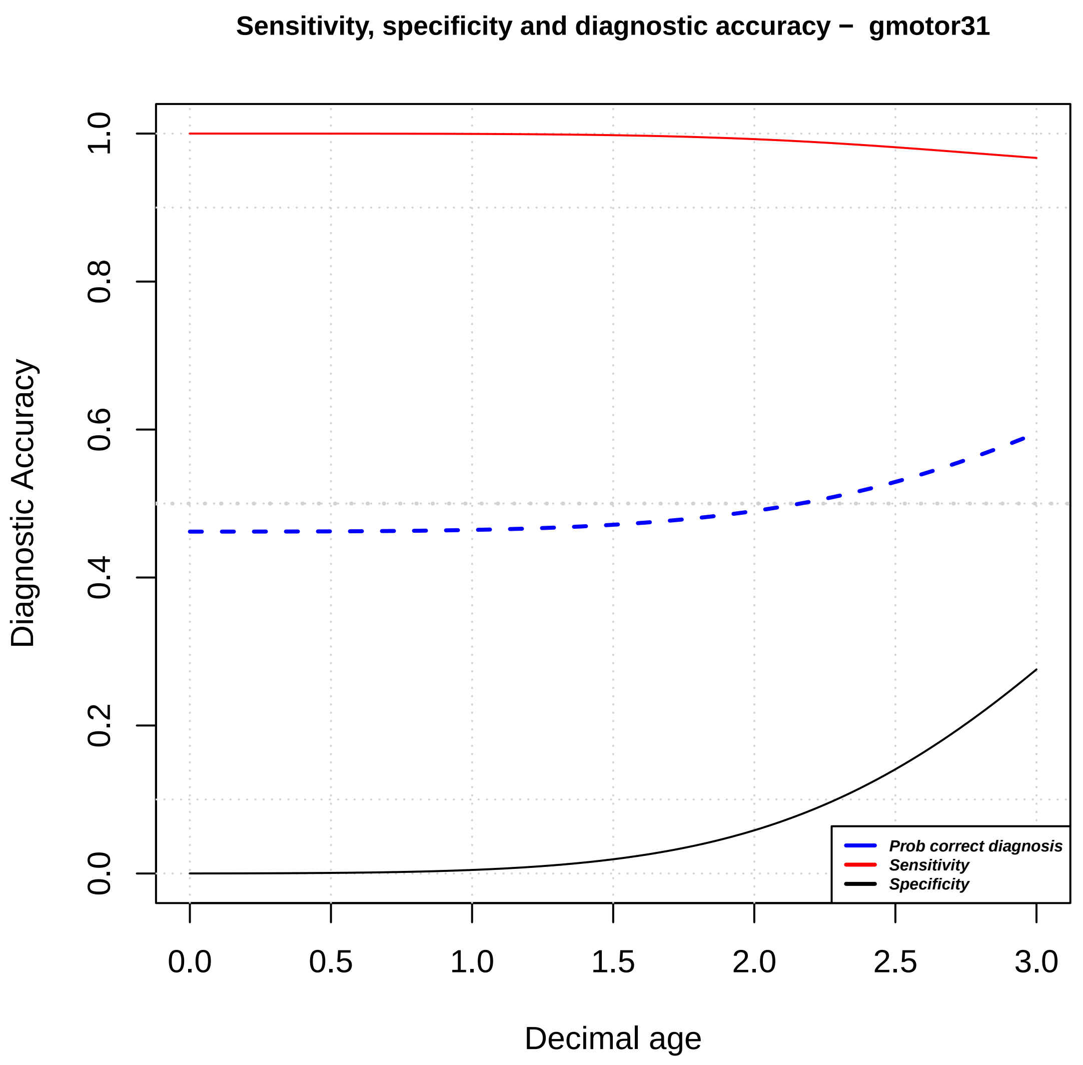
Sensitivity, specificity and diagnostic accuracy − gmotor31
Diagnostic Accuracy
Decimal age
0.0
0.2
0.4
0.6
0.8
1.0
0.0
0.5
1.0
1.5
2.0
2.5
3.0
Prob correct diagnosis
Sensitivity
Specificity

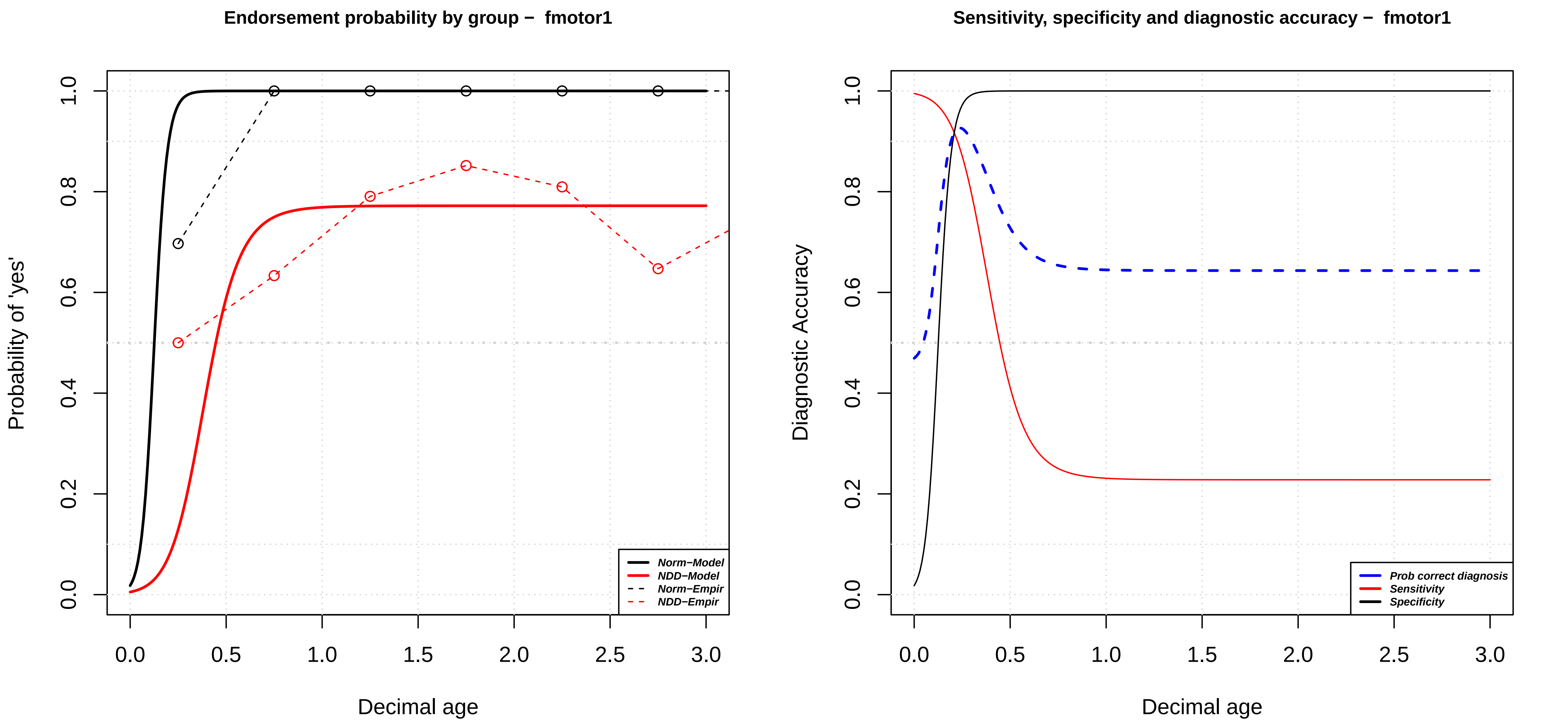
Endorsement probability by group – fmotor1
Probability of 'yes'
Decimal age
0.0
0.2
0.4
0.6
0.8
1.0
0.0
0.5
1.0
1.5
2.0
2.5
3.0
Norm–Model
NDD–Model
Norm–Empir
NDD–Empir
Sensitivity, specificity and diagnostic accuracy – fmotor1
Diagnostic Accuracy
Decimal age
Prob correct diagnosis
Sensitivity
Specificity

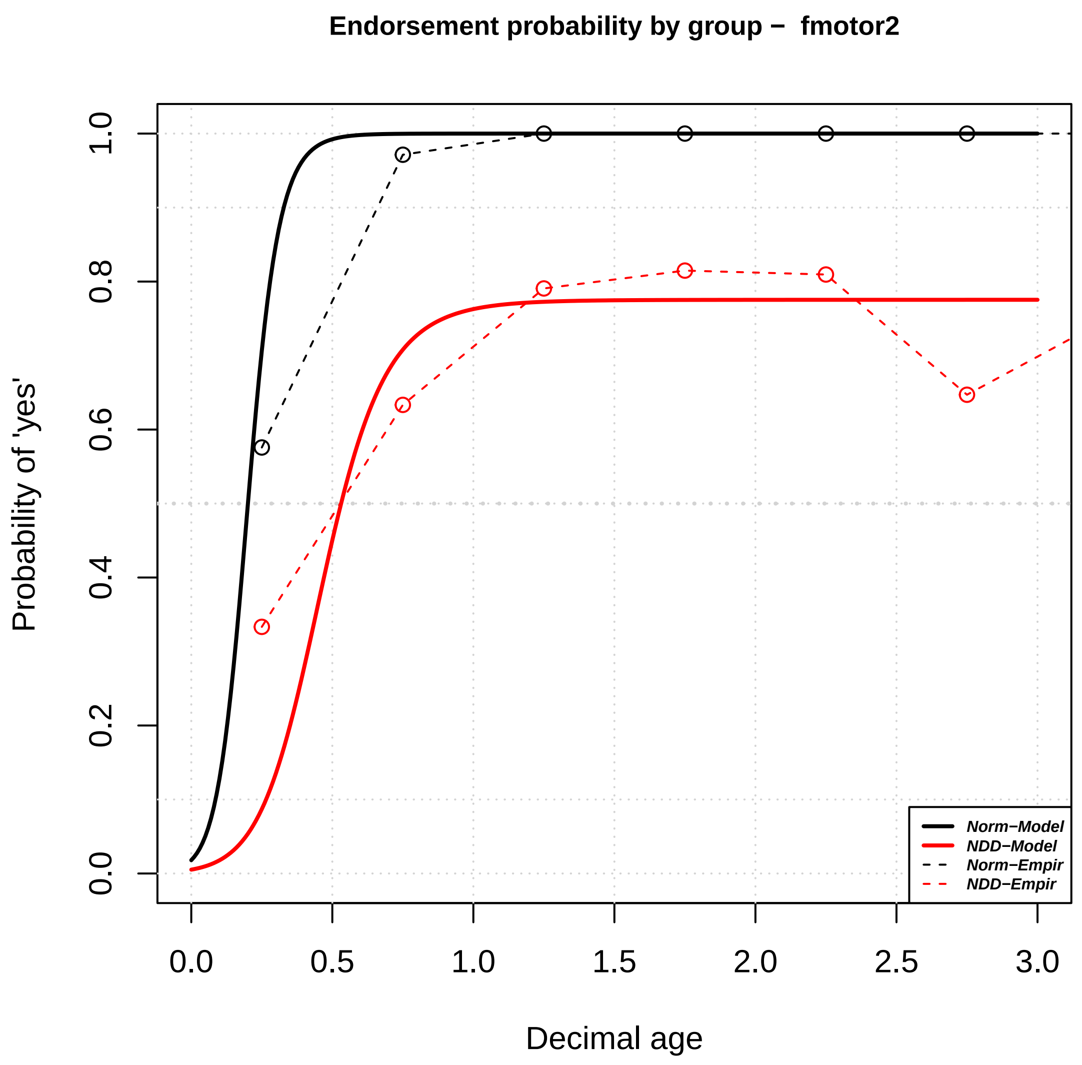
Endorsement probability by group – fmotor2
Probability of 'yes'
Decimal age
Norm–Model
NDD–Model
Norm–Empir
NDD–Empir
0.0
0.2
0.4
0.6
0.8
1.0
0.5
1.5
2.0
2.5
3.0

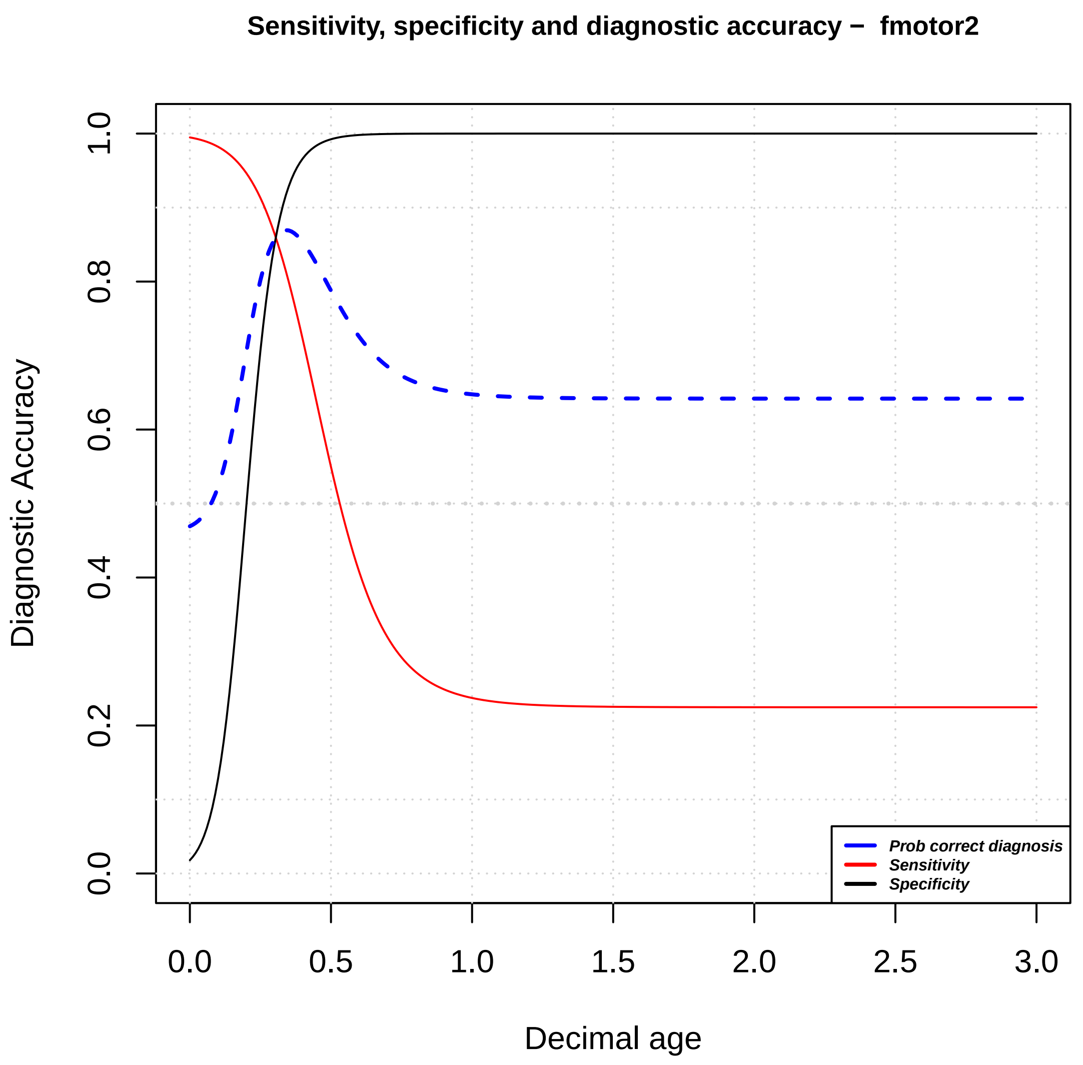
Sensitivity, specificity and diagnostic accuracy – fmotor2
Diagnostic Accuracy
Decimal age
Prob correct diagnosis
Sensitivity
Specificity
0.0
0.2
0.4
0.6
0.8
1.0
0.5
1.5
2.0
2.5
3.0

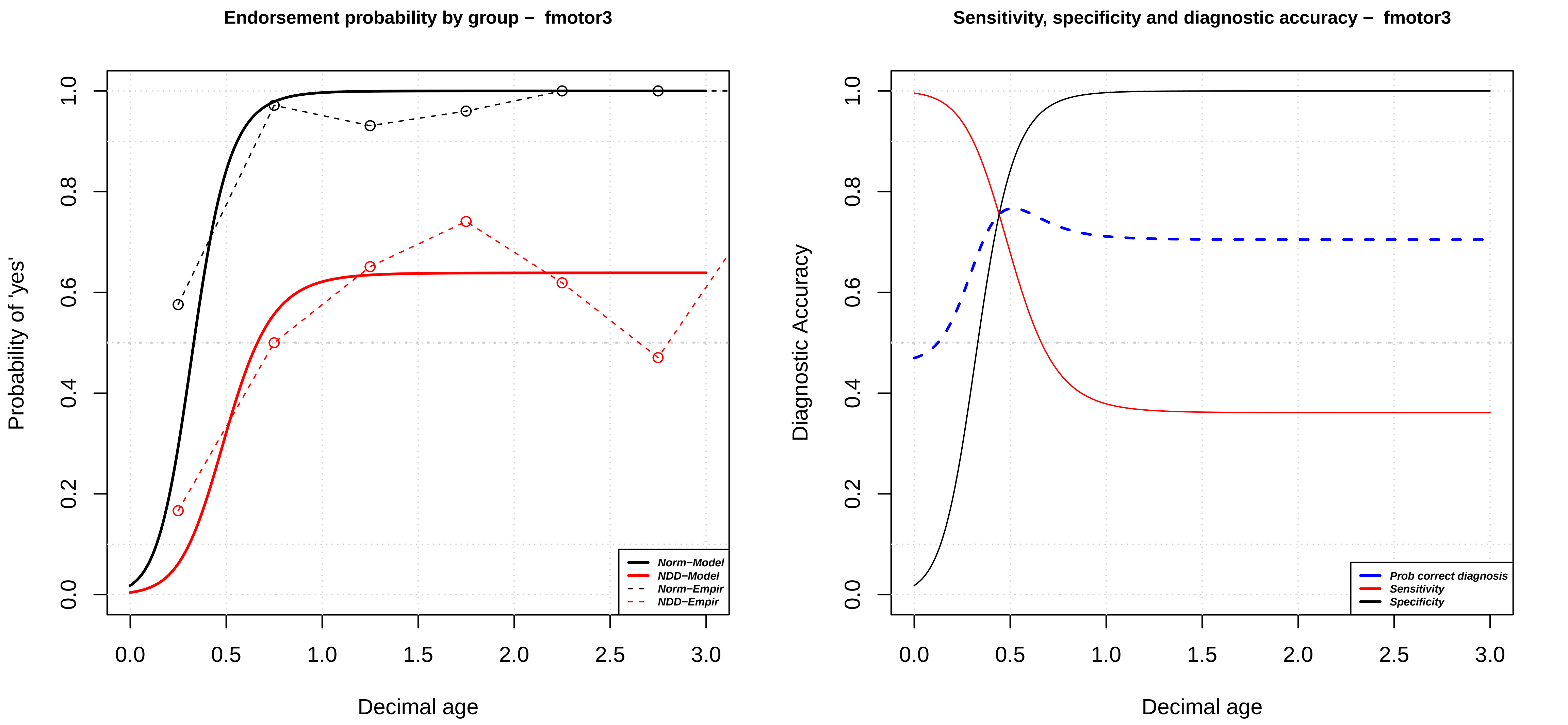

Endorsement probability by group – fmotor3
Probability of 'yes'
Decimal age
0.0
0.2
0.4
0.6
0.8
1.0
0.0
0.5
1.0
1.5
2.0
2.5
3.0
Norm–Model
NDD–Model
Norm–Empir
NDD–Empir
Sensitivity, specificity and diagnostic accuracy – fmotor3
Diagnostic Accuracy
Decimal age
Prob correct diagnosis
Sensitivity
Specificity

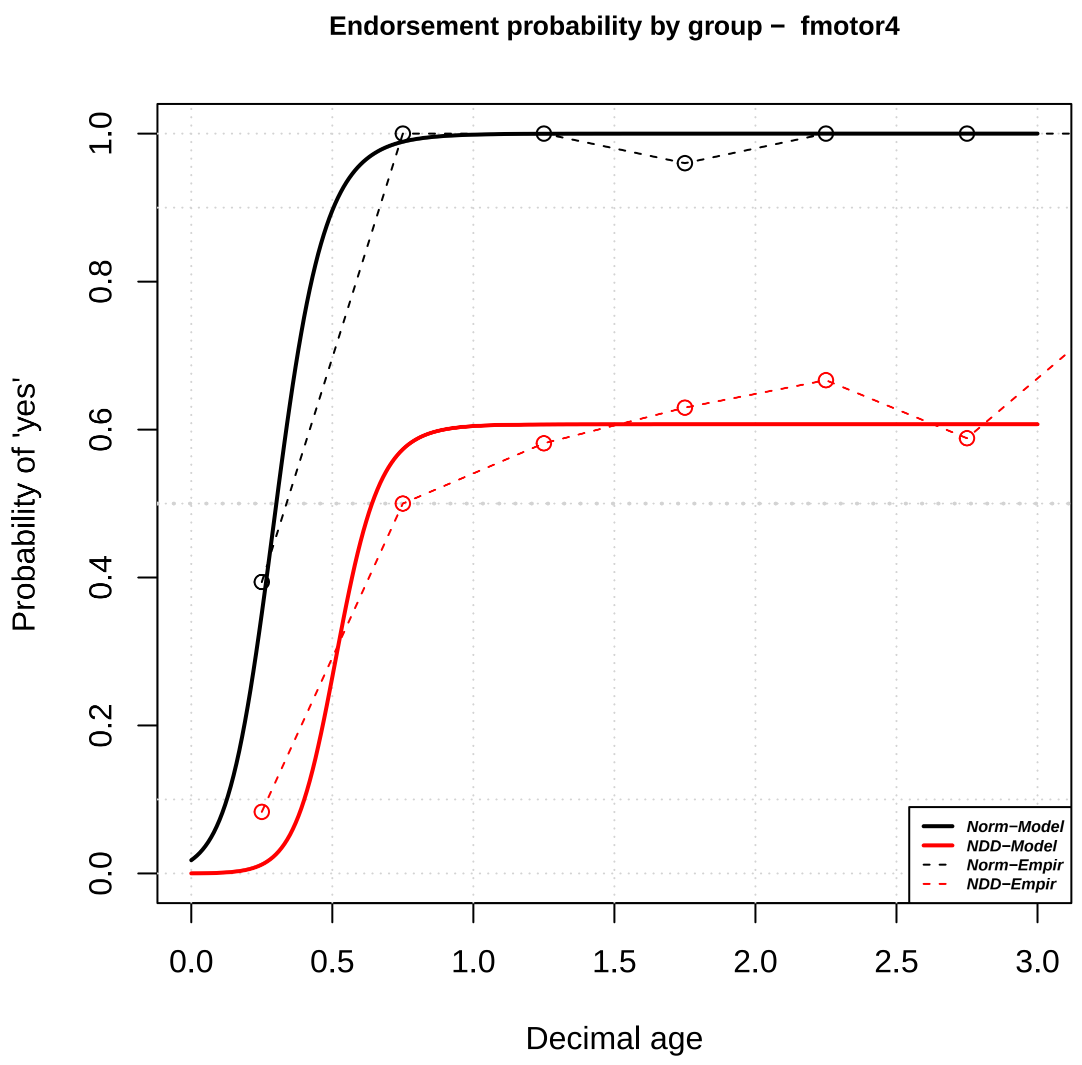

Endorsement probability by group – fmotor4
Probability of 'yes'
Decimal age
0.0
0.2
0.4
0.6
0.8
1.0
0.0
0.5
1.0
1.5
2.0
2.5
3.0
Norm–Model
NDD–Model
Norm–Empir
NDD–Empir


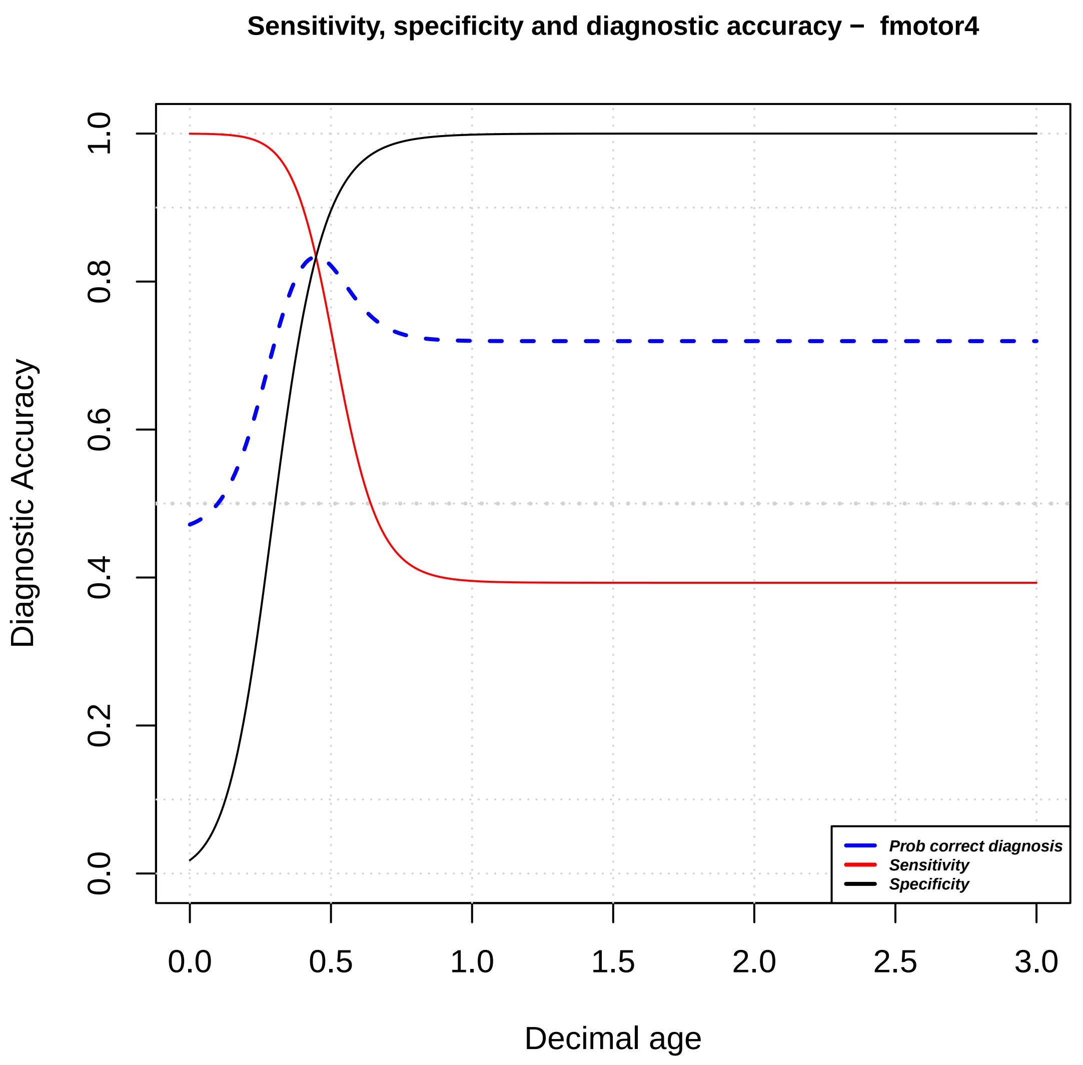

Sensitivity, specificity and diagnostic accuracy – fmotor4
Diagnostic Accuracy
Decimal age
0.0
0.2
0.4
0.6
0.8
1.0
0.0
0.5
1.0
1.5
2.0
2.5
3.0
Prob correct diagnosis
Sensitivity
Specificity

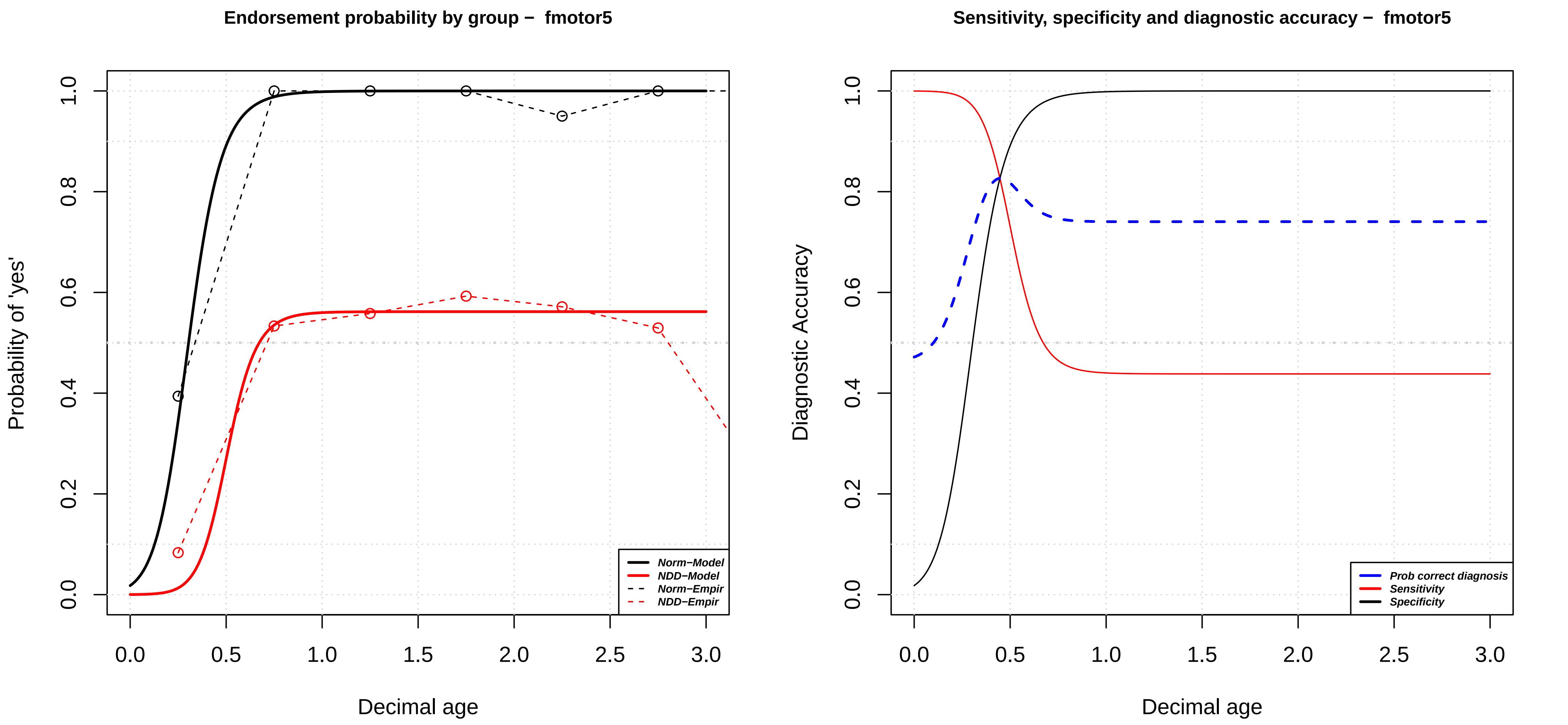

Endorsement probability by group – fmotor5
Probability of 'yes'
0.0
0.2
0.4
0.6
0.8
1.0
0.0
0.5
1.0
1.5
2.0
2.5
3.0
Decimal age
Norm−Model
NDD−Model
Norm−Empir
NDD−Empir
Sensitivity, specificity and diagnostic accuracy – fmotor5
Diagnostic Accuracy
0.0
0.2
0.4
0.6
0.8
1.0
0.0
0.5
1.0
1.5
2.0
2.5
3.0
Decimal age
Prob correct diagnosis
Sensitivity
Specificity

### Endorsement probability by group – fmotor6

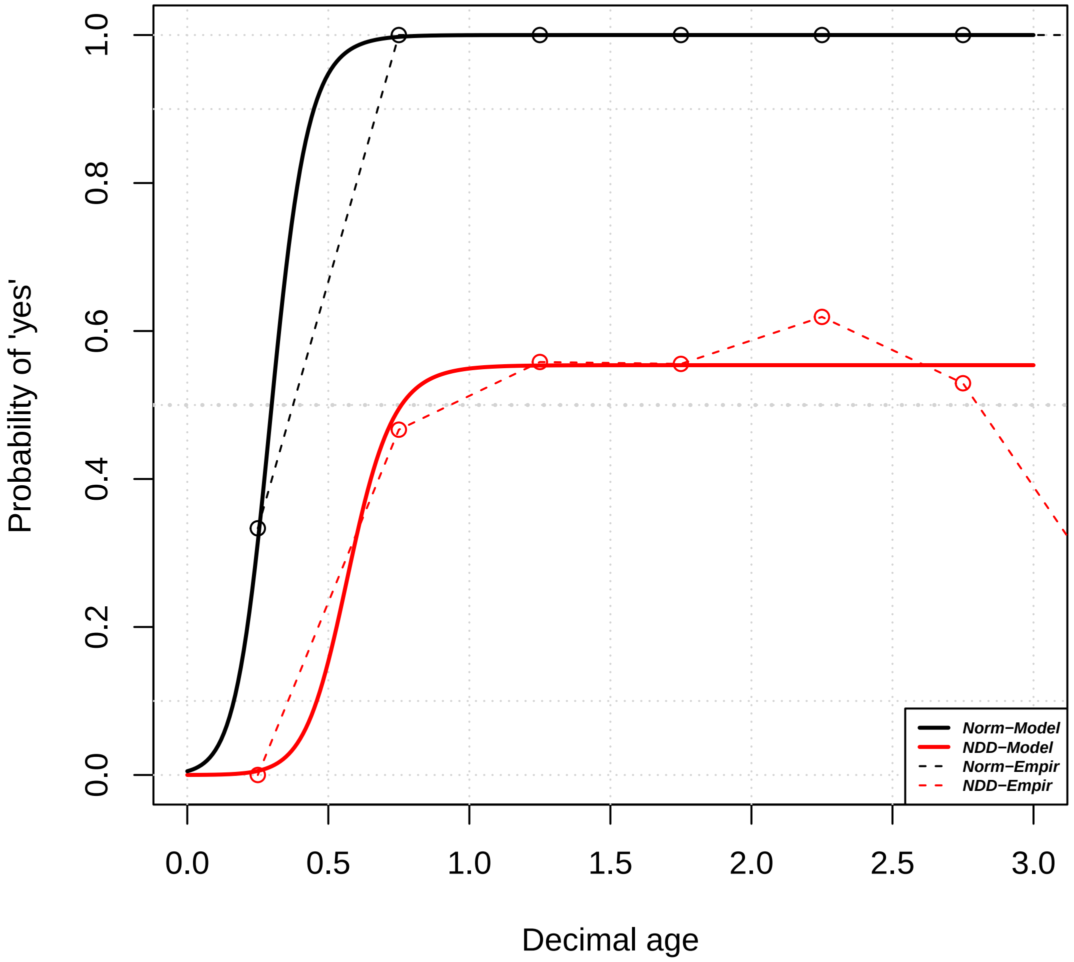


### Sensitivity, specificity and diagnostic accuracy – fmotor6

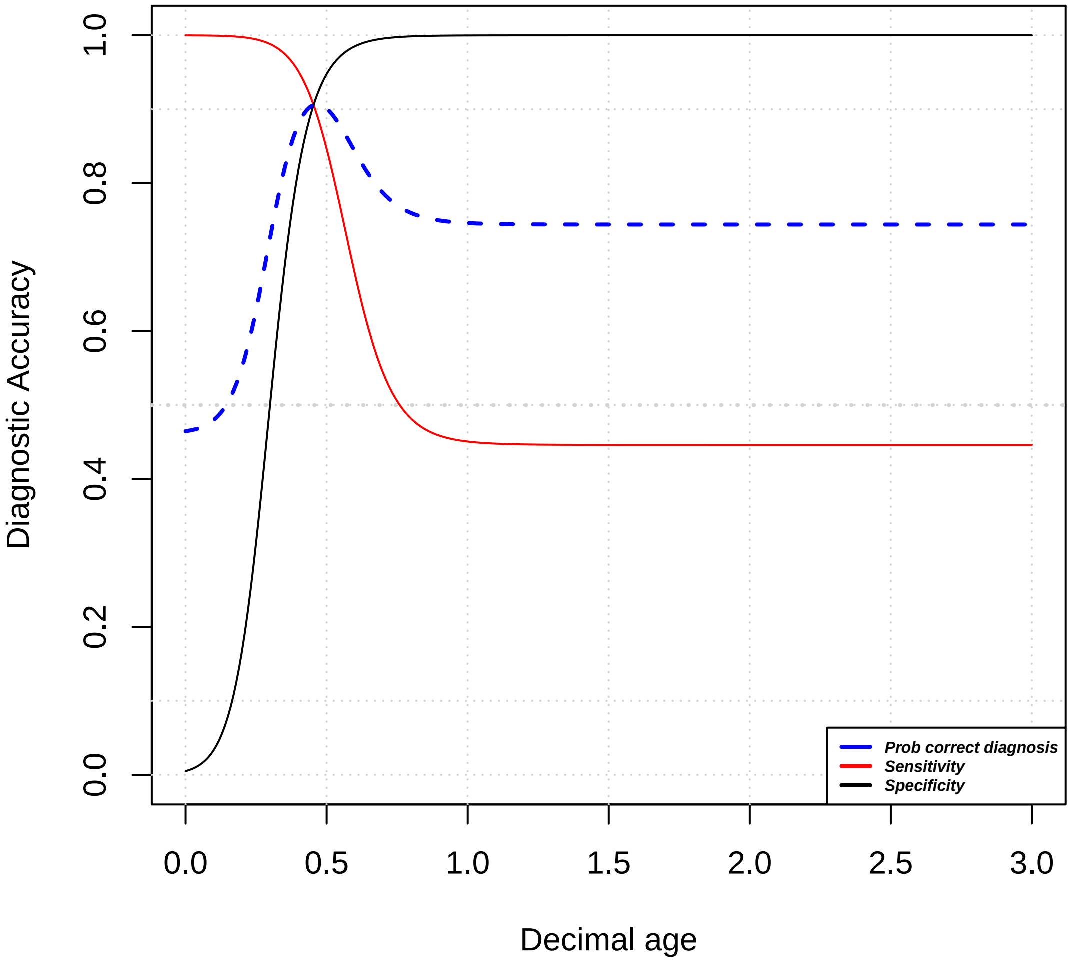

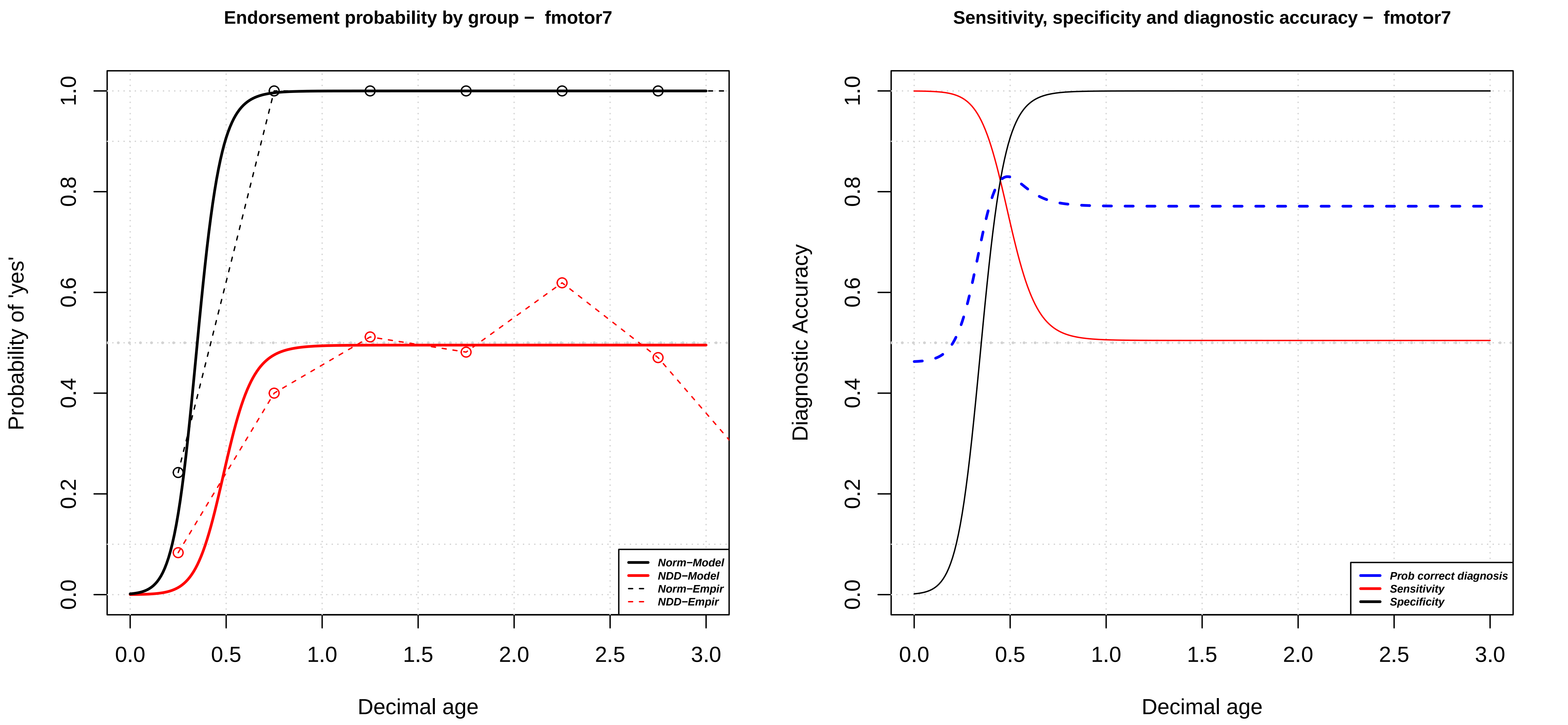
Endorsement probability by group – fmotor7
Probability of 'yes'
Decimal age
0.0
0.2
0.4
0.6
0.8
1.0
0.0
0.5
1.0
1.5
2.0
2.5
3.0
Norm–Model
NDD–Model
Norm–Empir
NDD–Empir
Sensitivity, specificity and diagnostic accuracy – fmotor7
Diagnostic Accuracy
Decimal age
Prob correct diagnosis
Sensitivity
Specificity

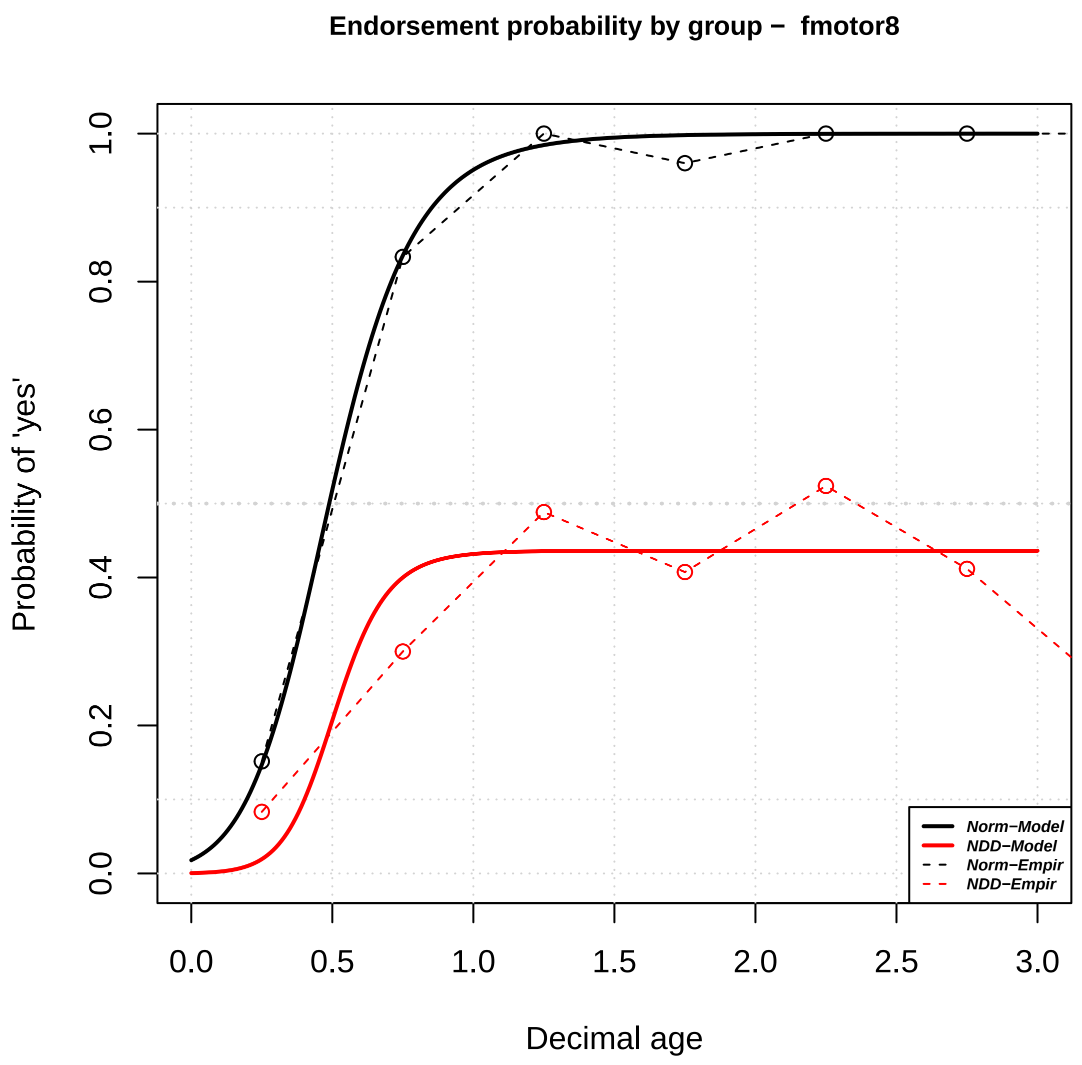

Endorsement probability by group – fmotor8
Probability of 'yes'
Decimal age
0.0
0.2
0.4
0.6
0.8
1.0
0.0
0.5
1.0
1.5
2.0
2.5
3.0
Norm–Model
NDD–Model
Norm–Empir
NDD–Empir


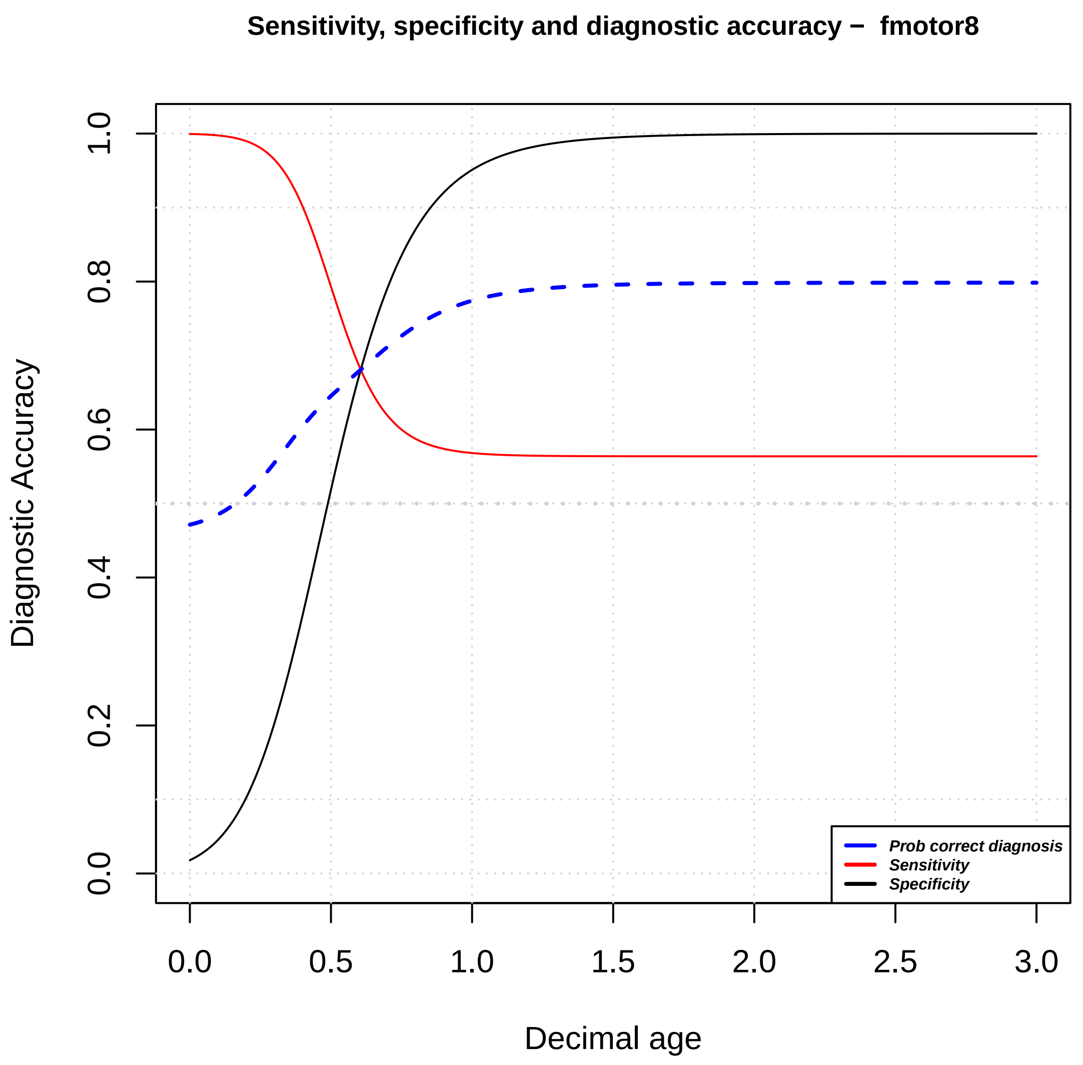

Sensitivity, specificity and diagnostic accuracy – fmotor8
Diagnostic Accuracy
Decimal age
0.0
0.2
0.4
0.6
0.8
1.0
0.0
0.5
1.0
1.5
2.0
2.5
3.0
Prob correct diagnosis
Sensitivity
Specificity

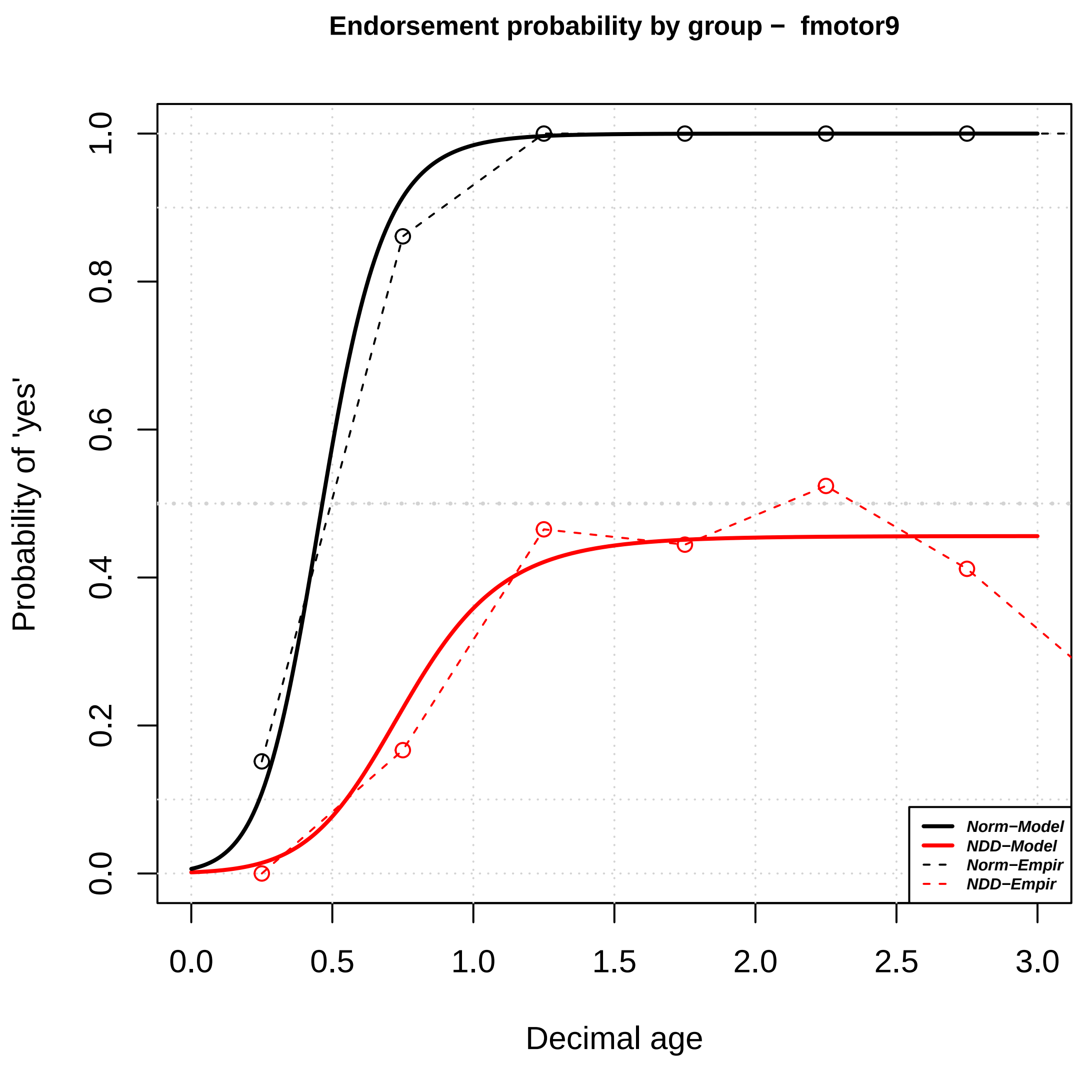

Endorsement probability by group – fmotor9
Probability of 'yes'
Decimal age
0.0
0.2
0.4
0.6
0.8
1.0
0.0
0.5
1.0
1.5
2.0
2.5
3.0
Norm–Model
NDD–Model
Norm–Empir
NDD–Empir


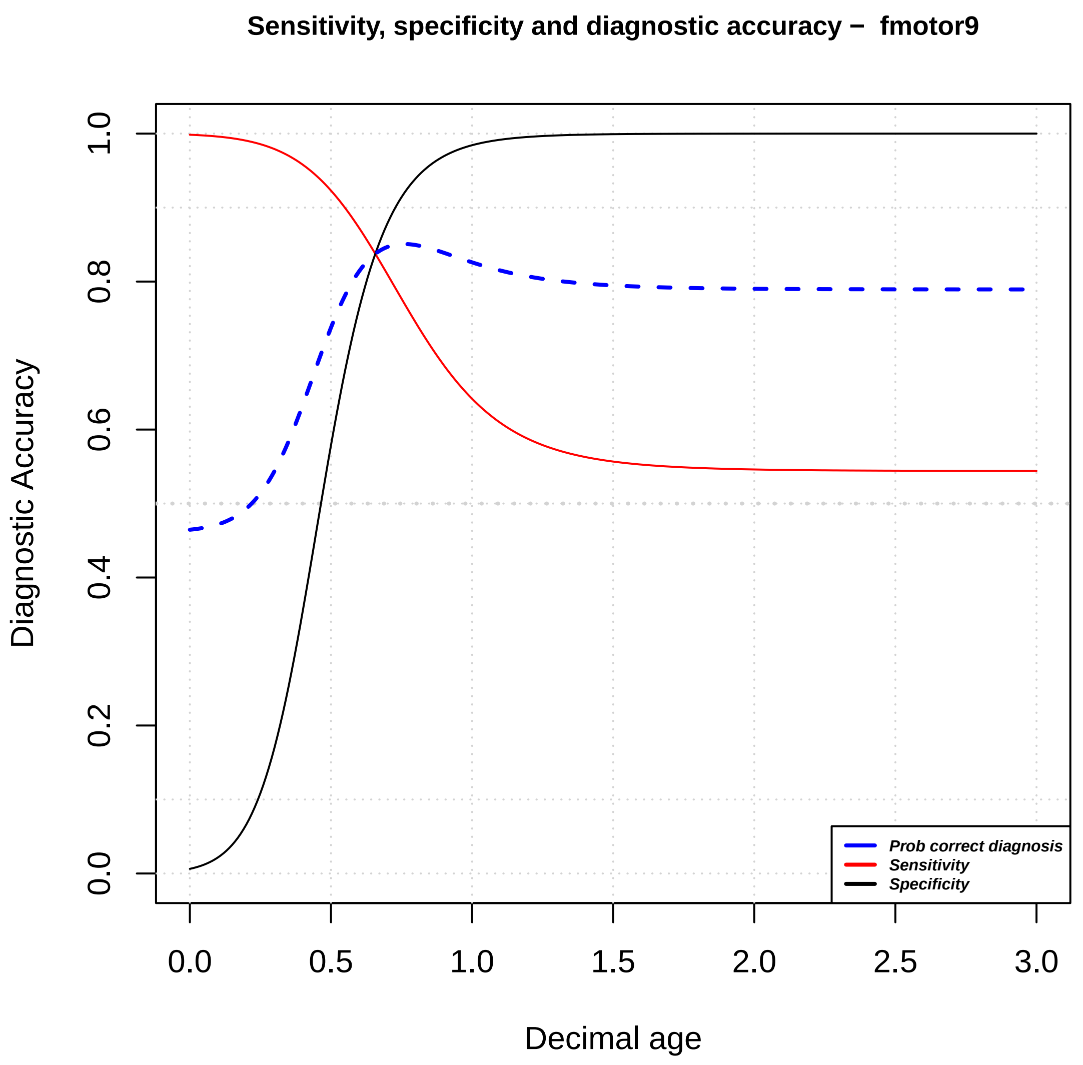

Sensitivity, specificity and diagnostic accuracy – fmotor9
Diagnostic Accuracy
Decimal age
0.0
0.2
0.4
0.6
0.8
1.0
0.0
0.5
1.0
1.5
2.0
2.5
3.0
Prob correct diagnosis
Sensitivity
Specificity

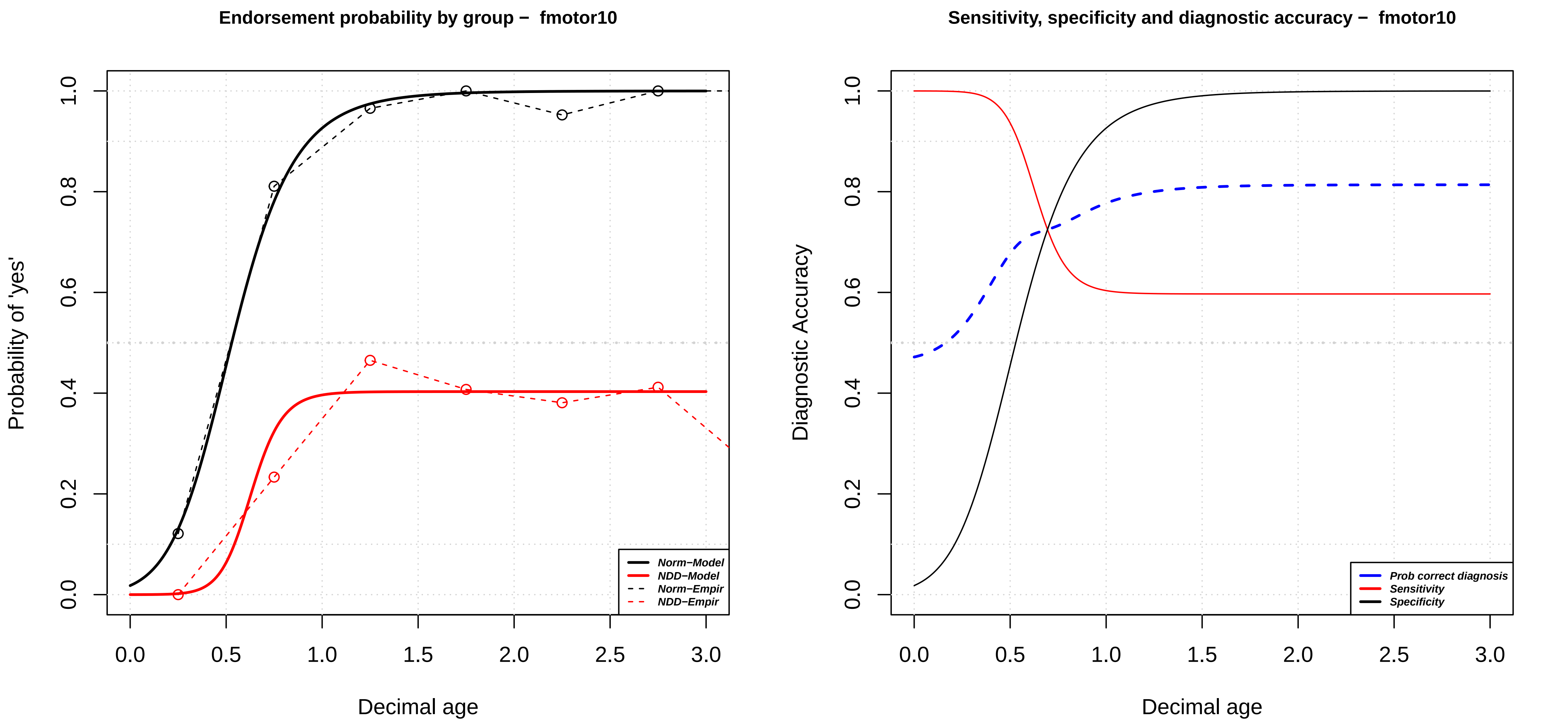

Endorsement probability by group – fmotor10
Probability of 'yes'
Decimal age
Norm−Model
NDD−Model
Norm−Empir
NDD−Empir
0.0
0.2
0.4
0.6
0.8
1.0
0.0
0.5
1.0
1.5
2.0
2.5
3.0
Sensitivity, specificity and diagnostic accuracy – fmotor10
Diagnostic Accuracy
Decimal age
Prob correct diagnosis
Sensitivity
Specificity

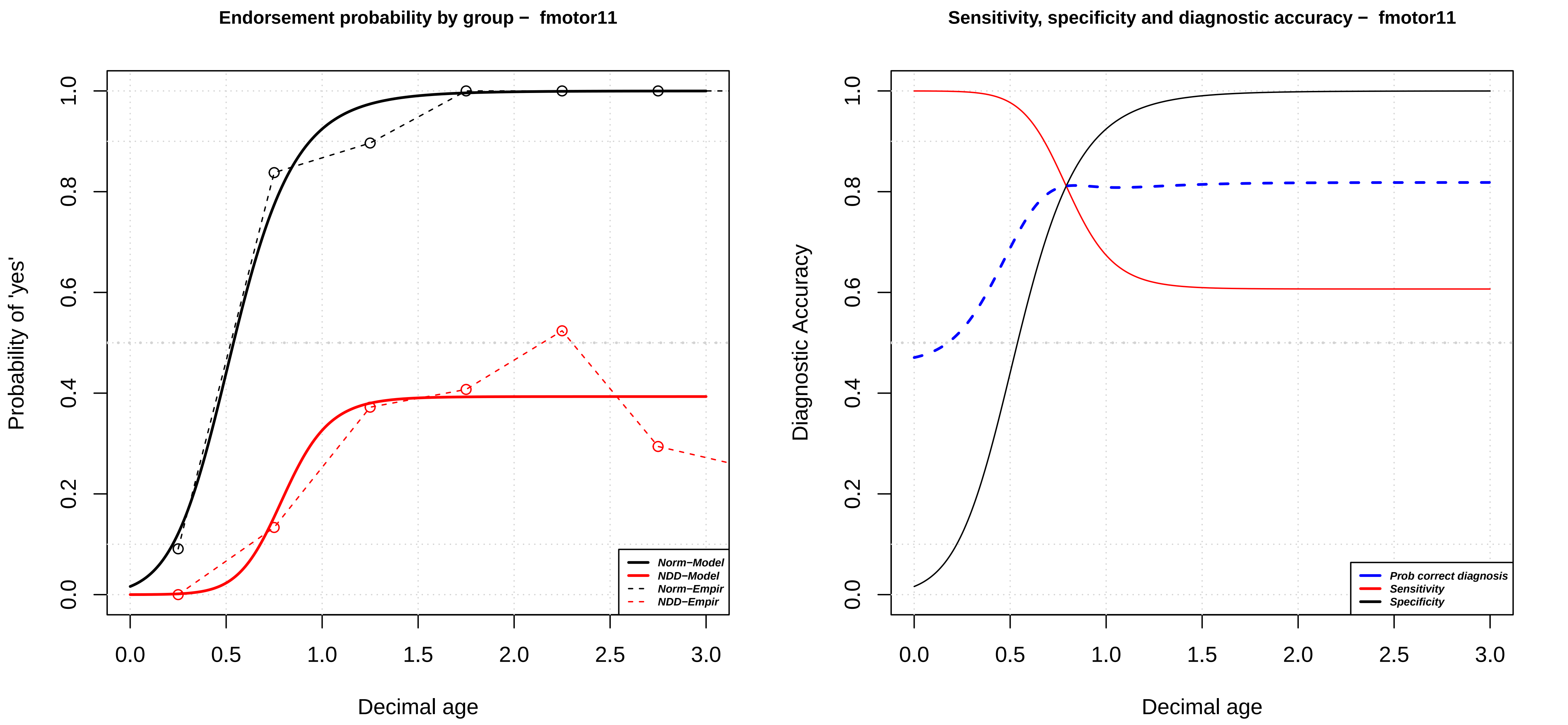
Endorsement probability by group – fmotor11
Probability of 'yes'
Decimal age
0.0
0.2
0.4
0.6
0.8
1.0
0.0
0.5
1.0
1.5
2.0
2.5
3.0
Norm−Model
NDD−Model
Norm−Empir
NDD−Empir
Sensitivity, specificity and diagnostic accuracy – fmotor11
Diagnostic Accuracy
Decimal age
Prob correct diagnosis
Sensitivity
Specificity

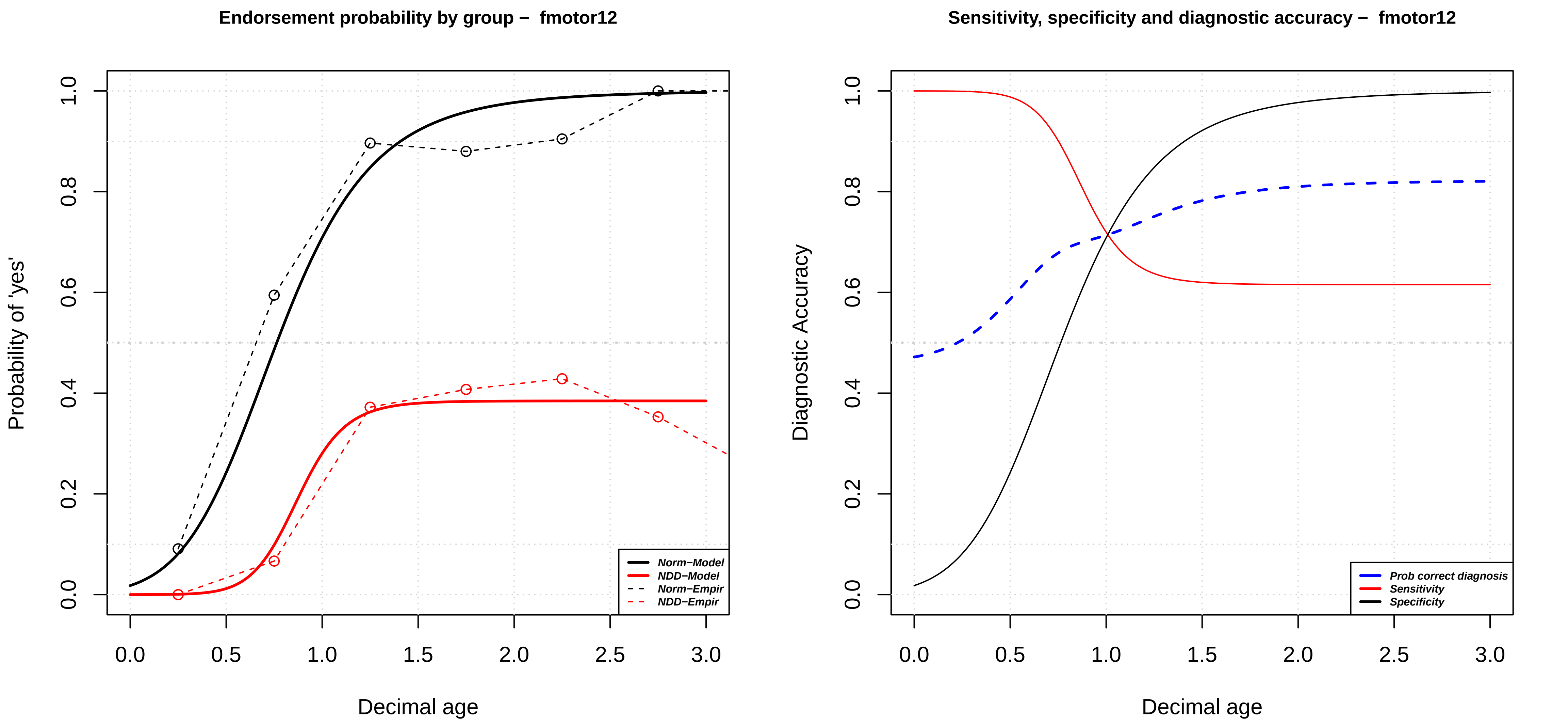

Endorsement probability by group – fmotor12
Probability of 'yes'
Decimal age
0.0
0.2
0.4
0.6
0.8
1.0
0.0
0.5
1.0
1.5
2.0
2.5
3.0
Norm–Model
NDD–Model
Norm–Empir
NDD–Empir
Sensitivity, specificity and diagnostic accuracy – fmotor12
Diagnostic Accuracy
Decimal age
Prob correct diagnosis
Sensitivity
Specificity

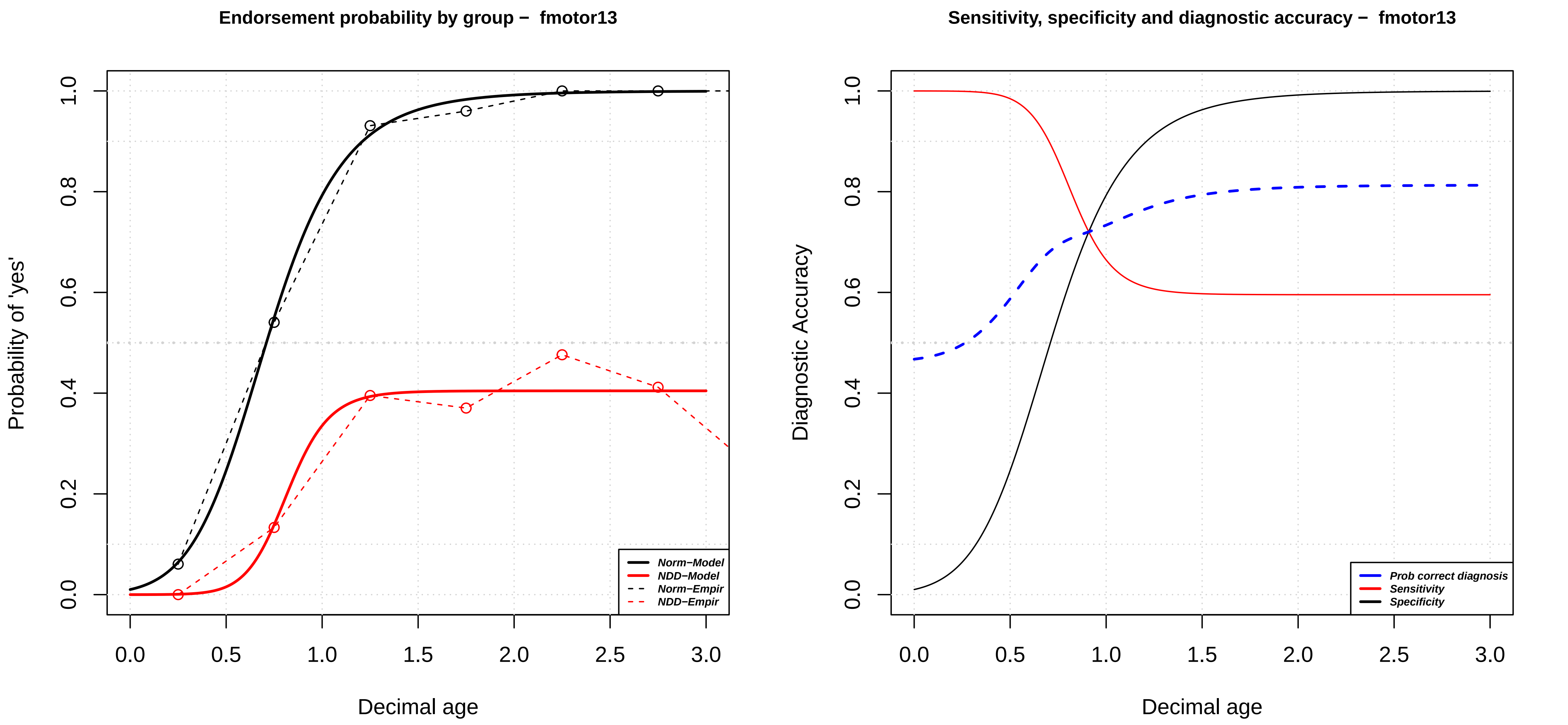

Endorsement probability by group – fmotor13
Probability of 'yes'
Decimal age
Norm−Model
NDD−Model
Norm−Empir
NDD−Empir
Sensitivity, specificity and diagnostic accuracy – fmotor13
Diagnostic Accuracy
Decimal age
Prob correct diagnosis
Sensitivity
Specificity

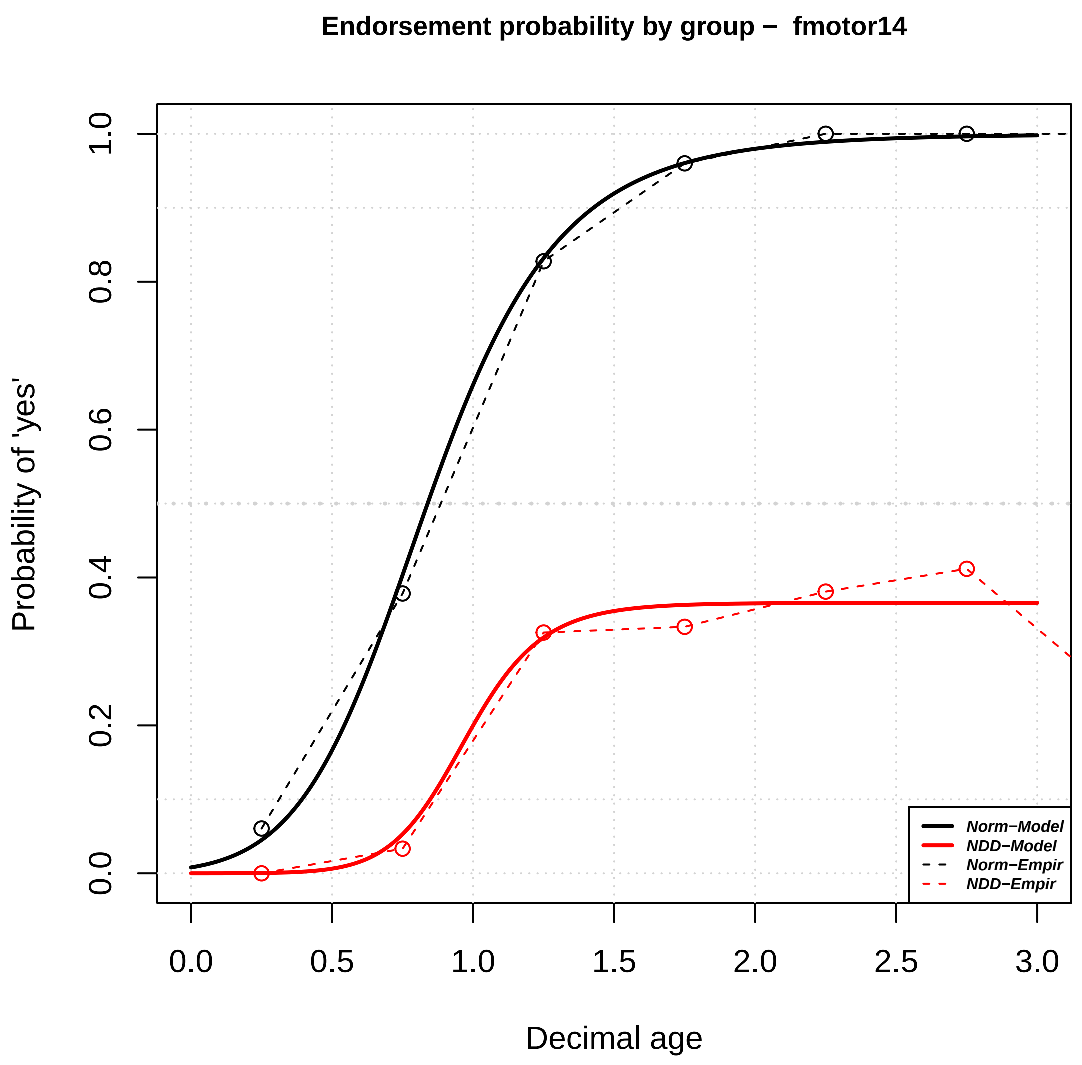

Endorsement probability by group – fmotor14
Probability of 'yes'
Decimal age
0.0
0.2
0.4
0.6
0.8
1.0
0.0
0.5
1.0
1.5
2.0
2.5
3.0
Norm−Model
NDD−Model
Norm−Empir
NDD−Empir


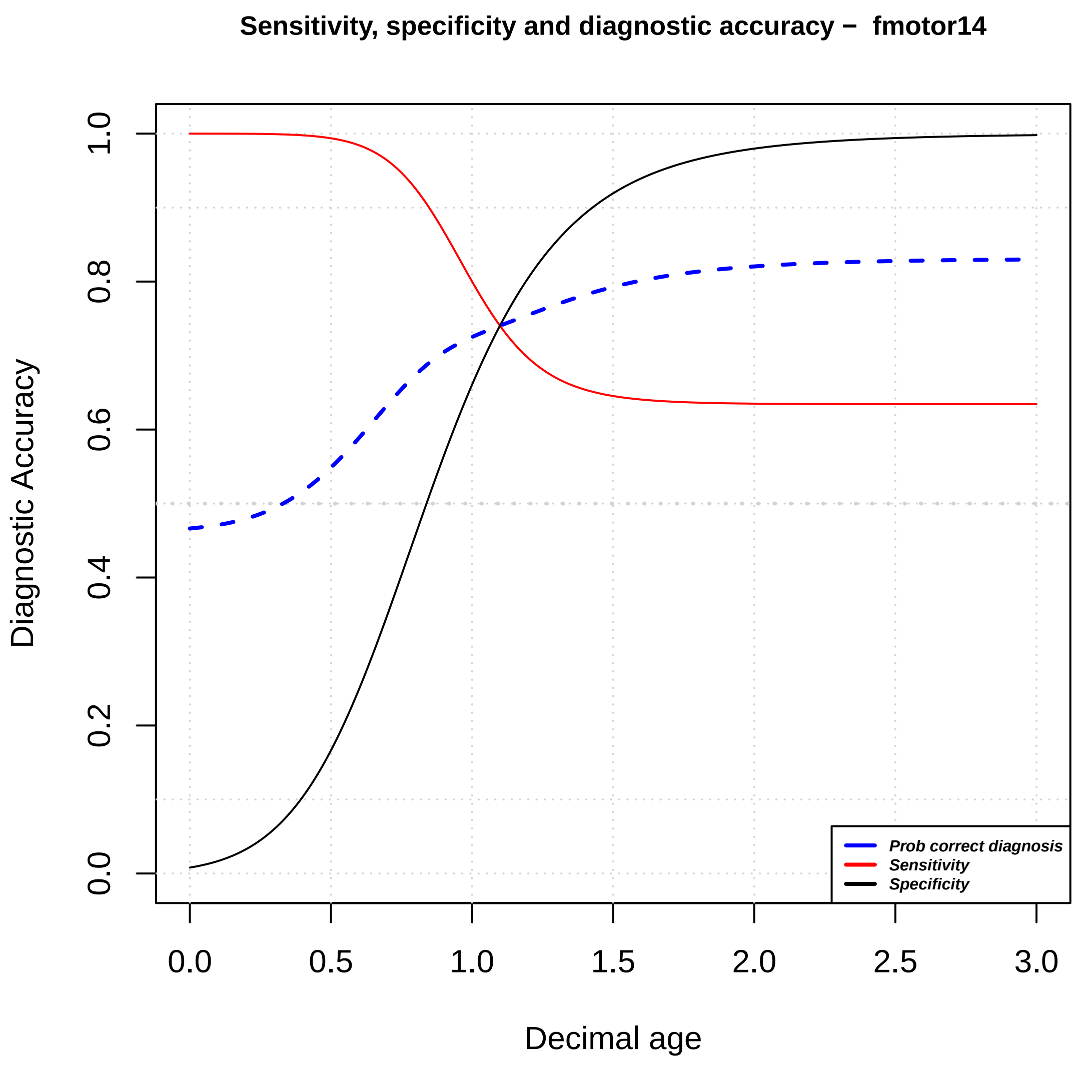

Sensitivity, specificity and diagnostic accuracy – fmotor14
Diagnostic Accuracy
Decimal age
0.0
0.2
0.4
0.6
0.8
1.0
0.0
0.5
1.0
1.5
2.0
2.5
3.0
Prob correct diagnosis
Sensitivity
Specificity

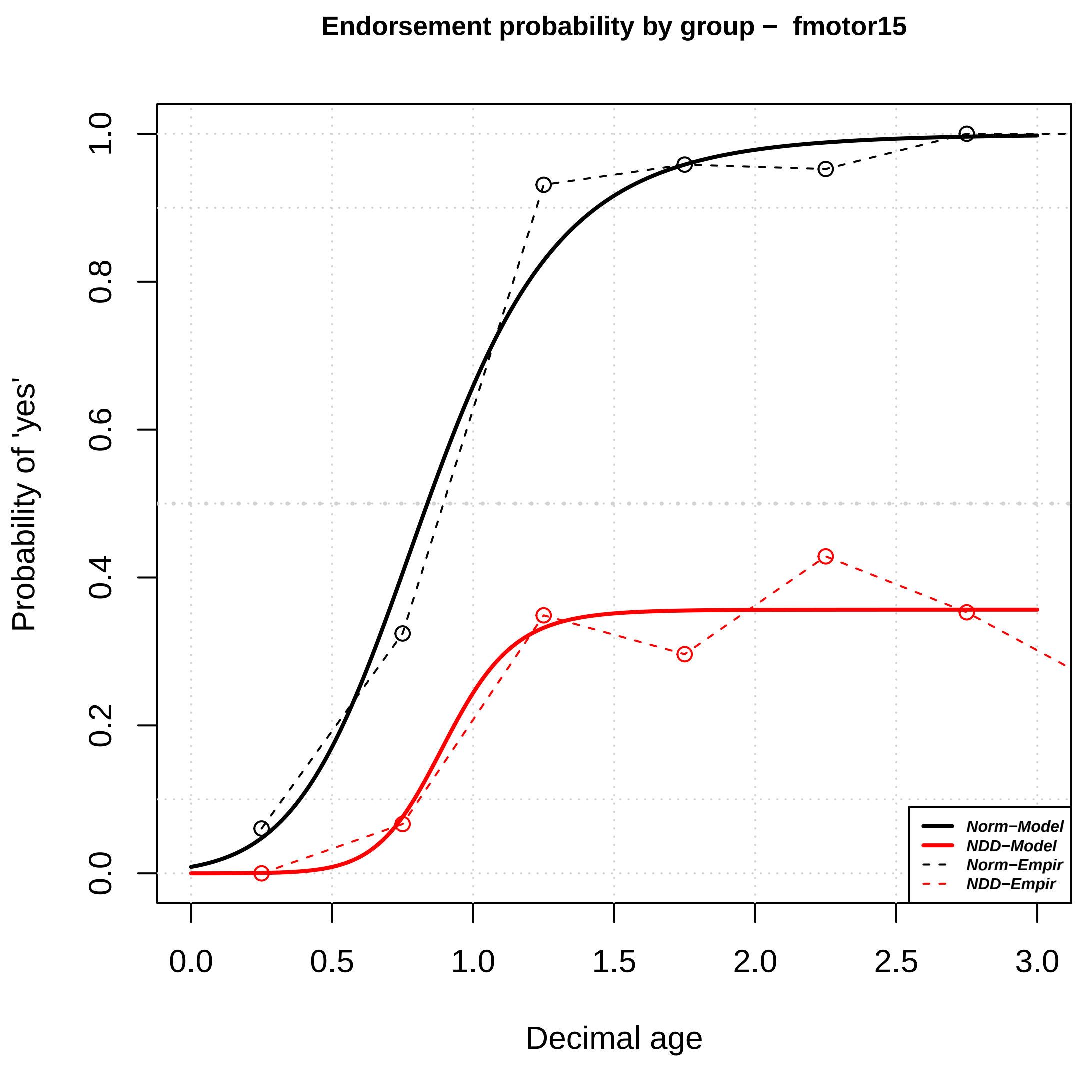

Endorsement probability by group – fmotor15
Probability of 'yes'
Decimal age
0.0
0.2
0.4
0.6
0.8
1.0
0.0
0.5
1.0
1.5
2.0
2.5
3.0
Norm–Model
NDD–Model
Norm–Empir
NDD–Empir


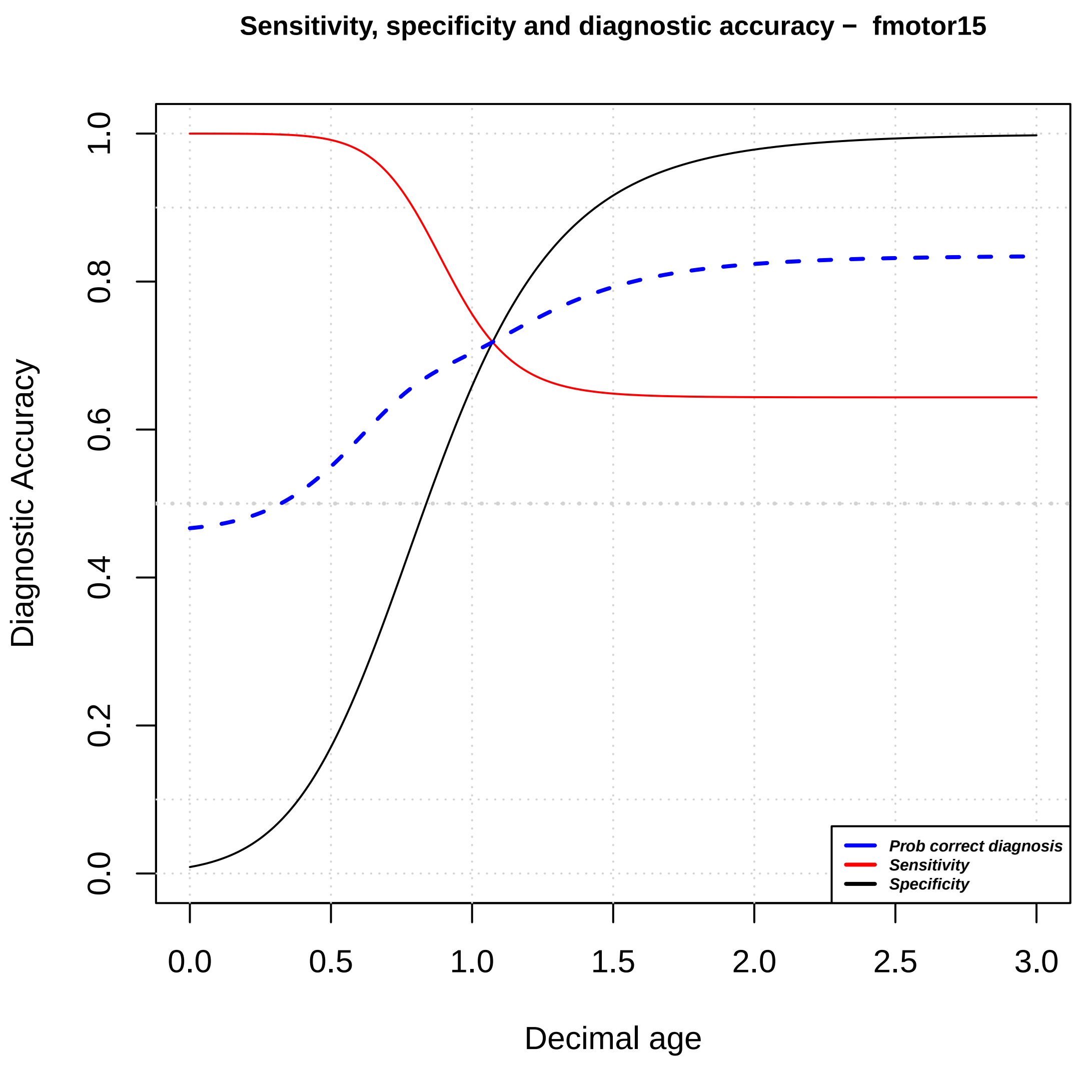

Sensitivity, specificity and diagnostic accuracy – fmotor15
Diagnostic Accuracy
Decimal age
0.0
0.2
0.4
0.6
0.8
1.0
0.0
0.5
1.0
1.5
2.0
2.5
3.0
Prob correct diagnosis
Sensitivity
Specificity

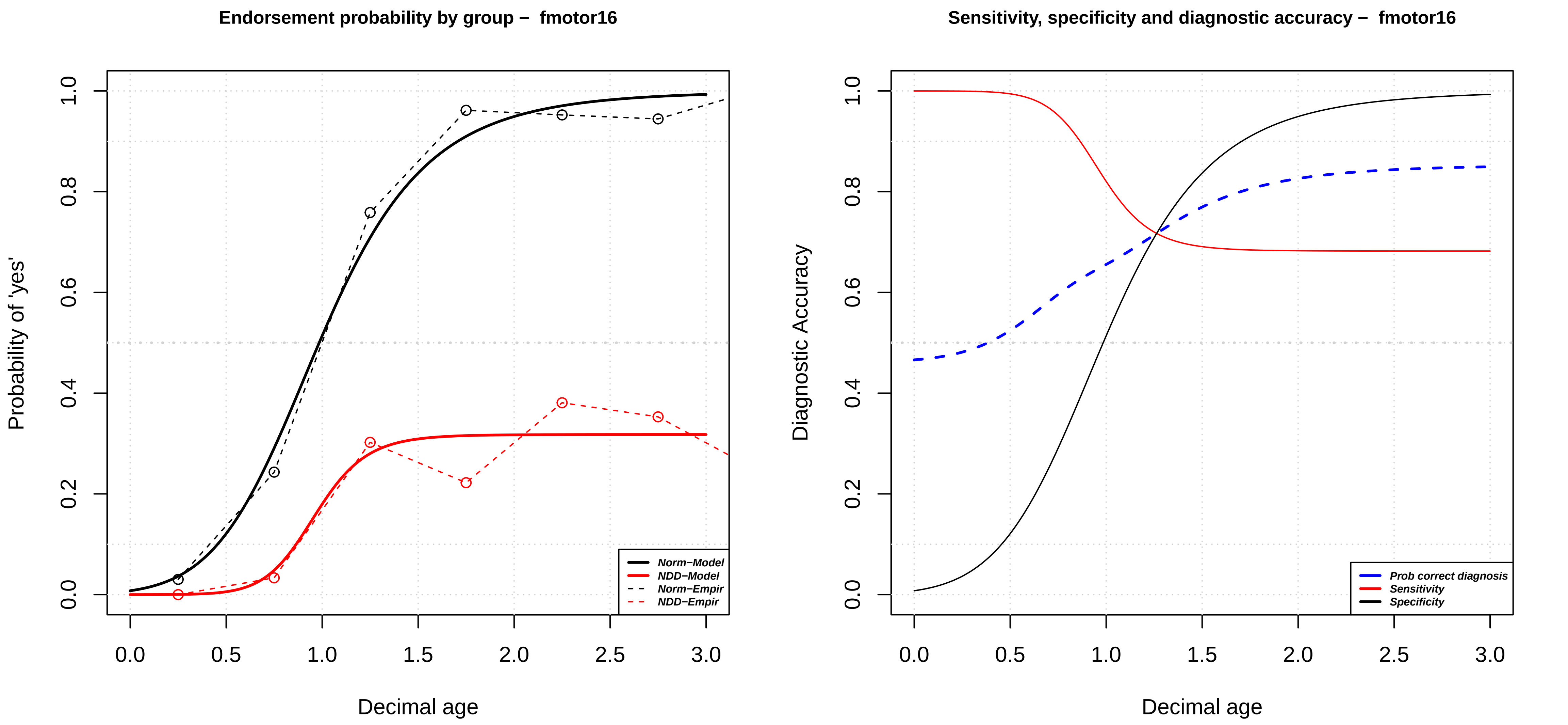

Endorsement probability by group – fmotor16
Probability of 'yes'
Decimal age
Norm−Model
NDD−Model
Norm−Empir
NDD−Empir
Sensitivity, specificity and diagnostic accuracy – fmotor16
Diagnostic Accuracy
Decimal age
Prob correct diagnosis
Sensitivity
Specificity

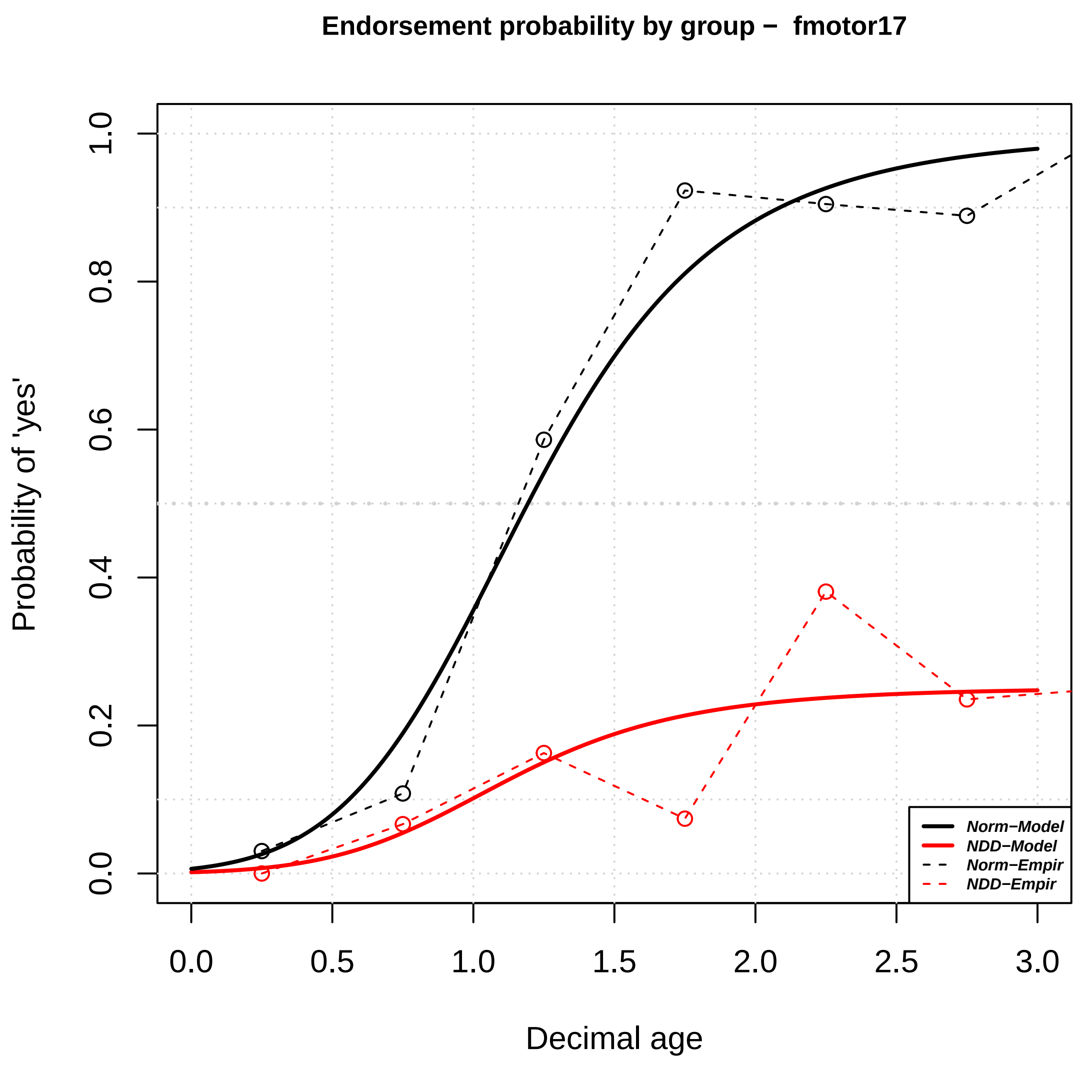

Endorsement probability by group – fmotor17
Probability of 'yes'
Decimal age
Norm−Model
NDD−Model
Norm−Empir
NDD−Empir


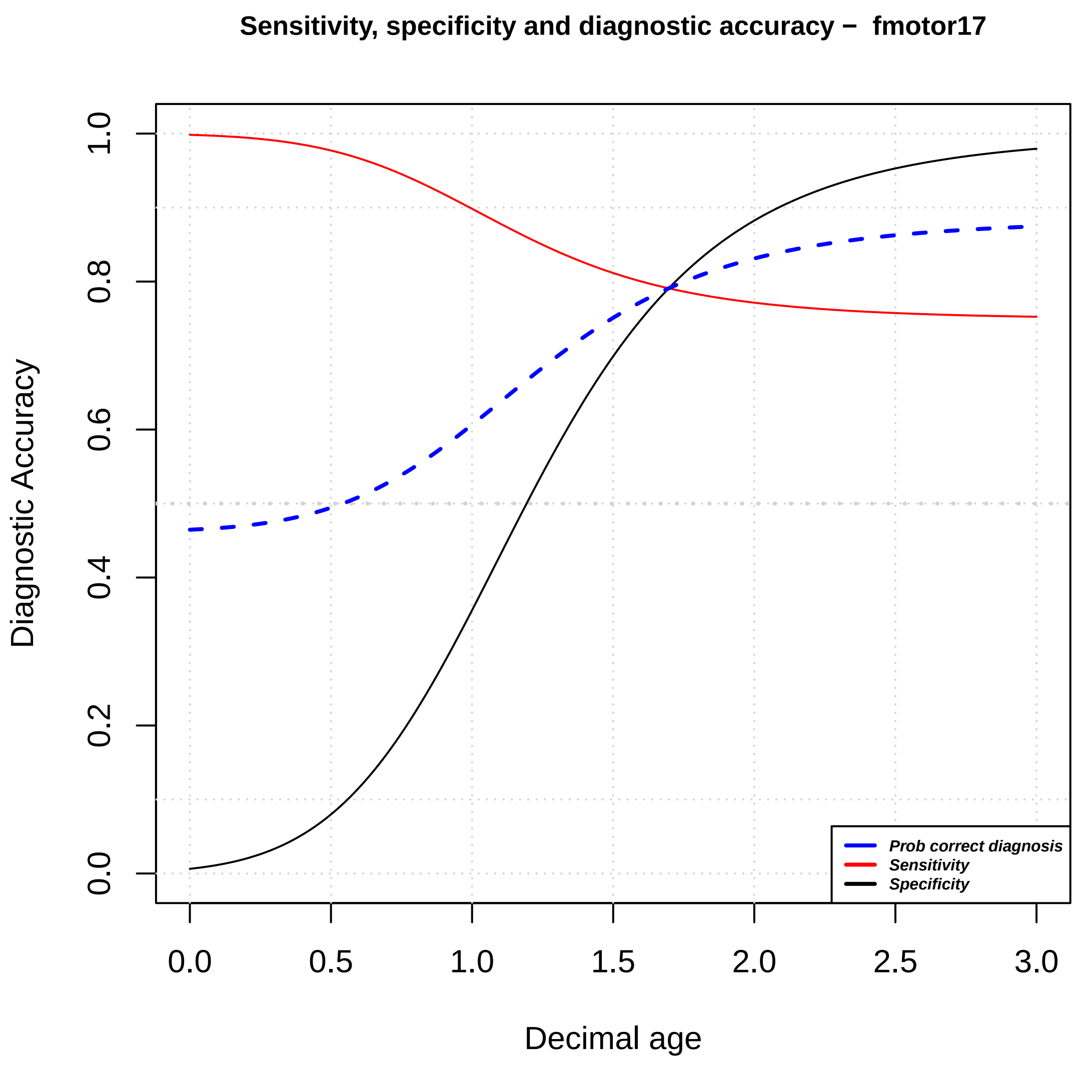

Sensitivity, specificity and diagnostic accuracy – fmotor17
Diagnostic Accuracy
Decimal age
Prob correct diagnosis
Sensitivity
Specificity

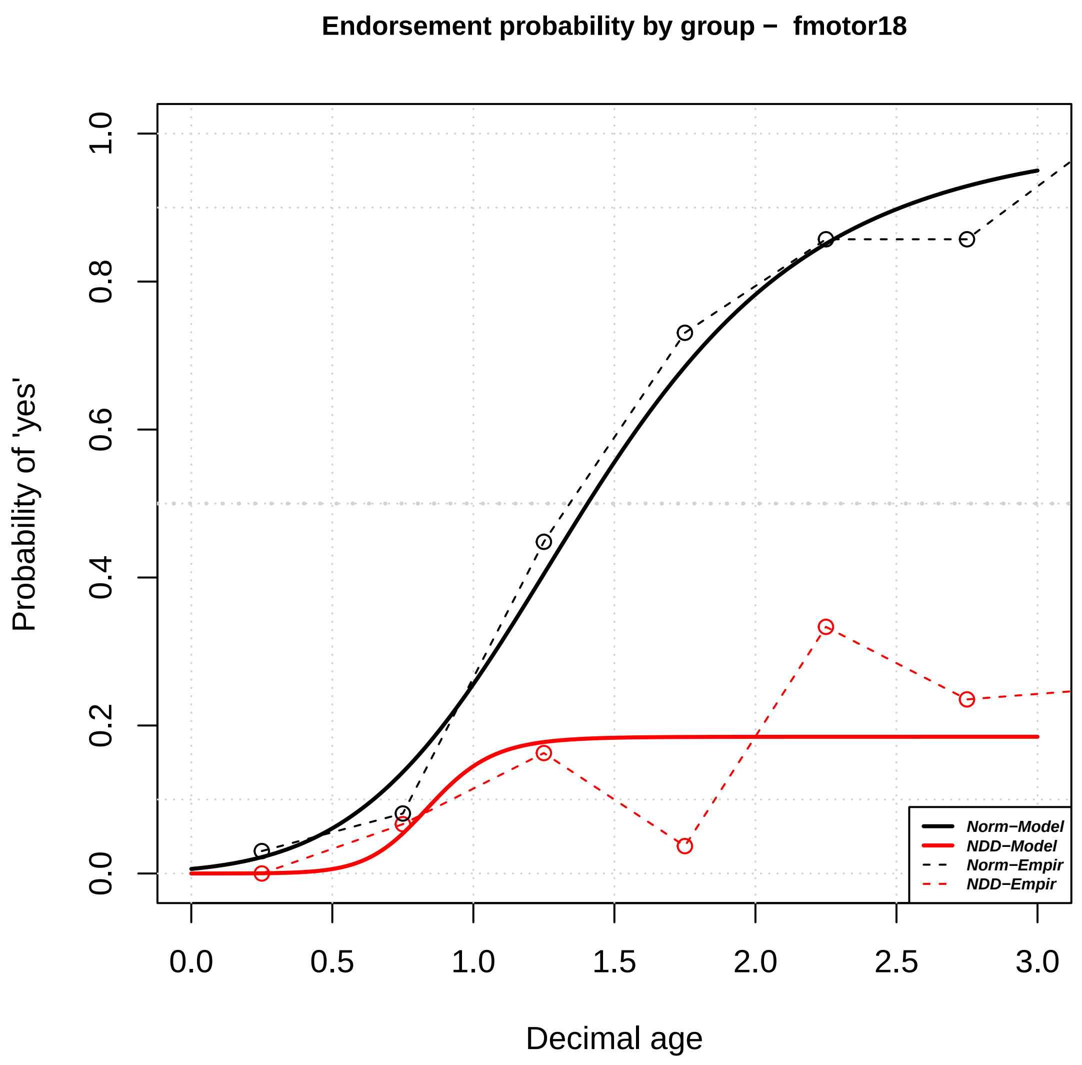
Endorsement probability by group – fmotor18
Probability of 'yes'
Decimal age
0.0
0.2
0.4
0.6
0.8
1.0
0.0
0.5
1.0
1.5
2.0
2.5
3.0
Norm−Model
NDD−Model
Norm−Empir
NDD−Empir

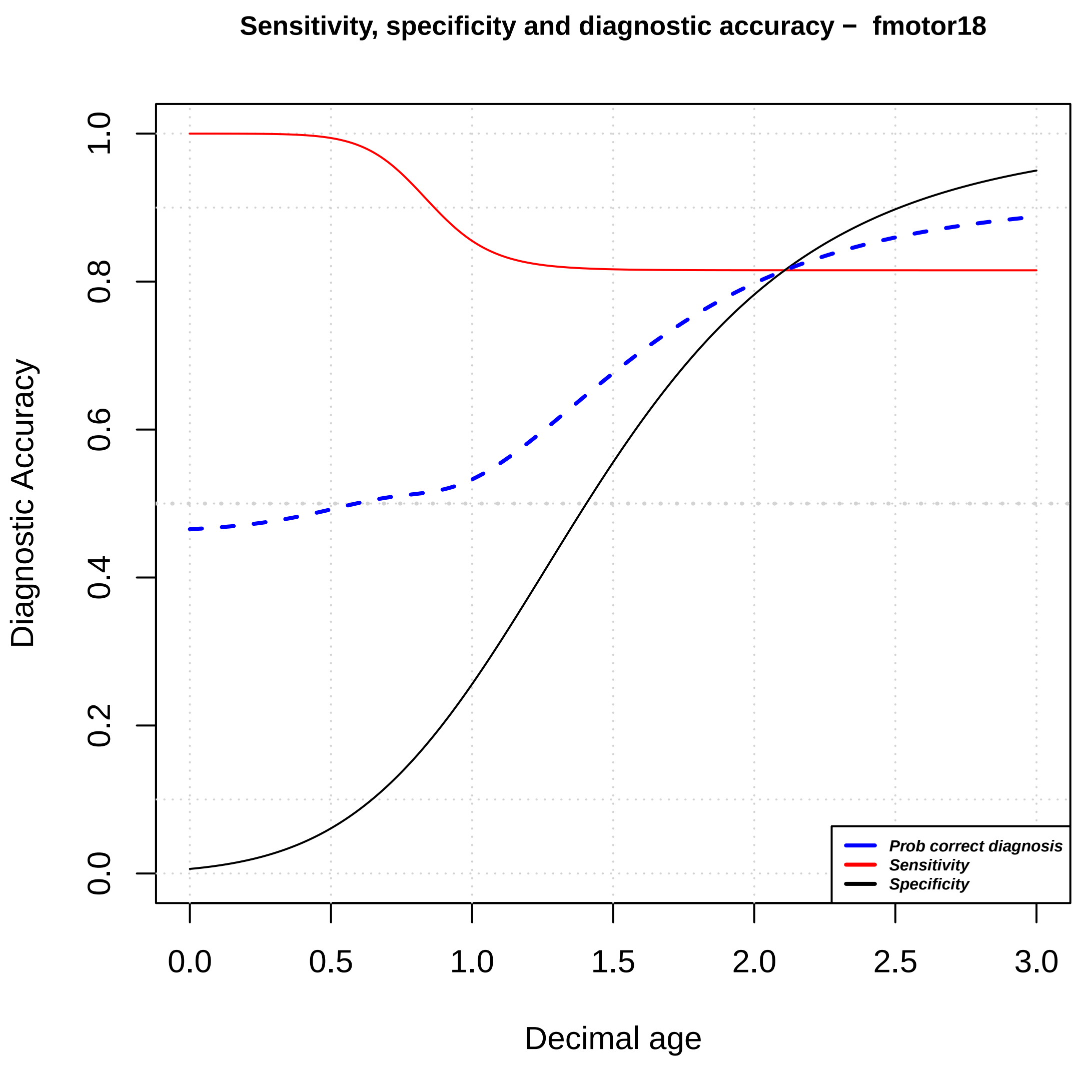
Sensitivity, specificity and diagnostic accuracy – fmotor18
Diagnostic Accuracy
Decimal age
0.0
0.2
0.4
0.6
0.8
1.0
0.0
0.5
1.0
1.5
2.0
2.5
3.0
Prob correct diagnosis
Sensitivity
Specificity

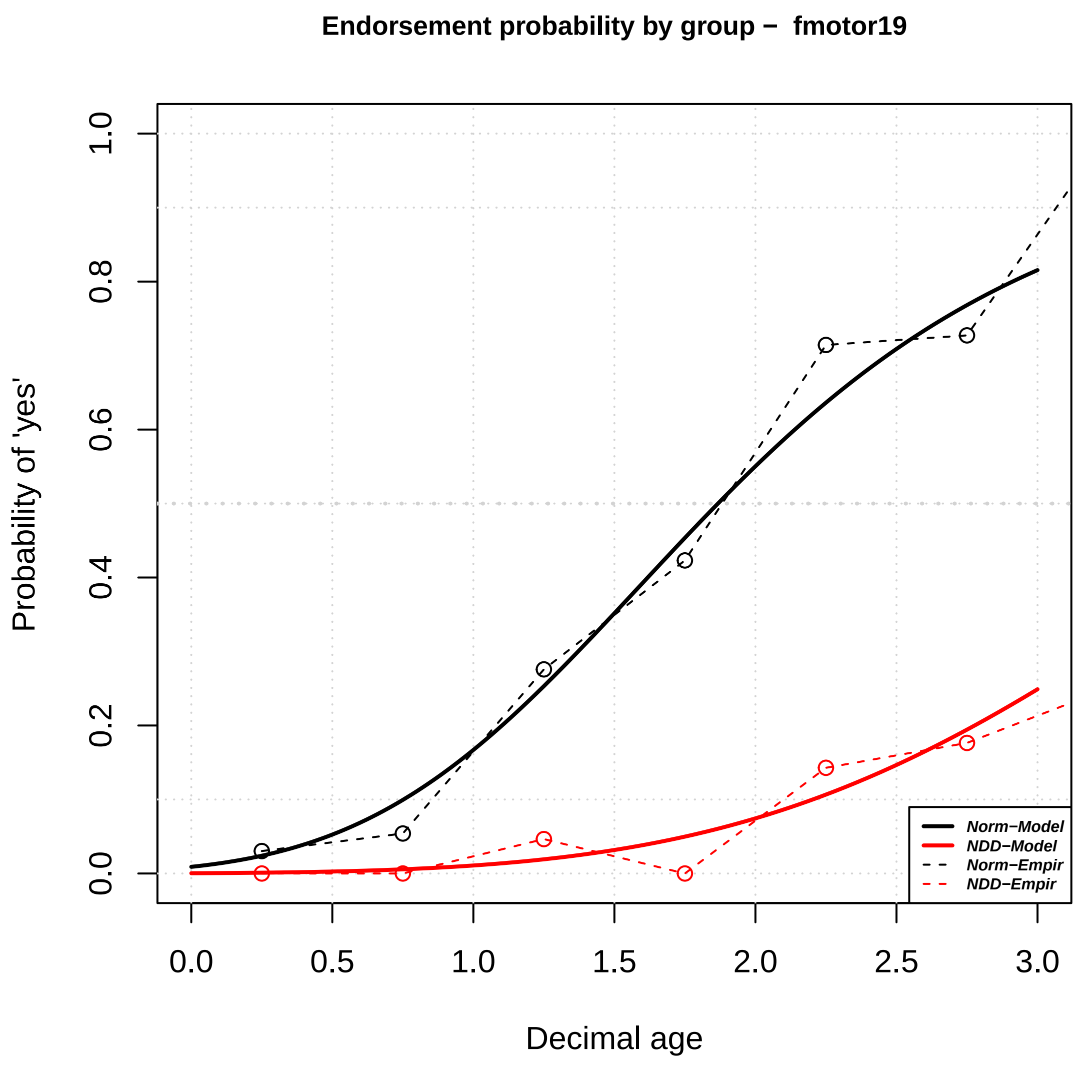

Endorsement probability by group − fmotor19
Probability of 'yes'
Decimal age
0.0
0.2
0.4
0.6
0.8
1.0
0.0
0.5
1.0
1.5
2.0
2.5
3.0
Norm−Model
NDD−Model
Norm−Empir
NDD−Empir


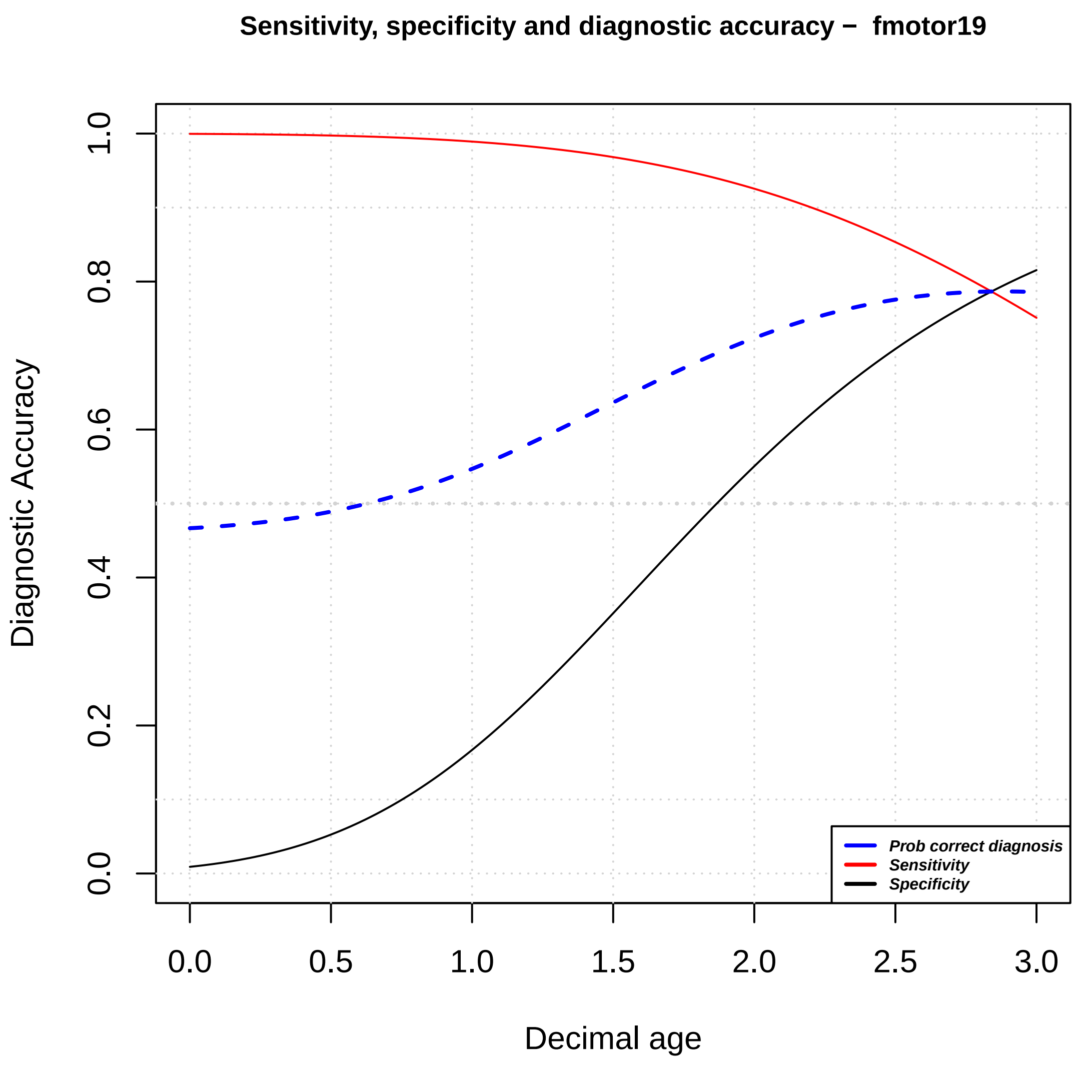

Sensitivity, specificity and diagnostic accuracy − fmotor19
Diagnostic Accuracy
Decimal age
0.0
0.2
0.4
0.6
0.8
1.0
0.0
0.5
1.0
1.5
2.0
2.5
3.0
Prob correct diagnosis
Sensitivity
Specificity

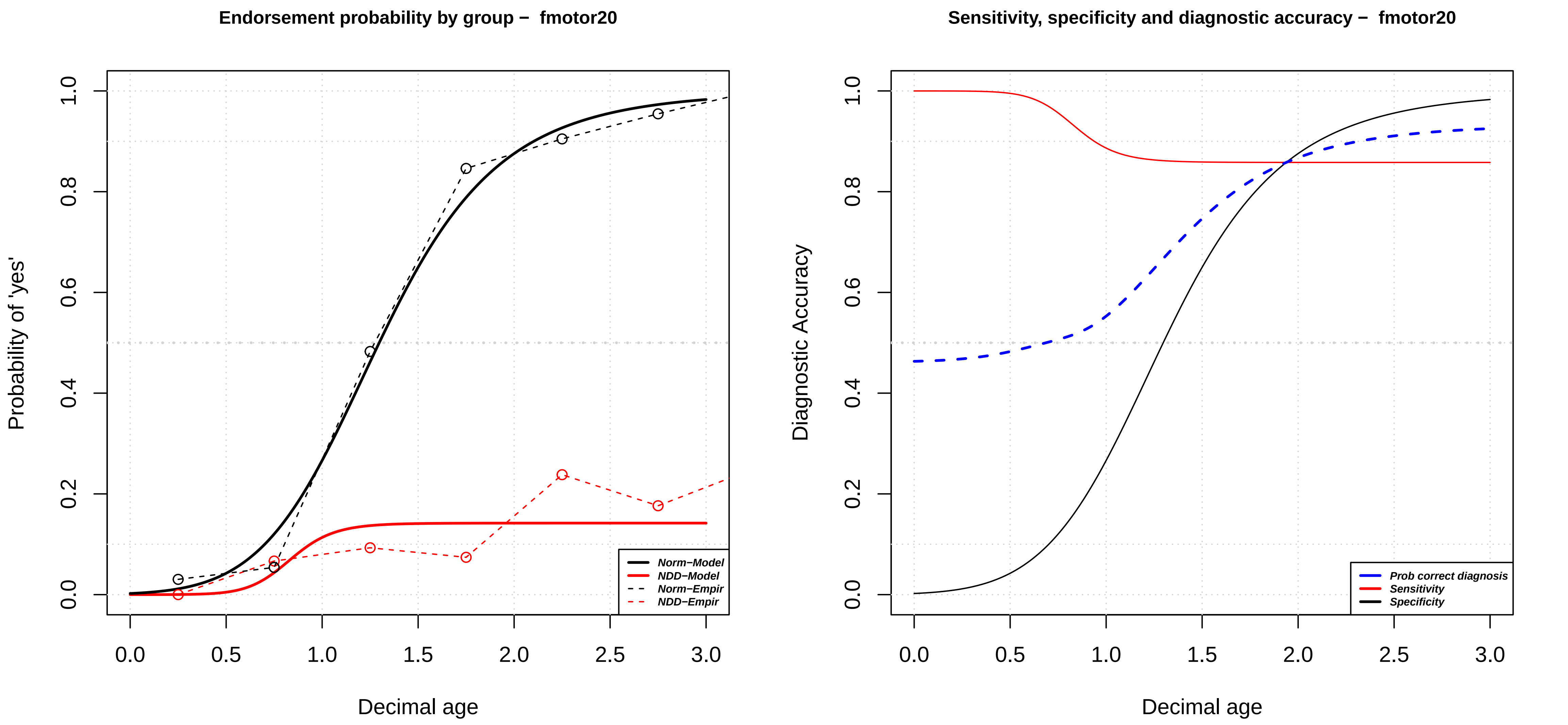

Endorsement probability by group – fmotor20
Probability of 'yes'
Decimal age
Norm−Model
NDD−Model
Norm−Empir
NDD−Empir
Sensitivity, specificity and diagnostic accuracy – fmotor20
Diagnostic Accuracy
Decimal age
Prob correct diagnosis
Sensitivity
Specificity

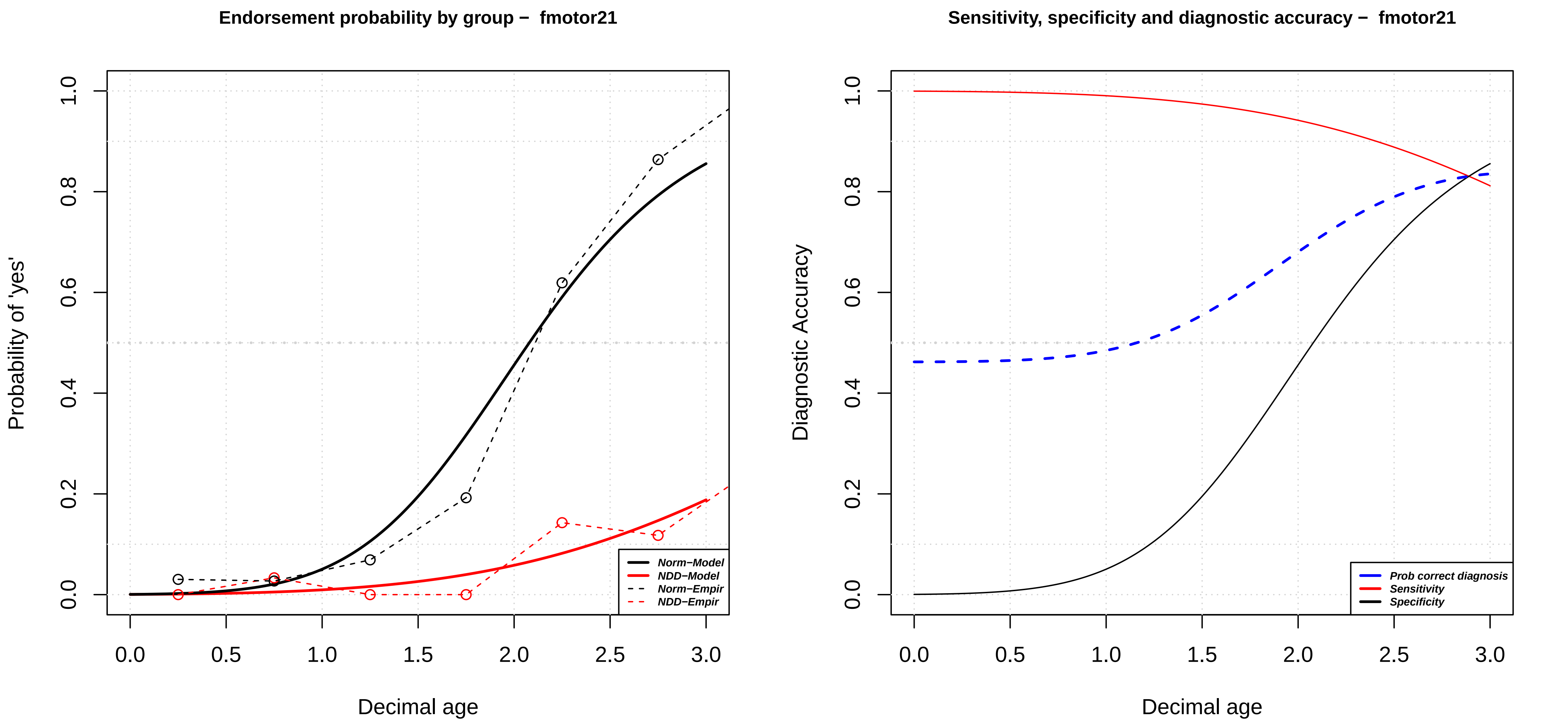
Endorsement probability by group – fmotor21
Probability of 'yes'
Decimal age
Norm−Model
NDD−Model
Norm−Empir
NDD−Empir
Sensitivity, specificity and diagnostic accuracy – fmotor21
Diagnostic Accuracy
Decimal age
Prob correct diagnosis
Sensitivity
Specificity

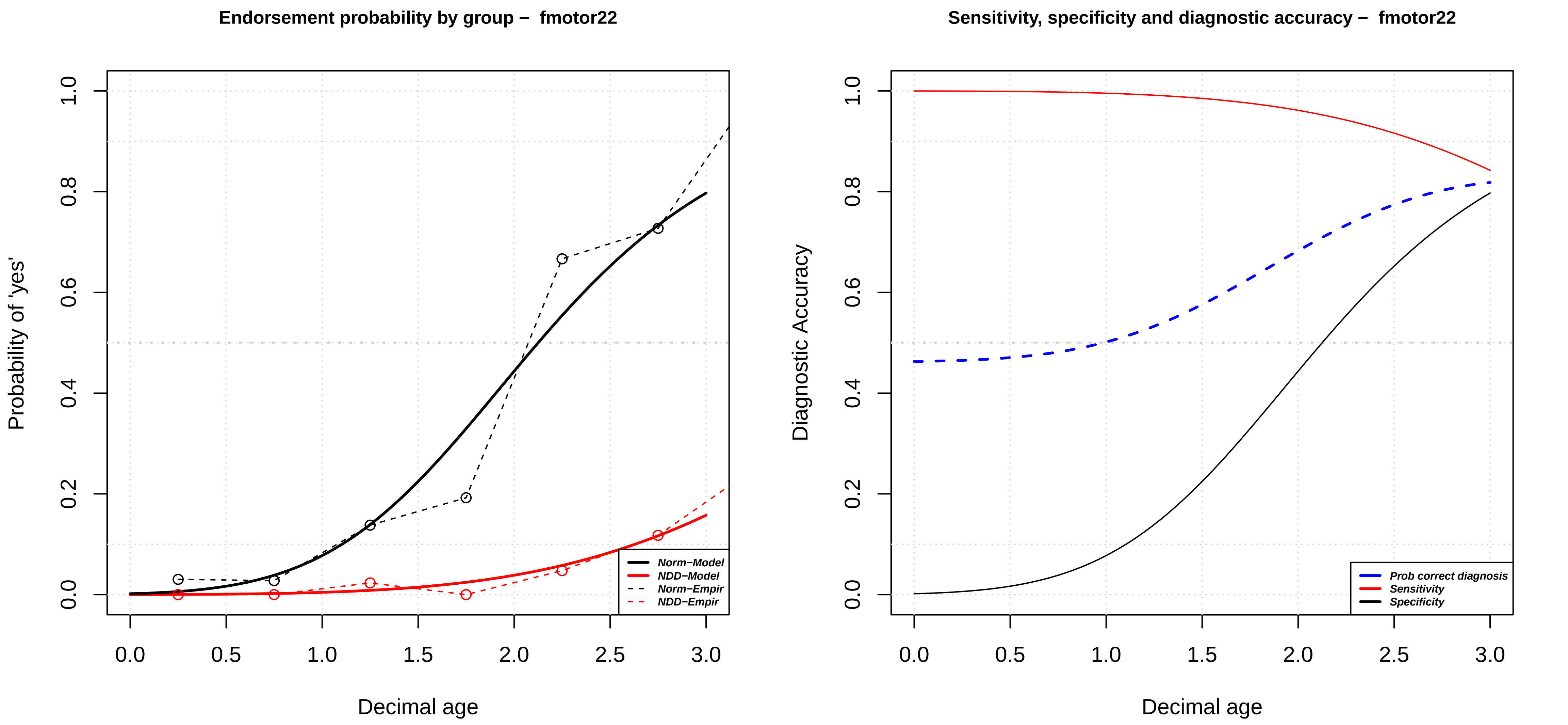
Endorsement probability by group – fmotor22
Probability of 'yes'
Decimal age
Norm−Model
NDD−Model
Norm−Empir
NDD−Empir
0.0
0.2
0.4
0.6
0.8
1.0
0.0
0.5
1.0
1.5
2.0
2.5
3.0
Sensitivity, specificity and diagnostic accuracy – fmotor22
Diagnostic Accuracy
Decimal age
Prob correct diagnosis
Sensitivity
Specificity

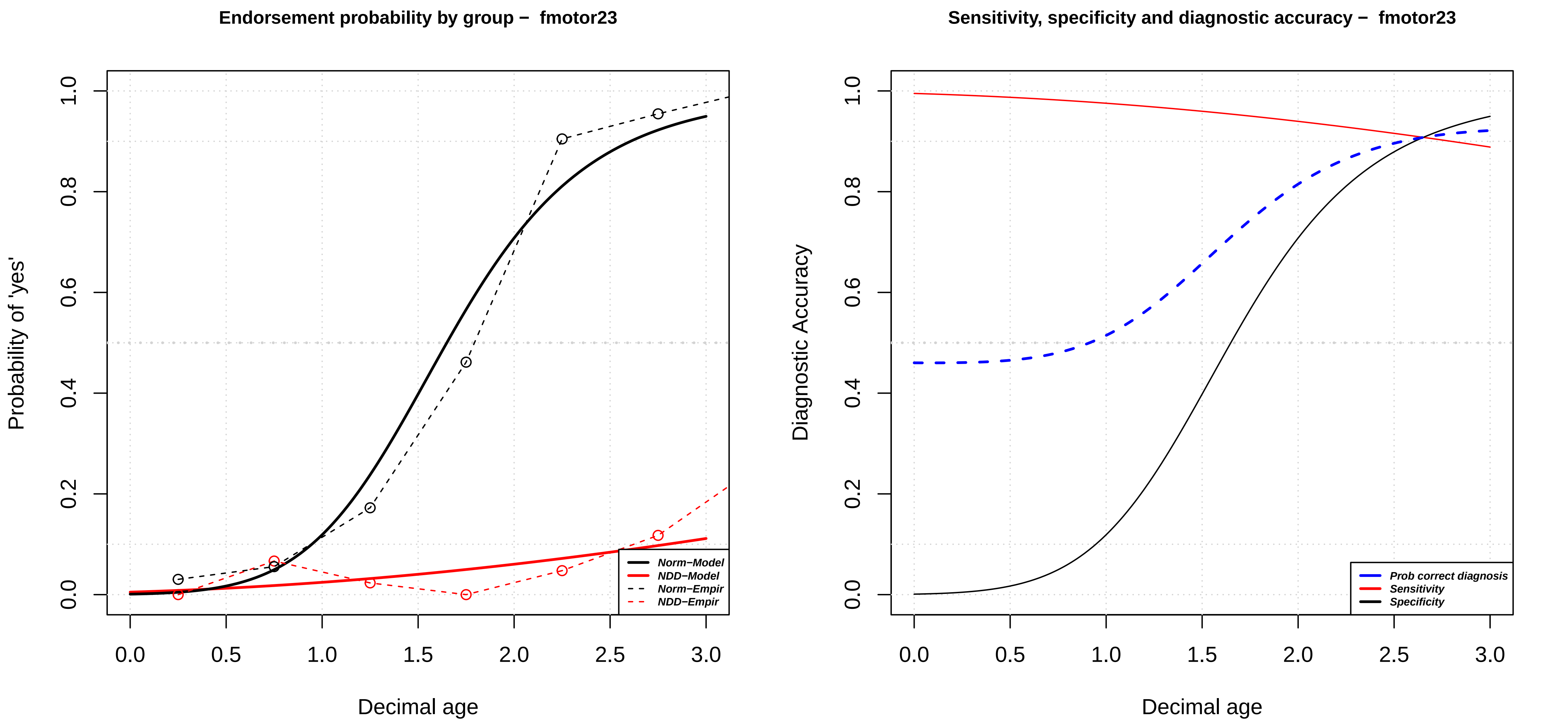

Endorsement probability by group – fmotor23
Probability of 'yes'
1.0
0.8
0.6
0.4
0.2
0.0
0.0
0.5
1.0
1.5
2.0
2.5
3.0
Decimal age
Norm−Model
NDD−Model
Norm−Empir
NDD−Empir
Sensitivity, specificity and diagnostic accuracy – fmotor23
Diagnostic Accuracy
1.0
0.8
0.6
0.4
0.2
0.0
0.0
0.5
1.0
1.5
2.0
2.5
3.0
Decimal age
Prob correct diagnosis
Sensitivity
Specificity

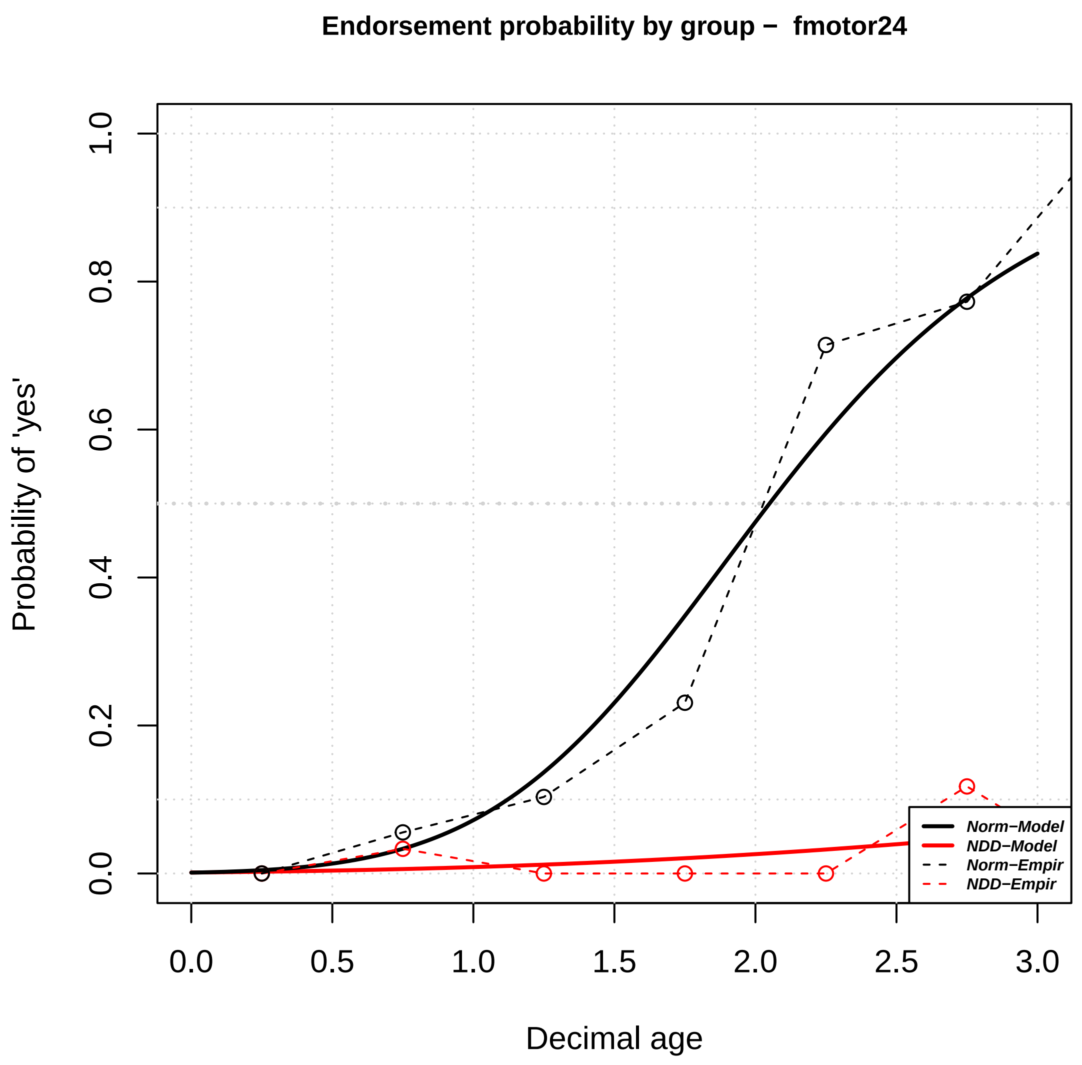
Endorsement probability by group − fmotor24
Probability of 'yes'
Decimal age
0.0
0.2
0.4
0.6
0.8
1.0
0.0
0.5
1.0
1.5
2.0
2.5
3.0
Norm−Model
NDD−Model
Norm−Empir
NDD−Empir

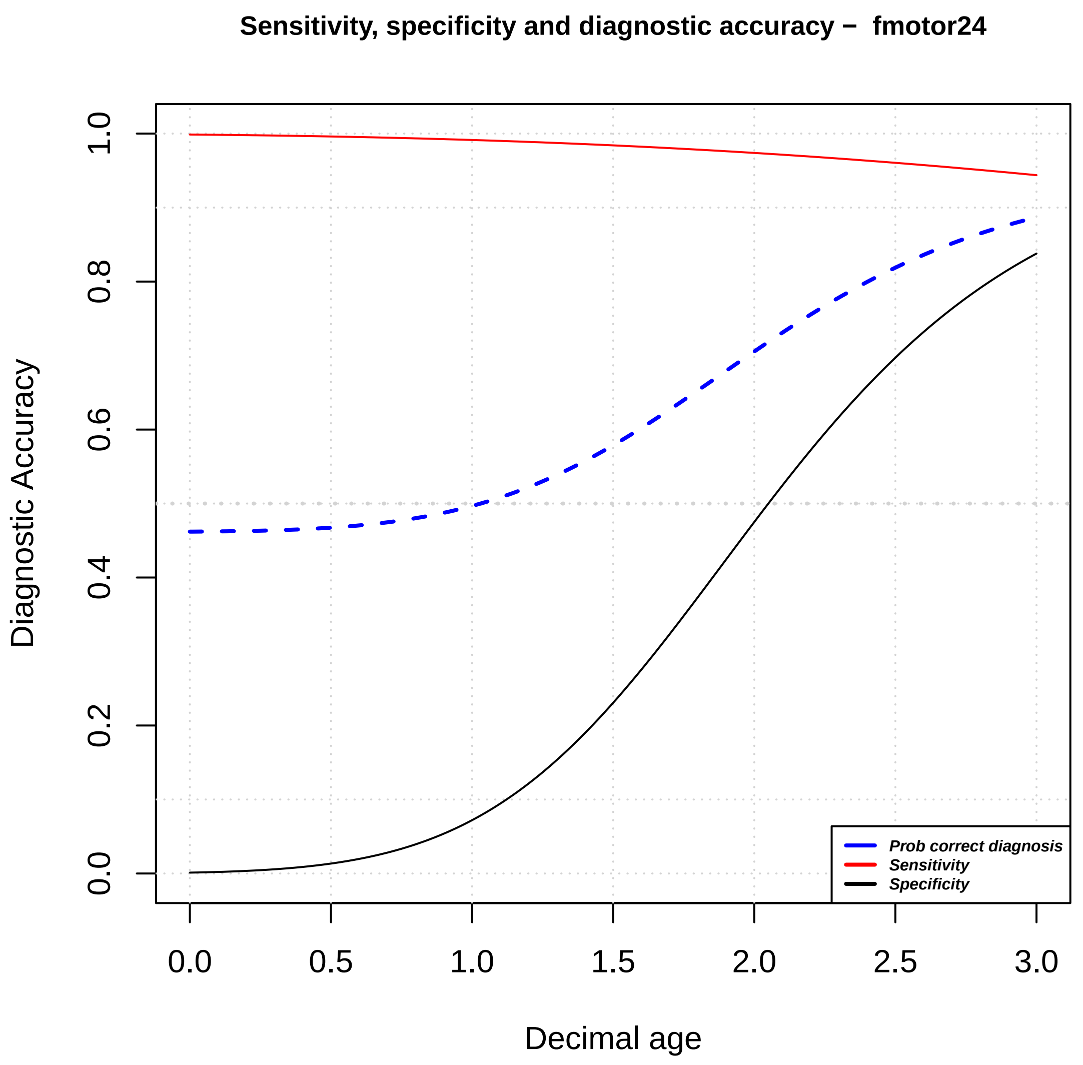
Sensitivity, specificity and diagnostic accuracy − fmotor24
Diagnostic Accuracy
Decimal age
0.0
0.2
0.4
0.6
0.8
1.0
0.0
0.5
1.0
1.5
2.0
2.5
3.0
Prob correct diagnosis
Sensitivity
Specificity

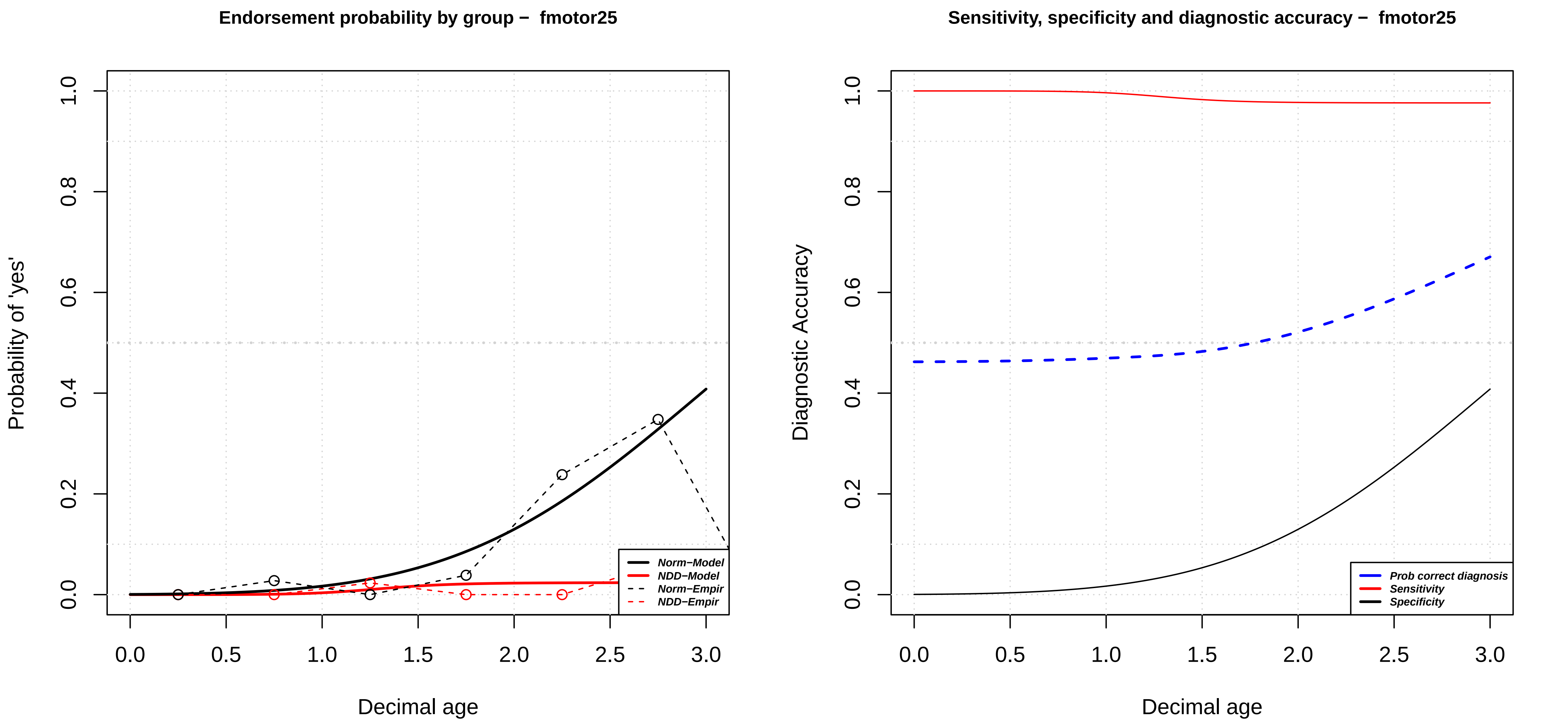

Endorsement probability by group – fmotor25
Probability of 'yes'
Decimal age
Norm−Model
NDD−Model
Norm−Empir
NDD−Empir
Sensitivity, specificity and diagnostic accuracy – fmotor25
Diagnostic Accuracy
Decimal age
Prob correct diagnosis
Sensitivity
Specificity

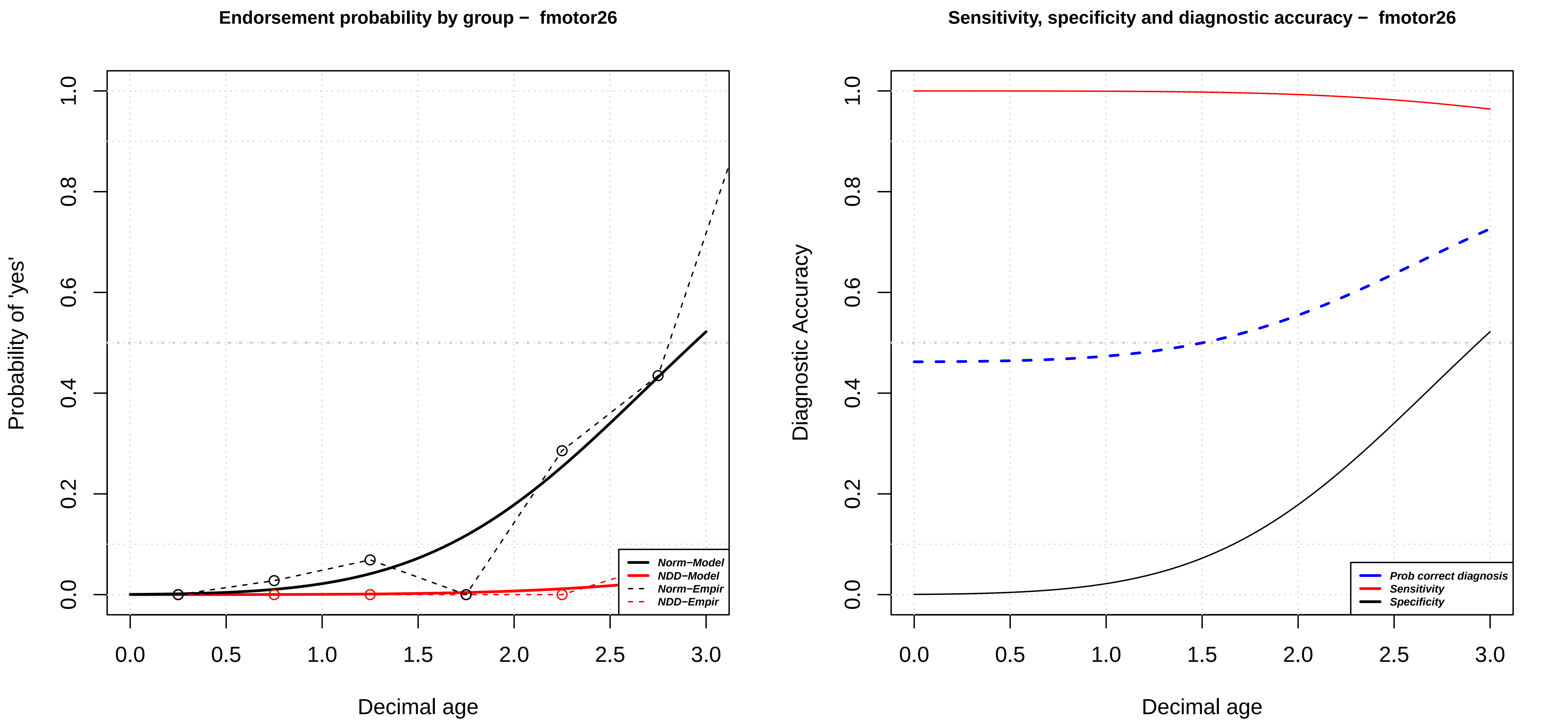

Endorsement probability by group – fmotor26
Probability of 'yes'
0.0
0.2
0.4
0.6
0.8
1.0
0.0
0.5
1.0
1.5
2.0
2.5
3.0
Decimal age
Norm–Model
NDD–Model
Norm–Empir
NDD–Empir
Sensitivity, specificity and diagnostic accuracy – fmotor26
Diagnostic Accuracy
Decimal age
Prob correct diagnosis
Sensitivity
Specificity

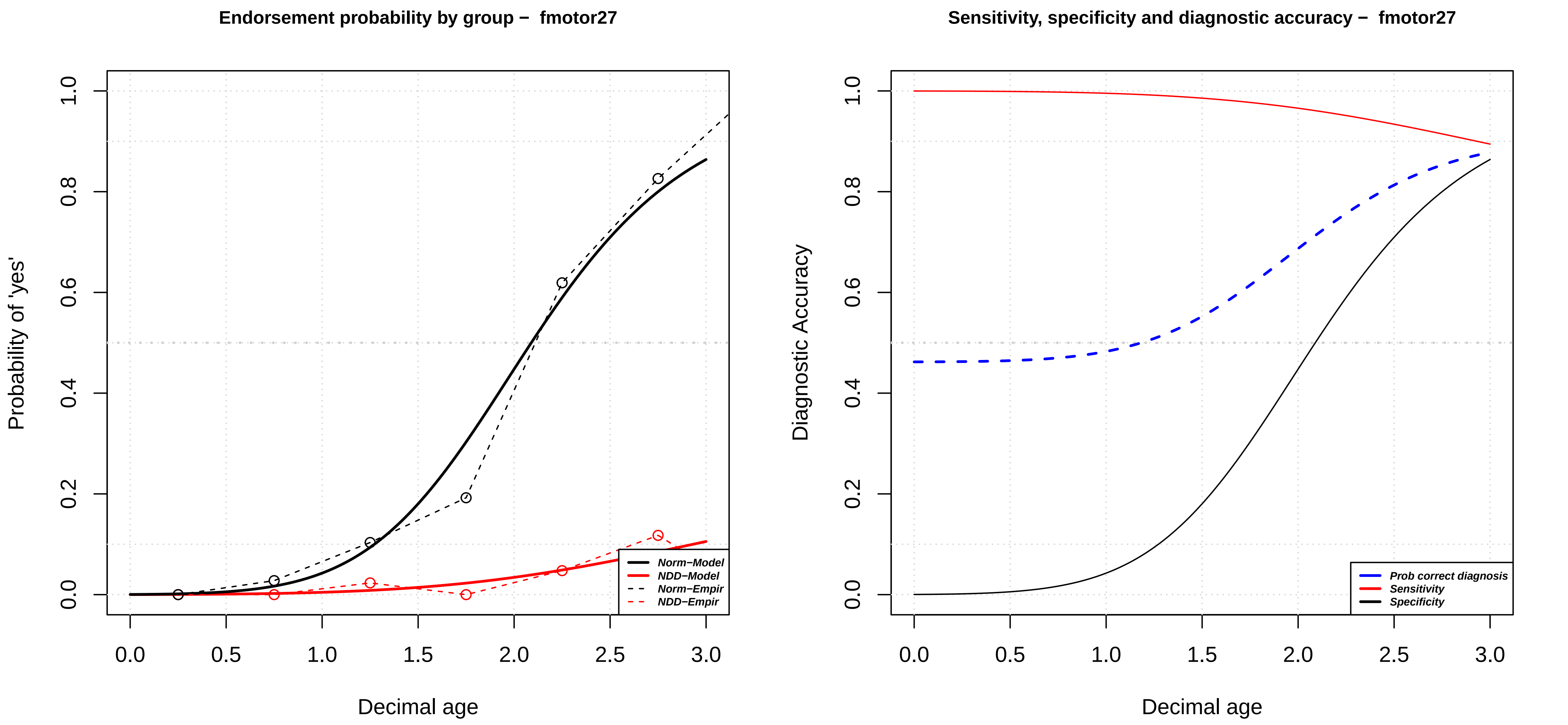

Endorsement probability by group – fmotor27
Probability of 'yes'
Decimal age
0.0
0.2
0.4
0.6
0.8
1.0
0.0
0.5
1.0
1.5
2.0
2.5
3.0
Norm−Model
NDD−Model
Norm−Empir
NDD−Empir
Sensitivity, specificity and diagnostic accuracy – fmotor27
Diagnostic Accuracy
Decimal age
Prob correct diagnosis
Sensitivity
Specificity

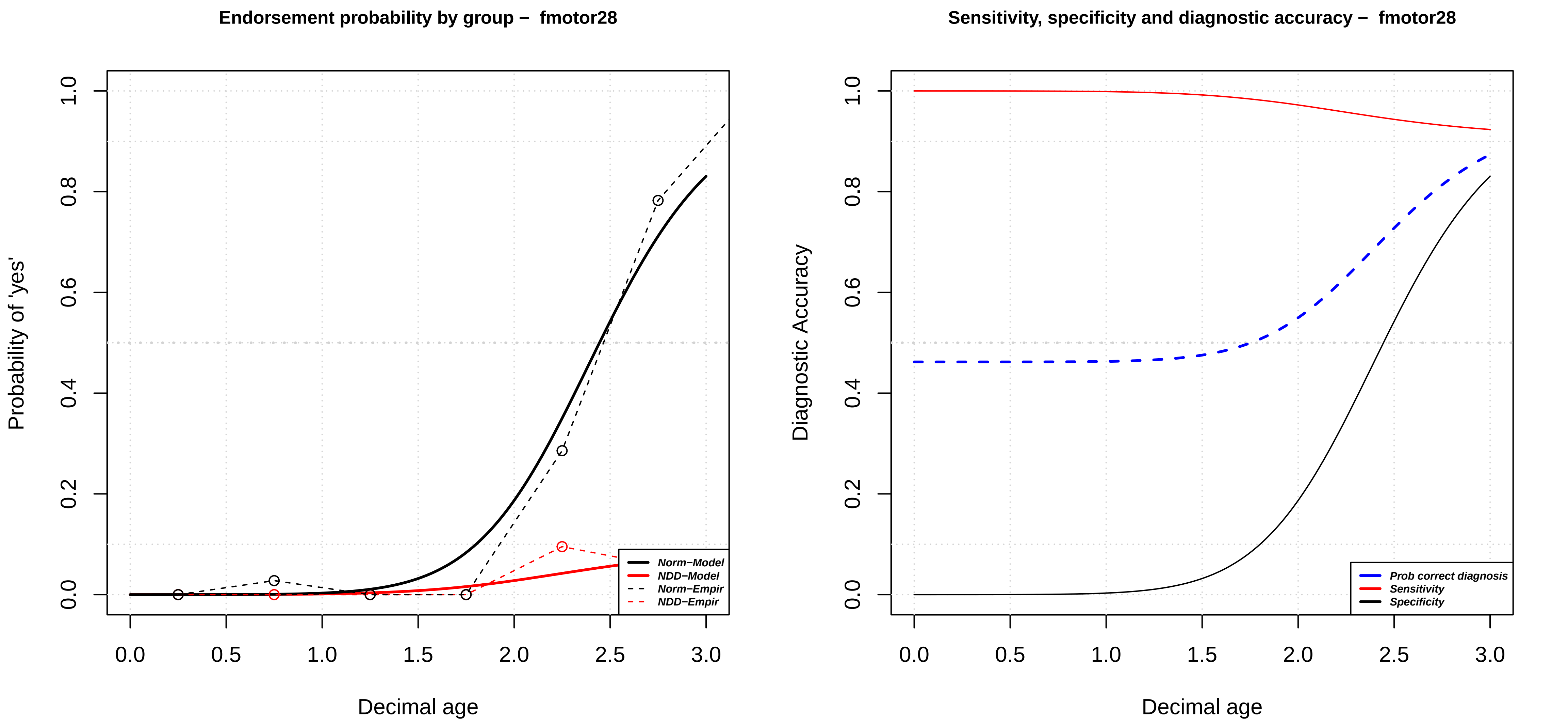
Endorsement probability by group – fmotor28
Probability of 'yes'
Decimal age
0.0
0.2
0.4
0.6
0.8
1.0
0.0
0.5
1.0
1.5
2.0
2.5
3.0
Norm–Model
NDD–Model
Norm–Empir
NDD–Empir
Sensitivity, specificity and diagnostic accuracy – fmotor28
Diagnostic Accuracy
Decimal age
Prob correct diagnosis
Sensitivity
Specificity

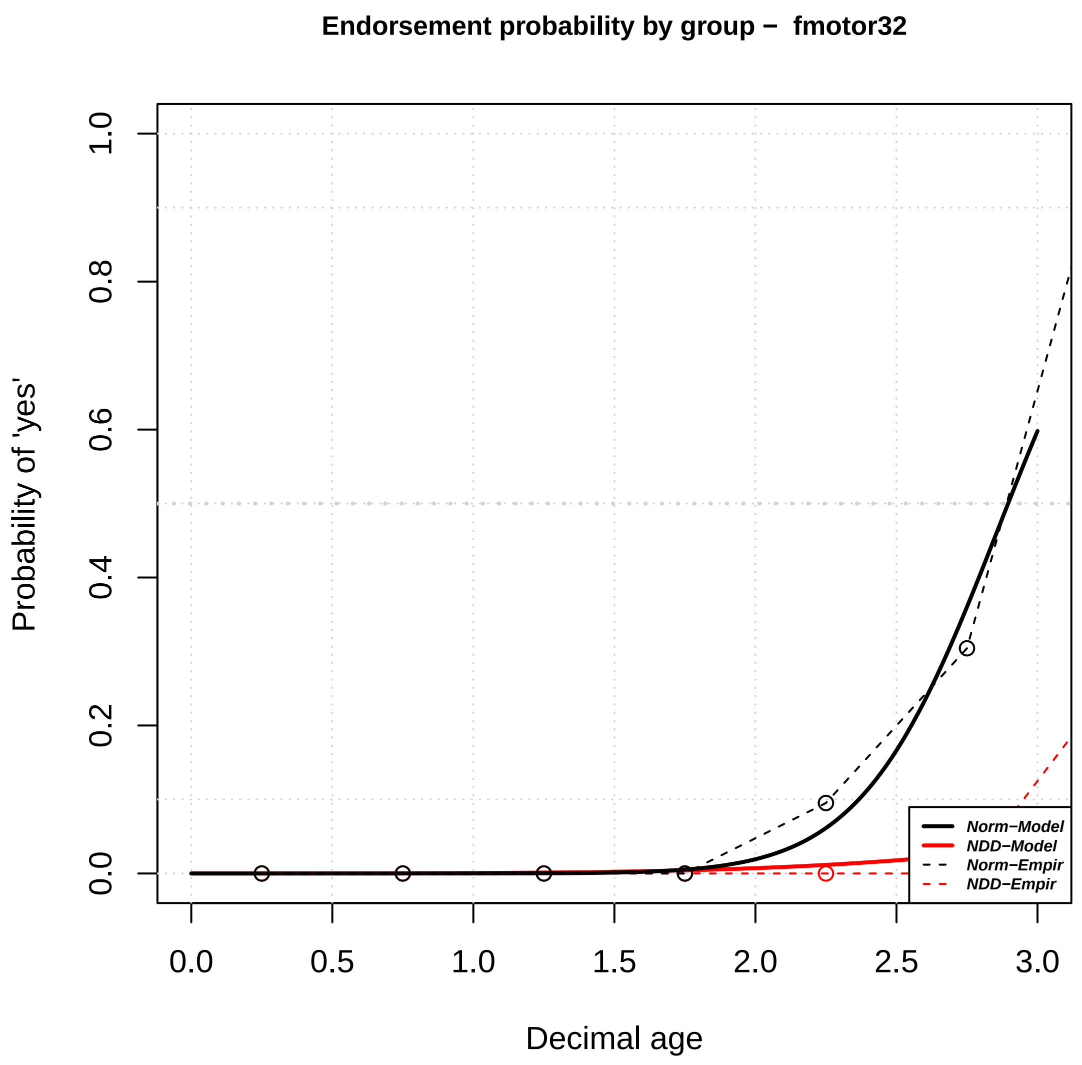
Endorsement probability by group − fmotor32
Probability of 'yes'
0.0
0.2
0.4
0.6
0.8
1.0
0.0
0.5
1.0
1.5
2.0
2.5
3.0
Decimal age
Norm−Model
NDD−Model
Norm−Empir
NDD−Empir

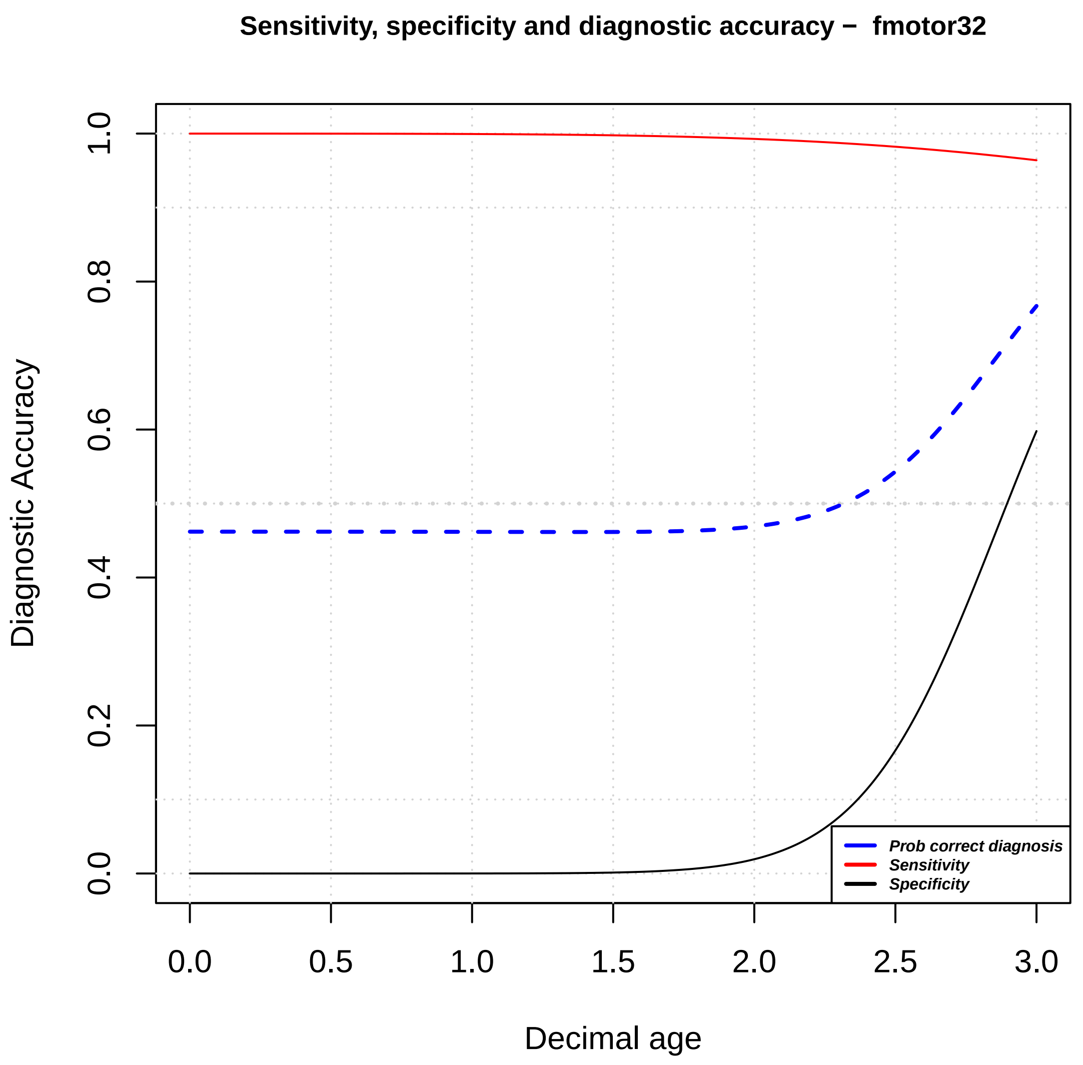
Sensitivity, specificity and diagnostic accuracy − fmotor32
Diagnostic Accuracy
0.0
0.2
0.4
0.6
0.8
1.0
0.0
0.5
1.0
1.5
2.0
2.5
3.0
Decimal age
Prob correct diagnosis
Sensitivity
Specificity

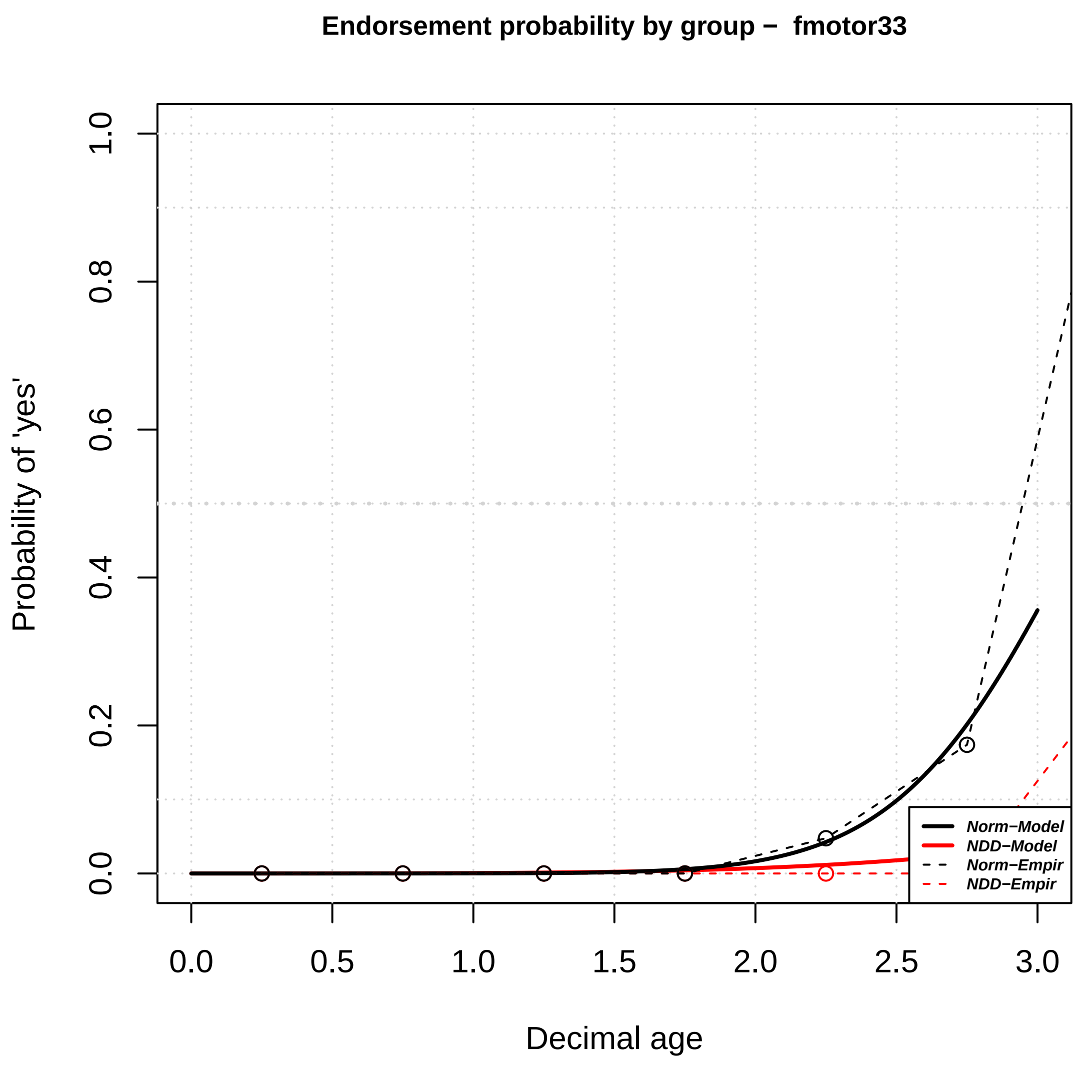

Endorsement probability by group – fmotor33
Probability of 'yes'
Decimal age
0.0
0.2
0.4
0.6
0.8
1.0
0.0
0.5
1.0
1.5
2.0
2.5
3.0
Norm–Model
NDD–Model
Norm–Empir
NDD–Empir


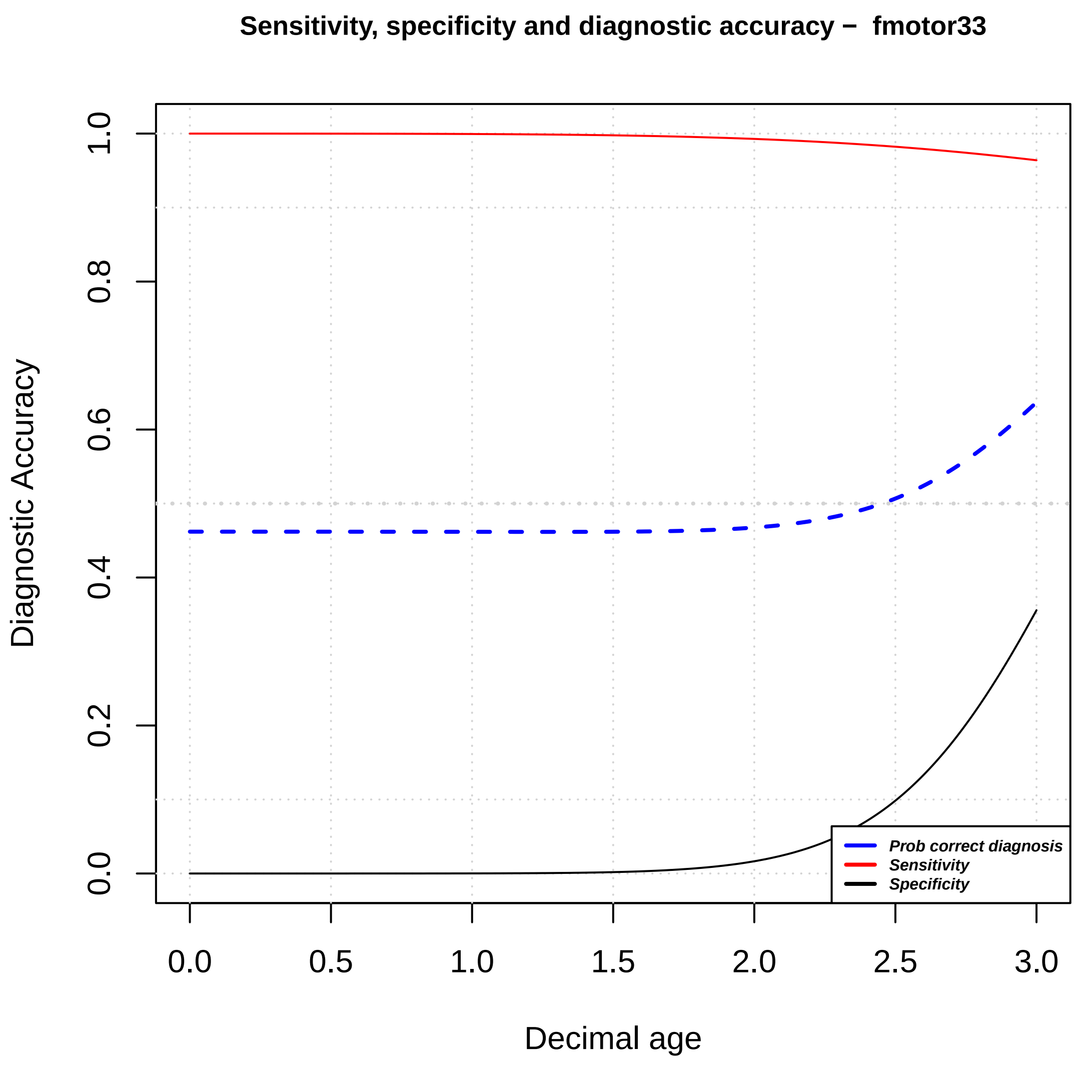

Sensitivity, specificity and diagnostic accuracy – fmotor33
Diagnostic Accuracy
Decimal age
0.0
0.2
0.4
0.6
0.8
1.0
0.0
0.5
1.0
1.5
2.0
2.5
3.0
Prob correct diagnosis
Sensitivity
Specificity

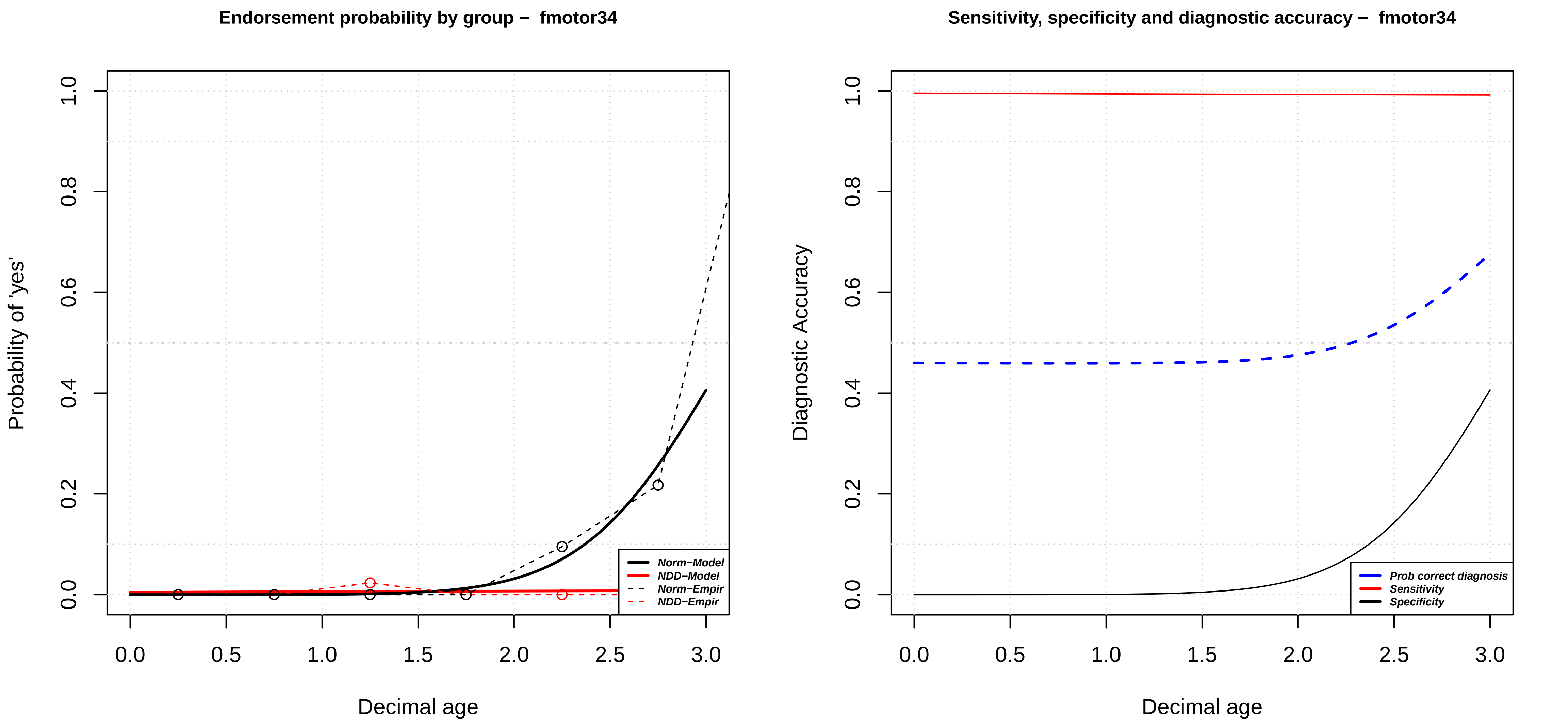

Endorsement probability by group – fmotor34
Probability of 'yes'
Decimal age
Norm−Model
NDD−Model
Norm−Empir
NDD−Empir
0.0
0.2
0.4
0.6
0.8
1.0
0.0
0.5
1.0
1.5
2.0
2.5
3.0
Sensitivity, specificity and diagnostic accuracy – fmotor34
Diagnostic Accuracy
Decimal age
Prob correct diagnosis
Sensitivity
Specificity

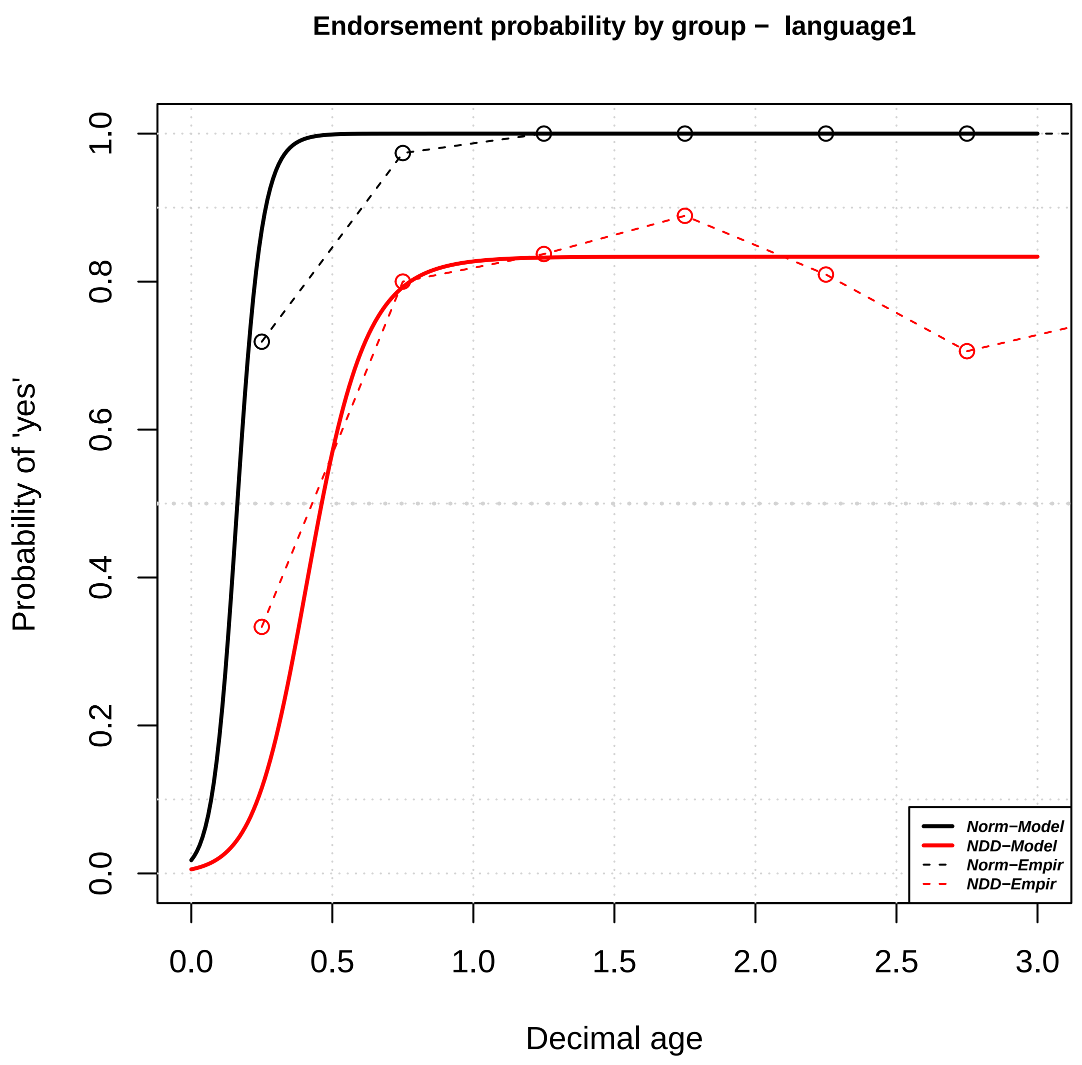

Endorsement probability by group − language1
Probability of 'yes'
Decimal age
0.0
0.2
0.4
0.6
0.8
1.0
0.0
0.5
1.0
1.5
2.0
2.5
3.0
Norm−Model
NDD−Model
Norm−Empir
NDD−Empir


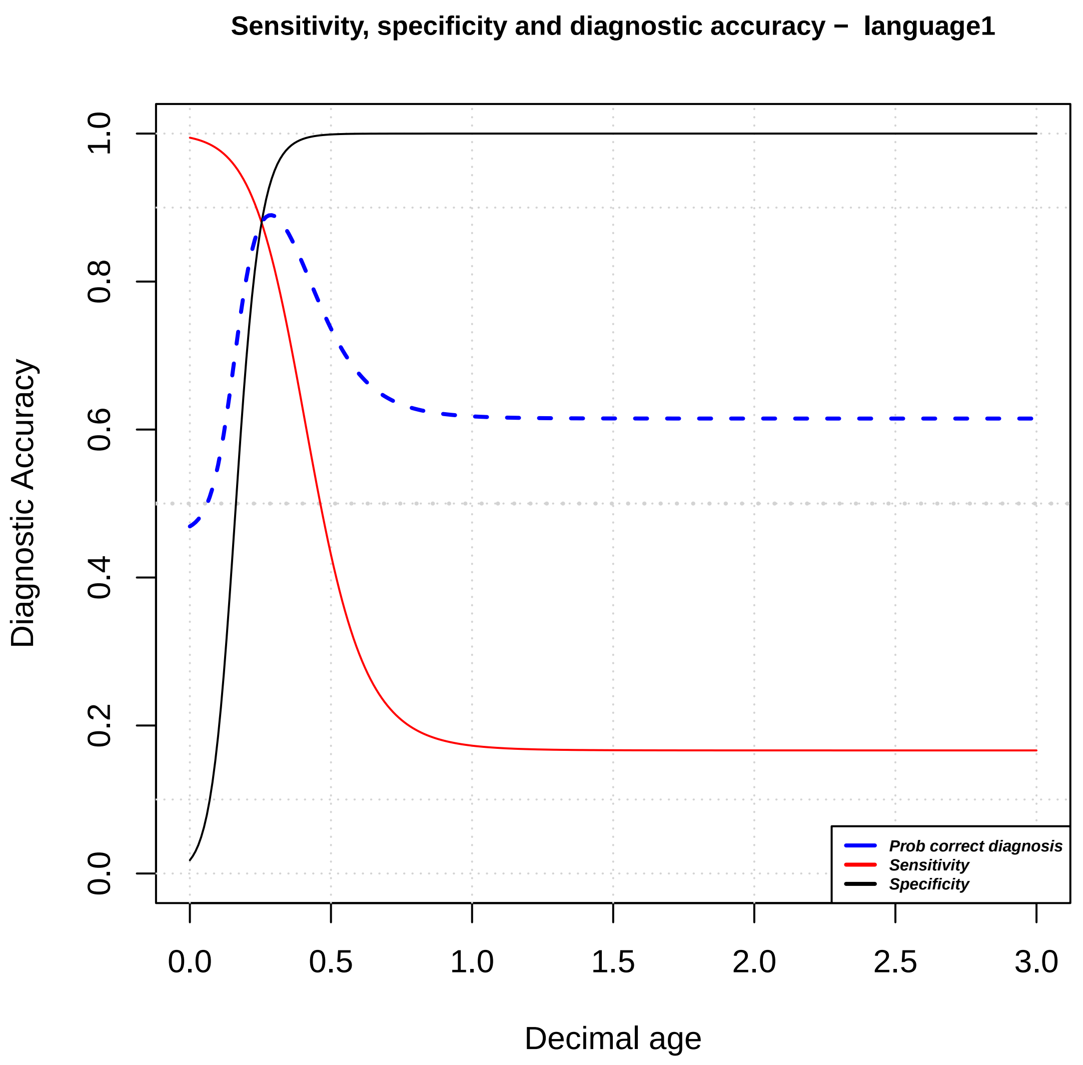

Sensitivity, specificity and diagnostic accuracy − language1
Diagnostic Accuracy
Decimal age
0.0
0.2
0.4
0.6
0.8
1.0
0.0
0.5
1.0
1.5
2.0
2.5
3.0
Prob correct diagnosis
Sensitivity
Specificity

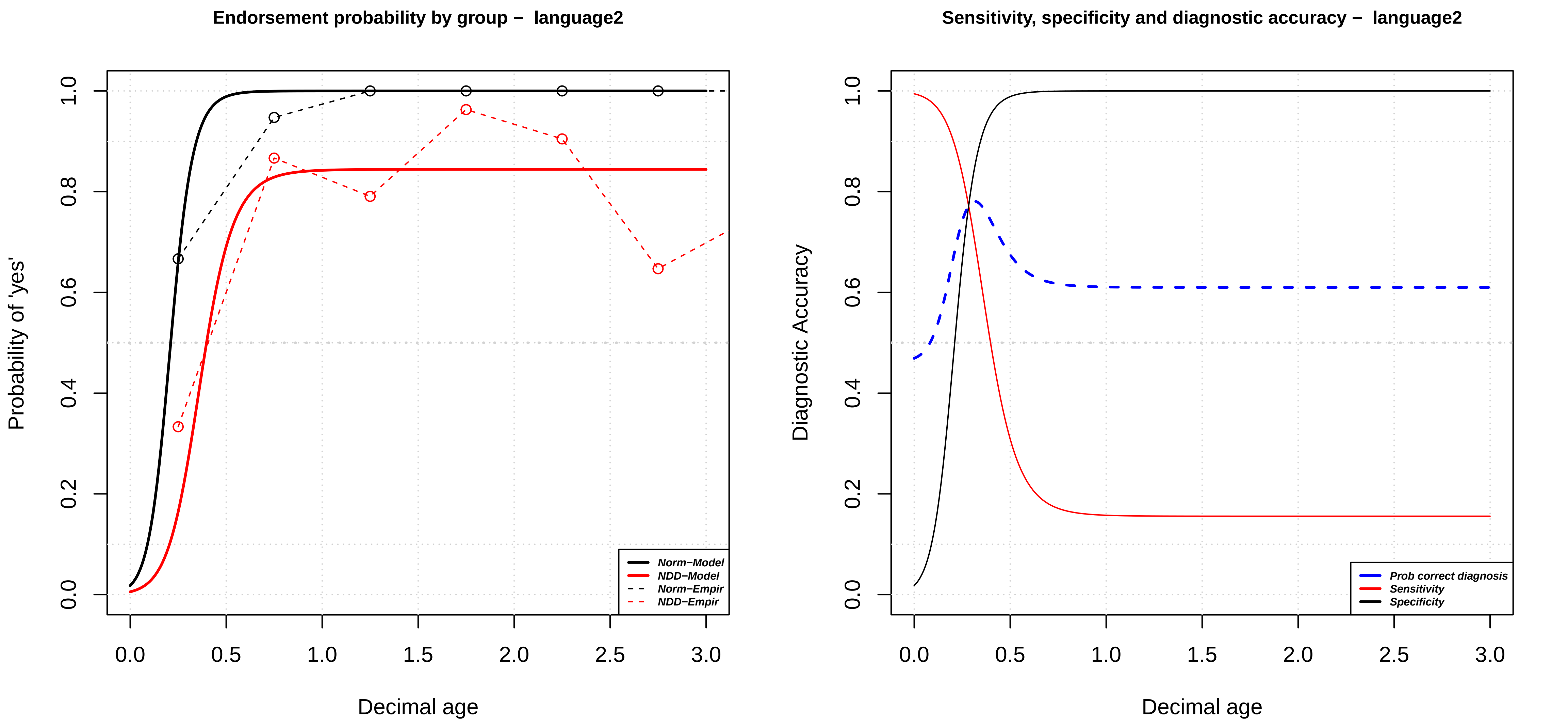

Endorsement probability by group – language2
Probability of 'yes'
Decimal age
Norm–Model
NDD–Model
Norm–Empir
NDD–Empir
Sensitivity, specificity and diagnostic accuracy – language2
Diagnostic Accuracy
Decimal age
Prob correct diagnosis
Sensitivity
Specificity

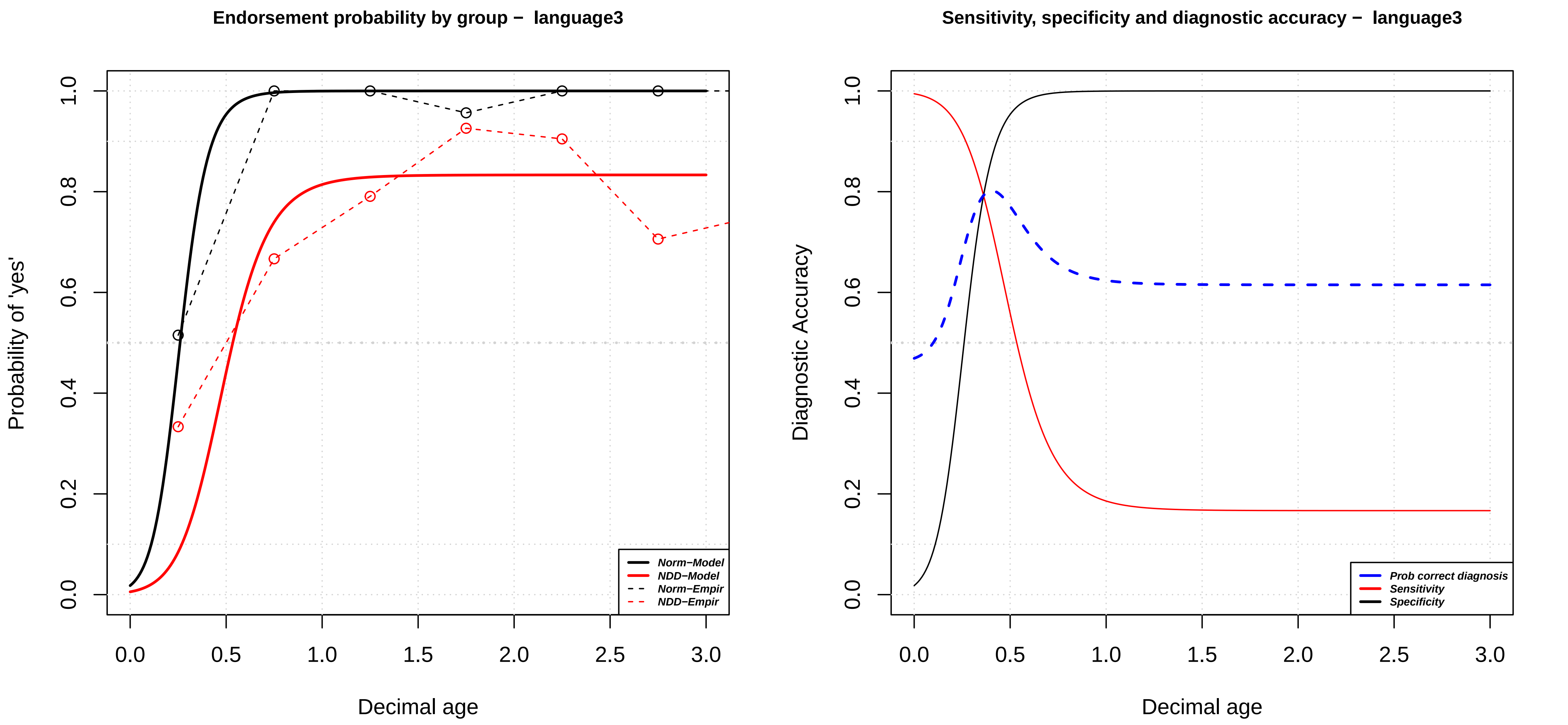

Endorsement probability by group – language3
Probability of 'yes'
Decimal age
Norm–Model
NDD–Model
Norm–Empir
NDD–Empir
0.0
0.2
0.4
0.6
0.8
1.0
0.0
0.5
1.0
1.5
2.0
2.5
3.0
Sensitivity, specificity and diagnostic accuracy – language3
Diagnostic Accuracy
Decimal age
Prob correct diagnosis
Sensitivity
Specificity

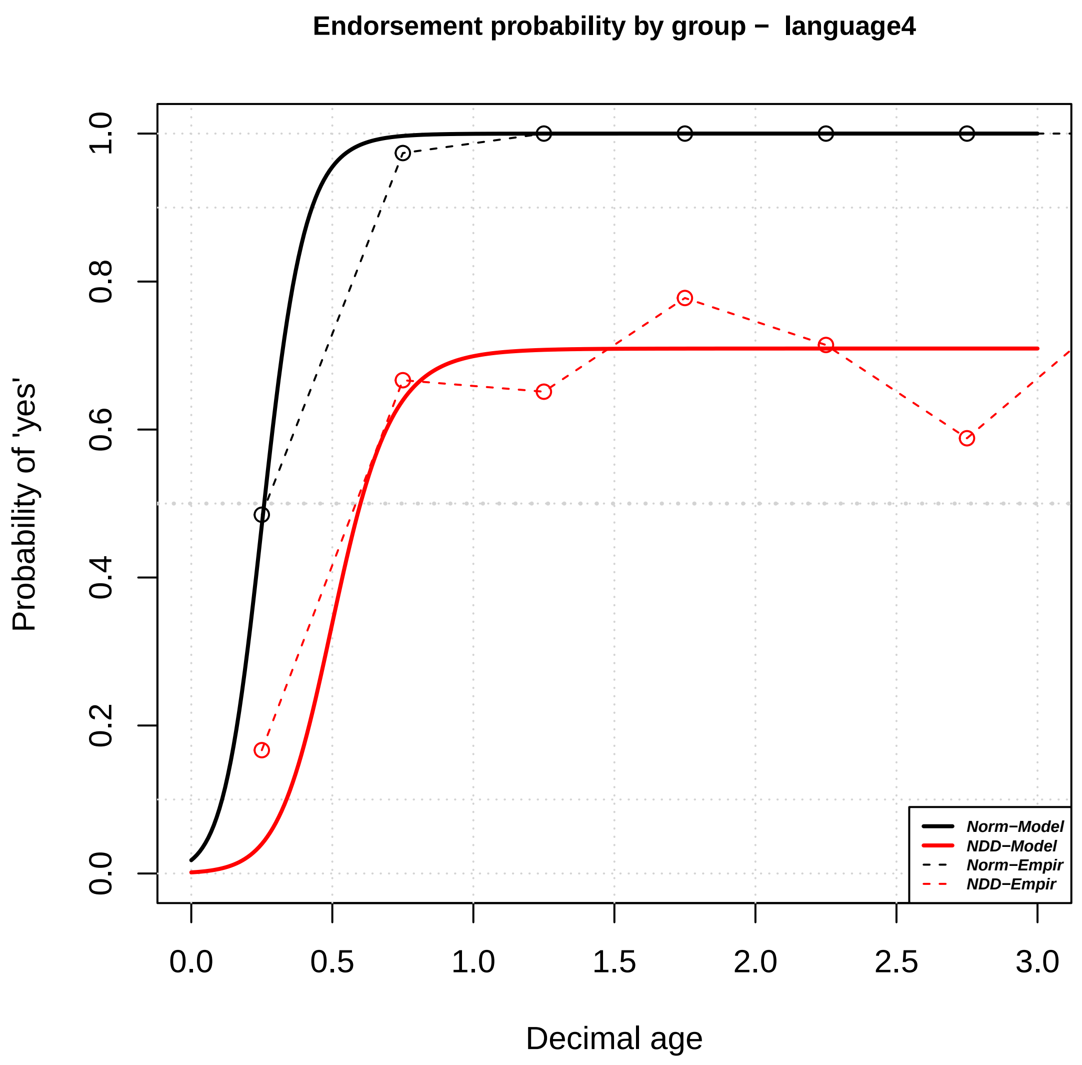
Endorsement probability by group − language4
Probability of 'yes'
Decimal age
0.0
0.2
0.4
0.6
0.8
1.0
0.0
0.5
1.0
1.5
2.0
2.5
3.0
Norm−Model
NDD−Model
Norm−Empir
NDD−Empir

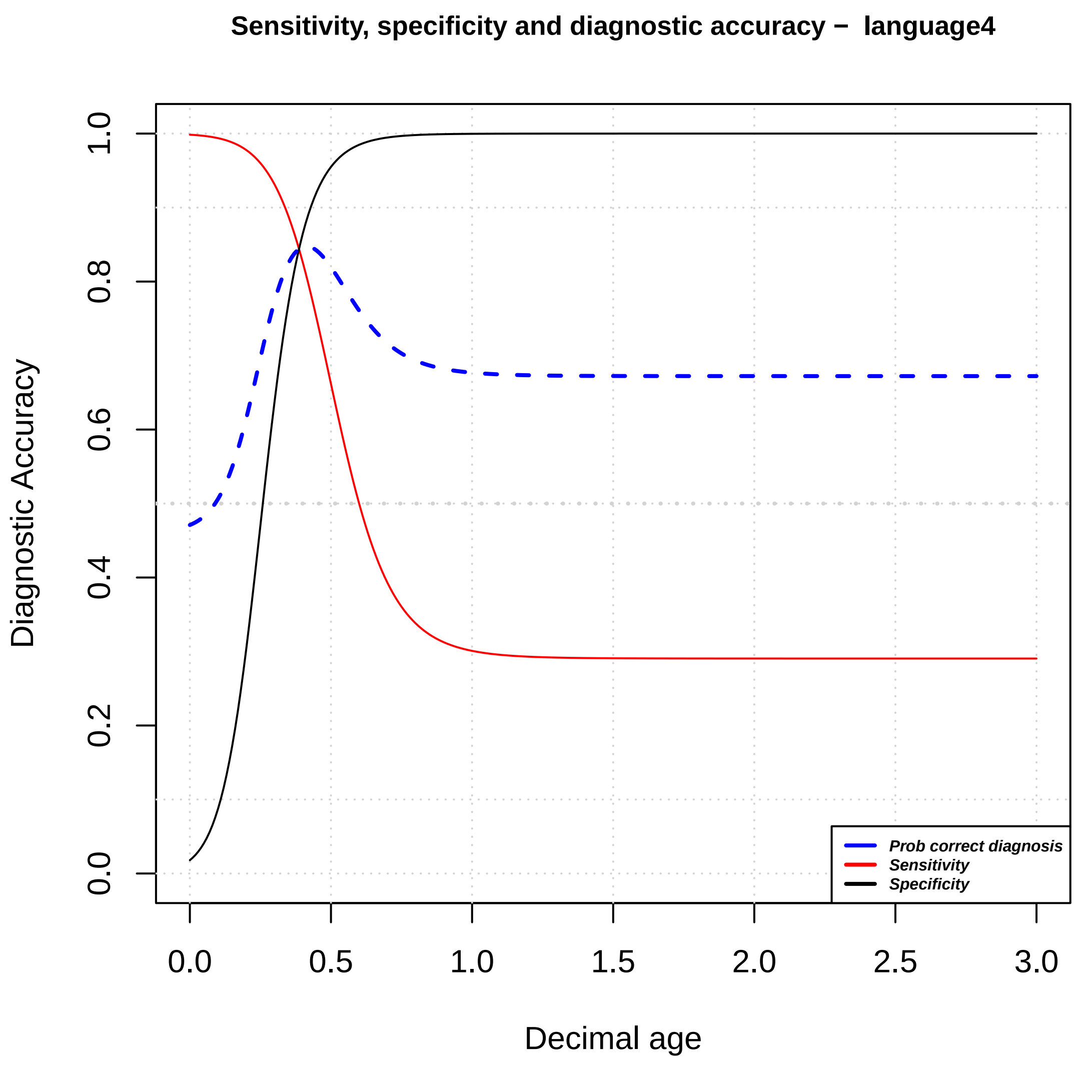
Sensitivity, specificity and diagnostic accuracy − language4
Diagnostic Accuracy
Decimal age
0.0
0.2
0.4
0.6
0.8
1.0
0.0
0.5
1.0
1.5
2.0
2.5
3.0
Prob correct diagnosis
Sensitivity
Specificity

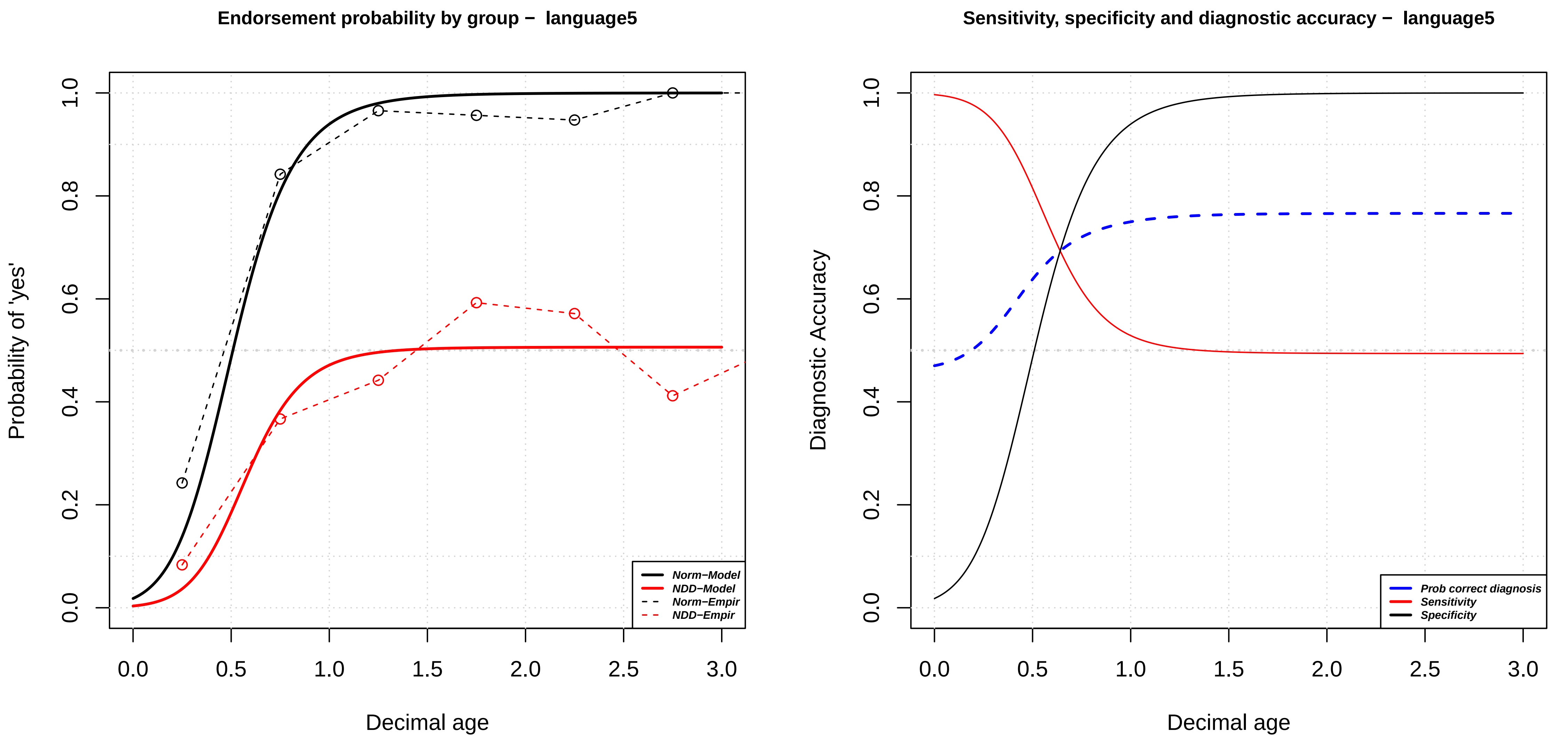

Endorsement probability by group – language5
Probability of 'yes'
Decimal age
0.0
0.2
0.4
0.6
0.8
1.0
0.0
0.5
1.0
1.5
2.0
2.5
3.0
Norm–Model
NDD–Model
Norm–Empir
NDD–Empir
Sensitivity, specificity and diagnostic accuracy – language5
Diagnostic Accuracy
Decimal age
Prob correct diagnosis
Sensitivity
Specificity

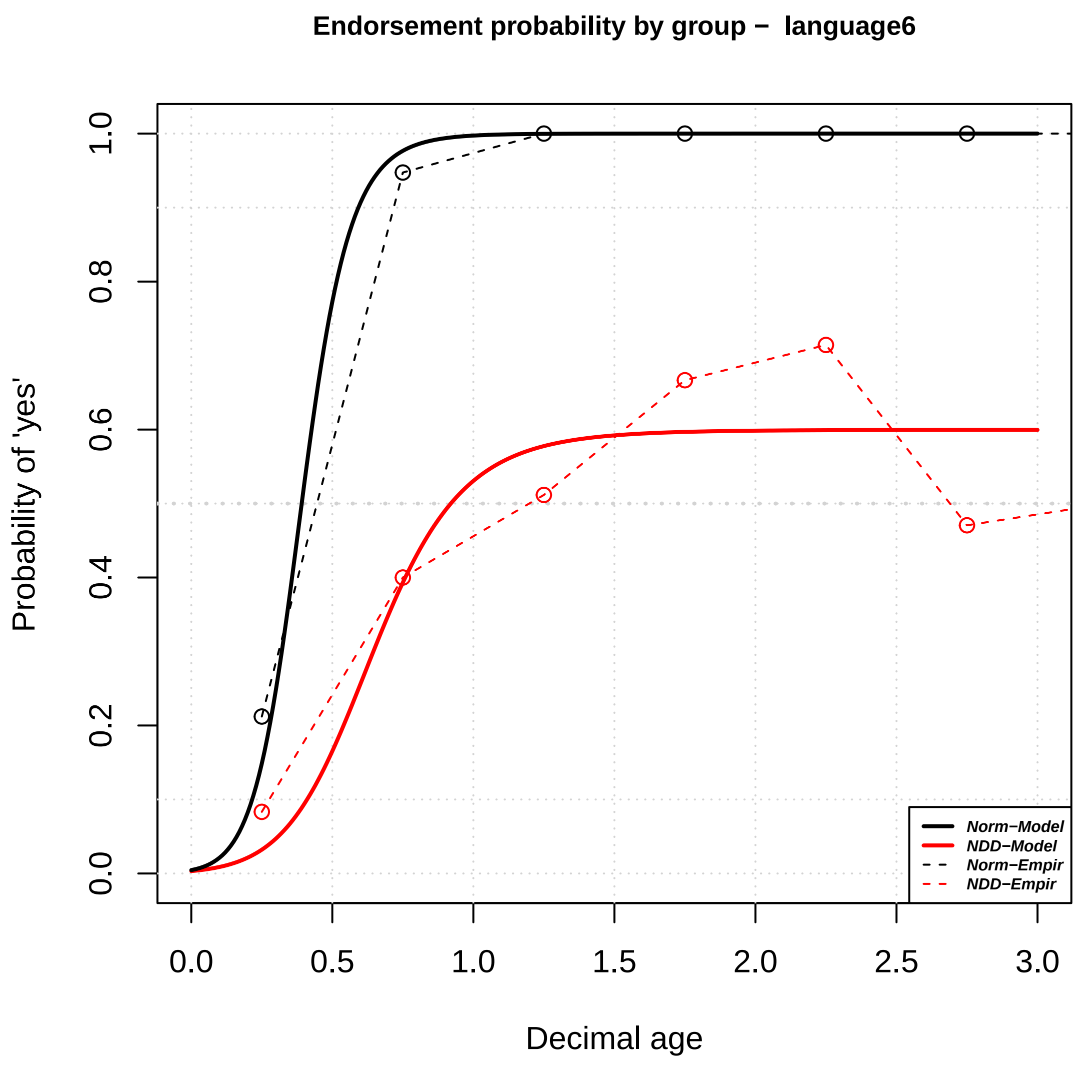

Endorsement probability by group − language6
Probability of 'yes'
0.0
0.2
0.4
0.6
0.8
1.0
0.0
0.5
1.0
1.5
2.0
2.5
3.0
Decimal age
Norm−Model
NDD−Model
Norm−Empir
NDD−Empir


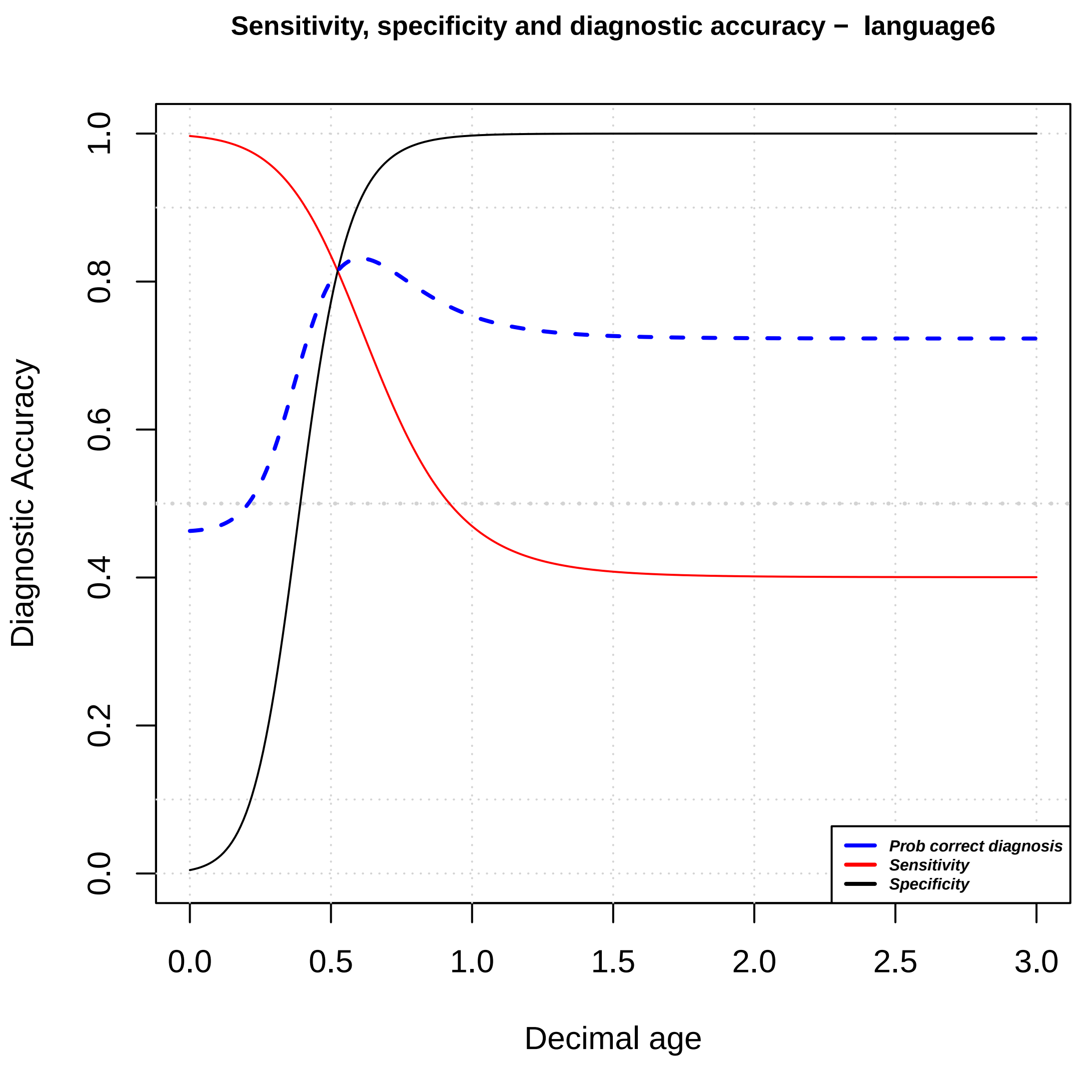

Sensitivity, specificity and diagnostic accuracy − language6
Diagnostic Accuracy
0.0
0.2
0.4
0.6
0.8
1.0
0.0
0.5
1.0
1.5
2.0
2.5
3.0
Decimal age
Prob correct diagnosis
Sensitivity
Specificity

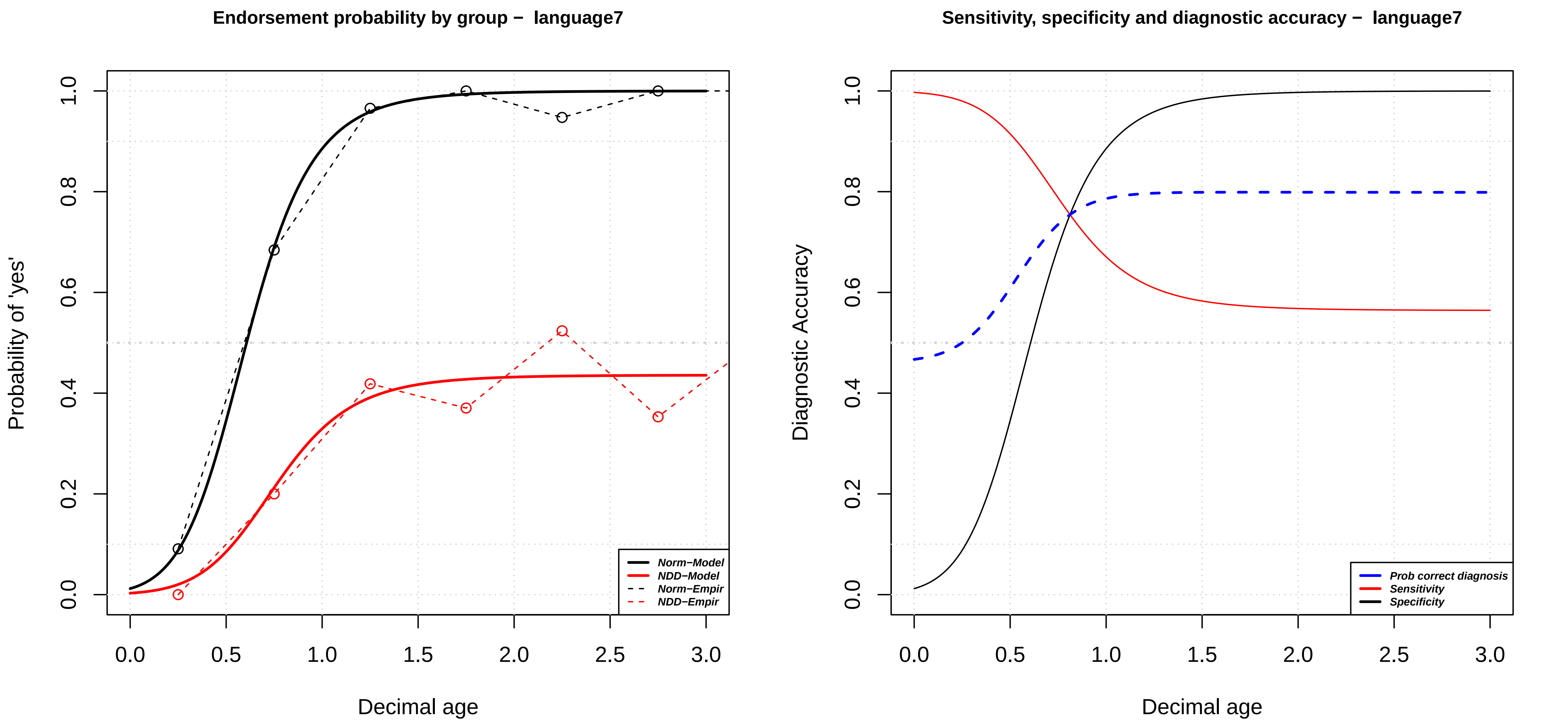
Endorsement probability by group − language7
Probability of 'yes'
Decimal age
0.0
0.2
0.4
0.6
0.8
1.0
0.0
0.5
1.0
1.5
2.0
2.5
3.0
Norm−Model
NDD−Model
Norm−Empir
NDD−Empir
Sensitivity, specificity and diagnostic accuracy − language7
Diagnostic Accuracy
Decimal age
Prob correct diagnosis
Sensitivity
Specificity

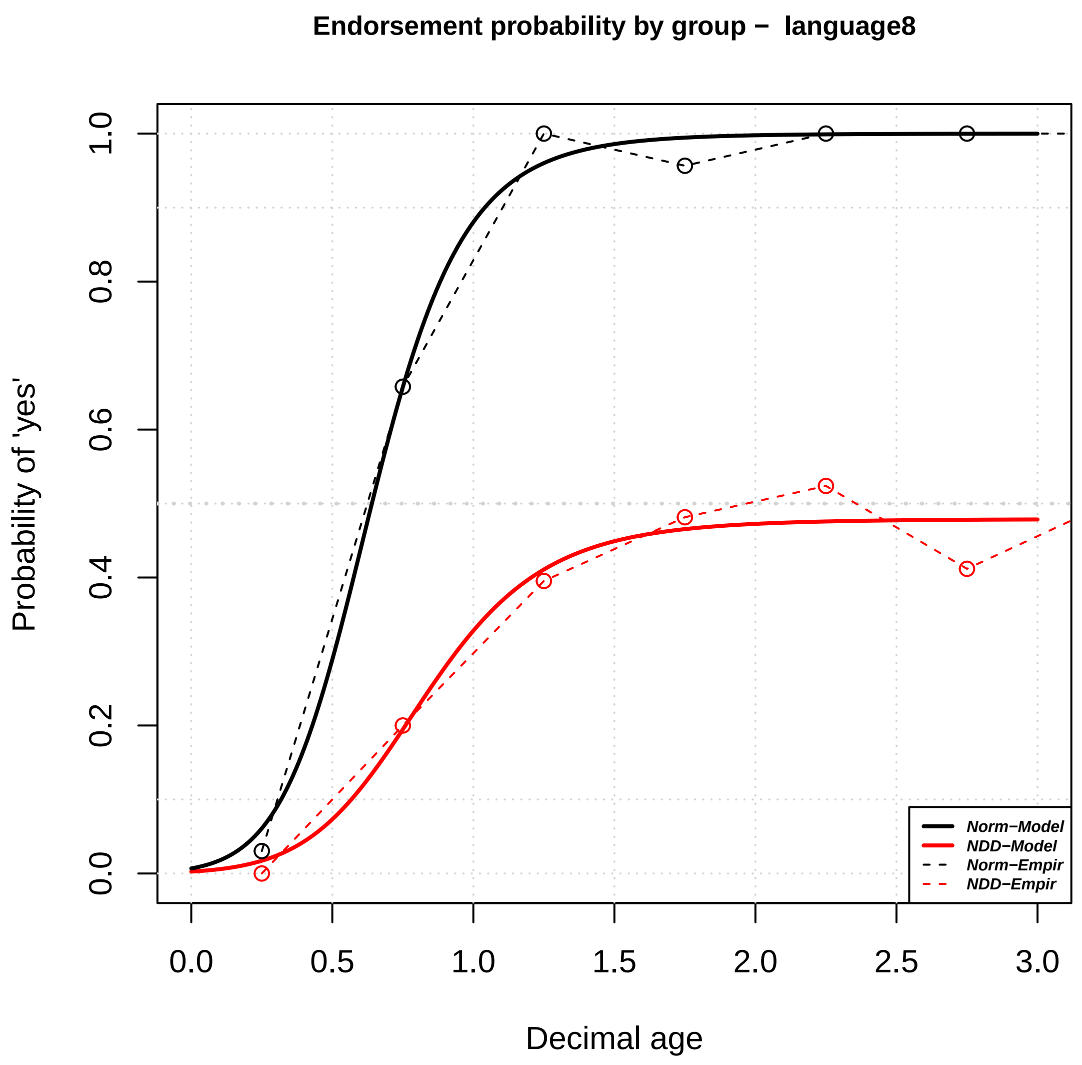
Endorsement probability by group – language8
Probability of 'yes'
Decimal age
0.0
0.2
0.4
0.6
0.8
1.0
0.0
0.5
1.0
1.5
2.0
2.5
3.0
Norm–Model
NDD–Model
Norm–Empir
NDD–Empir

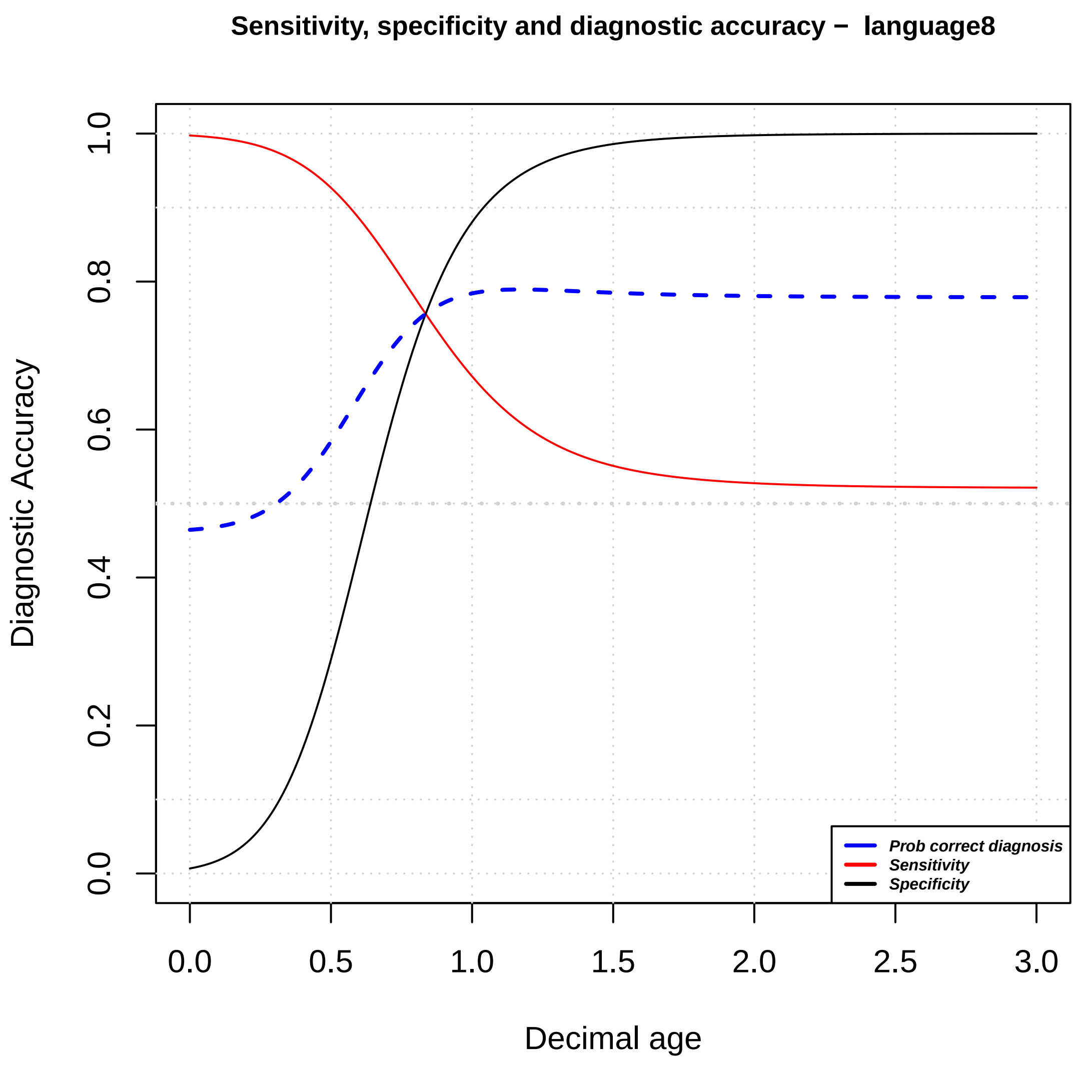
Sensitivity, specificity and diagnostic accuracy – language8
Diagnostic Accuracy
Decimal age
0.0
0.2
0.4
0.6
0.8
1.0
0.0
0.5
1.0
1.5
2.0
2.5
3.0
Prob correct diagnosis
Sensitivity
Specificity

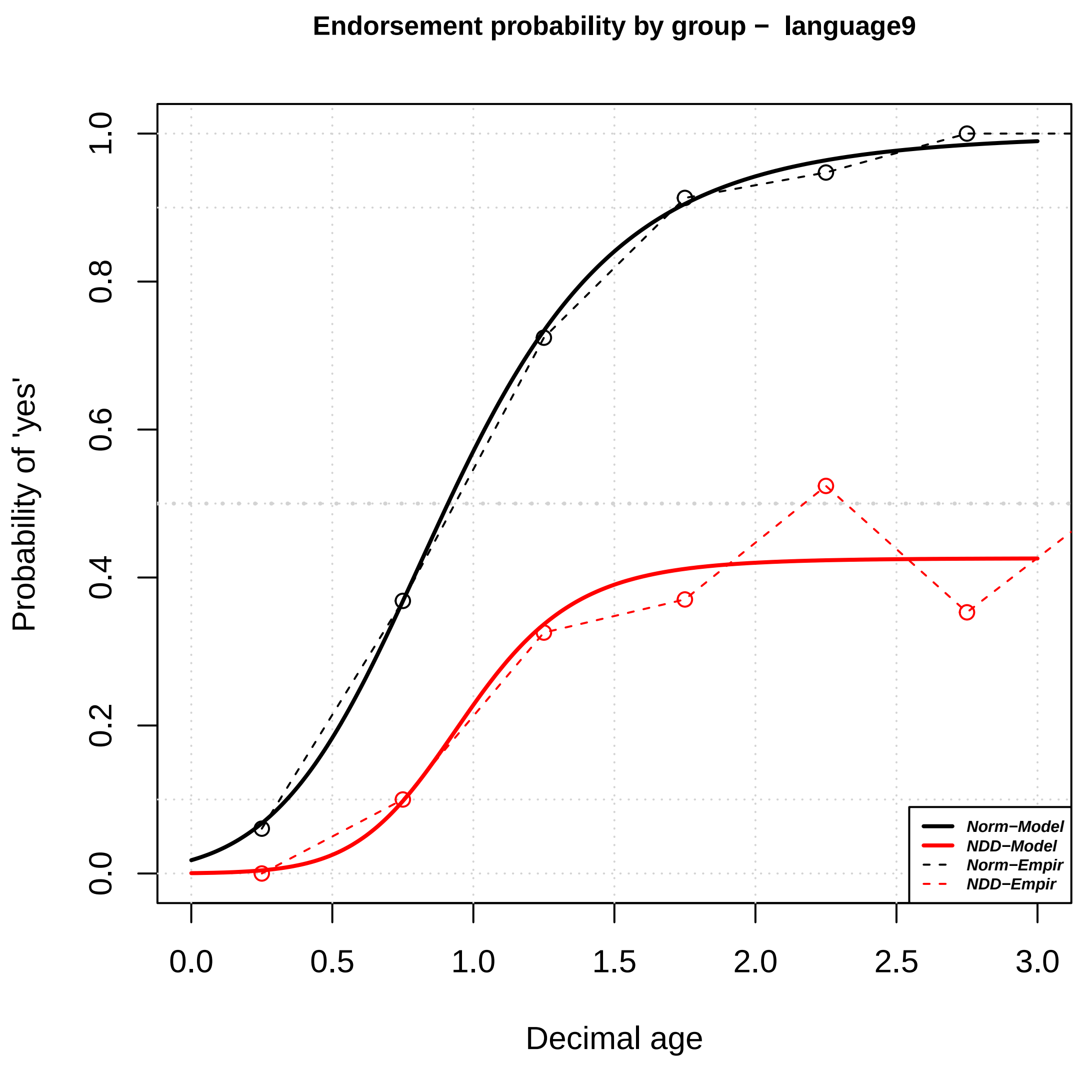
Endorsement probability by group – language9
Probability of 'yes'
Decimal age
0.0
0.2
0.4
0.6
0.8
1.0
0.0
0.5
1.0
1.5
2.0
2.5
3.0
Norm–Model
NDD–Model
Norm–Empir
NDD–Empir

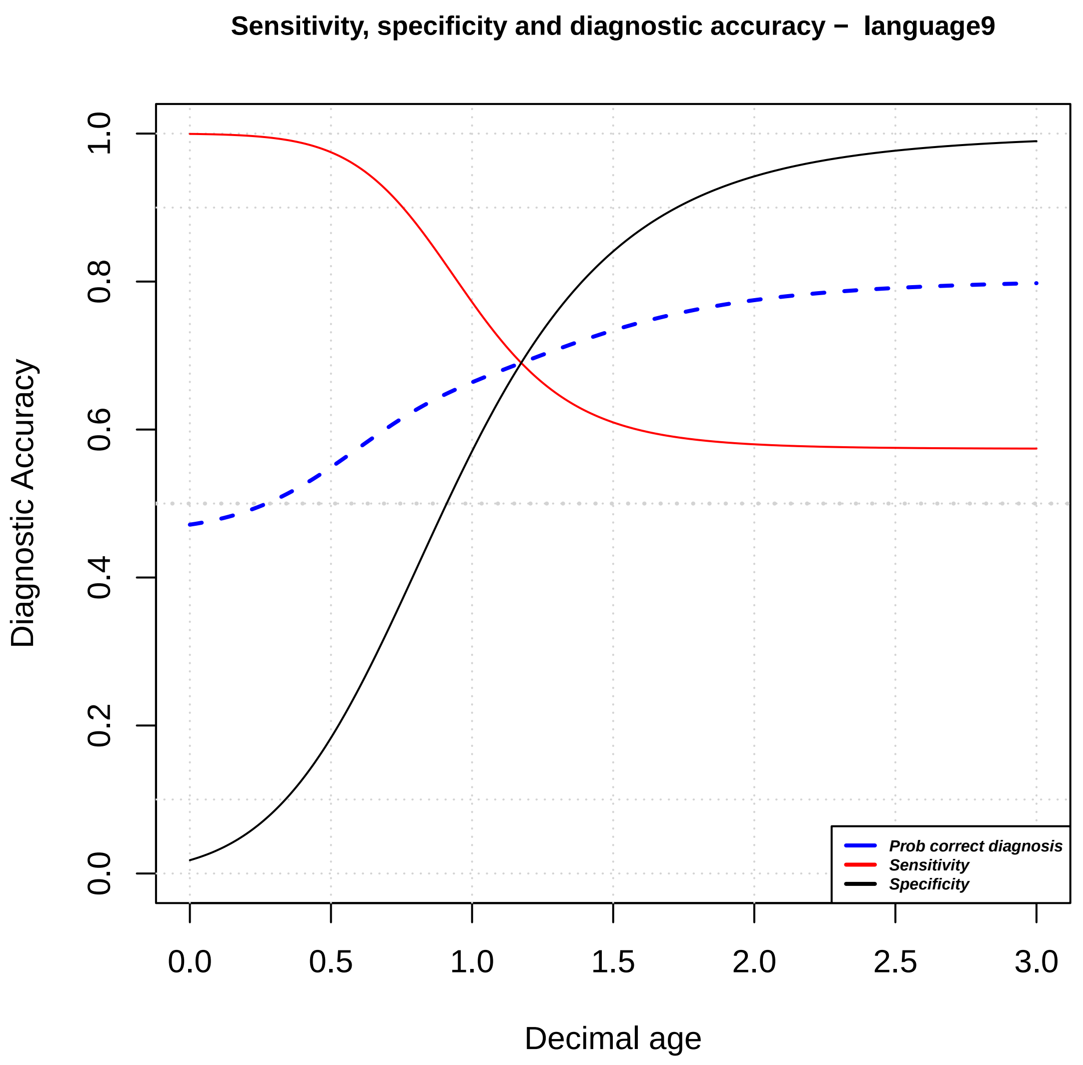
Sensitivity, specificity and diagnostic accuracy – language9
Diagnostic Accuracy
Decimal age
0.0
0.2
0.4
0.6
0.8
1.0
0.0
0.5
1.0
1.5
2.0
2.5
3.0
Prob correct diagnosis
Sensitivity
Specificity

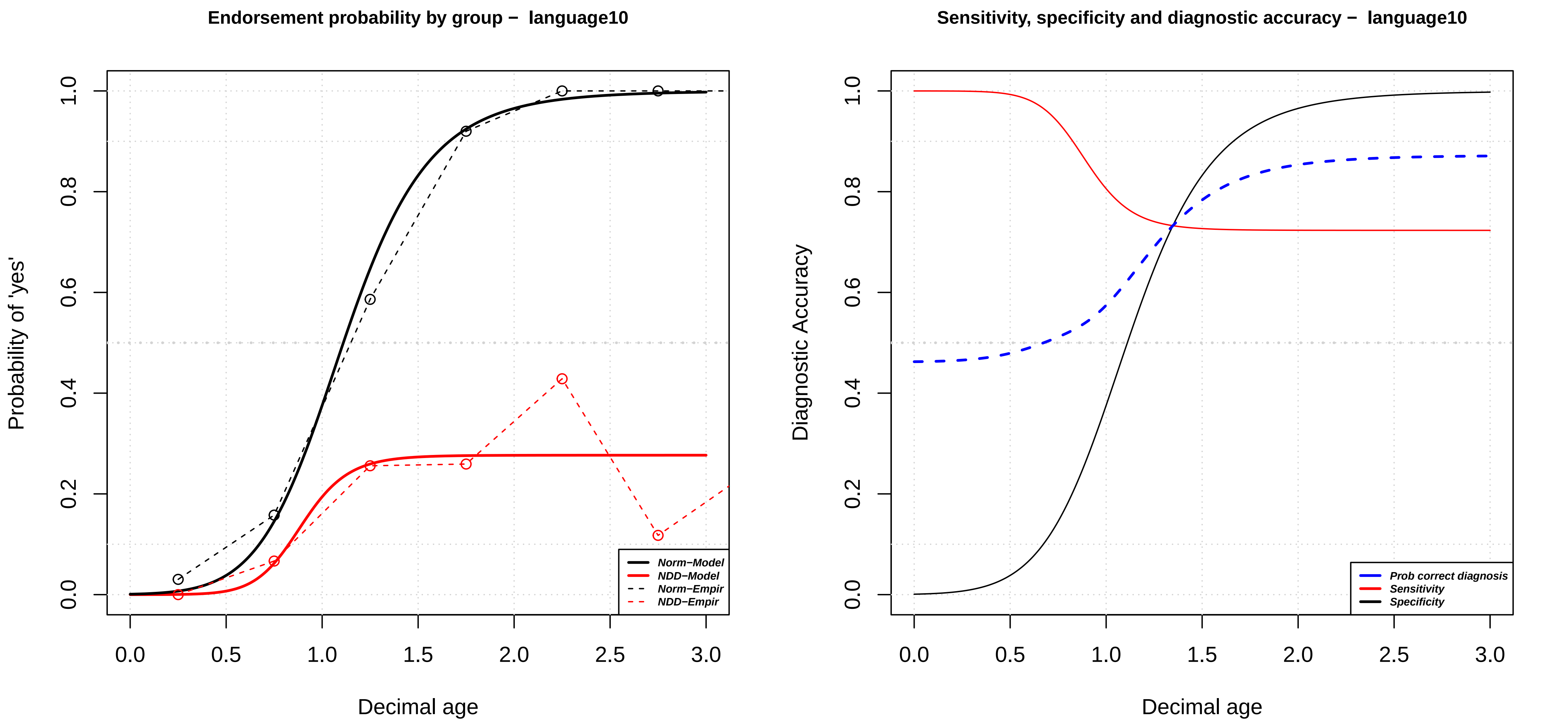

Endorsement probability by group – language10
Probability of 'yes'
0.0
0.2
0.4
0.6
0.8
1.0
0.0
0.5
1.0
1.5
2.0
2.5
3.0
Decimal age
Norm–Model
NDD–Model
Norm–Empir
NDD–Empir
Sensitivity, specificity and diagnostic accuracy – language10
Diagnostic Accuracy
0.0
0.2
0.4
0.6
0.8
1.0
0.0
0.5
1.0
1.5
2.0
2.5
3.0
Decimal age
Prob correct diagnosis
Sensitivity
Specificity

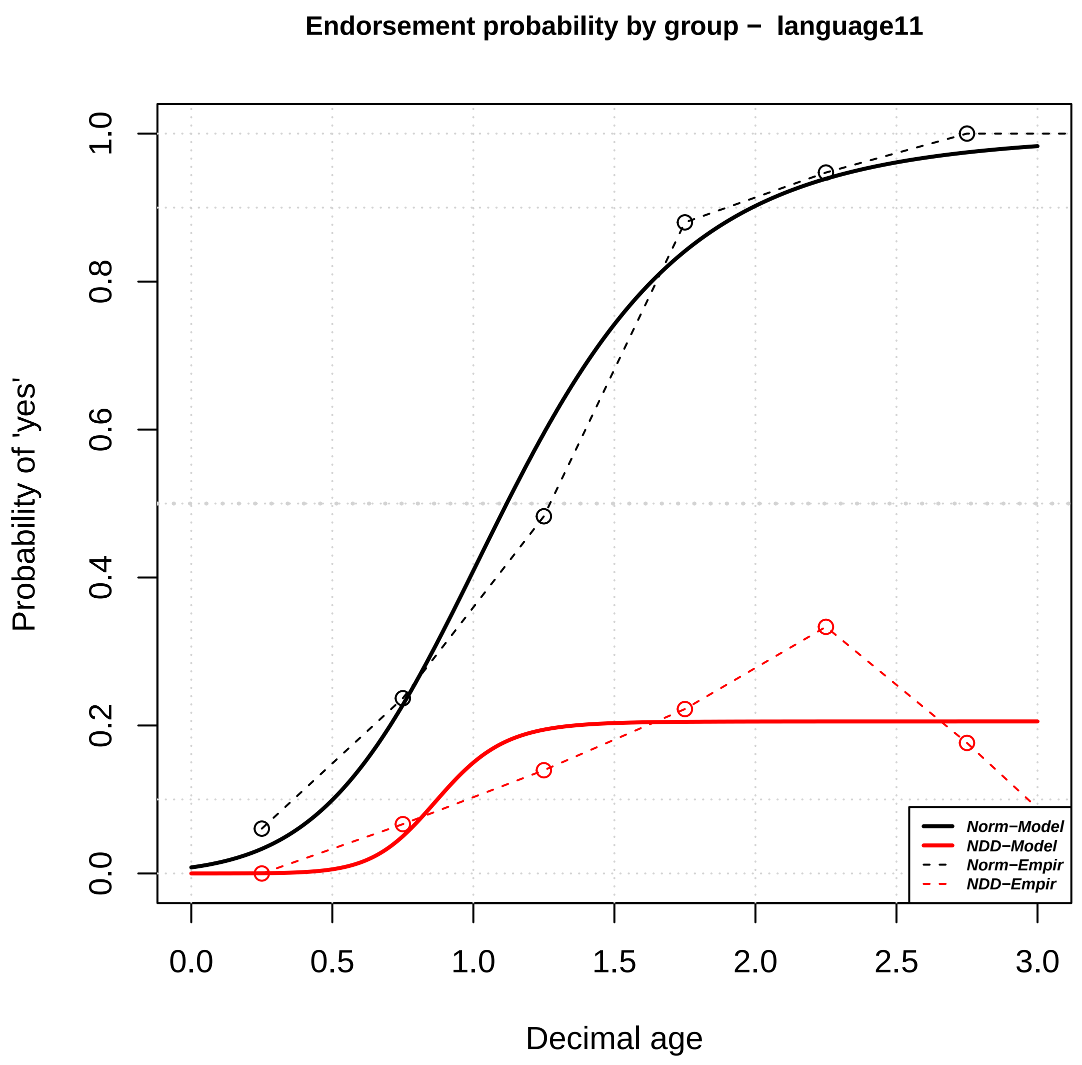
Endorsement probability by group – language11
Probability of 'yes'
Decimal age
0.0
0.2
0.4
0.6
0.8
1.0
0.0
0.5
1.0
1.5
2.0
2.5
3.0
Norm–Model
NDD–Model
Norm–Empir
NDD–Empir

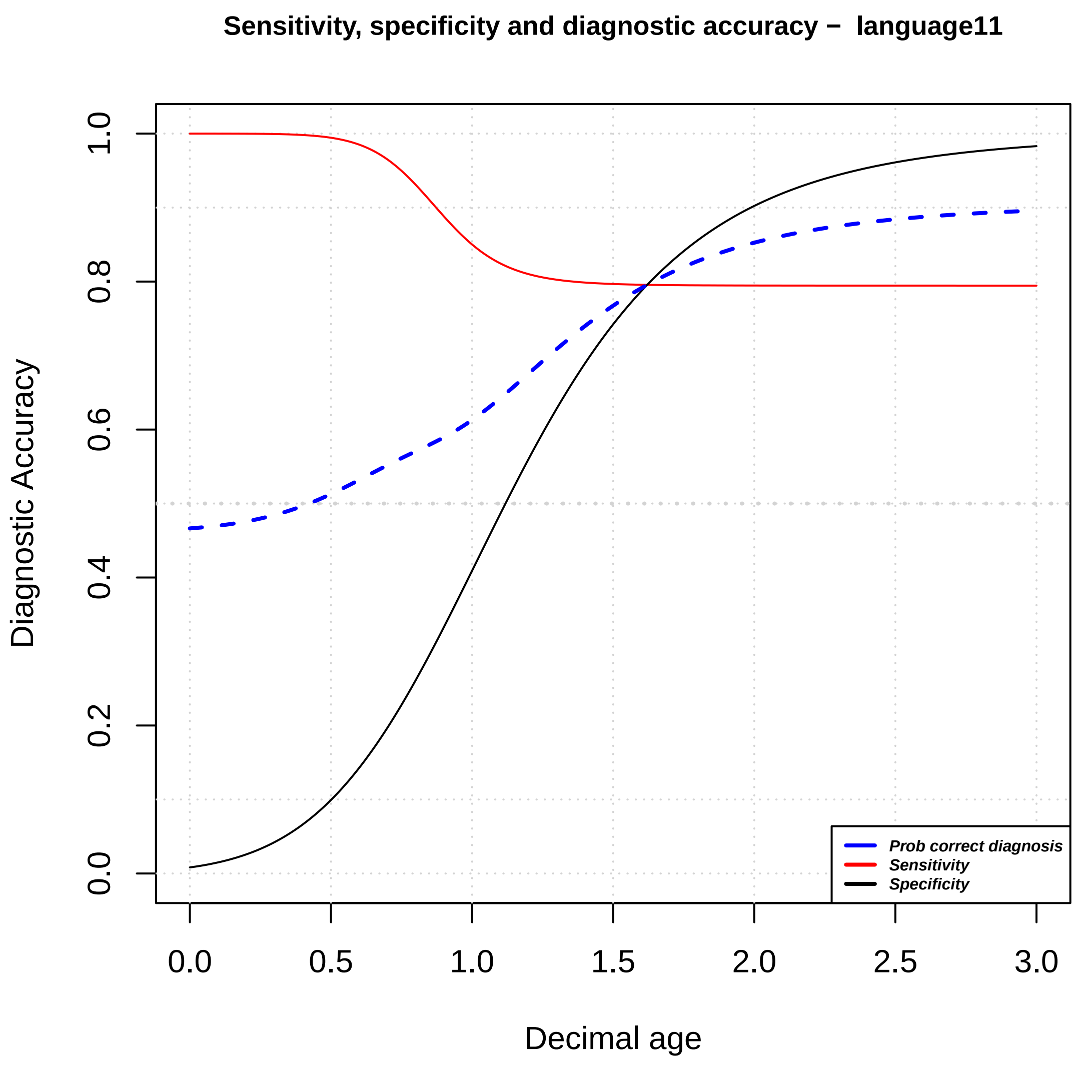
Sensitivity, specificity and diagnostic accuracy – language11
Diagnostic Accuracy
Decimal age
0.0
0.2
0.4
0.6
0.8
1.0
0.0
0.5
1.0
1.5
2.0
2.5
3.0
Prob correct diagnosis
Sensitivity
Specificity

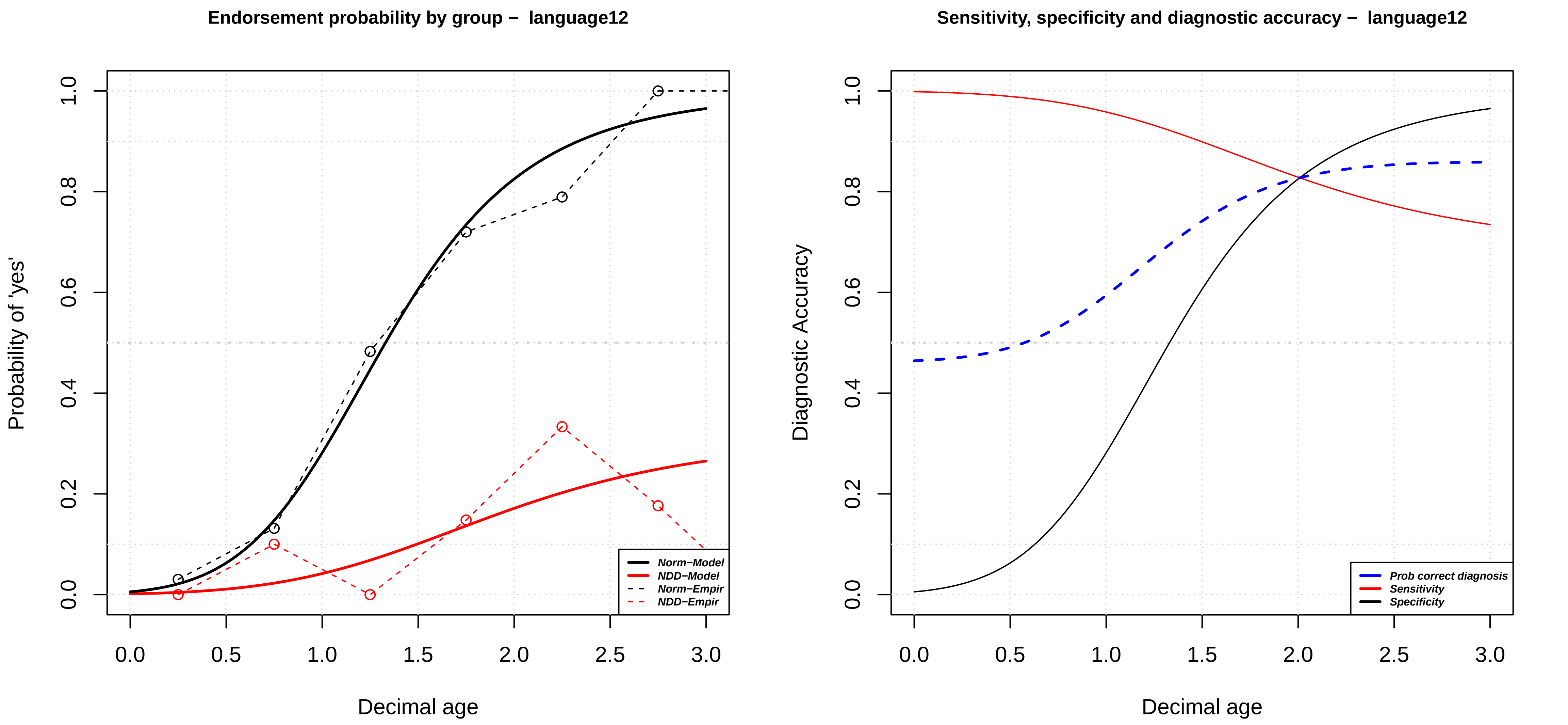

Endorsement probability by group − language12
Probability of 'yes'
Decimal age
Norm−Model
NDD−Model
Norm−Empir
NDD−Empir
Sensitivity, specificity and diagnostic accuracy − language12
Diagnostic Accuracy
Decimal age
Prob correct diagnosis
Sensitivity
Specificity

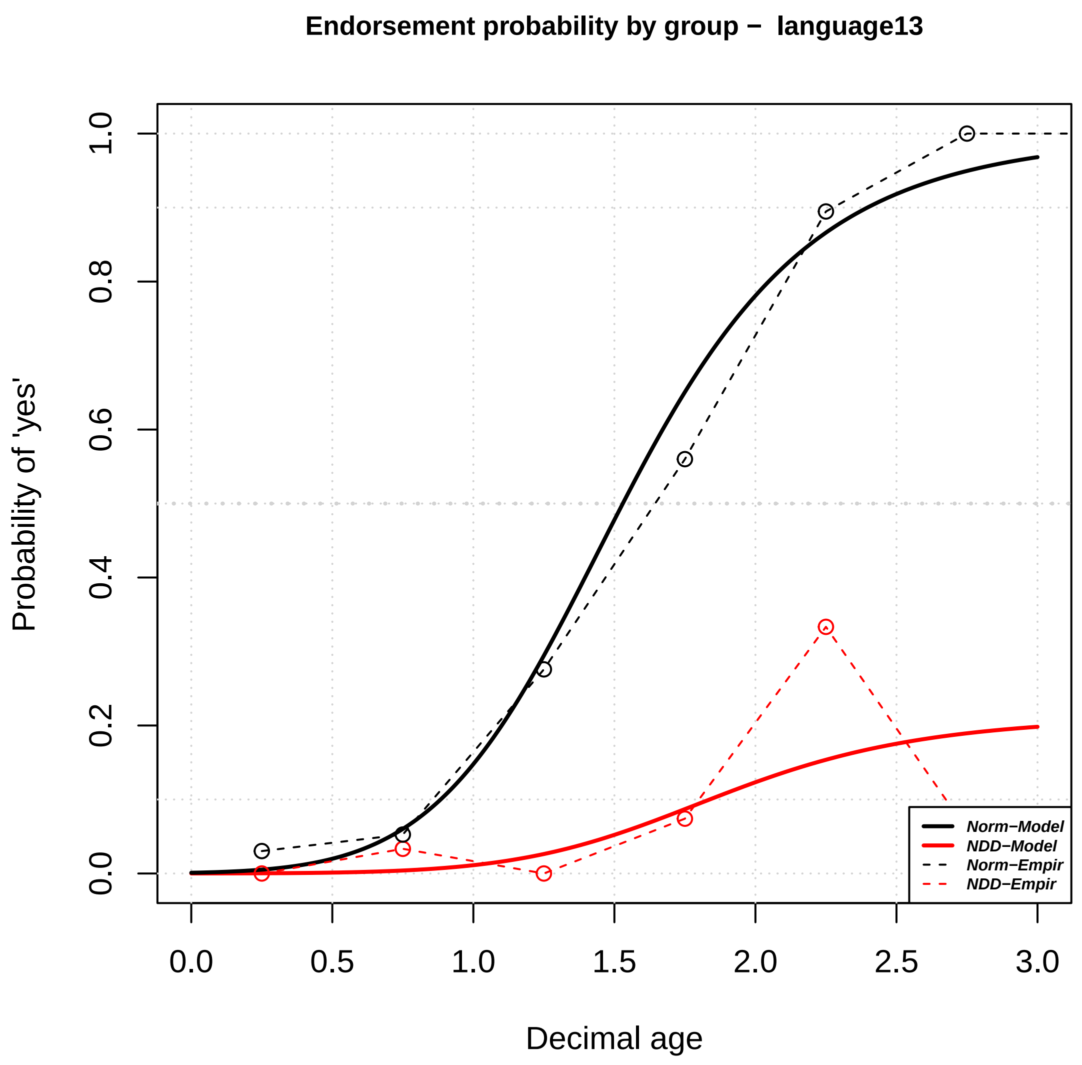

Endorsement probability by group – language13
Probability of 'yes'
Decimal age
Norm–Model
NDD–Model
Norm–Empir
NDD–Empir
0.0
0.2
0.4
0.6
0.8
1.0
0.0
0.5
1.0
1.5
2.0
2.5
3.0


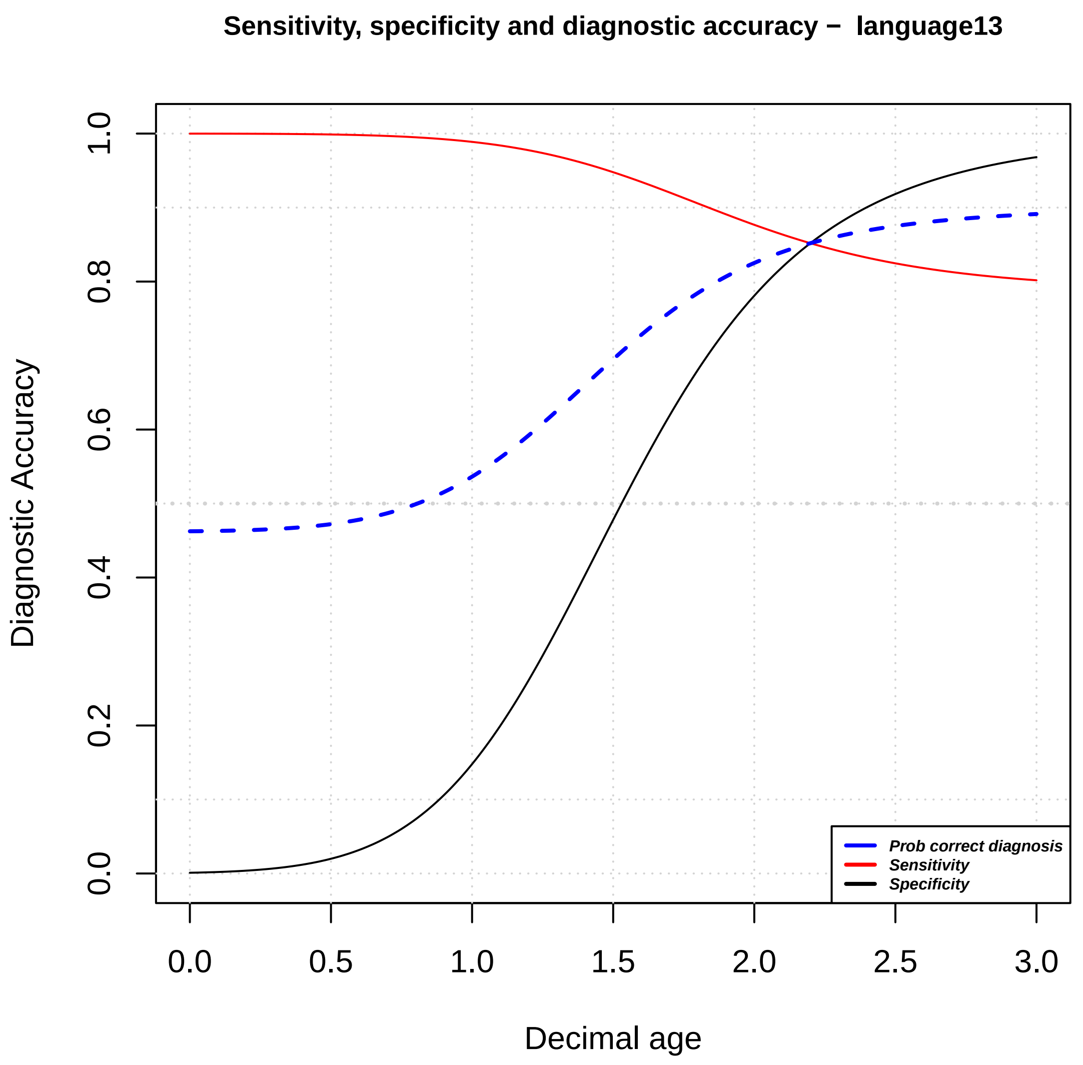

Sensitivity, specificity and diagnostic accuracy – language13
Diagnostic Accuracy
Decimal age
Prob correct diagnosis
Sensitivity
Specificity
0.0
0.2
0.4
0.6
0.8
1.0
0.0
0.5
1.0
1.5
2.0
2.5
3.0

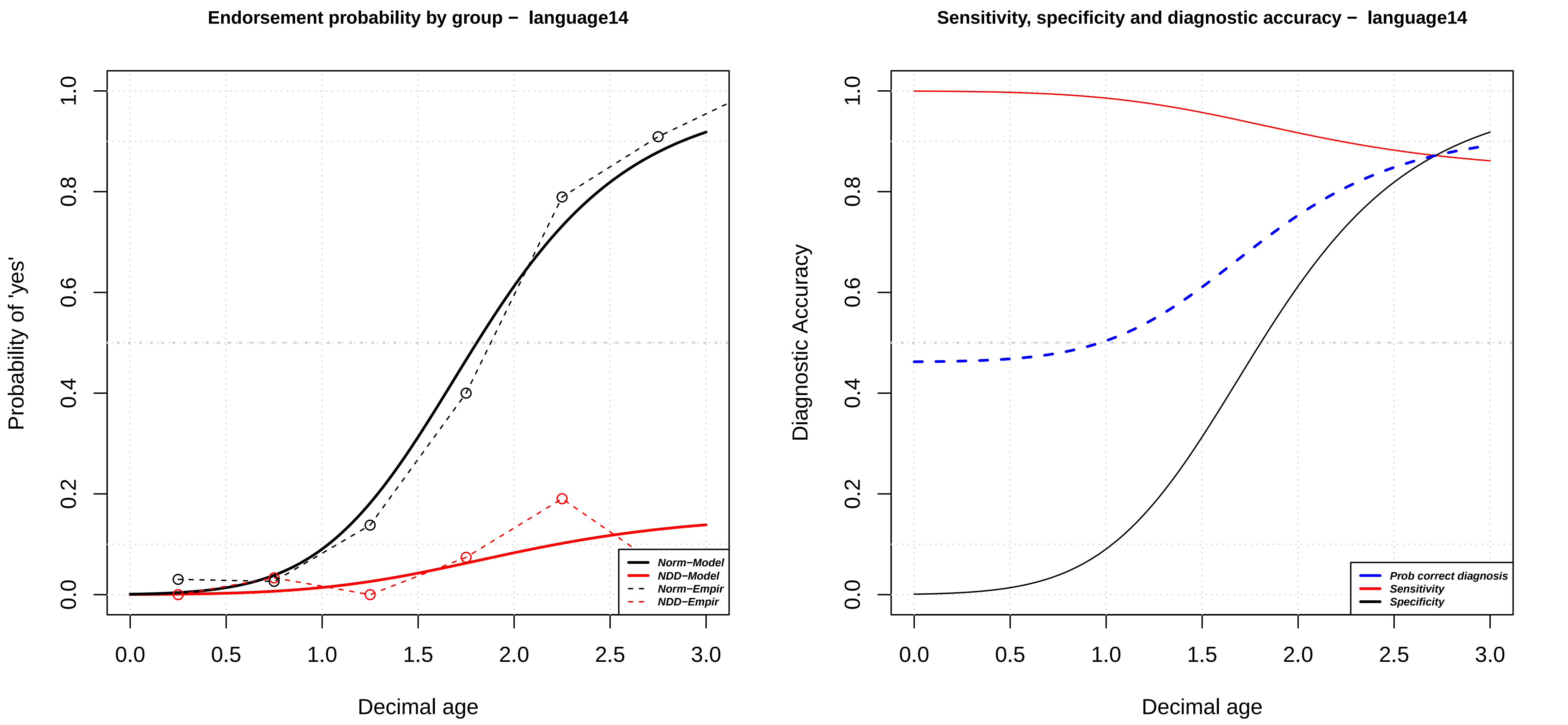
Endorsement probability by group − language14
Probability of 'yes'
Decimal age
Norm−Model
NDD−Model
Norm−Empir
NDD−Empir
Sensitivity, specificity and diagnostic accuracy − language14
Diagnostic Accuracy
Decimal age
Prob correct diagnosis
Sensitivity
Specificity

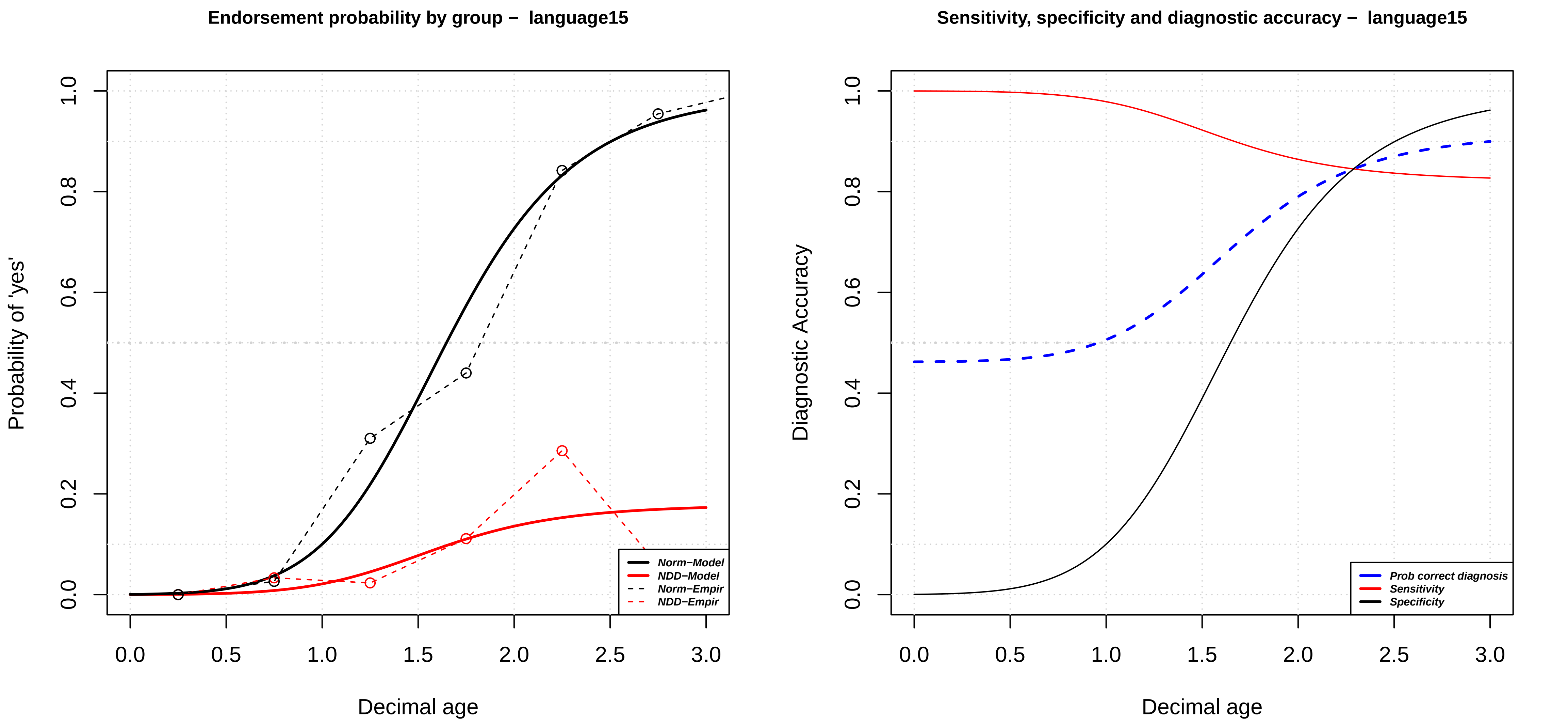

Endorsement probability by group − language15
Probability of 'yes'
Decimal age
0.0
0.2
0.4
0.6
0.8
1.0
0.0
0.5
1.0
1.5
2.0
2.5
3.0
Norm−Model
NDD−Model
Norm−Empir
NDD−Empir
Sensitivity, specificity and diagnostic accuracy − language15
Diagnostic Accuracy
Decimal age
Prob correct diagnosis
Sensitivity
Specificity

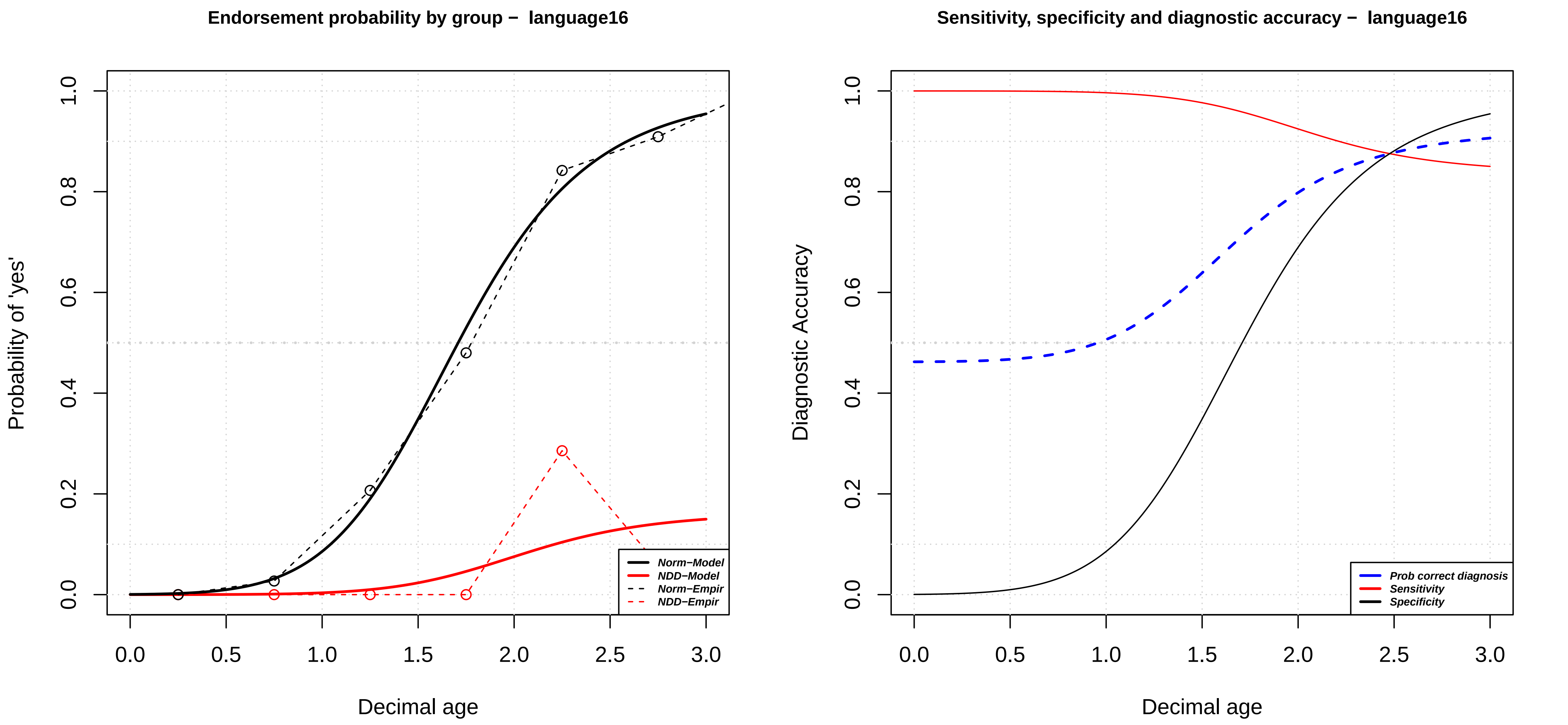

Endorsement probability by group – language16
Probability of 'yes'
Decimal age
0.0
0.2
0.4
0.6
0.8
1.0
0.0
0.5
1.0
1.5
2.0
2.5
3.0
Norm−Model
NDD−Model
Norm−Empir
NDD−Empir
Sensitivity, specificity and diagnostic accuracy – language16
Diagnostic Accuracy
Decimal age
Prob correct diagnosis
Sensitivity
Specificity

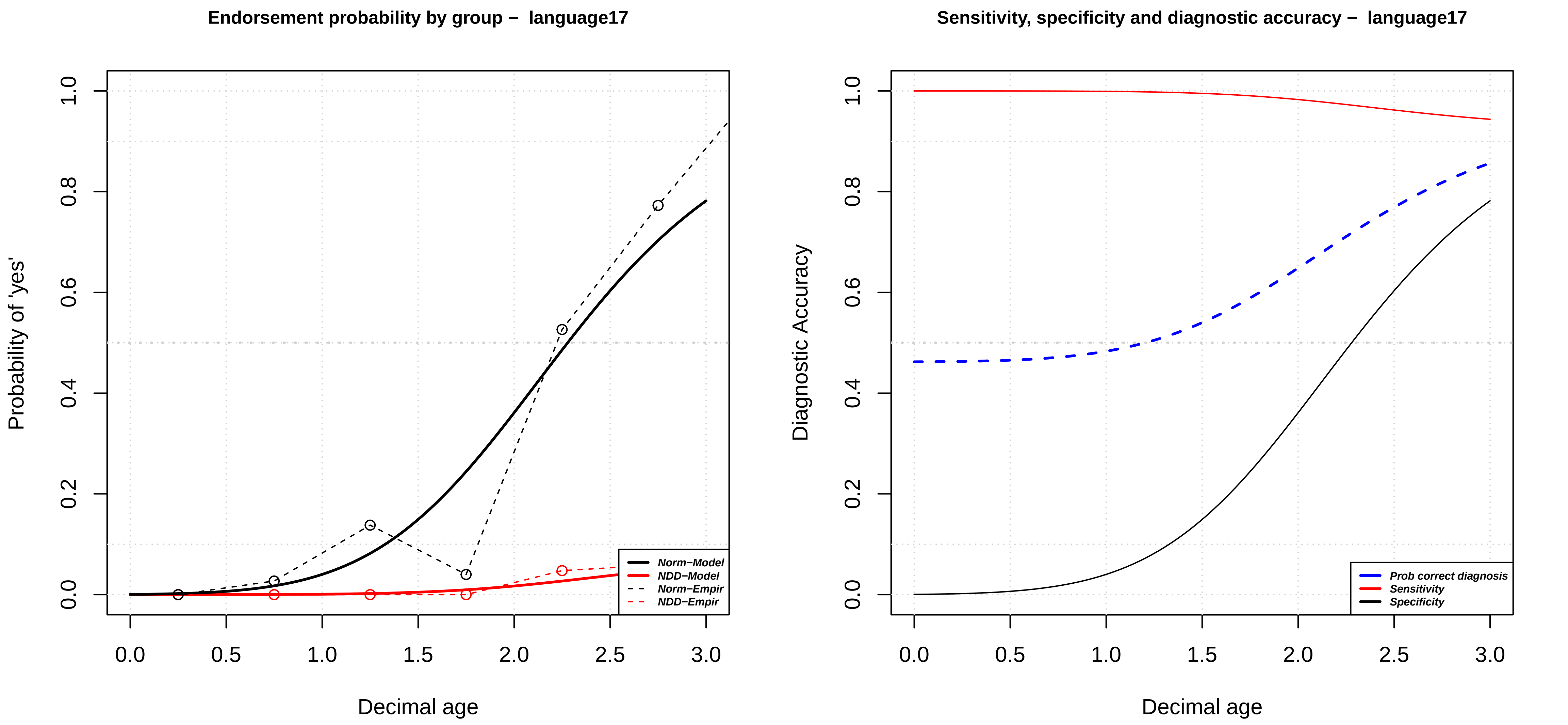
Endorsement probability by group − language17
Probability of 'yes'
Decimal age
0.0
0.2
0.4
0.6
0.8
1.0
0.0
0.5
1.0
1.5
2.0
2.5
3.0
Norm−Model
NDD−Model
Norm−Empir
NDD−Empir
Sensitivity, specificity and diagnostic accuracy − language17
Diagnostic Accuracy
Decimal age
Prob correct diagnosis
Sensitivity
Specificity

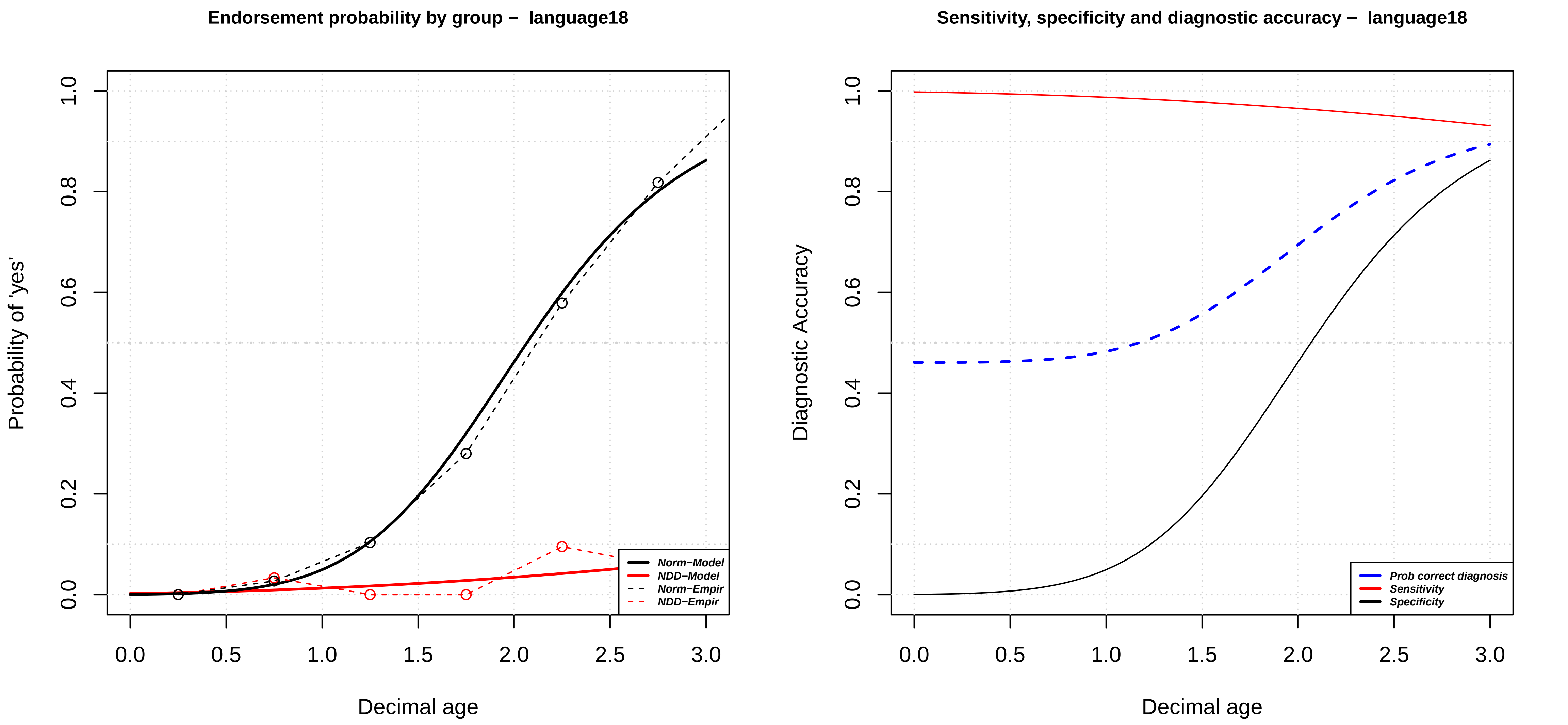

Endorsement probability by group – language18
Probability of 'yes'
Decimal age
Norm–Model
NDD–Model
Norm–Empir
NDD–Empir
Sensitivity, specificity and diagnostic accuracy – language18
Diagnostic Accuracy
Decimal age
Prob correct diagnosis
Sensitivity
Specificity

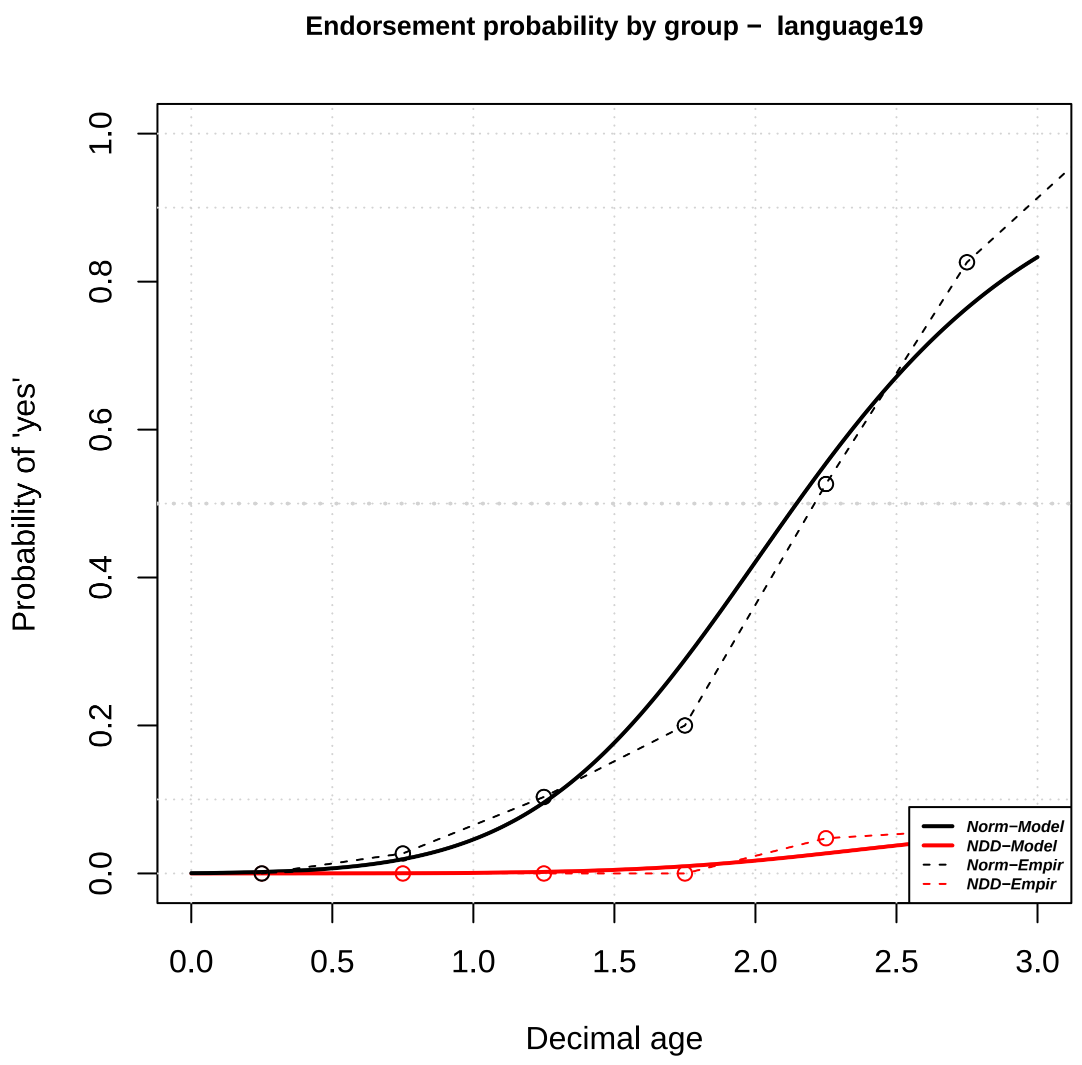
Endorsement probability by group − language19
Probability of 'yes'
Decimal age
0.0
0.2
0.4
0.6
0.8
1.0
0.0 0.5 1.0 1.5 2.0 2.5 3.0
Norm−Model
NDD−Model
Norm−Empir
NDD−Empir

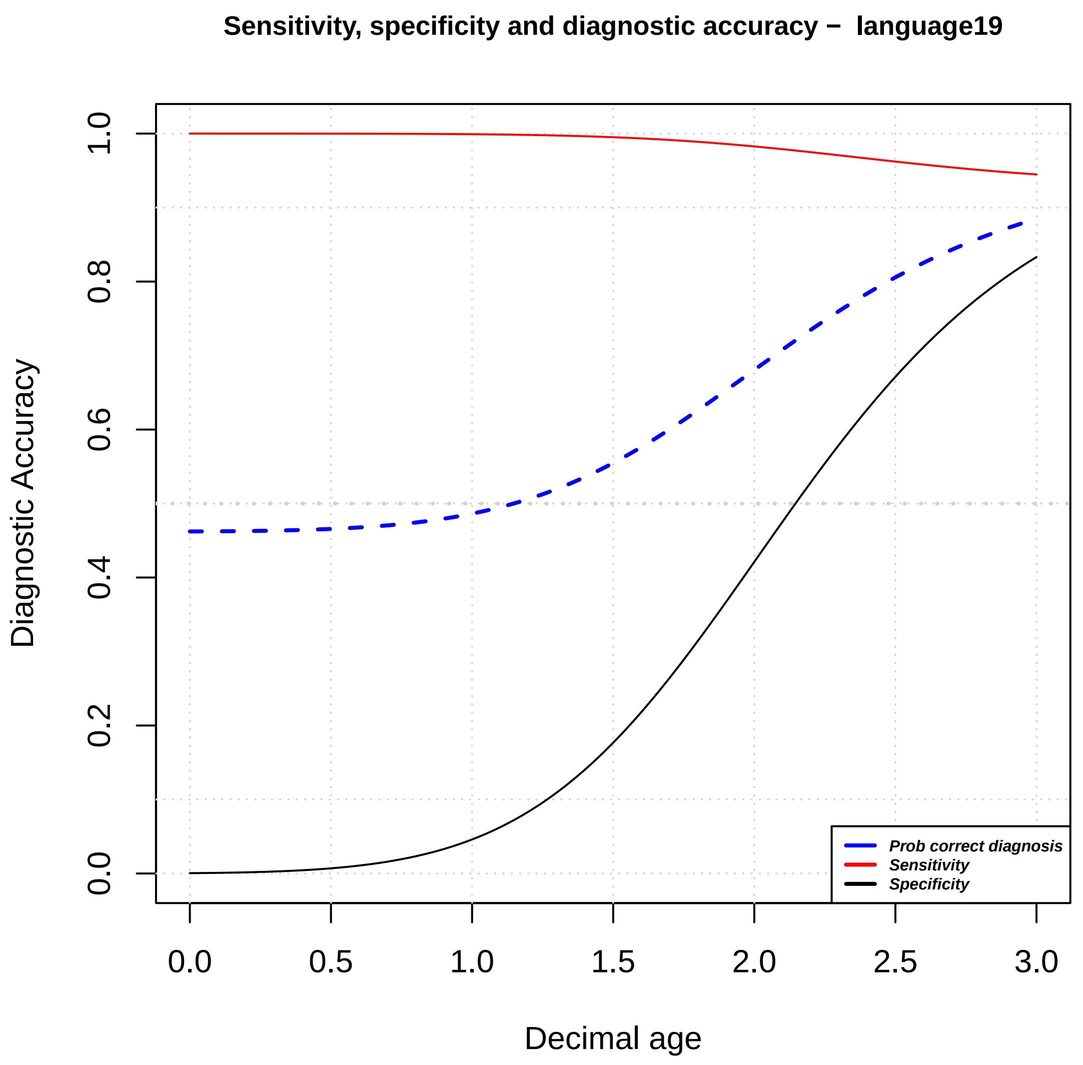
Sensitivity, specificity and diagnostic accuracy − language19
Diagnostic Accuracy
Decimal age
0.0
0.2
0.4
0.6
0.8
1.0
0.0 0.5 1.0 1.5 2.0 2.5 3.0
Prob correct diagnosis
Sensitivity
Specificity

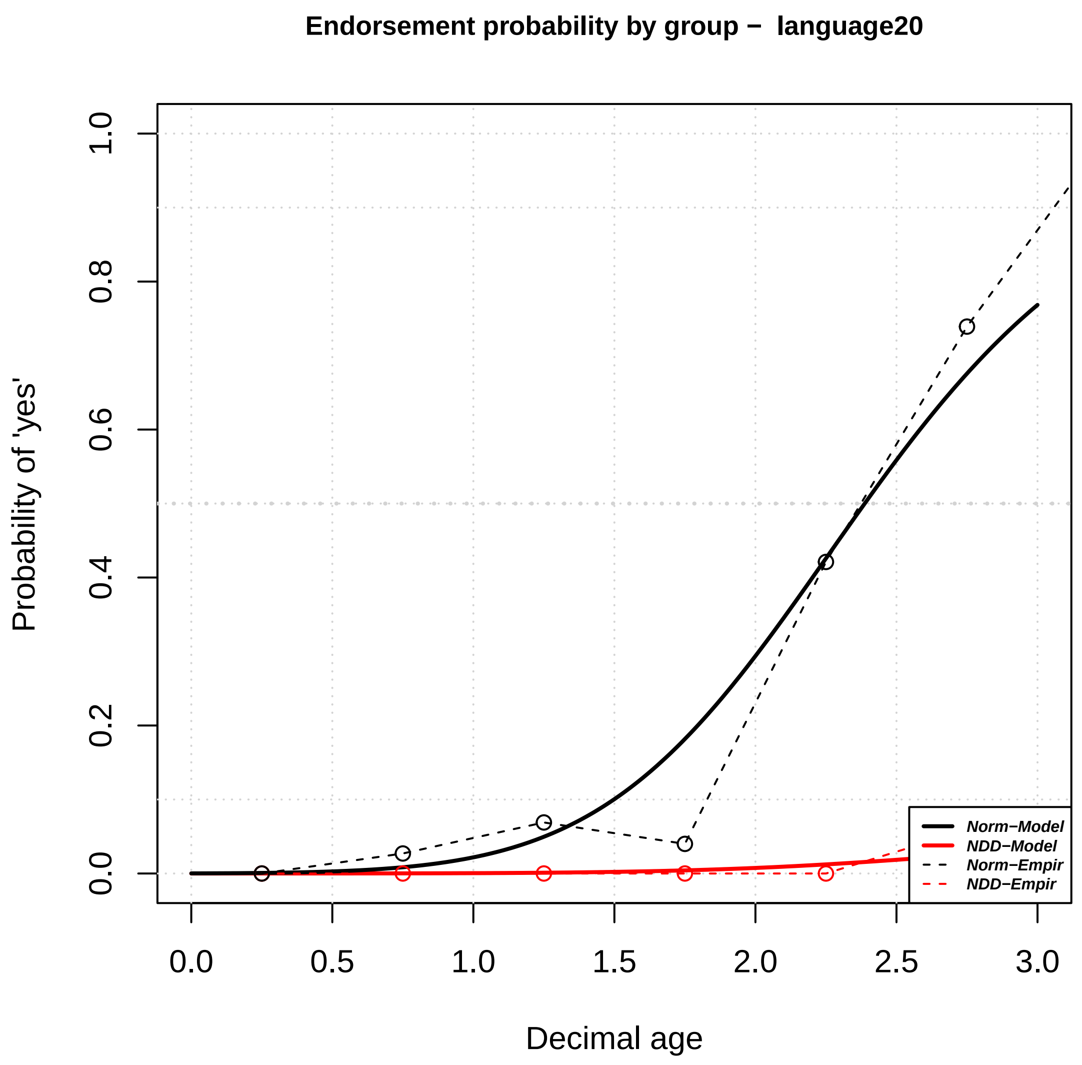

Endorsement probability by group – language20
Probability of 'yes'
Decimal age
Norm−Model
NDD−Model
Norm−Empir
NDD−Empir
0.0
0.2
0.4
0.6
0.8
1.0
0.0
0.5
1.0
1.5
2.0
2.5
3.0


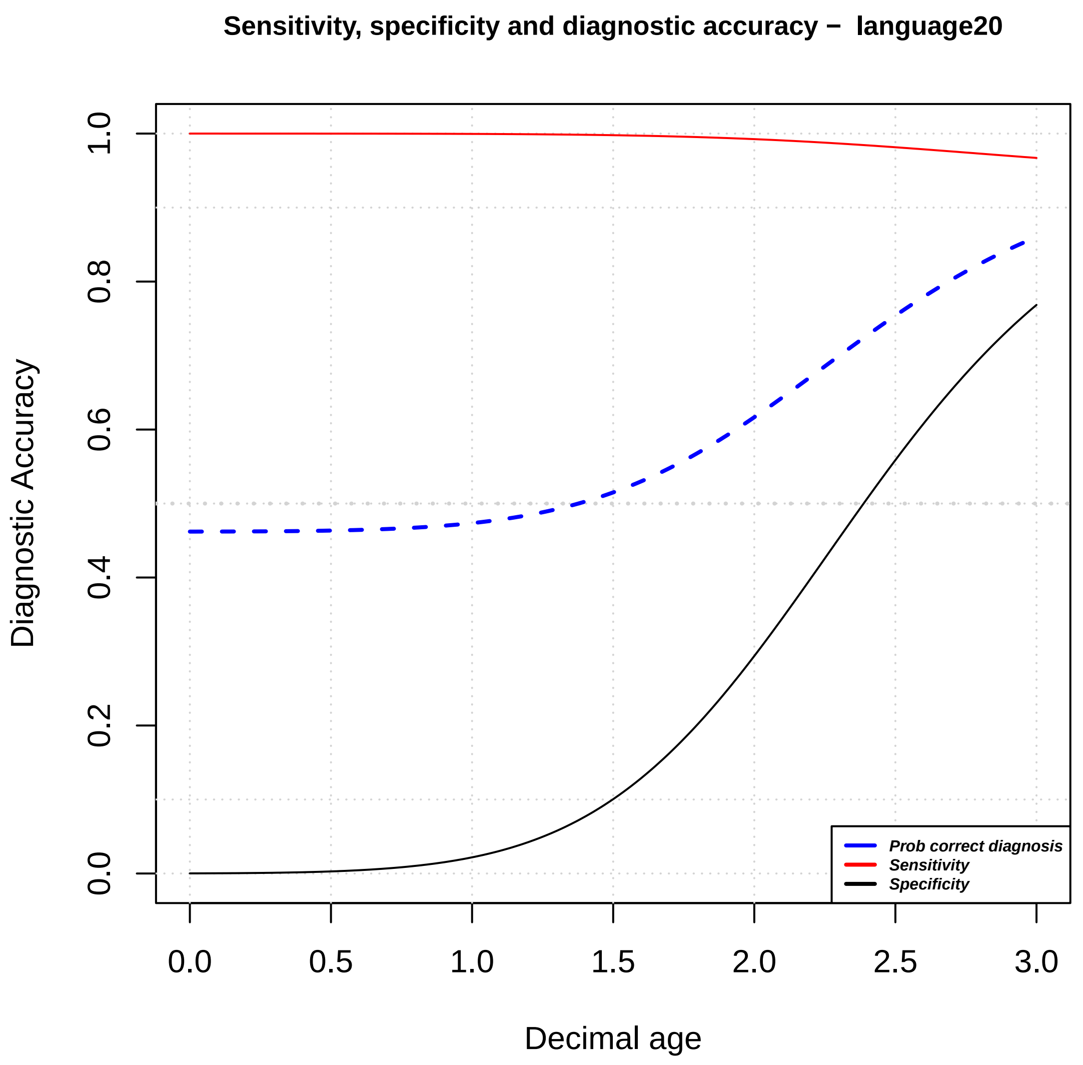

Sensitivity, specificity and diagnostic accuracy – language20
Diagnostic Accuracy
Decimal age
Prob correct diagnosis
Sensitivity
Specificity
0.0
0.2
0.4
0.6
0.8
1.0
0.0
0.5
1.0
1.5
2.0
2.5
3.0

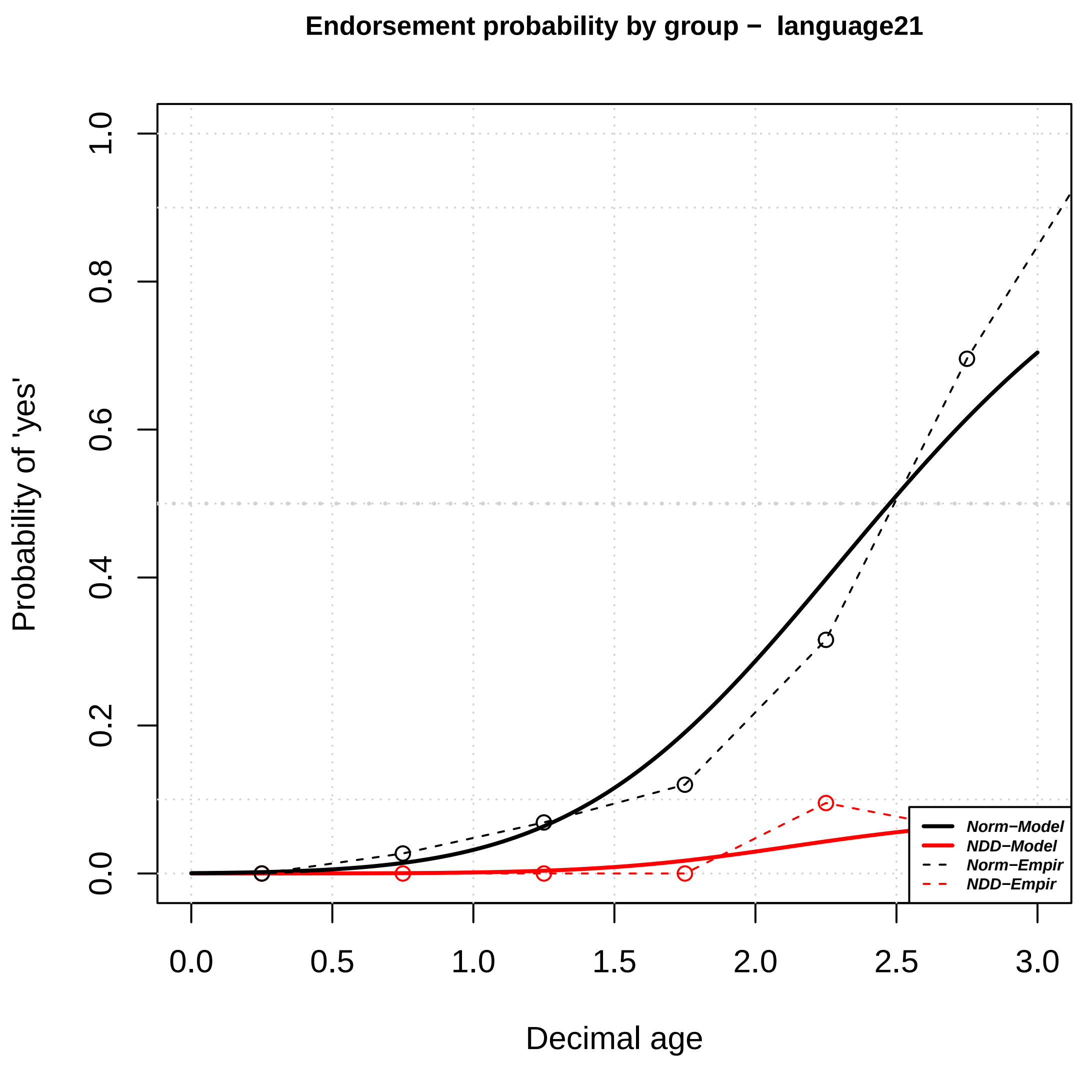

Endorsement probability by group – language21
Probability of 'yes'
Decimal age
Norm−Model
NDD−Model
Norm−Empir
NDD−Empir


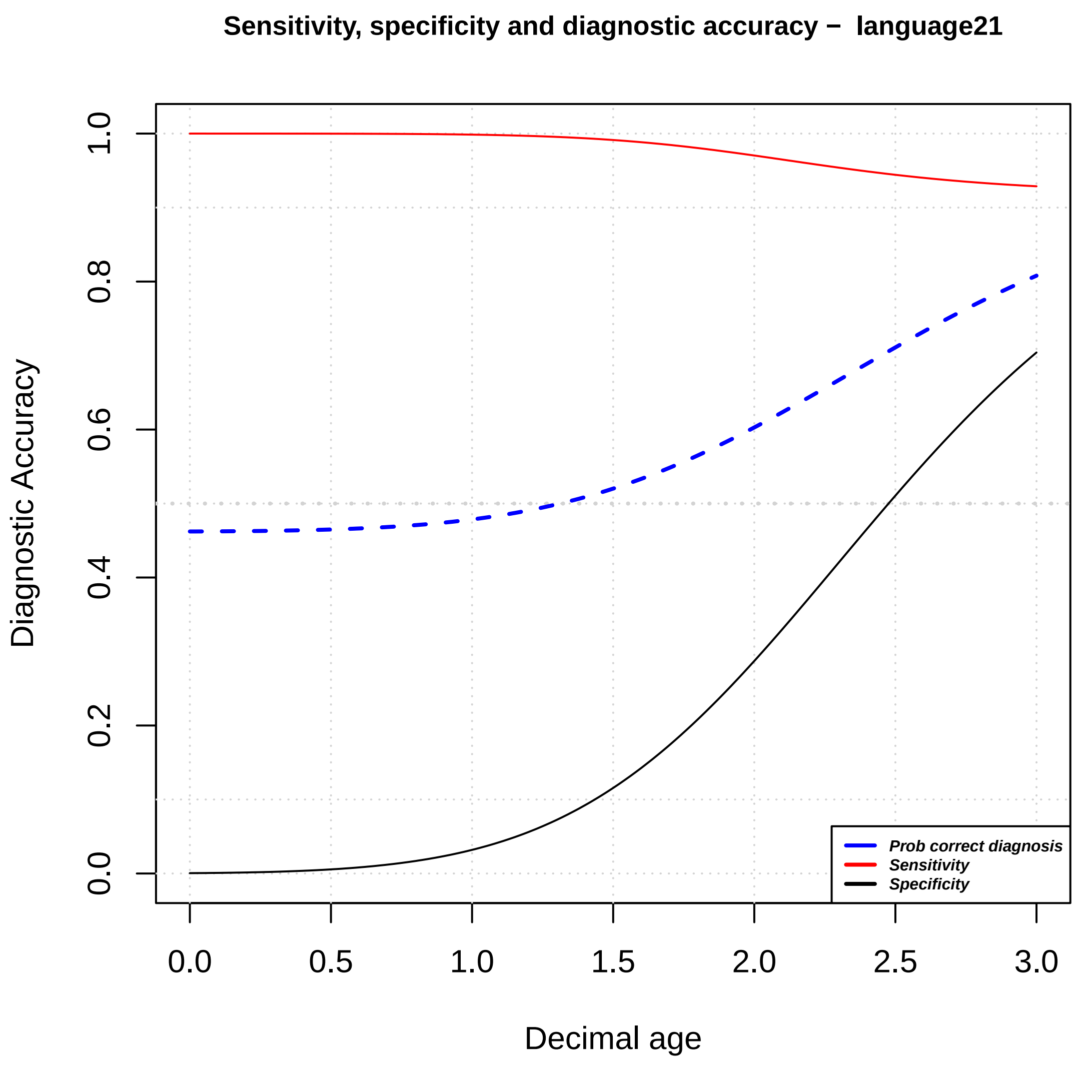

Sensitivity, specificity and diagnostic accuracy – language21
Diagnostic Accuracy
Decimal age
Prob correct diagnosis
Sensitivity
Specificity

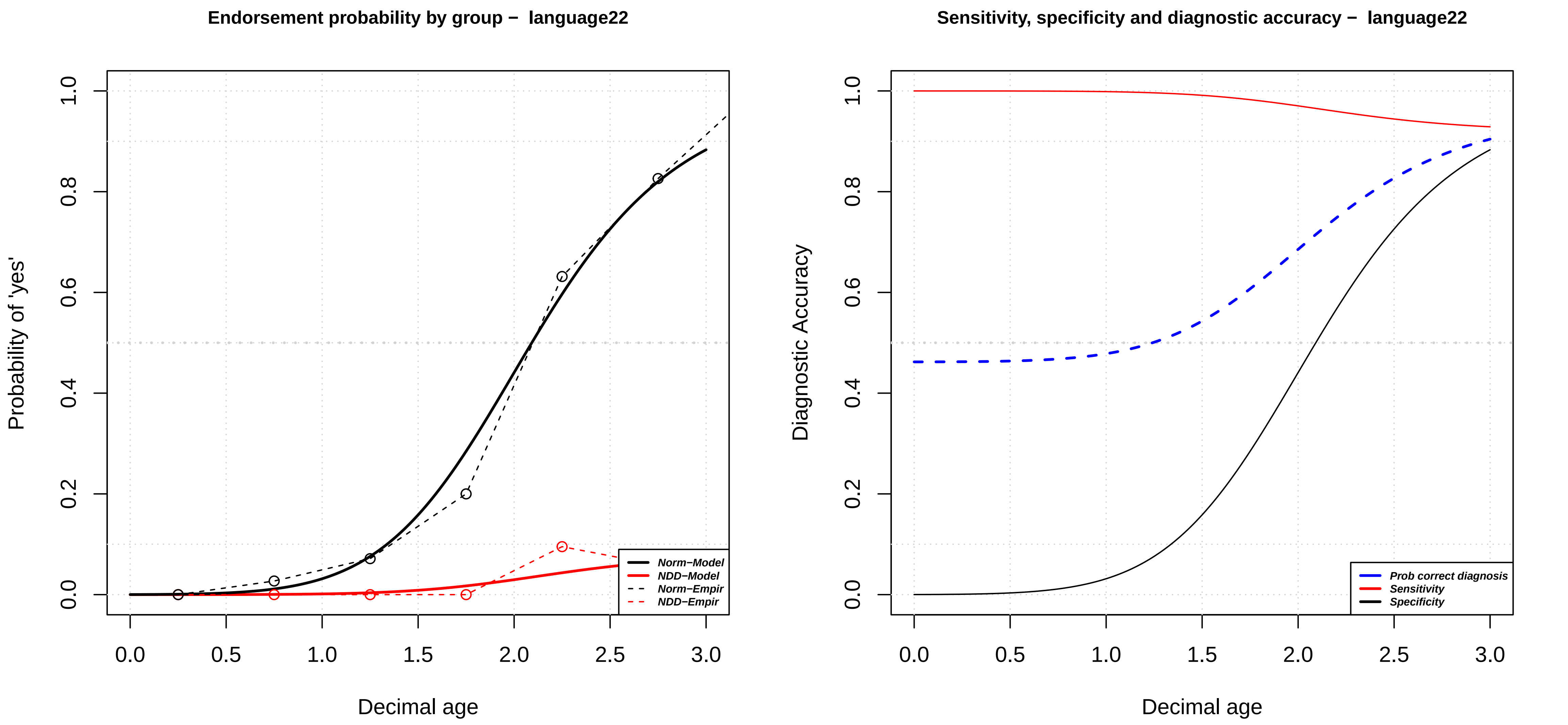
Endorsement probability by group – language22
Probability of 'yes'
Decimal age
0.0
0.2
0.4
0.6
0.8
1.0
0.0
0.5
1.0
1.5
2.0
2.5
3.0
Norm–Model
NDD–Model
Norm–Empir
NDD–Empir
Sensitivity, specificity and diagnostic accuracy – language22
Diagnostic Accuracy
Decimal age
Prob correct diagnosis
Sensitivity
Specificity

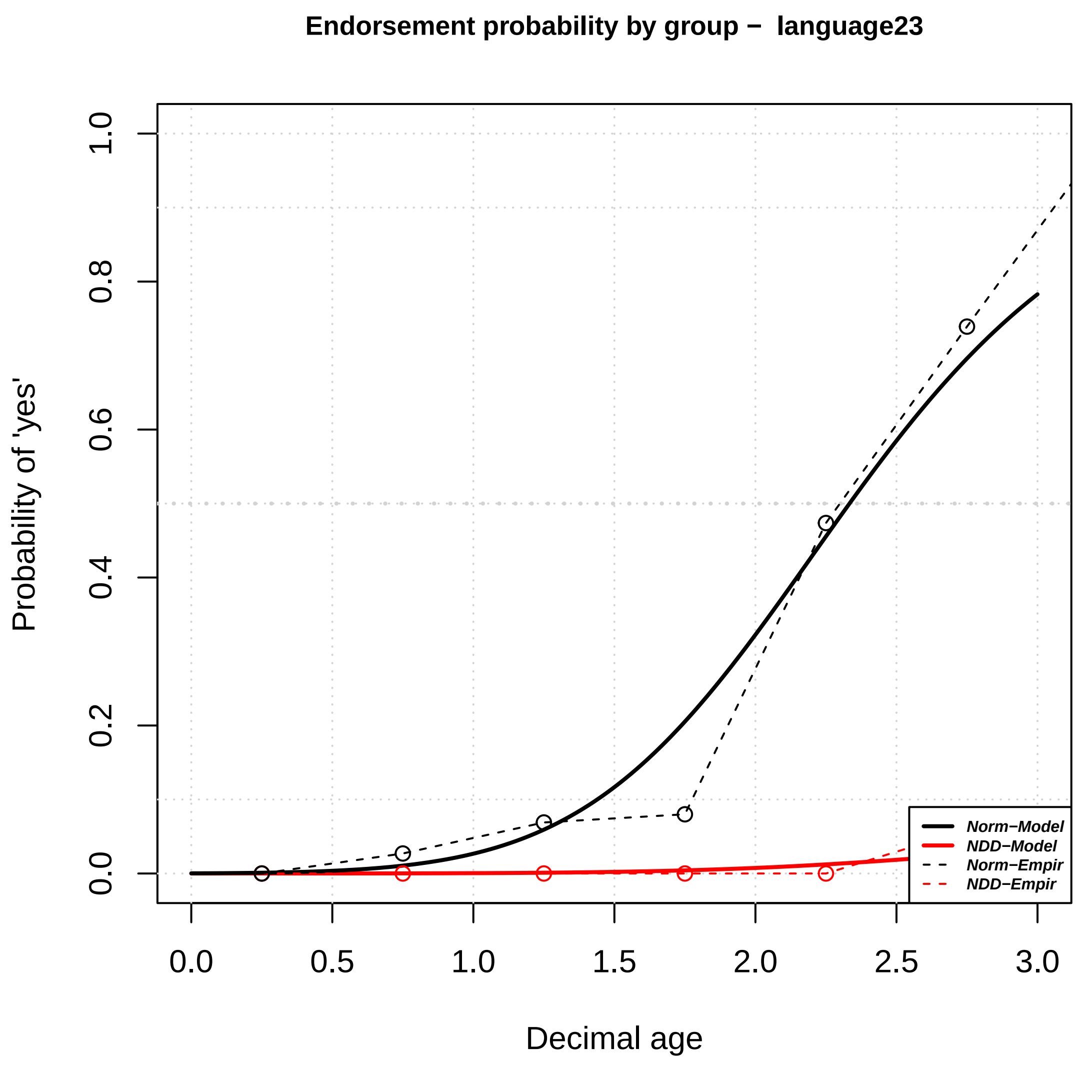
Endorsement probability by group – language23
Probability of 'yes'
0.0
0.2
0.4
0.6
0.8
1.0
0.0
0.5
1.0
1.5
2.0
2.5
3.0
Decimal age
Norm−Model
NDD−Model
Norm−Empir
NDD−Empir

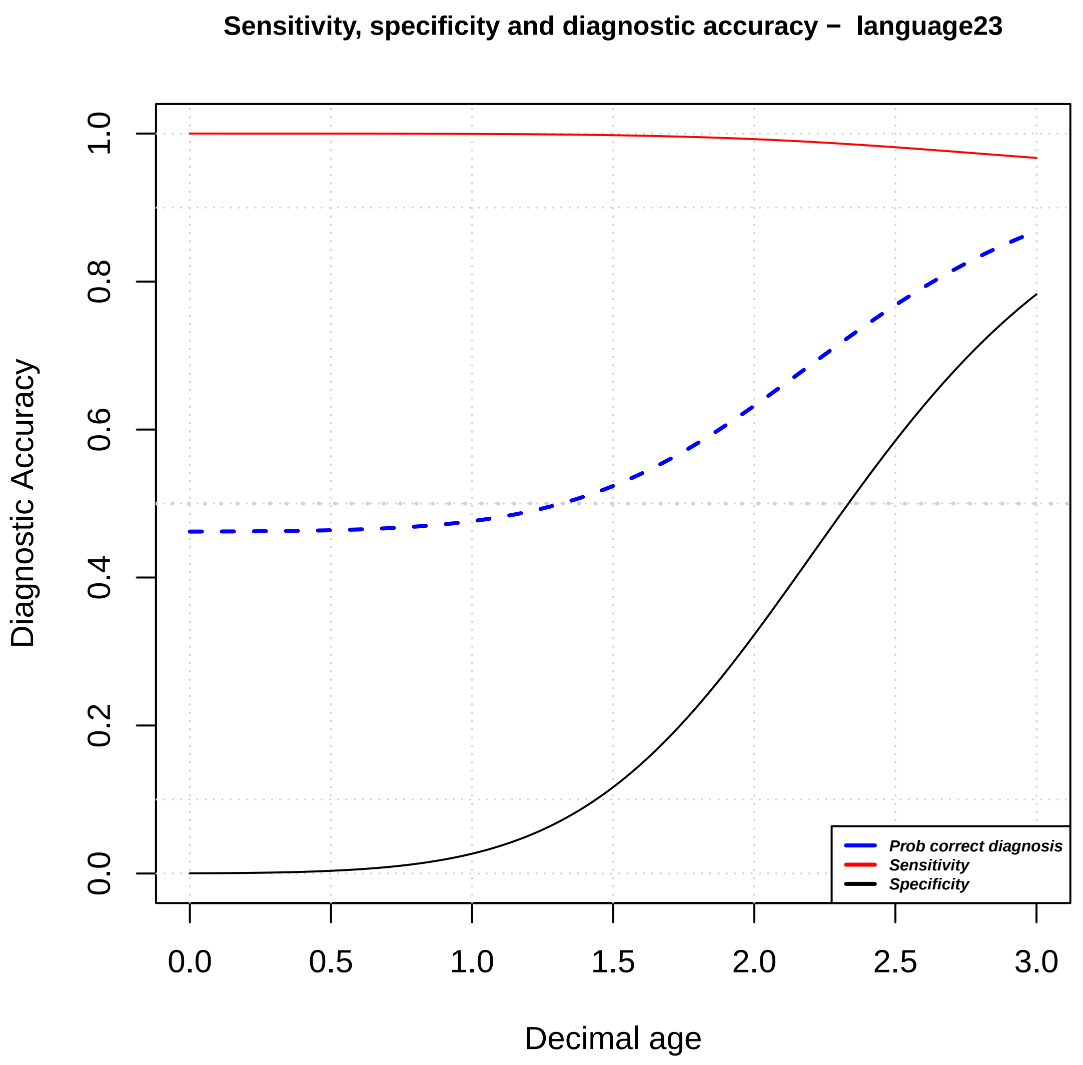
Sensitivity, specificity and diagnostic accuracy – language23
Diagnostic Accuracy
0.0
0.2
0.4
0.6
0.8
1.0
0.0
0.5
1.0
1.5
2.0
2.5
3.0
Decimal age
Prob correct diagnosis
Sensitivity
Specificity

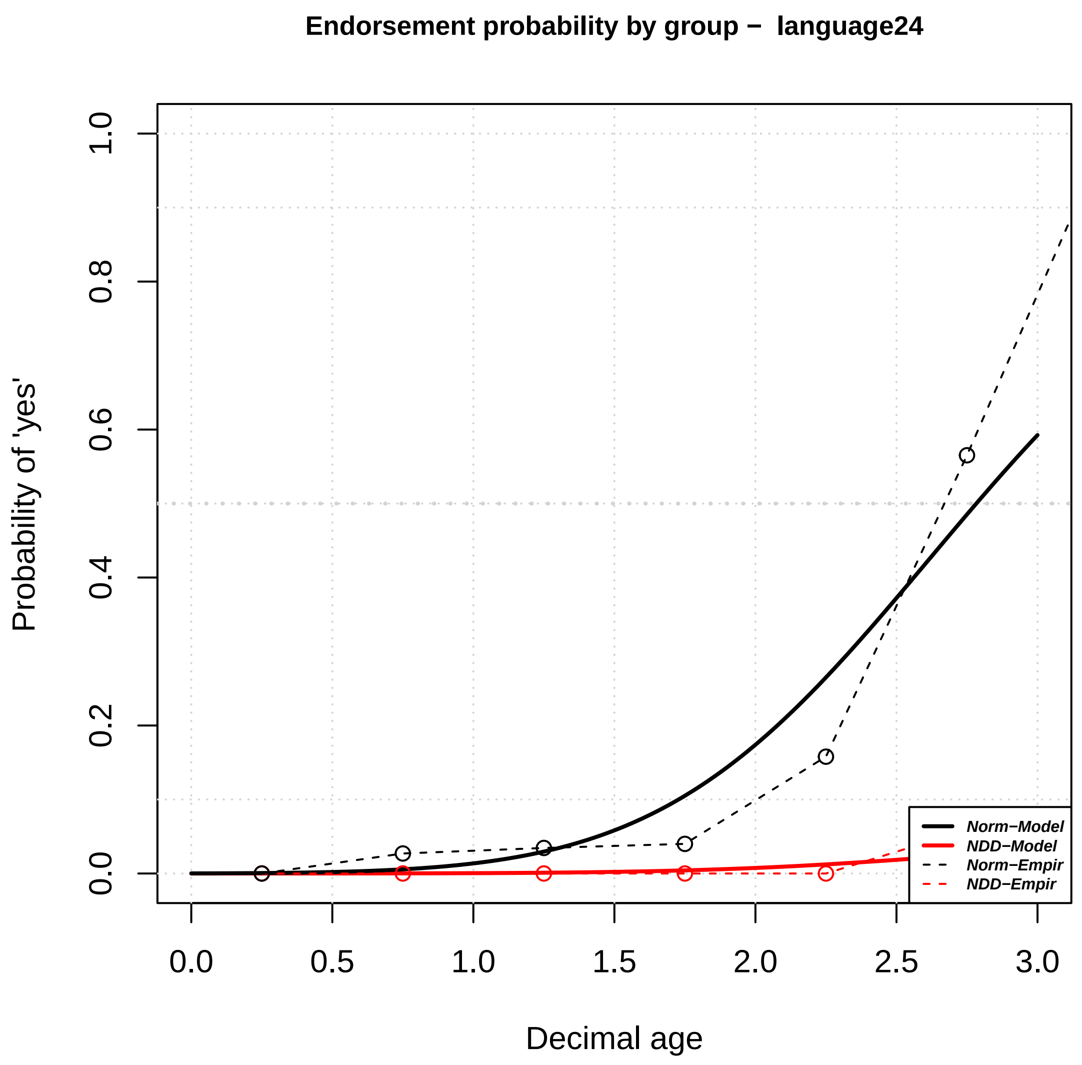
Endorsement probability by group – language24
Probability of 'yes'
Decimal age
0.0
0.2
0.4
0.6
0.8
1.0
0.0
0.5
1.0
1.5
2.0
2.5
3.0
Norm–Model
NDD–Model
Norm–Empir
NDD–Empir

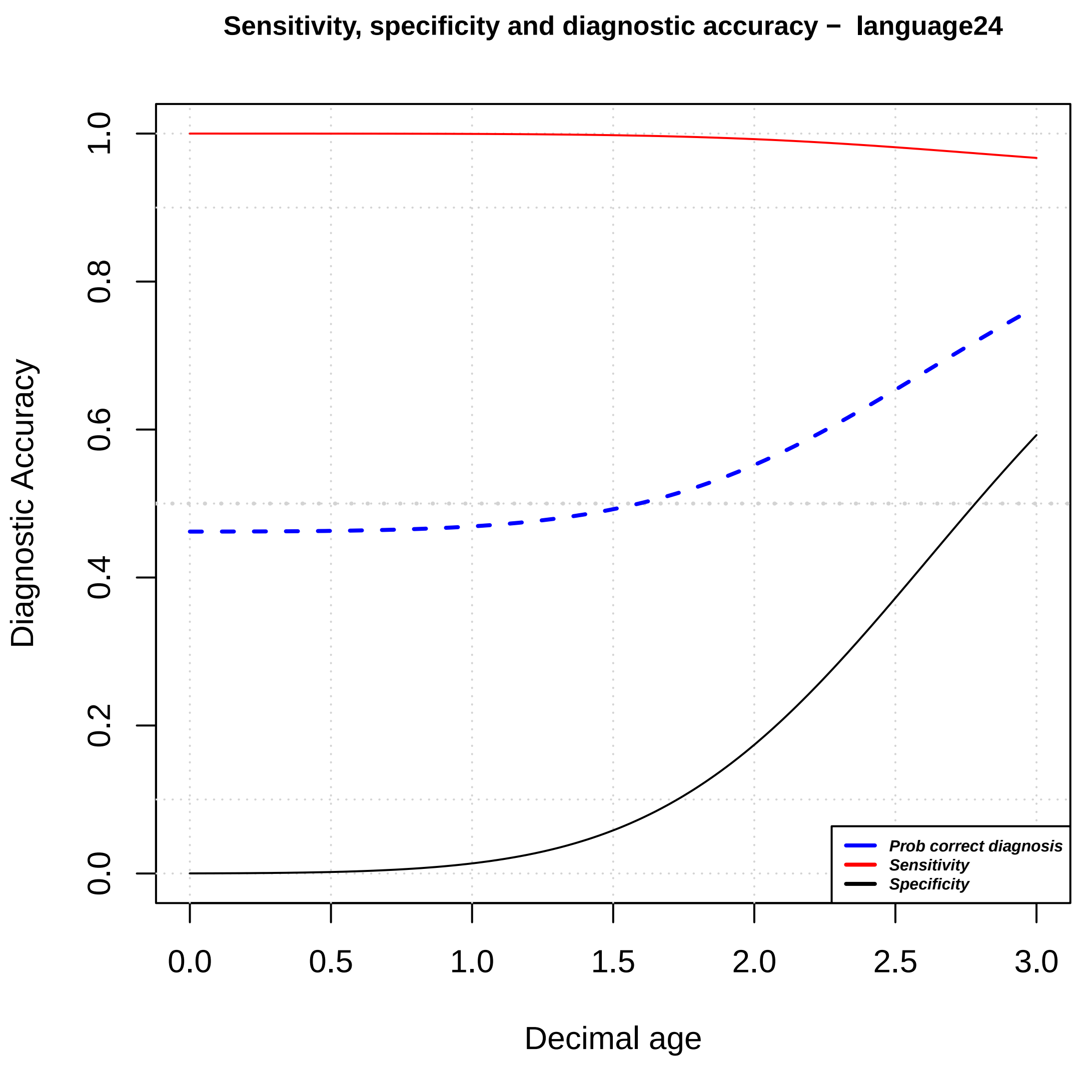
Sensitivity, specificity and diagnostic accuracy – language24
Diagnostic Accuracy
Decimal age
0.0
0.2
0.4
0.6
0.8
1.0
0.0
0.5
1.0
1.5
2.0
2.5
3.0
Prob correct diagnosis
Sensitivity
Specificity

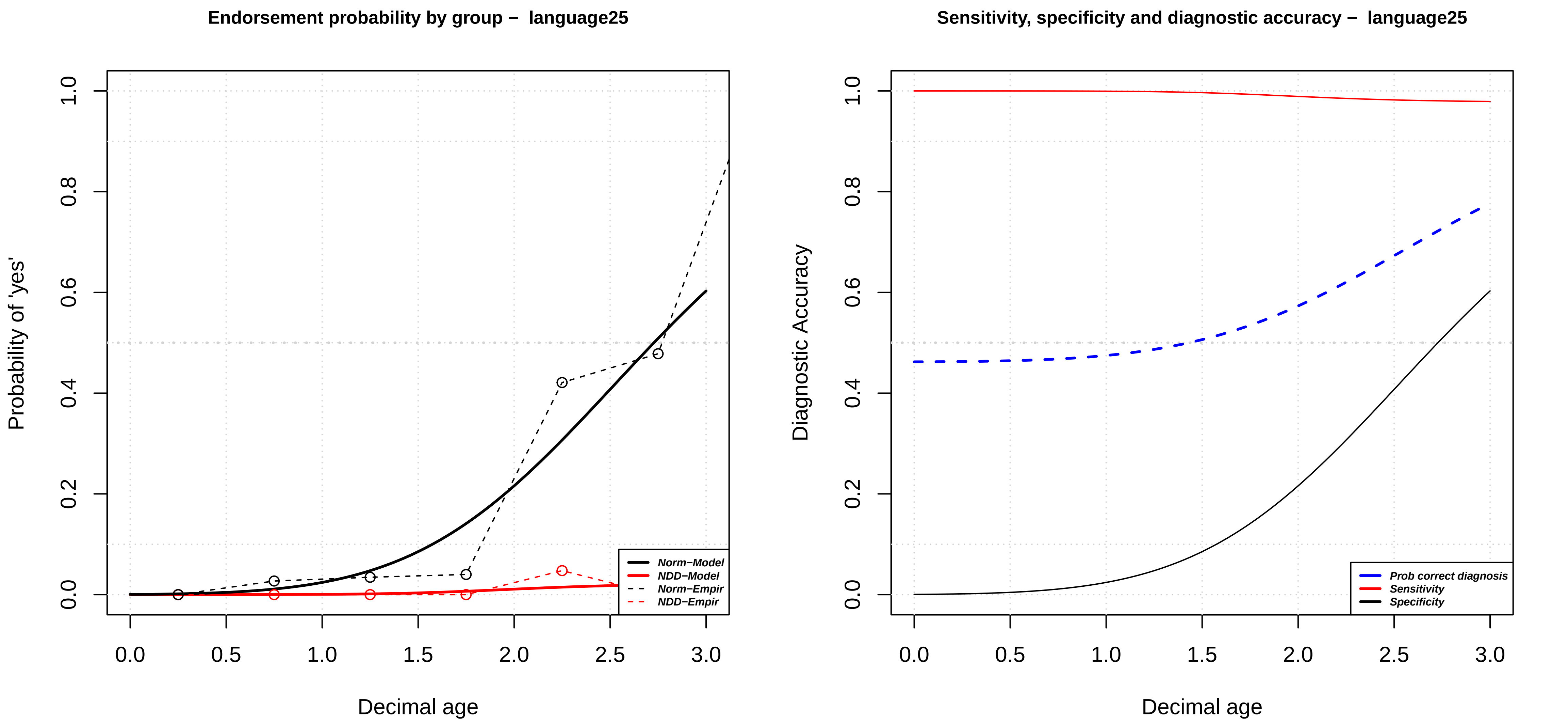

Endorsement probability by group – language25
Probability of 'yes'
Decimal age
0.0
0.2
0.4
0.6
0.8
1.0
0.0
0.5
1.0
1.5
2.0
2.5
3.0
Norm–Model
NDD–Model
Norm–Empir
NDD–Empir
Sensitivity, specificity and diagnostic accuracy – language25
Diagnostic Accuracy
Decimal age
Prob correct diagnosis
Sensitivity
Specificity

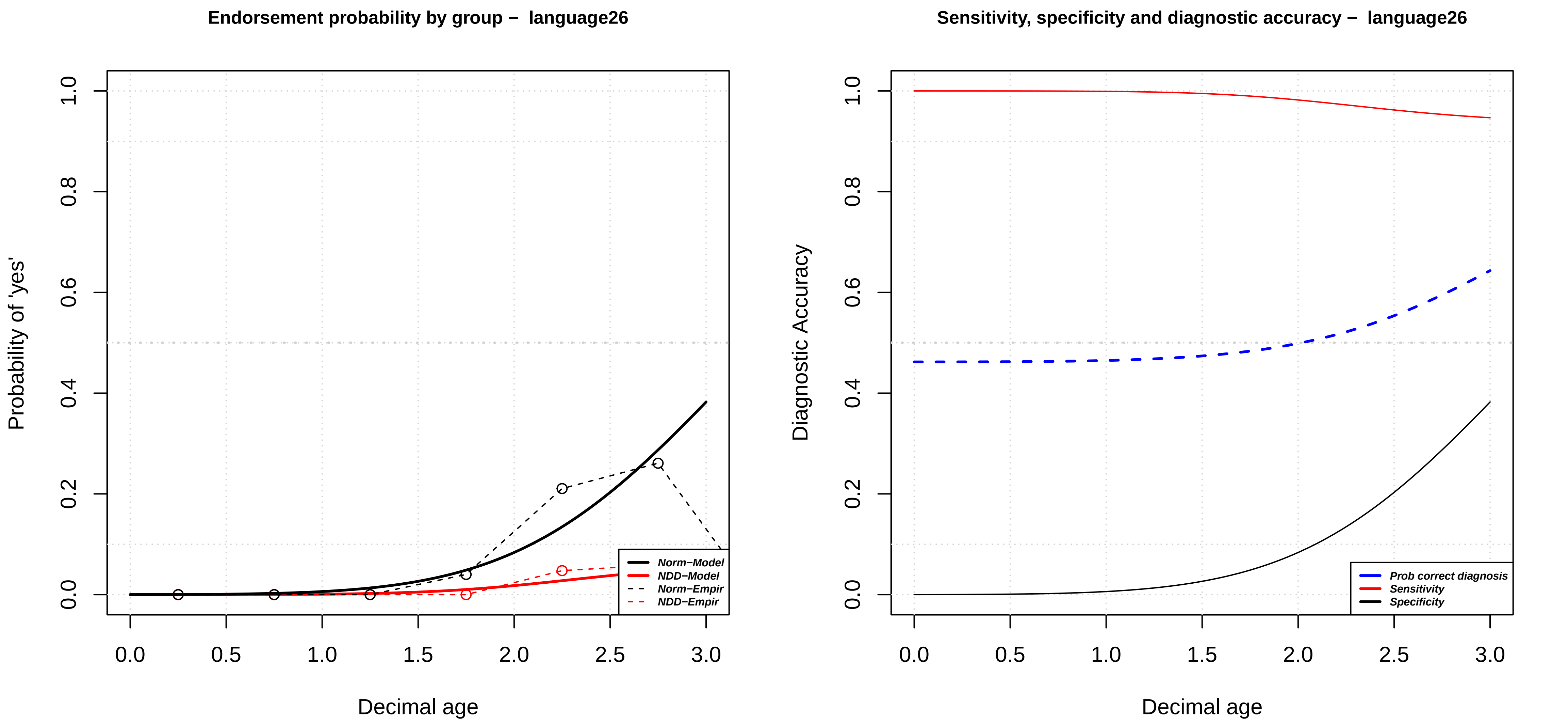
Endorsement probability by group – language26
Probability of 'yes'
Decimal age
0.0
0.2
0.4
0.6
0.8
1.0
0.0
0.5
1.0
1.5
2.0
2.5
3.0
Norm–Model
NDD–Model
Norm–Empir
NDD–Empir
Sensitivity, specificity and diagnostic accuracy – language26
Diagnostic Accuracy
Prob correct diagnosis
Sensitivity
Specificity

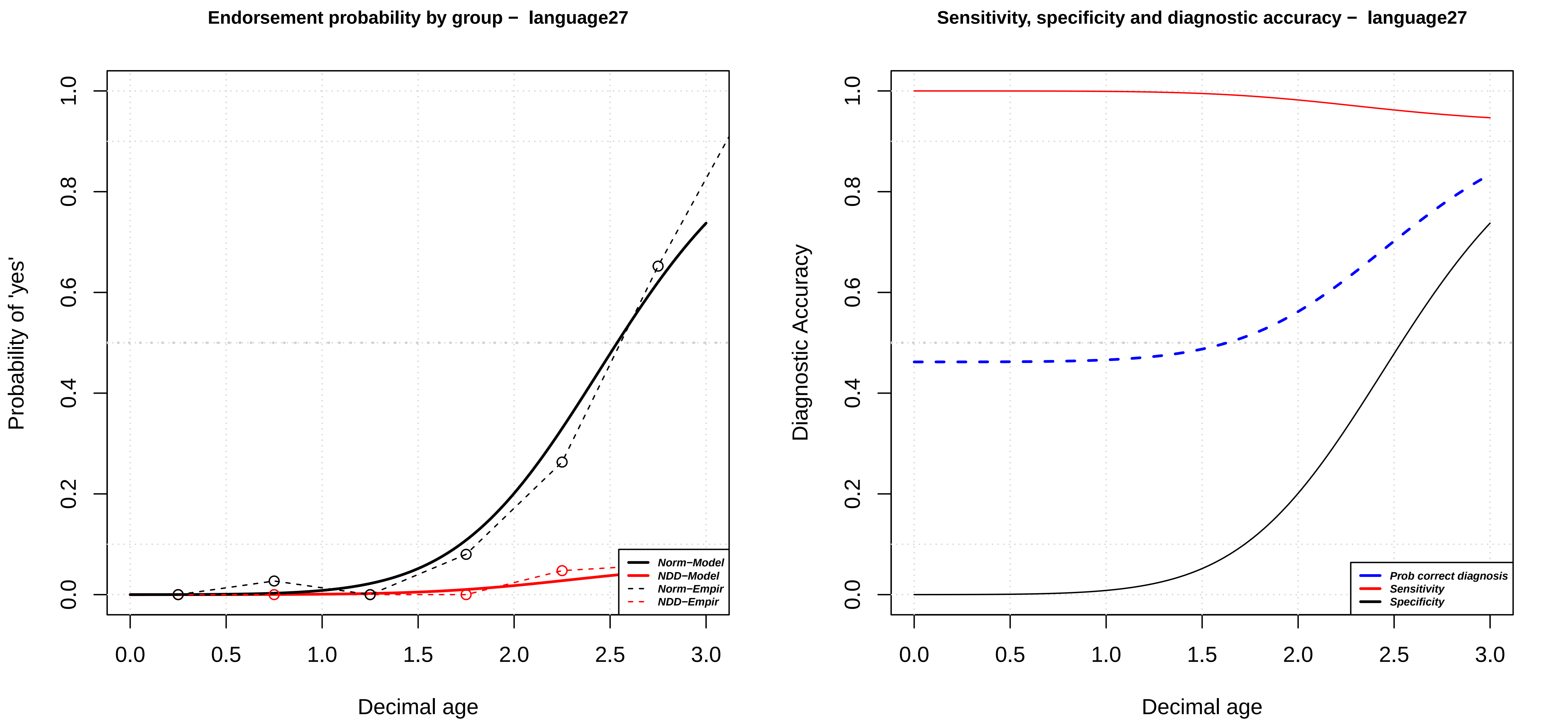

Endorsement probability by group − language27
Probability of 'yes'
Decimal age
0.0
0.2
0.4
0.6
0.8
1.0
0.0
0.5
1.0
1.5
2.0
2.5
3.0
Norm−Model
NDD−Model
Norm−Empir
NDD−Empir
Sensitivity, specificity and diagnostic accuracy − language27
Diagnostic Accuracy
Decimal age
Prob correct diagnosis
Sensitivity
Specificity

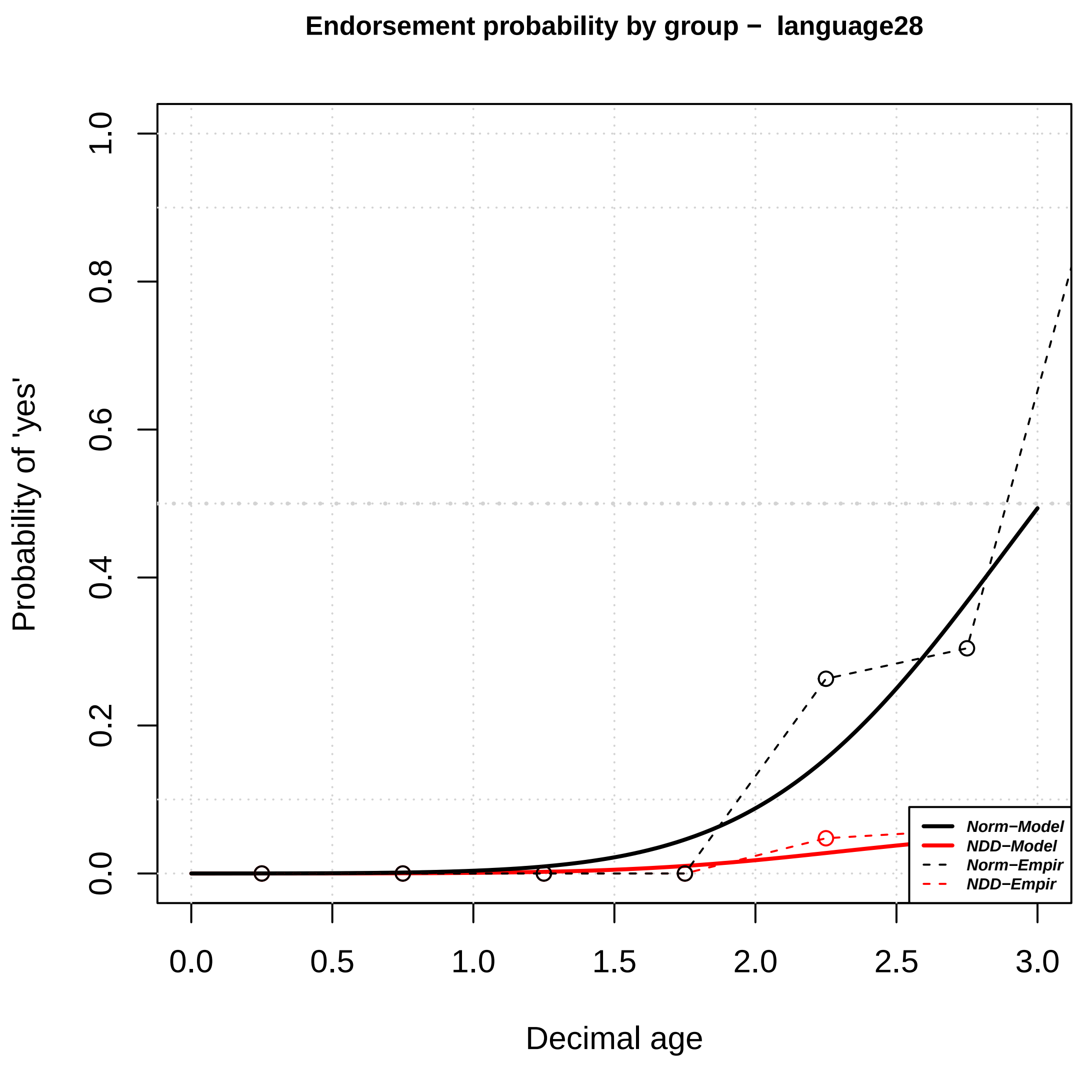
Endorsement probability by group – language28
Probability of 'yes'
Decimal age
0.0
0.2
0.4
0.6
0.8
1.0
0.0
0.5
1.0
1.5
2.0
2.5
3.0
Norm–Model
NDD–Model
Norm–Empir
NDD–Empir

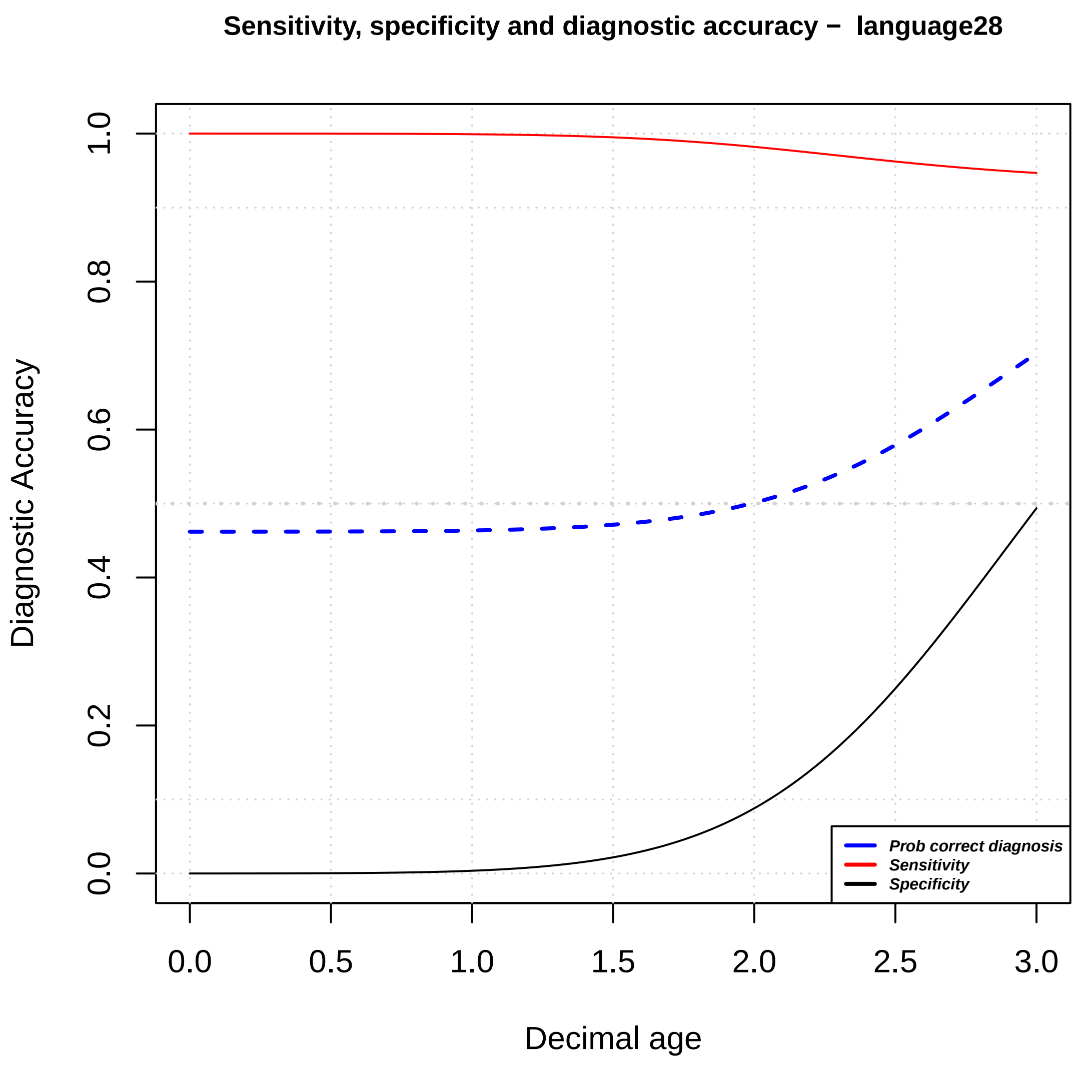
Sensitivity, specificity and diagnostic accuracy – language28
Diagnostic Accuracy
Decimal age
0.0
0.2
0.4
0.6
0.8
1.0
0.0
0.5
1.0
1.5
2.0
2.5
3.0
Prob correct diagnosis
Sensitivity
Specificity

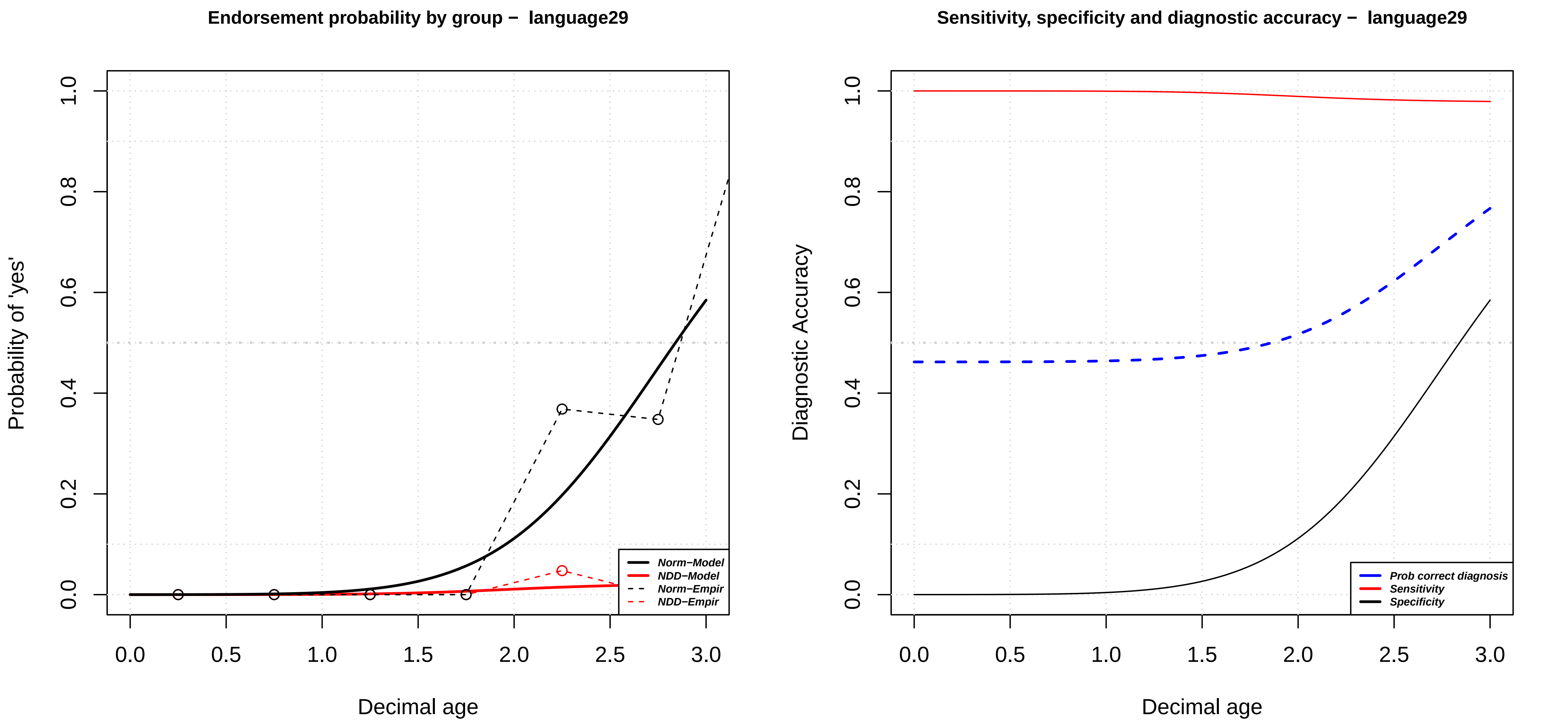

Endorsement probability by group – language29
Probability of 'yes'
Decimal age
Norm–Model
NDD–Model
Norm–Empir
NDD–Empir
Sensitivity, specificity and diagnostic accuracy – language29
Diagnostic Accuracy
Decimal age
Prob correct diagnosis
Sensitivity
Specificity

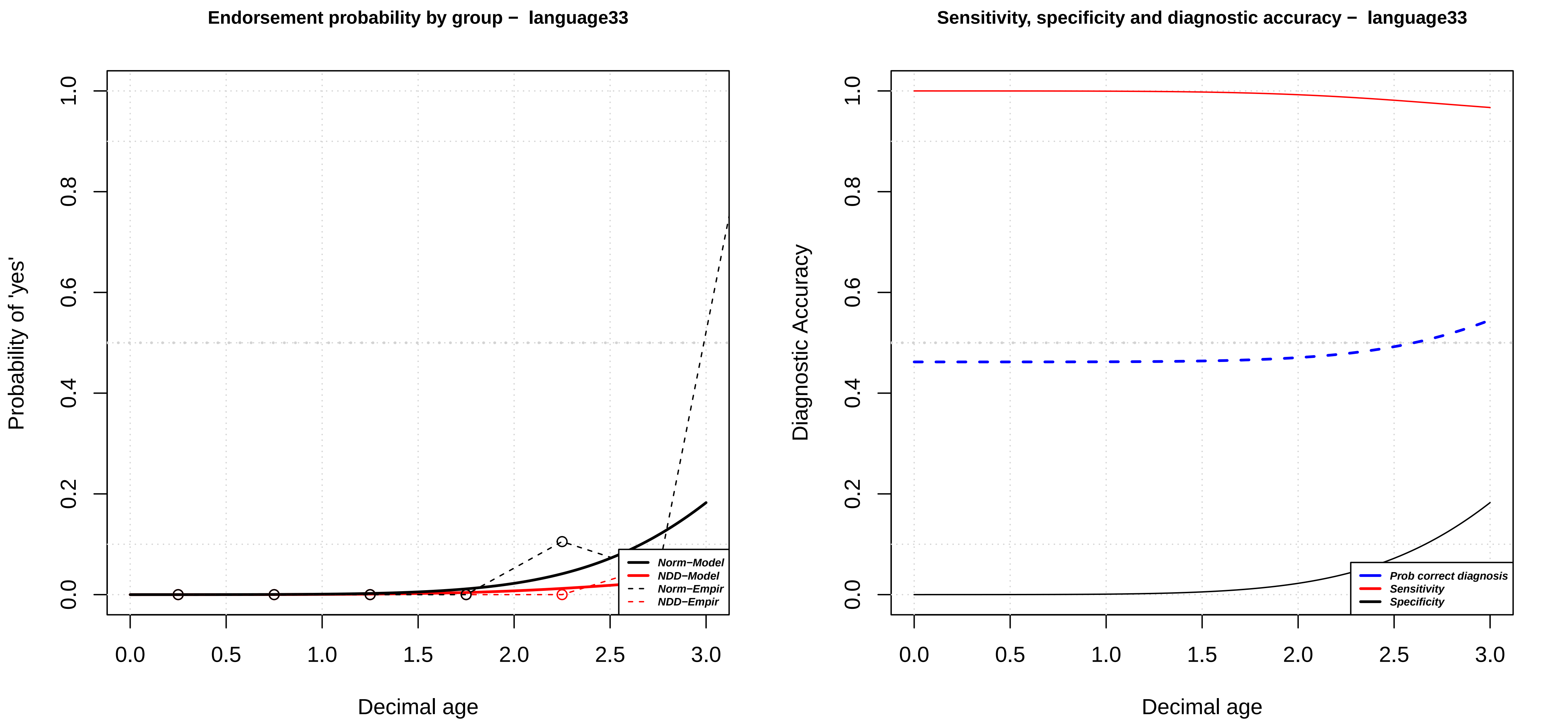

Endorsement probability by group – language33
Probability of 'yes'
Decimal age
Norm–Model
NDD–Model
Norm–Empir
NDD–Empir
Sensitivity, specificity and diagnostic accuracy – language33
Diagnostic Accuracy
Decimal age
Prob correct diagnosis
Sensitivity
Specificity

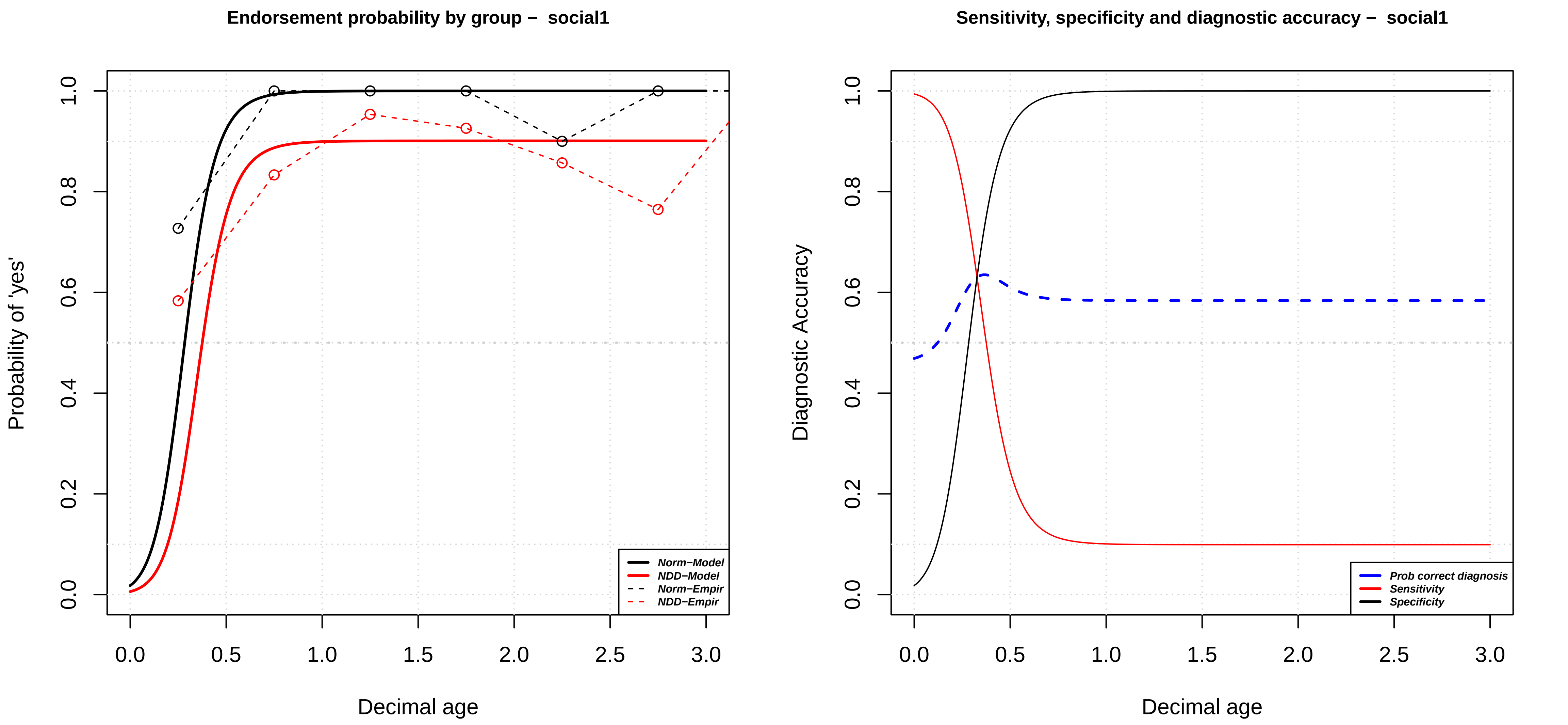

Endorsement probability by group − social1
Probability of 'yes'
Decimal age
Norm−Model
NDD−Model
Norm−Empir
NDD−Empir
0.0
0.2
0.4
0.6
0.8
1.0
0.0
0.5
1.0
1.5
2.0
2.5
3.0
Sensitivity, specificity and diagnostic accuracy − social1
Diagnostic Accuracy
Decimal age
Prob correct diagnosis
Sensitivity
Specificity

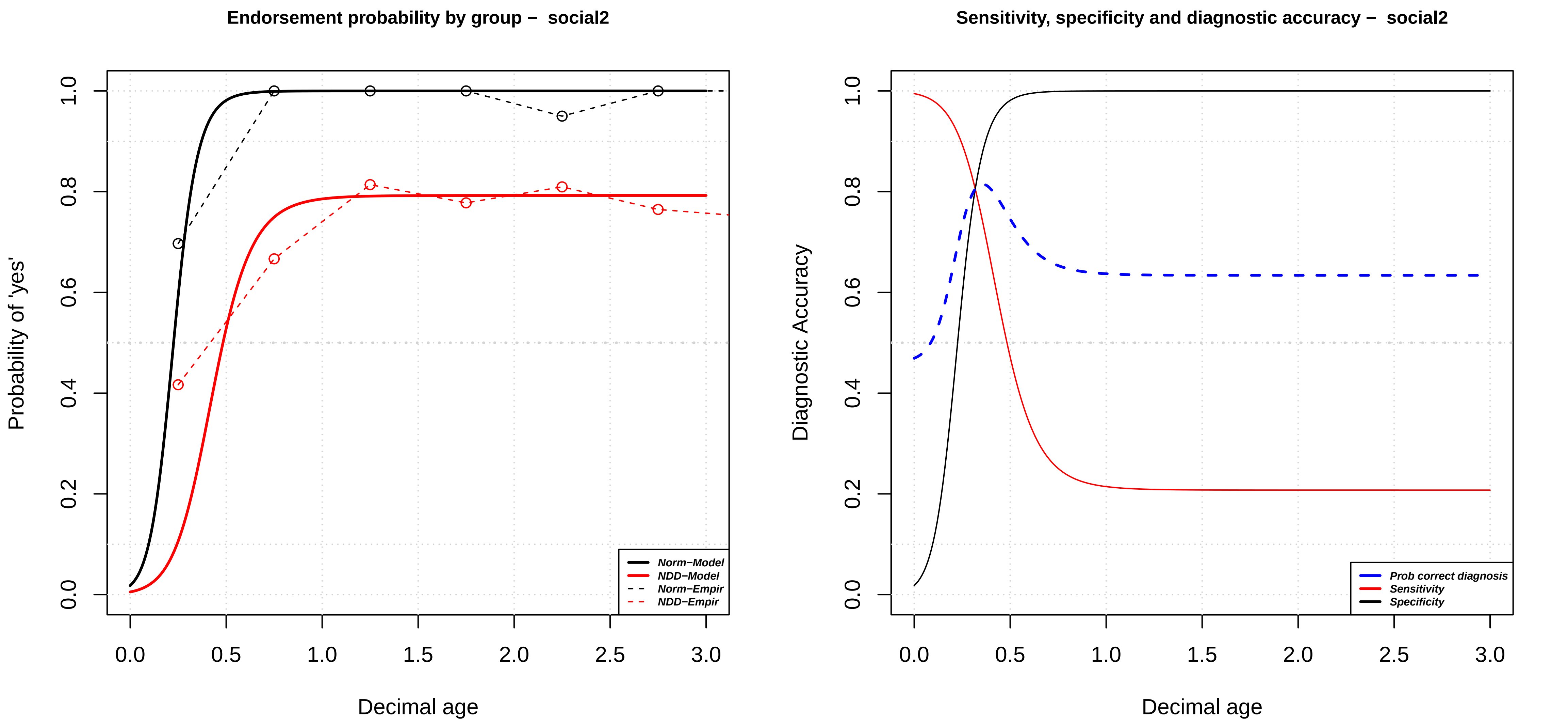

Endorsement probability by group – social2
Probability of 'yes'
Decimal age
Norm–Model
NDD–Model
Norm–Empir
NDD–Empir
0.0
0.2
0.4
0.6
0.8
1.0
0.0
0.5
1.0
1.5
2.0
2.5
3.0
Sensitivity, specificity and diagnostic accuracy – social2
Diagnostic Accuracy
Decimal age
Prob correct diagnosis
Sensitivity
Specificity

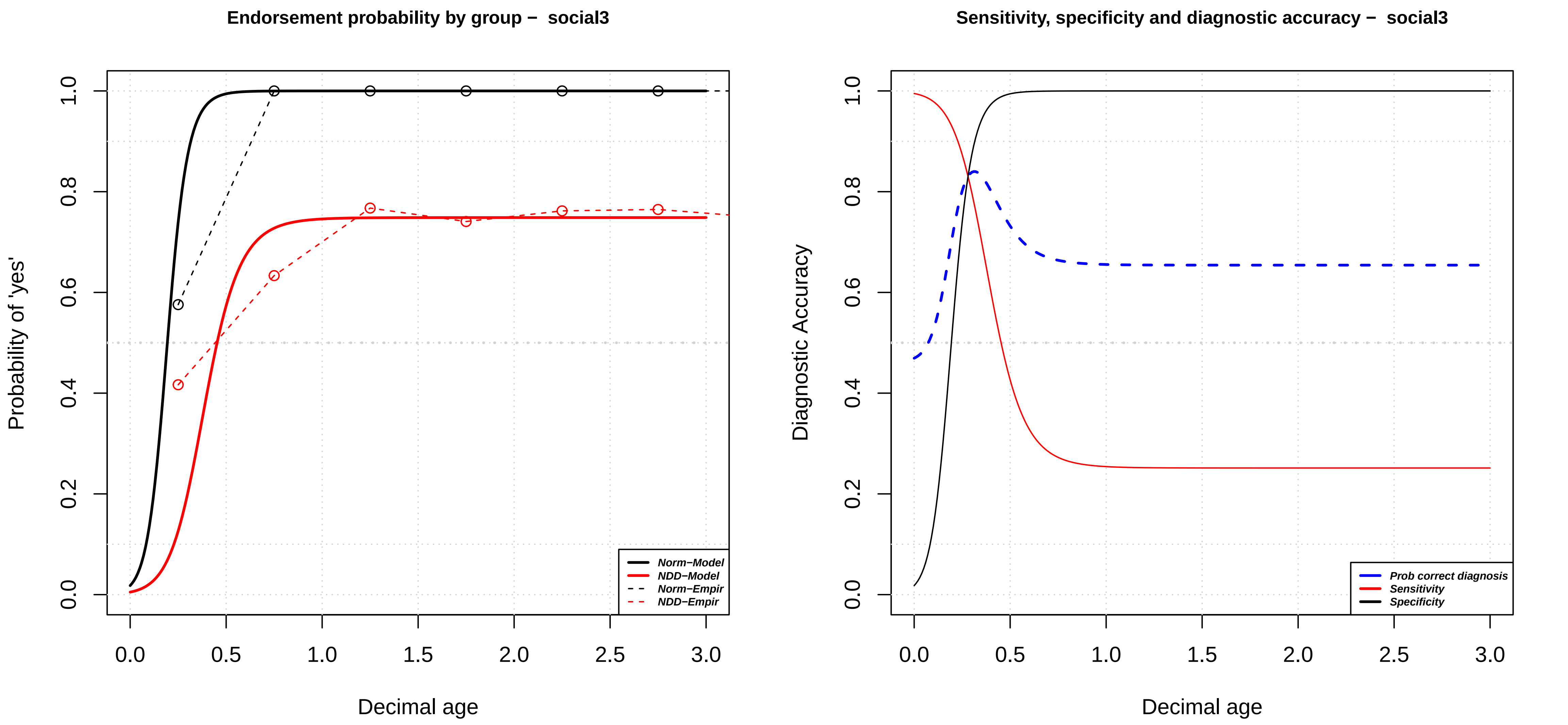
Endorsement probability by group – social3
Probability of 'yes'
Decimal age
0.0
0.2
0.4
0.6
0.8
1.0
0.0
0.5
1.0
1.5
2.0
2.5
3.0
Norm–Model
NDD–Model
Norm–Empir
NDD–Empir
Sensitivity, specificity and diagnostic accuracy – social3
Diagnostic Accuracy
Decimal age
Prob correct diagnosis
Sensitivity
Specificity

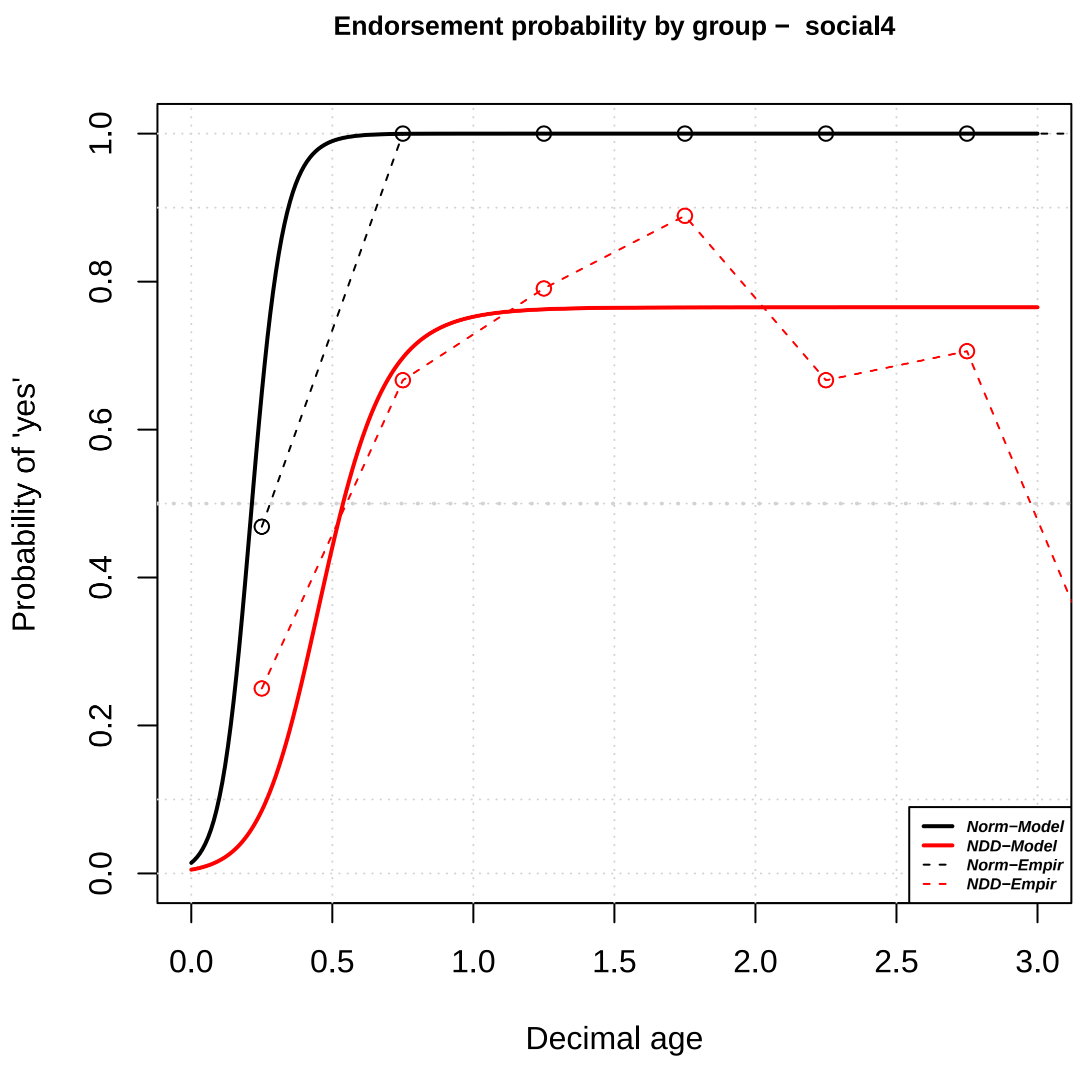

Endorsement probability by group – social4
Probability of 'yes'
0.0
0.2
0.4
0.6
0.8
1.0
0.0
0.5
1.0
1.5
2.0
2.5
3.0
Decimal age
Norm−Model
NDD−Model
Norm−Empir
NDD−Empir


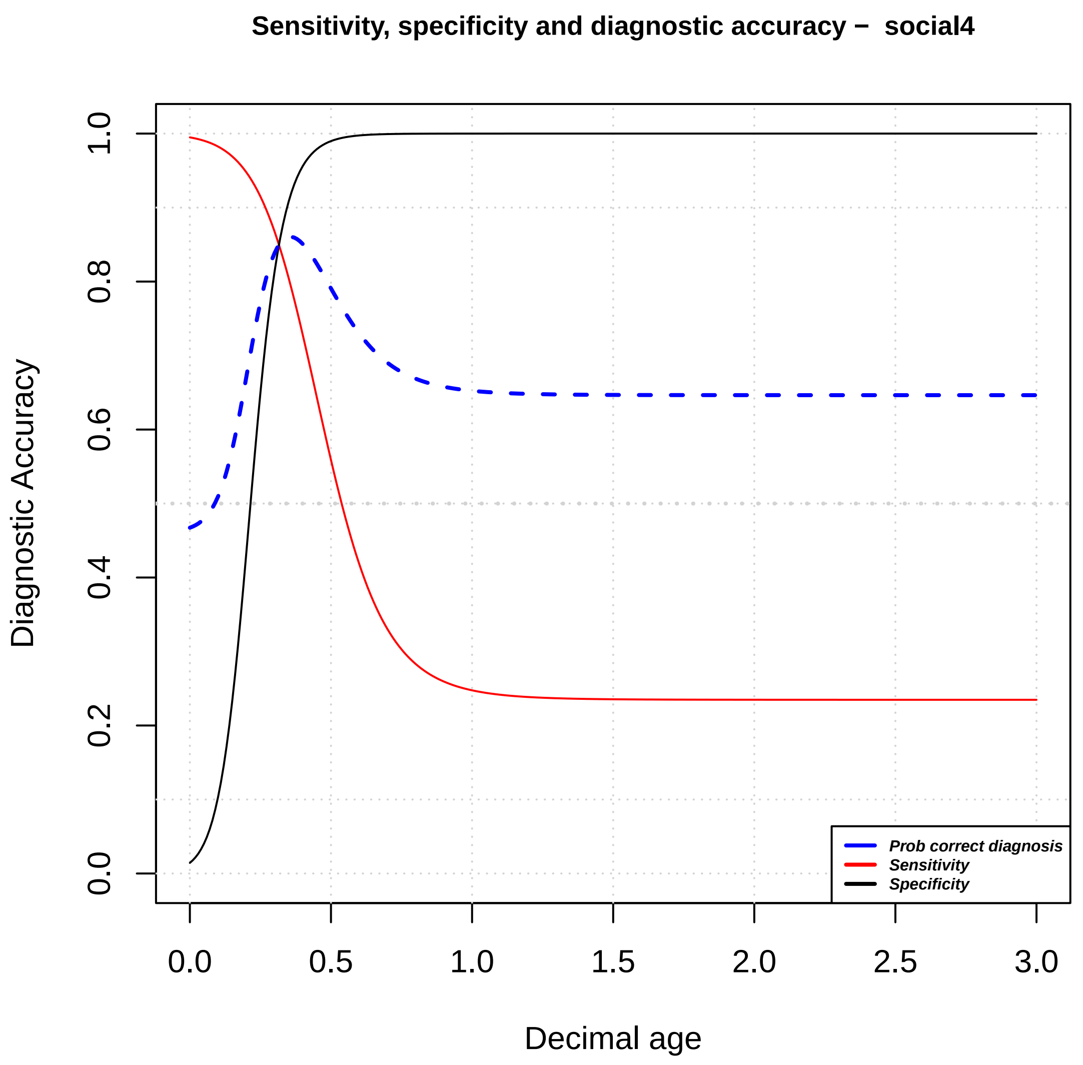

Sensitivity, specificity and diagnostic accuracy – social4
Diagnostic Accuracy
0.0
0.2
0.4
0.6
0.8
1.0
0.0
0.5
1.0
1.5
2.0
2.5
3.0
Decimal age
Prob correct diagnosis
Sensitivity
Specificity

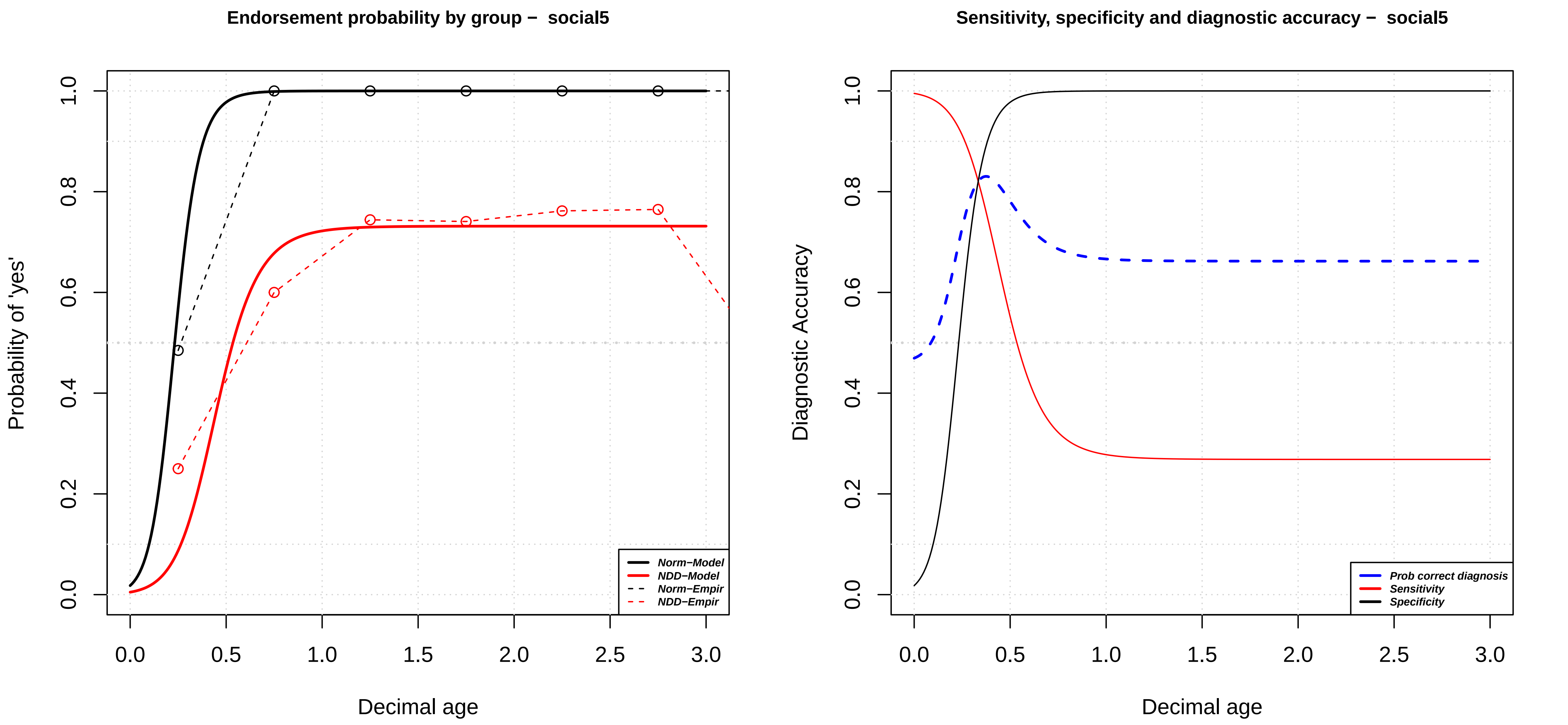

Endorsement probability by group – social5
Probability of 'yes'
Decimal age
0.0
0.2
0.4
0.6
0.8
1.0
0.0
0.5
1.0
1.5
2.0
2.5
3.0
Norm–Model
NDD–Model
Norm–Empir
NDD–Empir
Sensitivity, specificity and diagnostic accuracy – social5
Diagnostic Accuracy
Decimal age
0.0
0.2
0.4
0.6
0.8
1.0
0.0
0.5
1.0
1.5
2.0
2.5
3.0
Prob correct diagnosis
Sensitivity
Specificity

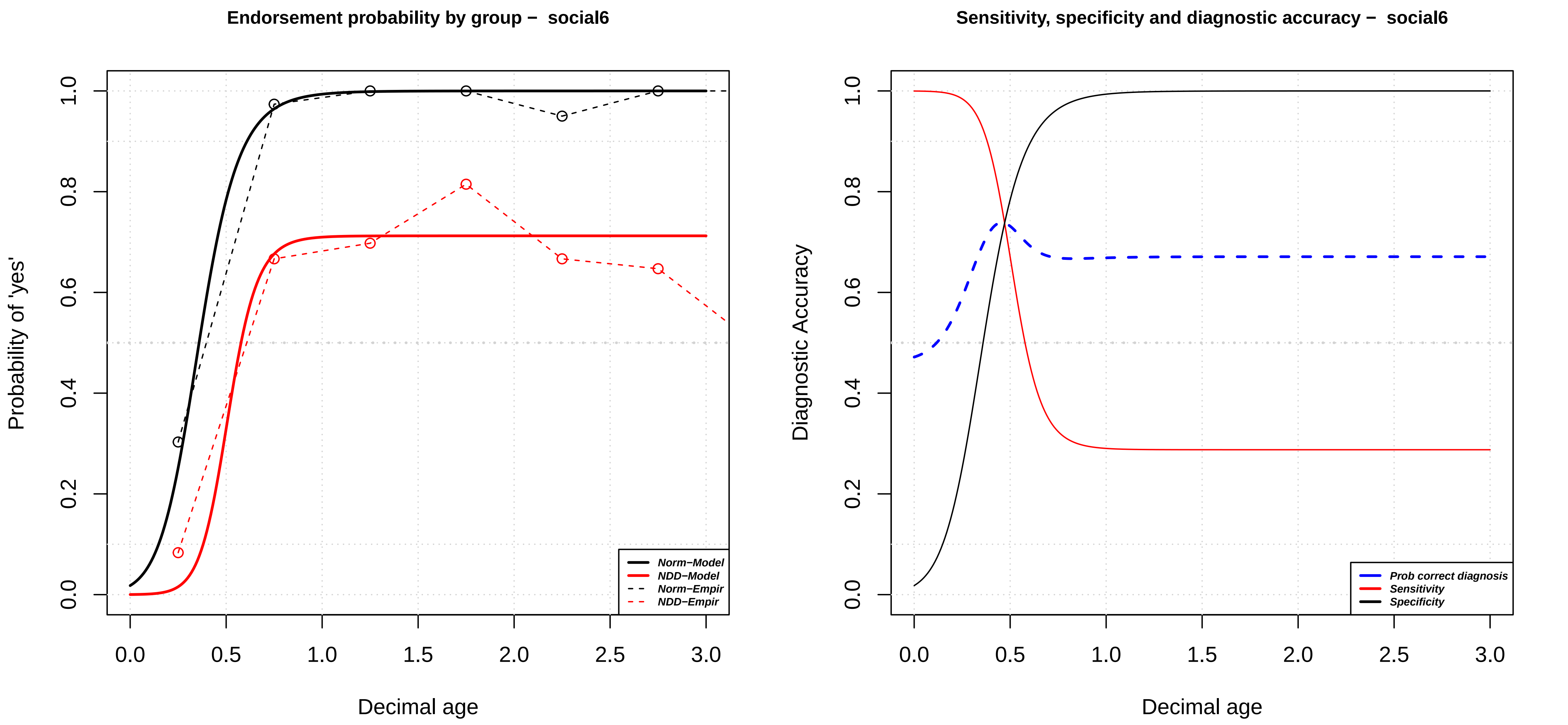
Endorsement probability by group – social6
Probability of 'yes'
Decimal age
0.0
0.2
0.4
0.6
0.8
1.0
0.5
1.5
2.0
2.5
3.0
Norm–Model
NDD–Model
Norm–Empir
NDD–Empir
Sensitivity, specificity and diagnostic accuracy – social6
Diagnostic Accuracy
Decimal age
Prob correct diagnosis
Sensitivity
Specificity

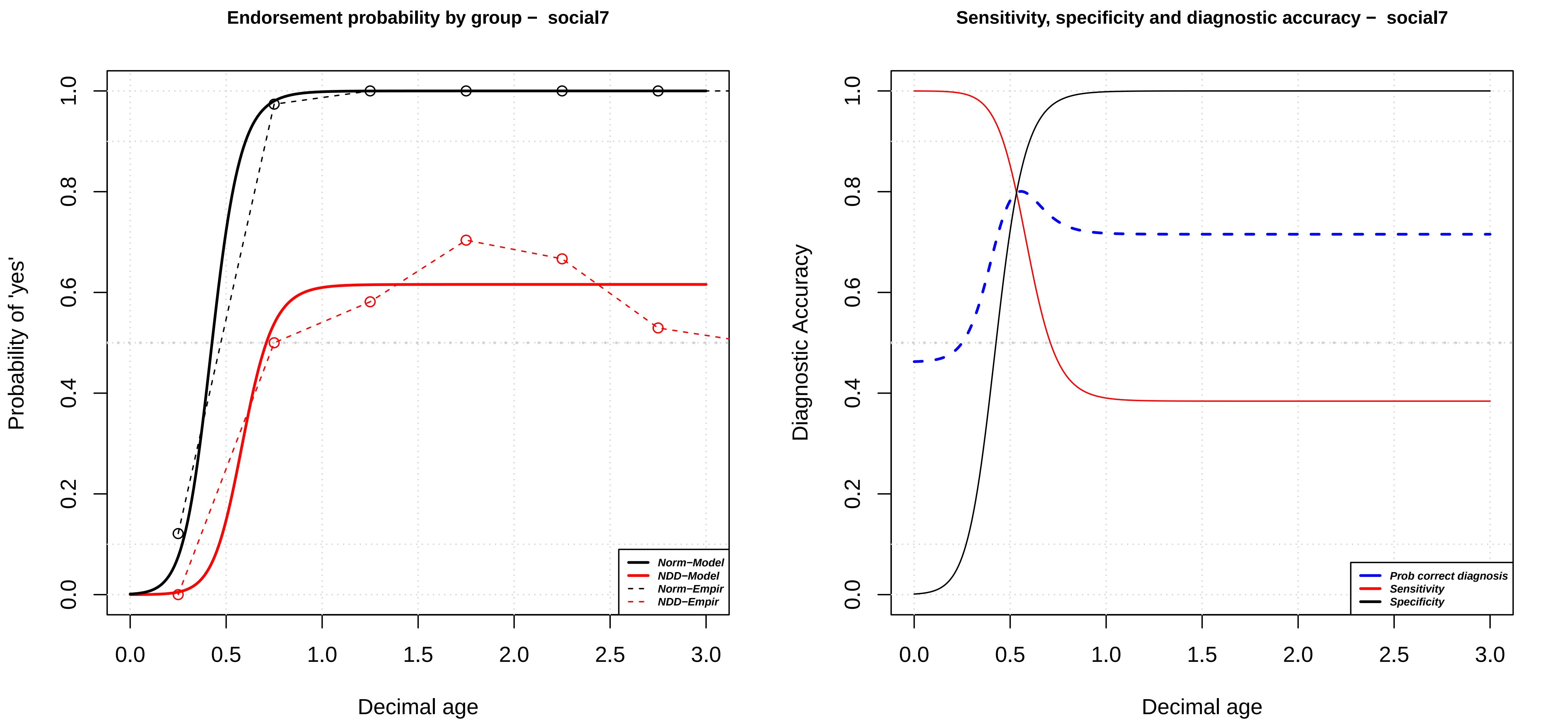

Endorsement probability by group – social7
Probability of 'yes'
Decimal age
Norm−Model
NDD−Model
Norm−Empir
NDD−Empir
0.0
0.2
0.4
0.6
0.8
1.0
0.0
0.5
1.0
1.5
2.0
2.5
3.0
Sensitivity, specificity and diagnostic accuracy – social7
Diagnostic Accuracy
Decimal age
Prob correct diagnosis
Sensitivity
Specificity

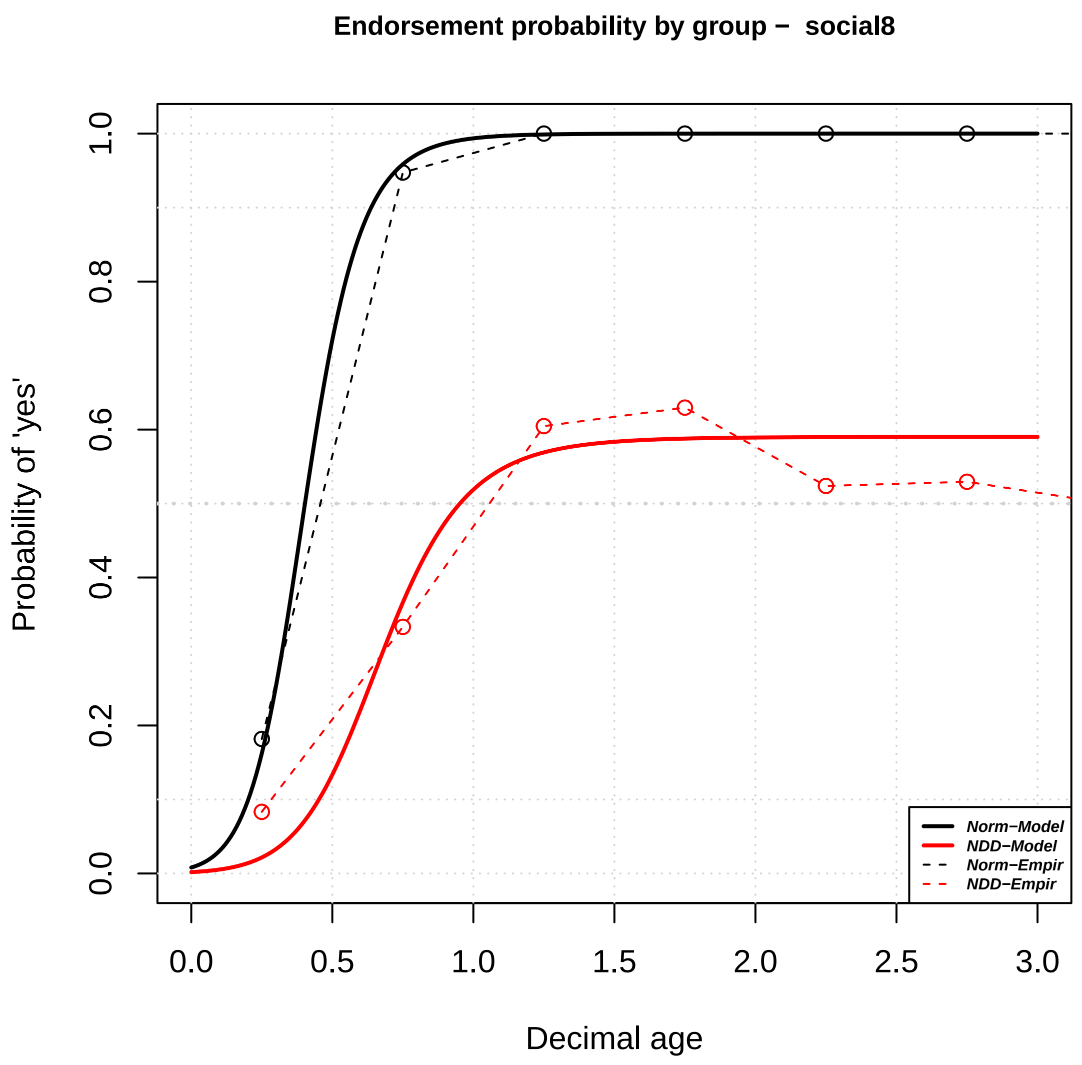
Endorsement probability by group − social8
Probability of 'yes'
Decimal age
0.0
0.2
0.4
0.6
0.8
1.0
0.0
0.5
1.0
1.5
2.0
2.5
3.0
Norm−Model
NDD−Model
Norm−Empir
NDD−Empir

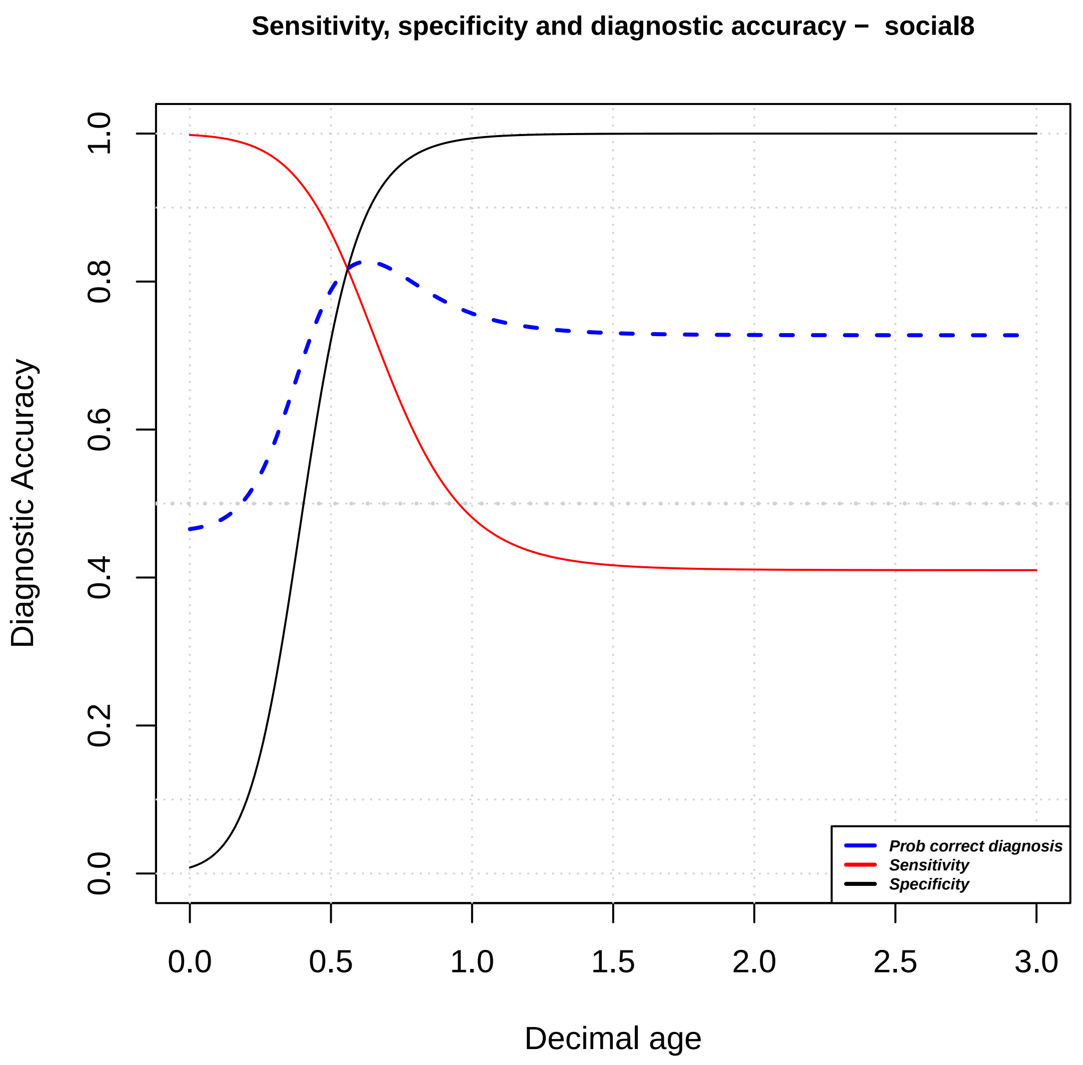
Sensitivity, specificity and diagnostic accuracy − social8
Diagnostic Accuracy
Decimal age
0.0
0.2
0.4
0.6
0.8
1.0
0.0
0.5
1.0
1.5
2.0
2.5
3.0
Prob correct diagnosis
Sensitivity
Specificity

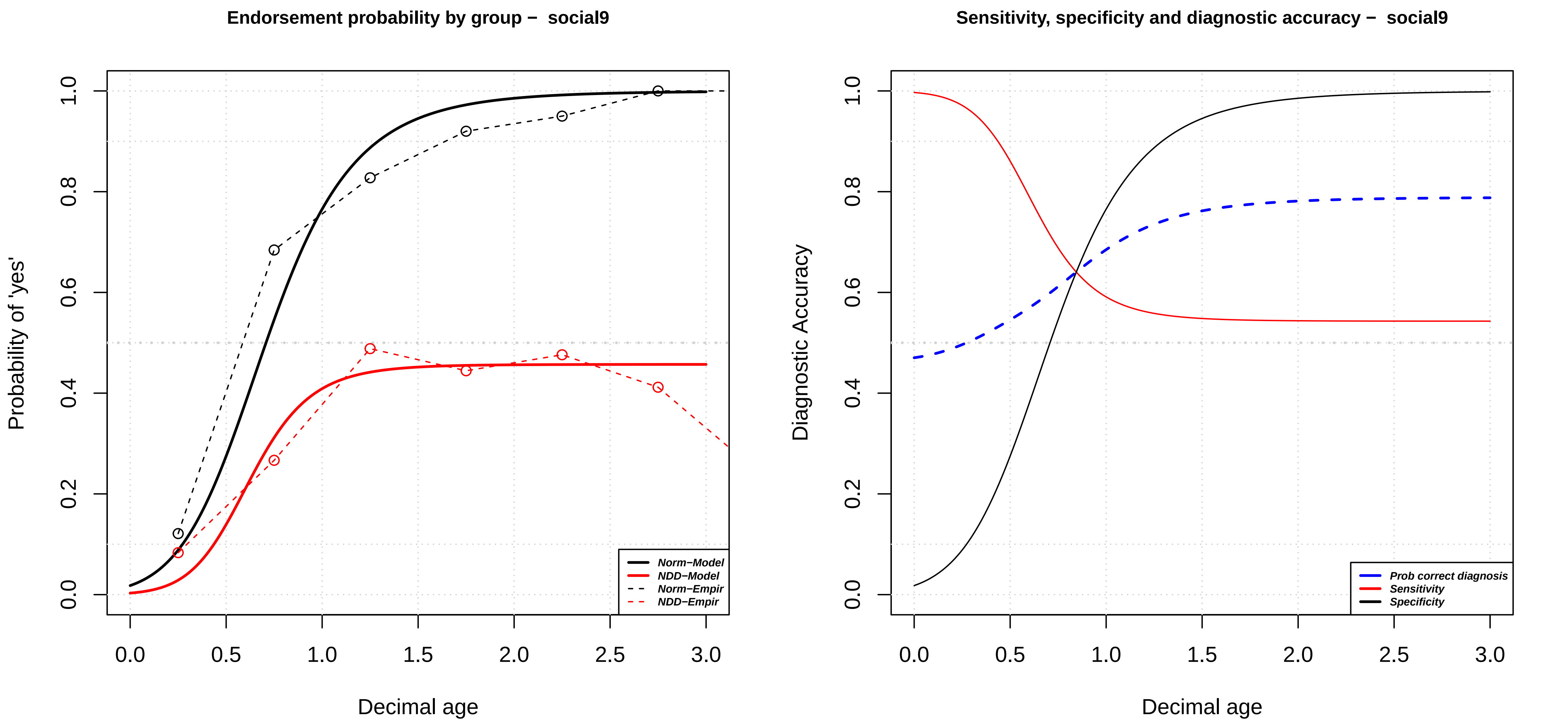
Endorsement probability by group – social9
Probability of 'yes'
Decimal age
0.0
0.2
0.4
0.6
0.8
1.0
0.5
1.5
2.0
2.5
3.0
Norm–Model
NDD–Model
Norm–Empir
NDD–Empir
Sensitivity, specificity and diagnostic accuracy – social9
Diagnostic Accuracy
Decimal age
Prob correct diagnosis
Sensitivity
Specificity

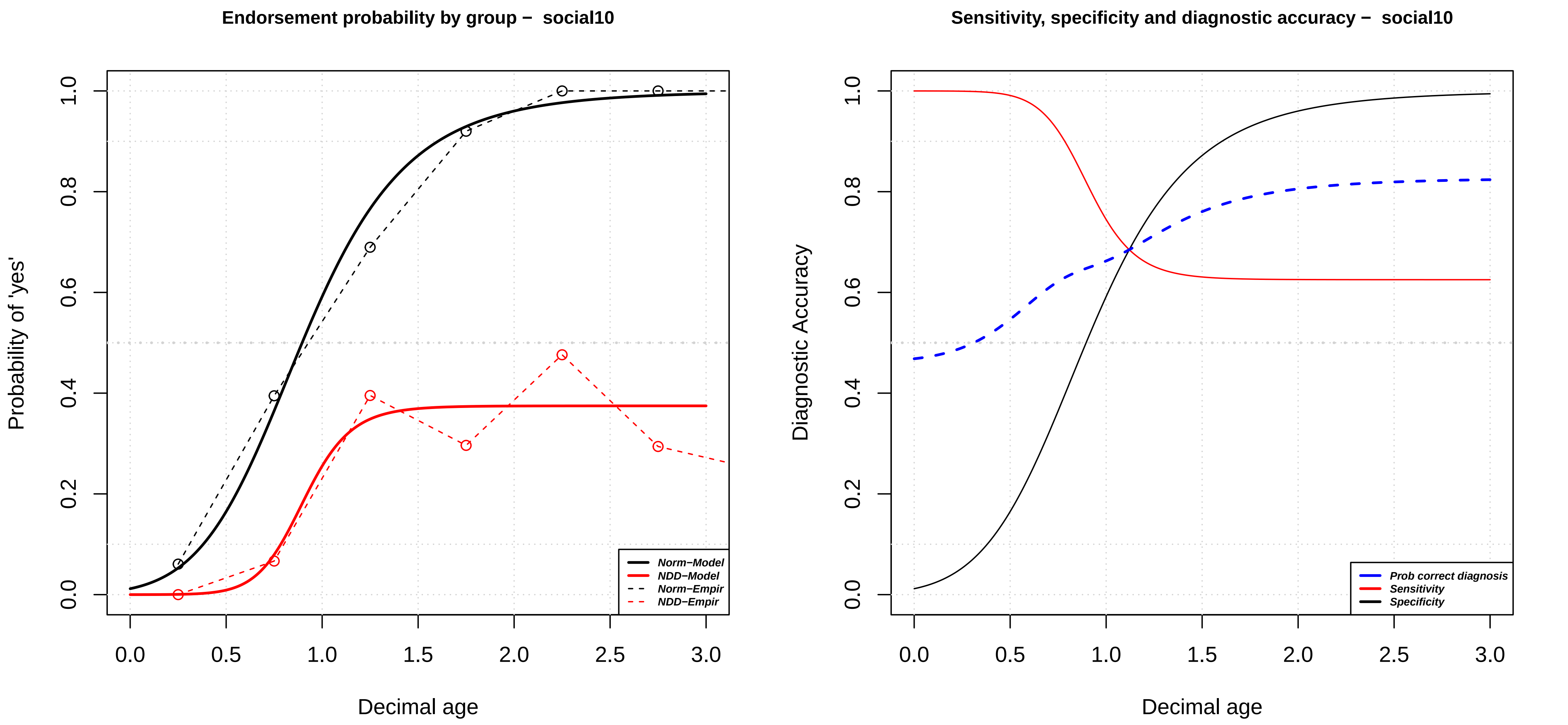

Endorsement probability by group – social10
Probability of 'yes'
Decimal age
Norm–Model
NDD–Model
Norm–Empir
NDD–Empir
Sensitivity, specificity and diagnostic accuracy – social10
Diagnostic Accuracy
Decimal age
Prob correct diagnosis
Sensitivity
Specificity

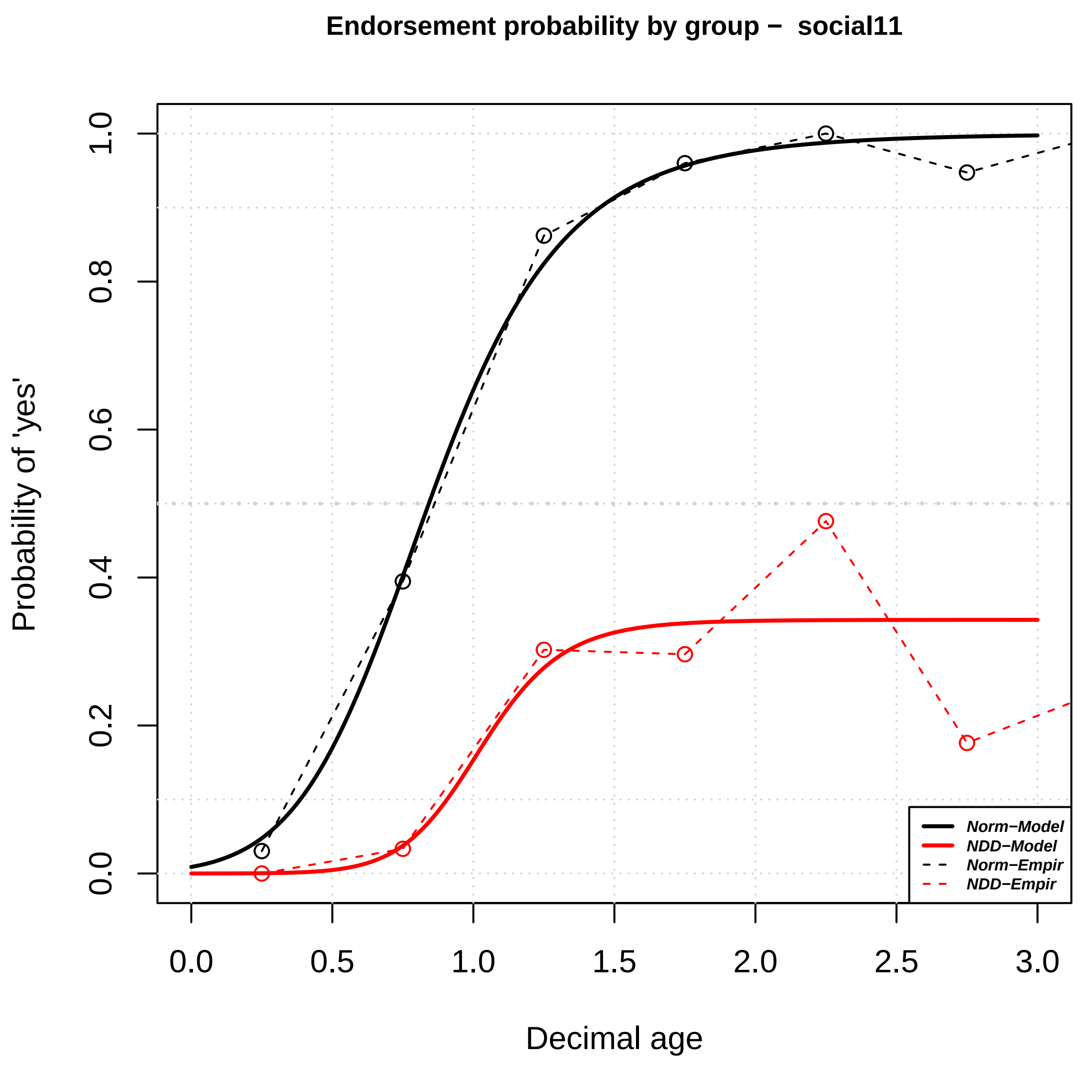
Endorsement probability by group − social11
Probability of 'yes'
Decimal age
0.0
0.2
0.4
0.6
0.8
1.0
0.0
0.5
1.0
1.5
2.0
2.5
3.0
Norm−Model
NDD−Model
Norm−Empir
NDD−Empir

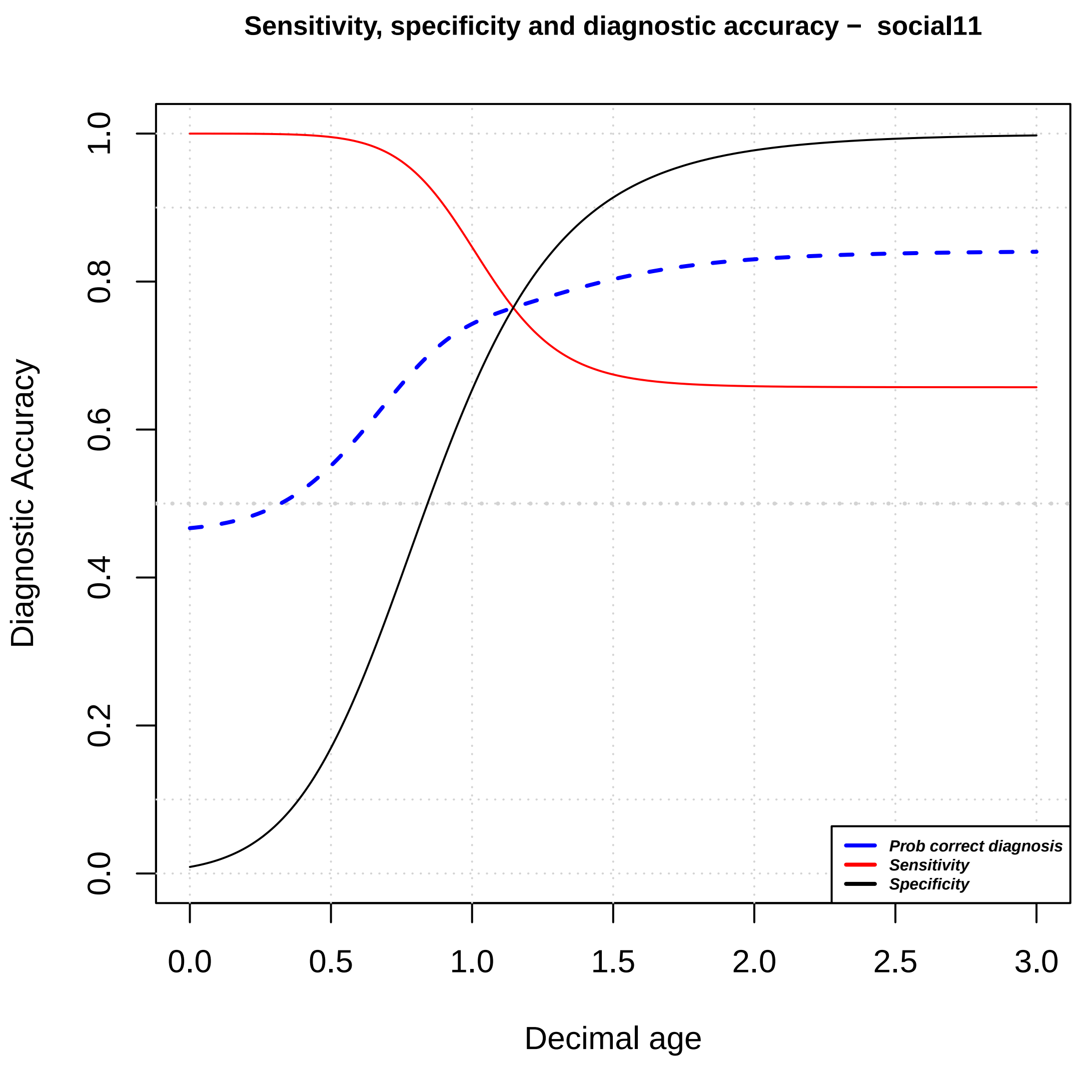
Sensitivity, specificity and diagnostic accuracy − social11
Diagnostic Accuracy
Decimal age
0.0
0.2
0.4
0.6
0.8
1.0
0.0
0.5
1.0
1.5
2.0
2.5
3.0
Prob correct diagnosis
Sensitivity
Specificity

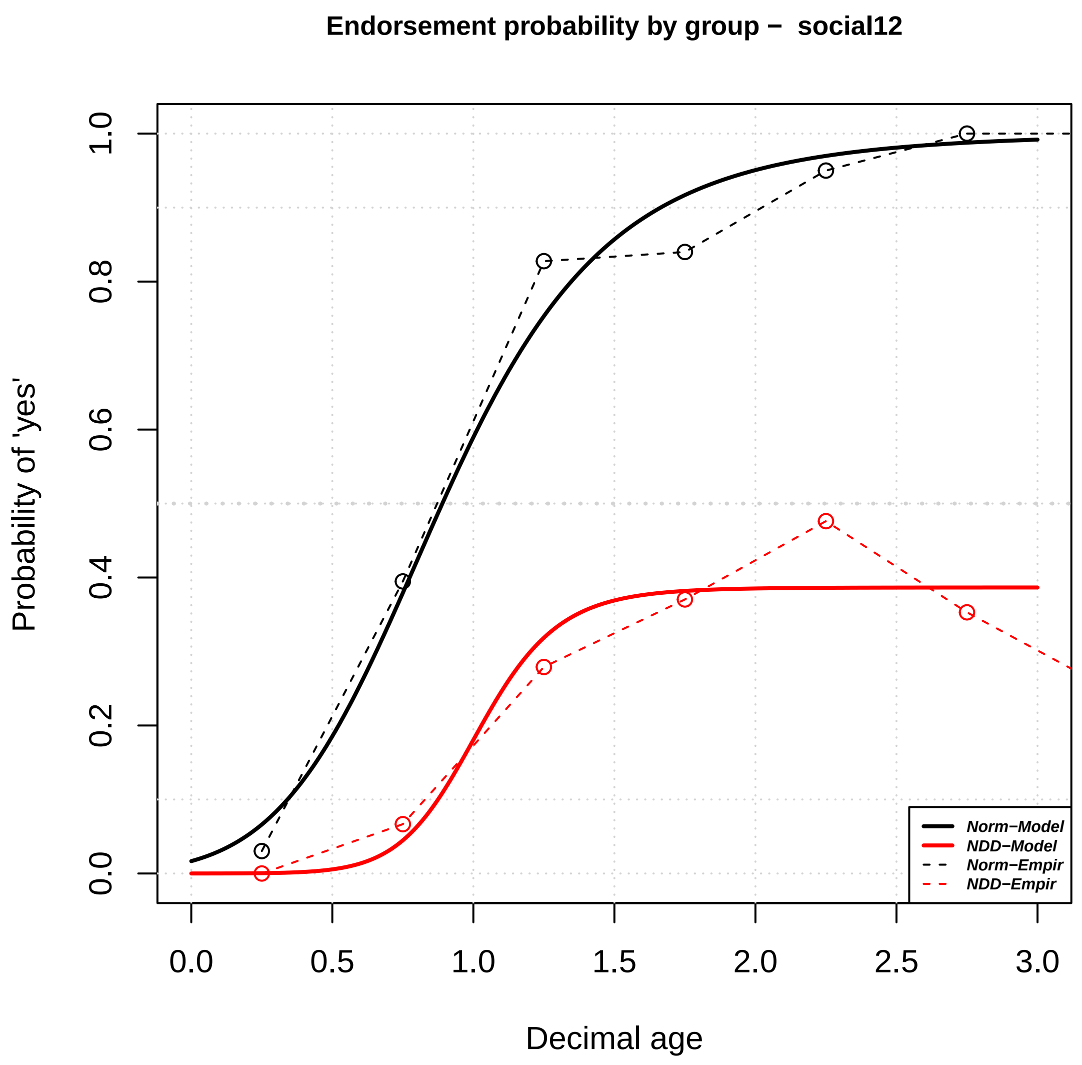

Endorsement probability by group – social12
Probability of 'yes'
Decimal age
Norm−Model
NDD−Model
Norm−Empir
NDD−Empir
0.0
0.2
0.4
0.6
0.8
1.0
0.0
0.5
1.0
1.5
2.0
2.5
3.0


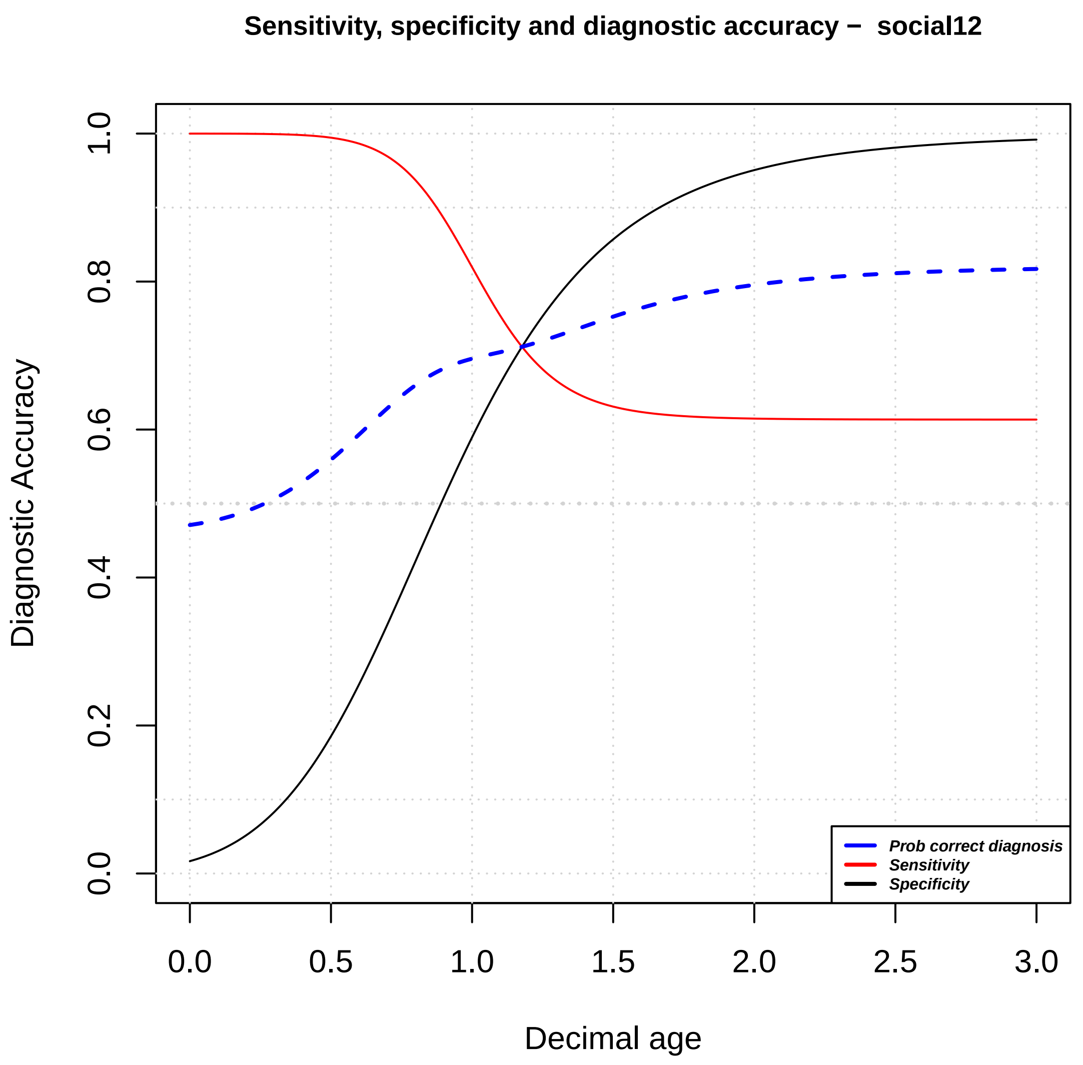

Sensitivity, specificity and diagnostic accuracy – social12
Diagnostic Accuracy
Decimal age
Prob correct diagnosis
Sensitivity
Specificity
0.0
0.2
0.4
0.6
0.8
1.0
0.0
0.5
1.0
1.5
2.0
2.5
3.0

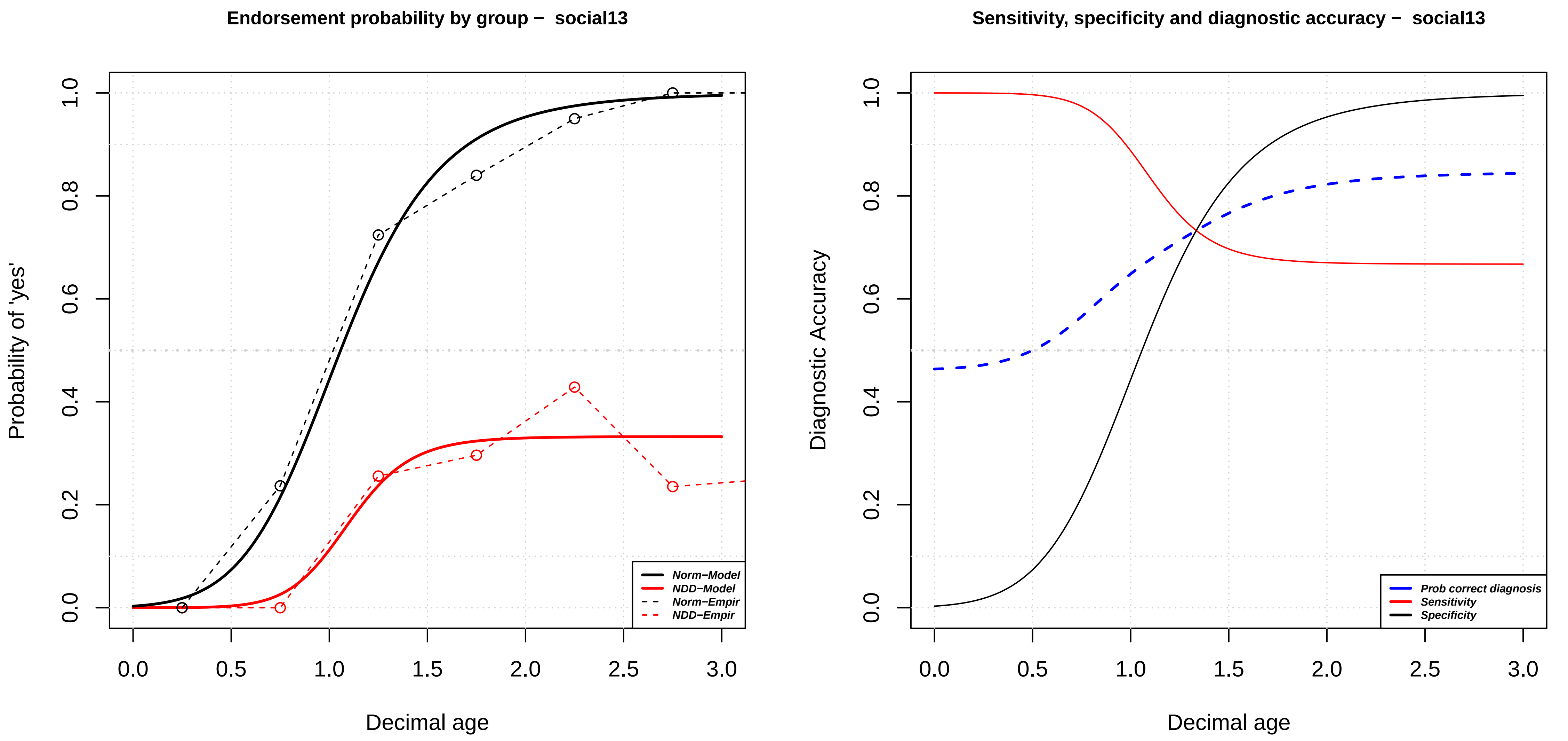

Endorsement probability by group – social13
Probability of 'yes'
Decimal age
0.0
0.2
0.4
0.6
0.8
1.0
0.0
0.5
1.0
1.5
2.0
2.5
3.0
Norm−Model
NDD−Model
Norm−Empir
NDD−Empir
Sensitivity, specificity and diagnostic accuracy – social13
Diagnostic Accuracy
Decimal age
Prob correct diagnosis
Sensitivity
Specificity

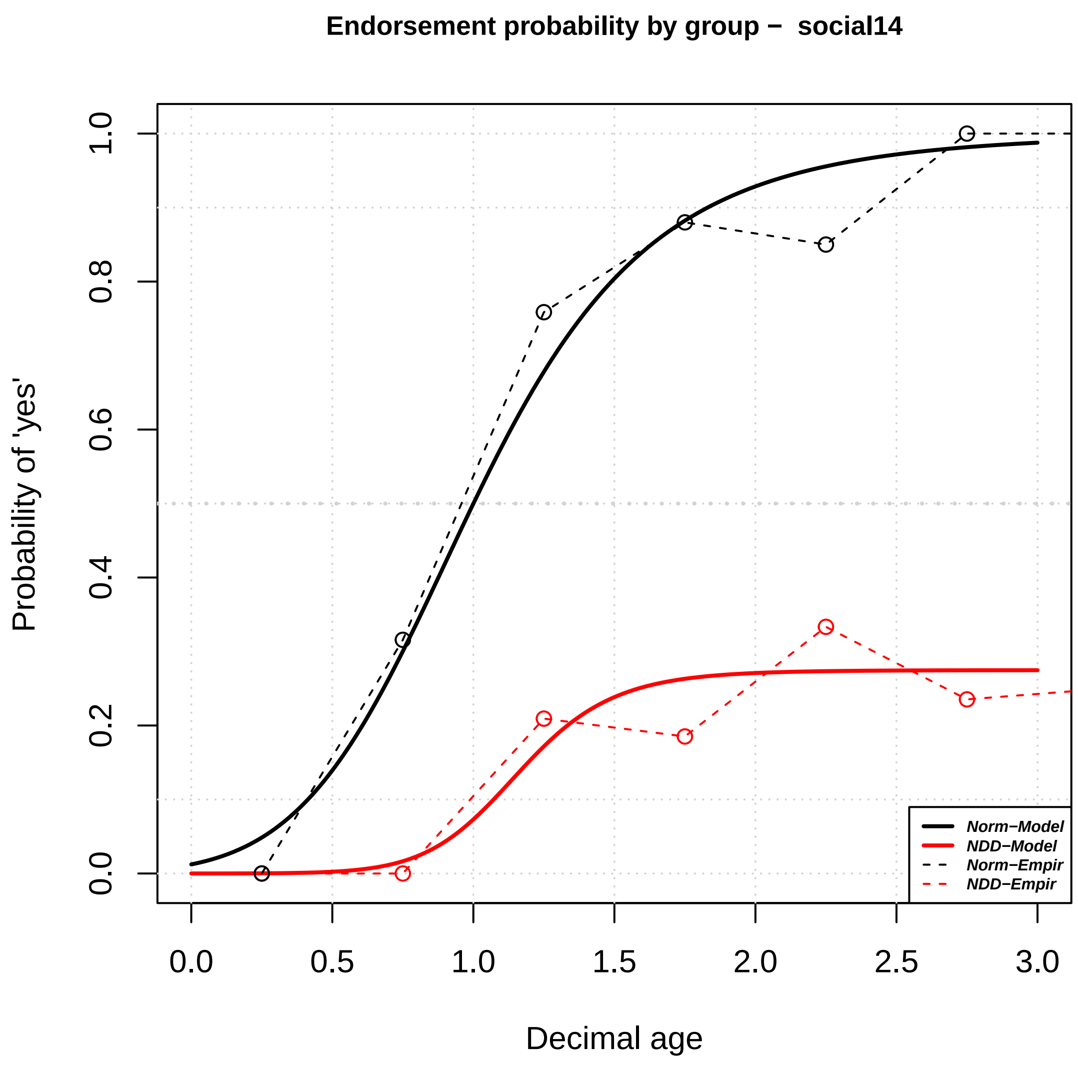

Endorsement probability by group − social14
Probability of 'yes'
Decimal age
0.0
0.2
0.4
0.6
0.8
1.0
0.0
0.5
1.0
1.5
2.0
2.5
3.0
Norm−Model
NDD−Model
Norm−Empir
NDD−Empir


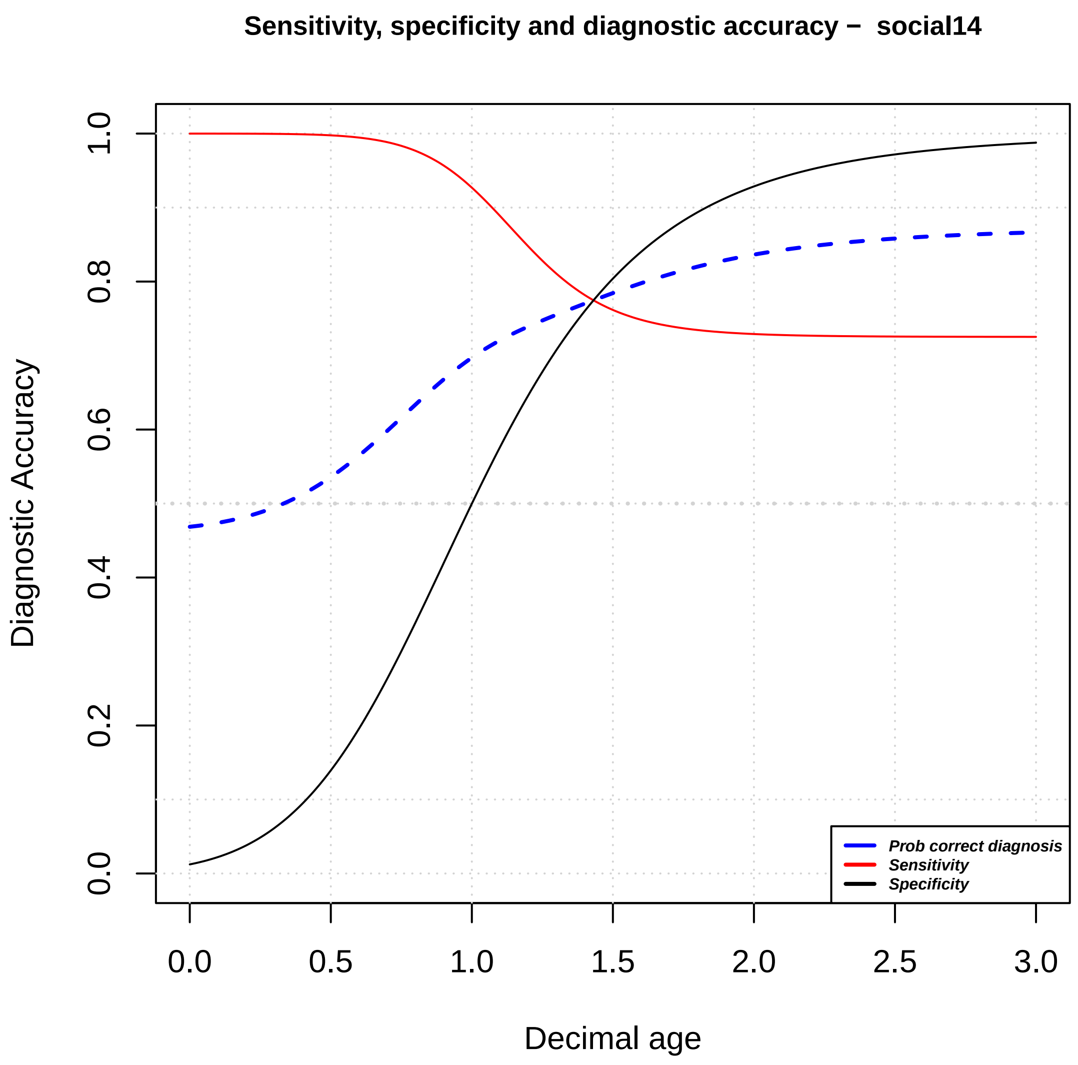

Sensitivity, specificity and diagnostic accuracy − social14
Diagnostic Accuracy
Decimal age
0.0
0.2
0.4
0.6
0.8
1.0
0.0
0.5
1.0
1.5
2.0
2.5
3.0
Prob correct diagnosis
Sensitivity
Specificity

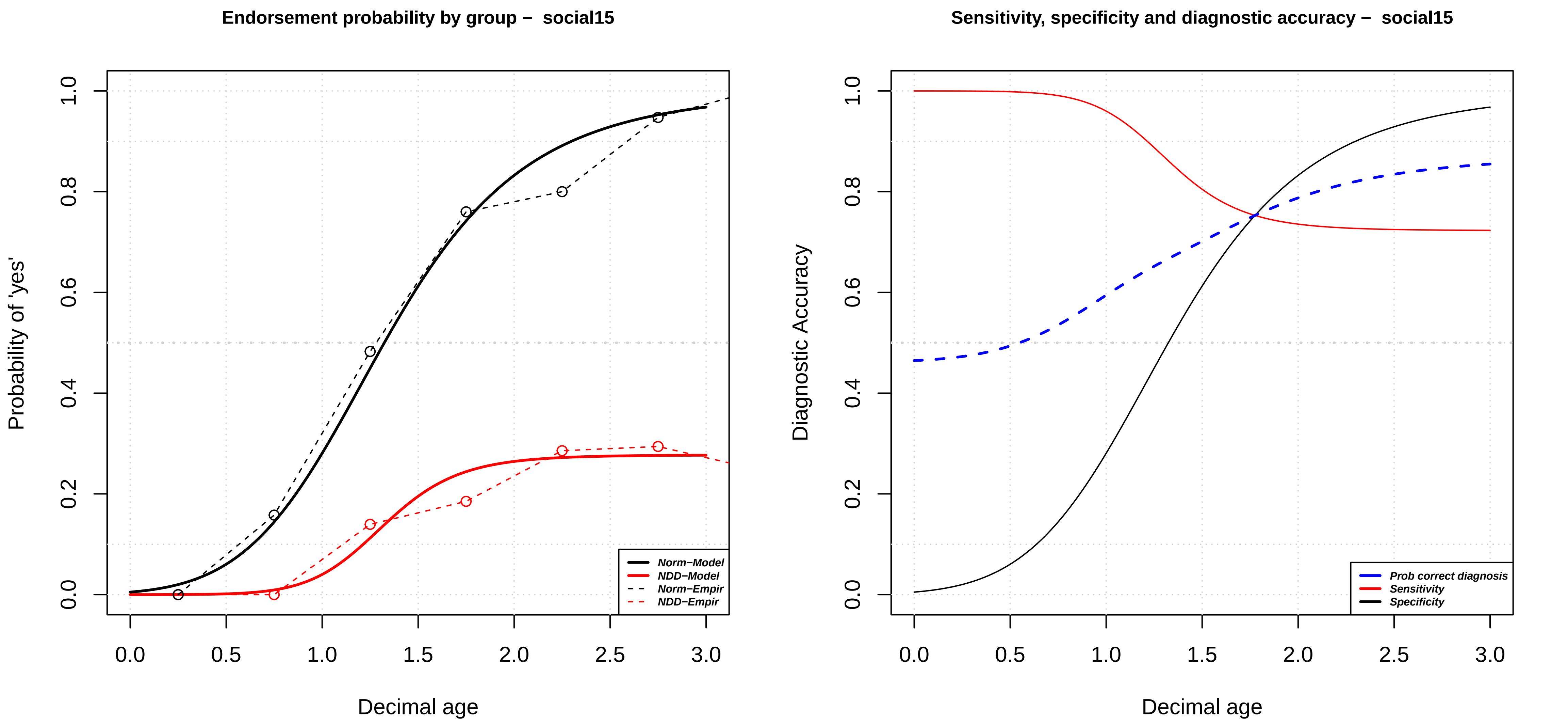
Endorsement probability by group – social15
Probability of 'yes'
Decimal age
Norm−Model
NDD−Model
Norm−Empir
NDD−Empir
Sensitivity, specificity and diagnostic accuracy – social15
Diagnostic Accuracy
Decimal age
Prob correct diagnosis
Sensitivity
Specificity
0.0
0.2
0.4
0.6
0.8
1.0
0.0
0.5
1.0
1.5
2.0
2.5
3.0

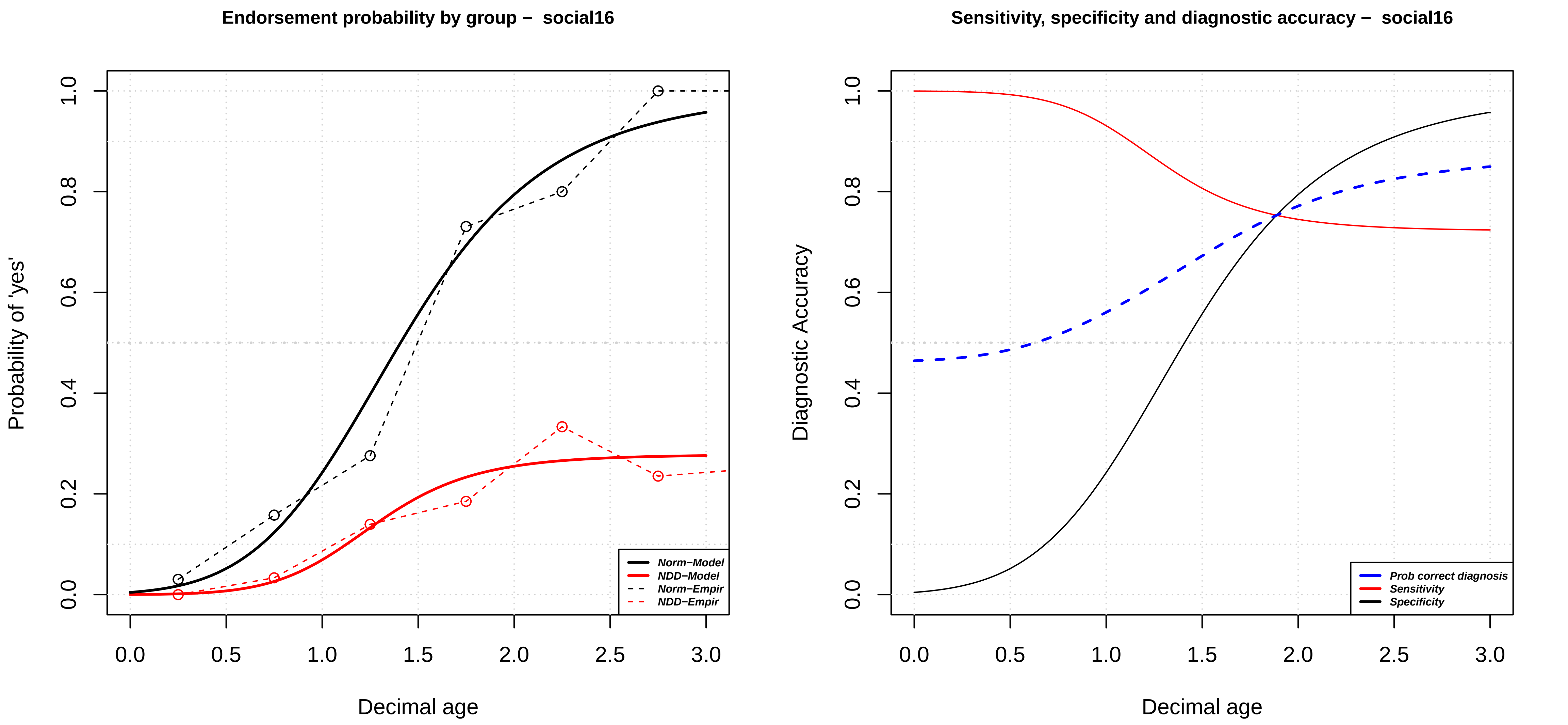
Endorsement probability by group – social16
Probability of 'yes'
Decimal age
0.0
0.2
0.4
0.6
0.8
1.0
0.0
0.5
1.0
1.5
2.0
2.5
3.0
Norm–Model
NDD–Model
Norm–Empir
NDD–Empir
Sensitivity, specificity and diagnostic accuracy – social16
Diagnostic Accuracy
Decimal age
Prob correct diagnosis
Sensitivity
Specificity

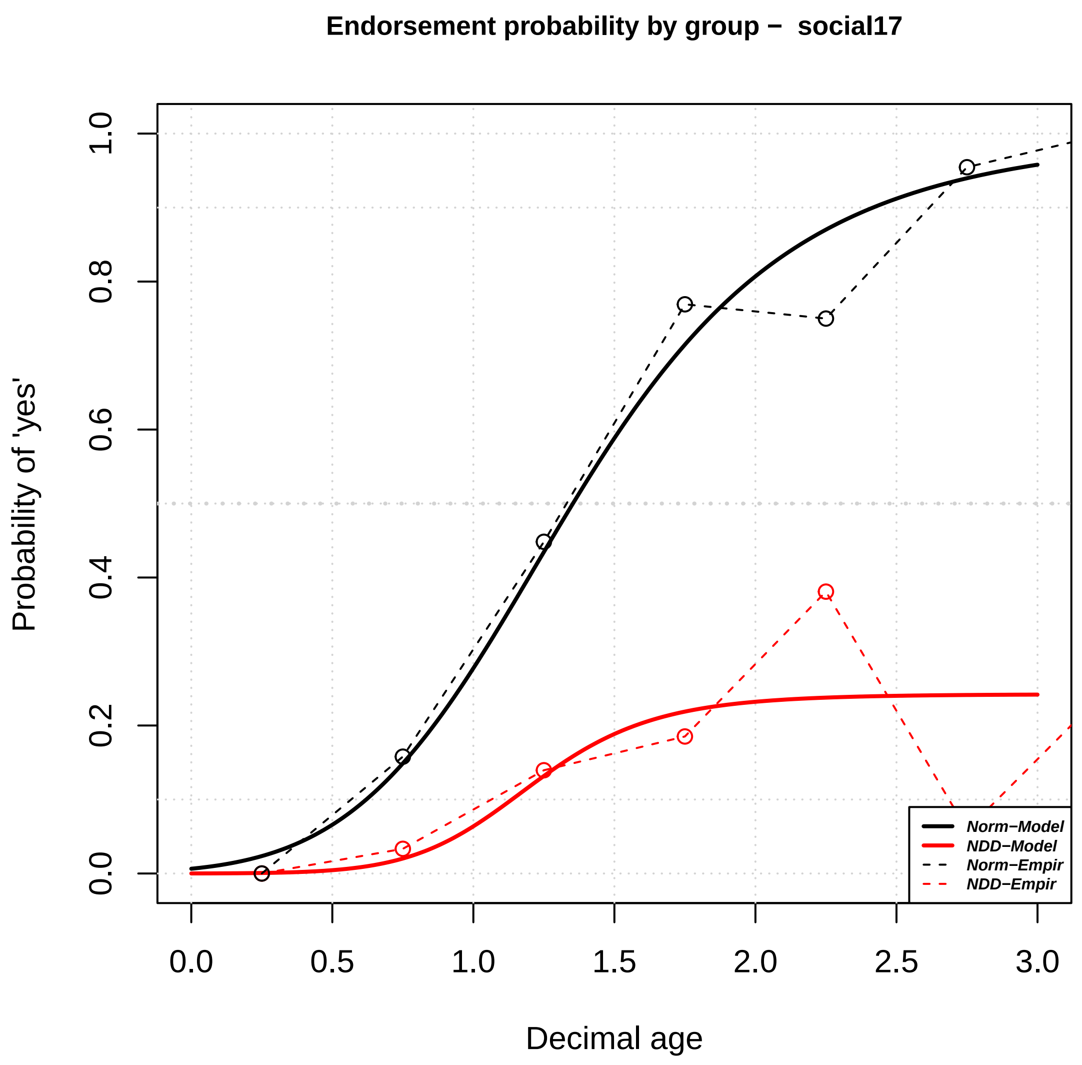
Endorsement probability by group − social17
Probability of 'yes'
Decimal age
0.0
0.2
0.4
0.6
0.8
1.0
0.0
0.5
1.0
1.5
2.0
2.5
3.0
Norm−Model
NDD−Model
Norm−Empir
NDD−Empir

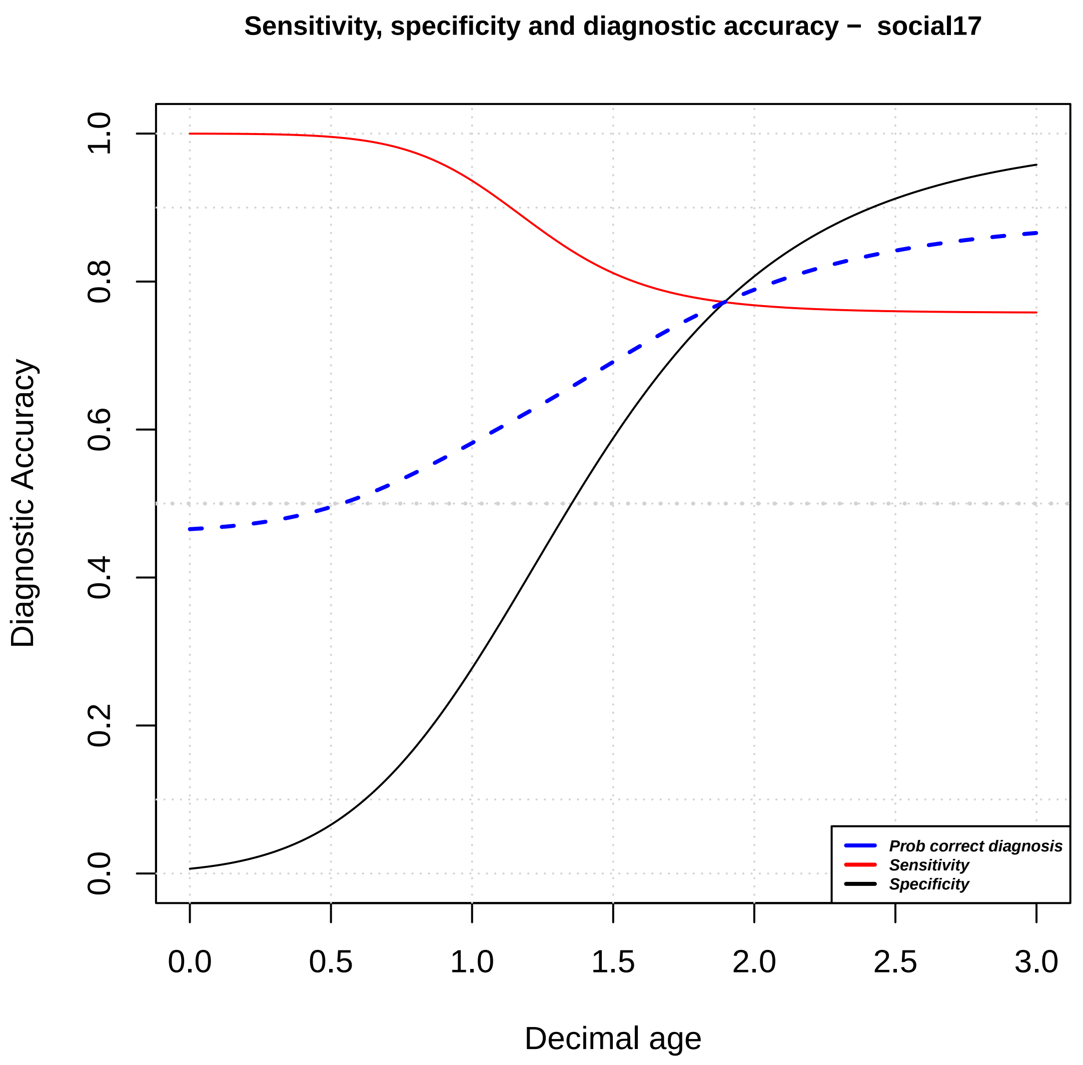
Sensitivity, specificity and diagnostic accuracy − social17
Diagnostic Accuracy
Decimal age
0.0
0.2
0.4
0.6
0.8
1.0
0.0
0.5
1.0
1.5
2.0
2.5
3.0
Prob correct diagnosis
Sensitivity
Specificity

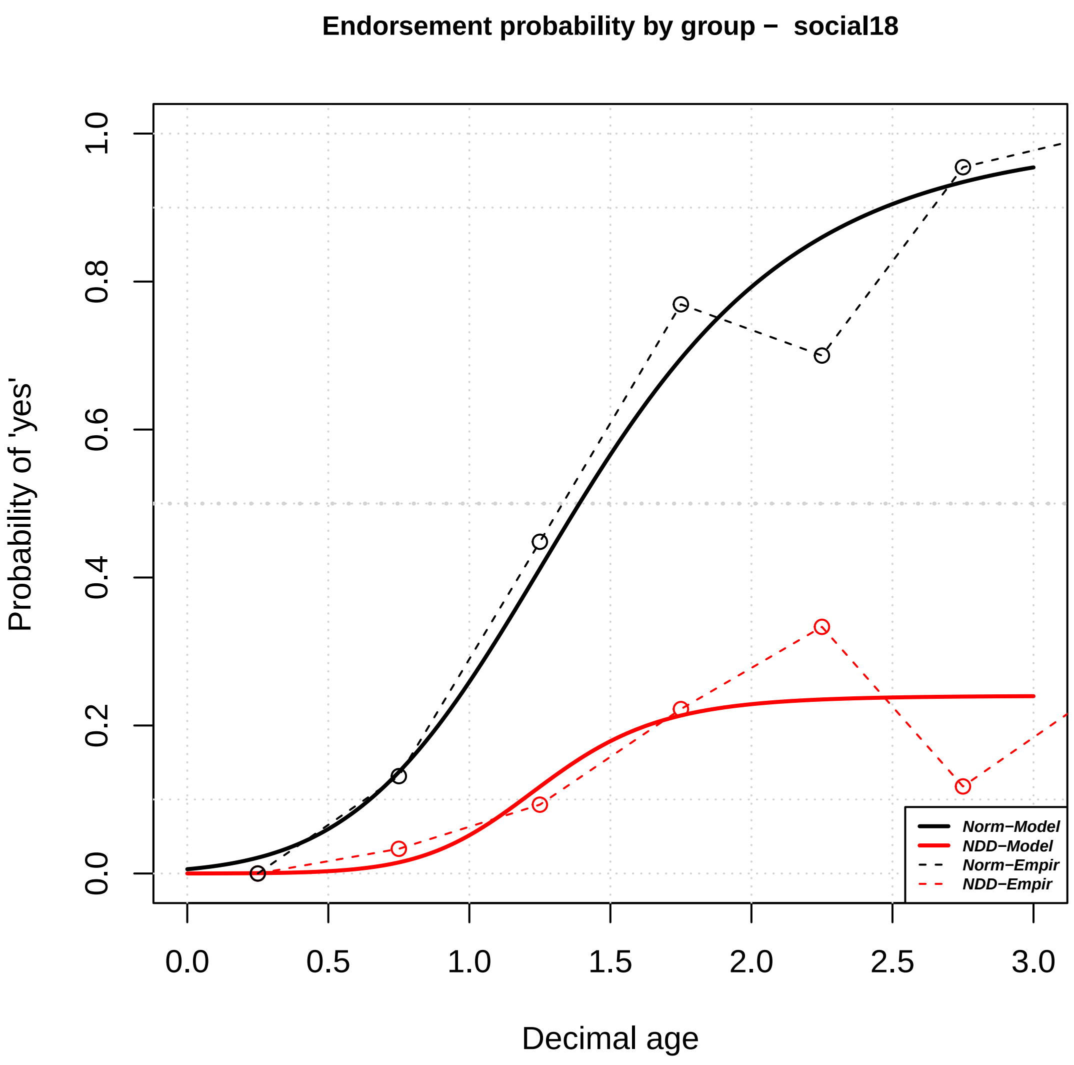

Endorsement probability by group – social18
Probability of 'yes'
Decimal age
0.0
0.2
0.4
0.6
0.8
1.0
0.0
0.5
1.0
1.5
2.0
2.5
3.0
Norm−Model
NDD−Model
Norm−Empir
NDD−Empir


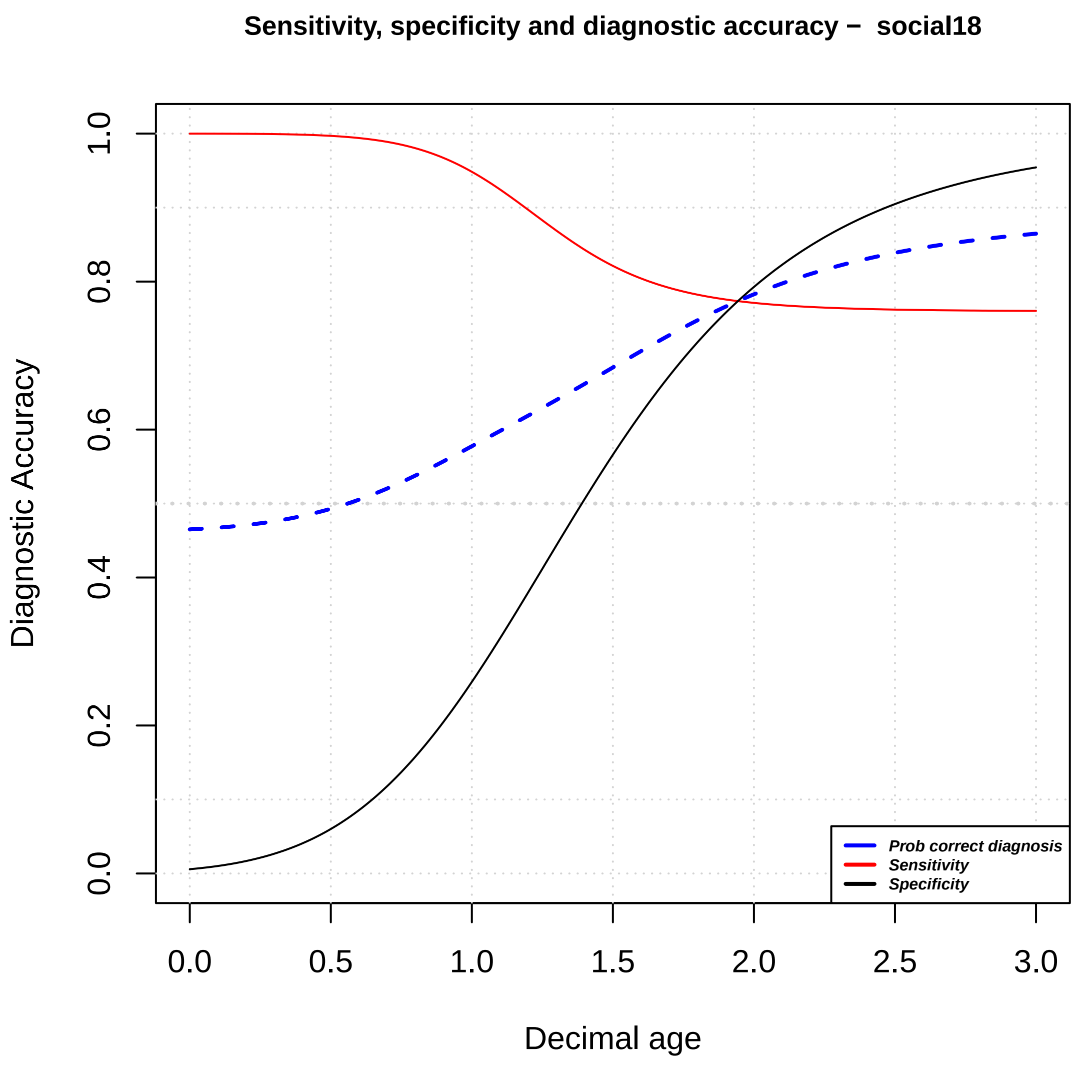

Sensitivity, specificity and diagnostic accuracy – social18
Diagnostic Accuracy
Decimal age
0.0
0.2
0.4
0.6
0.8
1.0
0.0
0.5
1.0
1.5
2.0
2.5
3.0
Prob correct diagnosis
Sensitivity
Specificity

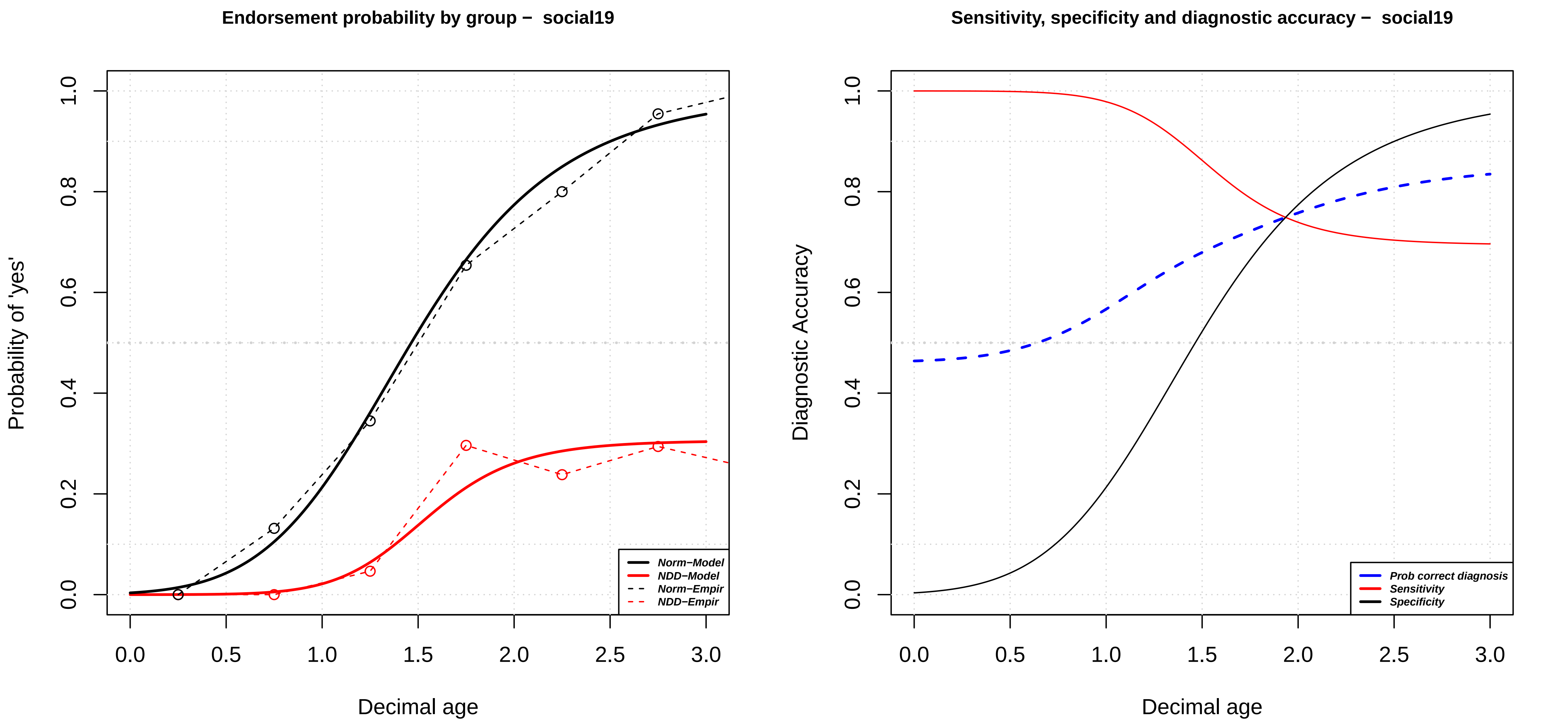

Endorsement probability by group − social19
Probability of 'yes'
Decimal age
0.0
0.2
0.4
0.6
0.8
1.0
0.5
1.5
2.0
2.5
3.0
Norm−Model
NDD−Model
Norm−Empir
NDD−Empir
Sensitivity, specificity and diagnostic accuracy − social19
Diagnostic Accuracy
Prob correct diagnosis
Sensitivity
Specificity

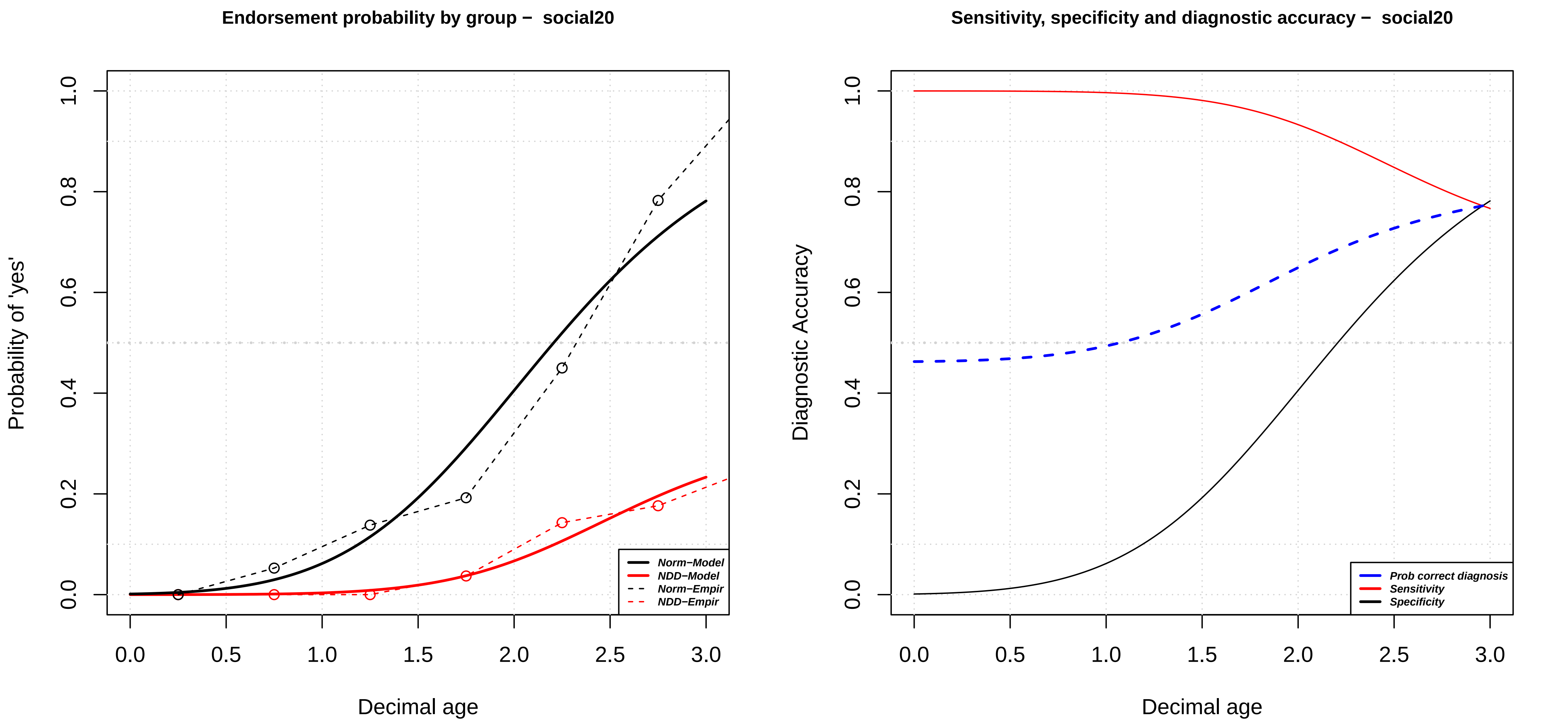

Endorsement probability by group − social20
Probability of 'yes'
Decimal age
Norm−Model
NDD−Model
Norm−Empir
NDD−Empir
0.0
0.2
0.4
0.6
0.8
1.0
0.0
0.5
1.0
1.5
2.0
2.5
3.0
Sensitivity, specificity and diagnostic accuracy − social20
Diagnostic Accuracy
Decimal age
Prob correct diagnosis
Sensitivity
Specificity

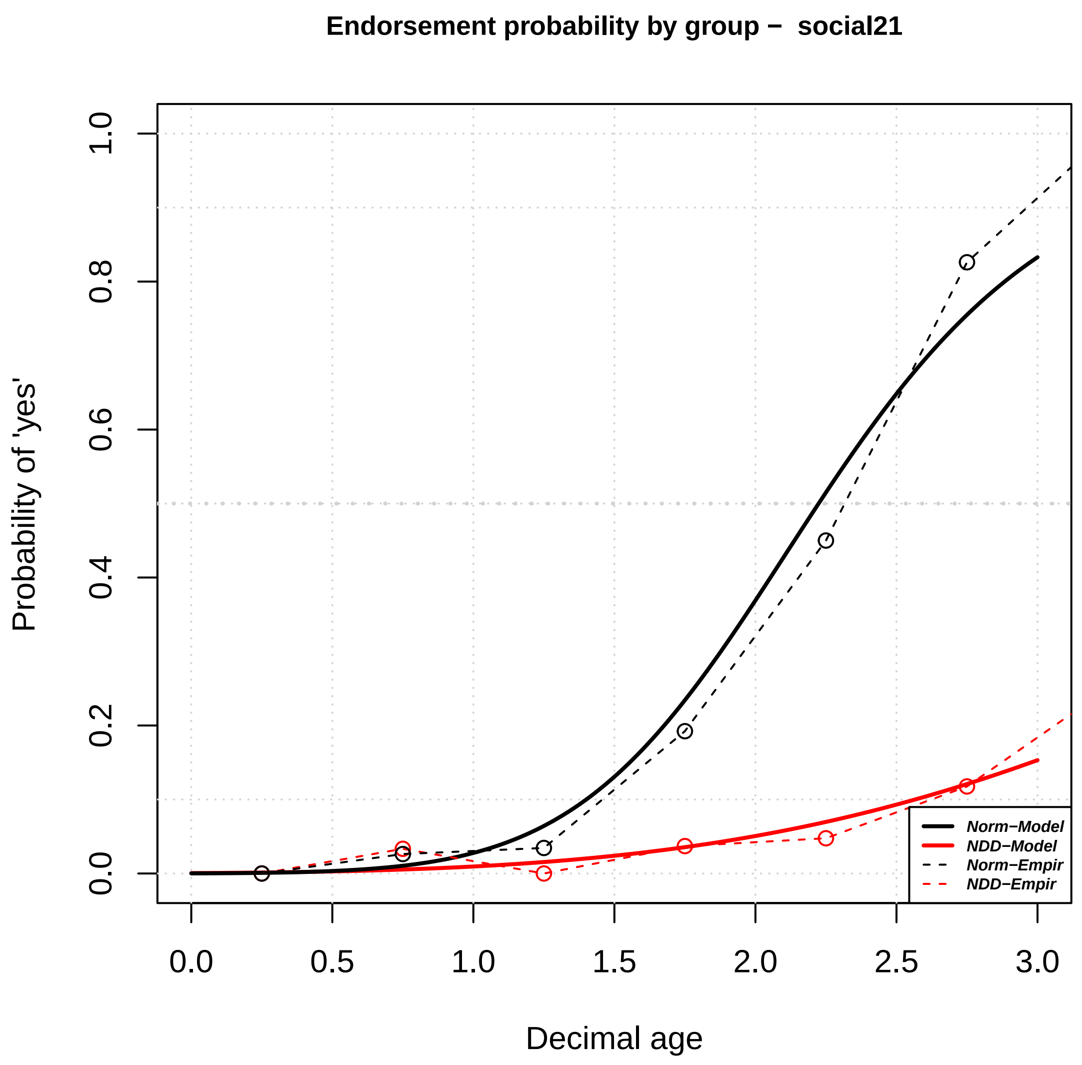

Endorsement probability by group − social21
Probability of 'yes'
Decimal age
0.0
0.2
0.4
0.6
0.8
1.0
0.0
0.5
1.0
1.5
2.0
2.5
3.0
Norm−Model
NDD−Model
Norm−Empir
NDD−Empir


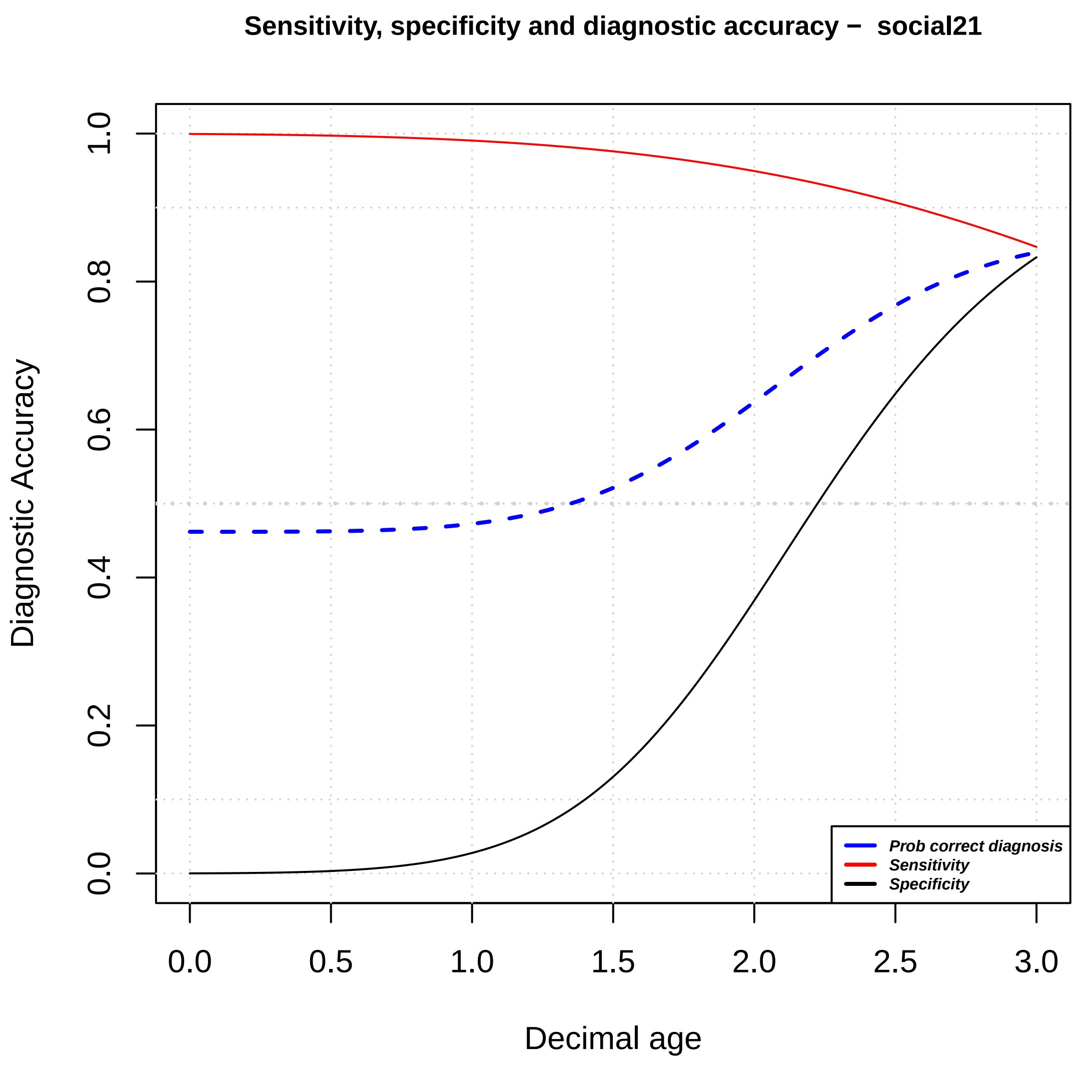

Sensitivity, specificity and diagnostic accuracy − social21
Diagnostic Accuracy
Decimal age
0.0
0.2
0.4
0.6
0.8
1.0
0.0
0.5
1.0
1.5
2.0
2.5
3.0
Prob correct diagnosis
Sensitivity
Specificity

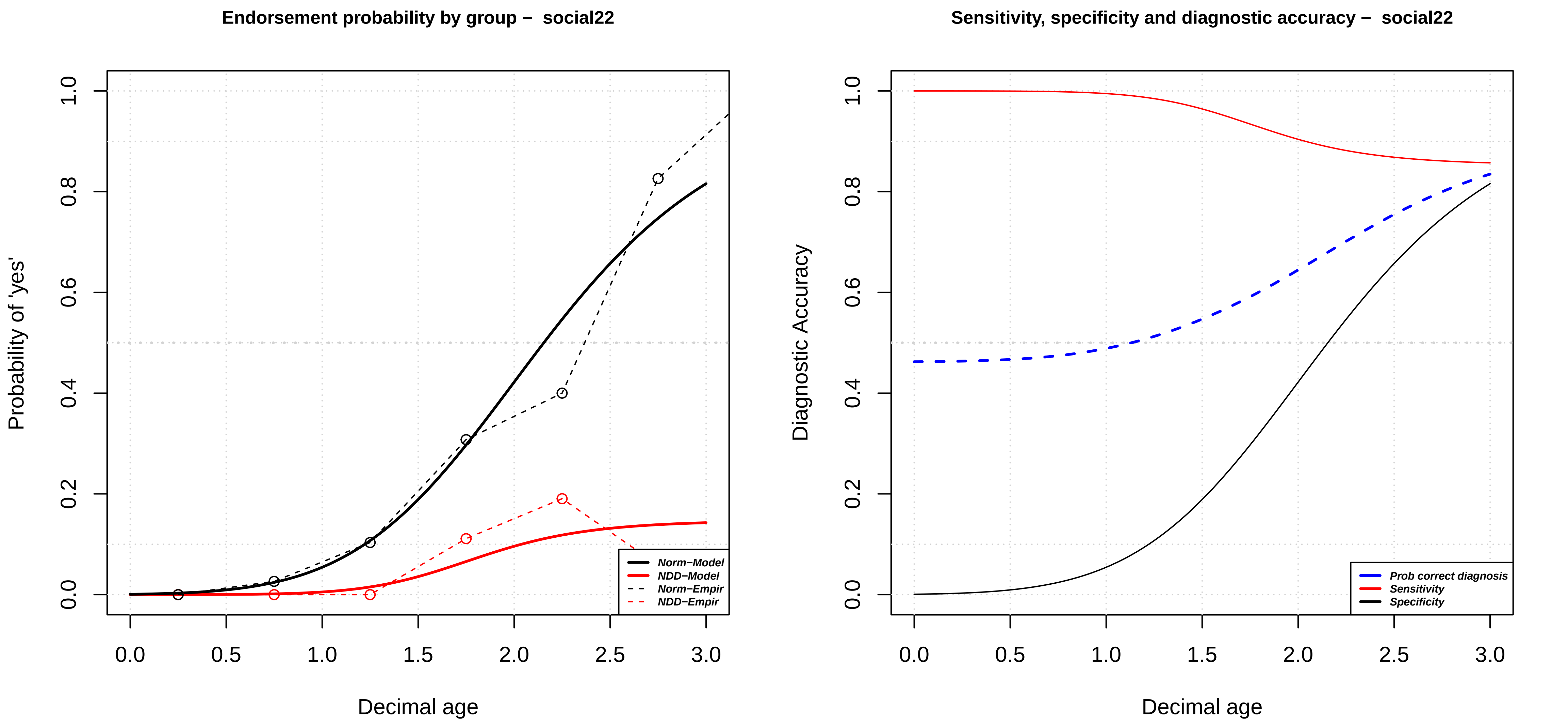

Endorsement probability by group − social22
Probability of 'yes'
Decimal age
Norm−Model
NDD−Model
Norm−Empir
NDD−Empir
0.0
0.2
0.4
0.6
0.8
1.0
0.0
0.5
1.0
1.5
2.0
2.5
3.0
Sensitivity, specificity and diagnostic accuracy − social22
Diagnostic Accuracy
Decimal age
Prob correct diagnosis
Sensitivity
Specificity

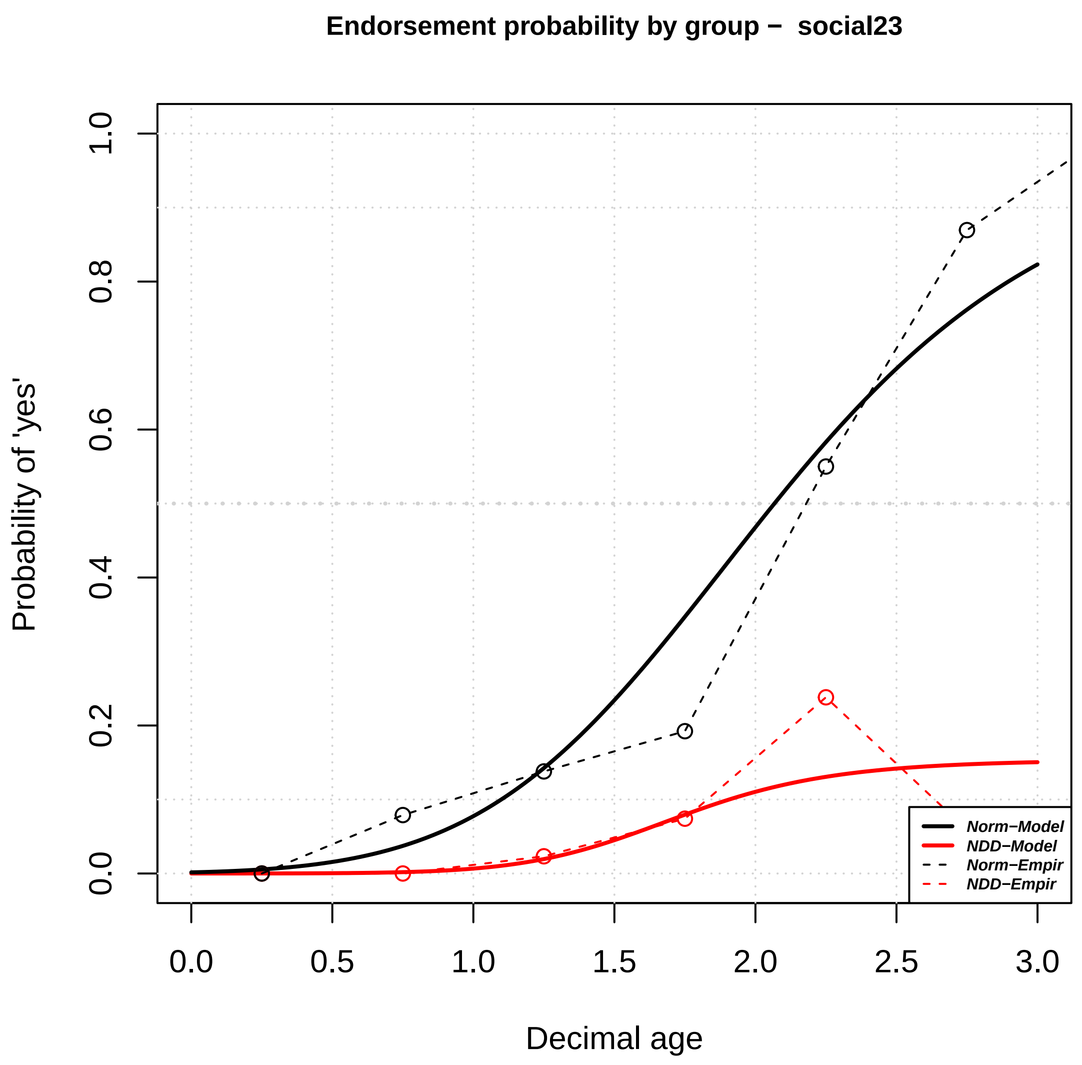

Endorsement probability by group – social23
Probability of 'yes'
Decimal age
0.0
0.2
0.4
0.6
0.8
1.0
0.0
0.5
1.0
1.5
2.0
2.5
3.0
Norm–Model
NDD–Model
Norm–Empir
NDD–Empir


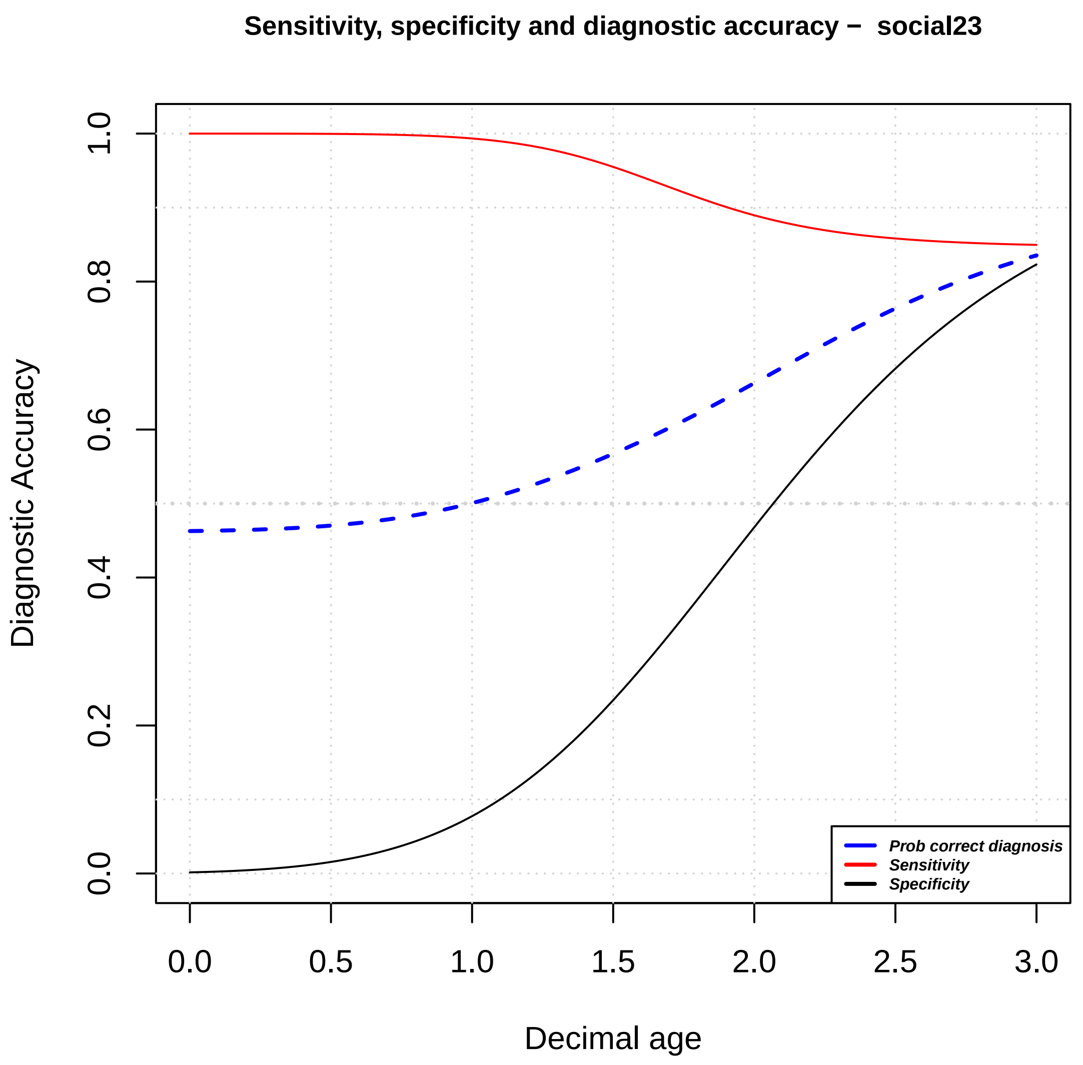

Sensitivity, specificity and diagnostic accuracy – social23
Diagnostic Accuracy
Decimal age
0.0
0.2
0.4
0.6
0.8
1.0
0.0
0.5
1.0
1.5
2.0
2.5
3.0
Prob correct diagnosis
Sensitivity
Specificity

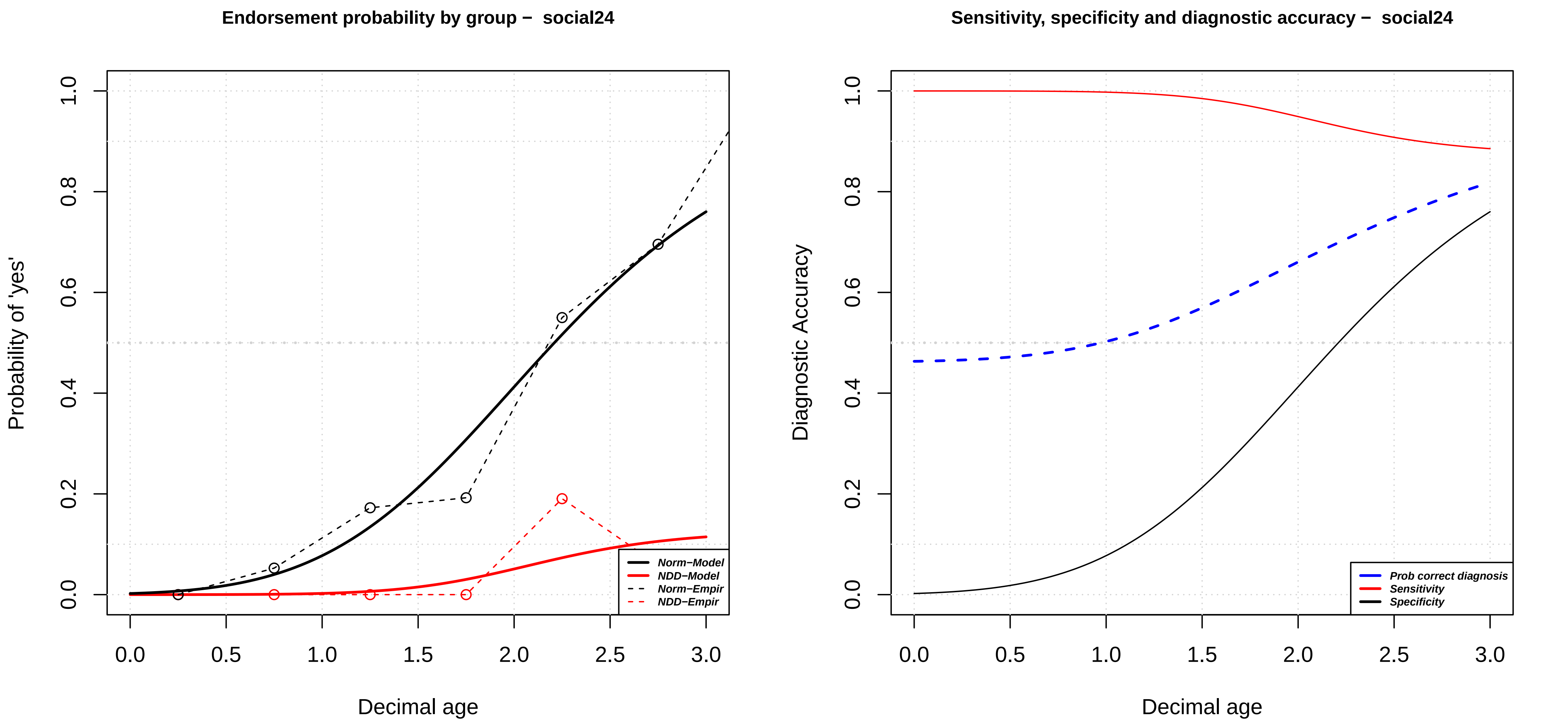

Endorsement probability by group − social24
Probability of 'yes'
Decimal age
0.0
0.2
0.4
0.6
0.8
1.0
0.0
0.5
1.0
1.5
2.0
2.5
3.0
Norm−Model
NDD−Model
Norm−Empir
NDD−Empir
Sensitivity, specificity and diagnostic accuracy − social24
Diagnostic Accuracy
Decimal age
Prob correct diagnosis
Sensitivity
Specificity

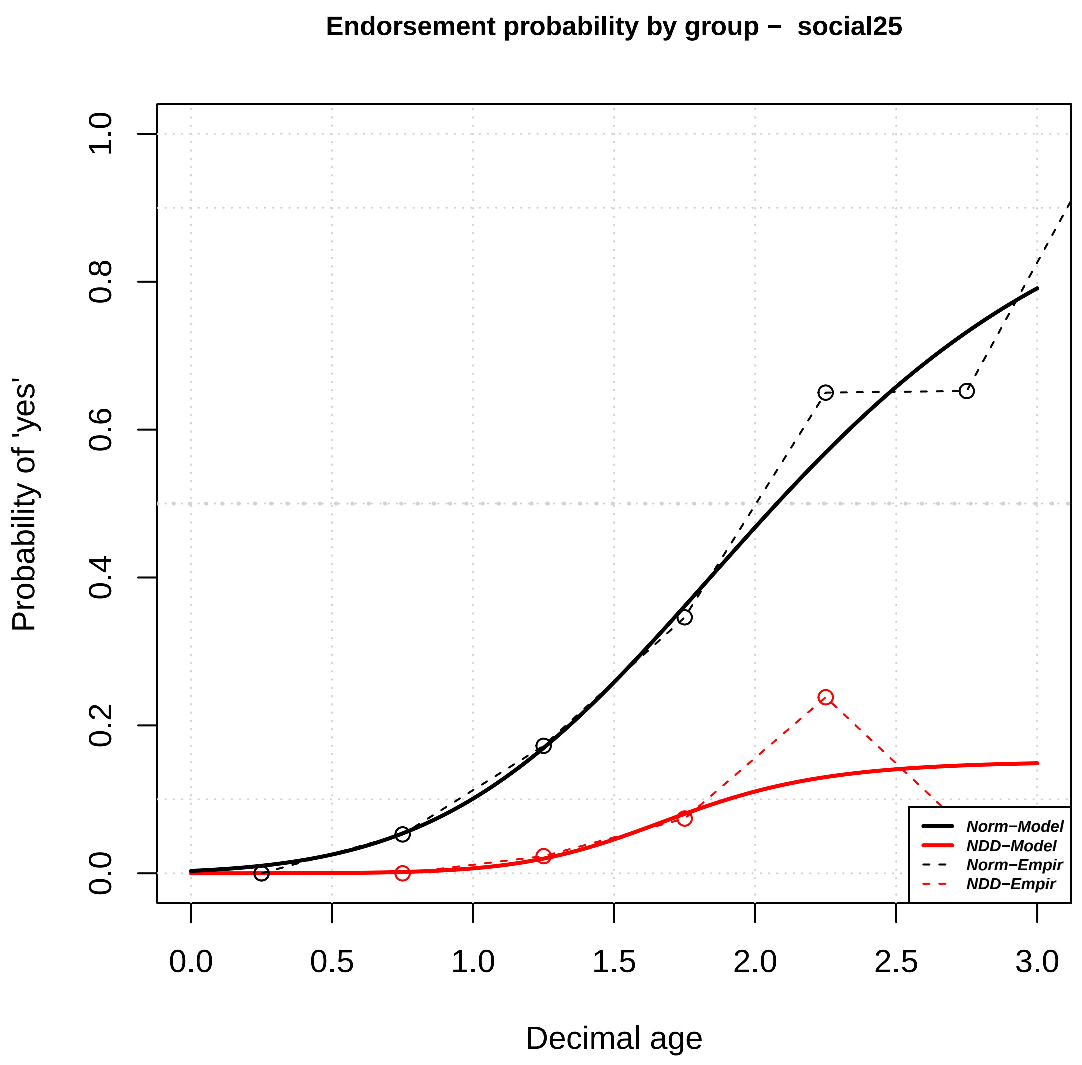

Endorsement probability by group − social25
Probability of 'yes'
Decimal age
0.0
0.2
0.4
0.6
0.8
1.0
0.0
0.5
1.0
1.5
2.0
2.5
3.0
Norm−Model
NDD−Model
Norm−Empir
NDD−Empir


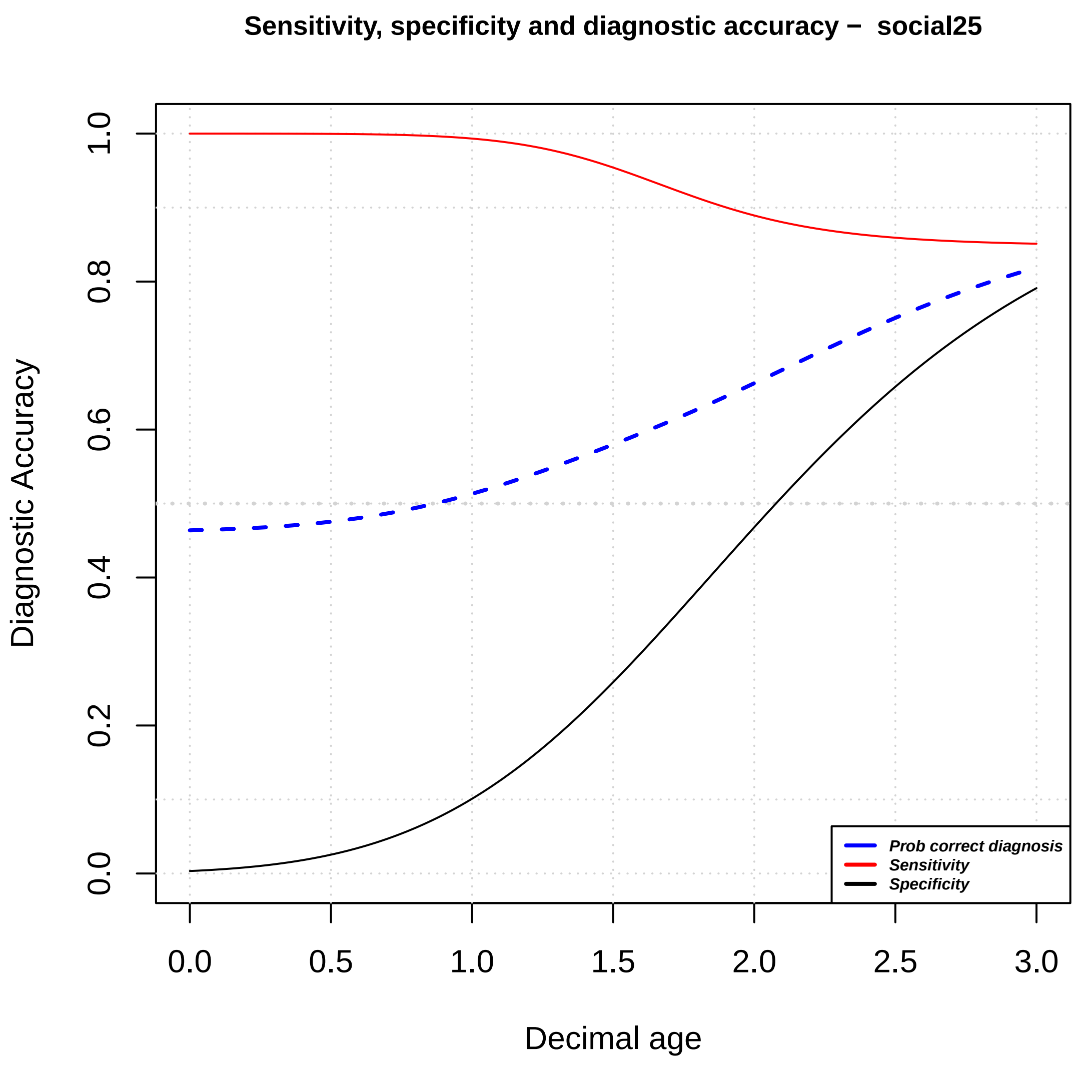

Sensitivity, specificity and diagnostic accuracy − social25
Diagnostic Accuracy
Decimal age
0.0
0.2
0.4
0.6
0.8
1.0
0.0
0.5
1.0
1.5
2.0
2.5
3.0
Prob correct diagnosis
Sensitivity
Specificity

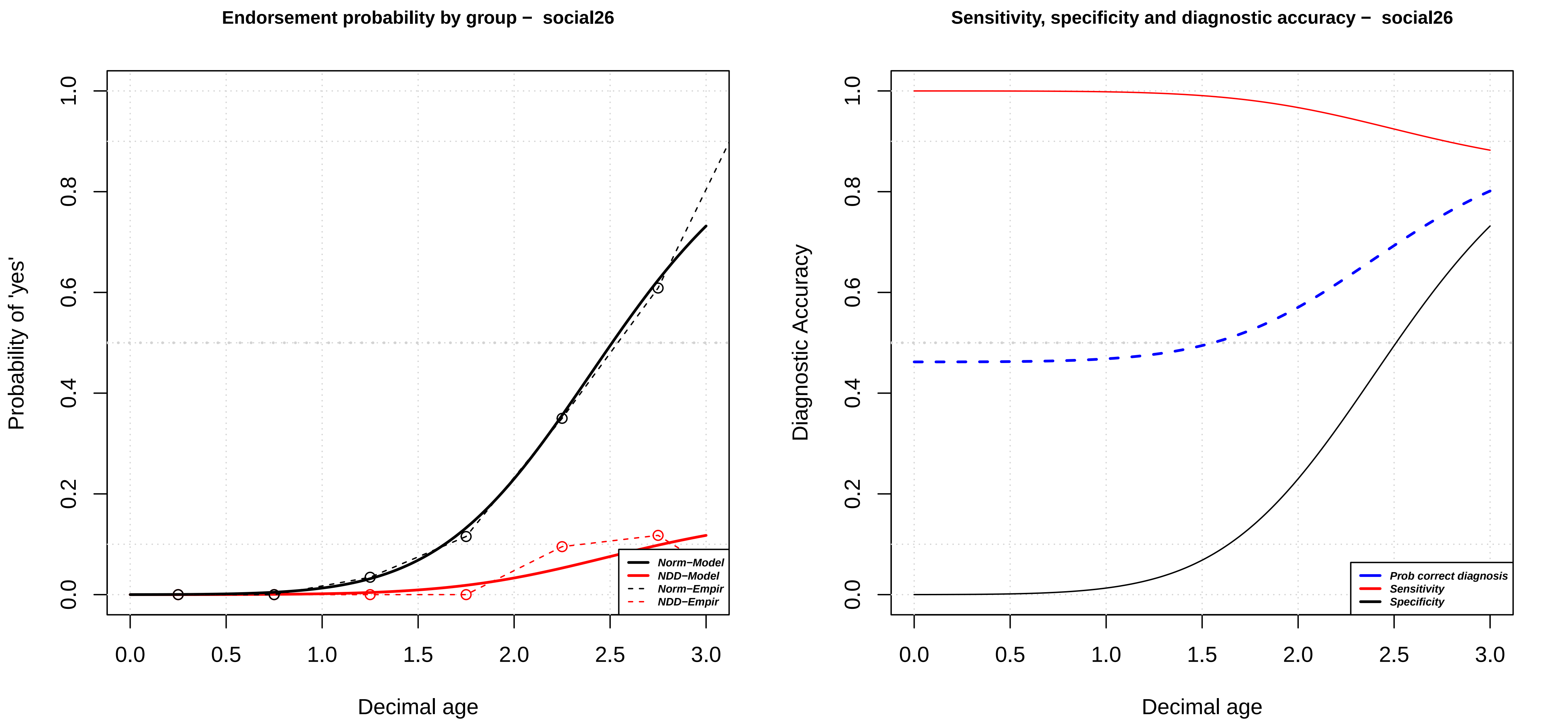

Endorsement probability by group − social26
Probability of 'yes'
Decimal age
Norm−Model
NDD−Model
Norm−Empir
NDD−Empir
Sensitivity, specificity and diagnostic accuracy − social26
Diagnostic Accuracy
Decimal age
Prob correct diagnosis
Sensitivity
Specificity

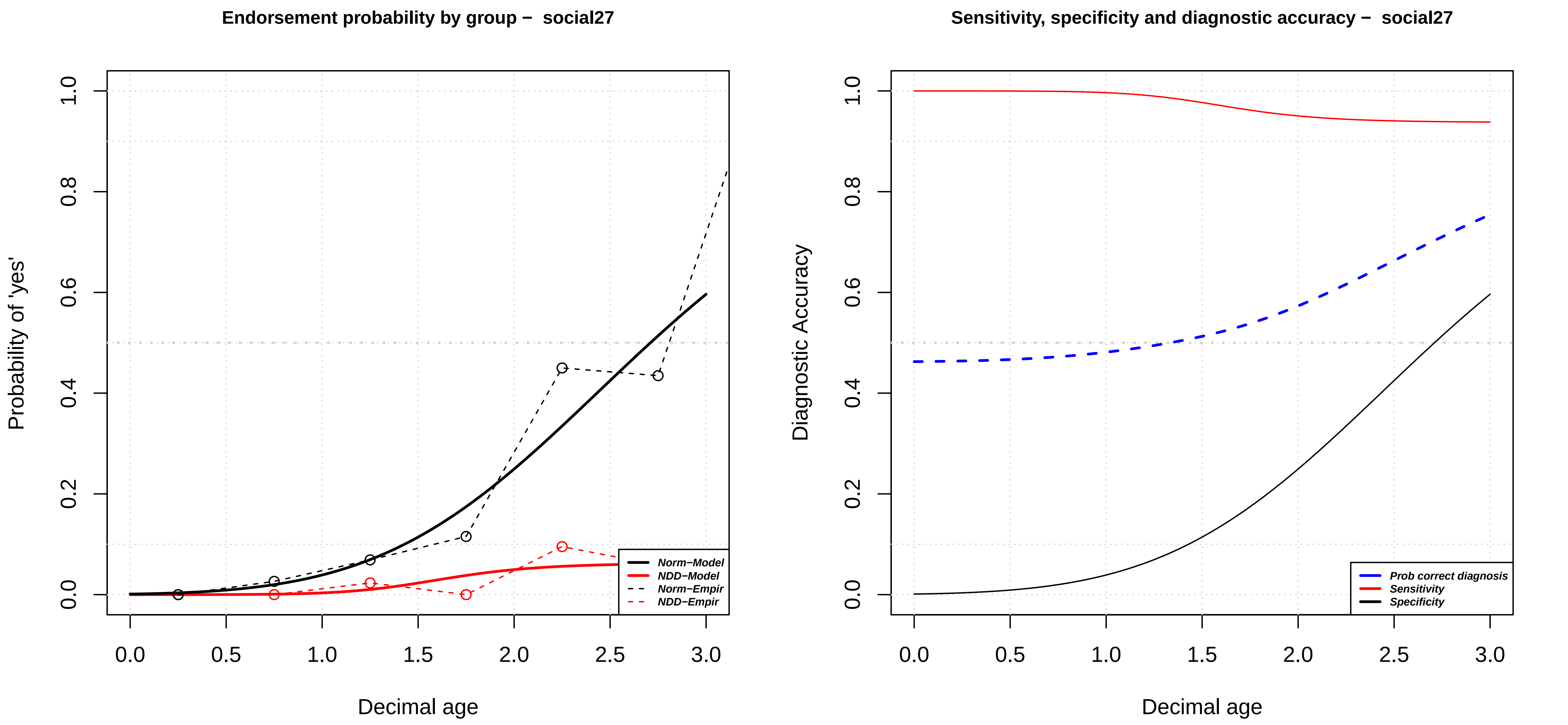
Endorsement probability by group – social27
Probability of 'yes'
Decimal age
Norm−Model
NDD−Model
Norm−Empir
NDD−Empir
Sensitivity, specificity and diagnostic accuracy – social27
Diagnostic Accuracy
Decimal age
Prob correct diagnosis
Sensitivity
Specificity

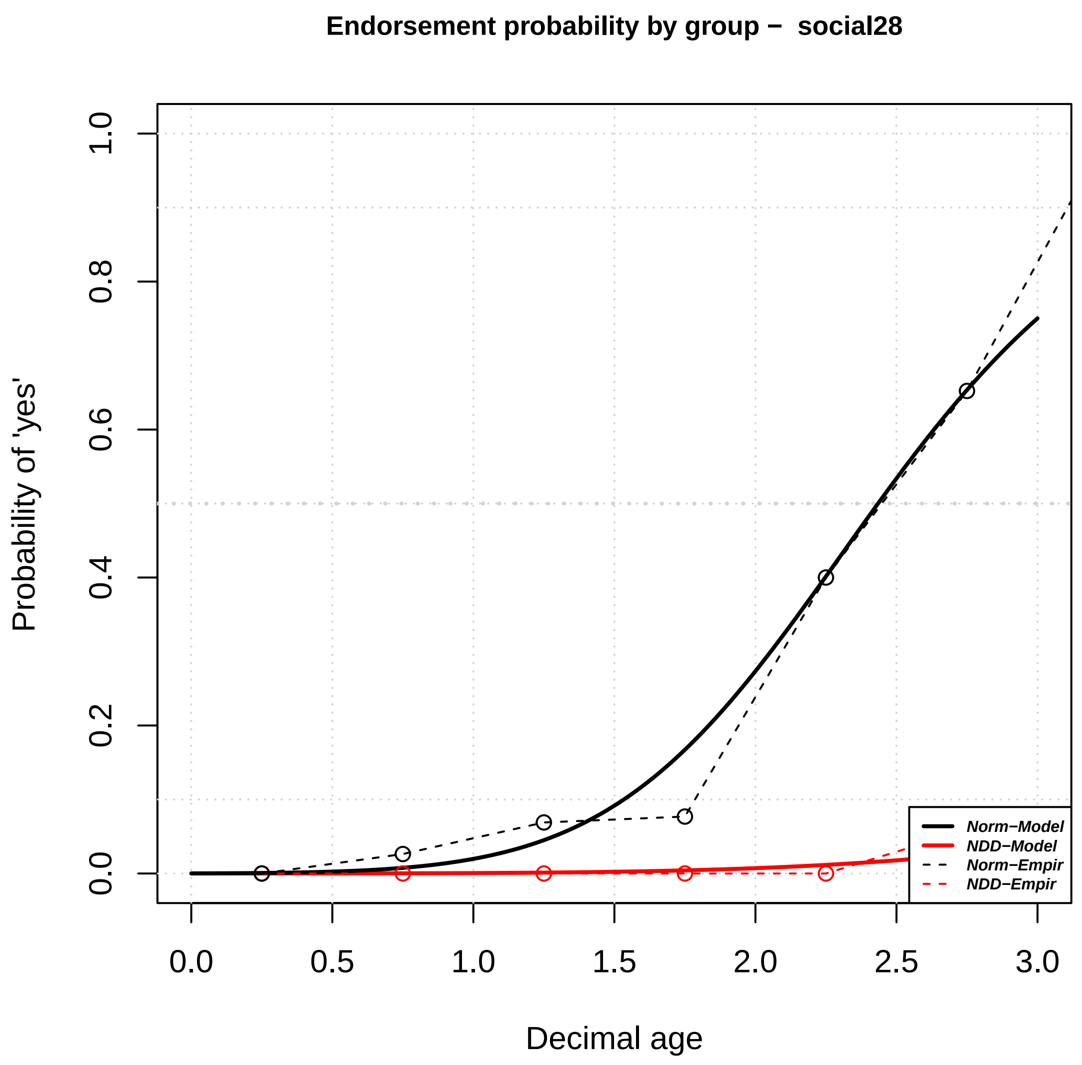

Endorsement probability by group – social28
Probability of 'yes'
Decimal age
Norm–Model
NDD–Model
Norm–Empir
NDD–Empir


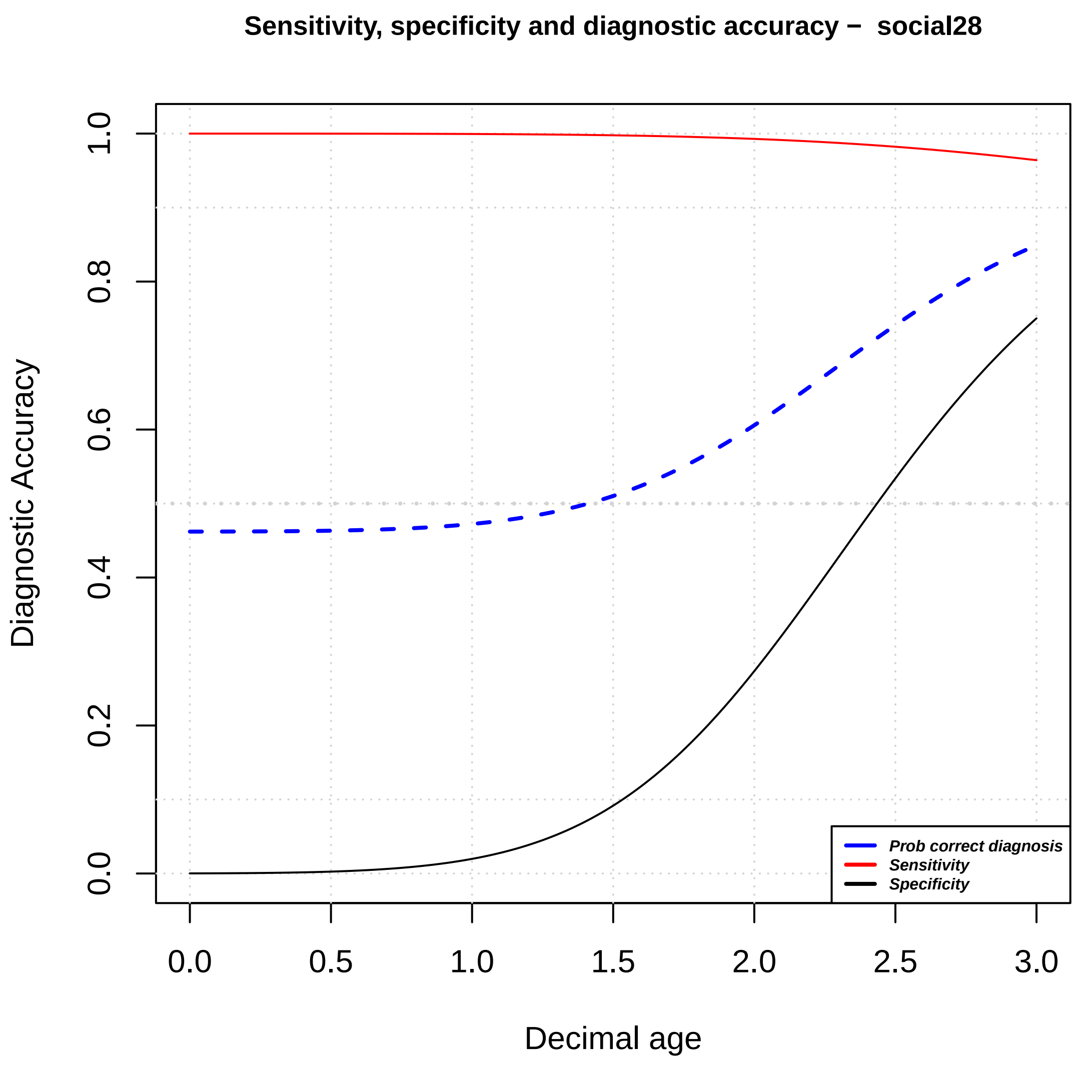

Sensitivity, specificity and diagnostic accuracy – social28
Diagnostic Accuracy
Decimal age
Prob correct diagnosis
Sensitivity
Specificity

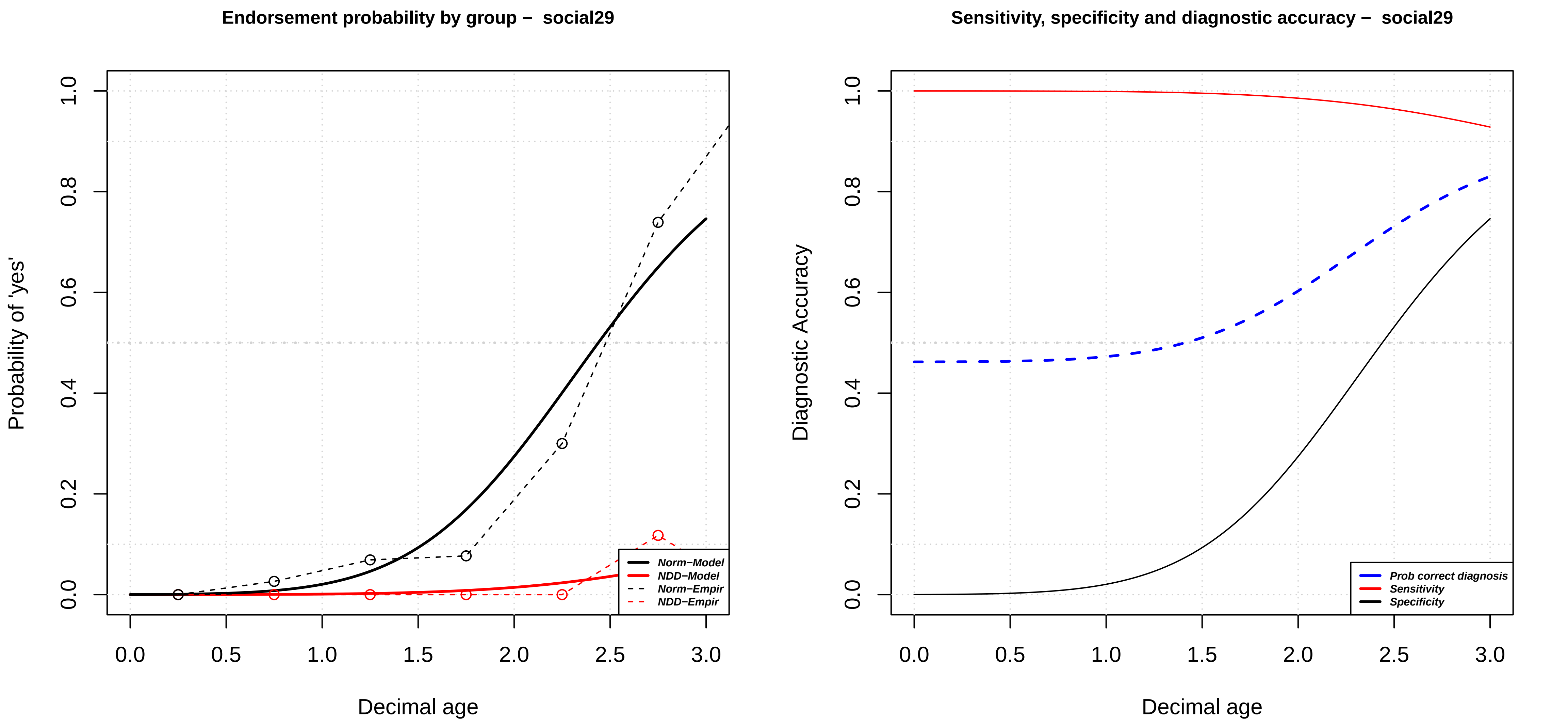

Endorsement probability by group – social29
Probability of 'yes'
Decimal age
Norm−Model
NDD−Model
Norm−Empir
NDD−Empir
0.0
0.2
0.4
0.6
0.8
1.0
0.0
0.5
1.0
1.5
2.0
2.5
3.0
Sensitivity, specificity and diagnostic accuracy – social29
Diagnostic Accuracy
Decimal age
Prob correct diagnosis
Sensitivity
Specificity

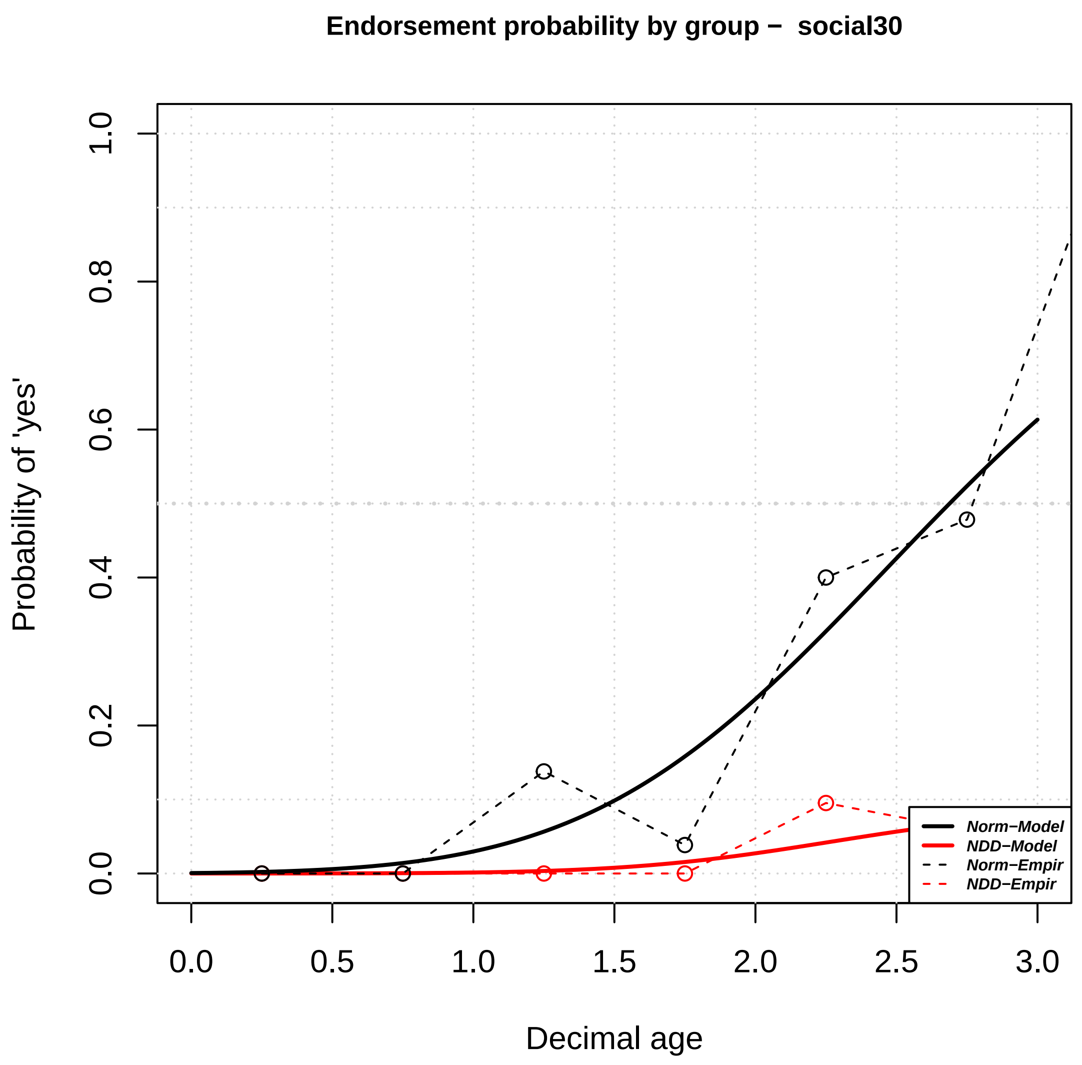

Endorsement probability by group – social30
Probability of 'yes'
Decimal age
0.0
0.2
0.4
0.6
0.8
1.0
0.0
0.5
1.0
1.5
2.0
2.5
3.0
Norm−Model
NDD−Model
Norm−Empir
NDD−Empir


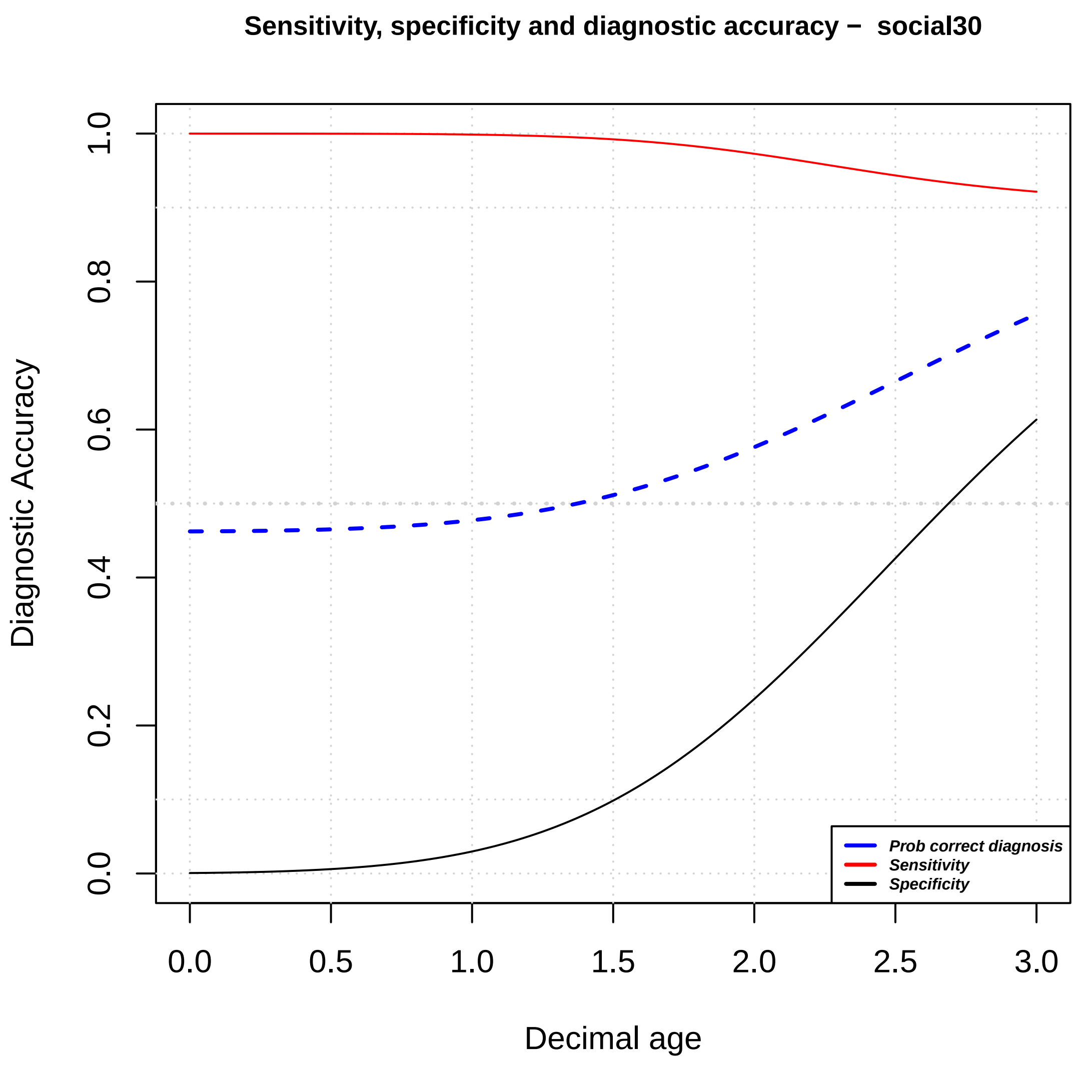

Sensitivity, specificity and diagnostic accuracy – social30
Diagnostic Accuracy
Decimal age
0.0
0.2
0.4
0.6
0.8
1.0
0.0
0.5
1.0
1.5
2.0
2.5
3.0
Prob correct diagnosis
Sensitivity
Specificity

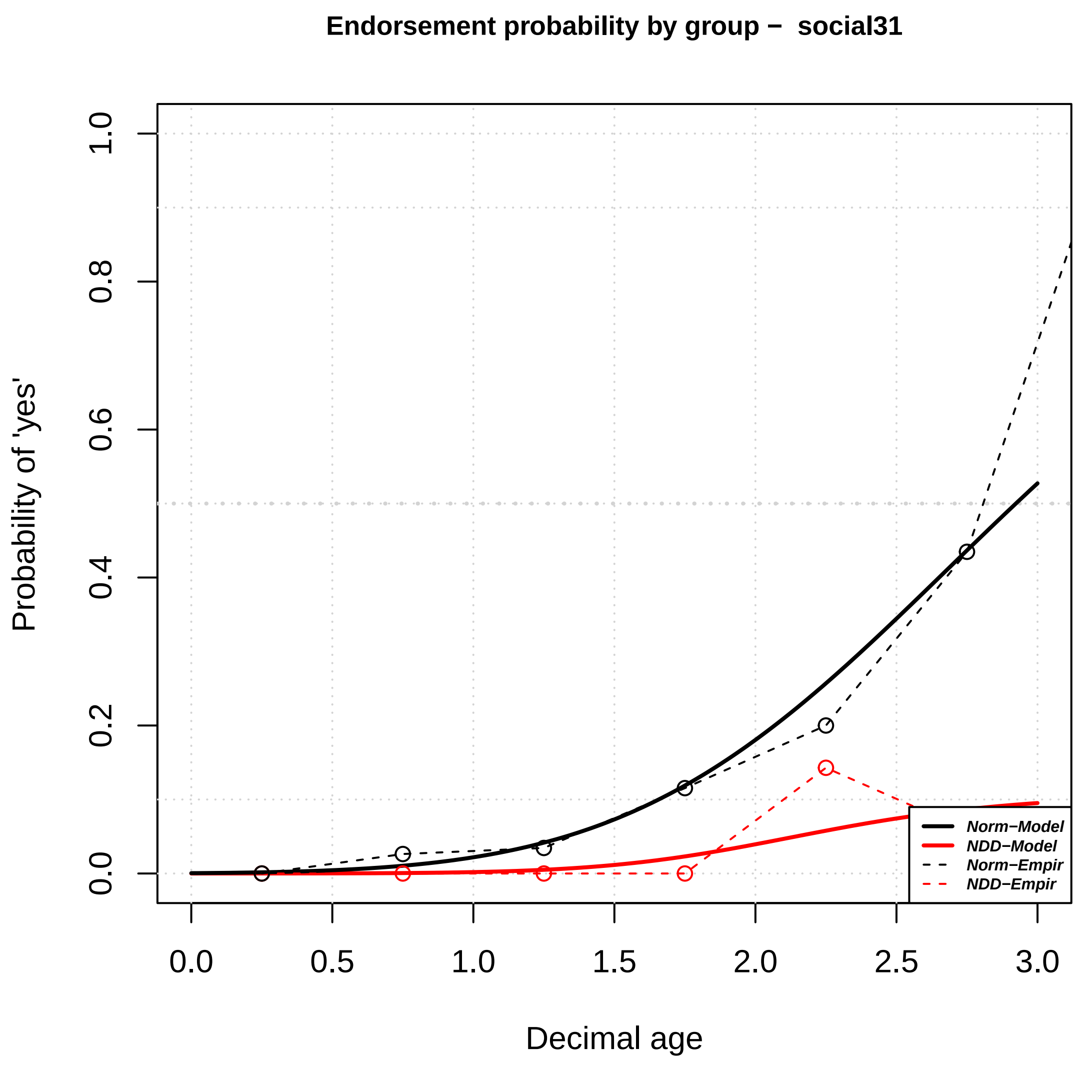

Endorsement probability by group − social31
Probability of 'yes'
0.0
0.2
0.4
0.6
0.8
1.0
0.0
0.5
1.0
1.5
2.0
2.5
3.0
Decimal age
Norm−Model
NDD−Model
Norm−Empir
NDD−Empir


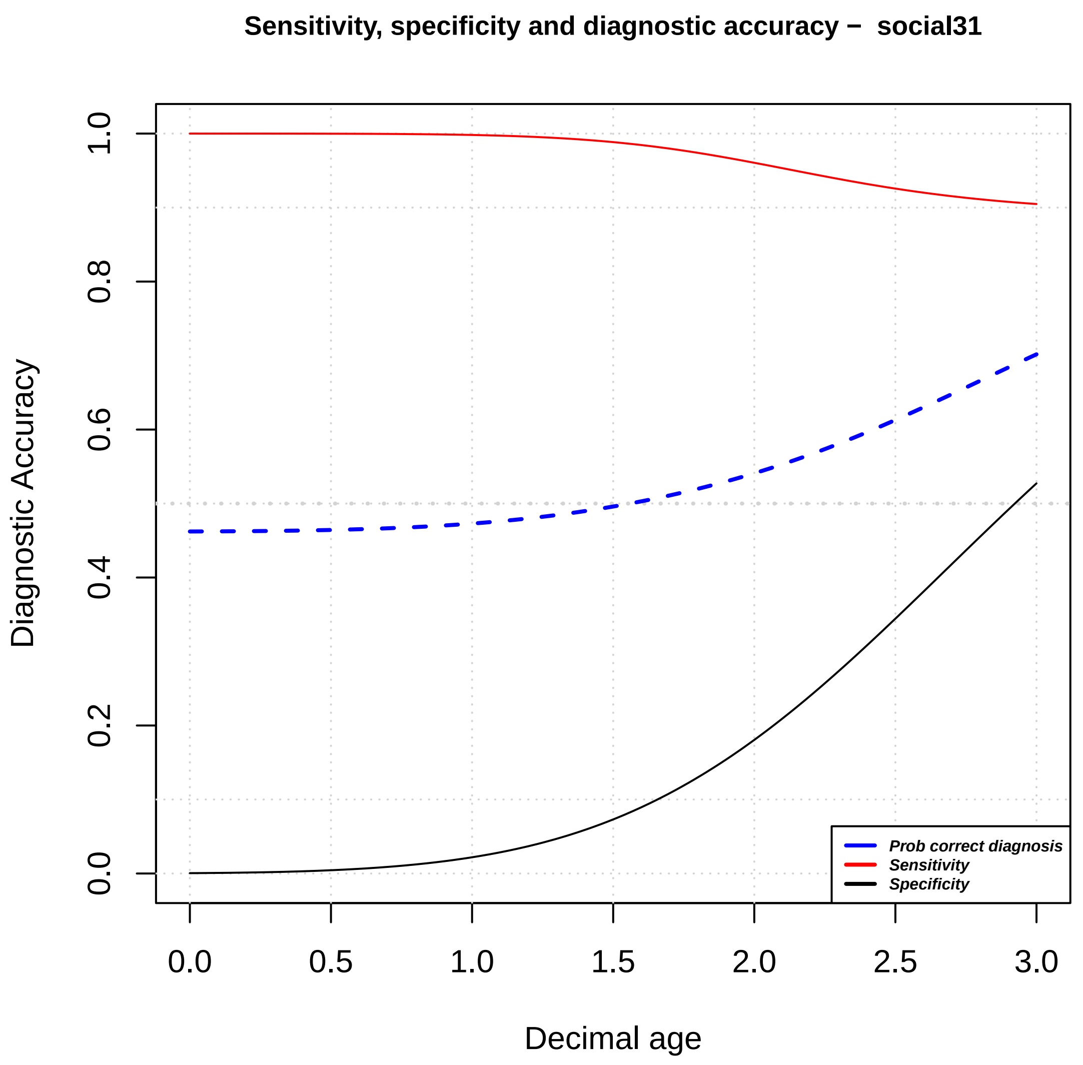

Sensitivity, specificity and diagnostic accuracy − social31
Diagnostic Accuracy
0.0
0.2
0.4
0.6
0.8
1.0
0.0
0.5
1.0
1.5
2.0
2.5
3.0
Decimal age
Prob correct diagnosis
Sensitivity
Specificity

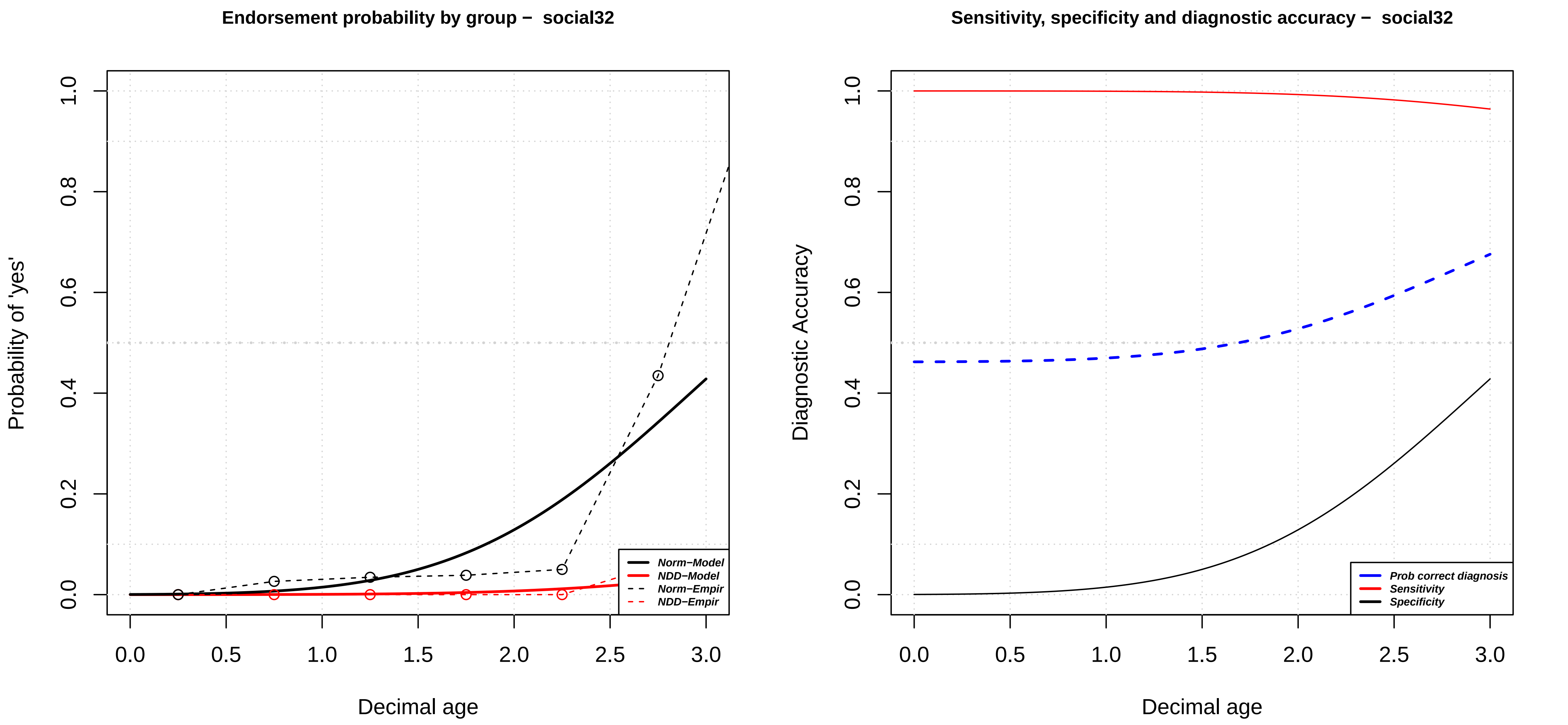

Endorsement probability by group – social32
Probability of 'yes'
Decimal age
Norm–Model
NDD–Model
Norm–Empir
NDD–Empir
Sensitivity, specificity and diagnostic accuracy – social32
Diagnostic Accuracy
Decimal age
Prob correct diagnosis
Sensitivity
Specificity

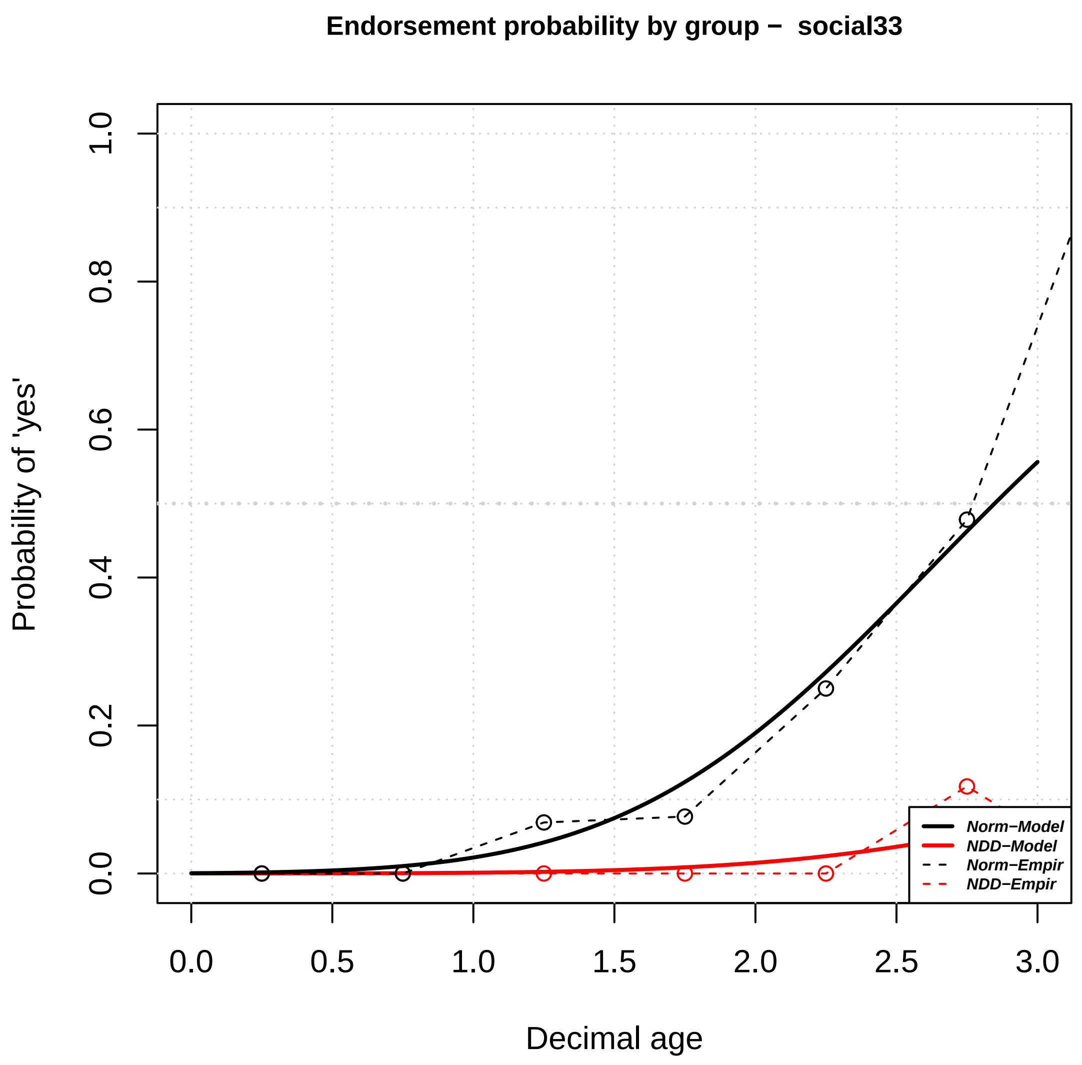
Endorsement probability by group − social33
Probability of 'yes'
Decimal age
Norm−Model
NDD−Model
Norm−Empir
NDD−Empir
0.0
0.2
0.4
0.6
0.8
1.0
0.0
0.5
1.0
1.5
2.0
2.5
3.0

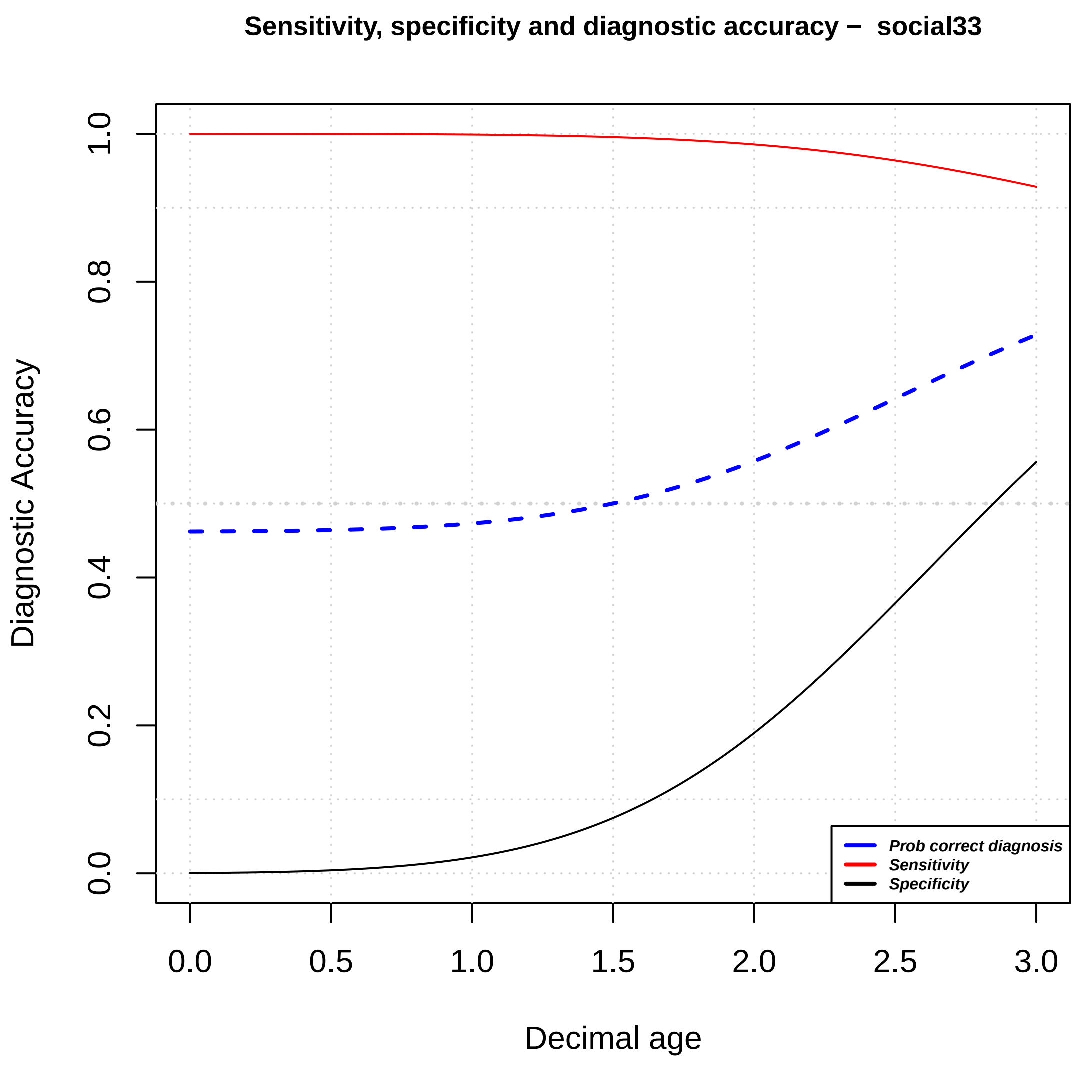
Sensitivity, specificity and diagnostic accuracy − social33
Diagnostic Accuracy
Decimal age
Prob correct diagnosis
Sensitivity
Specificity
0.0
0.2
0.4
0.6
0.8
1.0
0.0
0.5
1.0
1.5
2.0
2.5
3.0

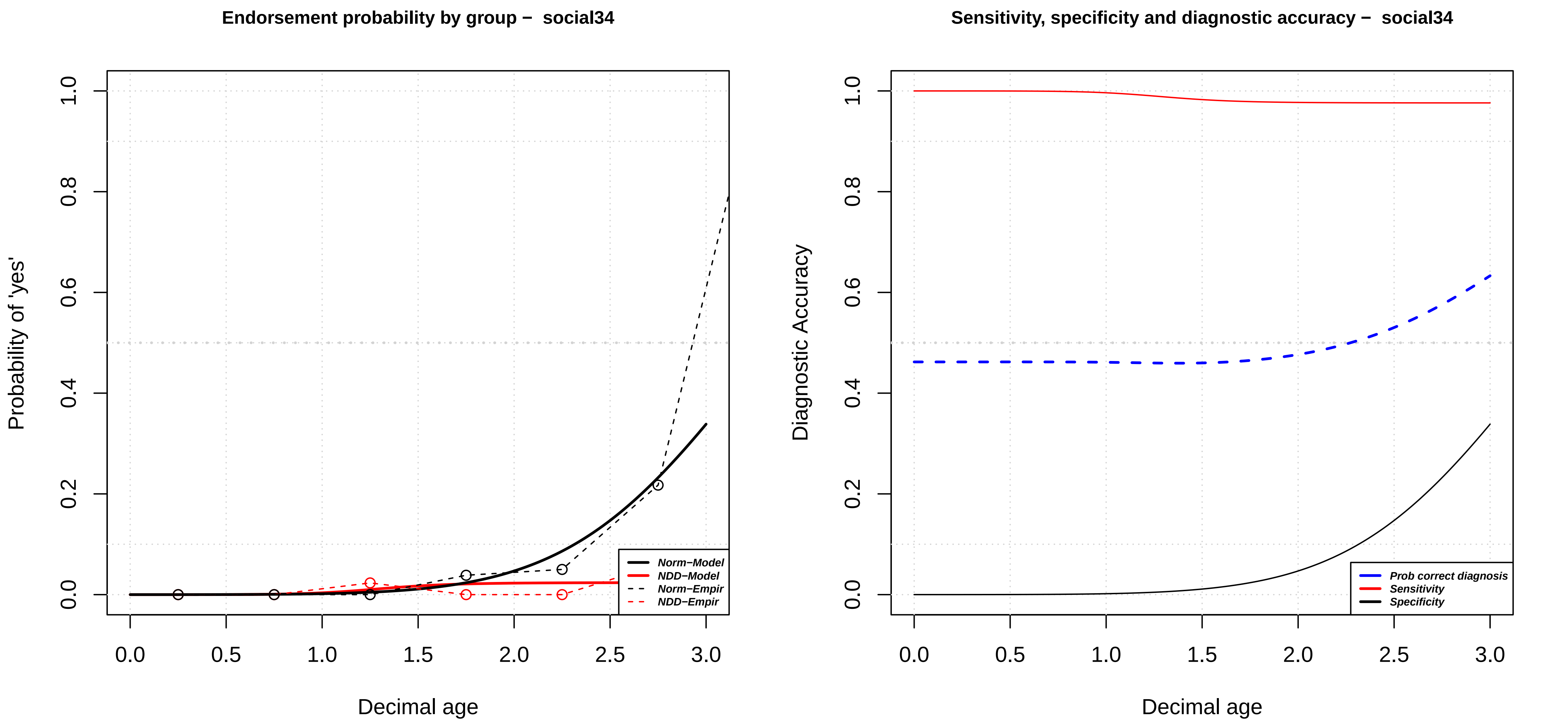
Endorsement probability by group – social34
Probability of 'yes'
Decimal age
0.0
0.2
0.4
0.6
0.8
1.0
0.0
0.5
1.0
1.5
2.0
2.5
3.0
Norm−Model
NDD−Model
Norm−Empir
NDD−Empir
Sensitivity, specificity and diagnostic accuracy – social34
Diagnostic Accuracy
Decimal age
Prob correct diagnosis
Sensitivity
Specificity

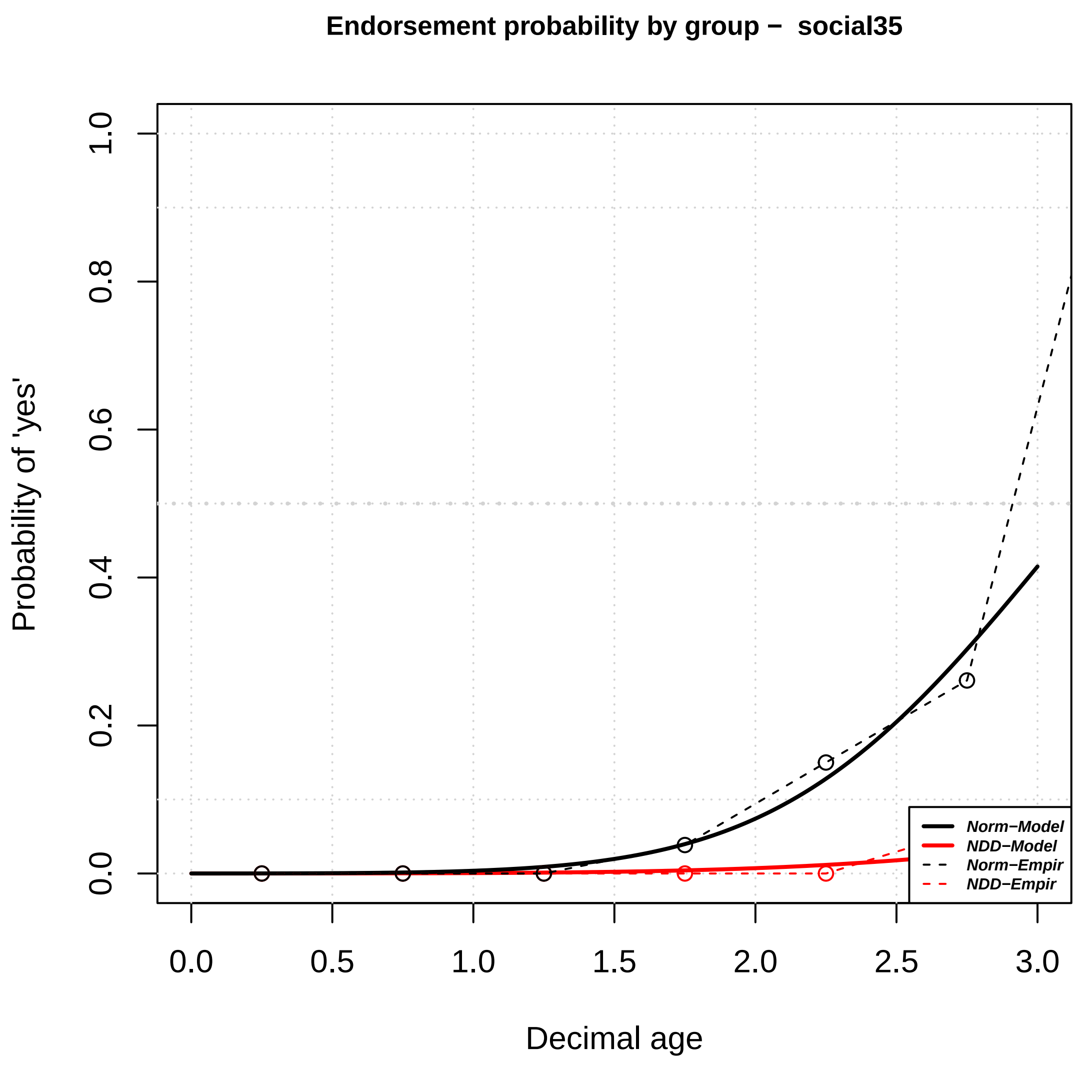

Endorsement probability by group – social35
Probability of 'yes'
1.0
0.8
0.6
0.4
0.2
0.0
0.0
0.5
1.0
1.5
2.0
2.5
3.0
Decimal age
Norm−Model
NDD−Model
Norm−Empir
NDD−Empir


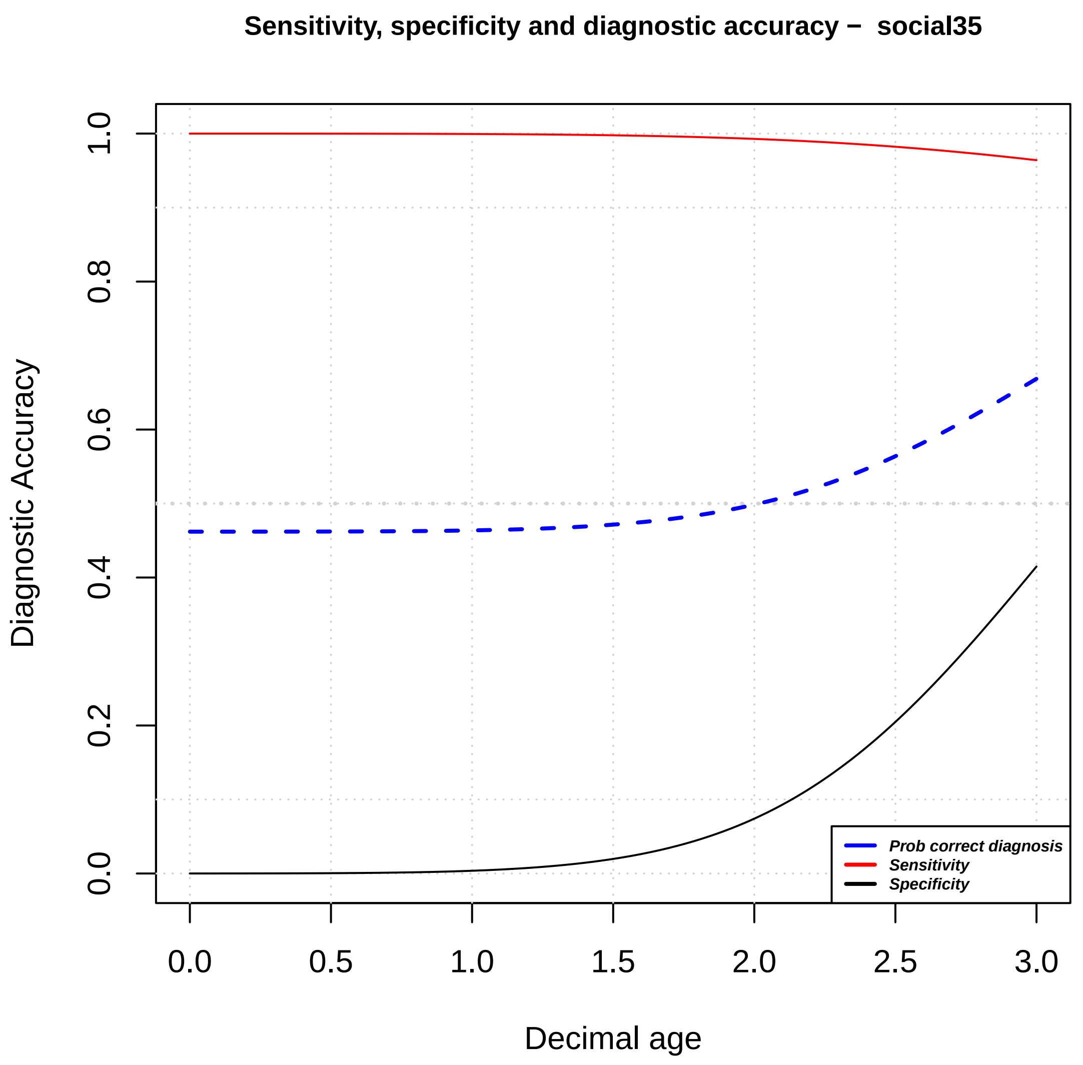

Sensitivity, specificity and diagnostic accuracy – social35
Diagnostic Accuracy
1.0
0.8
0.6
0.4
0.2
0.0
0.0
0.5
1.0
1.5
2.0
2.5
3.0
Decimal age
Prob correct diagnosis
Sensitivity
Specificity

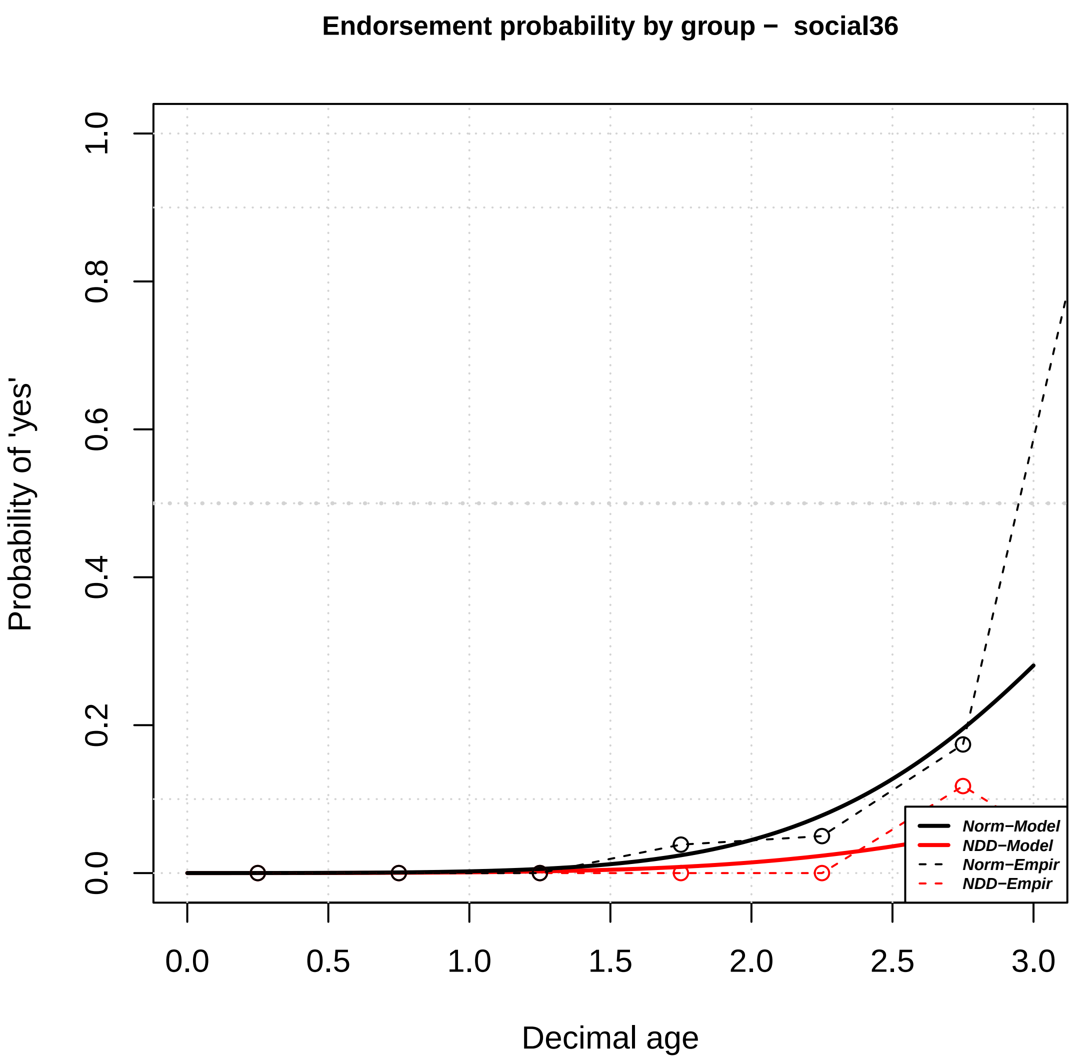

Endorsement probability by group – social36
Probability of 'yes'
Decimal age
0.0
0.2
0.4
0.6
0.8
1.0
0.0
0.5
1.0
1.5
2.0
2.5
3.0
Norm–Model
NDD–Model
Norm–Empir
NDD–Empir


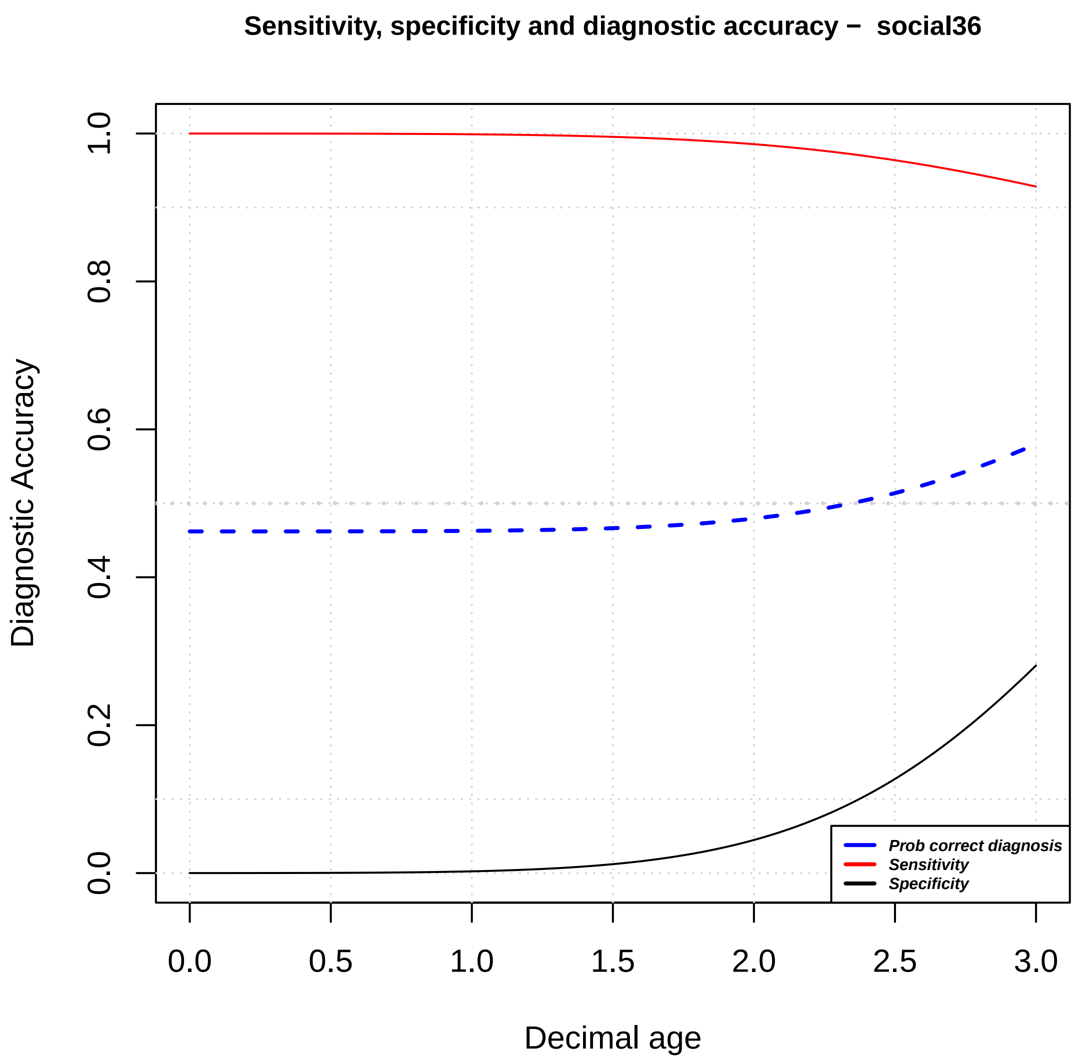

Sensitivity, specificity and diagnostic accuracy – social36
Diagnostic Accuracy
Decimal age
0.0
0.2
0.4
0.6
0.8
1.0
0.0
0.5
1.0
1.5
2.0
2.5
3.0
Prob correct diagnosis
Sensitivity
Specificity

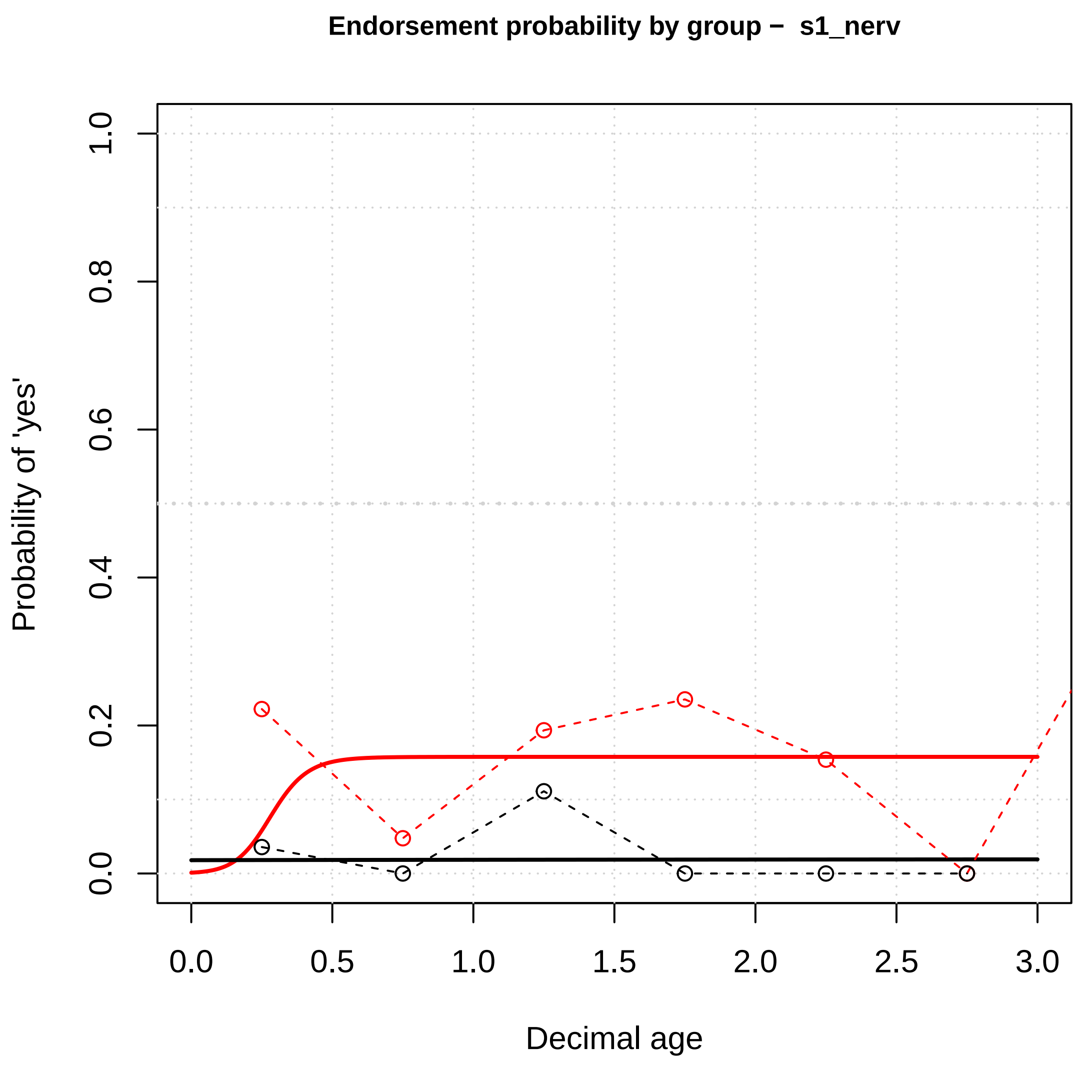

Endorsement probability by group – s1_nerv
Probability of 'yes'
0.0
0.2
0.4
0.6
0.8
1.0
0.0
0.5
1.0
1.5
2.0
2.5
3.0
Decimal age


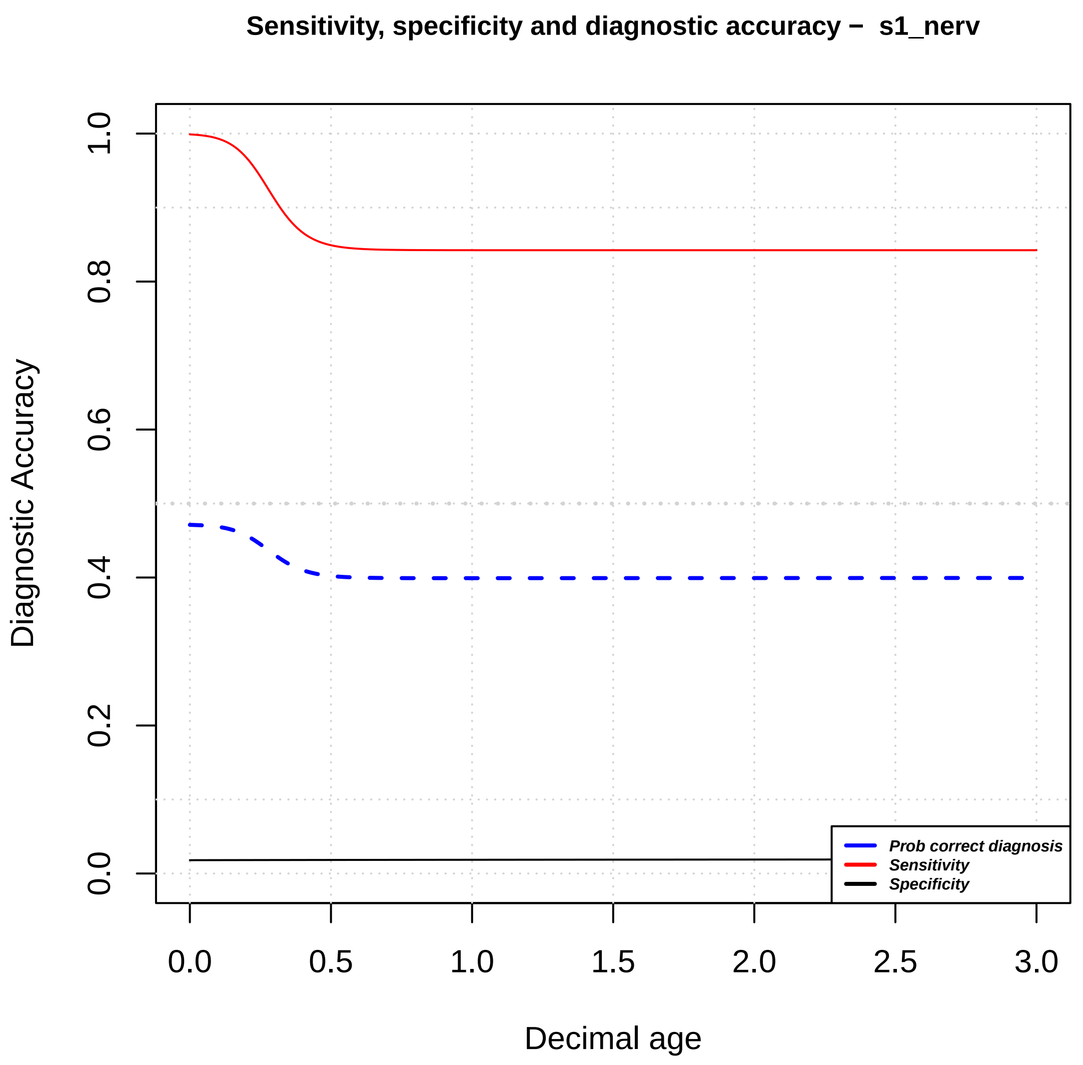

Sensitivity, specificity and diagnostic accuracy – s1_nerv
Diagnostic Accuracy
0.0
0.2
0.4
0.6
0.8
1.0
0.0
0.5
1.0
1.5
2.0
2.5
3.0
Decimal age
Prob correct diagnosis
Sensitivity
Specificity

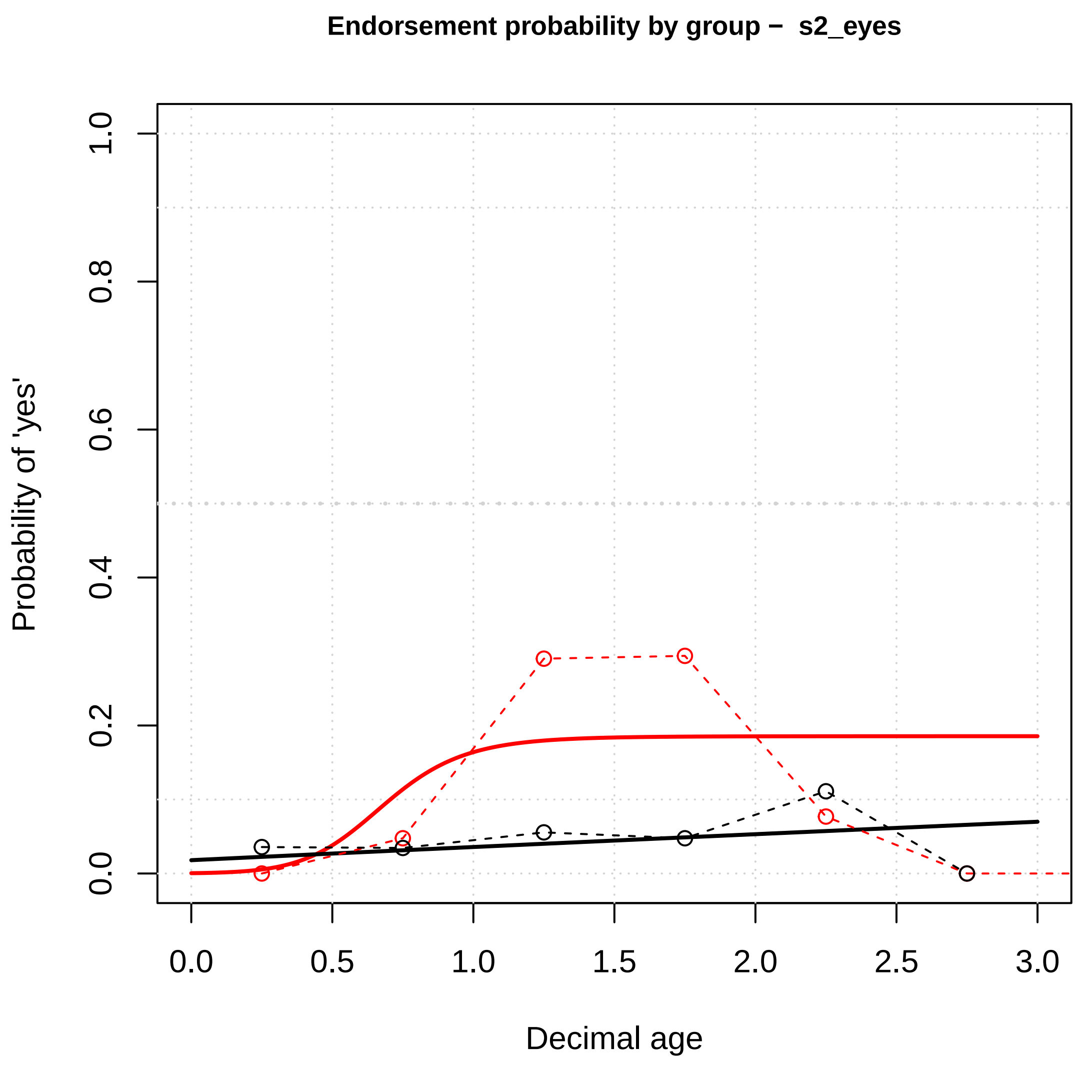

Endorsement probability by group – s2_eyes
Probability of 'yes'
Decimal age
0.0
0.2
0.4
0.6
0.8
1.0
0.0
0.5
1.0
1.5
2.0
2.5
3.0


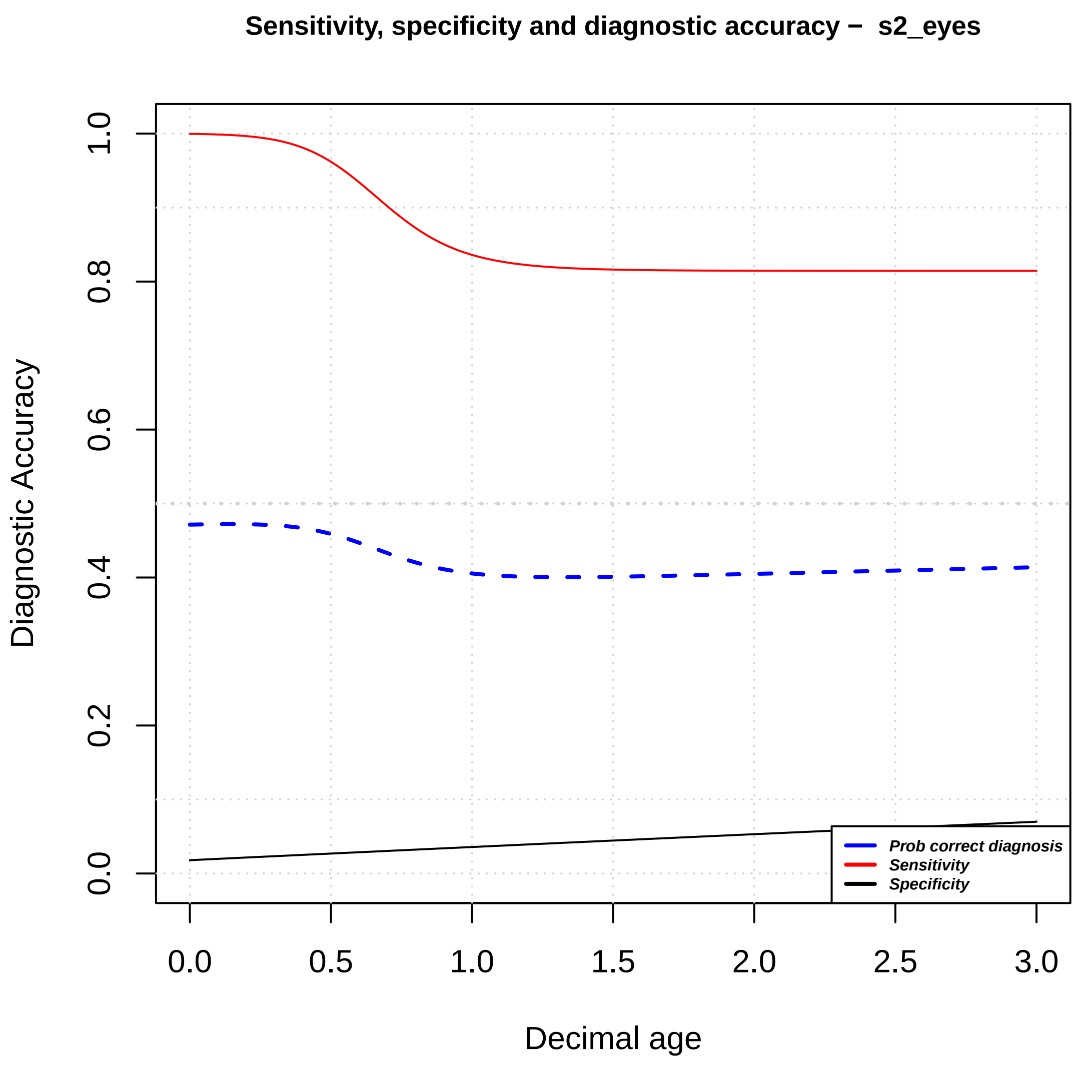

Sensitivity, specificity and diagnostic accuracy – s2_eyes
Diagnostic Accuracy
Decimal age
0.0
0.2
0.4
0.6
0.8
1.0
0.0
0.5
1.0
1.5
2.0
2.5
3.0
Prob correct diagnosis
Sensitivity
Specificity

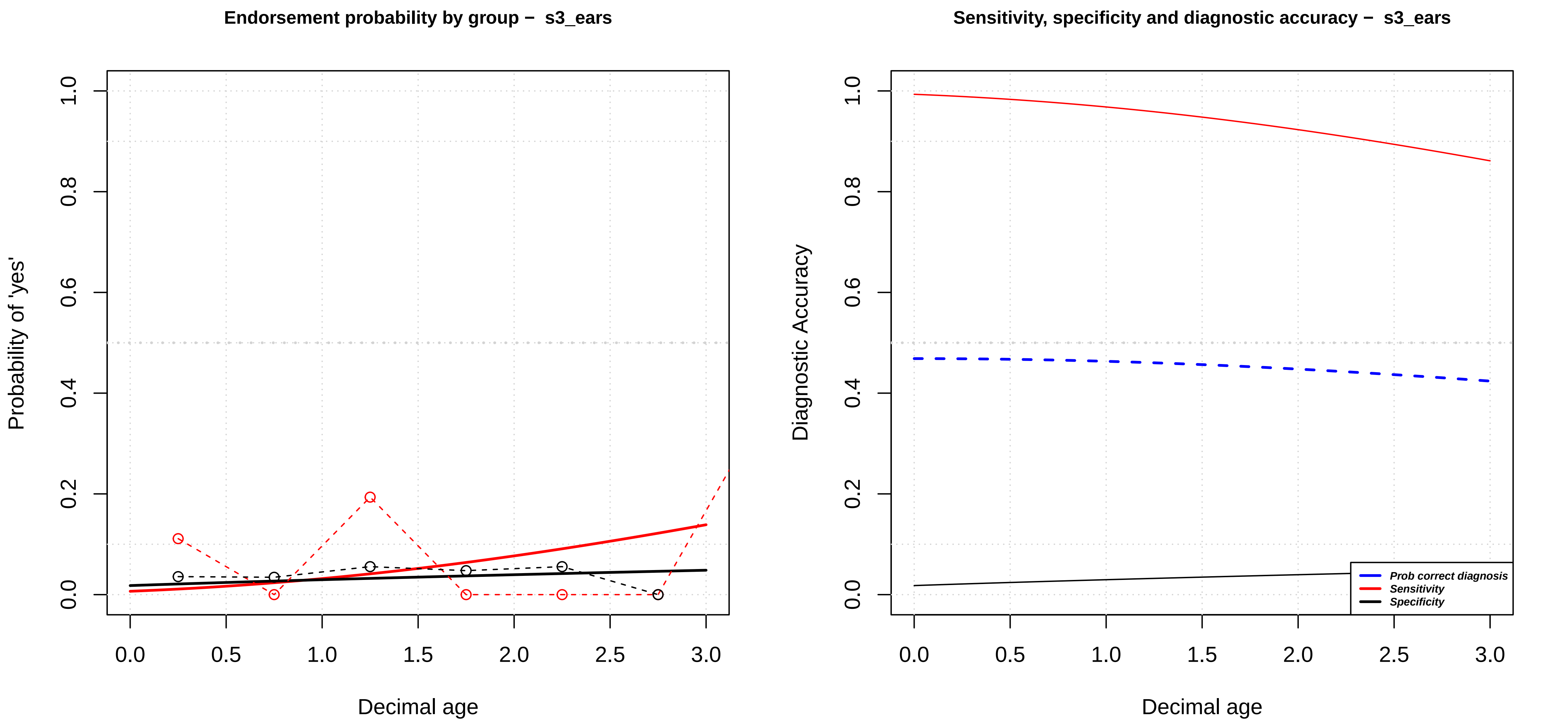

Endorsement probability by group – s3_ears
Probability of 'yes'
Decimal age
0.0
0.2
0.4
0.6
0.8
1.0
0.0
0.5
1.0
1.5
2.0
2.5
3.0
Sensitivity, specificity and diagnostic accuracy – s3_ears
Diagnostic Accuracy
Decimal age
Prob correct diagnosis
Sensitivity
Specificity

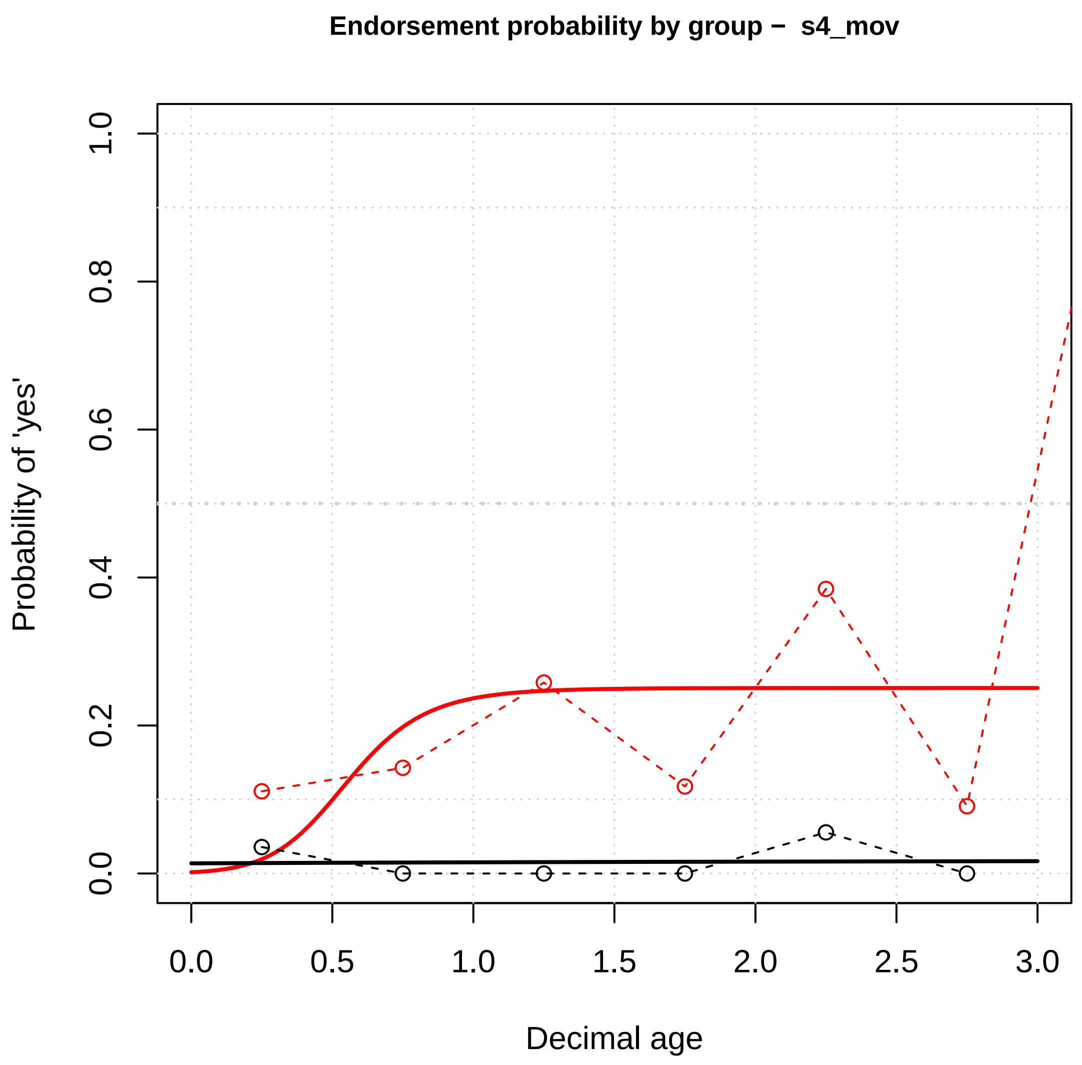
Endorsement probability by group – s4_mov
Probability of 'yes'
Decimal age
0.0
0.2
0.4
0.6
0.8
1.0
0.0
0.5
1.0
1.5
2.0
2.5
3.0

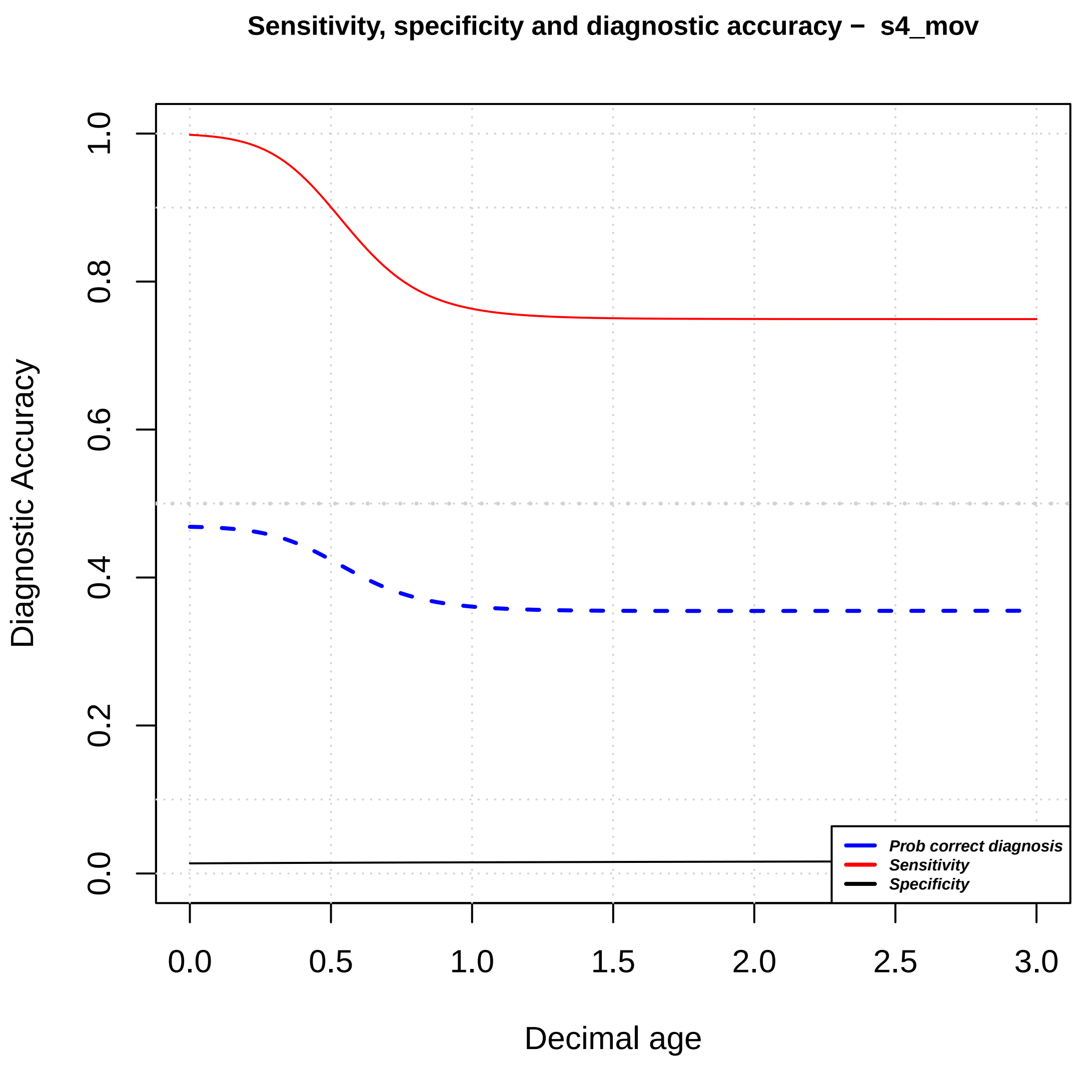
Sensitivity, specificity and diagnostic accuracy – s4_mov
Diagnostic Accuracy
Decimal age
0.0
0.2
0.4
0.6
0.8
1.0
0.0
0.5
1.0
1.5
2.0
2.5
3.0
Prob correct diagnosis
Sensitivity
Specificity

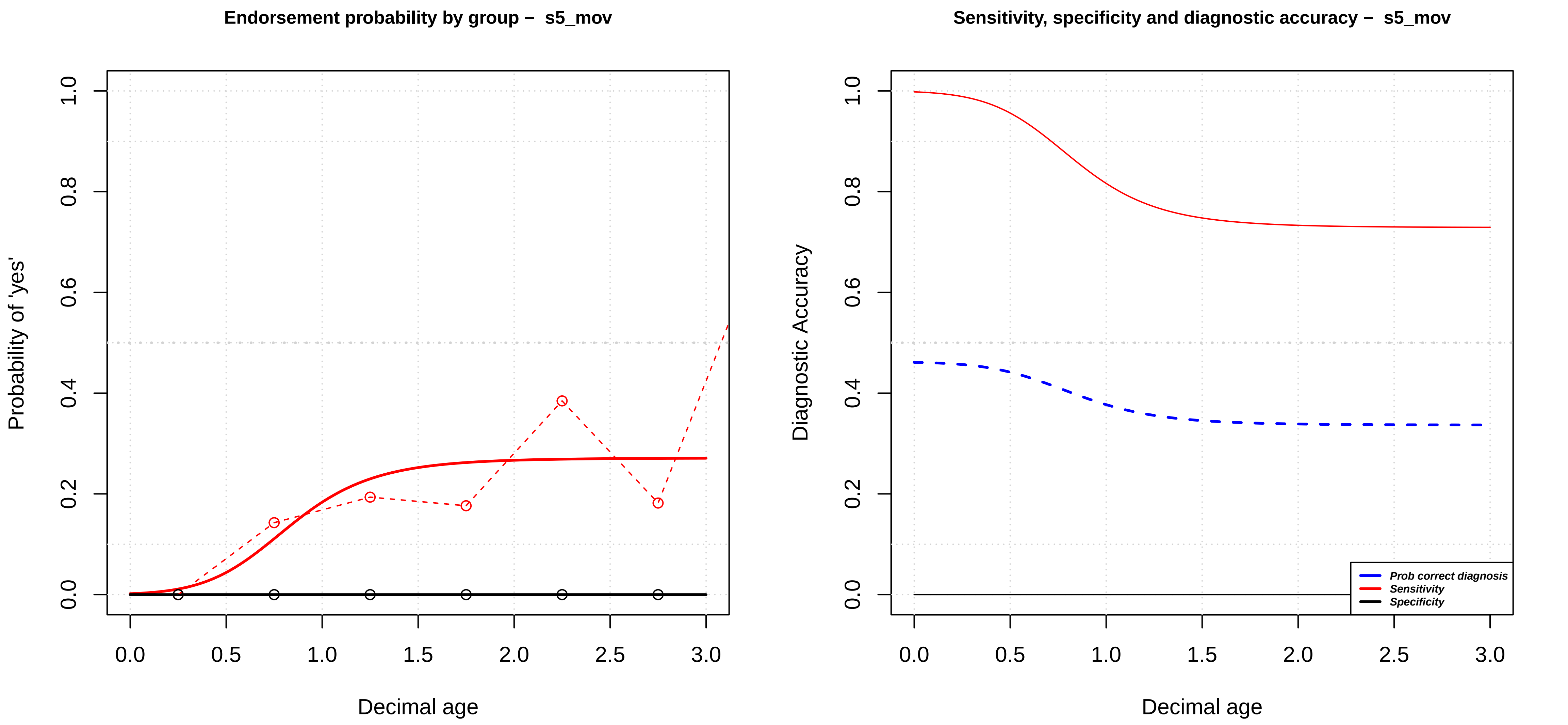
Endorsement probability by group − s5_mov
Probability of 'yes'
Decimal age
0.0
0.2
0.4
0.6
0.8
1.0
0.0
0.5
1.0
1.5
2.0
2.5
3.0
Sensitivity, specificity and diagnostic accuracy − s5_mov
Diagnostic Accuracy
Decimal age
Prob correct diagnosis
Sensitivity
Specificity

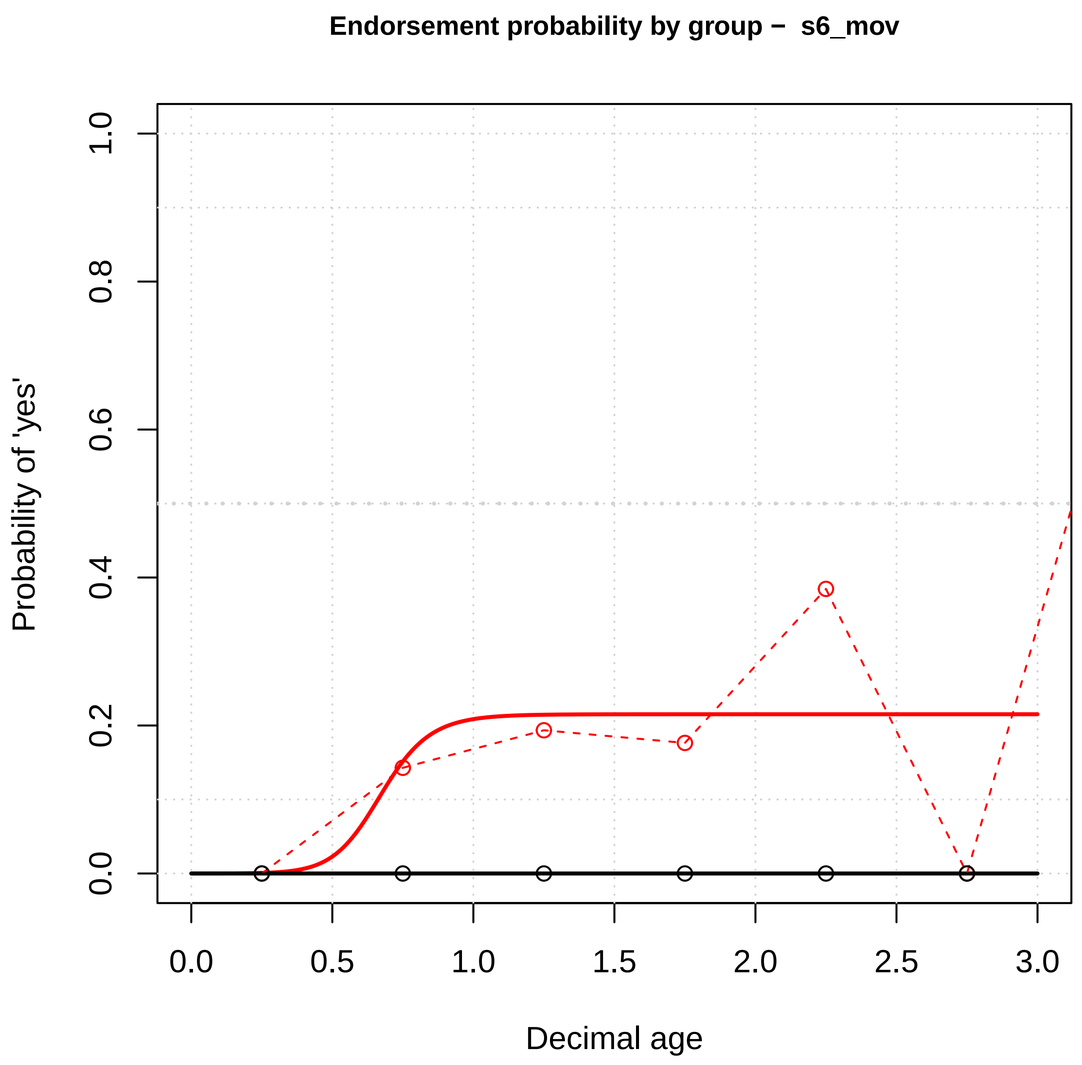

Endorsement probability by group – s6_mov
Probability of 'yes'
0.0
0.2
0.4
0.6
0.8
1.0
0.0
0.5
1.0
1.5
2.0
2.5
3.0
Decimal age


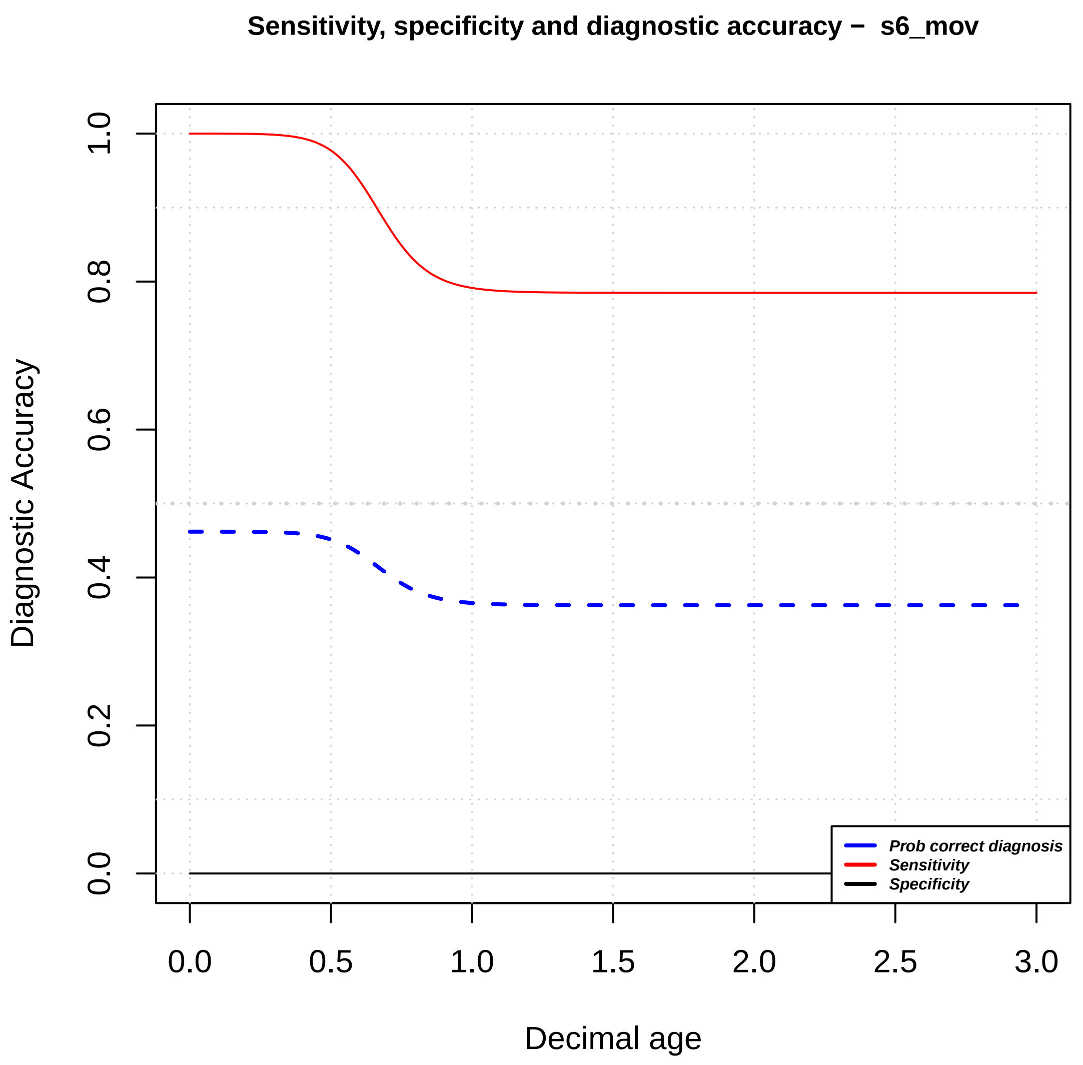

Sensitivity, specificity and diagnostic accuracy – s6_mov
Diagnostic Accuracy
0.0
0.2
0.4
0.6
0.8
1.0
0.0
0.5
1.0
1.5
2.0
2.5
3.0
Decimal age
Prob correct diagnosis
Sensitivity
Specificity

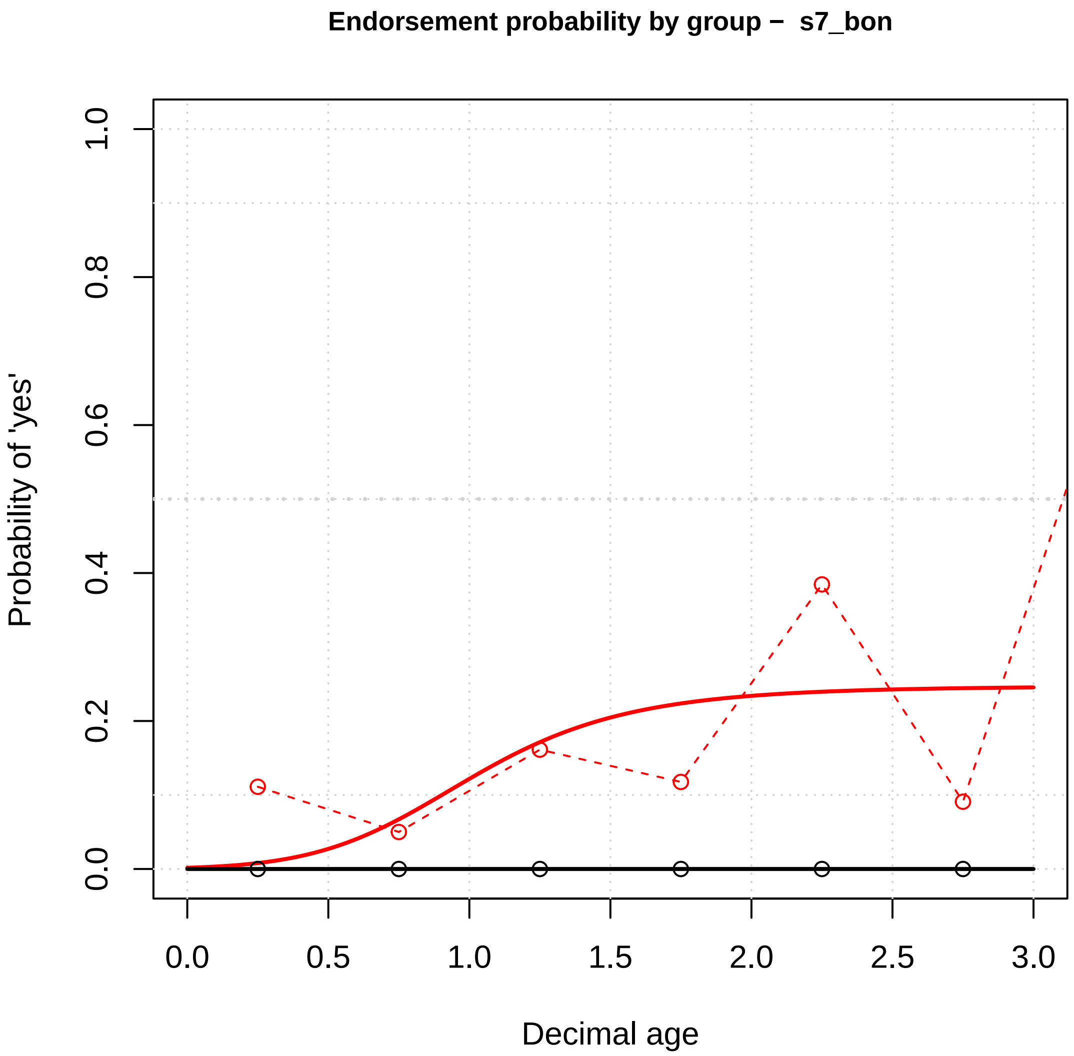

Endorsement probability by group – s7_bon
Probability of 'yes'
Decimal age
0.0
0.2
0.4
0.6
0.8
1.0
0.0
0.5
1.0
1.5
2.0
2.5
3.0


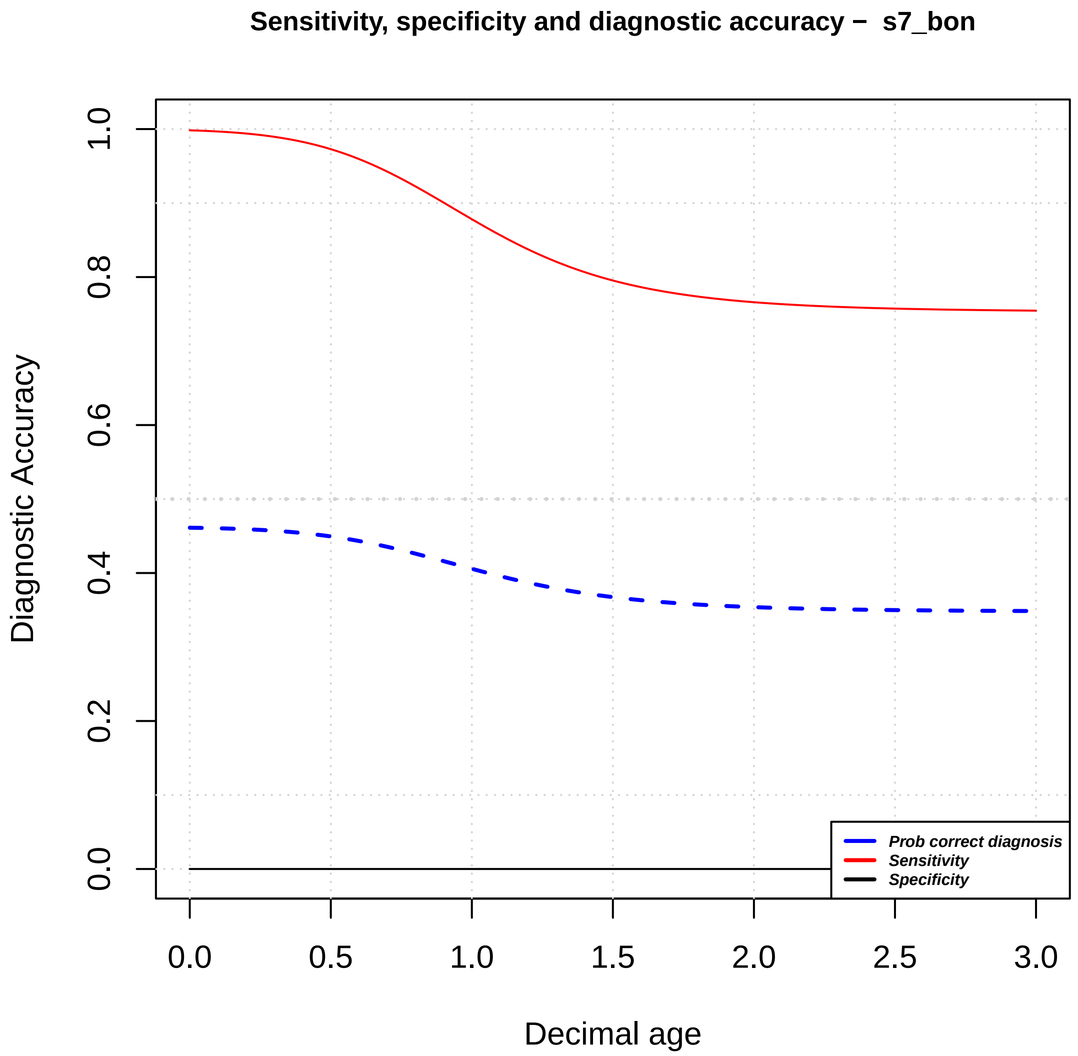

Sensitivity, specificity and diagnostic accuracy – s7_bon
Diagnostic Accuracy
Decimal age
0.0
0.2
0.4
0.6
0.8
1.0
0.0
0.5
1.0
1.5
2.0
2.5
3.0
Prob correct diagnosis
Sensitivity
Specificity

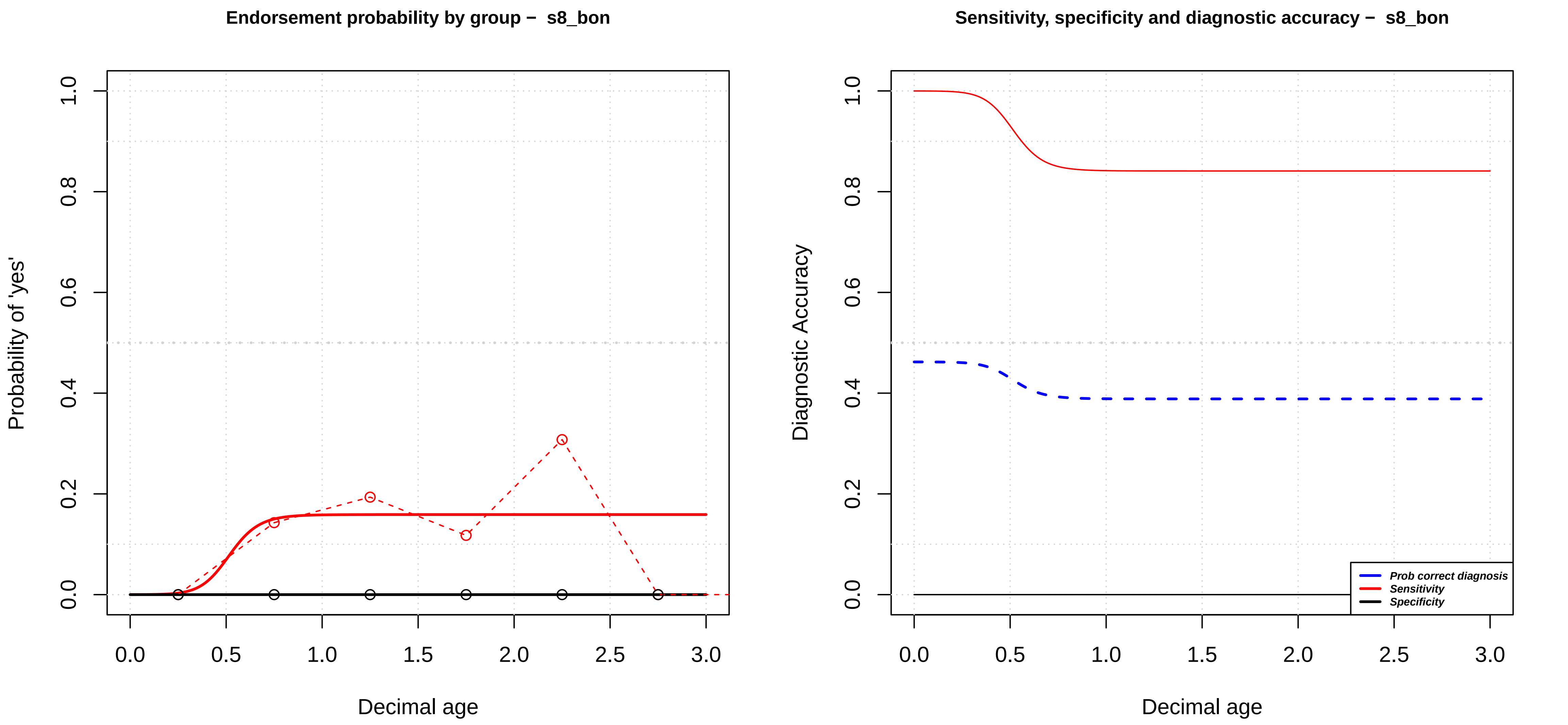

Endorsement probability by group – s8_bon
Probability of 'yes'
Decimal age
Sensitivity, specificity and diagnostic accuracy – s8_bon
Diagnostic Accuracy
Decimal age
Prob correct diagnosis
Sensitivity
Specificity

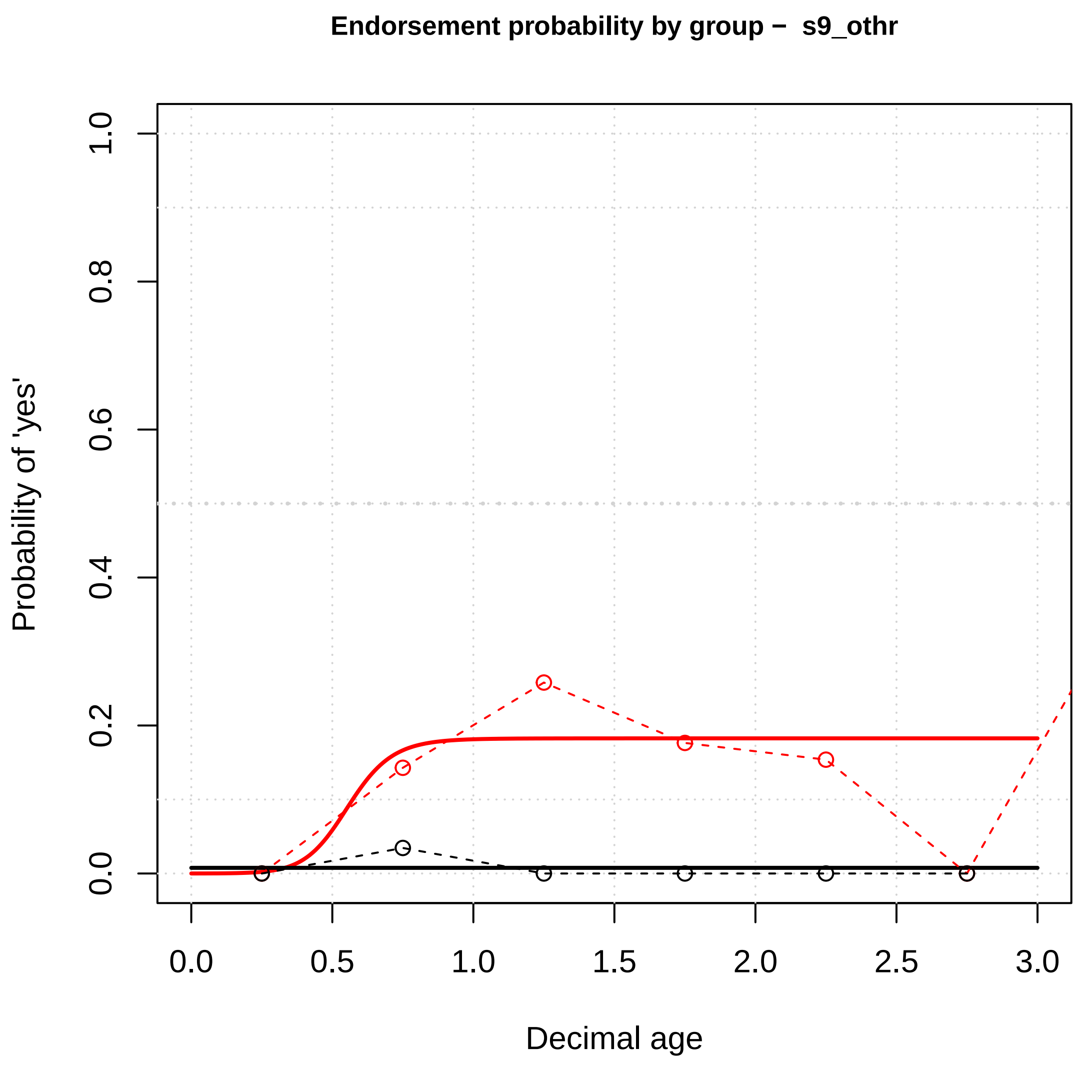

Endorsement probability by group – s9_othr
Probability of 'yes'
Decimal age
0.0
0.2
0.4
0.6
0.8
1.0
0.0
0.5
1.0
1.5
2.0
2.5
3.0


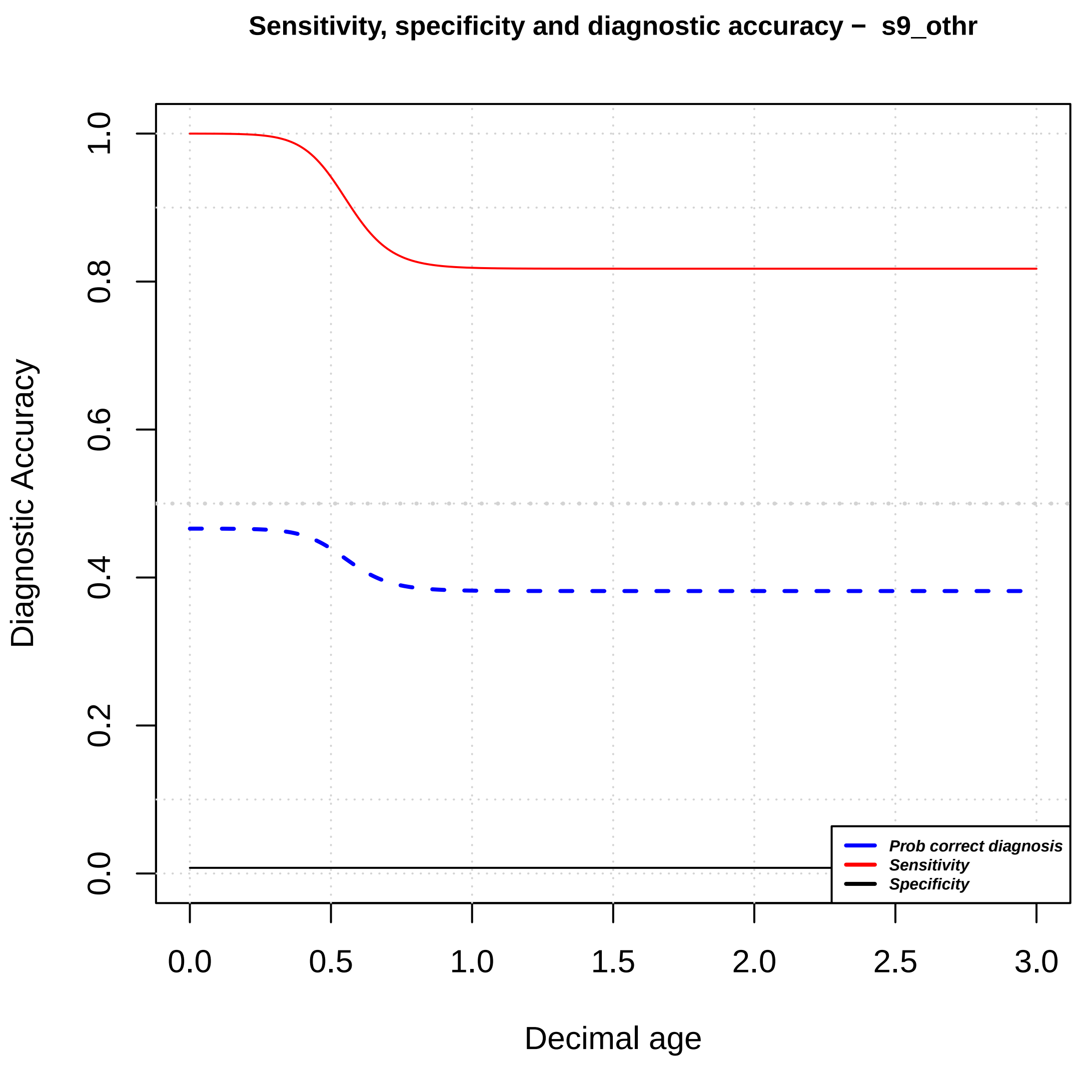

Sensitivity, specificity and diagnostic accuracy – s9_othr
Diagnostic Accuracy
Decimal age
0.0
0.2
0.4
0.6
0.8
1.0
0.0
0.5
1.0
1.5
2.0
2.5
3.0
Prob correct diagnosis
Sensitivity
Specificity

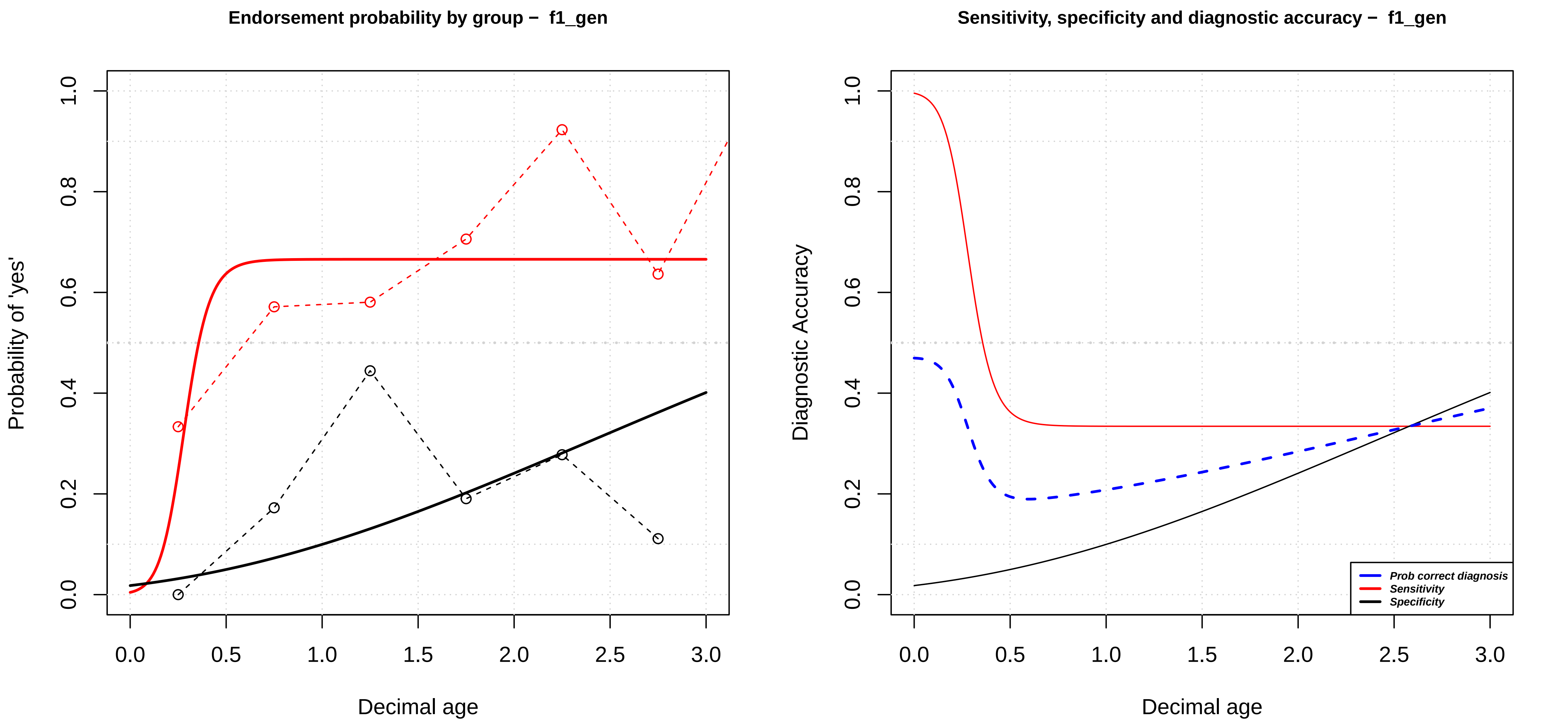

Endorsement probability by group – f1_gen
Probability of 'yes'
Decimal age
0.0
0.2
0.4
0.6
0.8
1.0
0.0
0.5
1.0
1.5
2.0
2.5
3.0
Sensitivity, specificity and diagnostic accuracy – f1_gen
Diagnostic Accuracy
Decimal age
Prob correct diagnosis
Sensitivity
Specificity

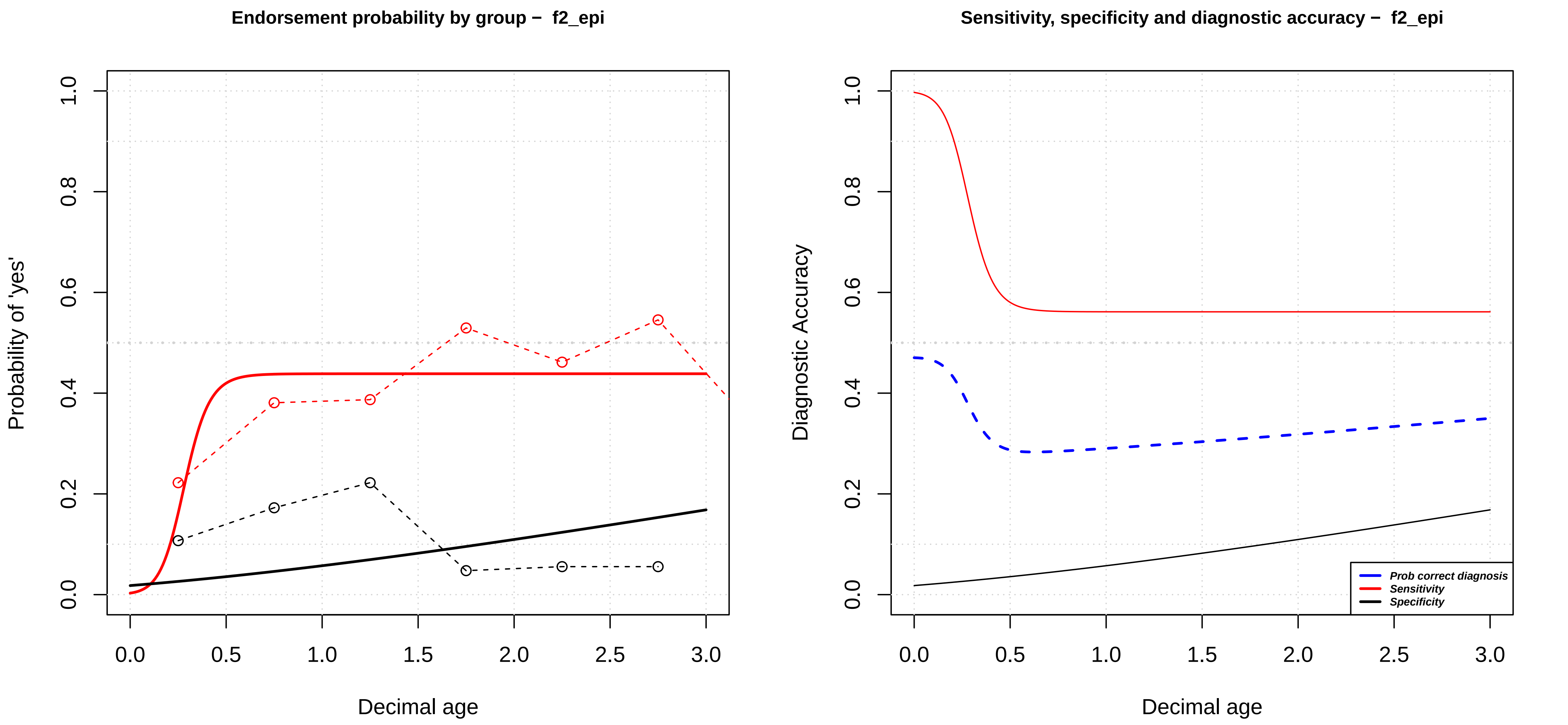

Endorsement probability by group – f2_epi
Probability of 'yes'
0.0
0.2
0.4
0.6
0.8
1.0
0.0
0.5
1.0
1.5
2.0
2.5
3.0
Decimal age
Sensitivity, specificity and diagnostic accuracy – f2_epi
Diagnostic Accuracy
Prob correct diagnosis
Sensitivity
Specificity
Decimal age

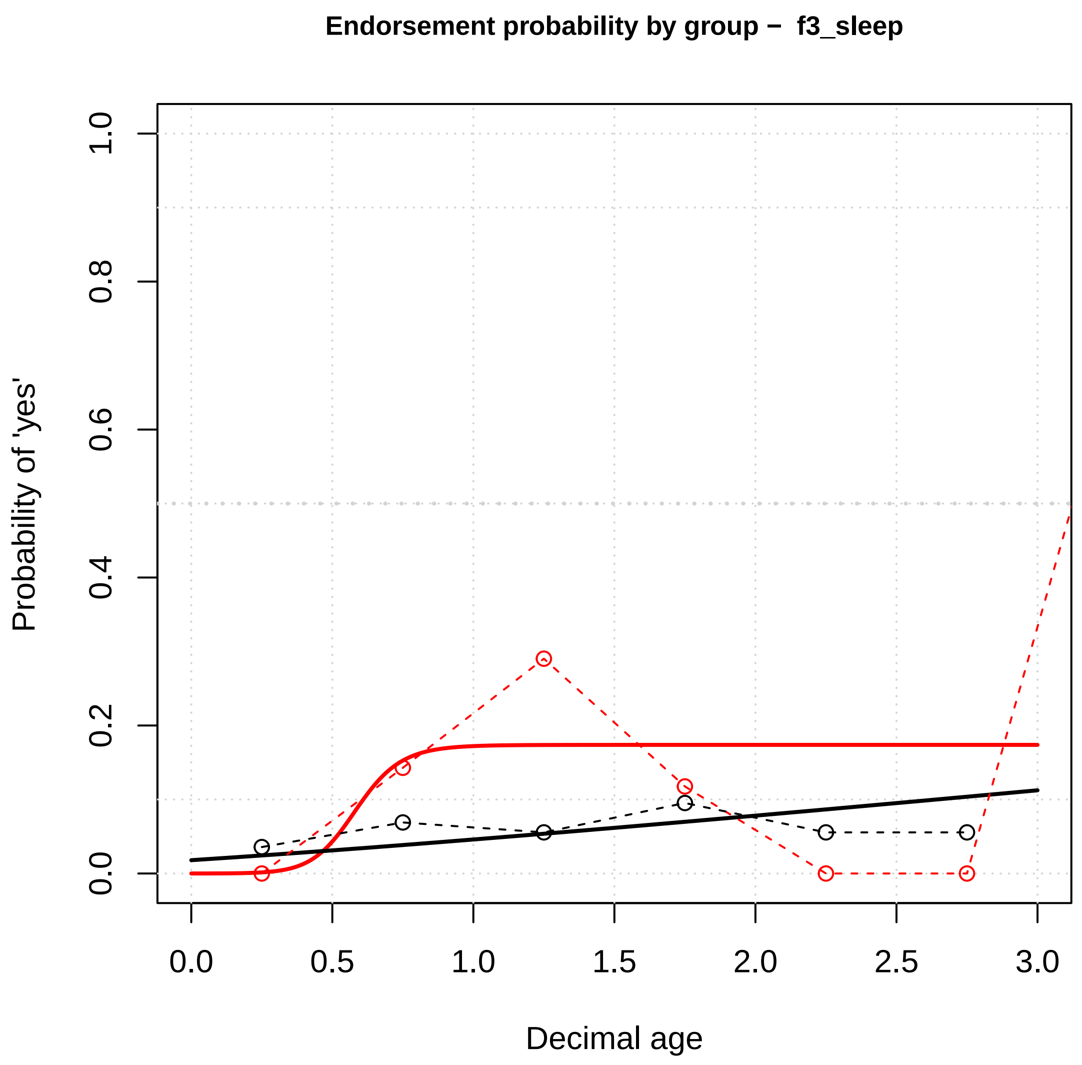
Endorsement probability by group – f3_sleep
Probability of 'yes'
1.0
0.8
0.6
0.4
0.2
0.0
0.0
0.5
1.0
1.5
2.0
2.5
3.0
Decimal age

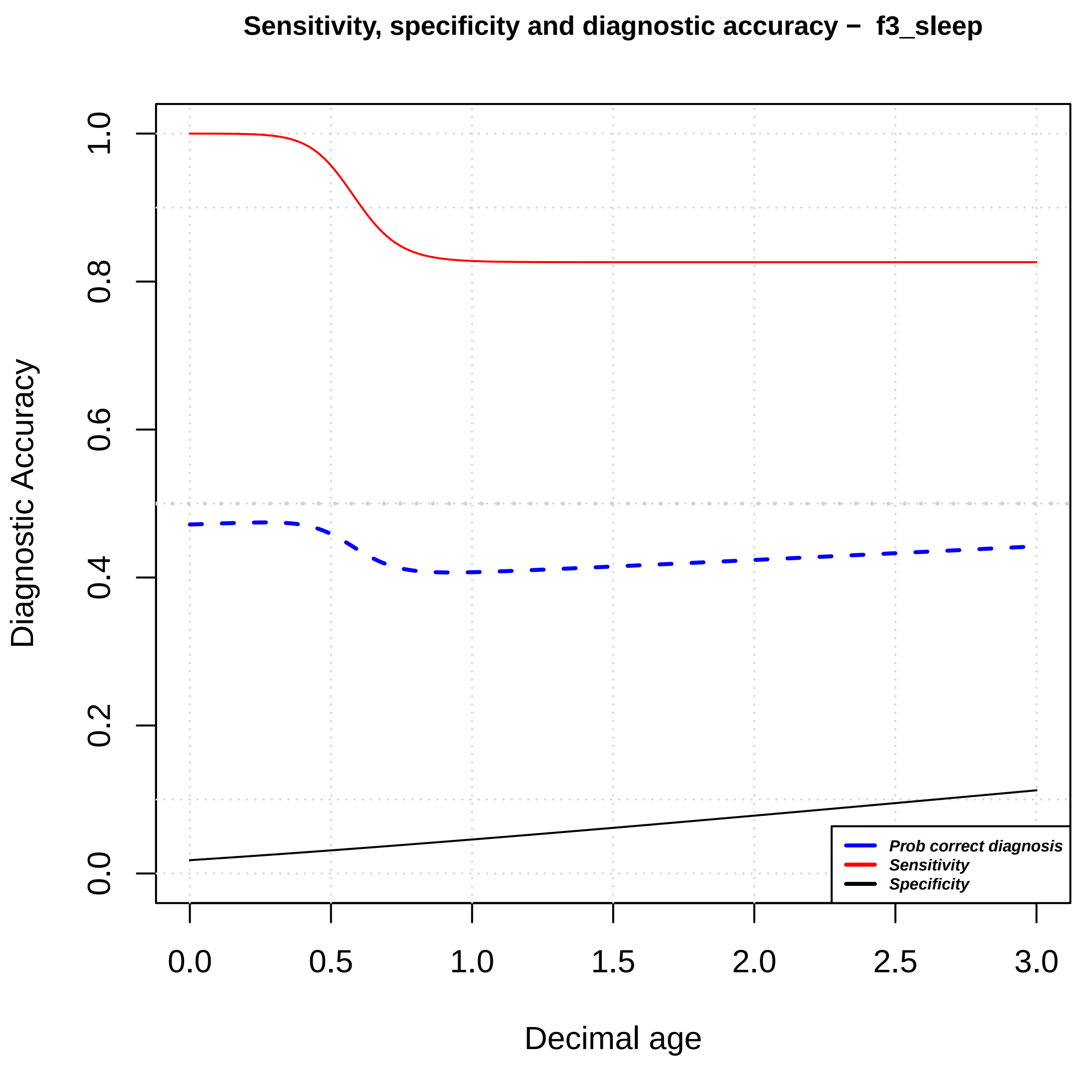
Sensitivity, specificity and diagnostic accuracy – f3_sleep
Diagnostic Accuracy
1.0
0.8
0.6
0.4
0.2
0.0
0.0
0.5
1.0
1.5
2.0
2.5
3.0
Decimal age
Prob correct diagnosis
Sensitivity
Specificity

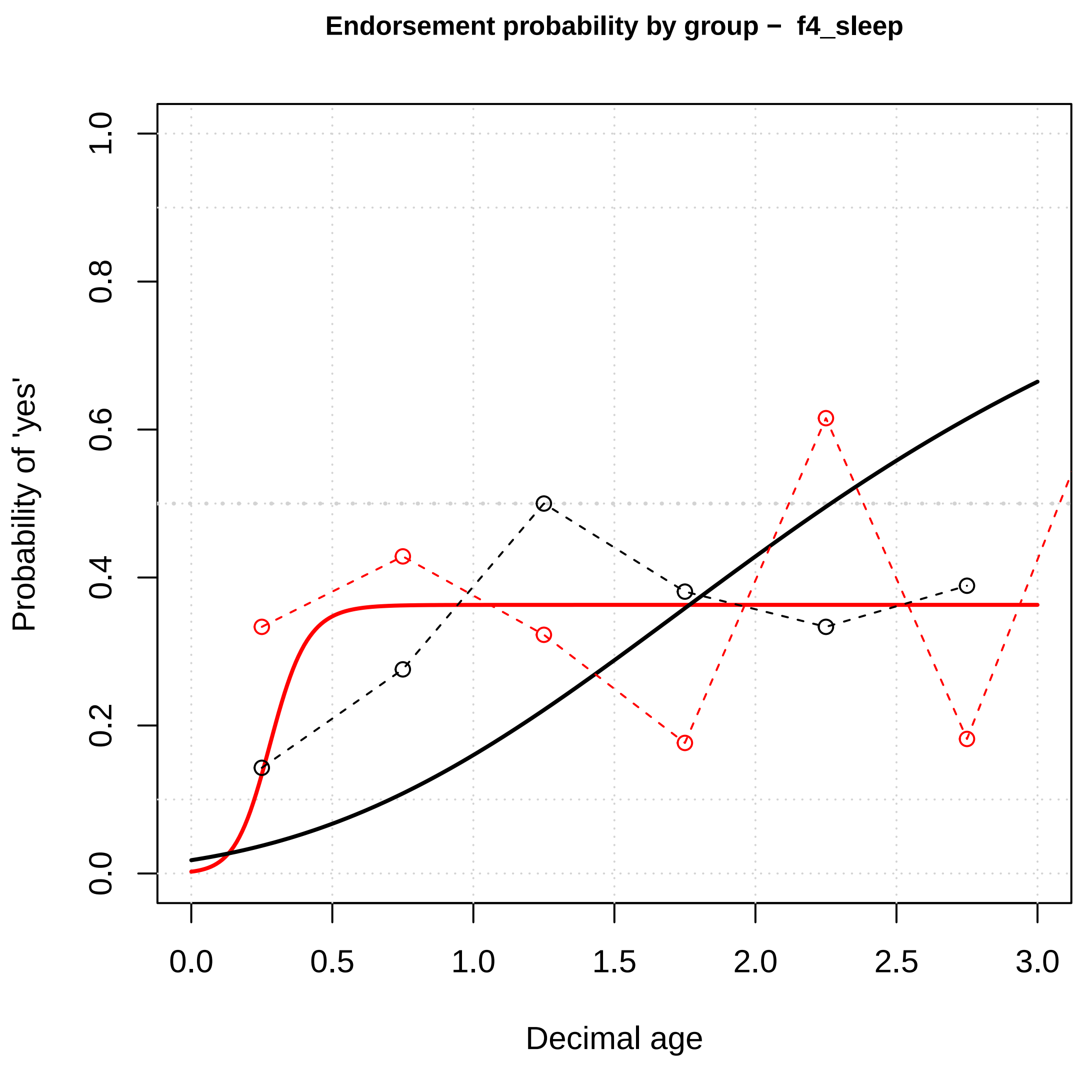

Endorsement probability by group – f4_sleep
Probability of 'yes'
Decimal age
0.0
0.2
0.4
0.6
0.8
1.0
0.0
0.5
1.0
1.5
2.0
2.5
3.0


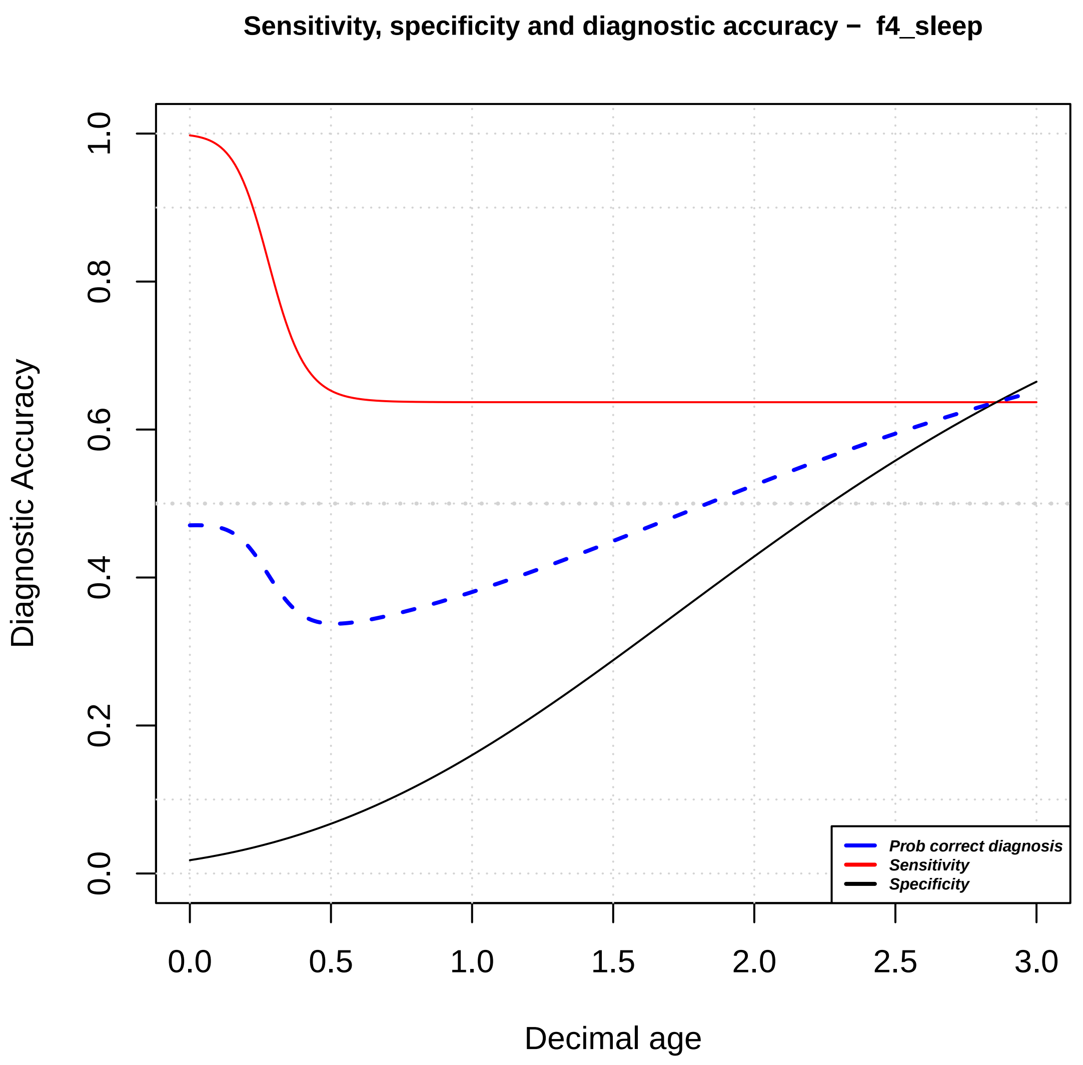

Sensitivity, specificity and diagnostic accuracy – f4_sleep
Diagnostic Accuracy
Decimal age
0.0
0.2
0.4
0.6
0.8
1.0
0.0
0.5
1.0
1.5
2.0
2.5
3.0
Prob correct diagnosis
Sensitivity
Specificity

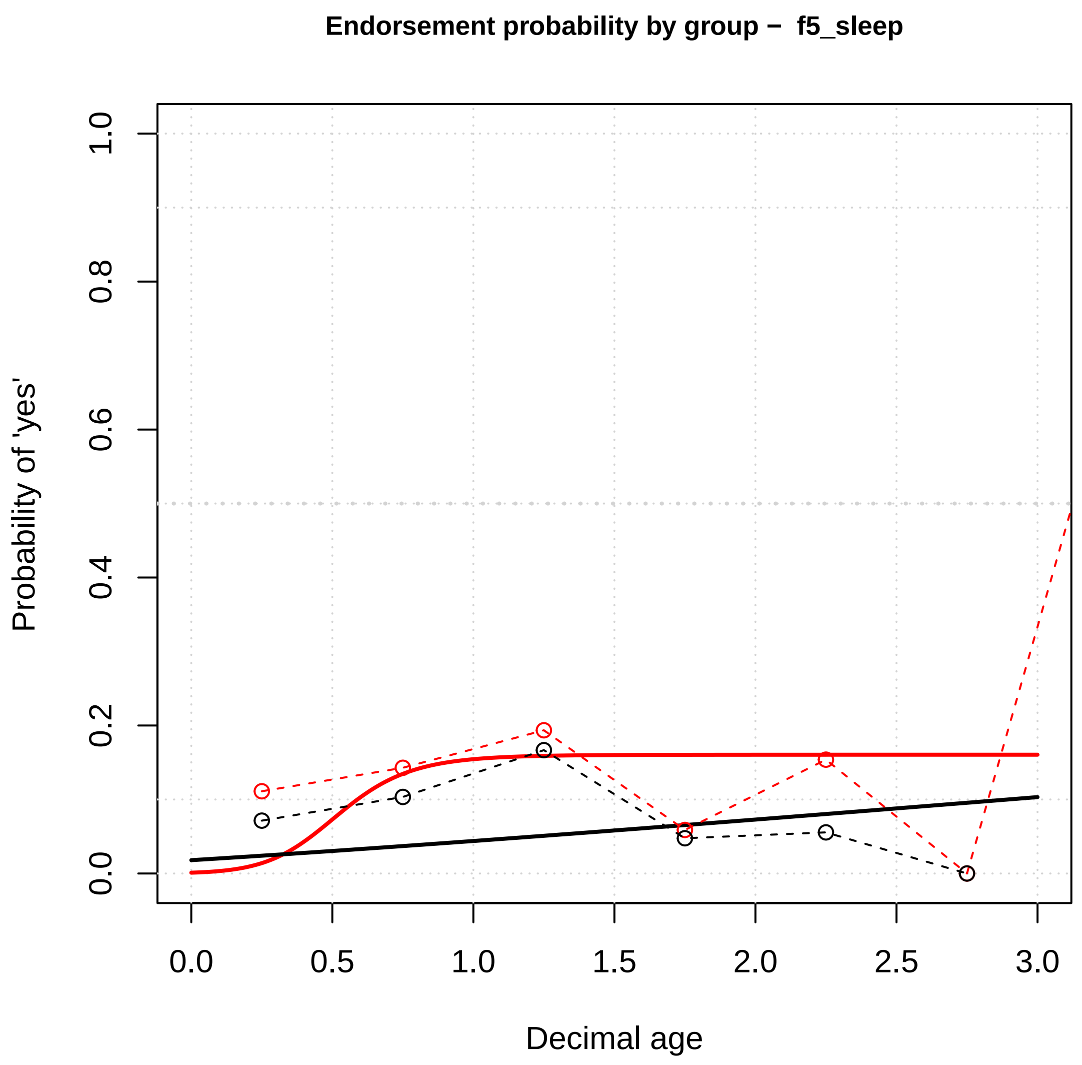

Endorsement probability by group – f5_sleep
Probability of 'yes'
0.0
0.2
0.4
0.6
0.8
1.0
0.0
0.5
1.0
1.5
2.0
2.5
3.0
Decimal age


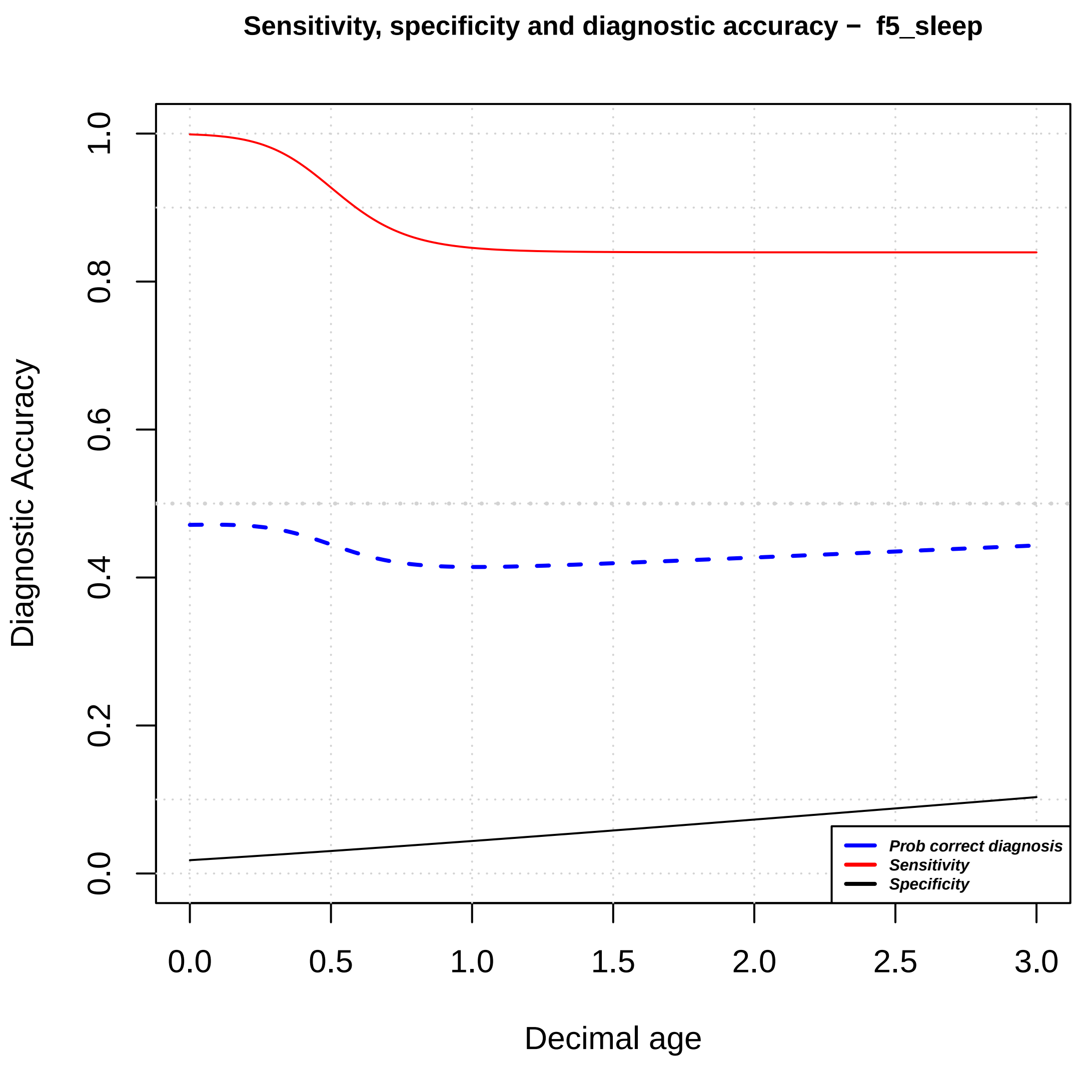

Sensitivity, specificity and diagnostic accuracy – f5_sleep
Diagnostic Accuracy
0.0
0.2
0.4
0.6
0.8
1.0
0.0
0.5
1.0
1.5
2.0
2.5
3.0
Decimal age
Prob correct diagnosis
Sensitivity
Specificity

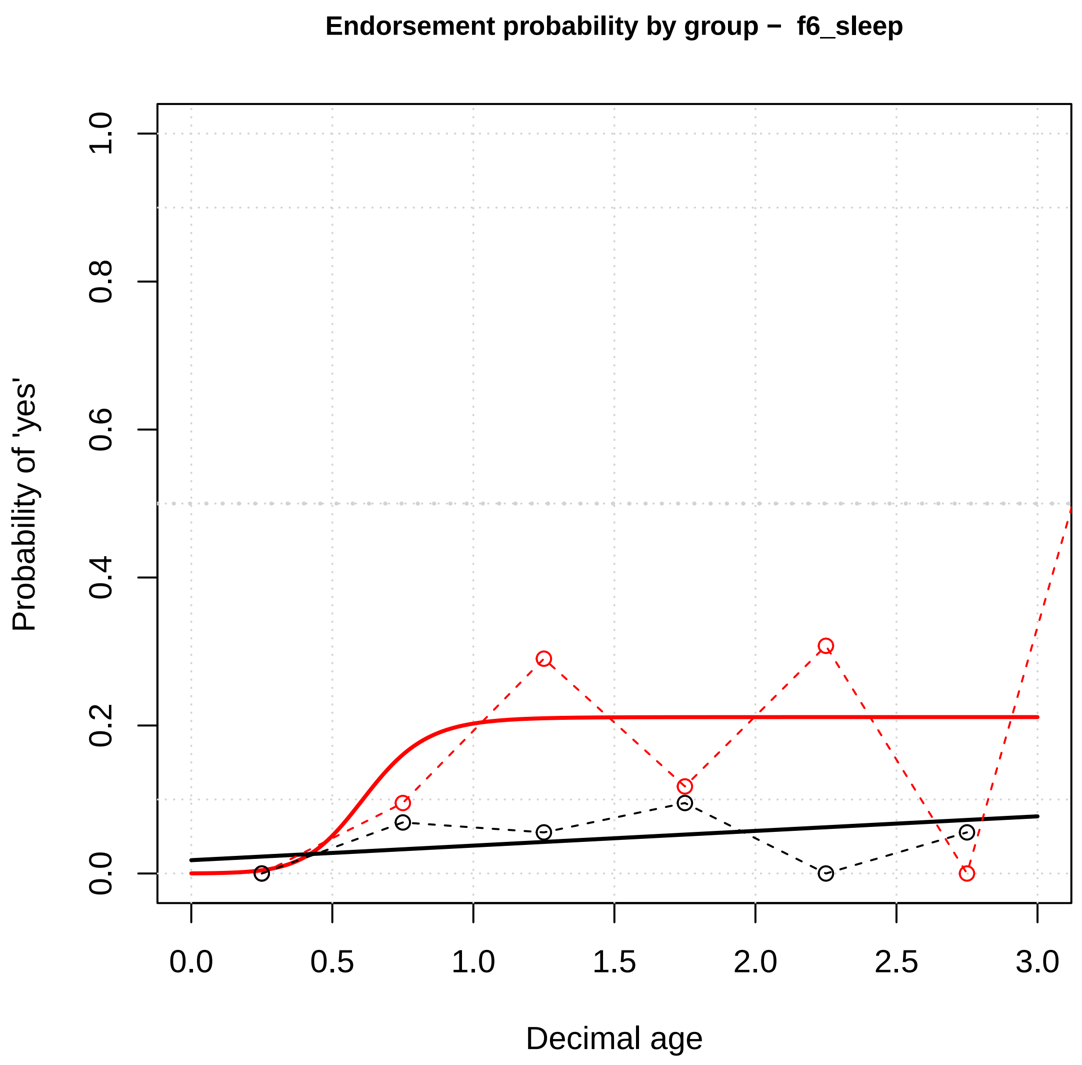

Endorsement probability by group – f6_sleep
Probability of 'yes'
1.0
0.8
0.6
0.4
0.2
0.0
0.0
0.5
1.0
1.5
2.0
2.5
3.0
Decimal age


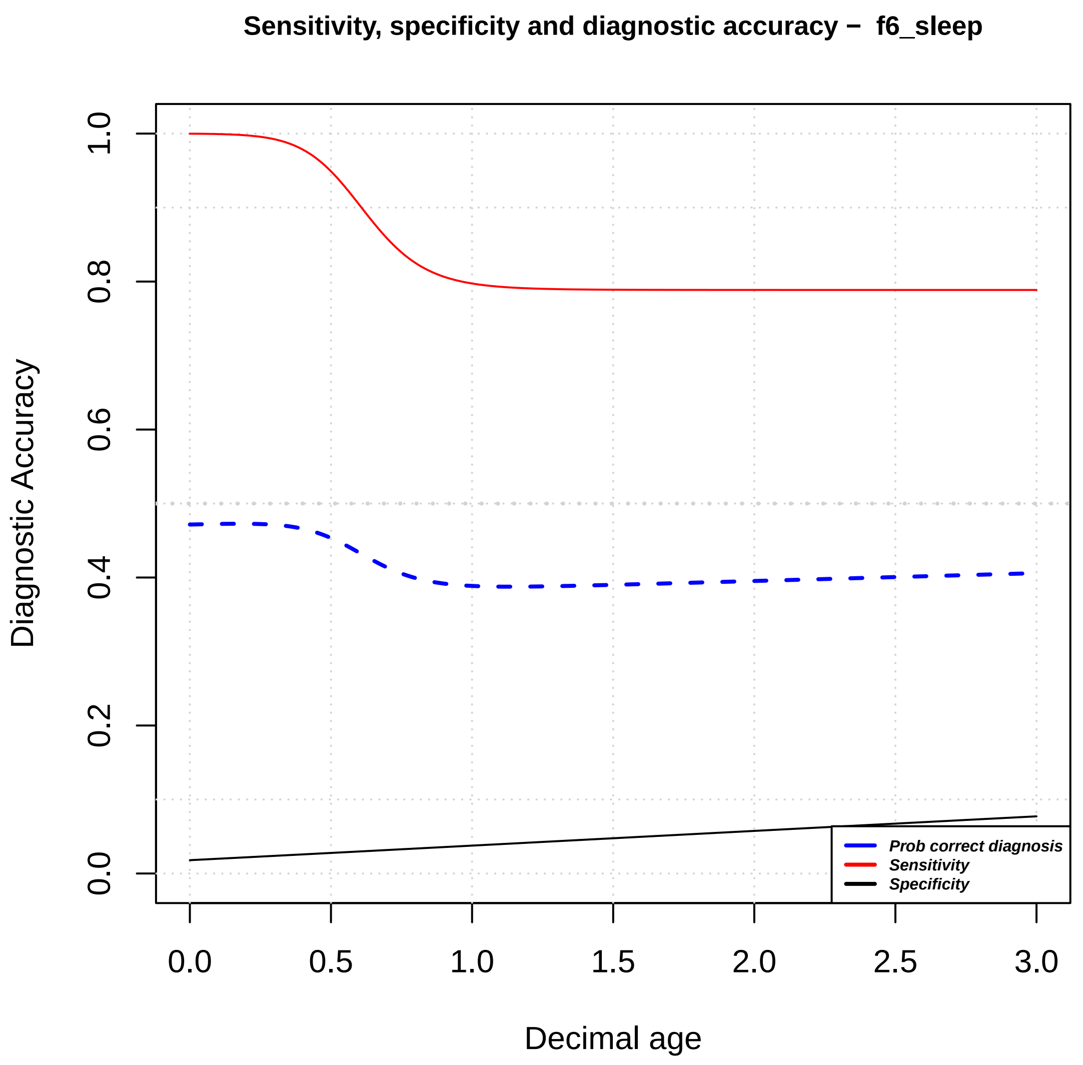

Sensitivity, specificity and diagnostic accuracy – f6_sleep
Diagnostic Accuracy
1.0
0.8
0.6
0.4
0.2
0.0
0.0
0.5
1.0
1.5
2.0
2.5
3.0
Decimal age
Prob correct diagnosis
Sensitivity
Specificity

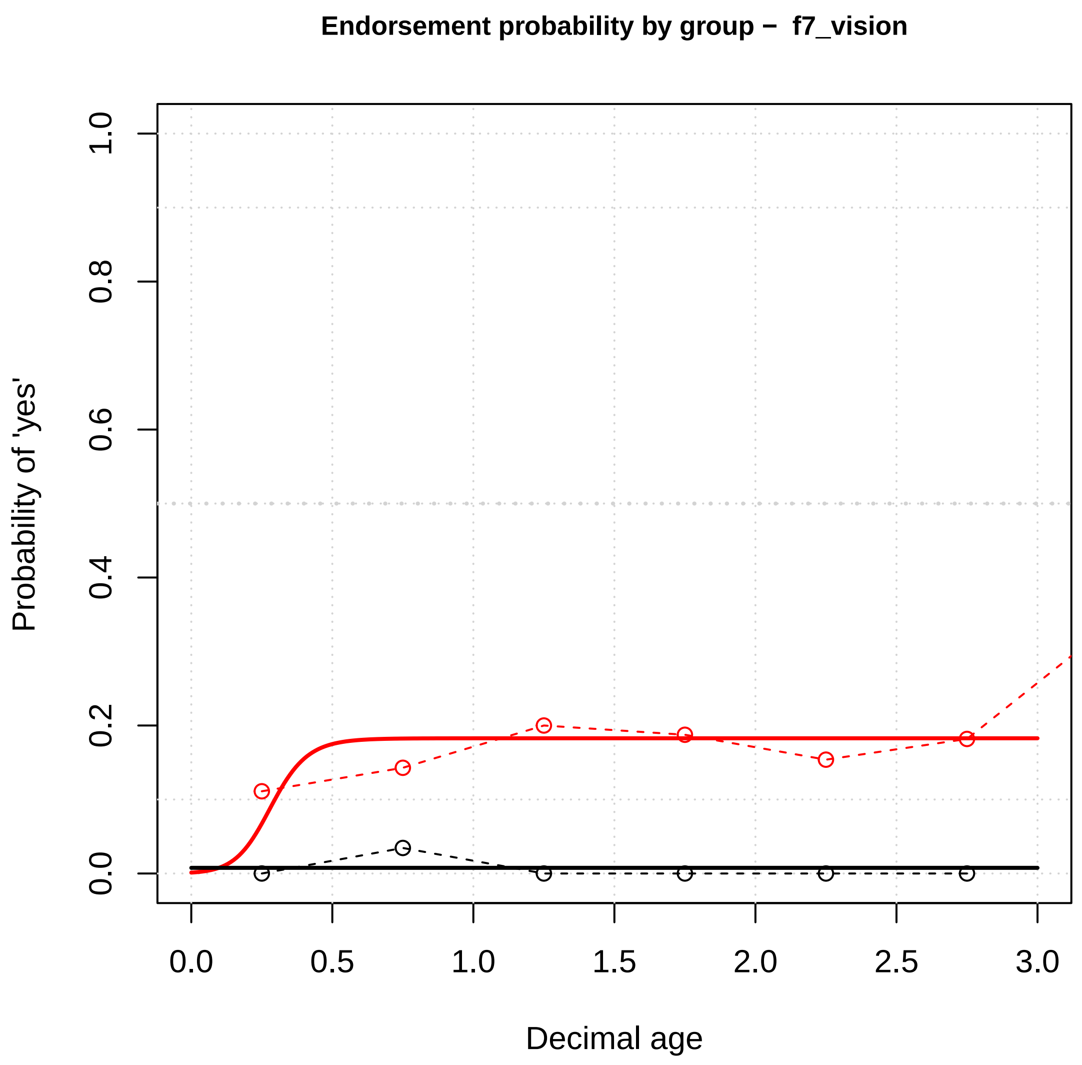

Endorsement probability by group – f7_vision
Probability of 'yes'
1.0
0.8
0.6
0.4
0.2
0.0
0.0
0.5
1.0
1.5
2.0
2.5
3.0
Decimal age


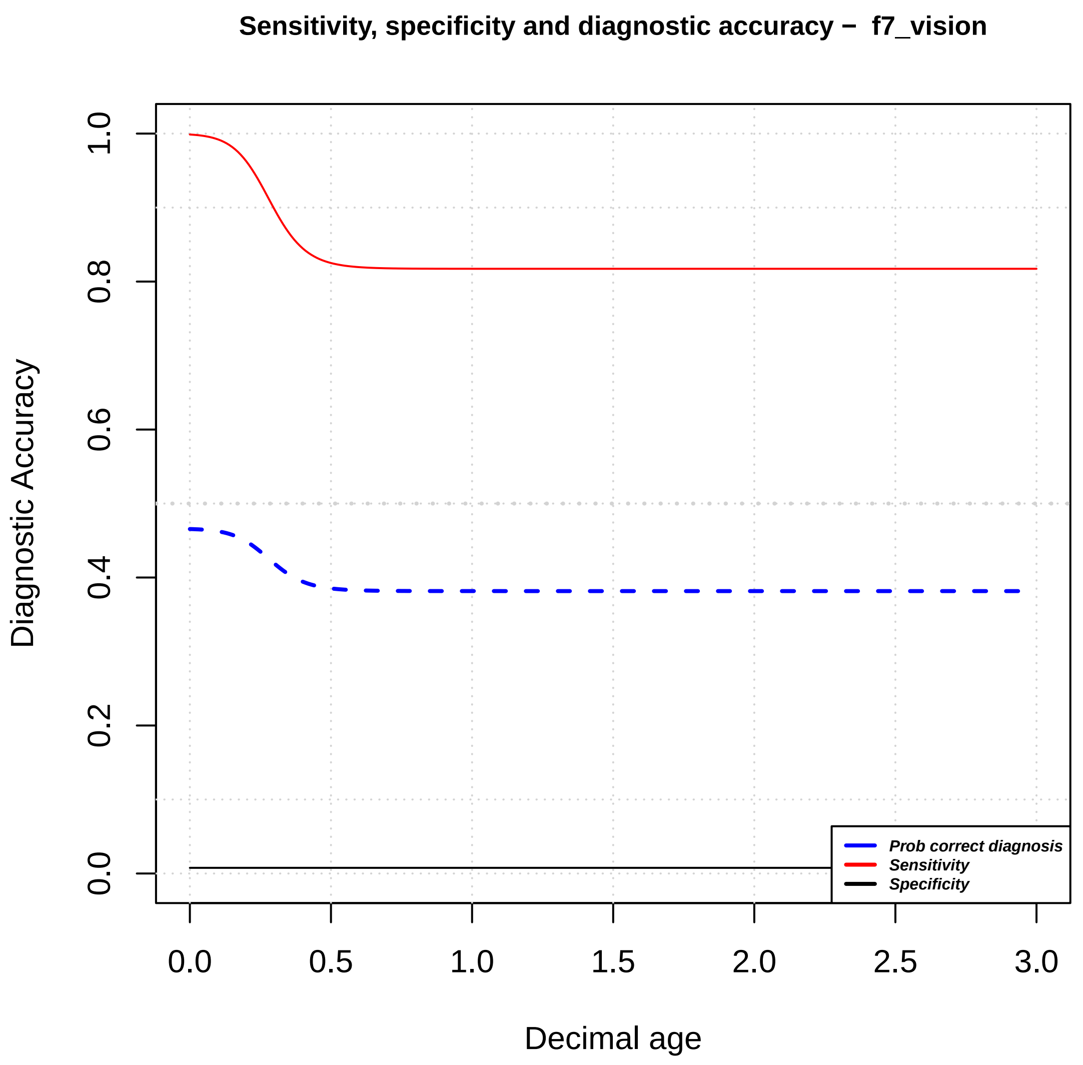

Sensitivity, specificity and diagnostic accuracy – f7_vision
Diagnostic Accuracy
1.0
0.8
0.6
0.4
0.2
0.0
0.0
0.5
1.0
1.5
2.0
2.5
3.0
Decimal age
Prob correct diagnosis
Sensitivity
Specificity

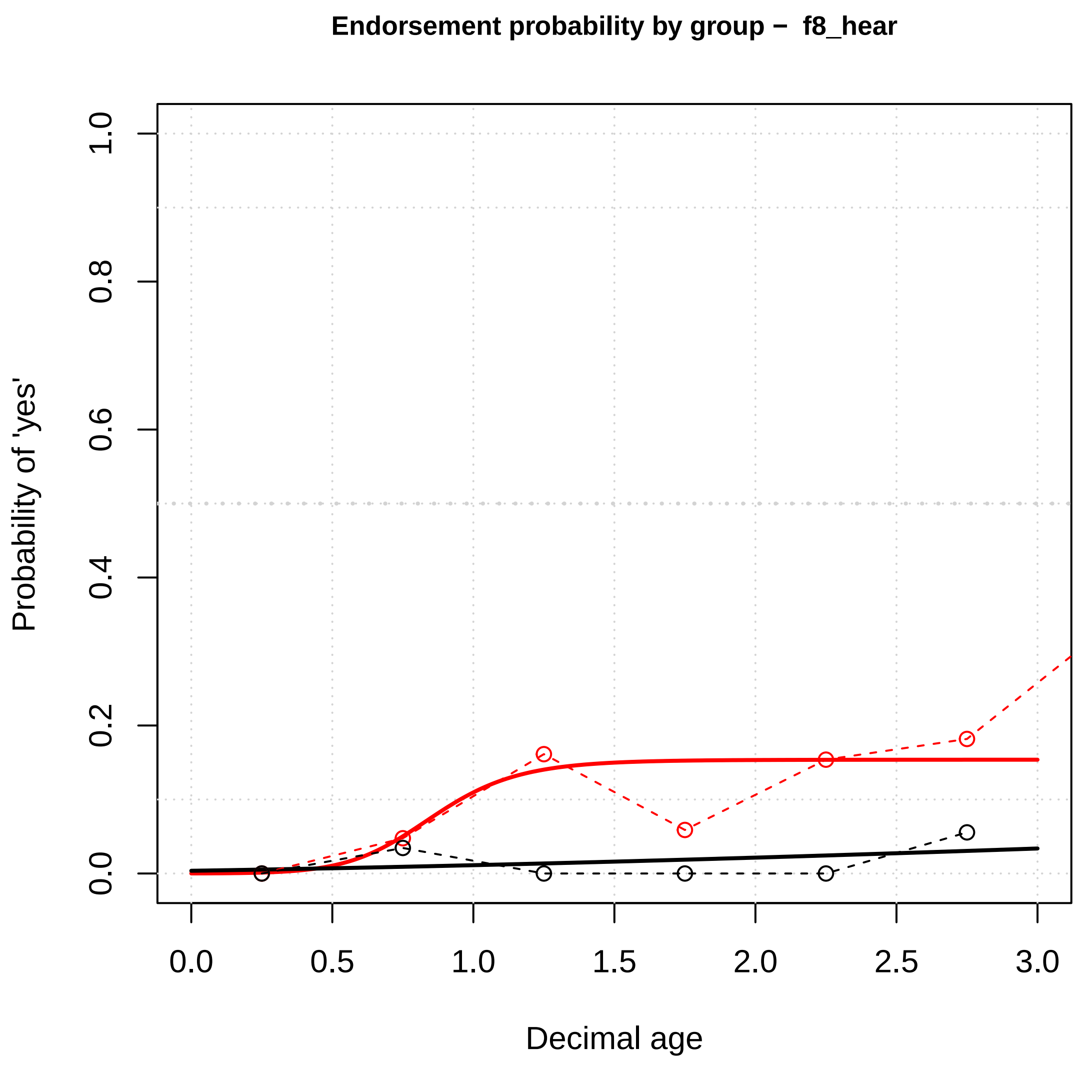

Endorsement probability by group – f8_hear
Probability of 'yes'
Decimal age
0.0
0.2
0.4
0.6
0.8
1.0
0.0
0.5
1.0
1.5
2.0
2.5
3.0


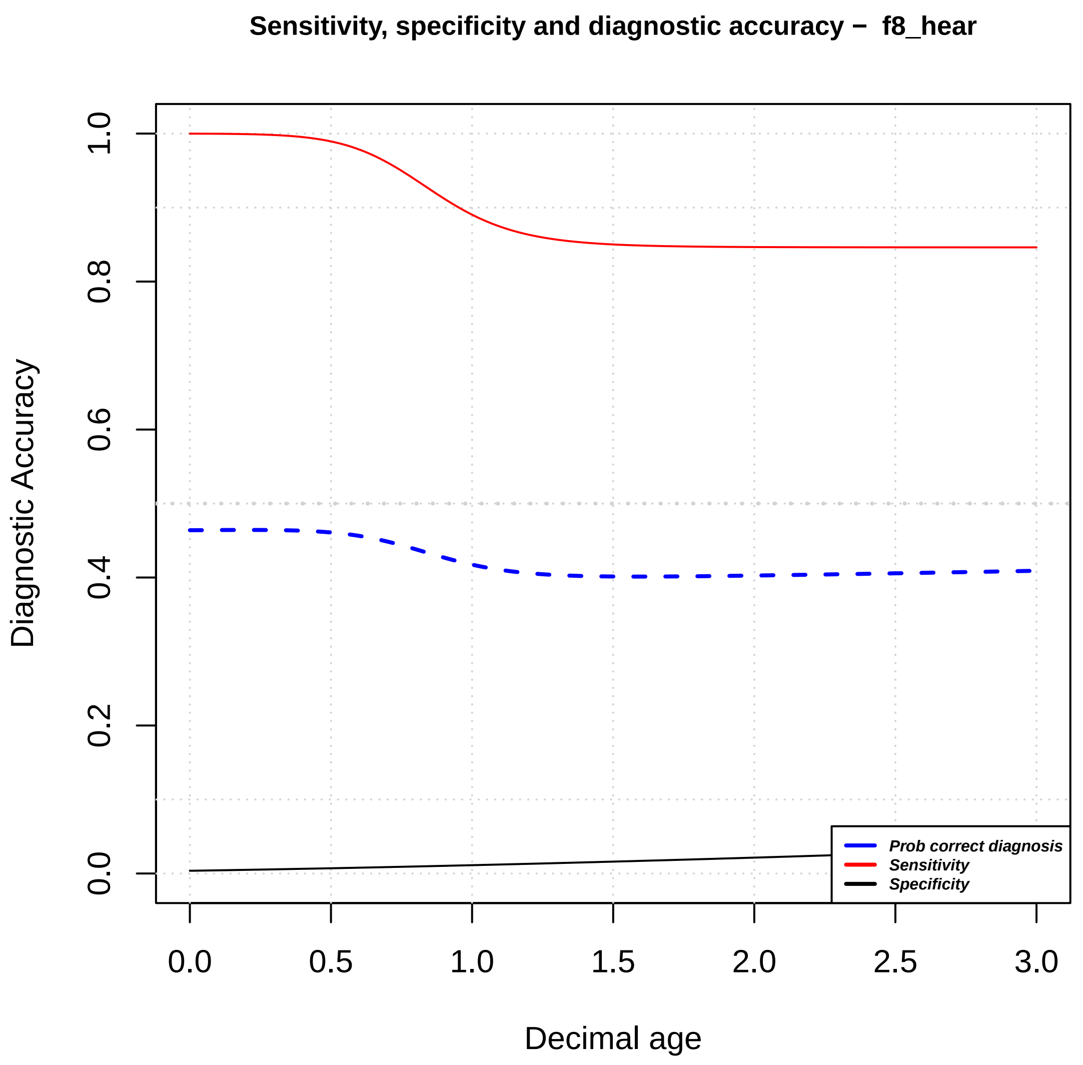

Sensitivity, specificity and diagnostic accuracy – f8_hear
Diagnostic Accuracy
Decimal age
0.0
0.2
0.4
0.6
0.8
1.0
0.0
0.5
1.0
1.5
2.0
2.5
3.0
Prob correct diagnosis
Sensitivity
Specificity

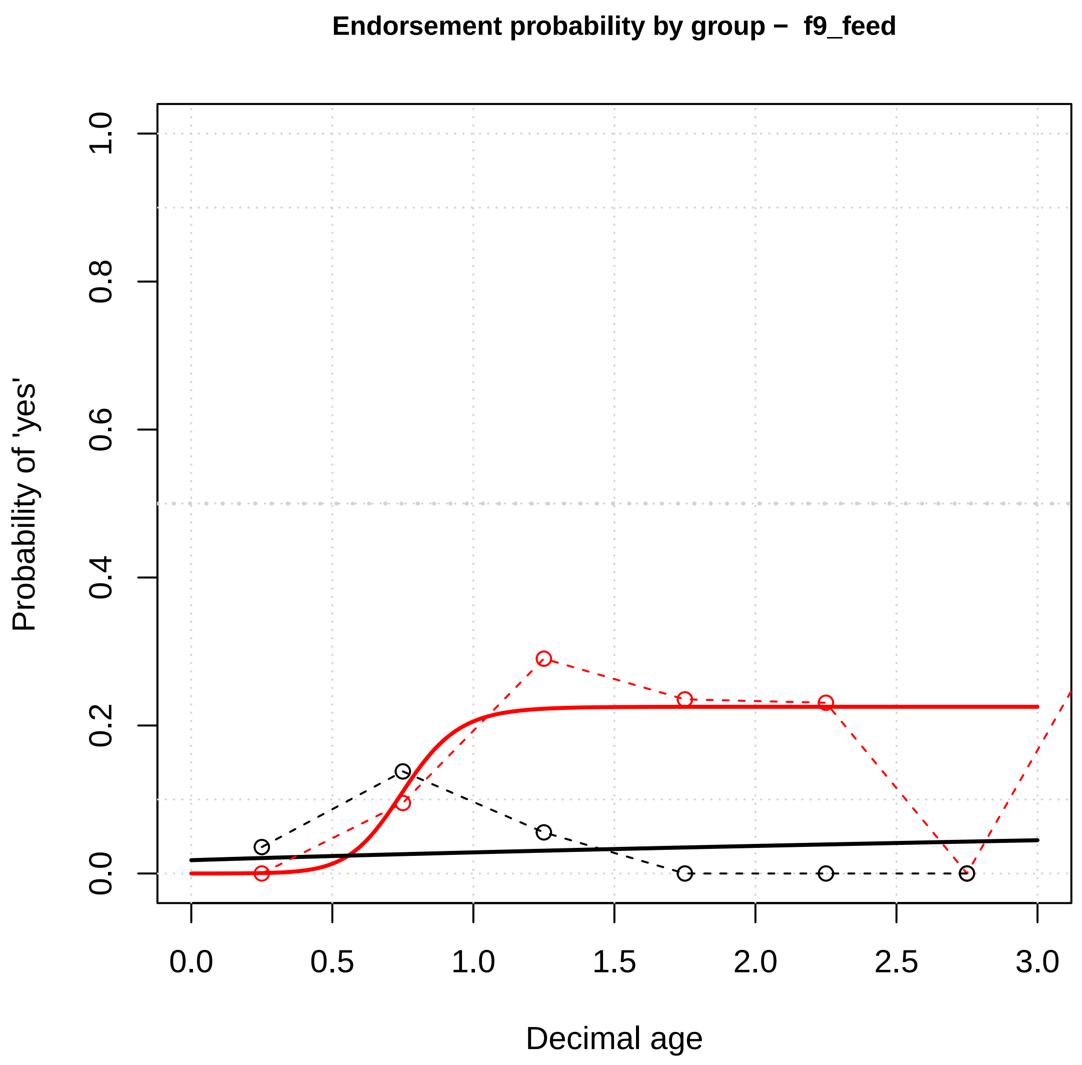

Endorsement probability by group – f9_feed
Probability of 'yes'
Decimal age
0.0
0.2
0.4
0.6
0.8
1.0
0.0
0.5
1.0
1.5
2.0
2.5
3.0


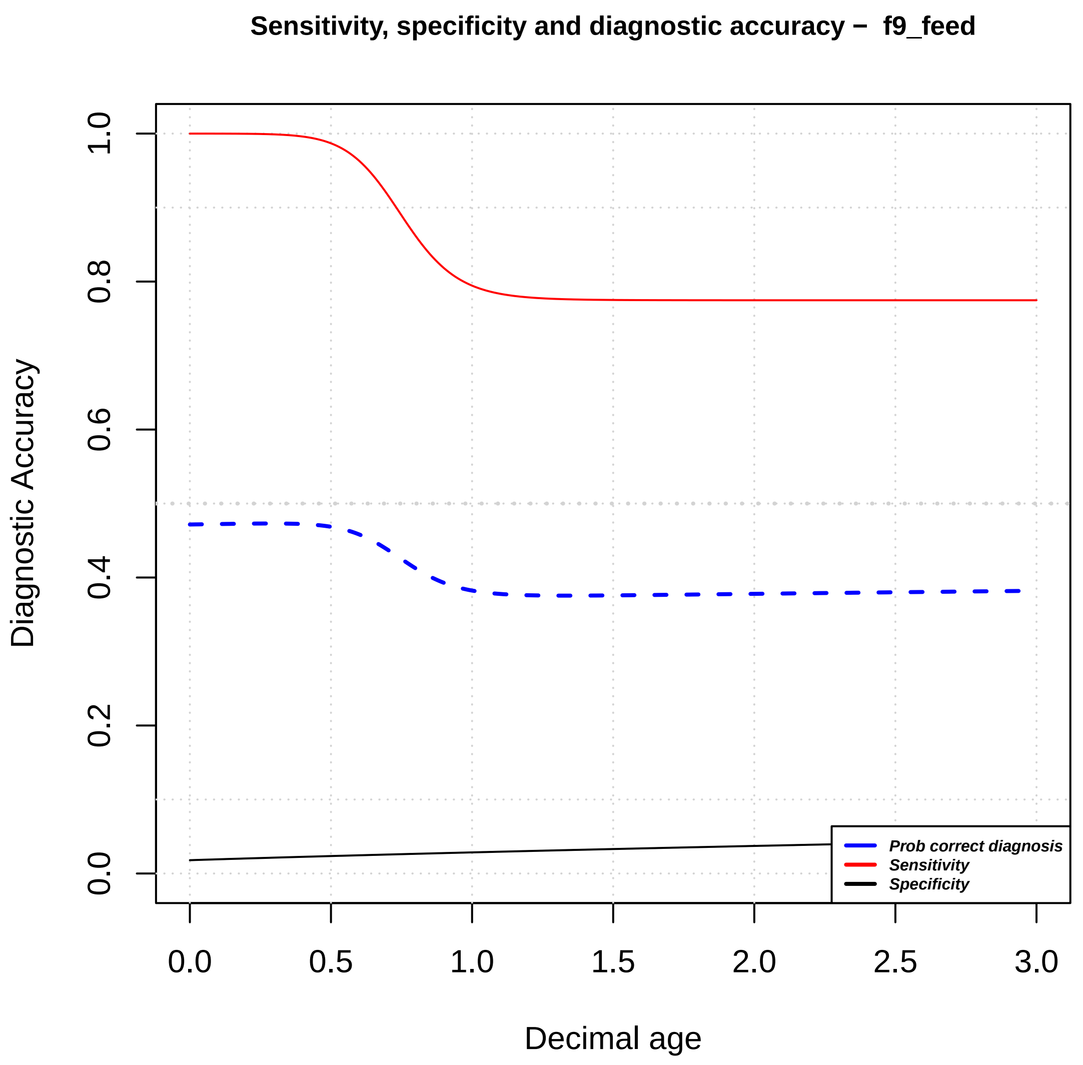

Sensitivity, specificity and diagnostic accuracy – f9_feed
Diagnostic Accuracy
Decimal age
0.0
0.2
0.4
0.6
0.8
1.0
0.0
0.5
1.0
1.5
2.0
2.5
3.0
Prob correct diagnosis
Sensitivity
Specificity

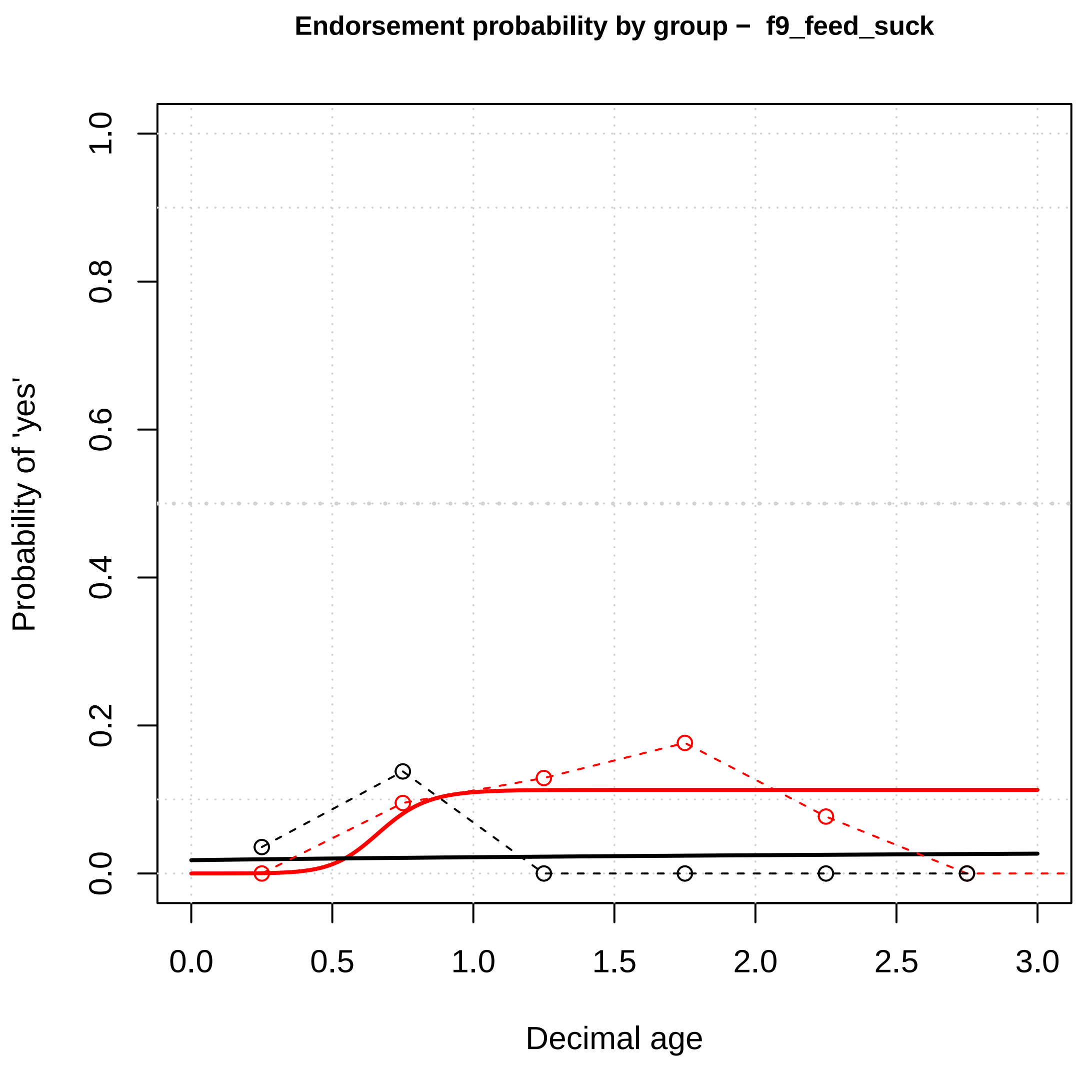
Endorsement probability by group – f9_feed_suck
Probability of 'yes'
Decimal age
0.0
0.2
0.4
0.6
0.8
1.0
0.5
1.5
2.0
2.5
3.0

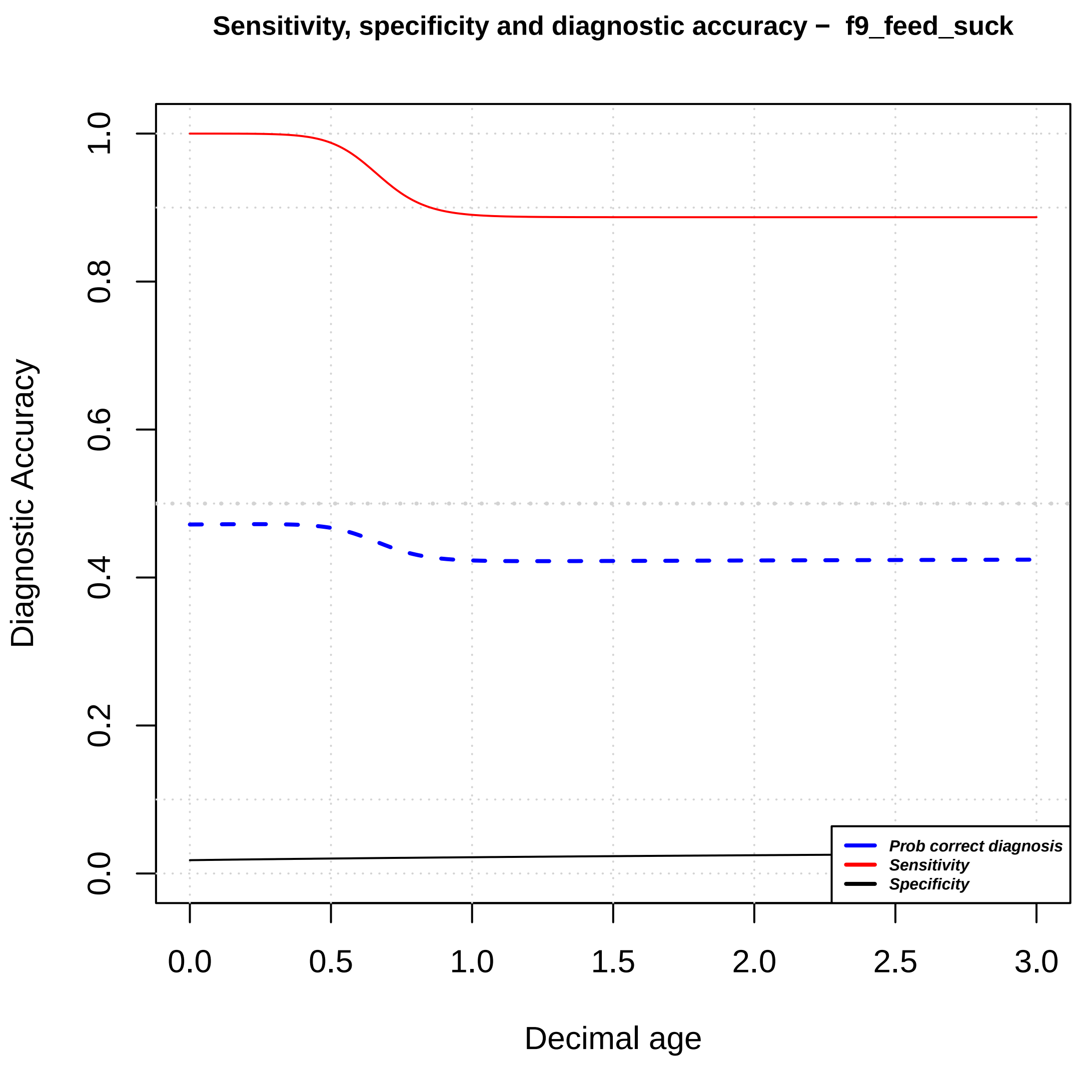
Sensitivity, specificity and diagnostic accuracy – f9_feed_suck
Diagnostic Accuracy
Decimal age
Prob correct diagnosis
Sensitivity
Specificity
0.0
0.2
0.4
0.6
0.8
1.0
0.5
1.5
2.0
2.5
3.0

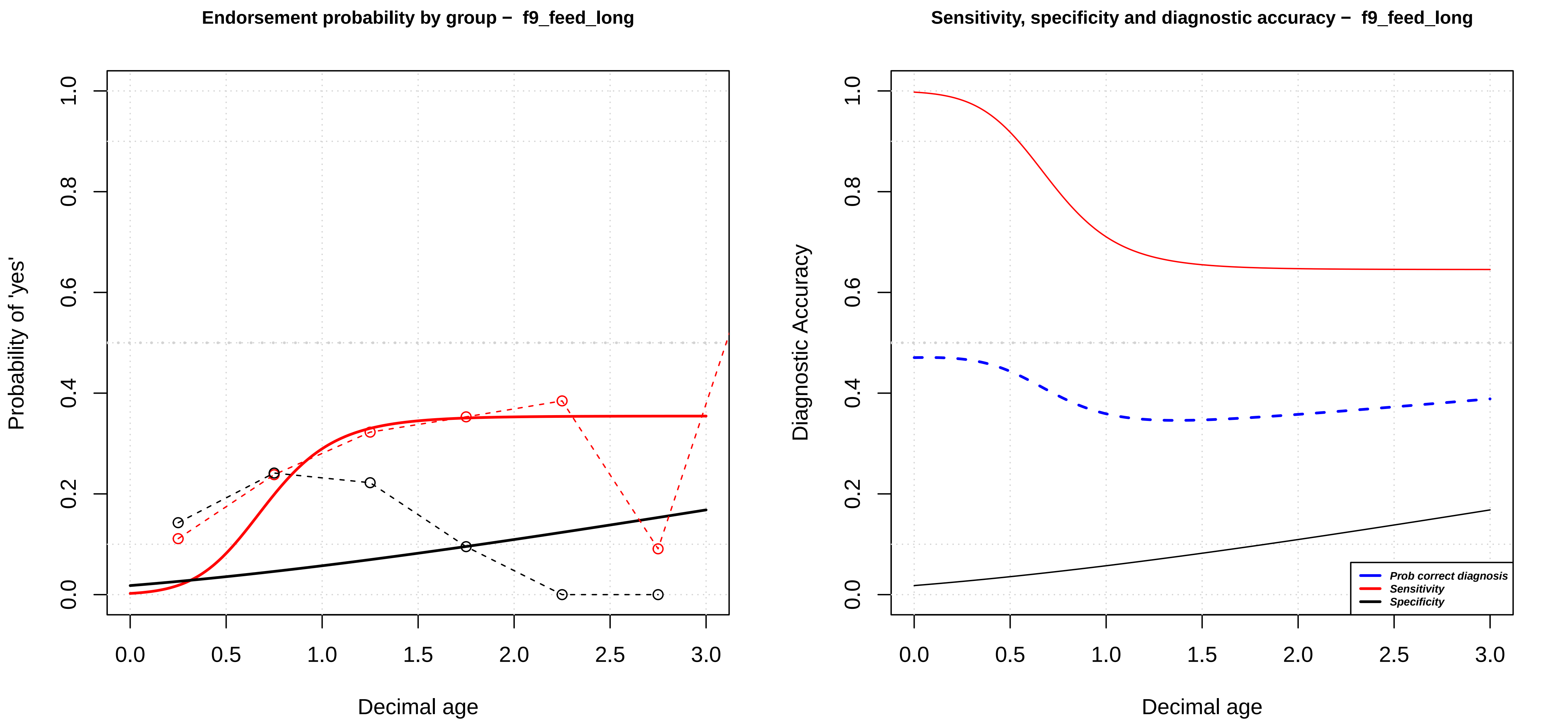

Endorsement probability by group – f9_feed_long
Probability of 'yes'
0.0
0.2
0.4
0.6
0.8
1.0
0.0
0.5
1.0
1.5
2.0
2.5
3.0
Decimal age
Sensitivity, specificity and diagnostic accuracy – f9_feed_long
Diagnostic Accuracy
0.0
0.2
0.4
0.6
0.8
1.0
0.0
0.5
1.0
1.5
2.0
2.5
3.0
Decimal age
Prob correct diagnosis
Sensitivity
Specificity

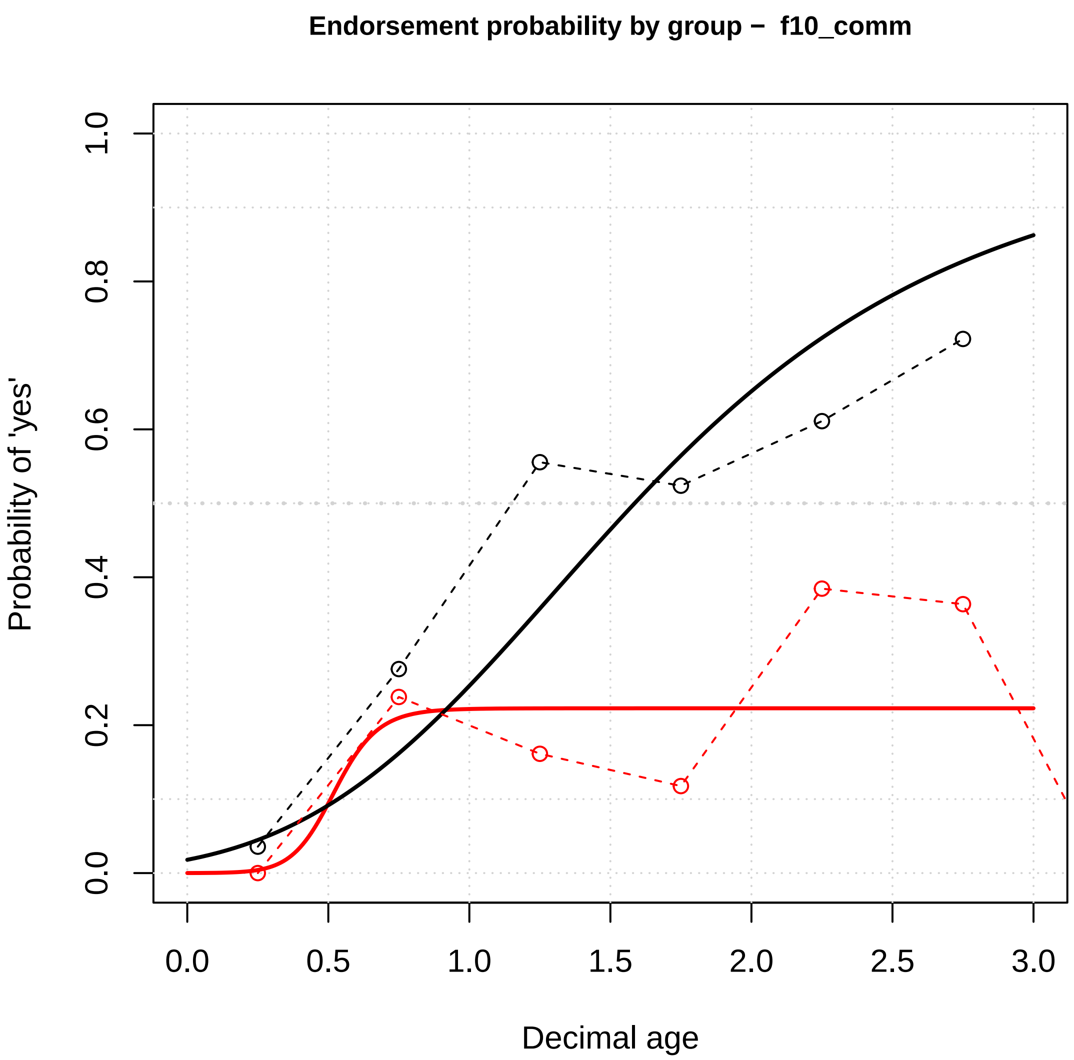

Endorsement probability by group − f10_comm
Probability of 'yes'
Decimal age
0.0
0.5
1.0
1.5
2.0
2.5
3.0
0.0
0.2
0.4
0.6
0.8
1.0


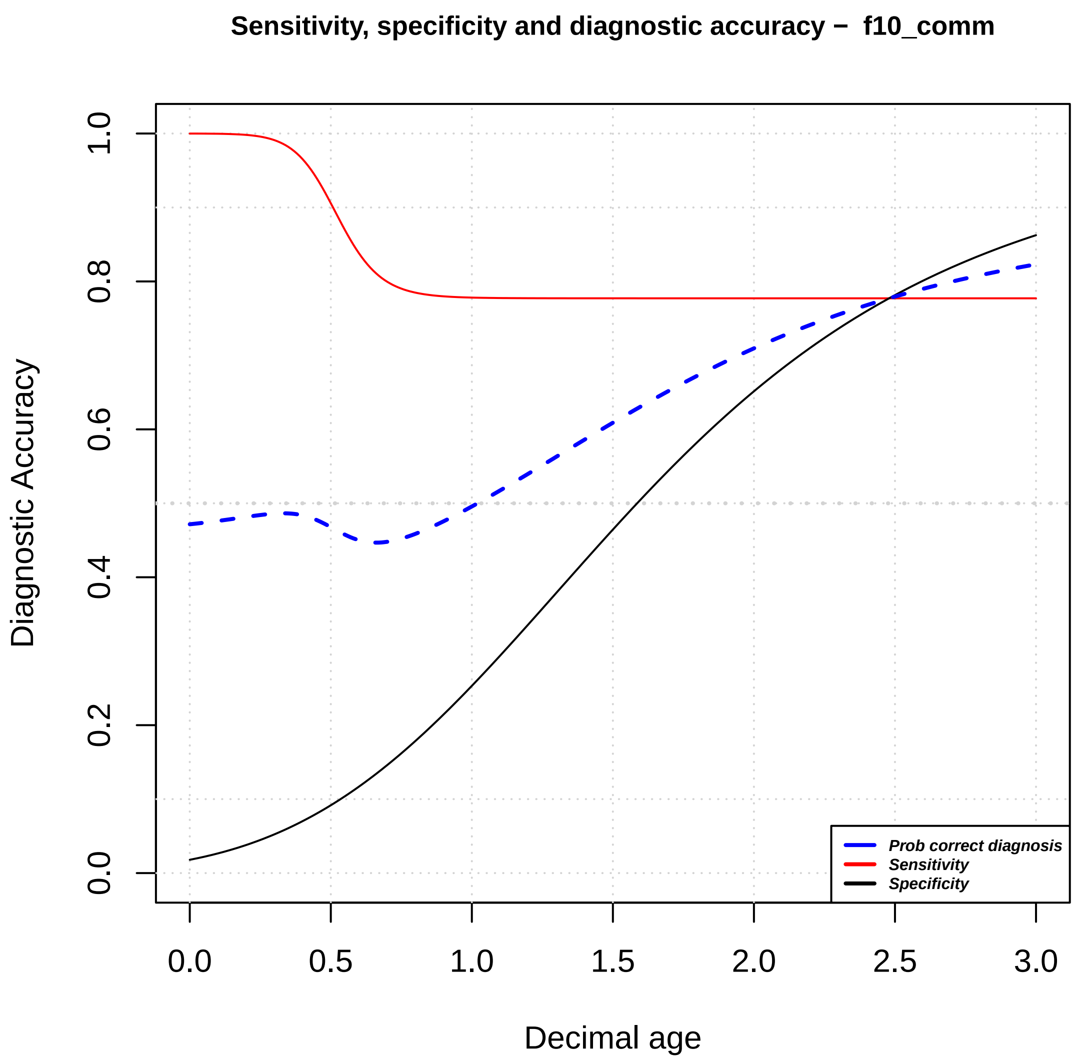

Sensitivity, specificity and diagnostic accuracy − f10_comm
Diagnostic Accuracy
Decimal age
Prob correct diagnosis
Sensitivity
Specificity
0.0
0.5
1.0
1.5
2.0
2.5
3.0
0.0
0.2
0.4
0.6
0.8
1.0

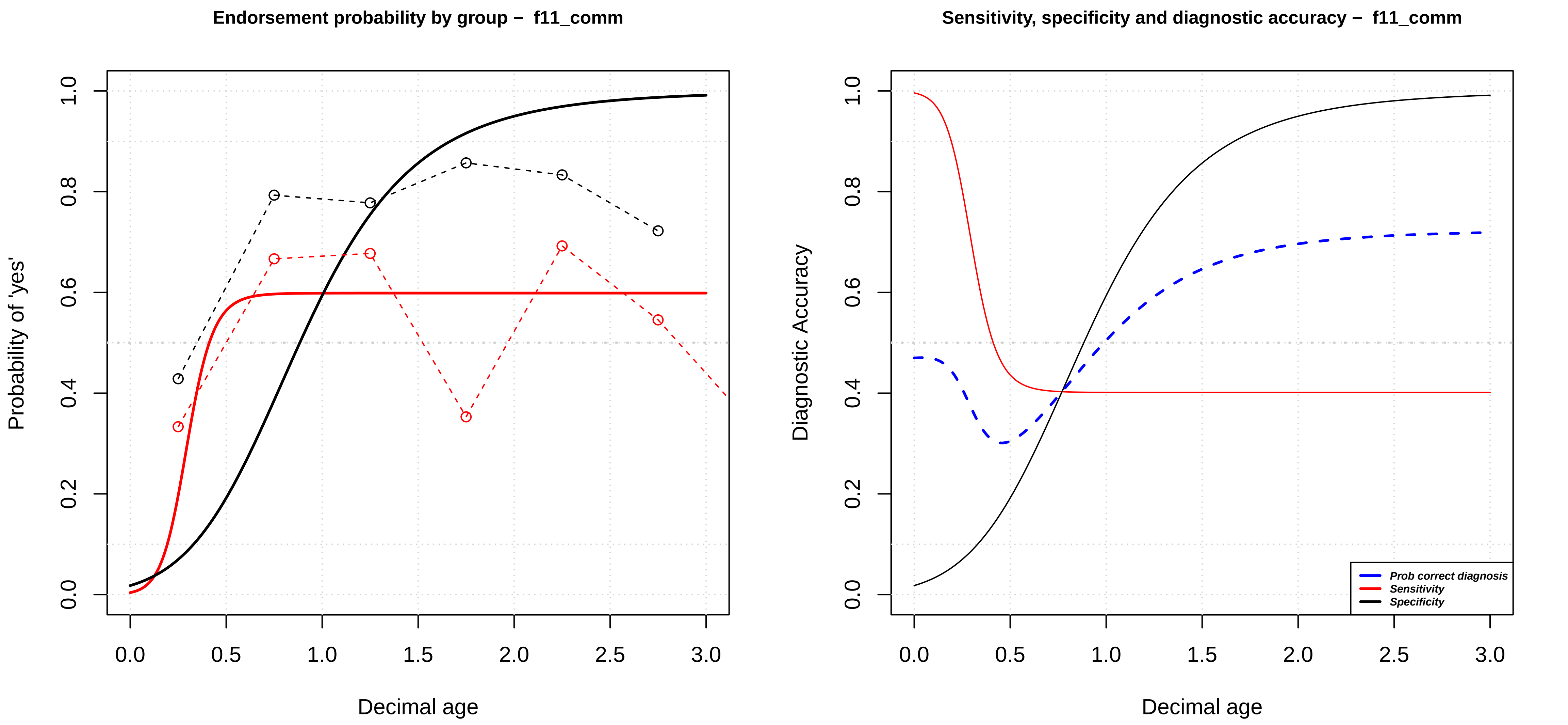

Endorsement probability by group – f11_comm
Probability of 'yes'
Decimal age
0.0
0.2
0.4
0.6
0.8
1.0
0.0
0.5
1.0
1.5
2.0
2.5
3.0
Sensitivity, specificity and diagnostic accuracy – f11_comm
Diagnostic Accuracy
Decimal age
Prob correct diagnosis
Sensitivity
Specificity

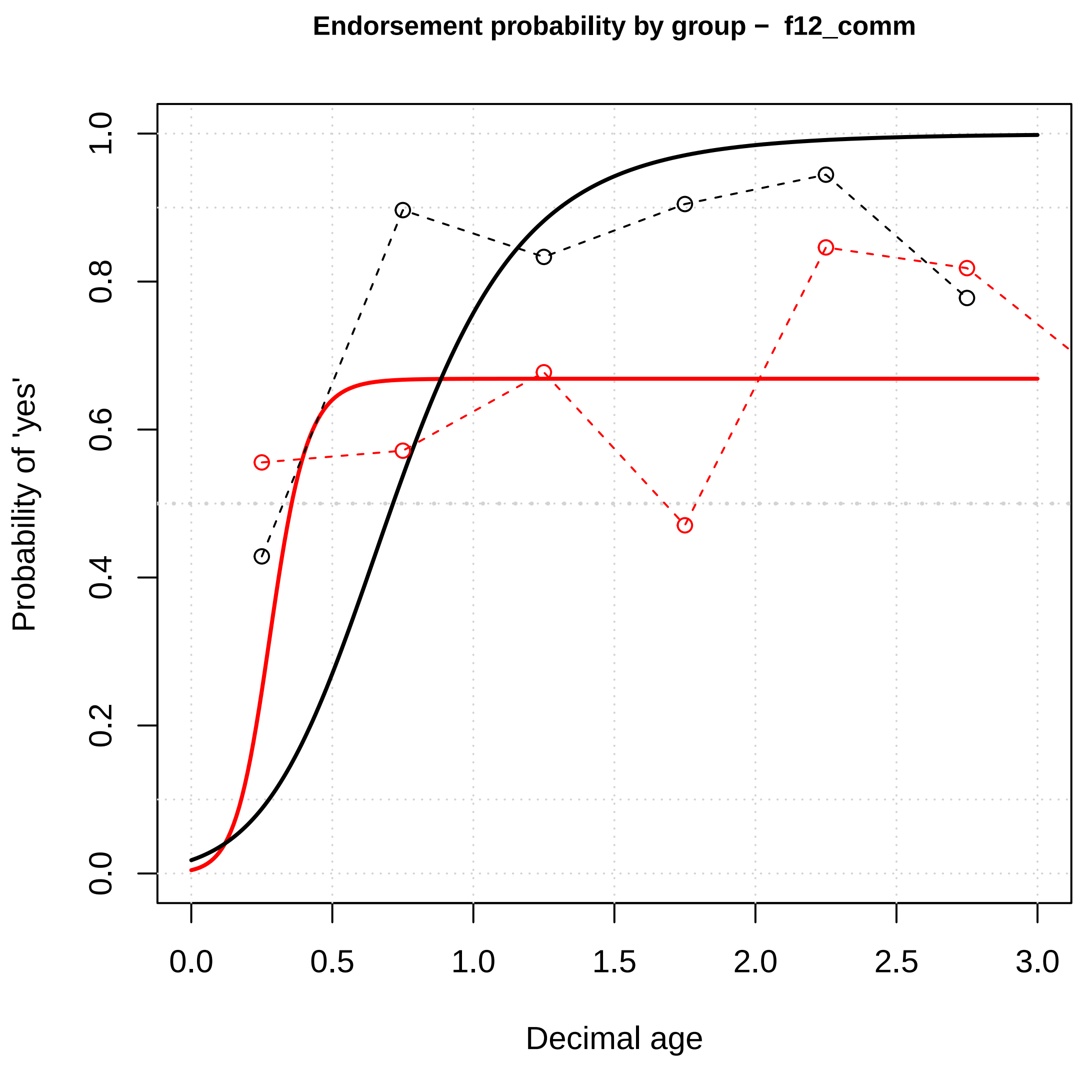

Endorsement probability by group – f12_comm
Probability of 'yes'
Decimal age
0.0
0.2
0.4
0.6
0.8
1.0
0.0
0.5
1.0
1.5
2.0
2.5
3.0


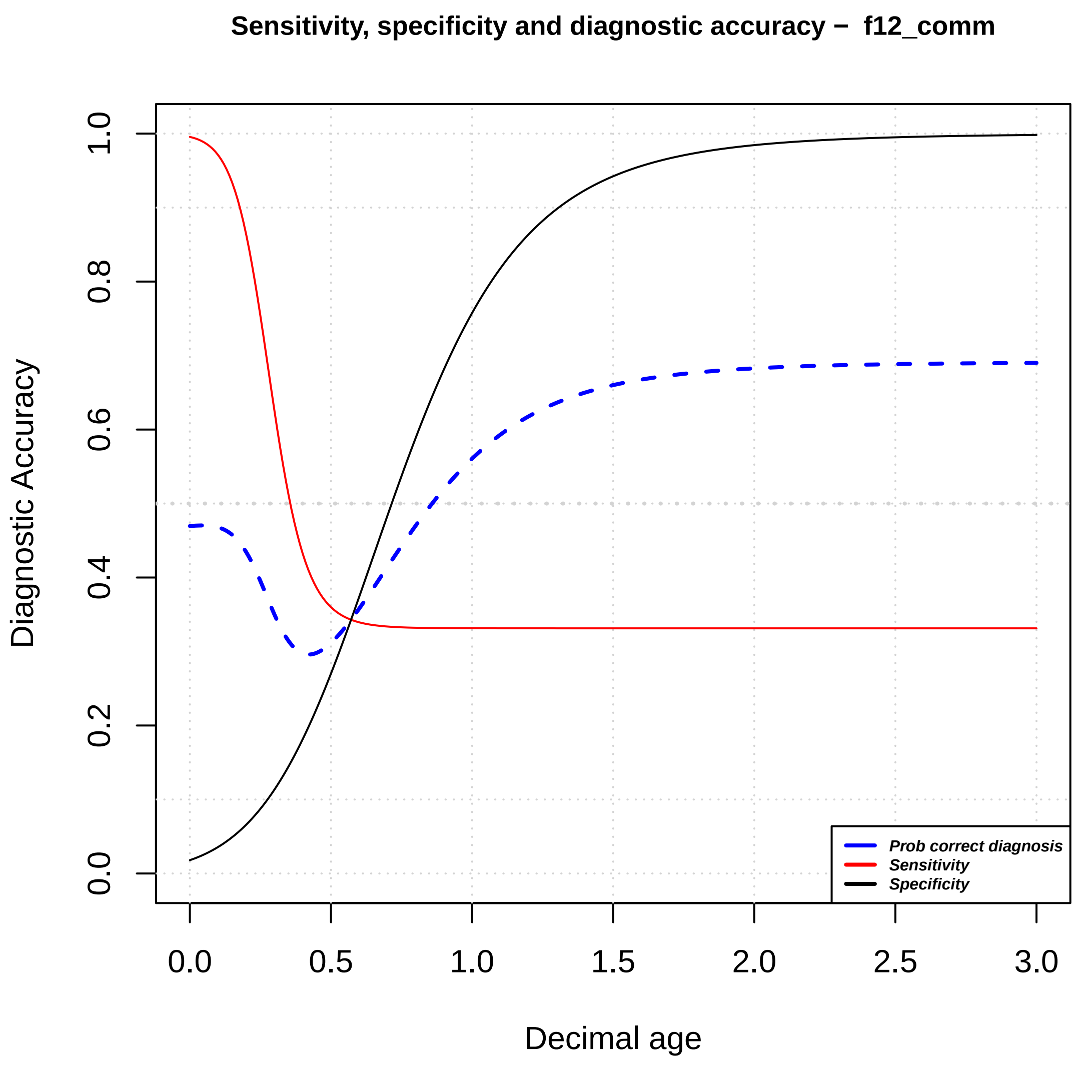

Sensitivity, specificity and diagnostic accuracy – f12_comm
Diagnostic Accuracy
Decimal age
0.0
0.2
0.4
0.6
0.8
1.0
0.0
0.5
1.0
1.5
2.0
2.5
3.0
Prob correct diagnosis
Sensitivity
Specificity

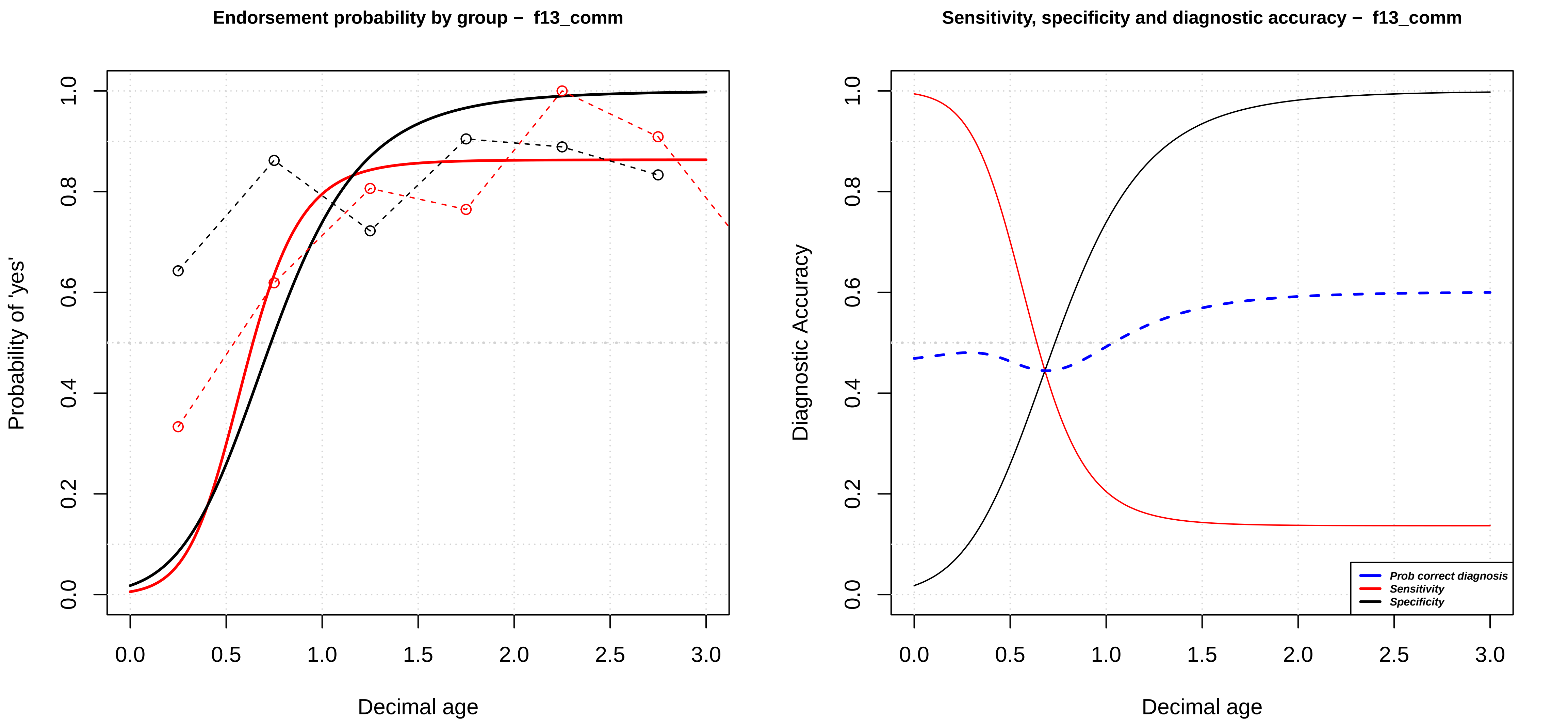
Endorsement probability by group – f13_comm
Probability of 'yes'
Decimal age
0.0
0.2
0.4
0.6
0.8
1.0
0.0
0.5
1.0
1.5
2.0
2.5
3.0
Sensitivity, specificity and diagnostic accuracy – f13_comm
Diagnostic Accuracy
Decimal age
Prob correct diagnosis
Sensitivity
Specificity

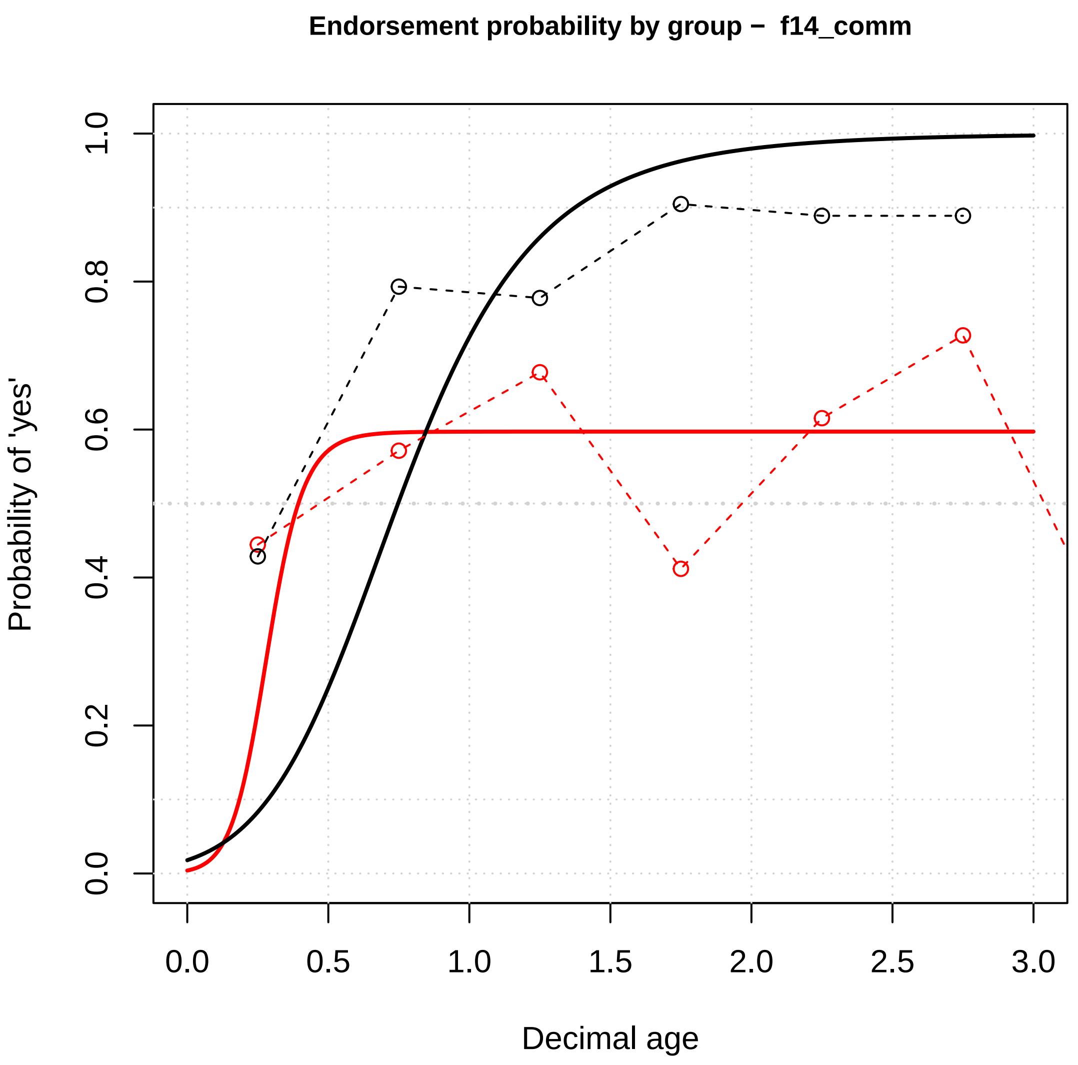
Endorsement probability by group – f14_comm
Probability of 'yes'
Decimal age
0.0
0.2
0.4
0.6
0.8
1.0
0.0
0.5
1.0
1.5
2.0
2.5
3.0

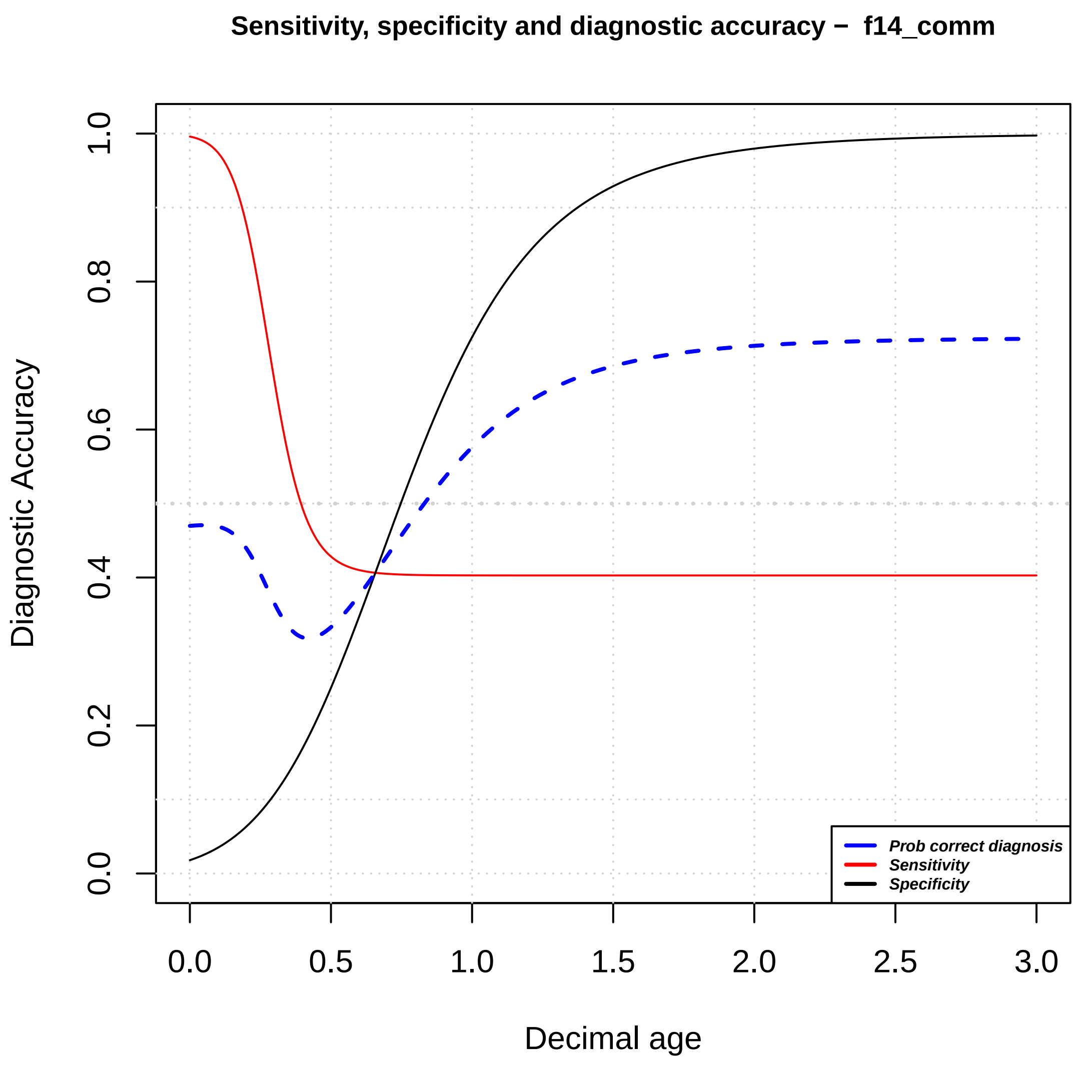
Sensitivity, specificity and diagnostic accuracy – f14_comm
Diagnostic Accuracy
Decimal age
0.0
0.2
0.4
0.6
0.8
1.0
0.0
0.5
1.0
1.5
2.0
2.5
3.0
Prob correct diagnosis
Sensitivity
Specificity

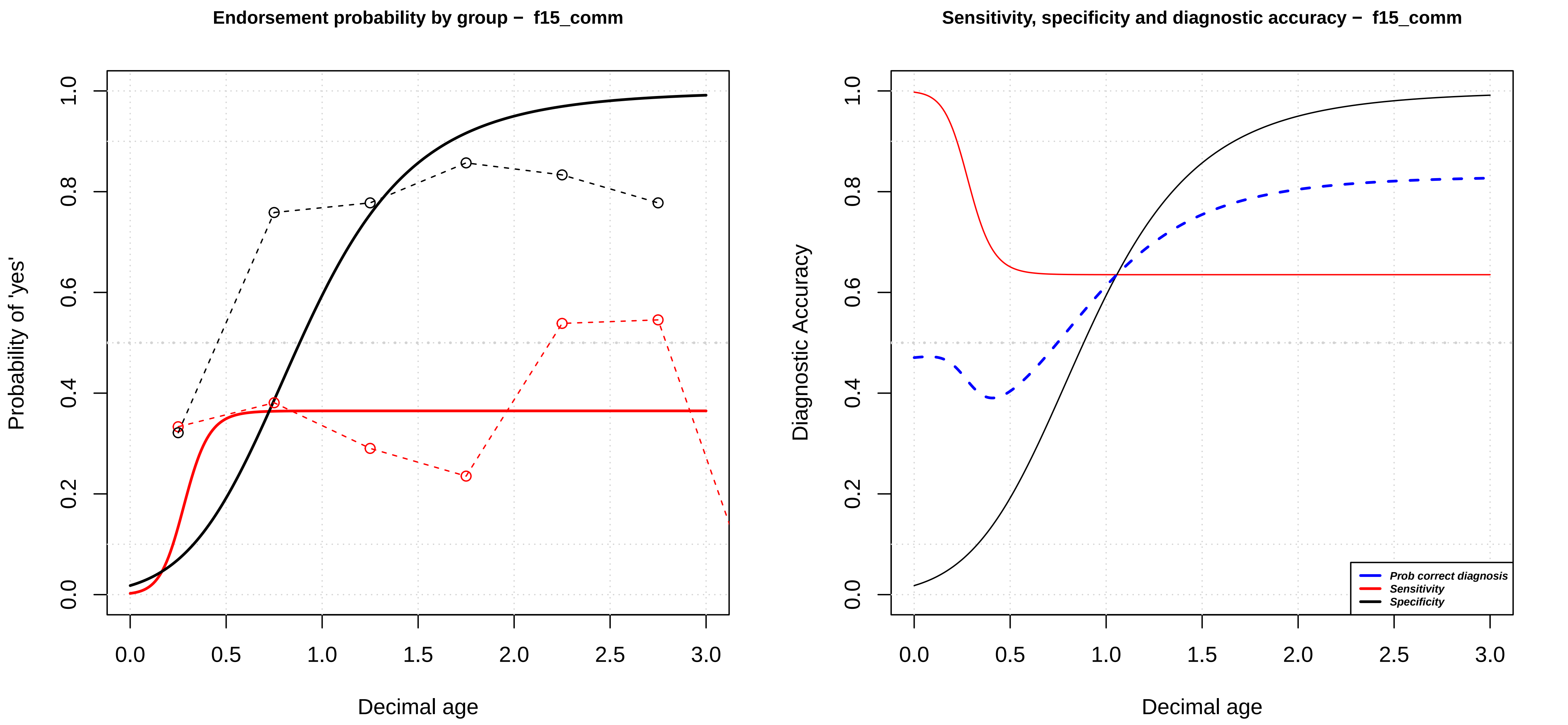

Endorsement probability by group − f15_comm
Probability of 'yes'
Decimal age
0.0
0.2
0.4
0.6
0.8
1.0
0.0
0.5
1.0
1.5
2.0
2.5
3.0
Sensitivity, specificity and diagnostic accuracy − f15_comm
Diagnostic Accuracy
Decimal age
Prob correct diagnosis
Sensitivity
Specificity

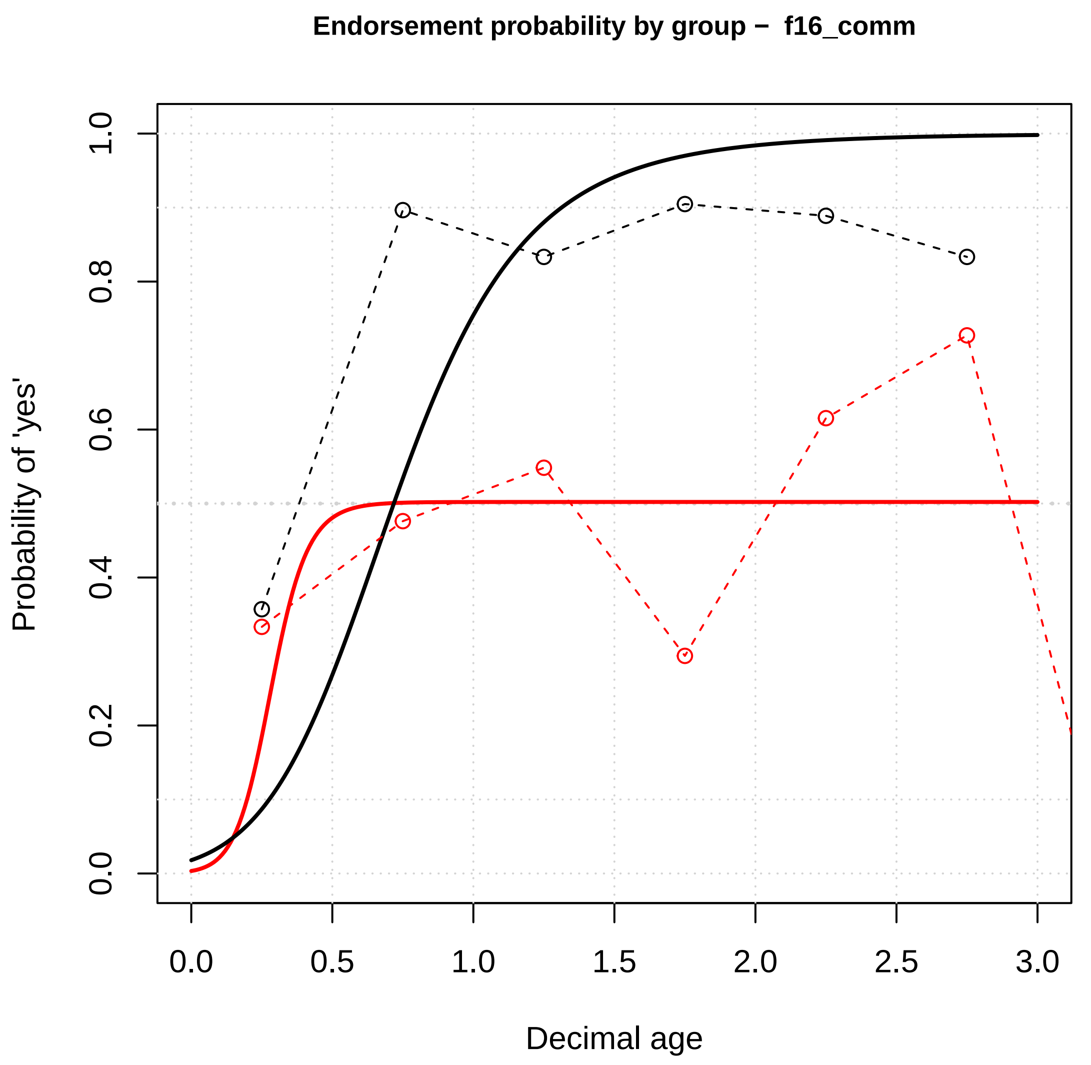

Endorsement probability by group – f16_comm
Probability of 'yes'
Decimal age
0.0
0.2
0.4
0.6
0.8
1.0
0.0
0.5
1.0
1.5
2.0
2.5
3.0


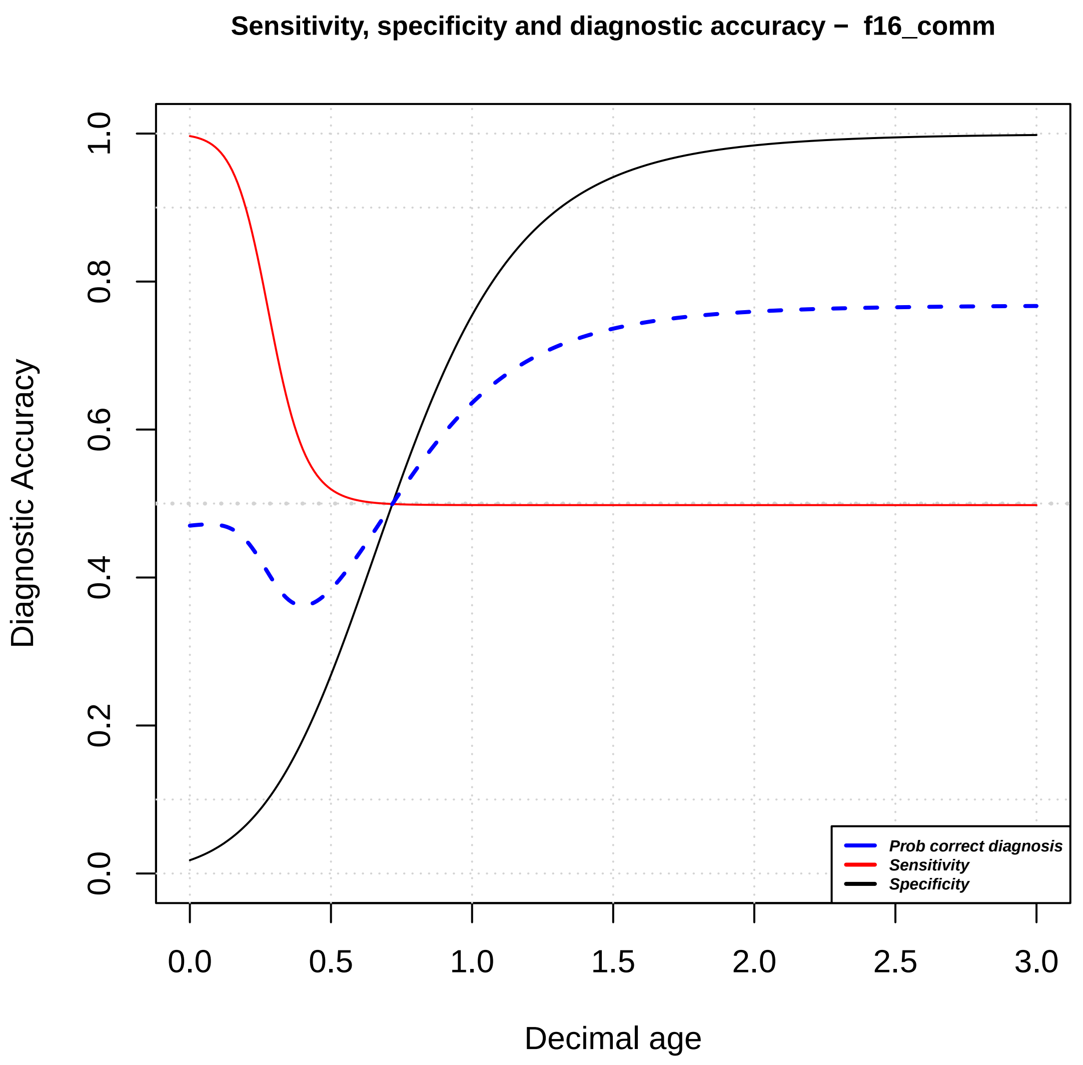

Sensitivity, specificity and diagnostic accuracy – f16_comm
Diagnostic Accuracy
Decimal age
0.0
0.2
0.4
0.6
0.8
1.0
0.0
0.5
1.0
1.5
2.0
2.5
3.0
Prob correct diagnosis
Sensitivity
Specificity

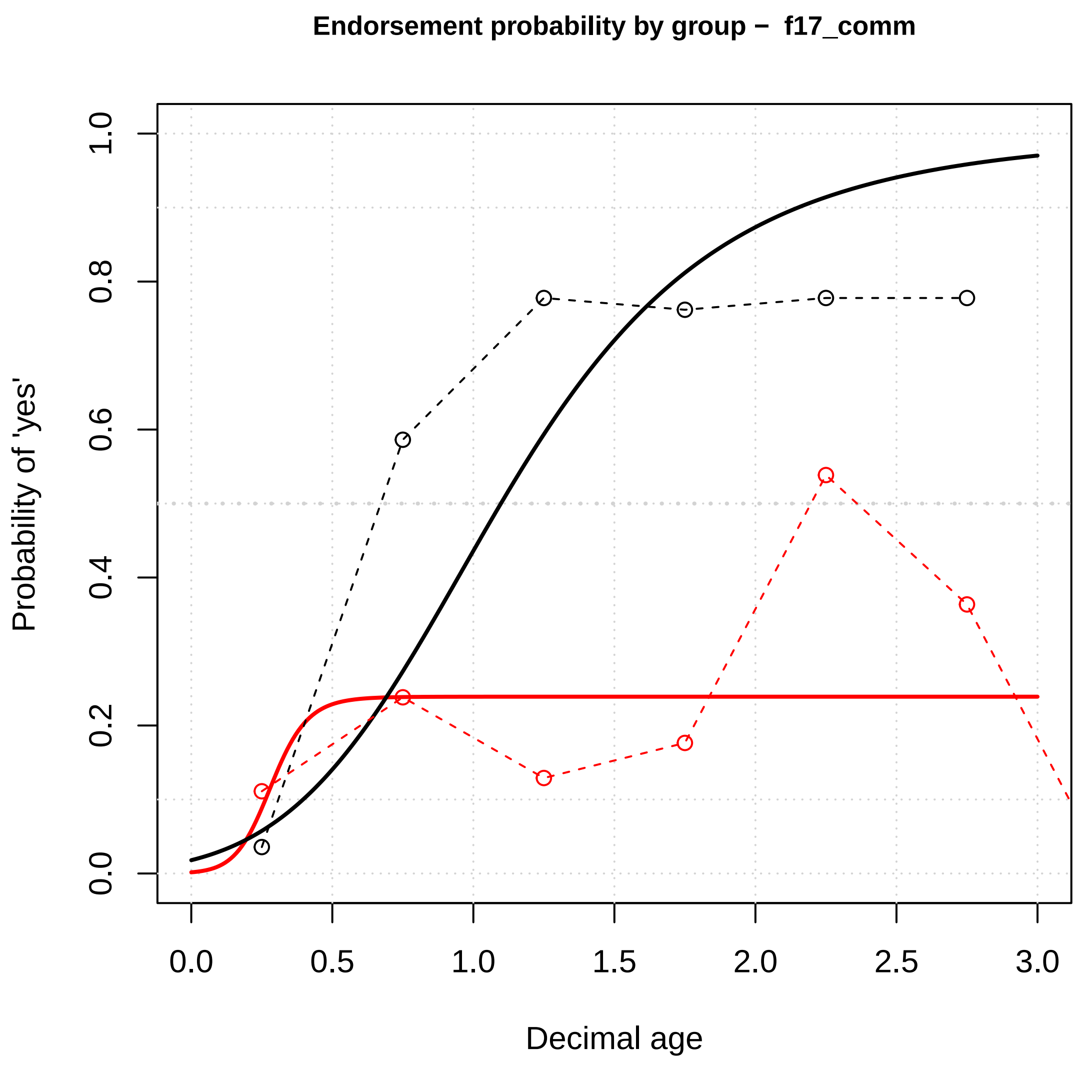
Endorsement probability by group – f17_comm
Probability of 'yes'
Decimal age
0.0
0.2
0.4
0.6
0.8
1.0
0.5
1.5
2.0
2.5
3.0

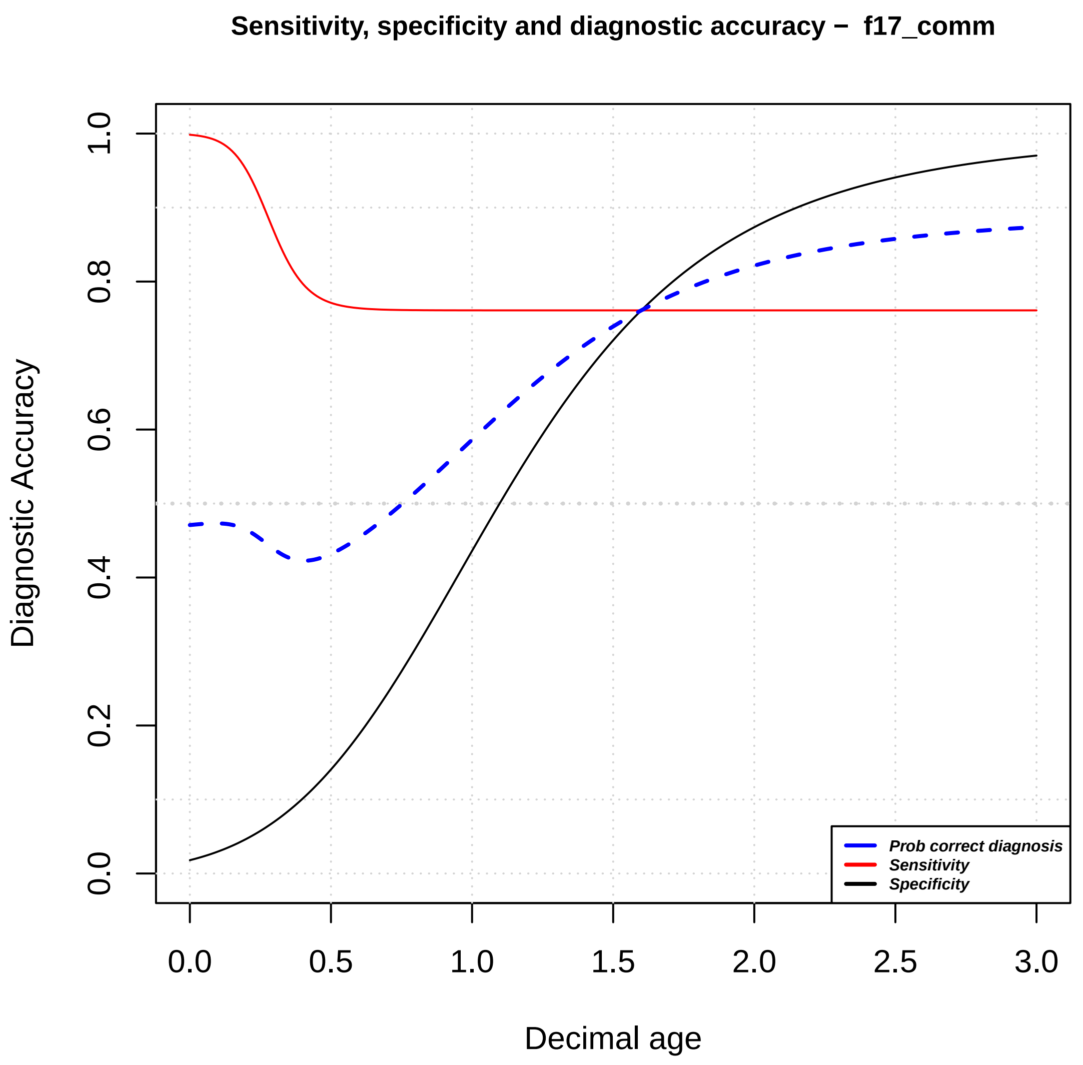
Sensitivity, specificity and diagnostic accuracy – f17_comm
Diagnostic Accuracy
Decimal age
Prob correct diagnosis
Sensitivity
Specificity
0.0
0.2
0.4
0.6
0.8
1.0
0.5
1.5
2.0
2.5
3.0

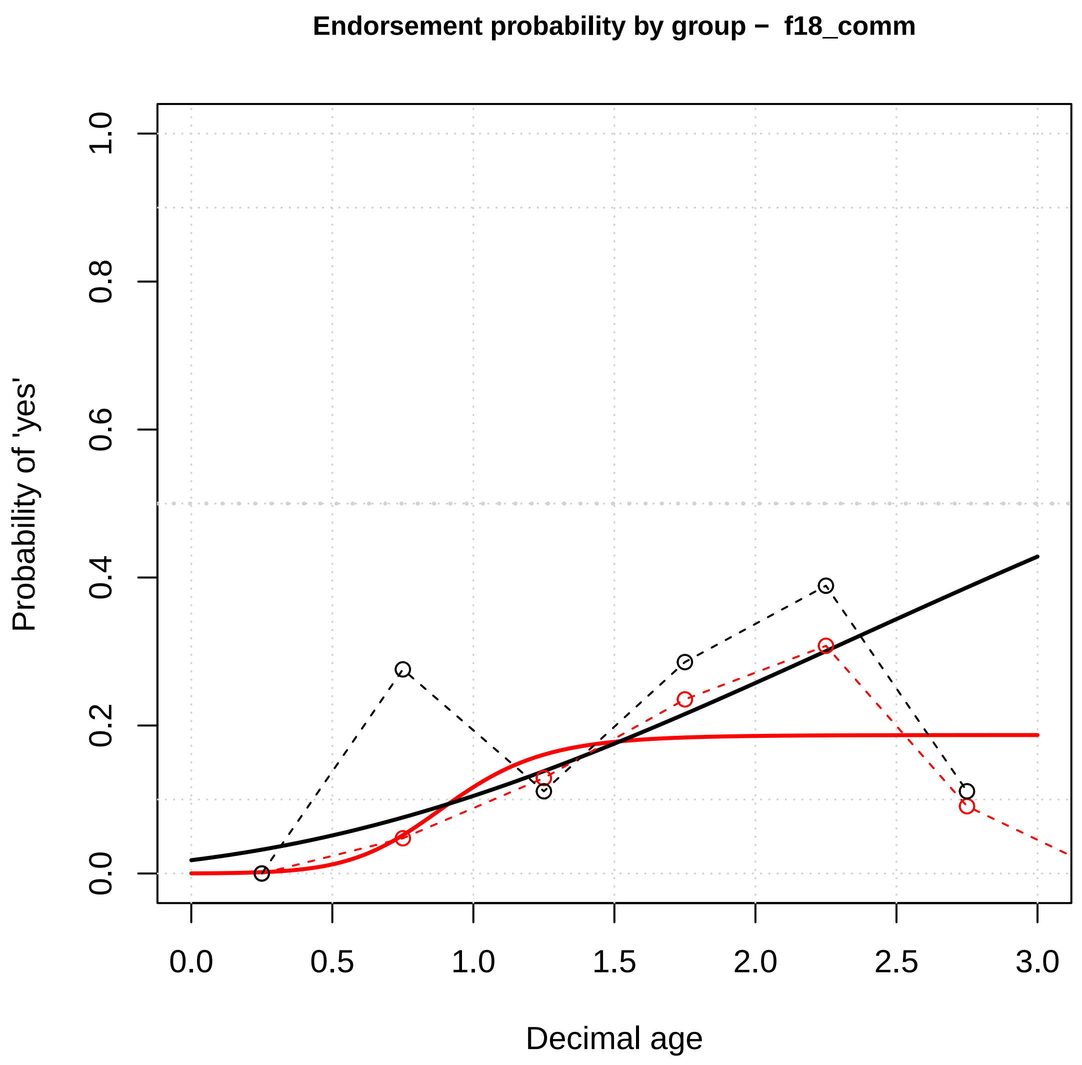

Endorsement probability by group – f18_comm
Probability of 'yes'
Decimal age
0.0
0.2
0.4
0.6
0.8
1.0
0.0
0.5
1.0
1.5
2.0
2.5
3.0


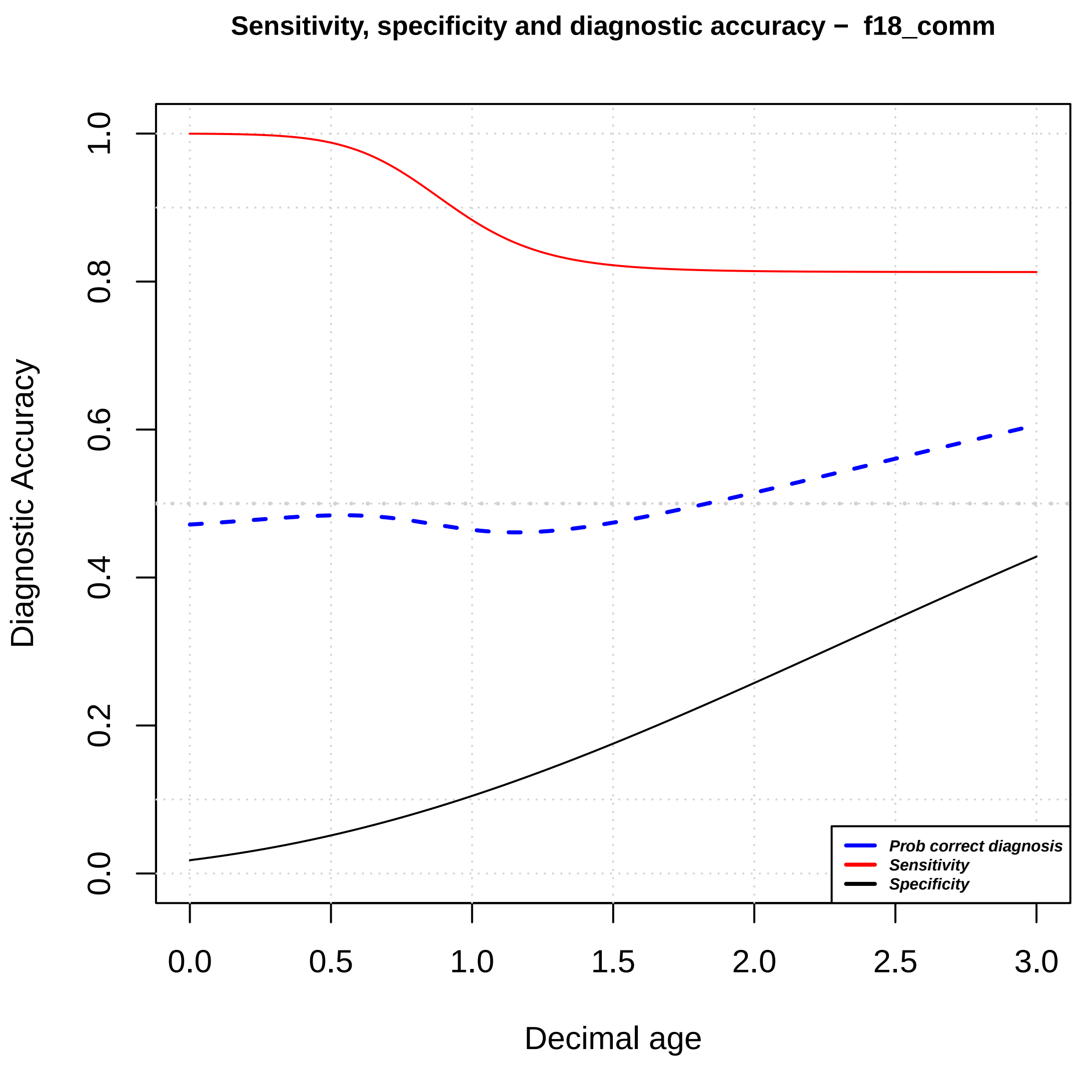

Sensitivity, specificity and diagnostic accuracy – f18_comm
Diagnostic Accuracy
Decimal age
Prob correct diagnosis
Sensitivity
Specificity
0.0
0.2
0.4
0.6
0.8
1.0
0.0
0.5
1.0
1.5
2.0
2.5
3.0

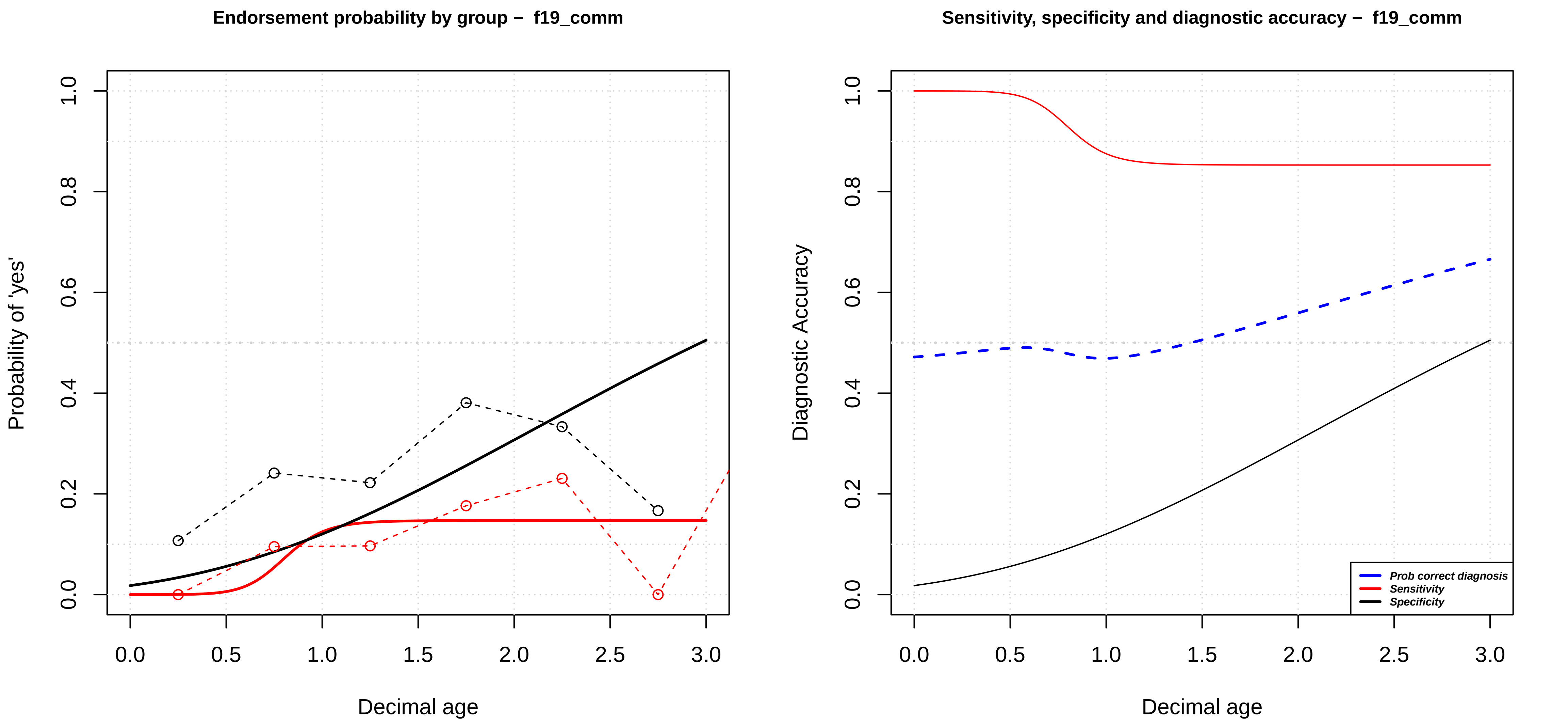

Endorsement probability by group – f19_comm
Probability of 'yes'
Decimal age
0.0
0.2
0.4
0.6
0.8
1.0
0.0
0.5
1.0
1.5
2.0
2.5
3.0
Sensitivity, specificity and diagnostic accuracy – f19_comm
Diagnostic Accuracy
Decimal age
Prob correct diagnosis
Sensitivity
Specificity

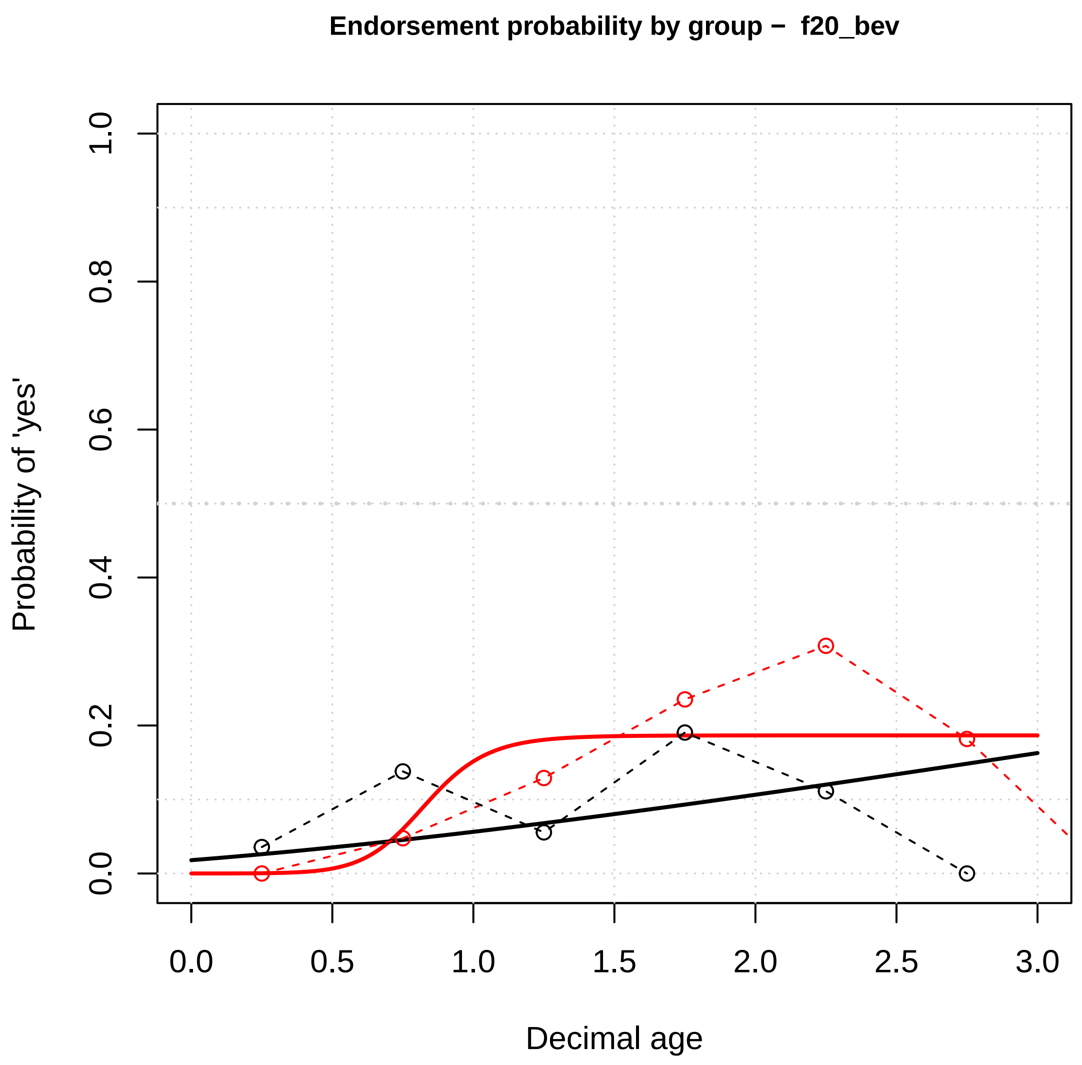
Endorsement probability by group – f20_bev
Probability of 'yes'
Decimal age
0.0
0.2
0.4
0.6
0.8
1.0
0.0
0.5
1.0
1.5
2.0
2.5
3.0

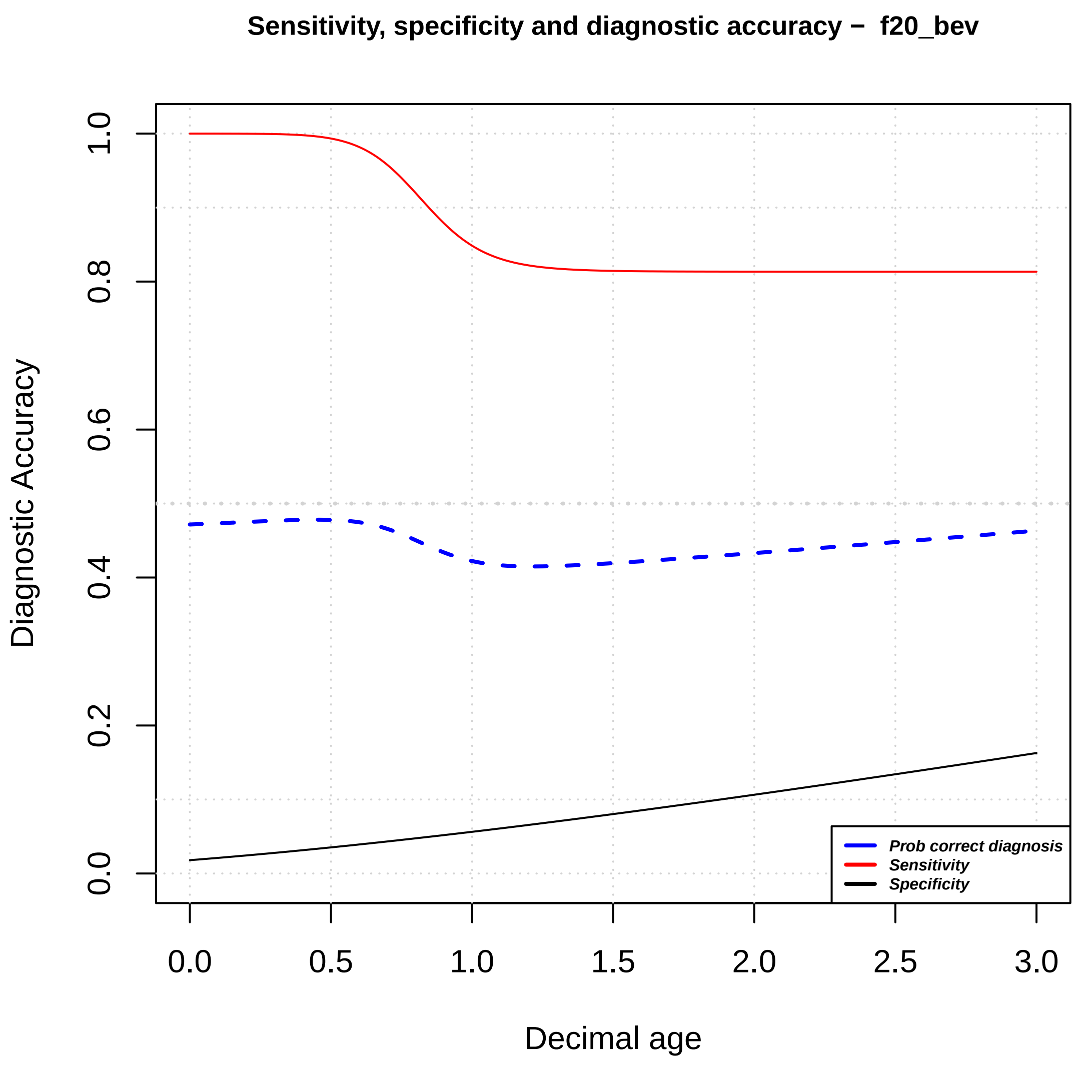
Sensitivity, specificity and diagnostic accuracy – f20_bev
Diagnostic Accuracy
Decimal age
0.0
0.2
0.4
0.6
0.8
1.0
0.0
0.5
1.0
1.5
2.0
2.5
3.0
Prob correct diagnosis
Sensitivity
Specificity

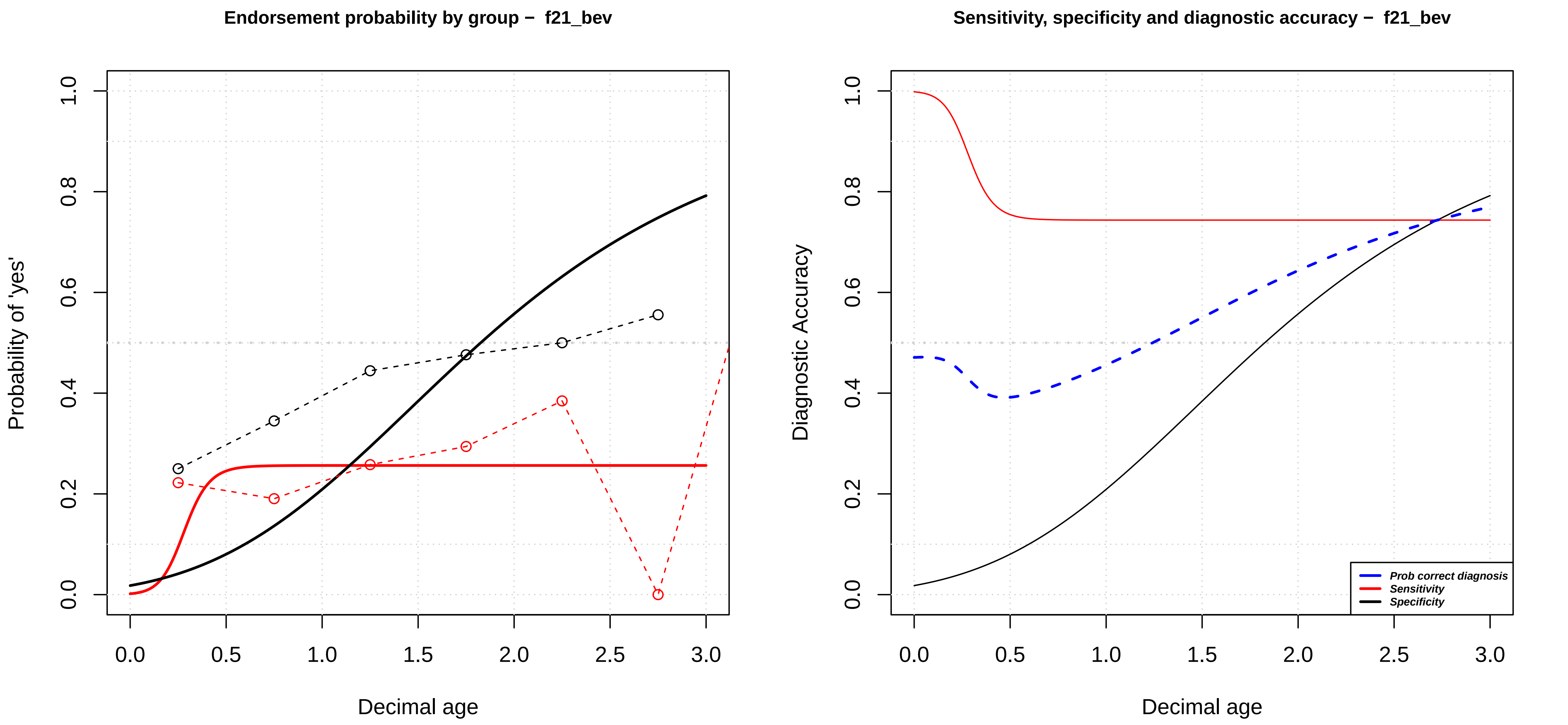

Endorsement probability by group – f21_bev
Probability of 'yes'
0.0
0.2
0.4
0.6
0.8
1.0
0.0
0.5
1.0
1.5
2.0
2.5
3.0
Decimal age
Sensitivity, specificity and diagnostic accuracy – f21_bev
Diagnostic Accuracy
Prob correct diagnosis
Sensitivity
Specificity
Decimal age

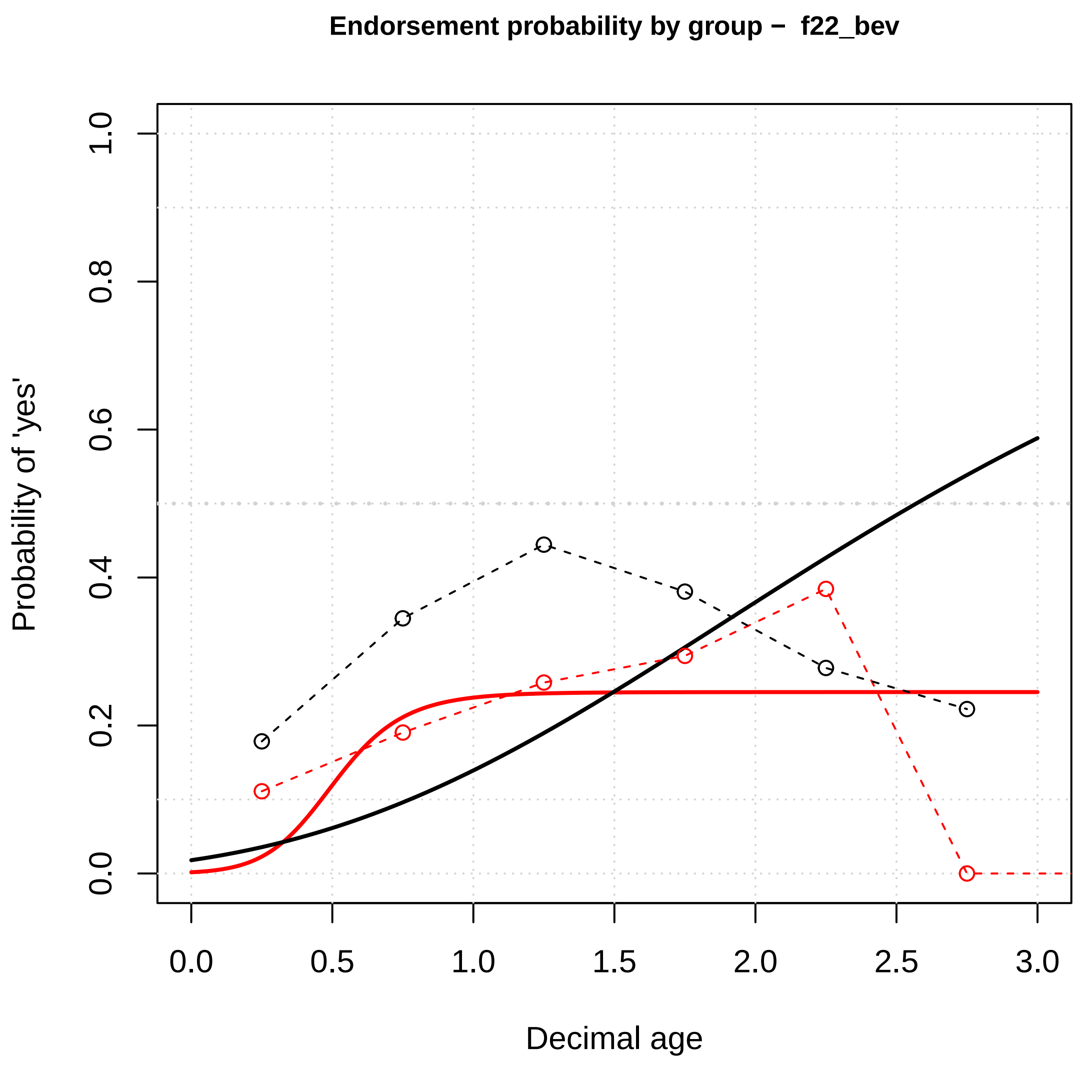

Endorsement probability by group – f22_bev
Probability of 'yes'
0.0
0.2
0.4
0.6
0.8
1.0
0.0
0.5
1.0
1.5
2.0
2.5
3.0
Decimal age


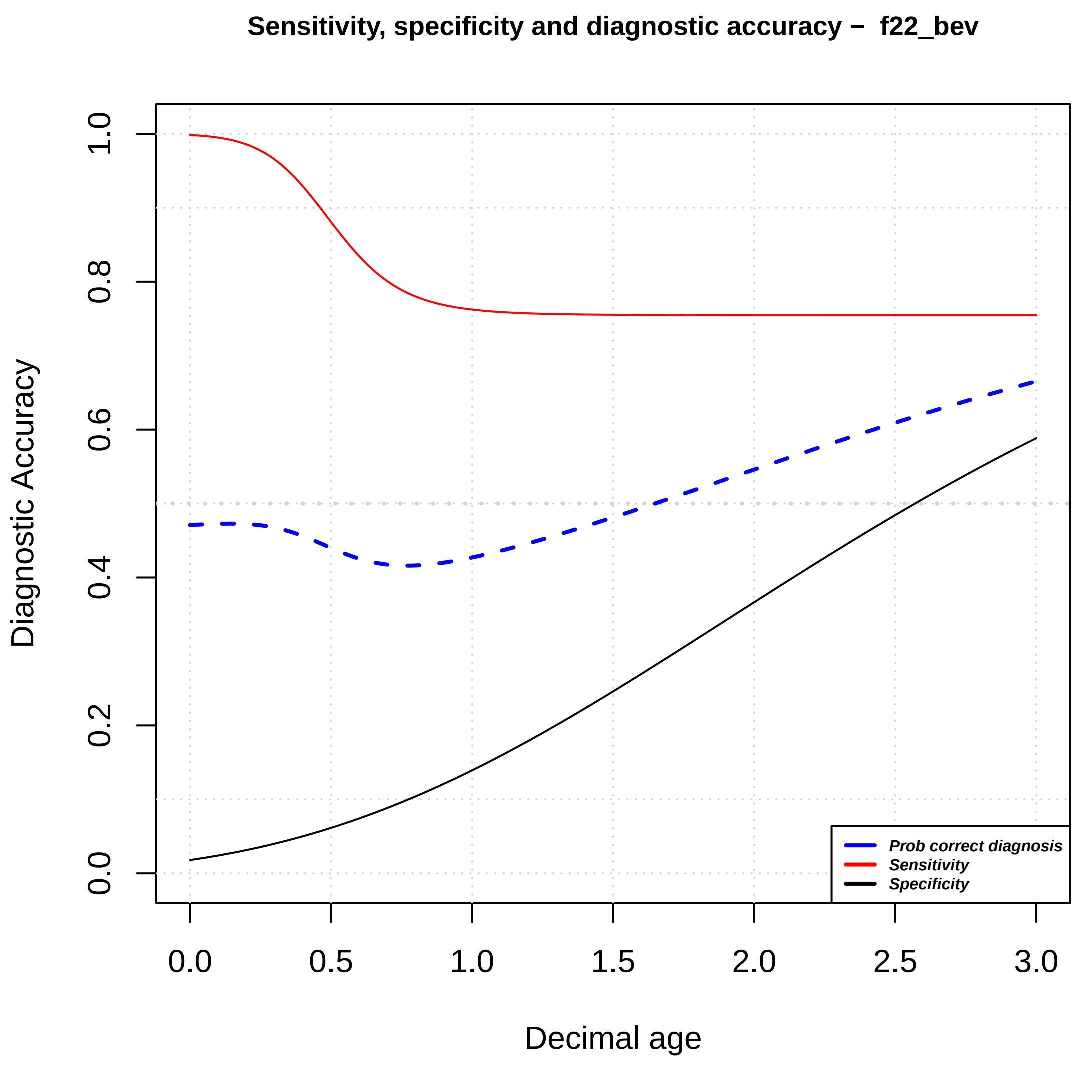

Sensitivity, specificity and diagnostic accuracy – f22_bev
Diagnostic Accuracy
0.0
0.2
0.4
0.6
0.8
1.0
0.0
0.5
1.0
1.5
2.0
2.5
3.0
Decimal age
Prob correct diagnosis
Sensitivity
Specificity

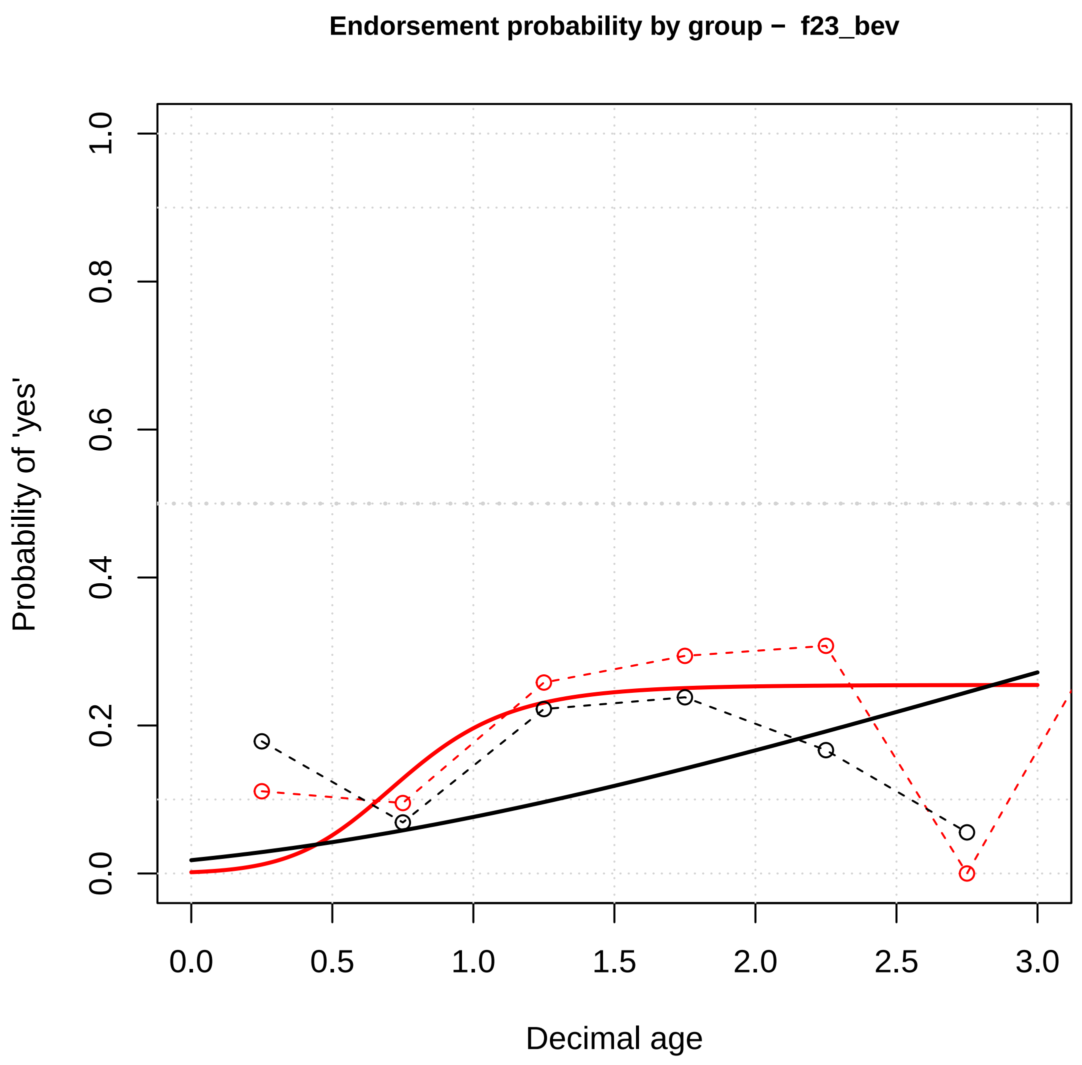
Endorsement probability by group – f23_bev
Probability of 'yes'
Decimal age
0.0
0.2
0.4
0.6
0.8
1.0
0.0
0.5
1.0
1.5
2.0
2.5
3.0

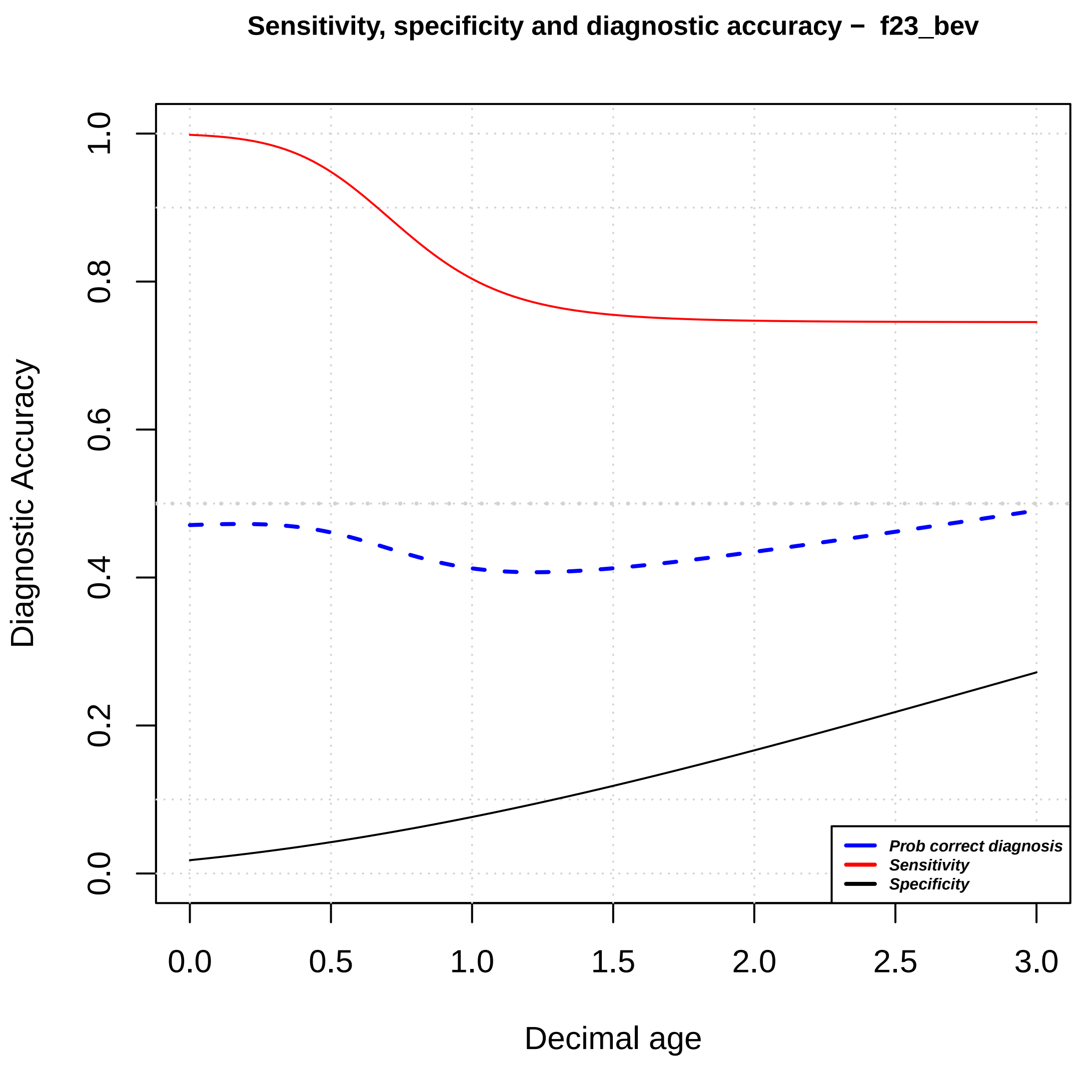
Sensitivity, specificity and diagnostic accuracy – f23_bev
Diagnostic Accuracy
Decimal age
Prob correct diagnosis
Sensitivity
Specificity
0.0
0.2
0.4
0.6
0.8
1.0
0.0
0.5
1.0
1.5
2.0
2.5
3.0

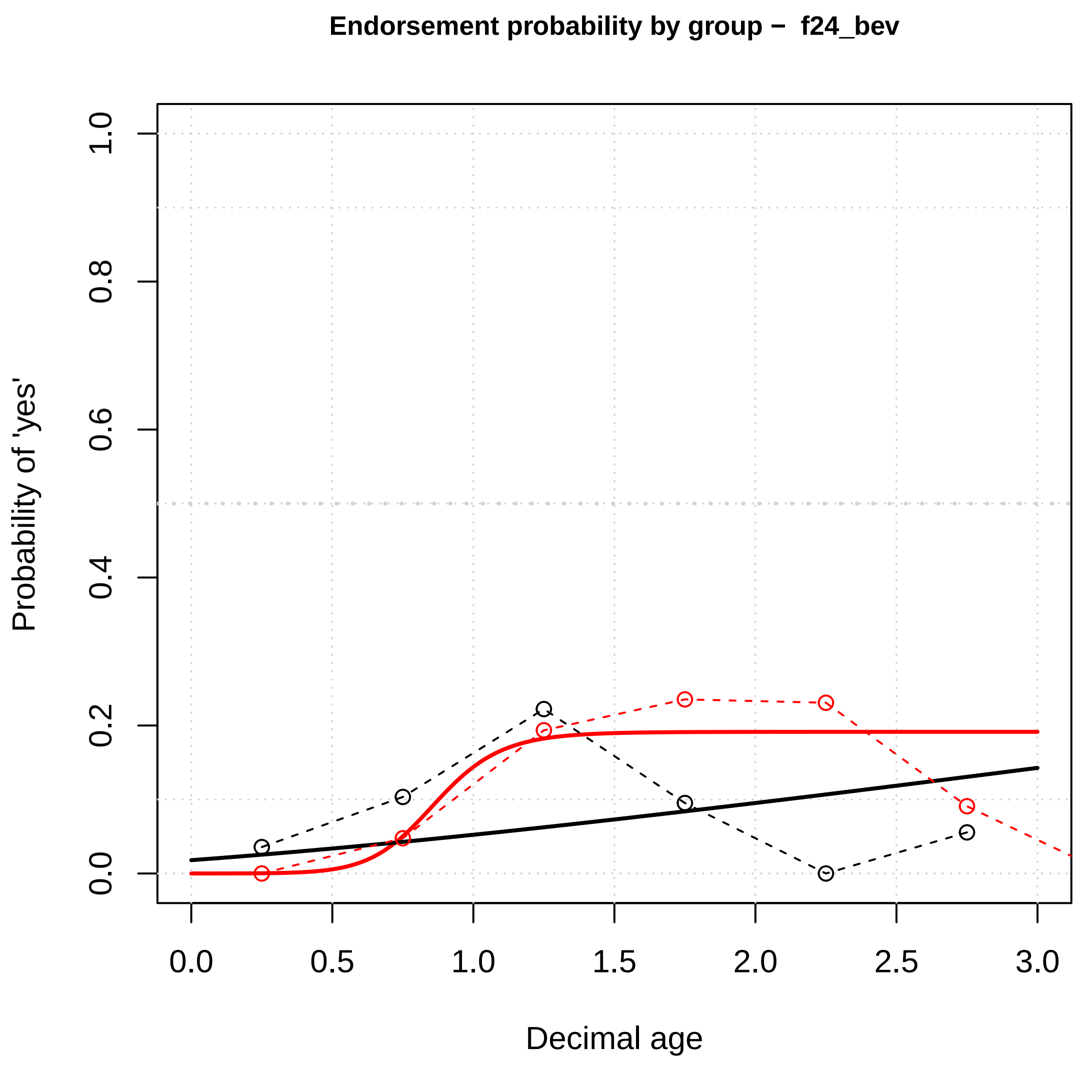
Endorsement probability by group – f24_bev
Probability of 'yes'
Decimal age
0.0
0.2
0.4
0.6
0.8
1.0
0.0
0.5
1.0
1.5
2.0
2.5
3.0

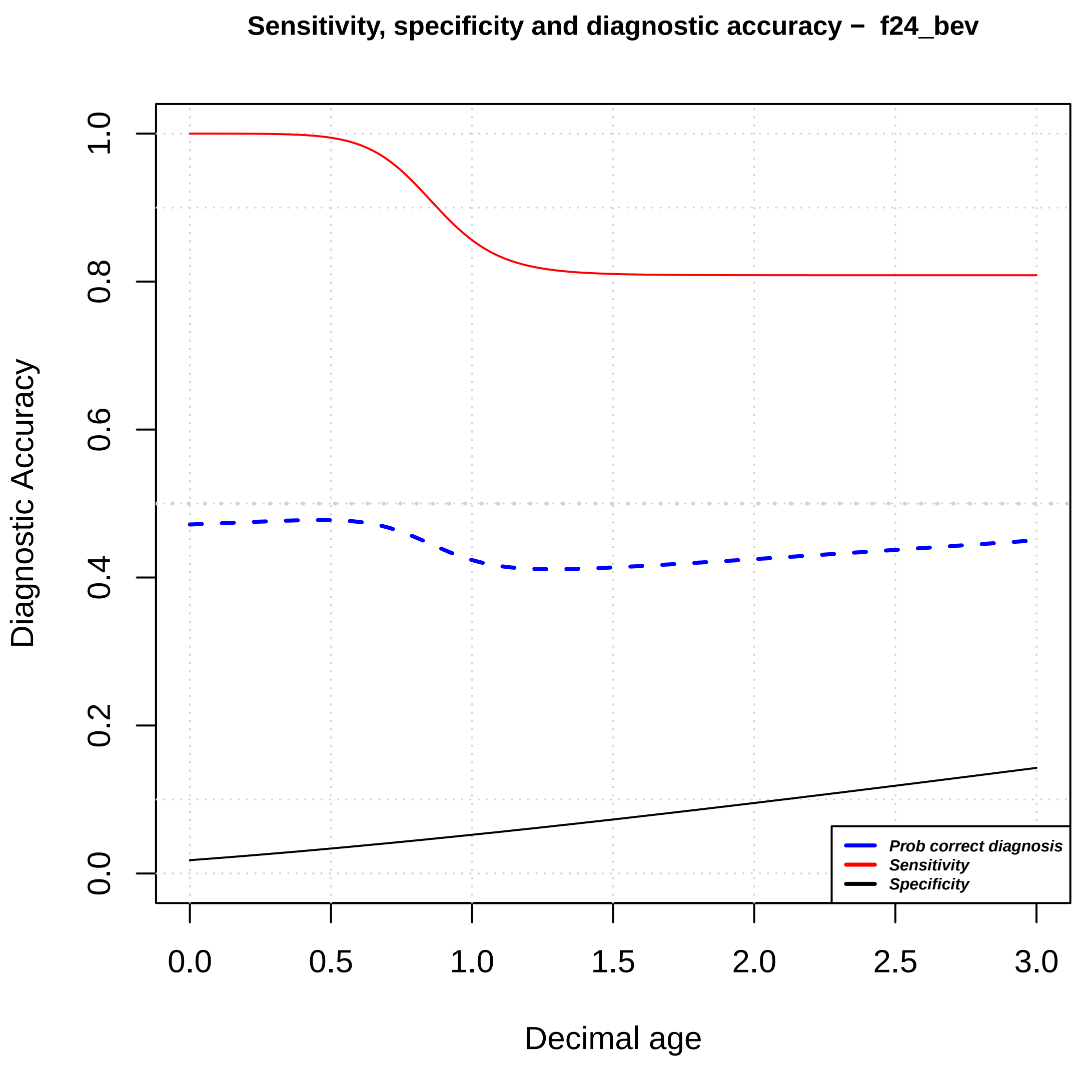
Sensitivity, specificity and diagnostic accuracy – f24_bev
Diagnostic Accuracy
Decimal age
Prob correct diagnosis
Sensitivity
Specificity
0.0
0.2
0.4
0.6
0.8
1.0
0.0
0.5
1.0
1.5
2.0
2.5
3.0

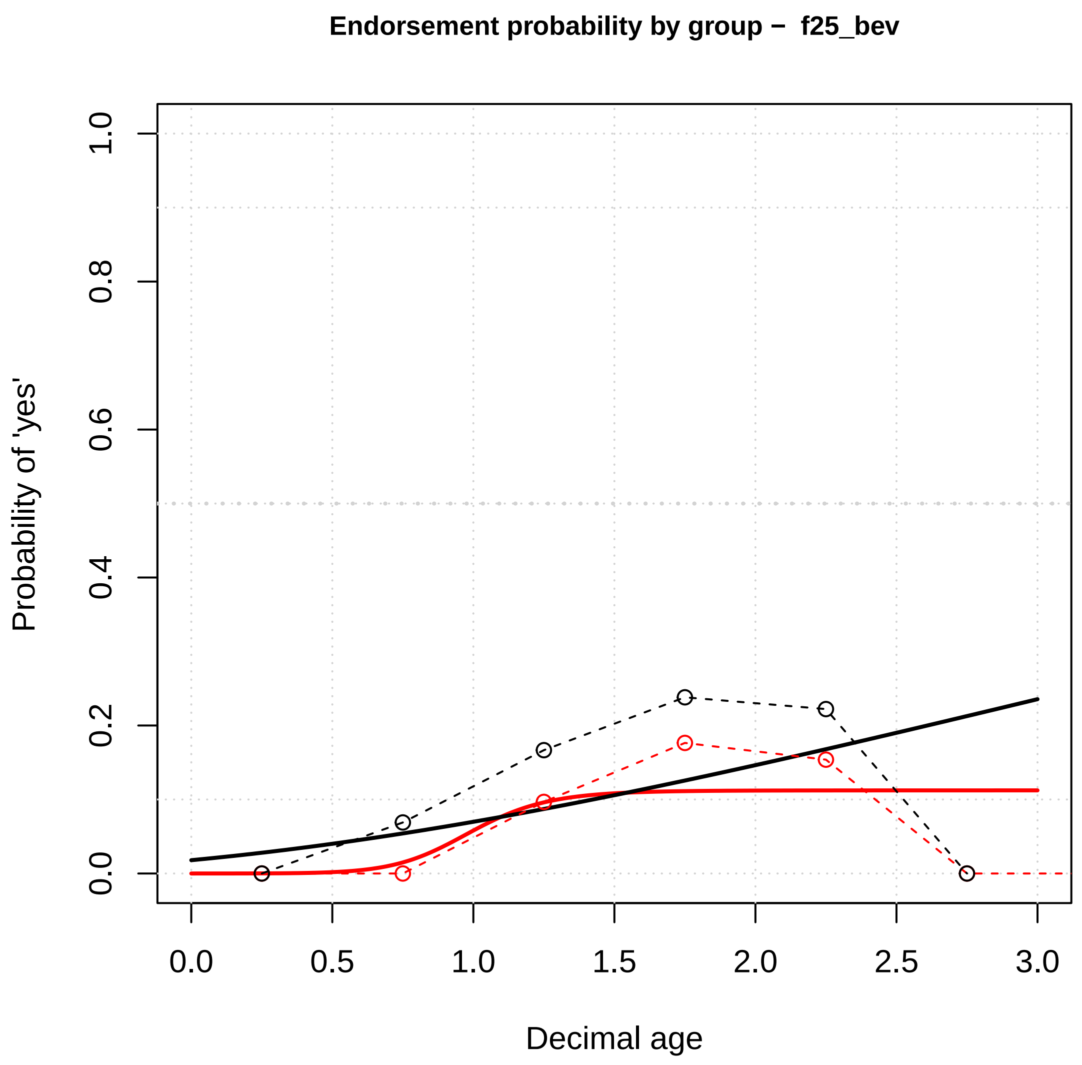

Endorsement probability by group – f25_bev
Probability of 'yes'
Decimal age
0.0
0.2
0.4
0.6
0.8
1.0
0.0
0.5
1.0
1.5
2.0
2.5
3.0


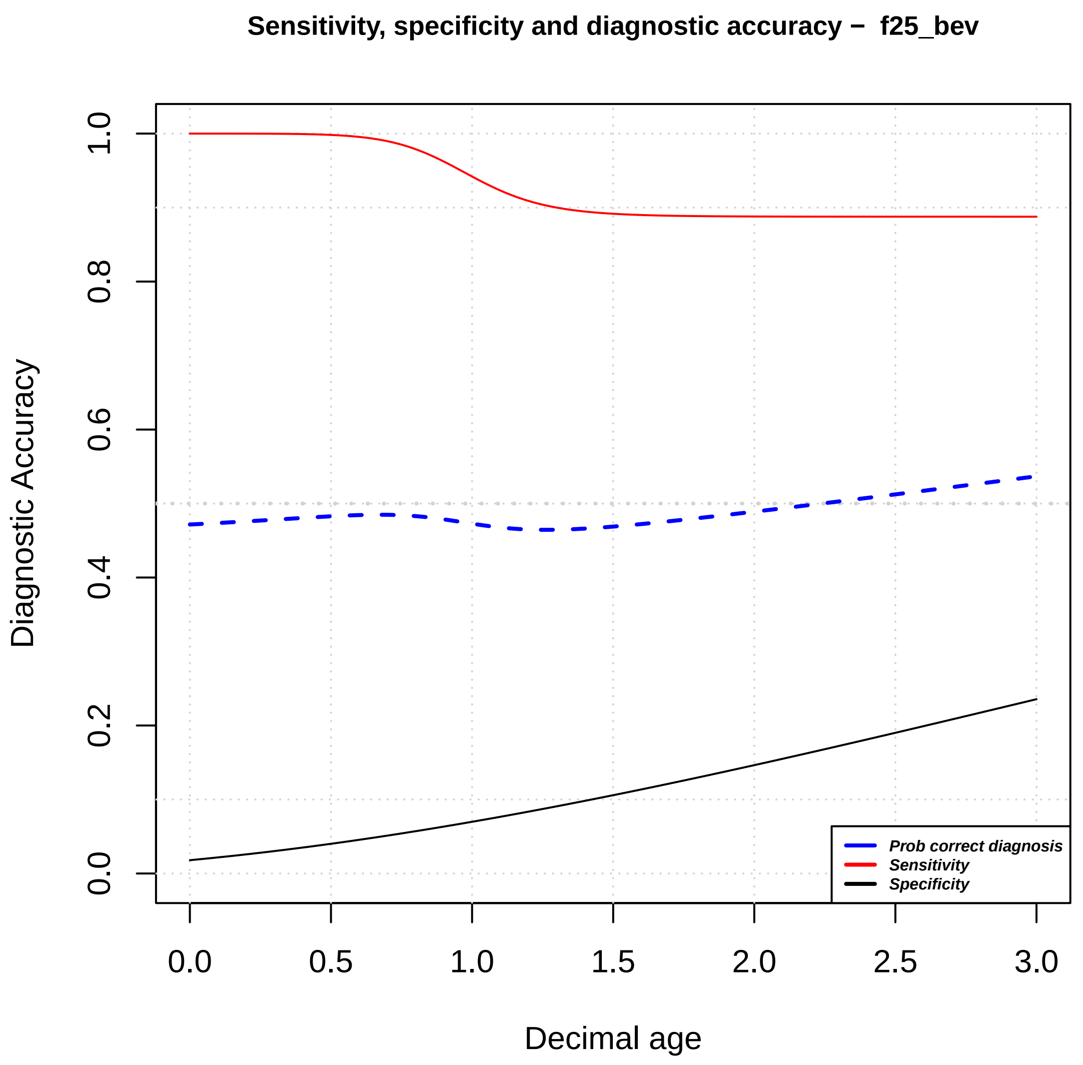

Sensitivity, specificity and diagnostic accuracy – f25_bev
Diagnostic Accuracy
Decimal age
0.0
0.2
0.4
0.6
0.8
1.0
0.0
0.5
1.0
1.5
2.0
2.5
3.0
Prob correct diagnosis
Sensitivity
Specificity

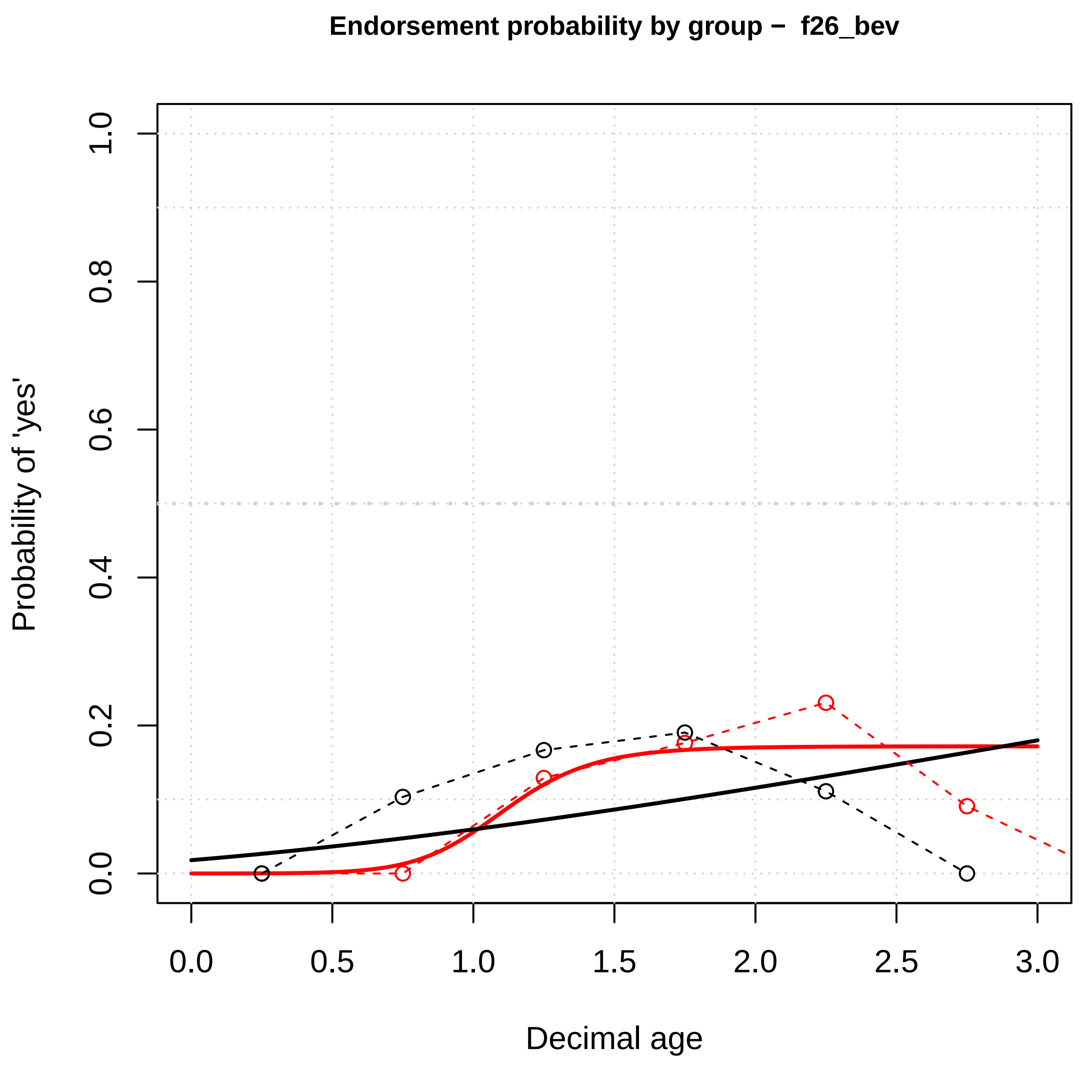

Endorsement probability by group – f26_bev
Probability of 'yes'
Decimal age
0.0
0.2
0.4
0.6
0.8
1.0
0.5
1.5
2.0
2.5
3.0


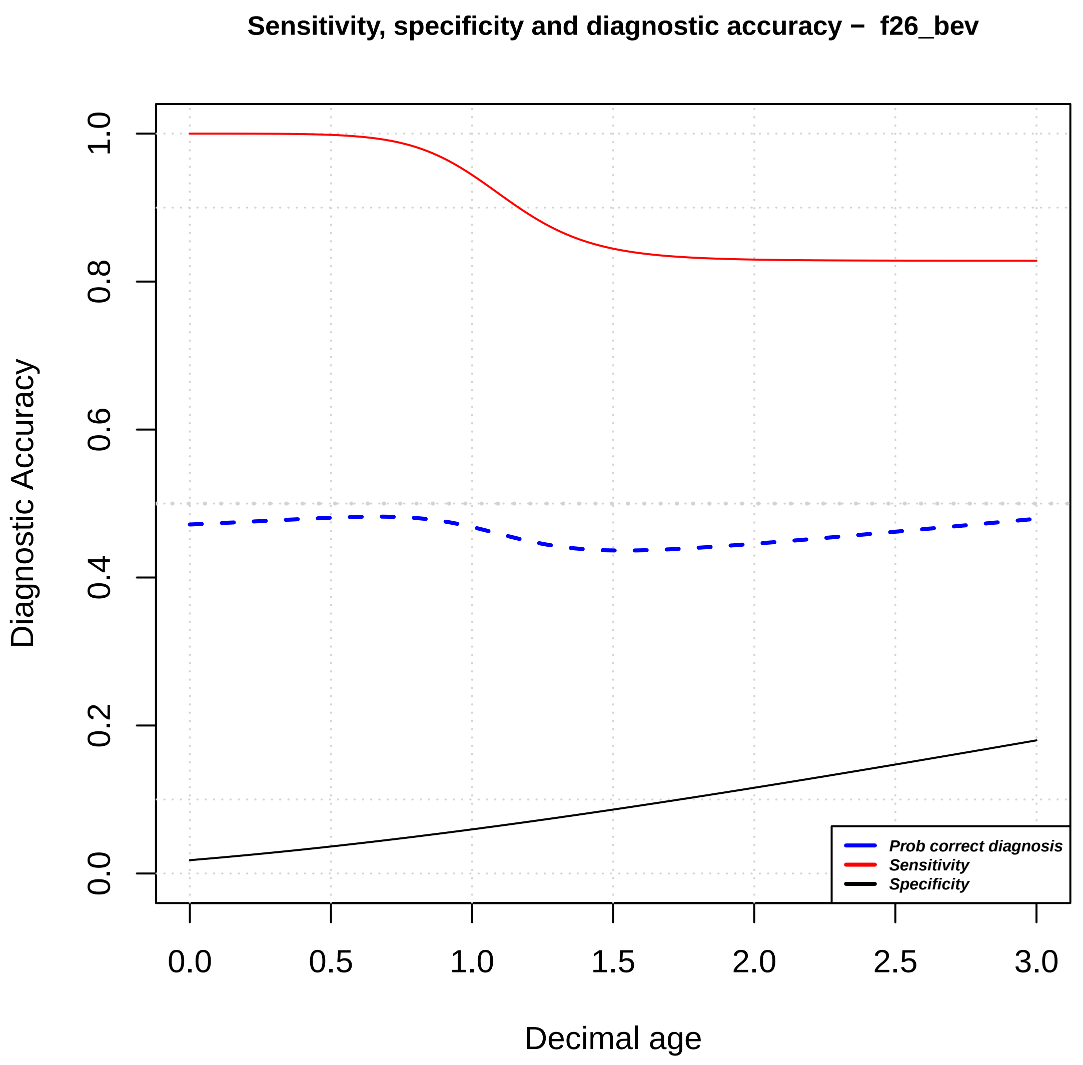

Sensitivity, specificity and diagnostic accuracy – f26_bev
Diagnostic Accuracy
Decimal age
Prob correct diagnosis
Sensitivity
Specificity
0.0
0.2
0.4
0.6
0.8
1.0
0.5
1.5
2.0
2.5
3.0

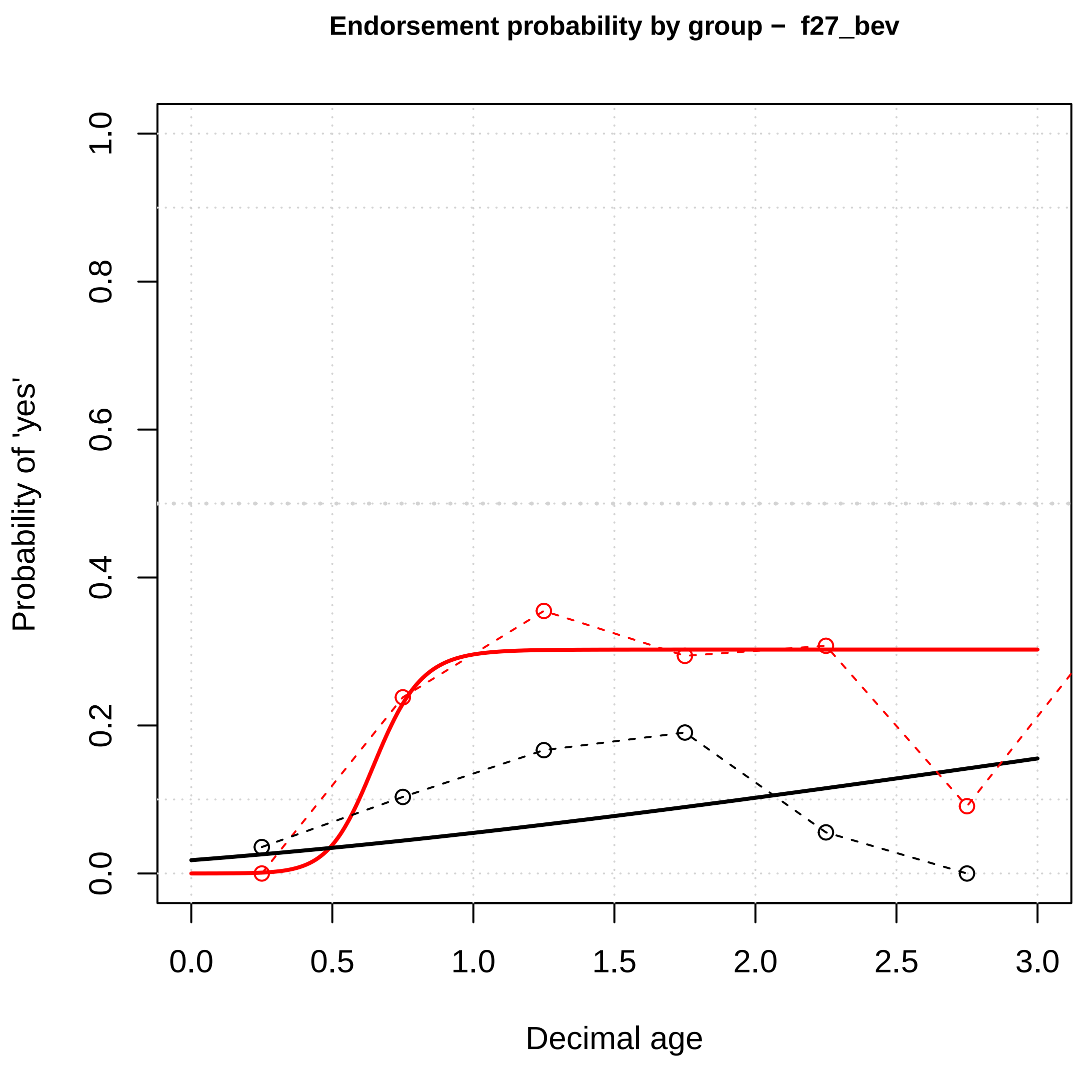

Endorsement probability by group – f27_bev
Probability of 'yes'
Decimal age
0.0
0.2
0.4
0.6
0.8
1.0
0.0
0.5
1.0
1.5
2.0
2.5
3.0


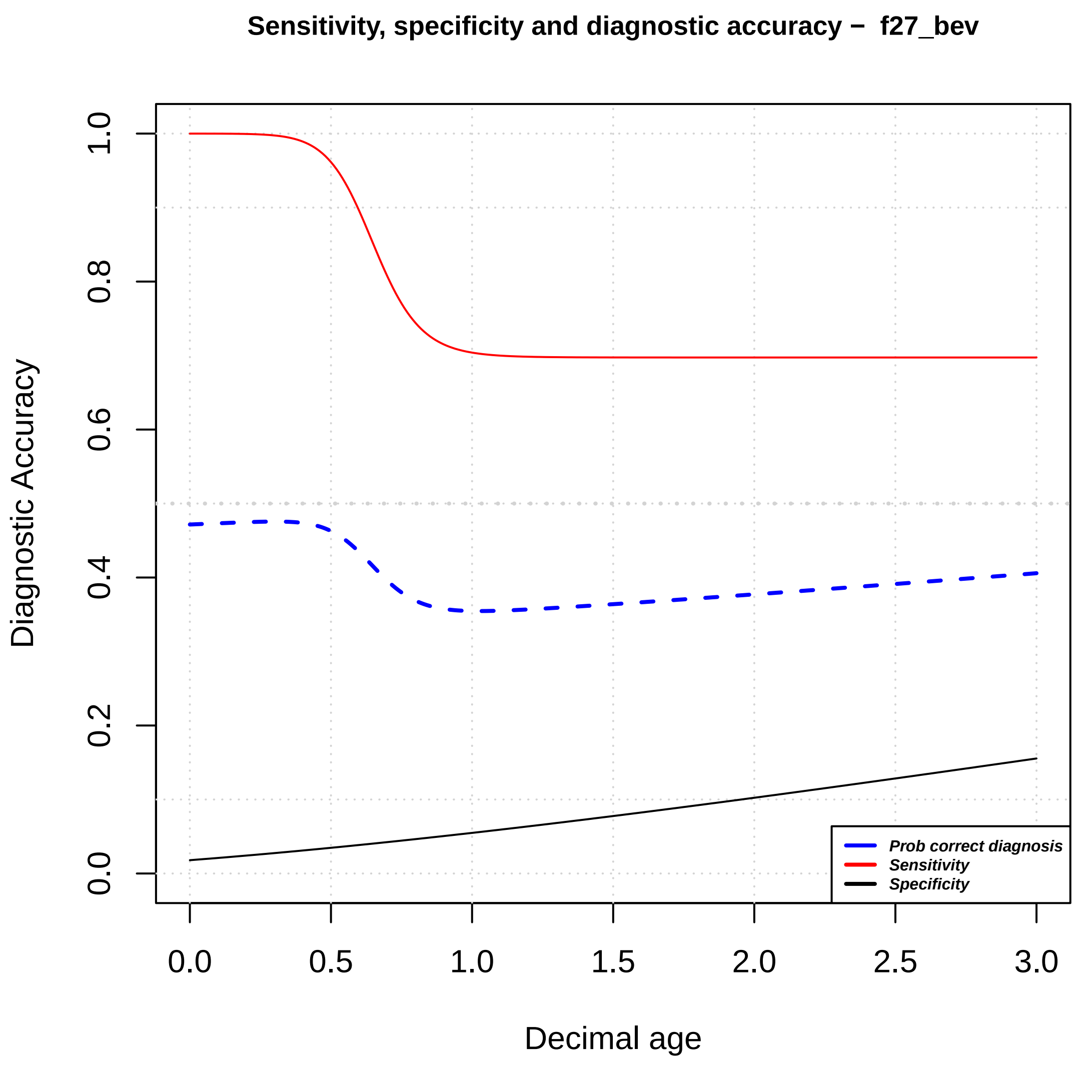

Sensitivity, specificity and diagnostic accuracy – f27_bev
Diagnostic Accuracy
Decimal age
0.0
0.2
0.4
0.6
0.8
1.0
0.0
0.5
1.0
1.5
2.0
2.5
3.0
Prob correct diagnosis
Sensitivity
Specificity

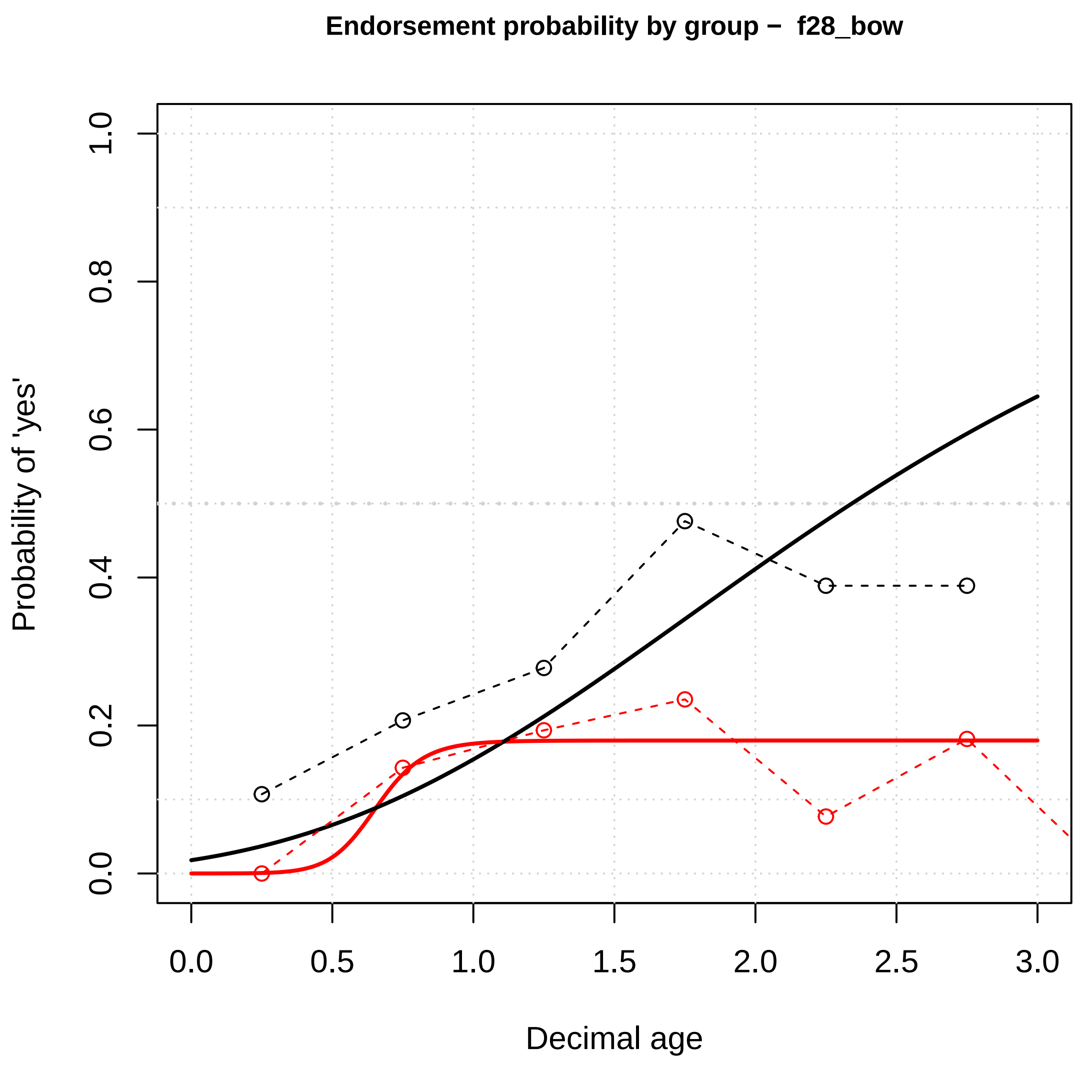

Endorsement probability by group – f28_bow
Probability of 'yes'
Decimal age
0.0
0.2
0.4
0.6
0.8
1.0
0.5
1.5
2.0
2.5
3.0


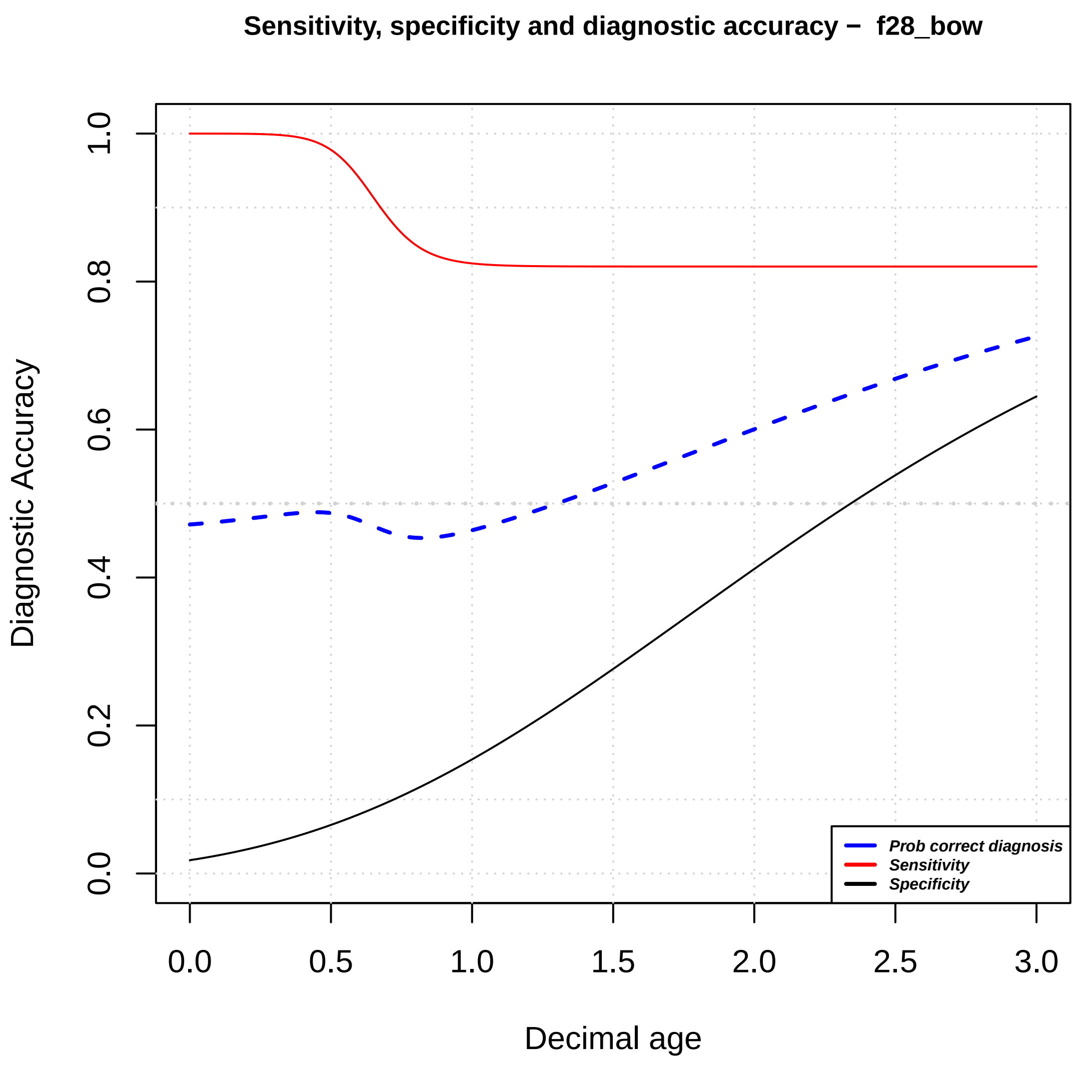

Sensitivity, specificity and diagnostic accuracy – f28_bow
Diagnostic Accuracy
Decimal age
Prob correct diagnosis
Sensitivity
Specificity
0.0
0.2
0.4
0.6
0.8
1.0
0.5
1.5
2.0
2.5
3.0

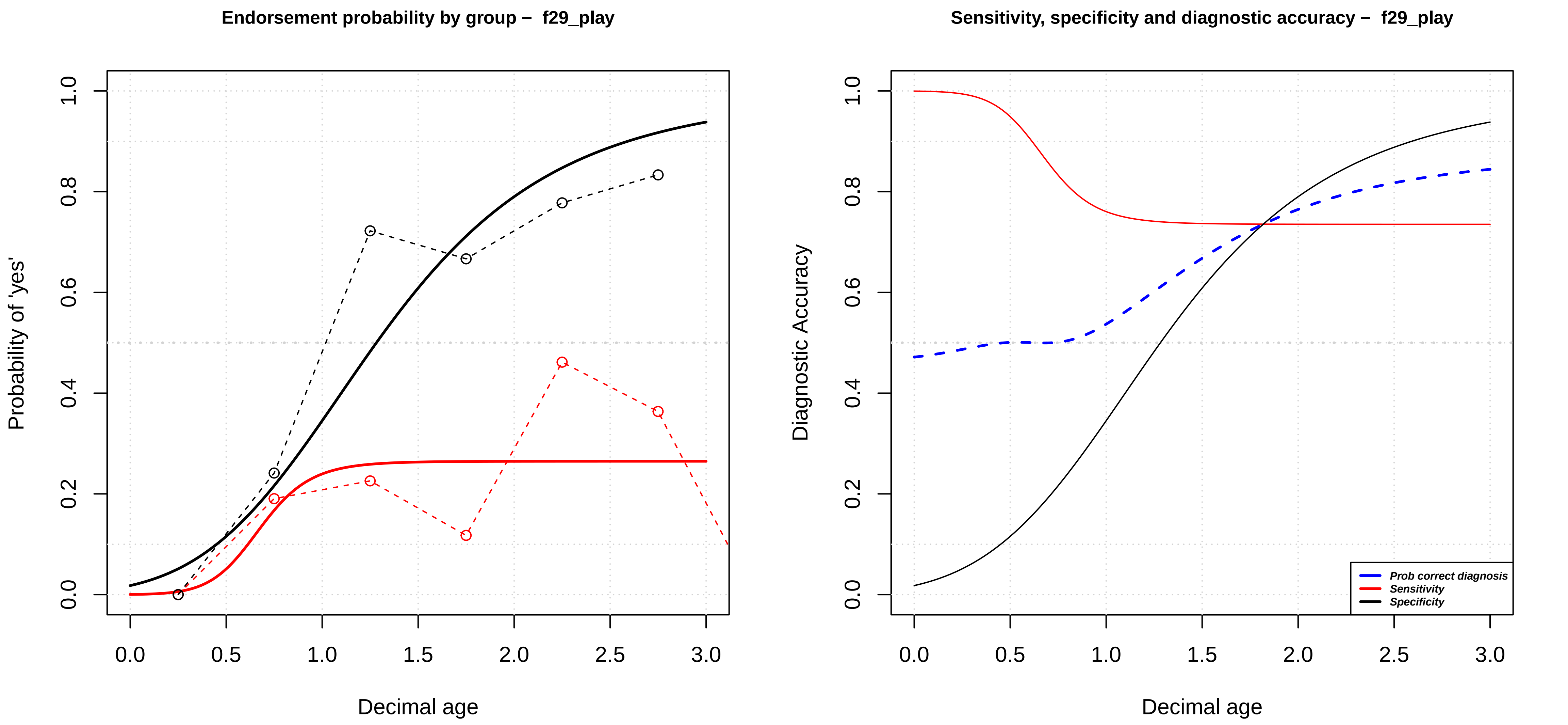

Endorsement probability by group – f29_play
Probability of 'yes'
Decimal age
0.0
0.2
0.4
0.6
0.8
1.0
0.0
0.5
1.0
1.5
2.0
2.5
3.0
Sensitivity, specificity and diagnostic accuracy – f29_play
Diagnostic Accuracy
Decimal age
Prob correct diagnosis
Sensitivity
Specificity

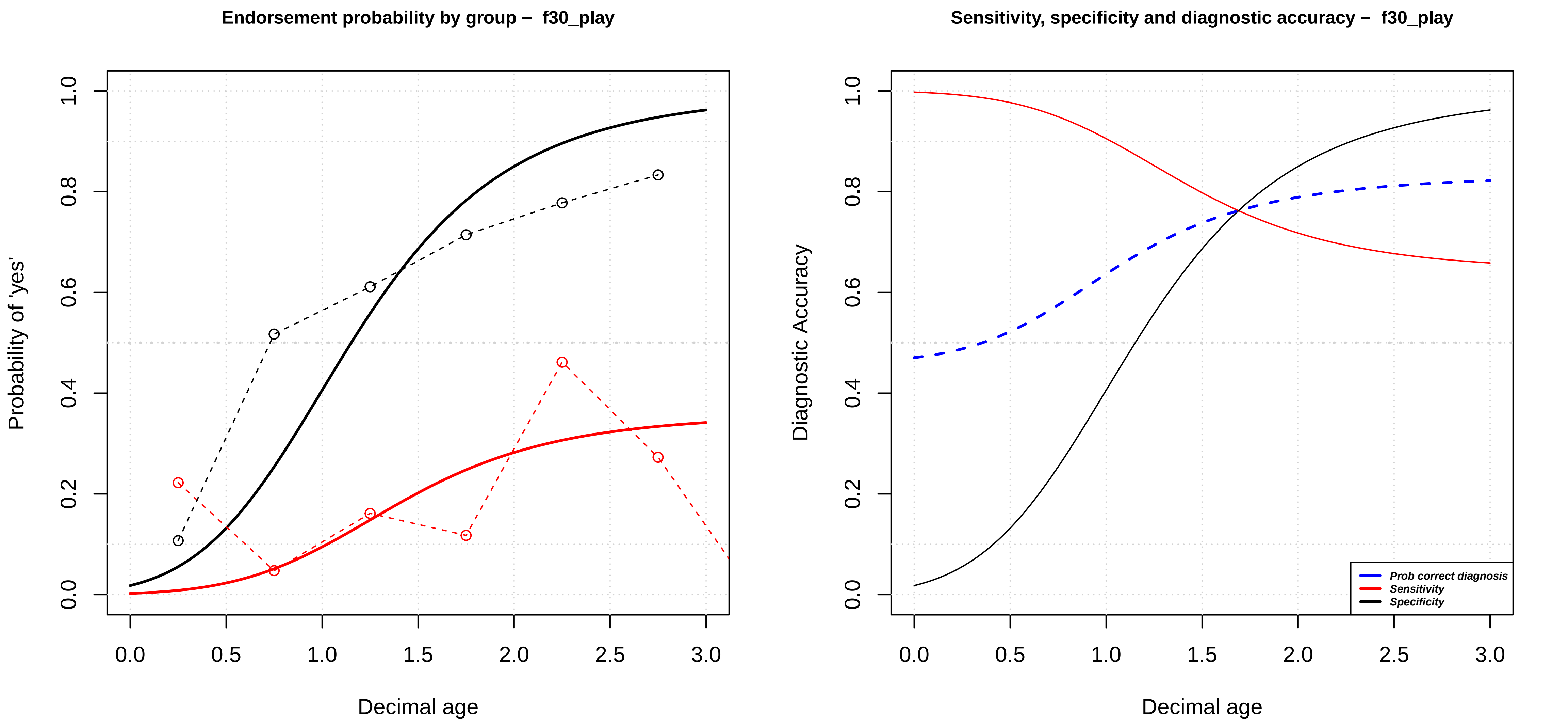

Endorsement probability by group – f30_play
Probability of 'yes'
Decimal age
0.0
0.2
0.4
0.6
0.8
1.0
0.0
0.5
1.0
1.5
2.0
2.5
3.0
Sensitivity, specificity and diagnostic accuracy – f30_play
Diagnostic Accuracy
Decimal age
Prob correct diagnosis
Sensitivity
Specificity

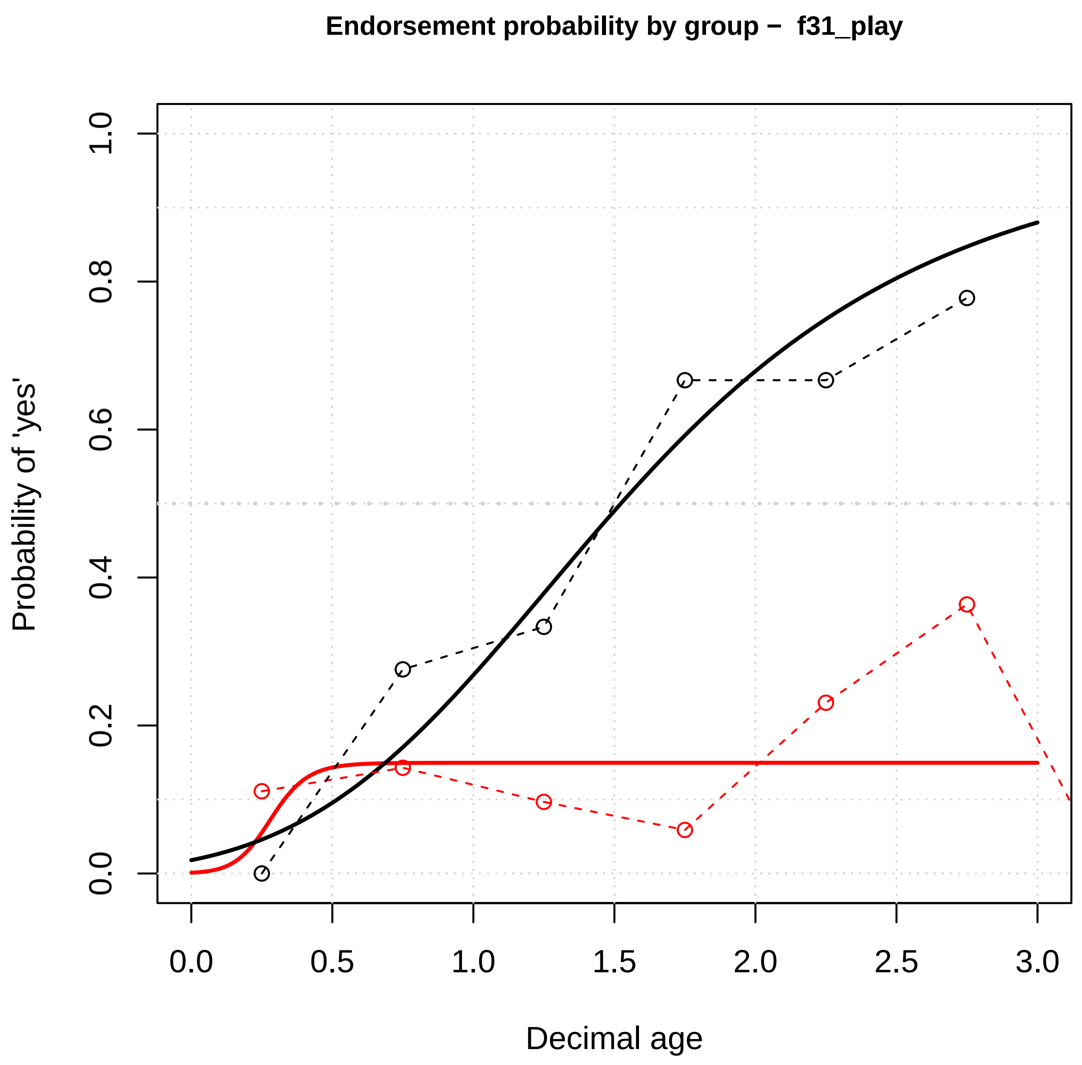

Endorsement probability by group – f31_play
Probability of 'yes'
Decimal age
0.0
0.2
0.4
0.6
0.8
1.0
0.0
0.5
1.0
1.5
2.0
2.5
3.0


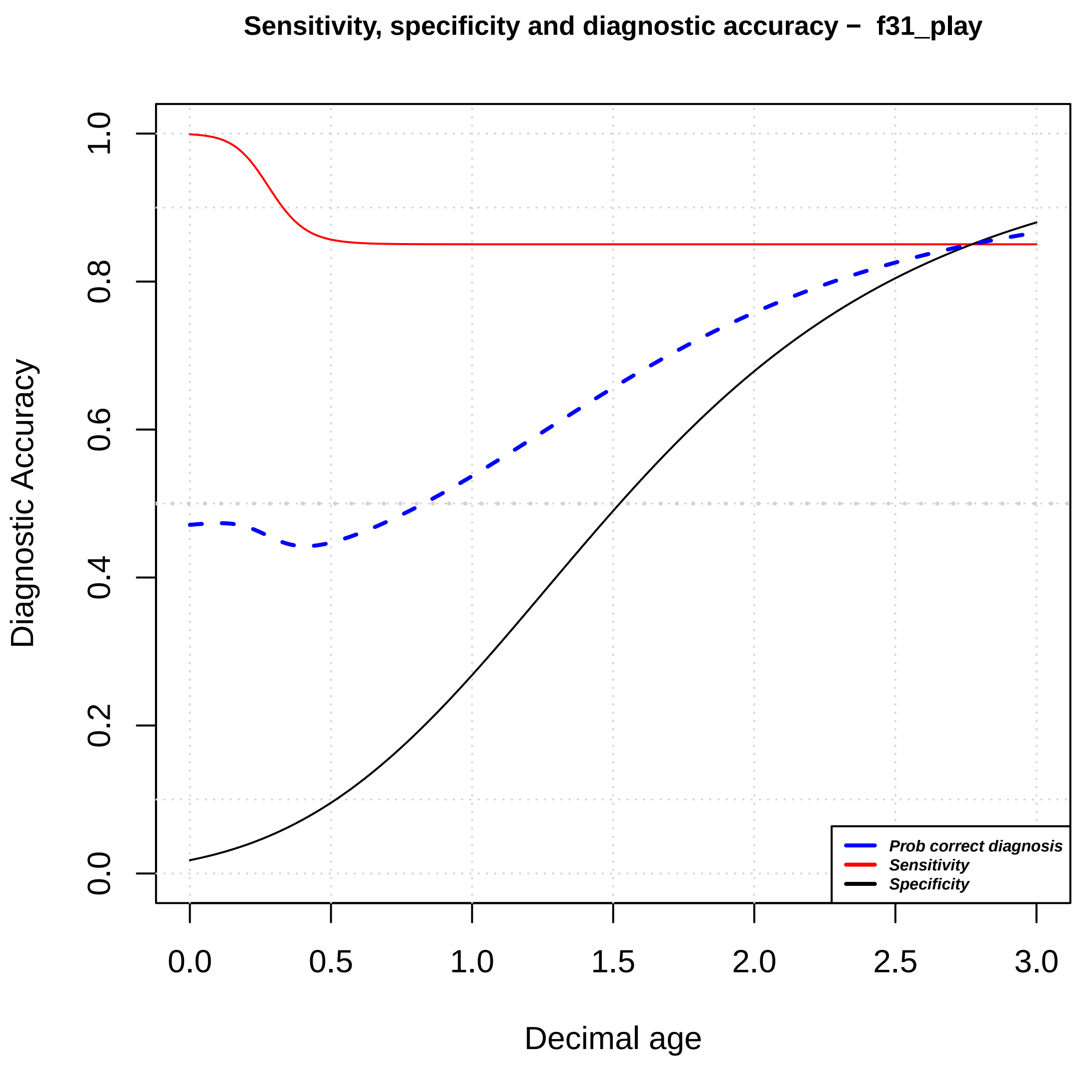

Sensitivity, specificity and diagnostic accuracy – f31_play
Diagnostic Accuracy
Decimal age
Prob correct diagnosis
Sensitivity
Specificity
0.0
0.2
0.4
0.6
0.8
1.0
0.0
0.5
1.0
1.5
2.0
2.5
3.0

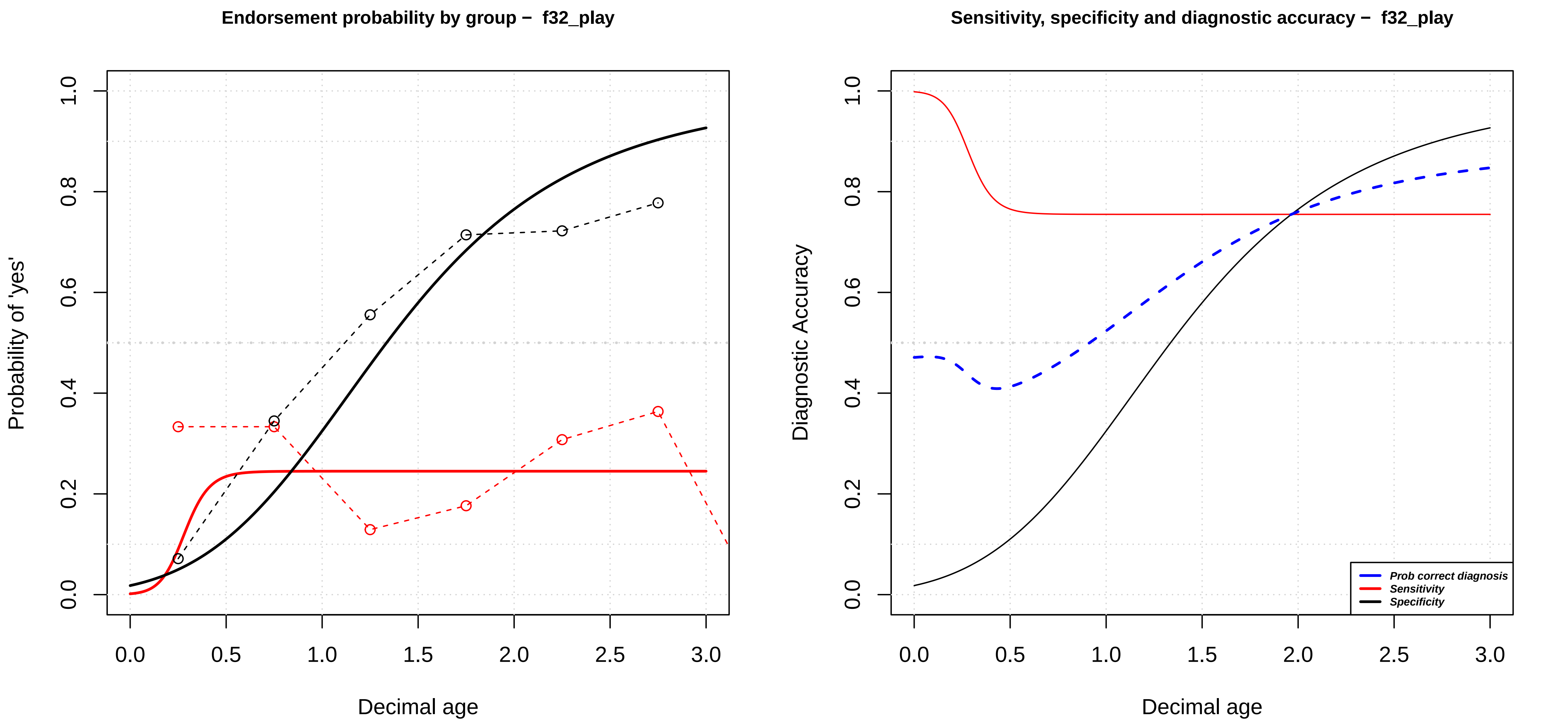

Endorsement probability by group − f32_play
Probability of 'yes'
Decimal age
0.0
0.2
0.4
0.6
0.8
1.0
0.0
0.5
1.0
1.5
2.0
2.5
3.0
Sensitivity, specificity and diagnostic accuracy − f32_play
Diagnostic Accuracy
Decimal age
Prob correct diagnosis
Sensitivity
Specificity

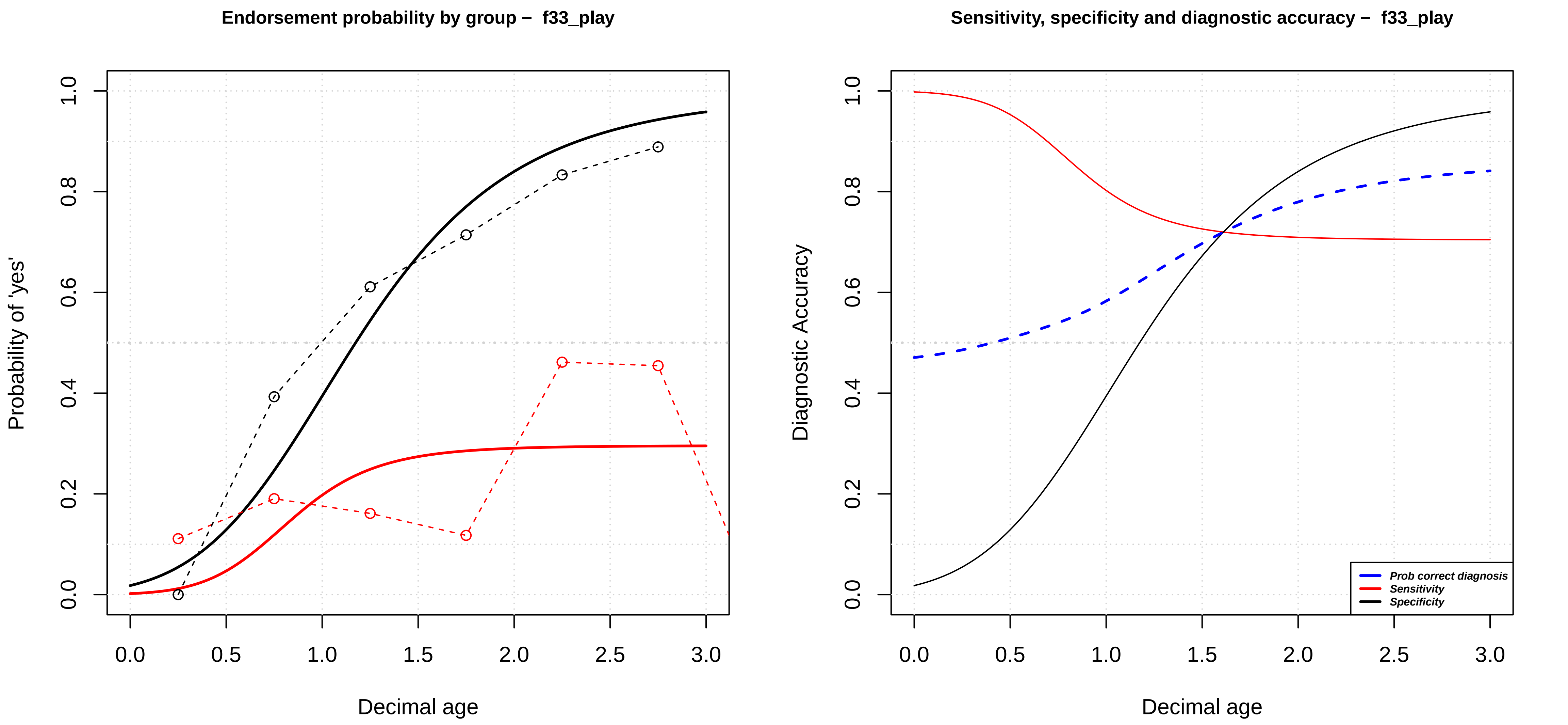

Endorsement probability by group – f33_play
Probability of 'yes'
Decimal age
0.0
0.2
0.4
0.6
0.8
1.0
0.0
0.5
1.0
1.5
2.0
2.5
3.0
Sensitivity, specificity and diagnostic accuracy – f33_play
Diagnostic Accuracy
Decimal age
Prob correct diagnosis
Sensitivity
Specificity

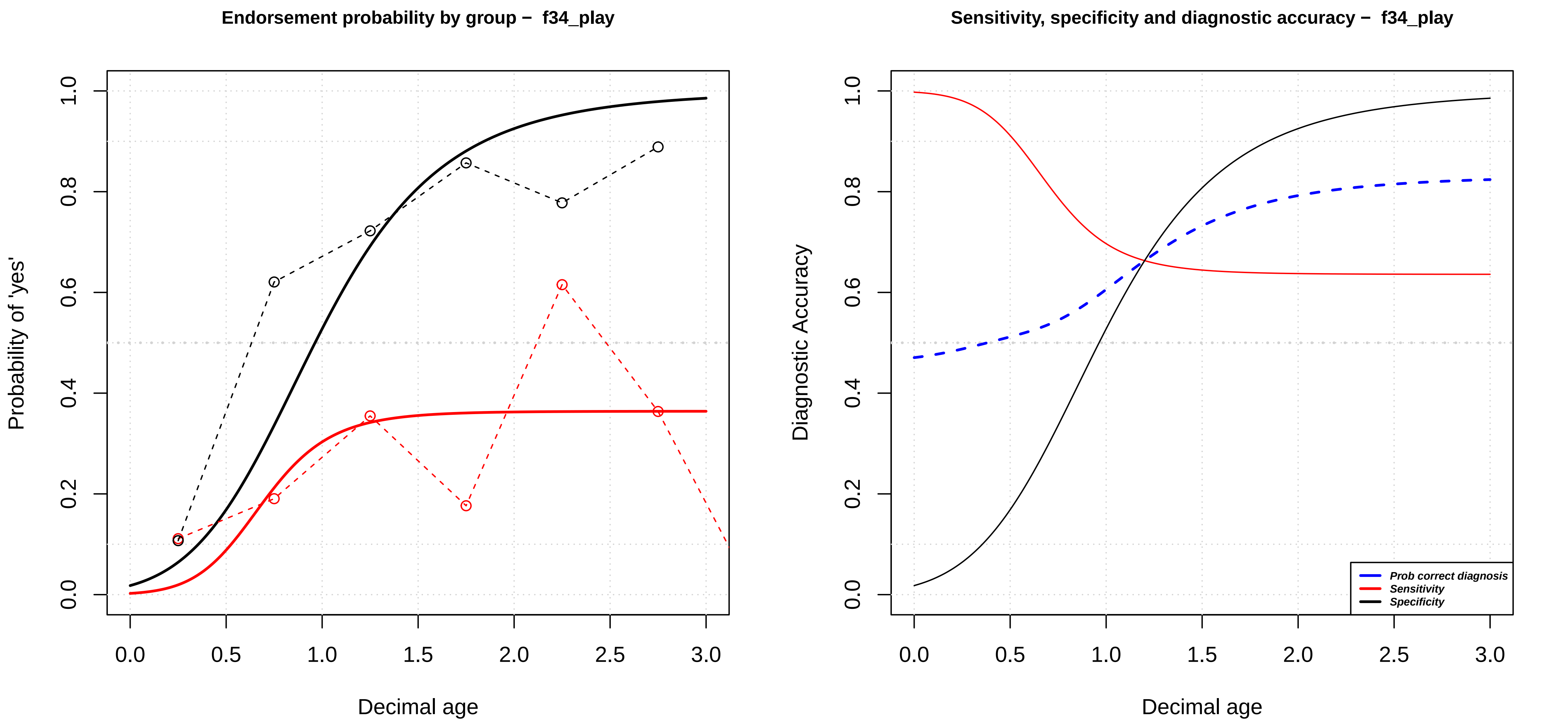
Endorsement probability by group – f34_play
Probability of 'yes'
Decimal age
0.0
0.2
0.4
0.6
0.8
1.0
0.0
0.5
1.0
1.5
2.0
2.5
3.0
Sensitivity, specificity and diagnostic accuracy – f34_play
Diagnostic Accuracy
Decimal age
Prob correct diagnosis
Sensitivity
Specificity

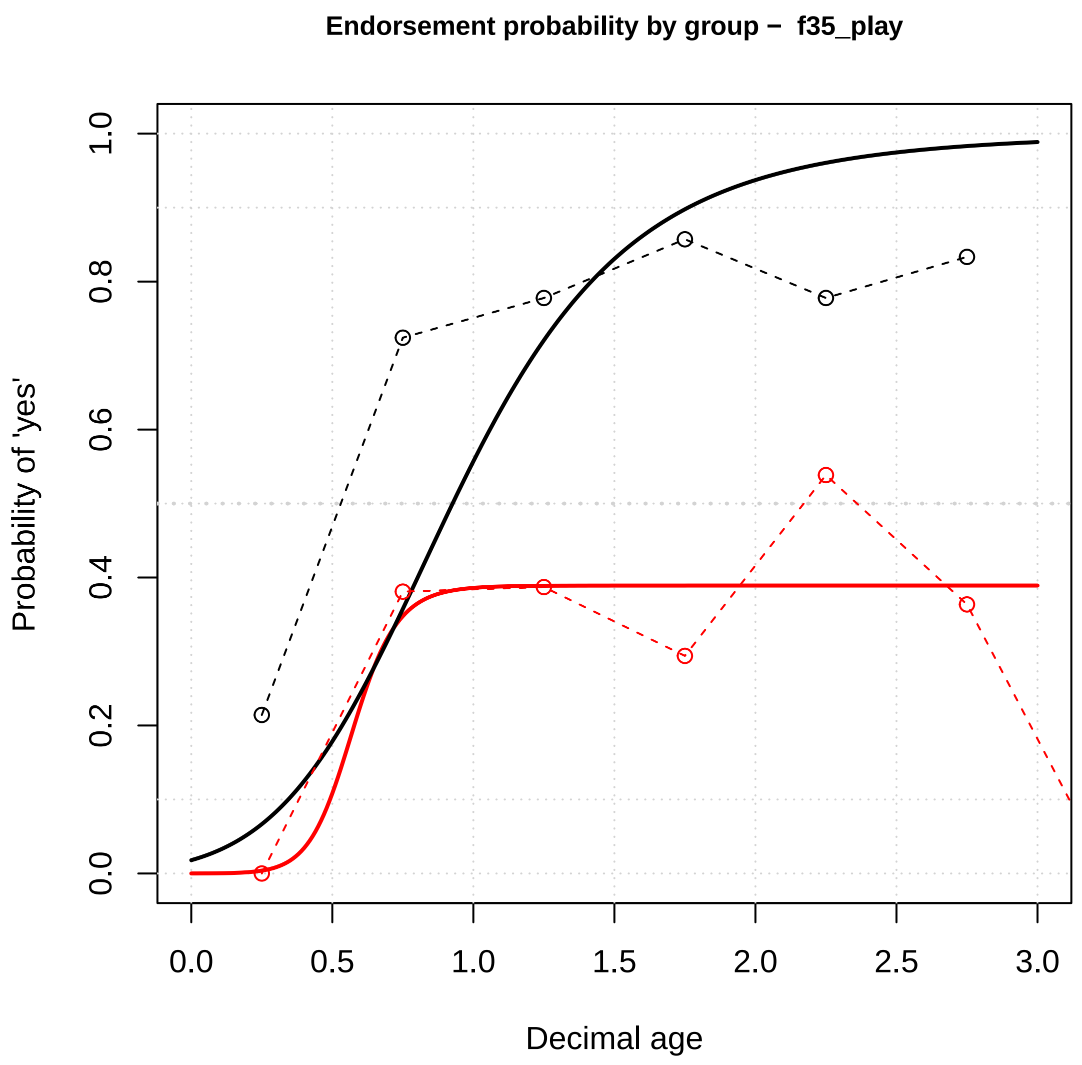

Endorsement probability by group – f35_play
Probability of 'yes'
Decimal age
0.0
0.2
0.4
0.6
0.8
1.0
0.0
0.5
1.0
1.5
2.0
2.5
3.0


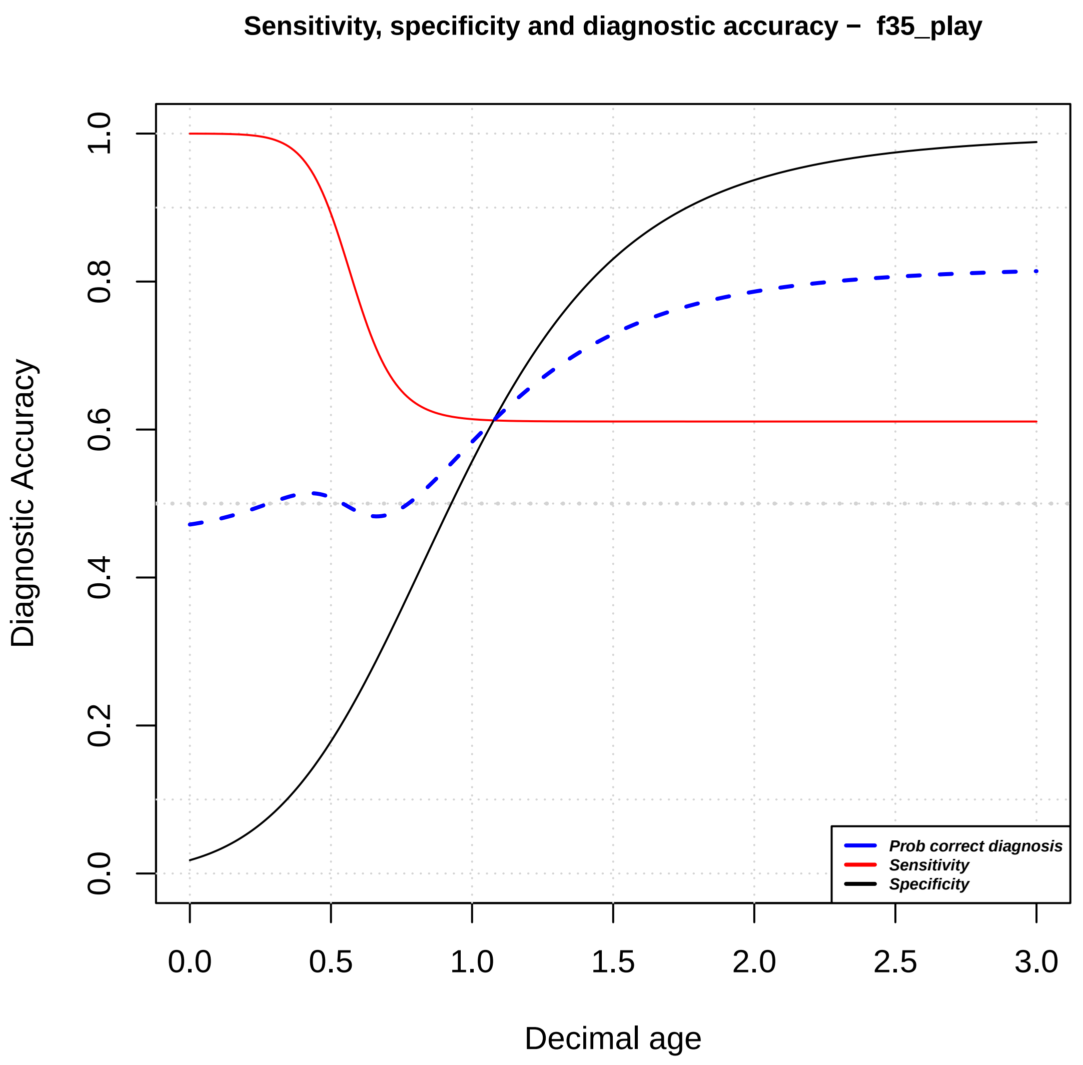

Sensitivity, specificity and diagnostic accuracy – f35_play
Diagnostic Accuracy
Decimal age
0.0
0.2
0.4
0.6
0.8
1.0
0.0
0.5
1.0
1.5
2.0
2.5
3.0
Prob correct diagnosis
Sensitivity
Specificity

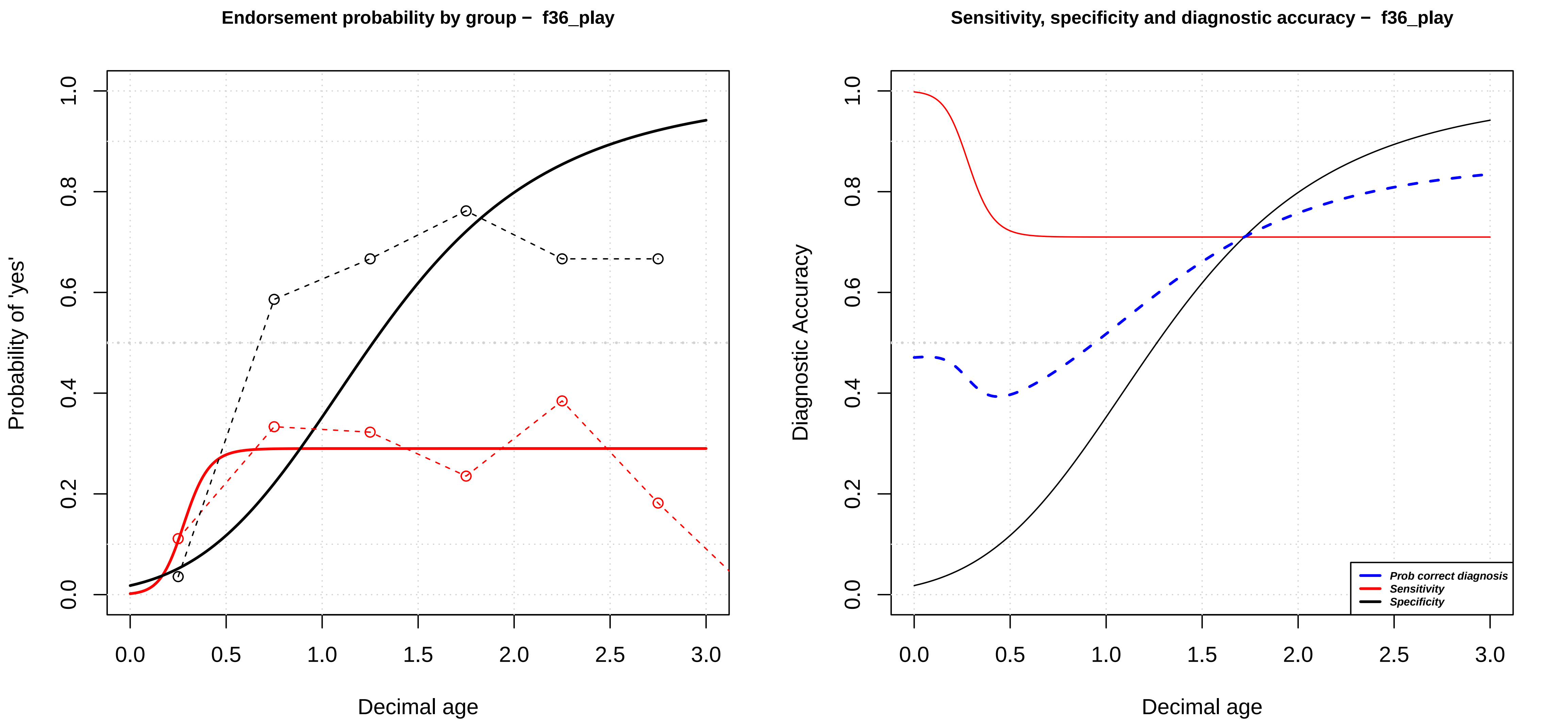

Endorsement probability by group – f36_play
Probability of 'yes'
0.0
0.2
0.4
0.6
0.8
1.0
0.0
0.5
1.0
1.5
2.0
2.5
3.0
Decimal age
Sensitivity, specificity and diagnostic accuracy – f36_play
Diagnostic Accuracy
Prob correct diagnosis
Sensitivity
Specificity
Decimal age

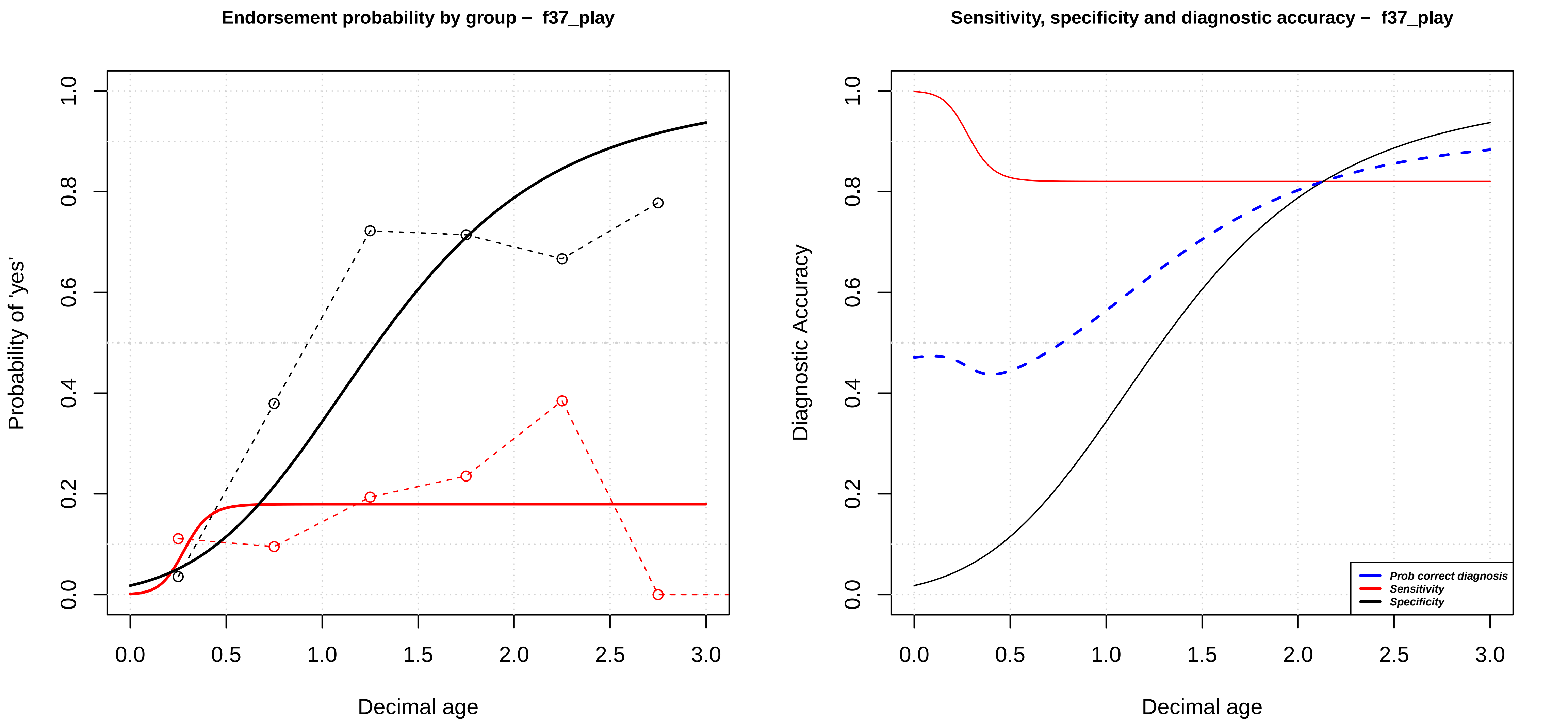

Endorsement probability by group – f37_play
Probability of 'yes'
Decimal age
0.0
0.2
0.4
0.6
0.8
1.0
0.0
0.5
1.0
1.5
2.0
2.5
3.0
Sensitivity, specificity and diagnostic accuracy – f37_play
Diagnostic Accuracy
Decimal age
Prob correct diagnosis
Sensitivity
Specificity

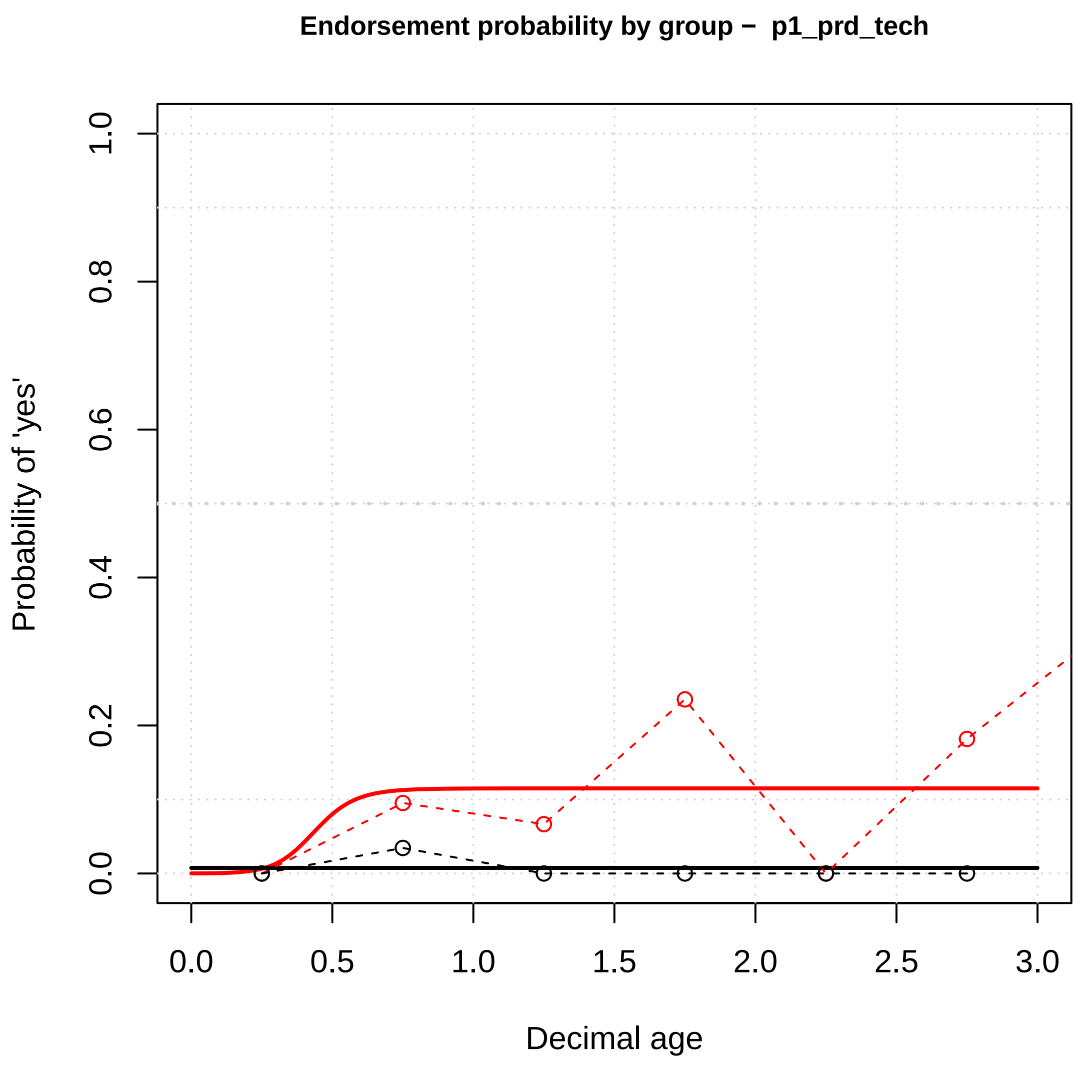

Endorsement probability by group – p1_prd_tech
Probability of 'yes'
Decimal age
0.0
0.2
0.4
0.6
0.8
1.0
0.5
1.5
2.0
2.5
3.0


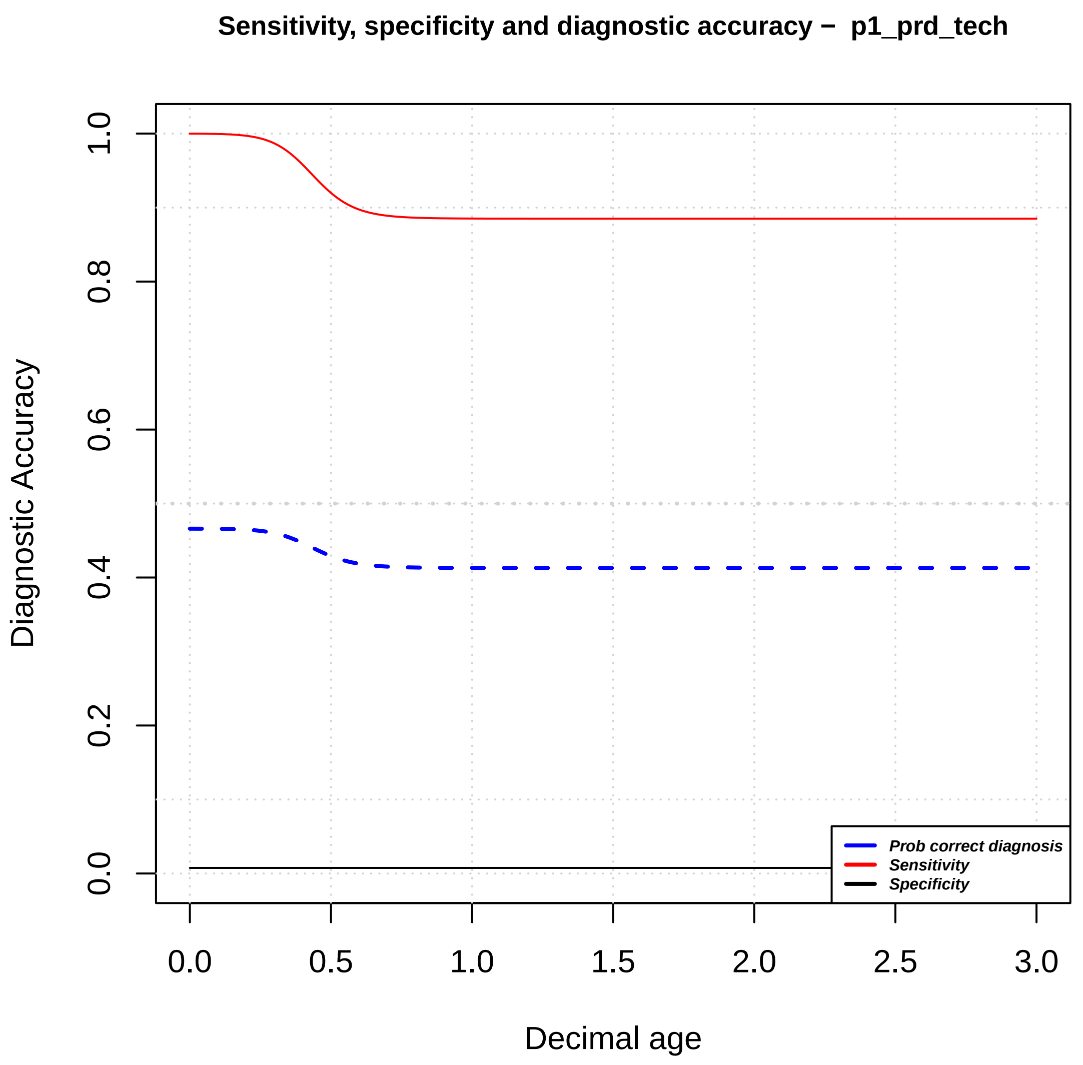

Sensitivity, specificity and diagnostic accuracy – p1_prd_tech
Diagnostic Accuracy
Decimal age
Prob correct diagnosis
Sensitivity
Specificity
0.0
0.2
0.4
0.6
0.8
1.0
0.5
1.5
2.0
2.5
3.0

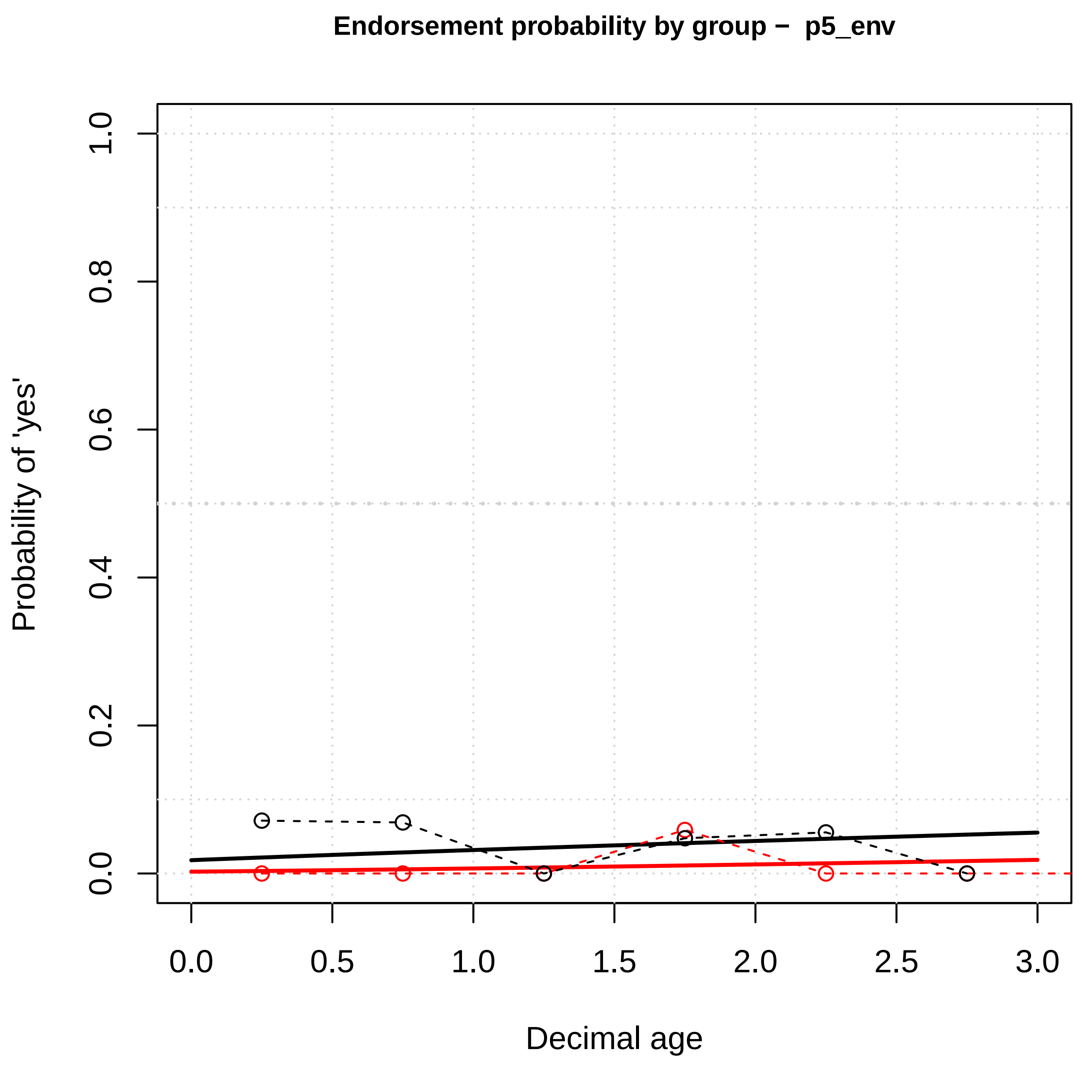
Endorsement probability by group – p5_env
Probability of 'yes'
Decimal age
0.0
0.2
0.4
0.6
0.8
1.0
0.5
1.5
2.0
2.5
3.0

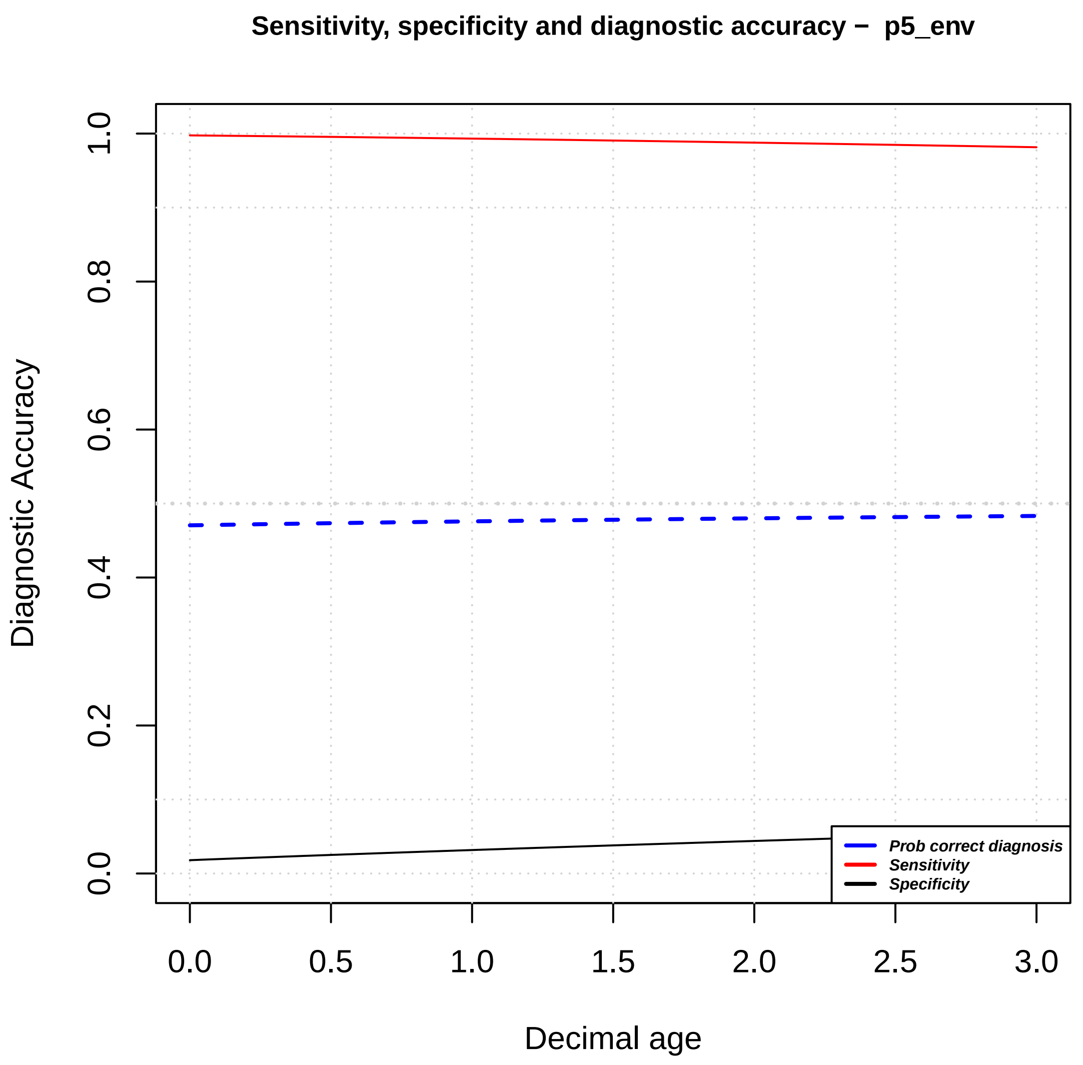
Sensitivity, specificity and diagnostic accuracy – p5_env
Diagnostic Accuracy
Decimal age
Prob correct diagnosis
Sensitivity
Specificity
0.0
0.2
0.4
0.6
0.8
1.0
0.5
1.5
2.0
2.5
3.0

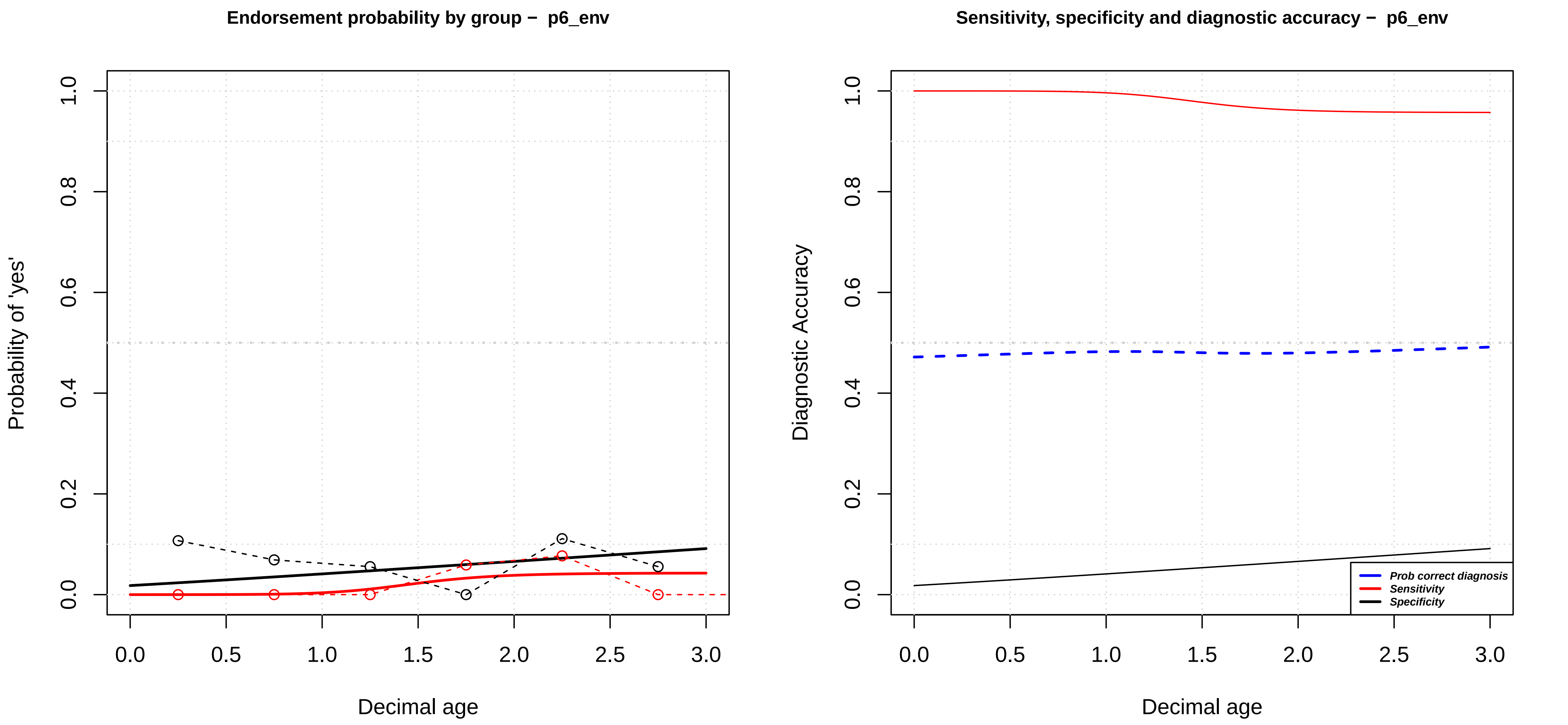
Endorsement probability by group – p6_env
Probability of 'yes'
0.0
0.2
0.4
0.6
0.8
1.0
0.0
0.5
1.0
1.5
2.0
2.5
3.0
Decimal age
Sensitivity, specificity and diagnostic accuracy – p6_env
Diagnostic Accuracy
Prob correct diagnosis
Sensitivity
Specificity
Decimal age

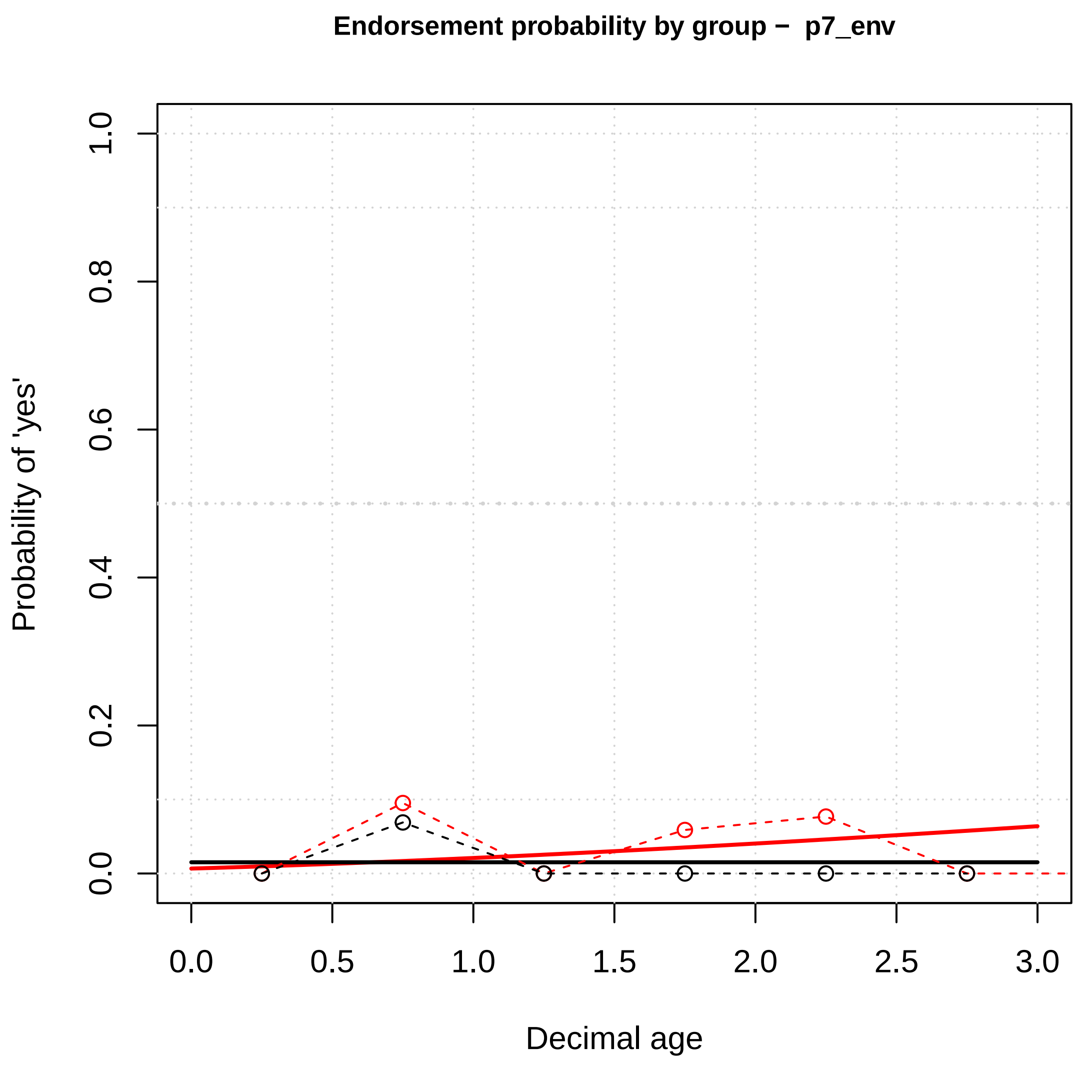

Endorsement probability by group − p7_env
Probability of 'yes'
0.0
0.2
0.4
0.6
0.8
1.0
0.0
0.5
1.0
1.5
2.0
2.5
3.0
Decimal age


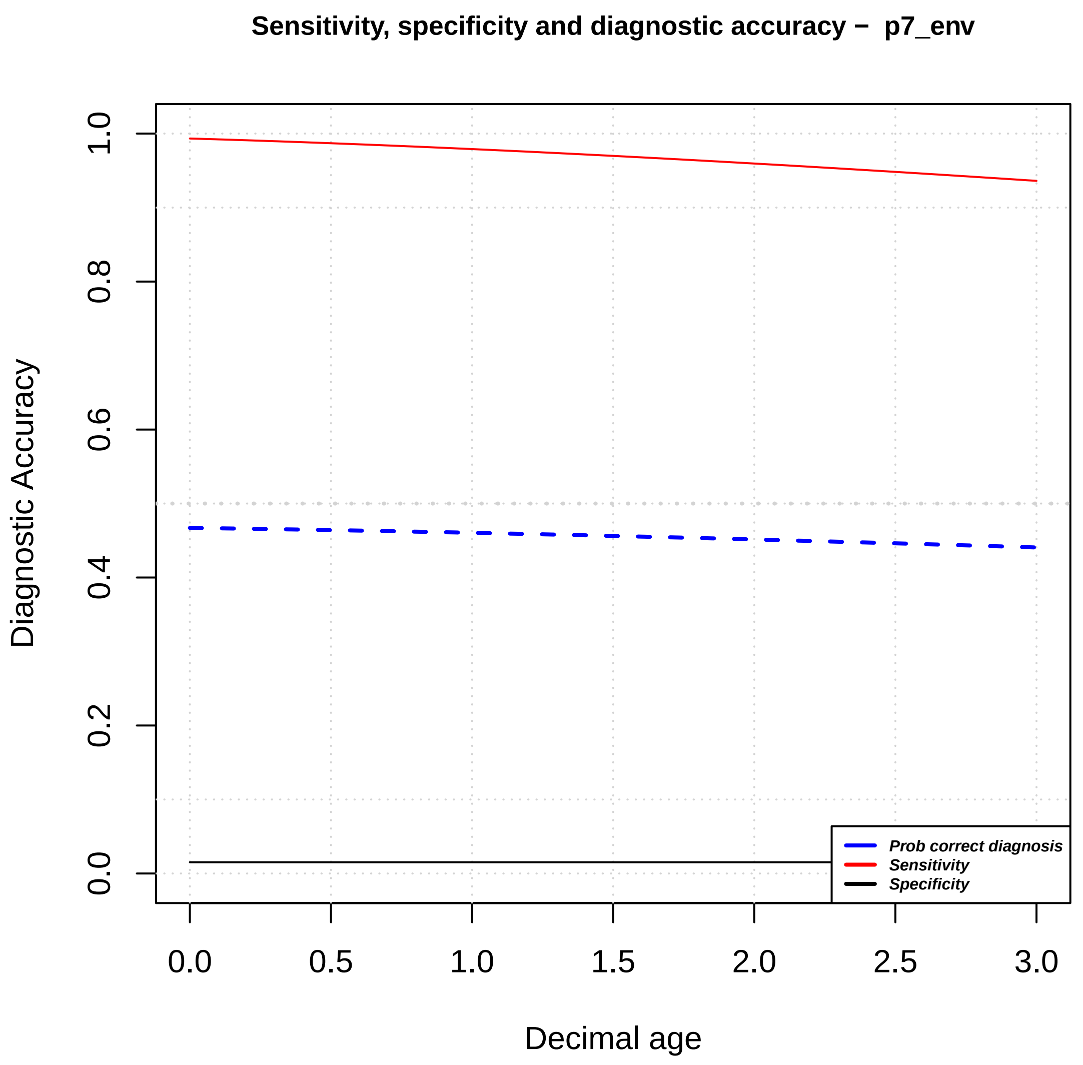

Sensitivity, specificity and diagnostic accuracy − p7_env
Diagnostic Accuracy
0.0
0.2
0.4
0.6
0.8
1.0
0.0
0.5
1.0
1.5
2.0
2.5
3.0
Decimal age
Prob correct diagnosis
Sensitivity
Specificity

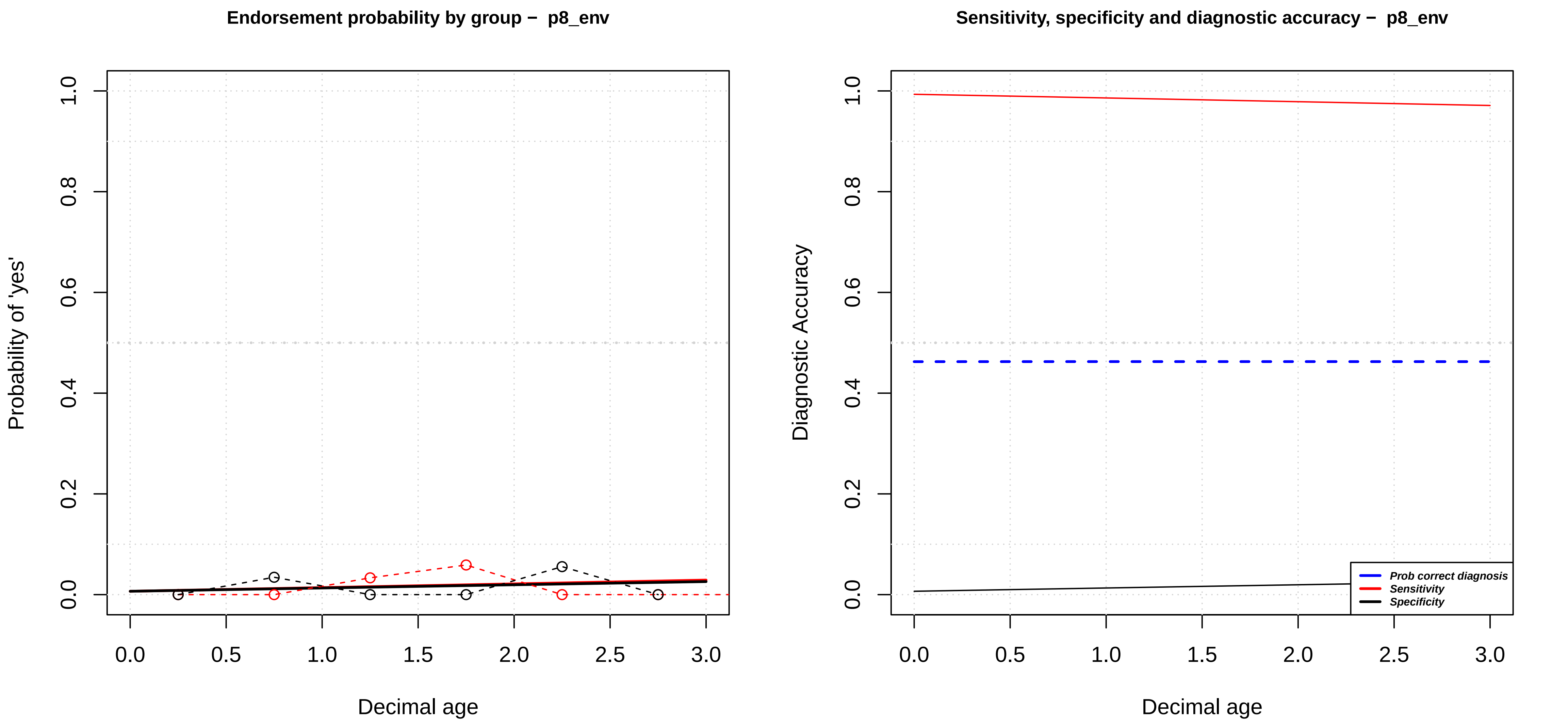

Endorsement probability by group − p8_env
Probability of 'yes'
0.0
0.2
0.4
0.6
0.8
1.0
0.0
0.5
1.0
1.5
2.0
2.5
3.0
Decimal age
Sensitivity, specificity and diagnostic accuracy − p8_env
Diagnostic Accuracy
Prob correct diagnosis
Sensitivity
Specificity
Decimal age

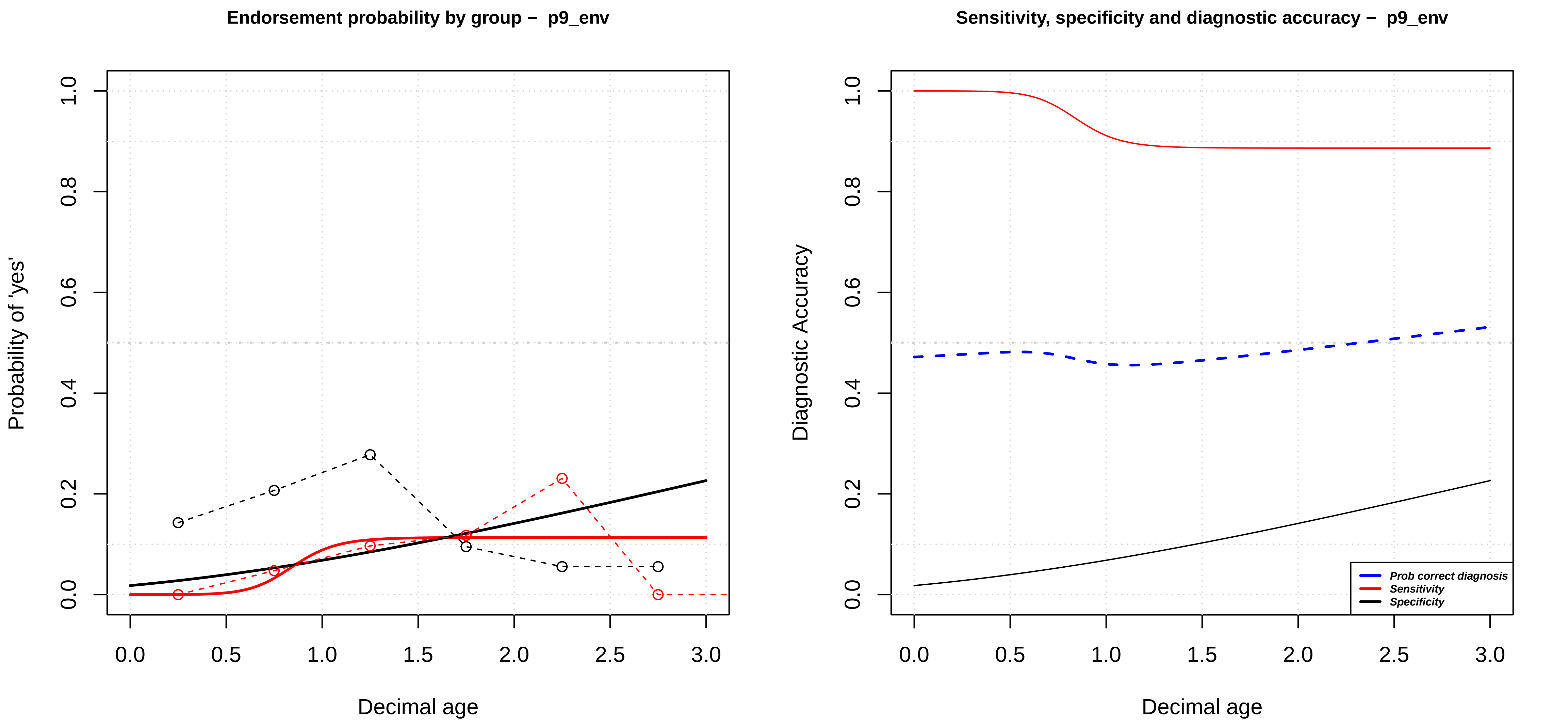

Endorsement probability by group – p9_env
Probability of 'yes'
Decimal age
0.0
0.2
0.4
0.6
0.8
1.0
0.0
0.5
1.0
1.5
2.0
2.5
3.0
Sensitivity, specificity and diagnostic accuracy – p9_env
Diagnostic Accuracy
Decimal age
Prob correct diagnosis
Sensitivity
Specificity

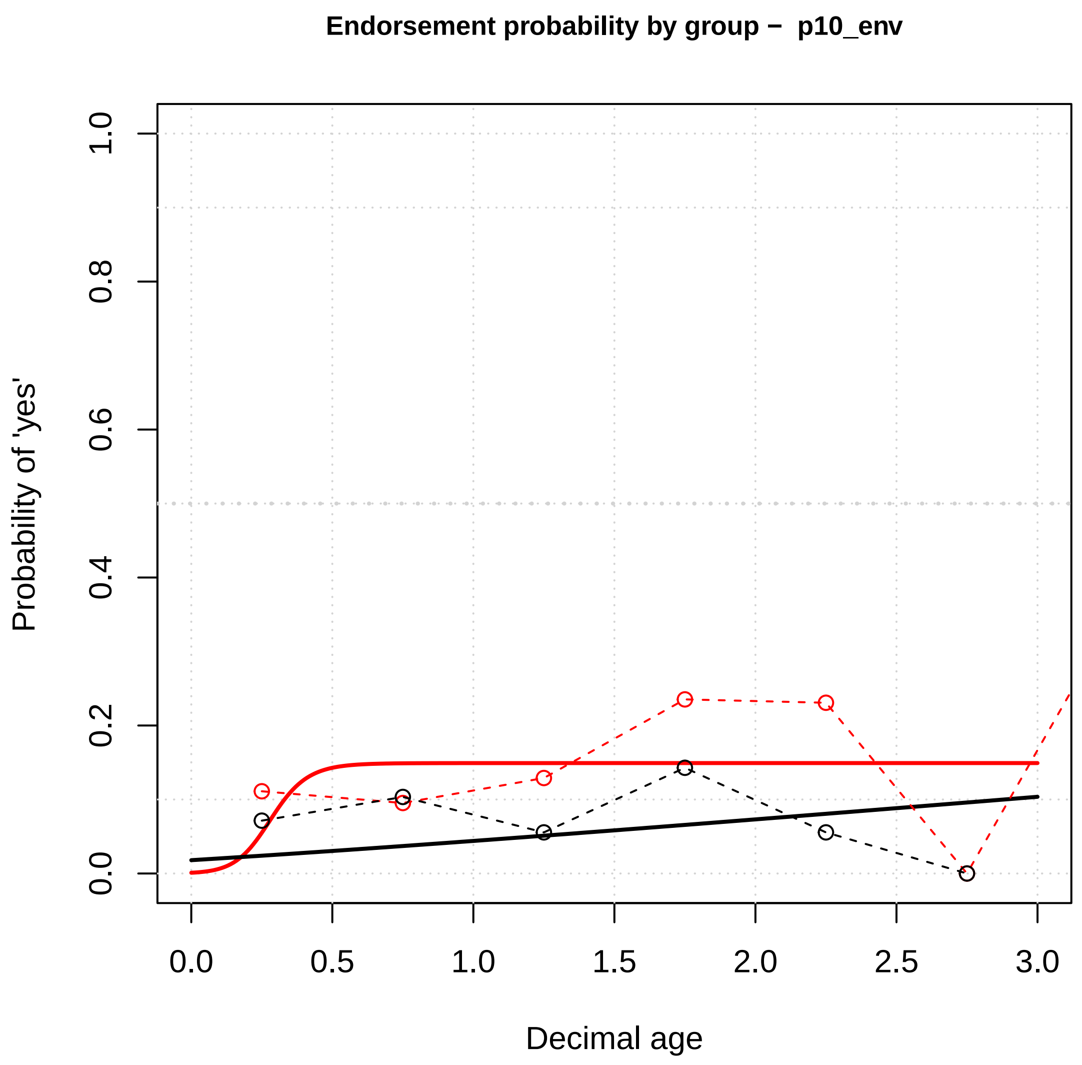

Endorsement probability by group – p10_env
Probability of 'yes'
Decimal age
0.0
0.2
0.4
0.6
0.8
1.0
0.0
0.5
1.0
1.5
2.0
2.5
3.0


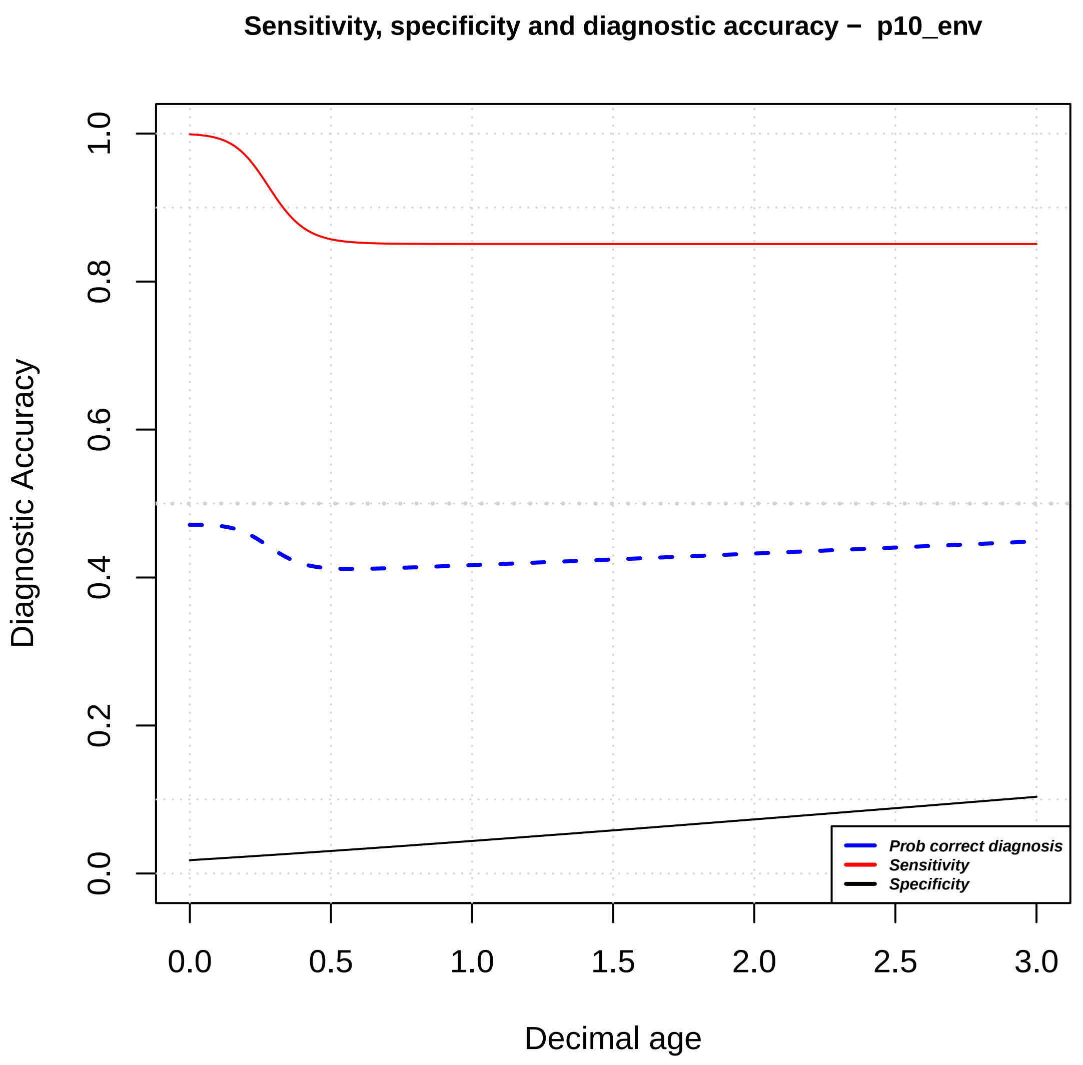

Sensitivity, specificity and diagnostic accuracy – p10_env
Diagnostic Accuracy
Decimal age
Prob correct diagnosis
Sensitivity
Specificity
0.0
0.2
0.4
0.6
0.8
1.0
0.0
0.5
1.0
1.5
2.0
2.5
3.0

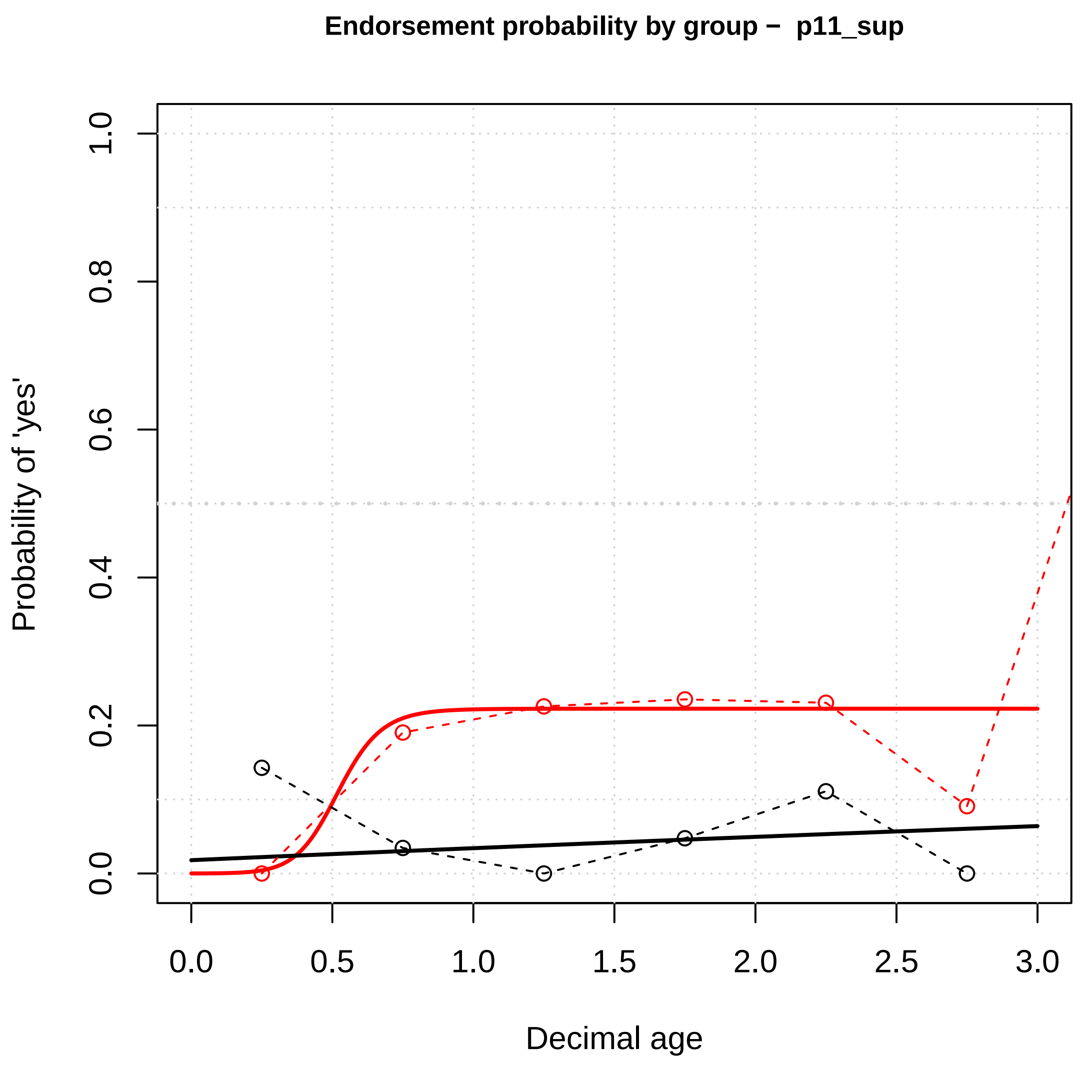
Endorsement probability by group – p11_sup
Probability of 'yes'
Decimal age
0.0
0.2
0.4
0.6
0.8
1.0
0.0
0.5
1.0
1.5
2.0
2.5
3.0

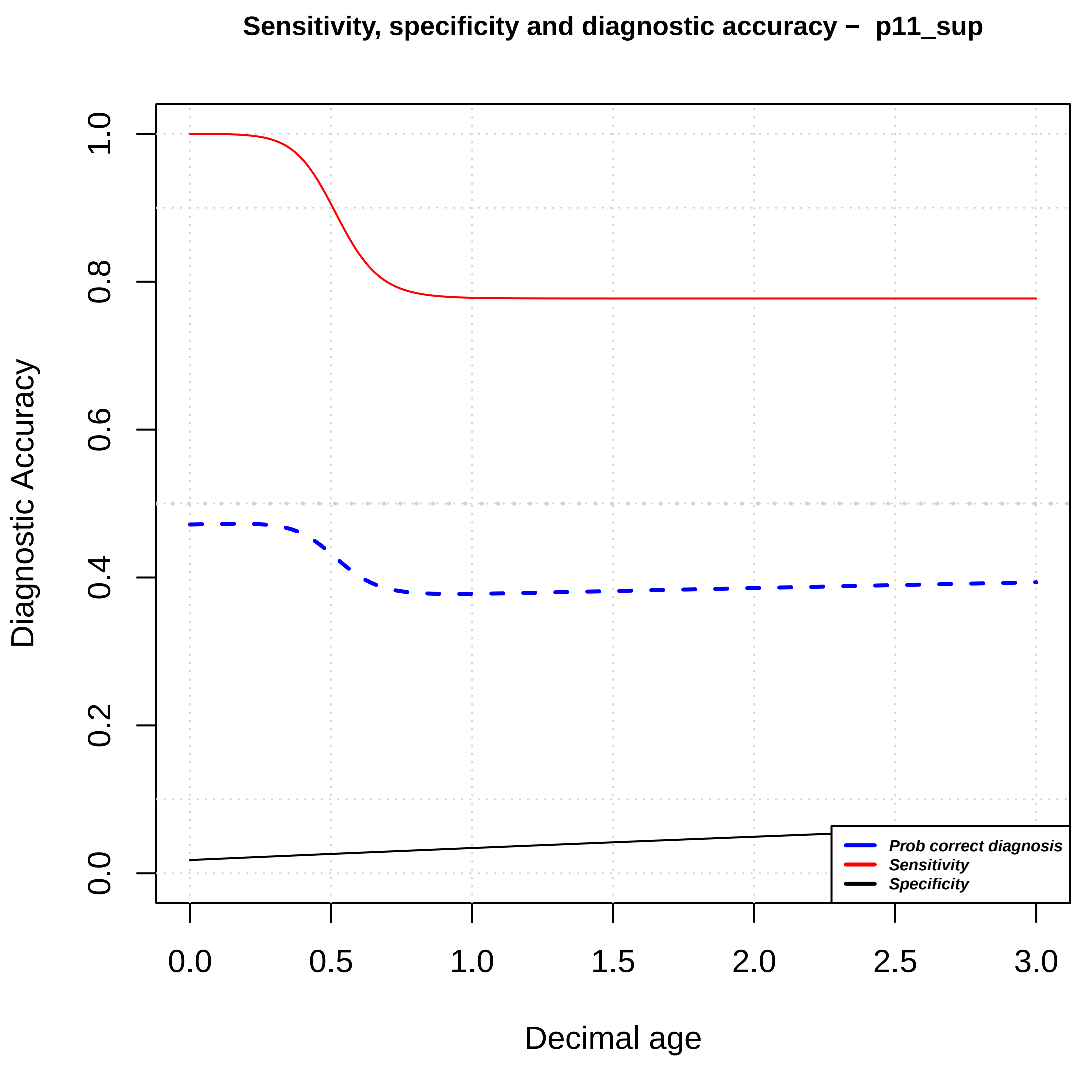
Sensitivity, specificity and diagnostic accuracy – p11_sup
Diagnostic Accuracy
Decimal age
0.0
0.2
0.4
0.6
0.8
1.0
0.0
0.5
1.0
1.5
2.0
2.5
3.0
Prob correct diagnosis
Sensitivity
Specificity

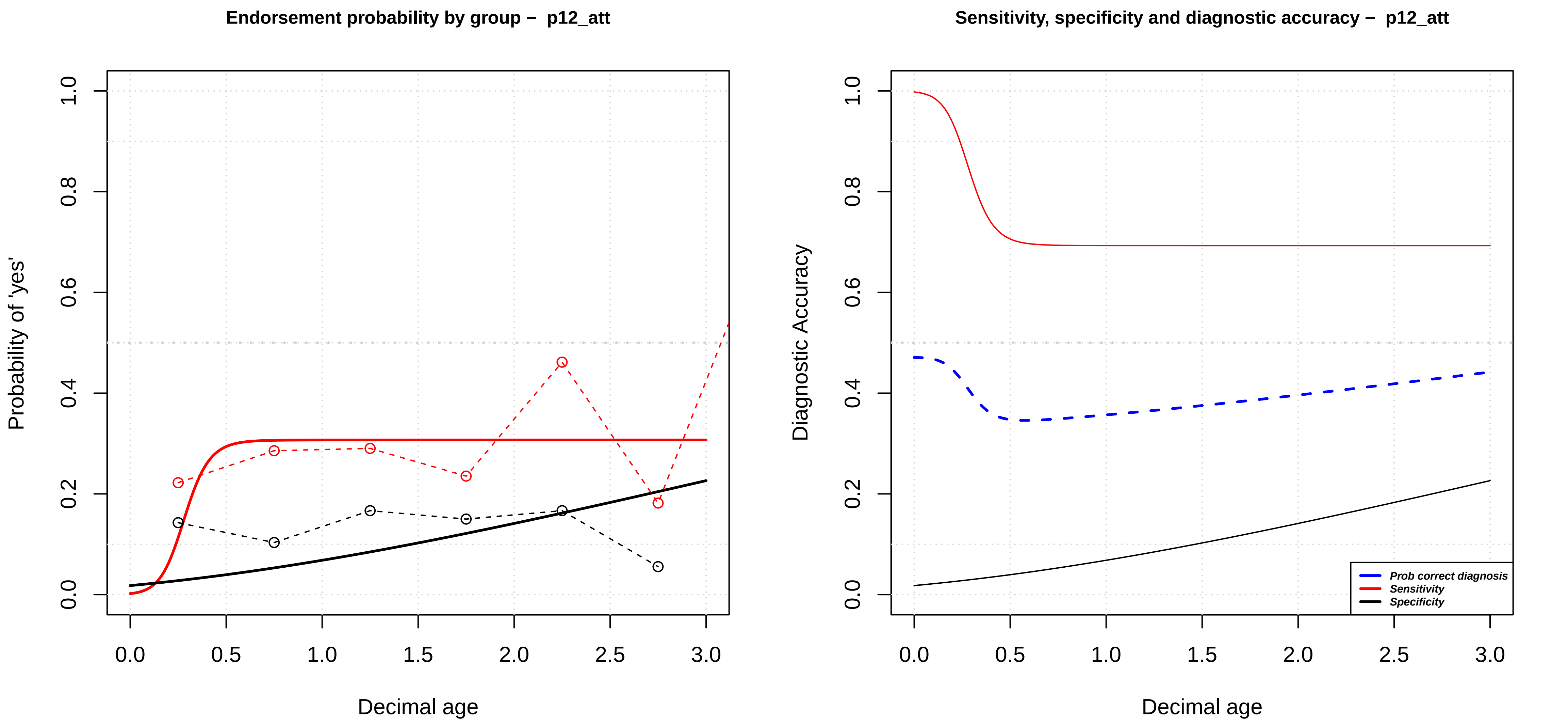

Endorsement probability by group − p12_att
Probability of 'yes'
Decimal age
Sensitivity, specificity and diagnostic accuracy − p12_att
Diagnostic Accuracy
Decimal age
Prob correct diagnosis
Sensitivity
Specificity
0.0
0.2
0.4
0.6
0.8
1.0
0.5
1.5
2.0
2.5
3.0

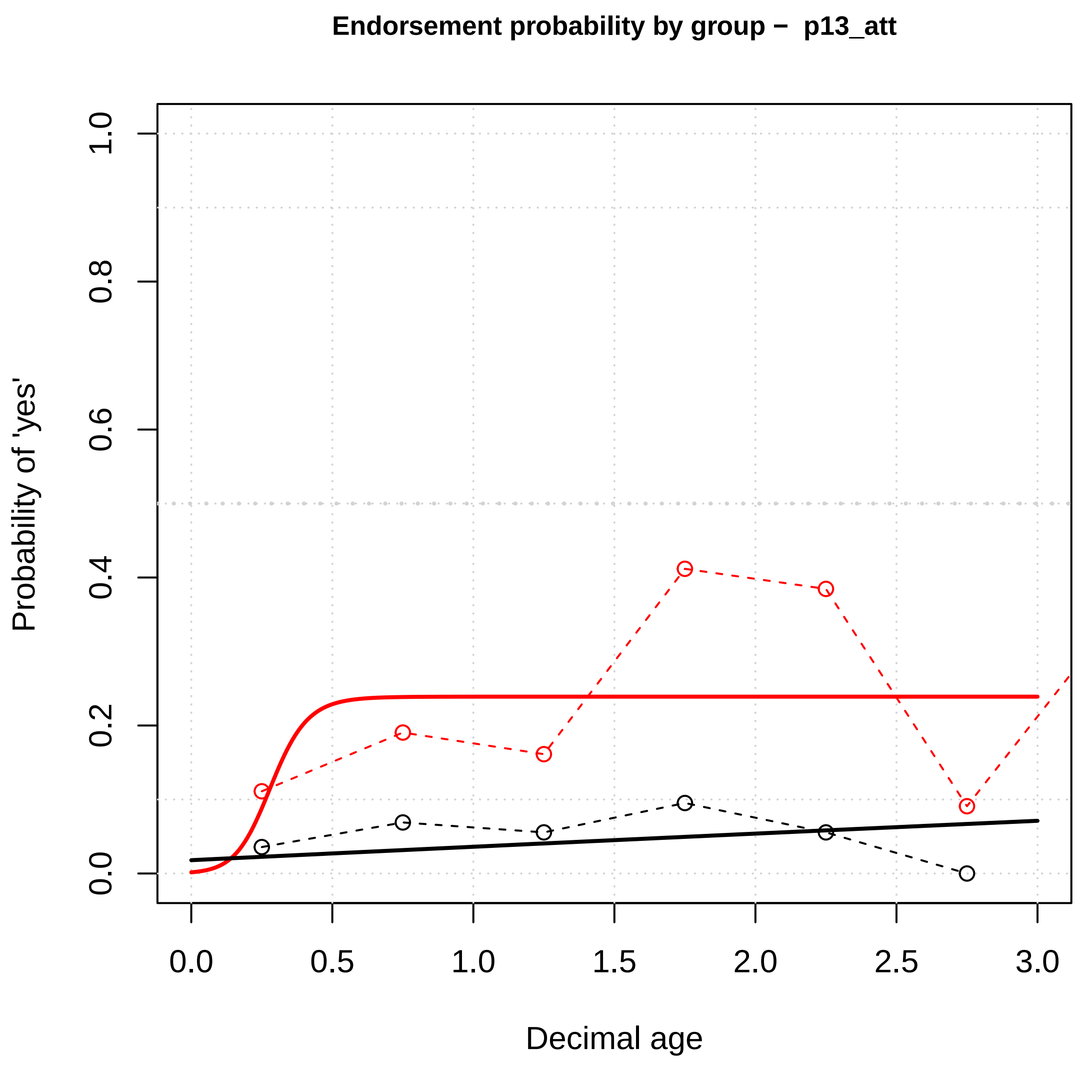

Endorsement probability by group – p13_att
Probability of 'yes'
0.0
0.2
0.4
0.6
0.8
1.0
0.0
0.5
1.0
1.5
2.0
2.5
3.0
Decimal age


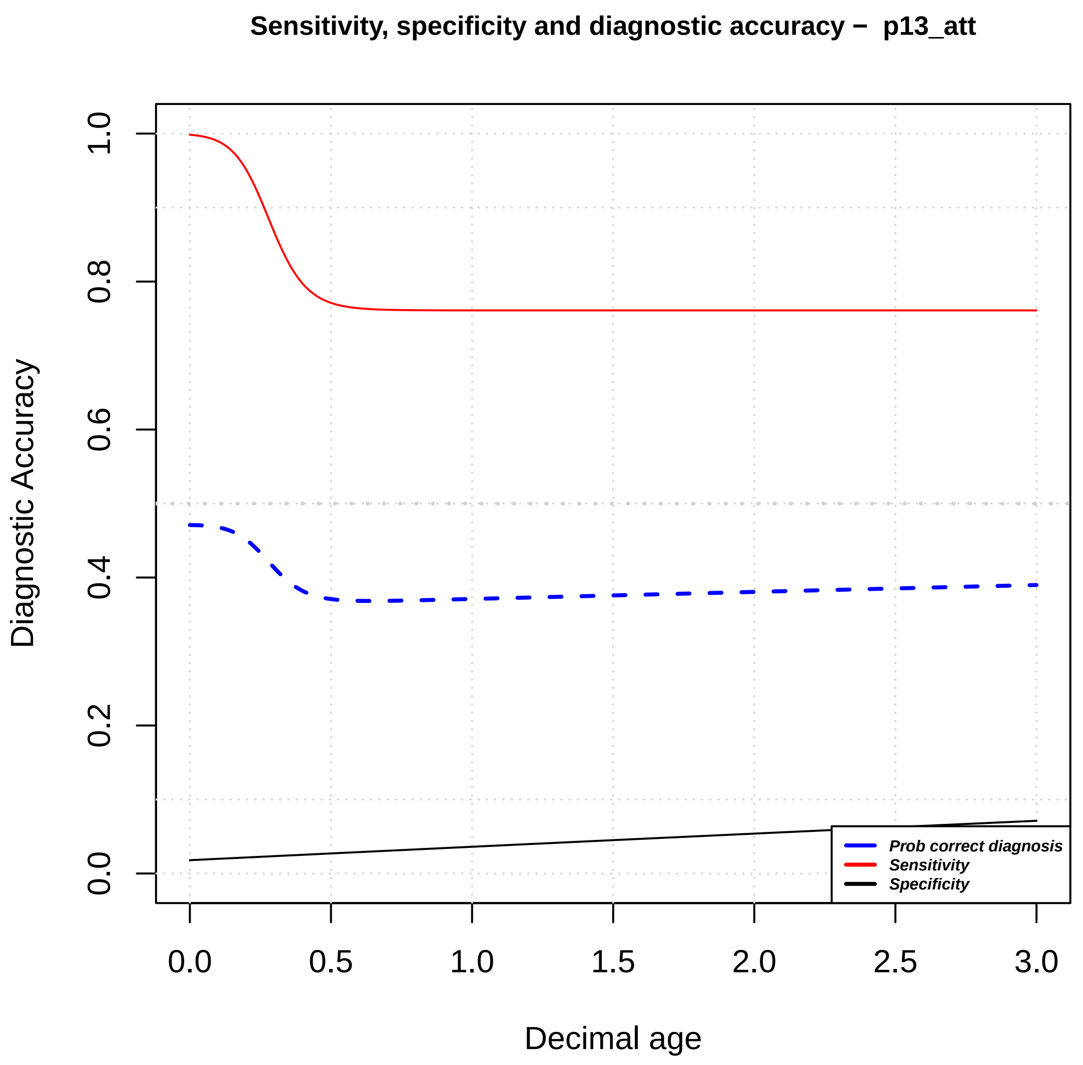

Sensitivity, specificity and diagnostic accuracy – p13_att
Diagnostic Accuracy
0.0
0.2
0.4
0.6
0.8
1.0
0.0
0.5
1.0
1.5
2.0
2.5
3.0
Decimal age
Prob correct diagnosis
Sensitivity
Specificity
